# Pharmacogenomics of steroid-induced ocular hypertension: relationship to high-tension glaucomas and new pathophysiologic insight

**DOI:** 10.1101/2025.08.11.25333245

**Authors:** Zeyuan Song, Satyabrata Pany, Shengru Guo, Arpan G. Mazumder, Tatsuo Itakura, Jonathan Huang, Evan Magarychoff, Anastasia Gurinovich, Penelope H. Benchek, W. Daniel Stamer, Francis W. Price, Colin E. Willoughby, Srinivasan Senthilkumari, Ronnie J. George, Srujana Chitipothu, Jonathan H. Lass, Sudha K. Iyengar, Stephen G. Schwartz, Anthony J. Griswold, Paola Sebastiani, Marianne O. Price, M. Elizabeth Fini

## Abstract

Adverse drug reactions are a frequent cause of worldwide morbidity and mortality. Glucocorticoids (GCs), commonly used to treat inflammatory diseases, alter gene expression with both beneficial and adverse consequences. When used in the eye, GCs cause steroid-induced ocular hypertension (SIOH) in 30-50% of patients, leading to steroid-induced glaucoma. Evidence suggests that predisposition to SIOH is genetically determined. Here we took a pharmacogenomic approach to discover DNA variants associated with SIOH. We identified 44 SNPs of genome-wide significance (p<5E-08) located at 26 risk loci out of a total of 531 SNPs of suggestive significance (p<5E-06) at 262 risk loci. Unlike SNPs identified in complex disease which are overwhelmingly common in frequency, most SNPs found here were rare or of low frequency, likely discoverable because of their large effect sizes. Follow-up analyses provide insight into the pathogenetic relationship of SIOH to high-tension glaucomas and suggest a new mechanistic paradigm for SIOH pathophysiology.

**Graphical Abstract:** 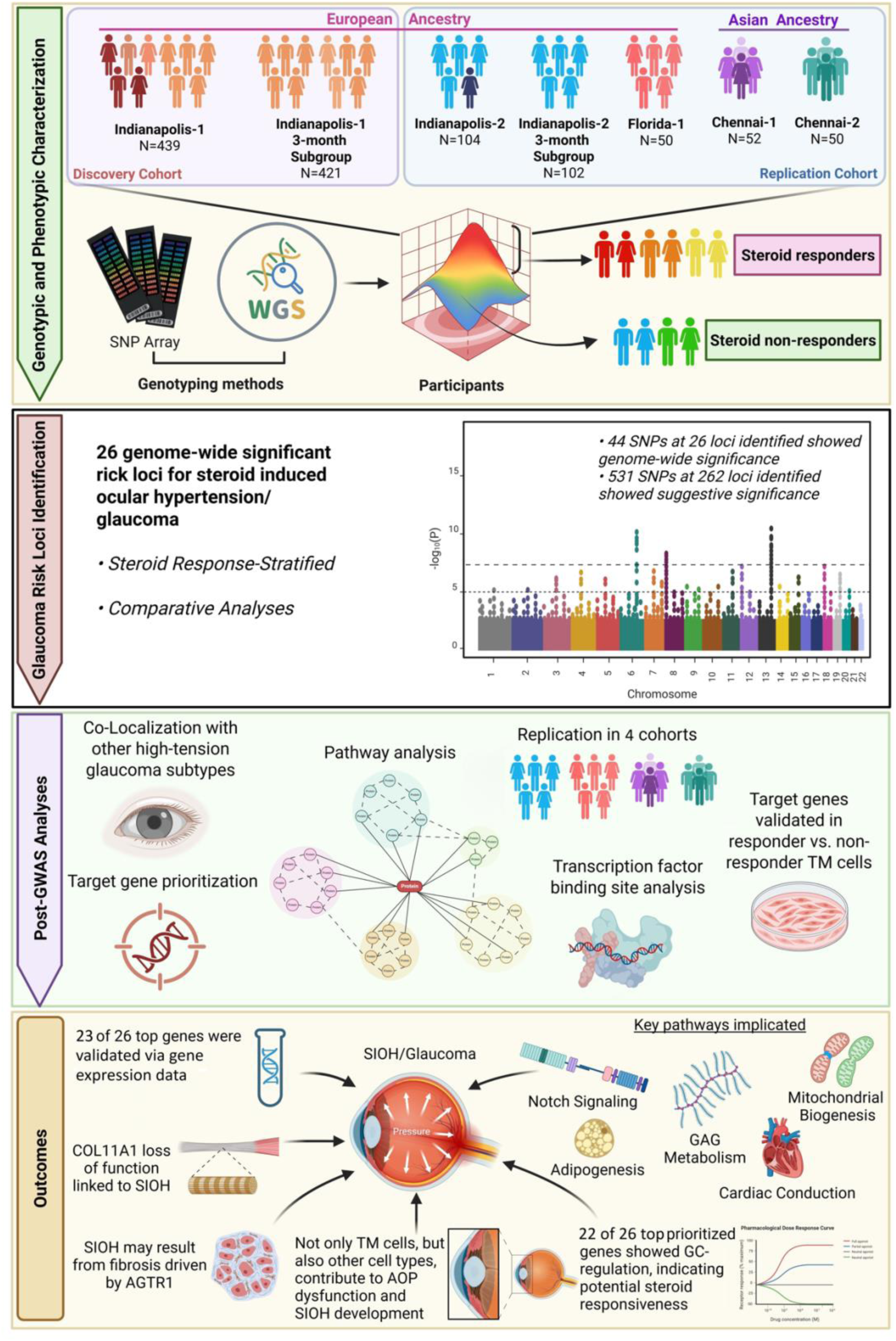

## INTRODUCTION

Adverse drug reactions are a frequent cause of worldwide morbidity and mortality^1^. A common example is response to glucocorticoids (GCs)^2,3^. GCs comprise a class of steroid hormones whose physiological role is to regulate carbohydrate, lipid and protein metabolism, electrolyte balance, and vascular tone. GCs also regulate the immune response and curtail inflammation, making them very effective therapeutics^4–8^. However, in predisposed individuals, systemic GC treatment can cause immunosuppression, muscle atrophy, central adiposity, hepatosteatosis, osteoporosis, insulin resistance, hypertension, depression and insomnia^2,3^. In addition, 30-50% of adults and almost all children develop elevated intraocular pressure (IOP), also known as ocular hypertension (OH), when treated with GCs in the eye^9,10^. If steroid-induced OH (SIOH) is untreated in these “steroid responders”, it can lead to steroid-induced glaucoma.

The glaucomas are a group of optic neuropathies characterized by optic nerve head “cupping” due to degeneration and loss of retinal ganglion cells. They comprise the leading cause of irreversible blindness worldwide among people over the age of 55^11^. Despite increasing recognition of a disease subtype called normal tension glaucoma (NTG), elevated IOP is still the major risk factor^12–15^ and lowering IOP is still the only effective treatment^16–18^, even in those with NTG^19,20^. IOP is the product of the rate of aqueous humor secretion by the ciliary body, resistance to its outflow from the eye localized to the juxtacanalicular tissue (JCT) of the trabecular meshwork (TM) that abuts Schlemm’s canal^21^, and episcleral venous pressure^22^. TM cells maintain contractile tissue tone in conjunction with the ciliary muscle to regulate outflow^23^. All forms of OH are caused by an increase in outflow resistance^24,25^. In primary angle closure glaucoma (PACG), outflow dysfunction is primarily due to physical obstruction by nearby tissues, while dysfunction in high-tension forms of primary open angle glaucoma (POAG) is a result of molecular, cellular and tissue changes in the aqueous outflow pathways (AOPs). This is also true for SIOH, a secondary OAG^9,10^.

The primary action of GCs is to regulate transcription^7^. GCs diffuse into cells and bind the glucocorticoid receptor (GR) resulting in its activation and translocation from the cytoplasm to the nucleus. There the activated GR acts as a transcription factor (TF) with two mechanistic options. As a monomer, it may bind with other TFs to interfere with their activation of specific genes. This is called transrepression, and is the major mechanism by which GCs curtail inflammation. As a dimer (or two monomers), the GR may also bind directly to consensus GC response elements on DNA, leading to transcriptional transactivation (or inhibition) of nearby genes. This second option has been linked to adverse reactions in susceptible individuals^26–28^, including when GCs are delivered directly to the eye^29^.

Differences in systemic sensitivity to GCs are determined by a combination of mutations in genes involved in GC response mechanics. This includes alterations in levels and isoforms of *NR3C1* encoding the GR, as well as its coactivators/corepressors; genes involved in metabolic clearance rates; mutations affecting 11β-hydroxysteroid dehydrogenase activity, and mutations in GC-binding proteins^30^. Disappointingly, these mutations do not appear to be particularly relevant to SIOH^31–35^. Coding mutations in *MYOC*, a GR target gene expressed in TM cell cultures^36,37^, causes high tension POAG^38–40^. However, *MYOC* does not contribute to SIOH^41,42^.

Treatment of cultured TM cells with GCs increases biosynthetic activity and leads to fibrosis^10^. Similar changes are seen in the AOPs when GCs are used in the eye^43–55^. TM cells also become less phagocytic and their numbers decline^10^. Together these changes lead to tissue stiffening^9,10,56^. Mouse studies also implicate the unfolded protein response (UPR) activated by fibrosis in the pathophysiology of SIOH^58–61^. Such fibrotic effects of GC treatment occur in all individuals, however it is suggested that excessive GC-induced fibrosis may tilt the balance to pathology in steroid responders^57^. Very similar changes are observed in high tension POAG^10^. Significantly, almost all POAG patients are steroid responders^62–69^. Moreover, steroid responders who do not have POAG are at higher risk of developing POAG compared with non-responders, and individuals with a family history of POAG are more likely to develop SIOH^65,70^. One suggested explanation for these findings is that SIOH susceptibility reflects underlying high-tension POAG^10^. Yet, while the the pathophysiology of these two diseases share many similarities, there are also distinct differences, most notably in the location and composition of the ECM deposits^9^.

Glaucomas of all subtypes are highly heritable^71^ and the genome-wide association study (GWAS) approach has been used very successfully to discover risk loci^72,73^. Typical of complex disease, SNPs associated with POAG are found almost entirely in non-coding regions of the genome and likely regulate gene expression^74–76^. Complex diseases like the glaucomas exhibit relatively small effect sizes meaning that very large cohorts must be enrolled. SNPs discovered thus far are of common frequency, accounting only for a small percentage of the estimated heritability in the case of POAG^77^. Rare variants have been proposed as a source of the missing heritability, but finding them is a challenge.

To identify GC-induced changes in gene expression that might be responsible for tipping the balance to pathology in SIOH, much previous work has employed a gene profiling approach^92,119–124^. Most recently a paired eye study design has been used that couples gene expression profiling to aqueous outflow measurement to identify steroid responders^96–98^. However the GWAS is a superior approach since it can identify causative genes, while gene profiling can only make correlations. Drug response typically manifests much larger effect sizes over a much shorter observation period than complex disease^78^ and this feature increases the potential to reveal rare variants^79^. Thus a study of SIOH can uncover rare SNPs linked to many new target genes, providing new hypothesis-generating information about aqueous outflow and its deficiency. Importantly, measurement of IOP, a routine component of clinical care, provides the basis for a quantitative trait (QT) that can be treated as a continuous variable, more powerful statistically than case-control study designs^80^.

Enrollment of large and uniform cohorts of GC-treated patients has proven elusive^9^, however small pilot studies conducted independently by different members of our team yielded variants of genome-wide significance^81,82^. Recognizing the opportunity of a “pharmacogenomics” approach to discover rare variants, our teams combined resources for a larger study, the successful results of which are reported here. We identified a remarkable 44 SNPs of genome-wide significance (p<5E-08) located at 26 risk loci, out of a total of 531 SNPs of suggestive significance (p<5E-06) at 262 risk loci. The majority of the SNPs discovered were rare. Follow-up analyses provide insight into the pathogenetic relationship of SIOH to high-tension glaucomas and suggest a new mechanistic paradigm for SIOH pathophysiology.

## RESULTS

### Cohorts and calculation of quantitative traits

Demographic and clinical data, as well as GWAS specific information on discovery and replication cohorts are shown in Table 1. Maximum change in IOP from baseline (deltaIOPmax) was used as the QT. The Indianapolis-1 discovery and Indianapolis-2 replication cohorts were stratified for the calculation of deltaIOPmax at 12 months (*n*=439) and deltaIOPmax at 3 months (*n*=421).

**Table 1.**
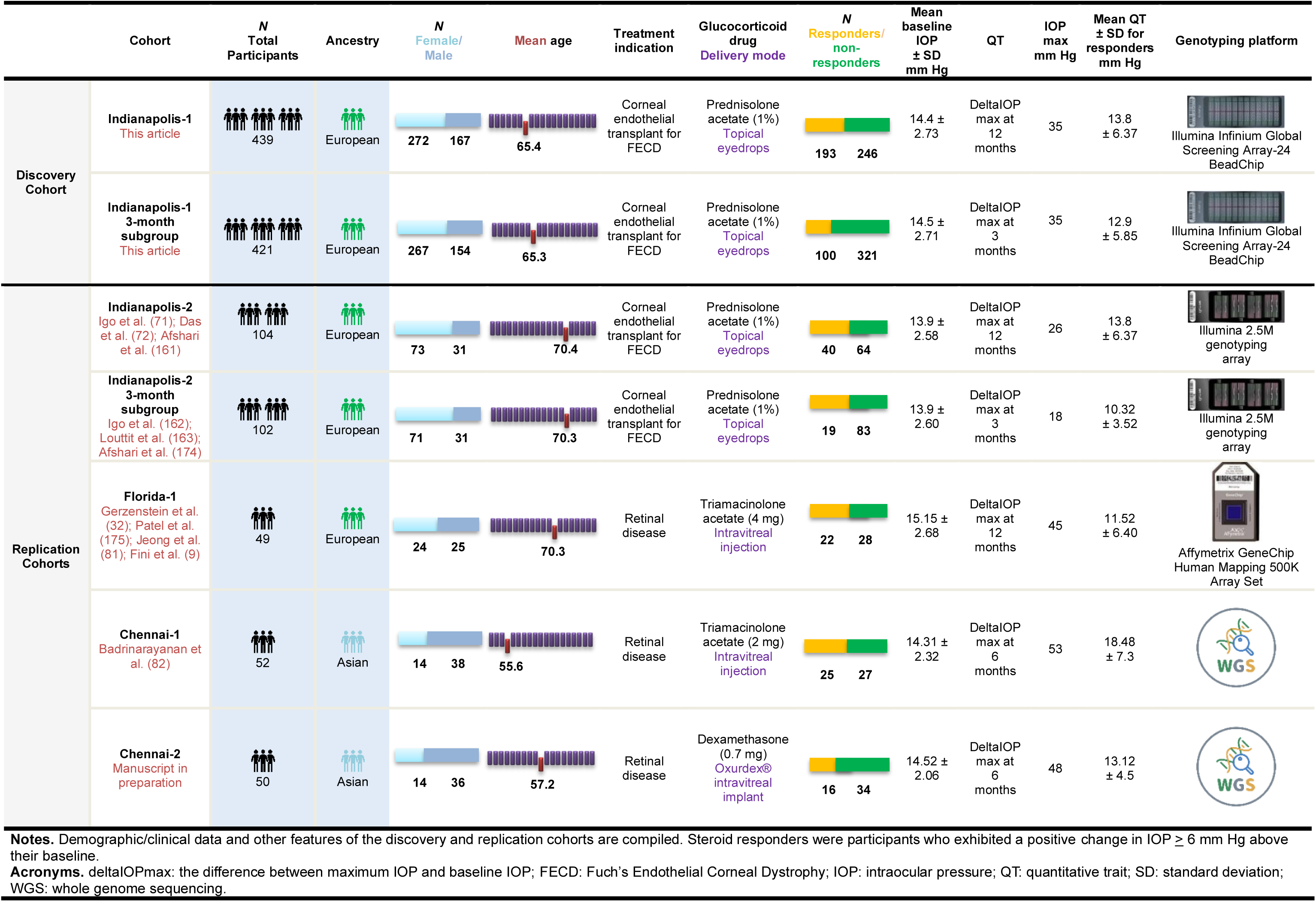
Cohorts.

|  | Cohort | N<br>Total<br>Participants | Ancestry | N<br>Female/<br>Male | Mean age | Treatment<br>indication | Glucocorticoid<br>drug<br>Delivery mode | N<br>Responders/<br>non-responders | Mean<br>baseline<br>IOP<br>± SD<br>mm Hg | QT | IOP<br>max<br>mm Hg | Mean QT<br>± SD for<br>responders<br>mm Hg | Genotyping platform |
| --- | --- | --- | --- | --- | --- | --- | --- | --- | --- | --- | --- | --- | --- |
| Discovery<br>Cohort | Indianapolis-1<br>*This article* | 439 | European | 272 167 | 65.4 | Corneal<br>endothelial<br>transplant for<br>FECD | Prednisolone<br>acetate (1%)<br>Topical<br>eyedrops | 193 246 | 14.4 ±<br>2.73 | DeltaIOP<br>max at<br>12<br>months | 35 | 13.8<br>± 6.37 | Illumina Infinium Global<br>Screening Array-24<br>BeadChip |
|  | Indianapolis-1<br>3-month<br>subgroup<br>*This article* | 421 | European | 267 154 | 65.3 | Corneal<br>endothelial<br>transplant for<br>FECD | Prednisolone<br>acetate (1%)<br>Topical<br>eyedrops | 100 321 | 14.5 ±<br>2.71 | DeltaIOP<br>max at<br>3<br>months | 35 | 12.9<br>± 5.85 | Illumina Infinium Global<br>Screening Array-24<br>BeadChip |
| Replication<br>Cohorts | Indianapolis-2<br>*Igo et al. (71); Das*<br>*et al. (72); Afshari*<br>*et al. (161)* | 104 | European | 73 31 | 70.4 | Corneal<br>endothelial<br>transplant for<br>FECD | Prednisolone<br>acetate (1%)<br>Topical<br>eyedrops | 40 64 | 13.9 ±<br>2.58 | DeltaIOP<br>max at<br>12<br>months | 26 | 13.8<br>± 6.37 | Illumina 2.5M<br>genotyping<br>array |
|  | Indianapolis-2<br>3-month<br>subgroup<br>*Igo et al. (162);*<br>*Louttit et al. (163);*<br>*Afshari et al. (174)* | 102 | European | 71 31 | 70.3 | Corneal<br>endothelial<br>transplant for<br>FECD | Prednisolone<br>acetate (1%)<br>Topical<br>eyedrops | 19 83 | 13.9 ±<br>2.60 | DeltaIOP<br>max at<br>3<br>months | 18 | 10.32<br>± 3.52 | Illumina 2.5M<br>genotyping<br>array |
|  | Florida-1<br>*Gerzenstein et al.*<br>*(32); Patel et al.*<br>*(175); Jeong et al.*<br>*(81); Fini et al. (9)* | 49 | European | 24 25 | 70.3 | Retinal<br>disease | Triamcinolone<br>acetate (4 mg)<br>Intravitreal<br>injection | 22 28 | 15.15 ±<br>2.68 | DeltaIOP<br>max at<br>12<br>months | 45 | 11.52<br>± 6.40 | Affymetrix GeneChip<br>Human Mapping 500K<br>Array Set |
|  | Chennai-1<br>*Badrinarayanan et*<br>*al. (82)* | 52 | Asian | 14 38 | 55.6 | Retinal<br>disease | Triamcinolone<br>acetate (2 mg)<br>Intravitreal<br>injection | 25 27 | 14.31 ±<br>2.32 | DeltaIOP<br>max at<br>6<br>months | 53 | 18.48<br>± 7.3 | WGS |
|  | Chennai-2<br>*Manuscript in*<br>*preparation* | 50 | Asian | 14 36 | 57.2 | Retinal<br>disease | Dexamethasone<br>(0.7 mg)<br>Oxurdex®<br>intravitreal<br>implant | 16 34 | 14.52 ±<br>2.06 | DeltaIOP<br>max at<br>6<br>months | 48 | 13.12<br>± 4.5 | WGS |

**Notes.** Demographic/clinical data and other features of the discovery and replication cohorts are compiled. Steroid responders were participants who exhibited a positive change in IOP $\geq 6$ mm Hg above their baseline.
**Acronyms.** deltaIOPmax: the difference between maximum IOP and baseline IOP; FECD: Fuch's Endothelial Corneal Dystrophy; IOP: intraocular pressure; QT: quantitative trait; SD: standard deviation; WGS: whole genome sequencing.

### SNP association analysis for discovery

The Manhattan plots for SNP association analysis of the discovery cohort are show in Figure 1. Quantile-quantile (Q-Q) plots are shown in Figure S1. The list of top SNPs resulting from linear regression analyses are provided in Table S1, ordered by P value. The 12 month and 3 month QT results were merged in Table S2, then sorted by chromosomal position to reveal individual risk loci that clustered multiple top SNPs.

**Figure 1.**
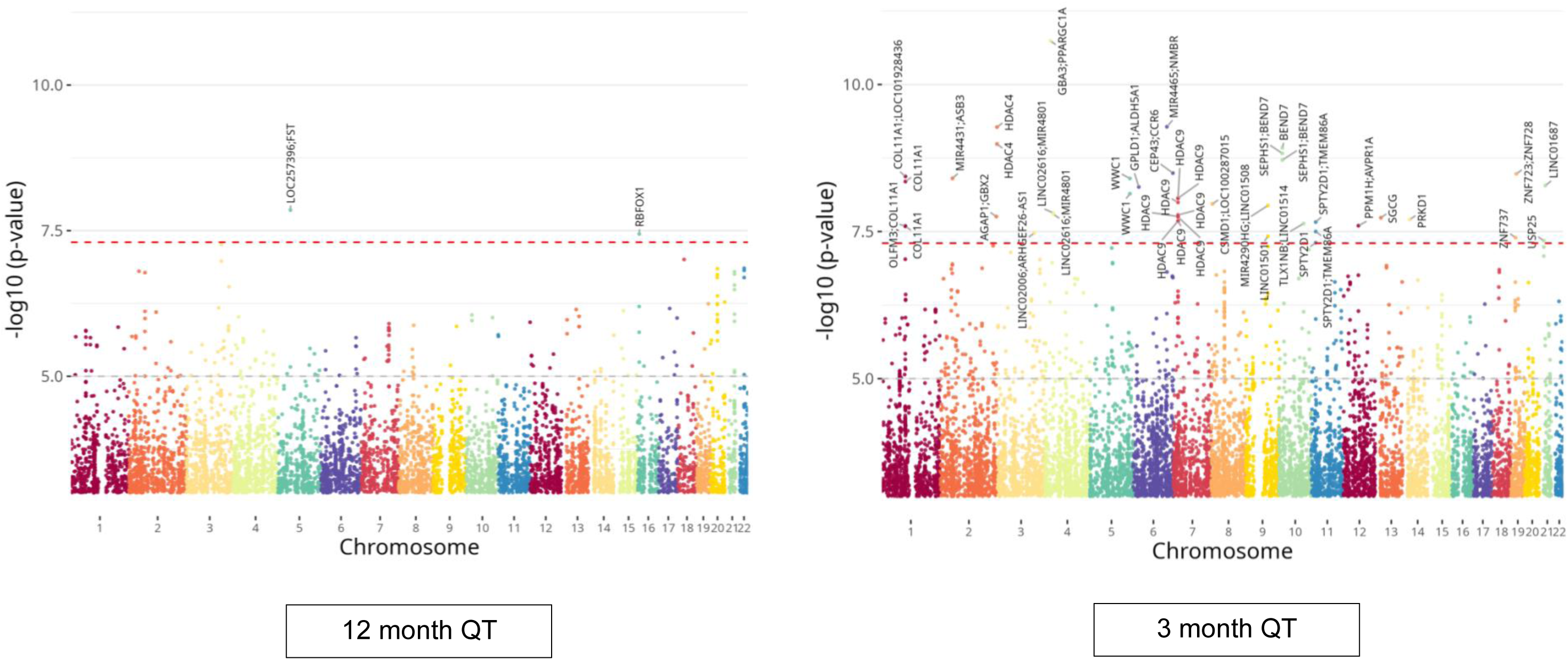
Manhattan plots for the Indianapolis-1 discovery GWASs. The x axis is the position on each chromosome and the y axis is the –log10 *P* value from the GWAS for each SNP. The red dashed line demarcates the threshold for genome-wide significance [−log10(5E-08); p=5.0E-08]. The risk locus for each SNP of genome-wide significance is defined by the nearest genes to the SNP, as delineated on the plots. The quantitative trait (QT) used for each GWAS is indicated.

Table 2 summarizes all results. Using the 12 month QT, we identified 2 SNPs at 2 different risk loci that exceeded the threshold for genome-wide significance and using the 3 month QT, we identified 42 SNPs at 24 different risk loci that exceeded the threshold for genome-wide significance (p<5E-08). Together, a total of 44 SNPs at 26 risk loci attained genome-wide significance and a total of 531 SNPs clustered at 262 risk loci attained suggestive significance (p<5E-06). Five risk loci clustered unique SNPs from the 12 month and 3 month analyses. Eleven SNPs were duplicated between the two analyses, but with different *P* values.

**Table 2.** GWAS summary findings.

| Group/subgroup | *N* SNPs<br>p<5E-08<br>(genome-wide) | *N* SNPs<br>p<5E-06<br>(suggestive) | *N* risk loci clustering<br>SNPs of p<5E-08<br>(genome-wide) | *N* risk loci clustering<br>SNPs p<5E-06<br>(suggestive) |
| --- | --- | --- | --- | --- |
| 12 month QT | 2 | 129 | 2 | 75 |
| 3 month QT | 42 | 413 | 24 | 225 |
| Overlap risk loci, same SNPs | 0 | 11 | 1* | 7 |
| Overlap risk loci, different SNPs | N/A | N/A | 1** | 5 |
| Merged | 44 | 531 | 26 | 262 |

**12 month QT**
| SNP Frequency | MAF ALPHA | Mean effect size<br>in mm Hg (range) | *N* SNPs | % Total |
| --- | --- | --- | --- | --- |
| Common | > 0.05 | 4.21 (-2.57 to 5.70) | 20 | 13.18% |
| Low Frequency | 0.01 to 0.05 | 10.11 (6.09 to 12.89) | 24 | 16.60% |
| Rare | < 0.01 | 19.77 (13.57 to 27.15) | 90 | 68.22% |
| All | All | 15.92 (-2.57 to 27.15) | 134 | 100% |

**3 month QT**
| SNP Frequency | MAF ALPHA | Mean effect size<br>in mm Hg (range) | *N* SNPs | % Total |
| --- | --- | --- | --- | --- |
| Common | > 0.05 | 2.25 (2.00 to 3.78) | 49 | 11.38% |
| Low Frequency | 0.01 to 0.05 | 7.67 (5.13 to 11.80) | 69 | 16.46% |
| Rare | < 0.01 | 17.29 (10.52 to 27.33) | 302 | 72.16% |
| All | All | 13.99 (2.00 to 27.33) | 420 | 100% |

**Notes.** \*The risk locus *PPARGC1A* clusters 1 SNP of suggestive significance by the 12 month QT and genome-wide significance by the 3 month QT. \*\*The risk locus *SGCG* clusters 2 SNPs of suggestive significance by the 12 month QT and a 3<sup>rd</sup> SNP of genome-wide significance by the 3 month QT.
**Acronyms.** QT: quantitative trait; MAF: minor allele frequency; the ALFA project provides aggregate allele frequency from NCBI's dbGaP.

Most SNPs discovered in the current study were rare or of low frequency; only ∼15% were common. Effect sizes were quite large overall, and inversely correlated with minor allele frequency (MAF). All SNPs had a positive effect on the QT, except one SNP with a negative effect: rs2427460. Only 6 SNPs were located within exonic regions of a gene, suggesting that essentially all have regulatory effects on gene expression.

### Co-localization of discovered risk loci with other high-tension ocular phenotypes

Co-localization of SIOH risk loci with those discovered for other high-tension ocular phenotypes was investigated by cross-referencing with published reports^72,73,83–87^. In addition to POAG and PACG, we included pseudoexfoliation syndrome (PEXS) a systemic condition characterized by the deposition of white dandruff-like material within various organs; when this occurs in the TM, it can lead to pseudoexfoliation glaucoma (PEXG). We also included central corneal thickness (CCT) and IOP, both endophenotypes underlying high-tension glaucomas^88^, as well as Type II Stickler syndrome, a congenital disorder that can exhibit a high-tension ocular phenotype (MedLine Plus).

Results are shown in Table S3. Of all risk loci discovered in this study, 48/262 (19%) co-localized with other high-tension ocular phenotypes. Of those 26 discovered risk loci clustering SNPs that exceed the threshold for genome-wide significance, 8/26 (31%) co-localizations were found. Notably, while risk loci matched, none of the associated SNPs were the same.

### Target gene prioritization

While the closest gene to a regulatory SNP is a likely target of that SNP, the closest gene might not be expressed in the relevant tissue. Moreover, regulatory SNPs can affect expression of more than one gene^89^. To prioritize likely target gene(s) of discovered SNPs clustered at each risk locus, we required them to be expressed in the AOPs. To determine AOP expression, we searched the two relevant human scRNA-seq datasets on the Spectacle portal^90^ for the closest expressed gene(s). Results are compiled in Table S4. Notably, of the 6 exonic SNPs discovered, only 2 were located in AOP-expressed genes (*ARVCF* and *LRP2*), suggesting the others, despite their location, are regulatory.

To identify additional target gene candidates, we searched SNPs at each risk locus on the Adult Genotype Tissue Expression (GTEx) Project portal^91^ to determine co-localization with expression or splicing quantitative trait loci (eQTLs/sQTLs). Results are shown in Table S4. Target genes of expression QTLs were searched on Spectacle and if AOP-expressed, this was indicated in Table S4. eQTL/sQTL target genes were, in some cases, the same as genes already prioritized by expression in the AOPs, thus providing cross-validation.

The lists of prioritized target genes identified in these analyses is compiled in Table S4.

### Prioritized target gene validation

Since the primary action of GCs is to regulate transcription, most of our prioritized target genes should be GC-regulated, either directly by the activated GR, or as downstream targets of upstream effectors regulated by the activated GR. To find both, we compared our prioritized target gene list to lists of GC regulated genes identified in cultures of primary TM cell strains in a study co-authored by a member of our team^92^. As shown in Table S5, we found a match to 33% (134/406) of our prioritized target genes.

We also compared our prioritized target gene list to lists of DEGs associated with steroid response in paired eye studies. For this analysis, we used the DEG list compiled in a human paired eye study conducted by members of our team^96^, as well as DEG lists from two studies conducted with bovine donor eyes^97,98^. Our results are shown in Table S5. We found matches to 26 of our prioritized target genes. Of these, 18 were associated with responders and 10 with non-responders (with 2 overlaps), most down-regulated by GC treatment.

To identify prioritized target genes that can be regulated directly by the GR, we turned to the NIH DAVID Bioinformatics functional annotation clustering tool ^93,94^, pairing it with the UCSC_TF Binding Site database that curates CHIP-seq data accessed from the ENCODE project^95^. When run at low classification stringency, the top-scoring cluster included the GR, as shown in Table S5. Of our total prioritized target genes, 58% (221/402) clustered with the GR.

Combining these analyses provided evidence for GC regulation of 68% (276/408) of our prioritized target genes, as tabulated in Table S5.

In our final analysis, we determined all upstream regulators of our prioritized target gene list, comparing to the paired eye study DEGs. To do this, we ran the gene lists through the DAVID plus UCSC_TF binding site analysis as above, but this time at moderate stringency. Results are shown in Table S5. Only a single cluster was identified, comprised of 3 TFs involved in eye development: SP8^99,100^, LHX3^101–103^ and CHX10^104^. A total of 63% (252/402) of these genes were identified as targets of at least one of the three TFs. The paired eye study steroid responder DEG list returned a single cluster comprised of the same three TFs, as well as a fourth eye development TF, CART1^105^. The single cluster for our prioritized target gene list received a very high enrichment score and high statistical significance; the score and significance was much lower for paired eye study DEGs.

### Summary of findings for the top 26 risk loci and additional prioritized target gene validation

Table 3 lists the 26 risk loci clustering SNPs of genome-wide significance, summarizes findings made thus far. As noted above, nearly 1/3^rd^ (31%; 8/26) of these top 26 risk loci co-localize with risk loci for other high-tension ocular phenotypes.

**Table 3.** Lead genomic variants for the 26 risk loci clustering SNPs of genome-wide significance.

| Chr:<br>Position<br>(GRCh38) | Lead variant<br>rsID | P value | Ref<br>allele | Effect<br>allele | Effect<br>Size<br>(mm Hg) | Minor<br>Allele<br>Count | MAF<br>ALPHA | Annotation | Risk locus | Closest AOP-expressed<br>gene | Evidence<br>for GC<br>regulation | Other<br>validation | Overlap with other<br>high-tension ocular<br>phenotypes |
| --- | --- | --- | --- | --- | --- | --- | --- | --- | --- | --- | --- | --- | --- |
| 1:103168079 | rs111928960 | 3.65E-09 | G | A | 8.6 | 22 | 0.016995 | Intronic | COL11A1 | COL11A1 | Paired eye<br>Chromatin | sQTL<br>upstream TFs | POAG, PACG, PEXS,<br>IOP, Stickler |
| 2:53132903 | rs187520610 | 3.93E-09 | G | A | 18.8 | 4 | 0.001116 | Intergenic | MIR4431;<br>ASB3 | ASB3 |  | upstream TFs |  |
| 2:236142879 | rs147559909 | 1.75E-08 | T | C | 17.08 | 5 | 0.008629 | Intergenic | AGAP1;<br>GBS2 | AGAP1 | In silico<br>Chromatin | upstream TFs |  |
| 2:239067423 | rs181217257 | 5.29E-10 | C | T | 21.57 | 4 | 0.000583 | Intronic | HDAC4 | HDAC4 | In silico<br>Chromatin | upstream TFs |  |
| 3:153939413 | rs16823323 | 3.35E-08 | G | A | 9.48 | 14 | 0.018517 | Intergenic | LINC02006;<br>ARHGEF26 | ARHGEF26 | TM cells<br>In silico<br>Paired eye | upstream TFs |  |
| 4:23507444 | rs113063005 | 1.79E-11 | T | C | 21.21 | 6 | 0.003864 | Intergenic | GBA3; PPARGC1A | PPARGC1A | TM cells<br>In silico | upstream TFs |  |
| 4:37052137 | rs77141817 | 1.52E-08 | T | C | 20.61 | 3 | 0.001376 | Intergenic | LINC02616;<br>MIR4801 | *C4orf19 | In silico |  |  |
| 5:53366842 | rs142934021 | 1.40E-08 | G | A | 18.82 | 7 | 0.00257 | Intergenic | LOC257396; FST | FST | TM cells<br>In silico<br>Paired eye | upstream TFs | CCT |
| 5:168394261 | rs545428520 | 3.96E-09 | T | C | 21.57 | 4 | 0.001271 | Intronic | WWC1 | WWC1 | TM cells<br>In silico | upstream TFs | POAG |
| 6:24491120 | rs150586237 | 5.51E-09 | C | T | 22.05 | 3 | 0.002382 | Intergenic | GPLD1; ALDH5A1 | GPLD1 | In silico<br>Chromatin |  |  |
| 6:141948748 | rs72986533 | 5.23E-10 | C | A | 22.83 | 8 | 0.00459 | Intergenic | MIR4465; NMBR | *VTA1 | TM cells<br>In silico | upstream TFs |  |
| 6:167501386 | rs148153037 | 3.20E-09 | G | A | 14.34 | 8 | 0.01395 | Intergenic | CEP43;<br>CCR6 | CCR6 | Chromatin |  |  |
| 6:18391487<br>6:18392161 | rs76526501<br>rs74455595 | 8.57E-09 | G<br>A | A<br>G | 14.84 | 6 | 0.019854<br>0.014293 | Intronic | HDAC9 | HDAC9 | In silico<br>Chromatin | upstream TFs |  |
| 8:5719972 | rs575473987 | 1.07E-08 | C | T | 22.22 | 3 | 0.001117 | Intergenic | CSMD1;<br>LOC100287015 | CSMD1 | In silico | upstream TFs | POAG |
| 9:90268417 | rs545690161 | 1.14E-08 | G | A | 17.14 | 6 | 0.005241 | Intergenic | MIR4290HG;<br>LINC01508 | DIRAS2 |  |  |  |
| 10:13455976 | rs117998251 | 1.19E-09 | C | T | 22.8 | 3 | 0.006514 | Intronic | BEND7 | BEND7 | TM cells<br>In silico<br>Paired eye | upstream TFs | POAG |
| 10:101158729 | rs117913371 | 2.31E-08 | G | A | 7.87 | 21 | 0.018916 | Intergenic | TLX1NB;<br>LINC01514 | *KAZALD1 | TM cells<br>In silico | upstream TFs |  |
| 11:18680239 | rs541653703 | 2.18E-08 | G | A | 18.32 | 4 | 0.004341 | Intergenic | SPTY2D1;<br>TMEM86A | SPTY2D1 | TM cells<br>Chromatin |  |  |
| 12:63051500 | rs191053292 | 2.51E-08 | T | C | 22.96 | 3 | 0.002275 | Intergenic | PPM1H; AVPR1A | PPM1H | TM cells<br>In silico<br>Chromatin | upstream TFs | POAG |
| 13:23312875 | rs139598422 | 1.84E-08 | A | G | 20.49 | 4 | 0.001704 | Intronic | SGCG | SGCG | TM cells<br>In silico | upstream TFs | CCT |
| 14:29692681 | rs74704551 | 1.98E-08 | C | T | 24.2 | 3 | 0.000624 | Intronic | *PRKD1* | *PRKD1* | TM cells<br>In silico | upstream TFs |  |
| 16:6808238 | rs138164904 | 3.60E-08 | C | T | 23.89 | 3 | 0.000136 | Intronic | *RBFOX1* | *RBFOX1* | TM cells<br>In silico<br>Paired eye | upstream TFs | POAG, PEXS |
| 19:20546292 | rs560206697 | 4.02E-08 | C | T | 25 | 3 | 0.001377 | Intronic | *ZNF737* | *ZNF737* |  |  |  |
| 19:22940396 | rs111285015 | 3.31E-09 | G | A | 27.33 | 3 | 0.009052 | Intergenic | *ZNF723; ZNF728* | *ZNF728* |  |  |  |
| 21:15872687 | rs79486609 | 4.53E-08 | G | A | 20.66 | 3 | 0.01061 | Intronic | *USP25* | *USP25* | In silico | upstream TFs |  |
| 21:22078395 | rs192134381 | 5.23E-09 | T | C | 21.79 | 3 | 0.03987 | ncRNA<br>intronic | *LINC01687* | *\*NCAM2* | In silico | upstream TFs |  |

**Notes.** The closest AOP-expressed gene was determined by identifying genes located up or downstream from the hit SNP using the UCSC Genome Browser, then determining expression of each via Spectacle. Except for the four genes marked with an asterisk, the identified gene was also one of the genes closest to the lead SNP, upstream or downstream. Two overlapping SNPs with equal p values were associated with *HDAC9*.
**Definitions.** Paired eye: responder or non-responder in one of the 3 paired eye studies discussed in the text. sQTL: colocalizes with an sQTL identified on the GTEx portal. GC-regulated TM cells: differentially expressed genes (DEGs) in the TM cell culture studies discussed in the text. GC-regulated In silico: identified in the DAVID Bioinformatics/ UCSC\_TFBS low stringency analysis. GC-regulated chromatin: identified in chromatin analysis using Regulome DB; Stickler: Type II Stickler syndrome; upstream TFs: clusters with eye development TFs identified in the DAVID Bioinformatics/ UCSC\_TFBS moderate stringency analysis.
**Acronyms.** AOP: aqueous outflow pathway; MAF ALPHA: minor allele frequency (the ALFA project provides aggregate allele frequency in NCBI dbGaP); POAG MA: primary open angle glaucoma, multi-ancestry; POAG AF: primary open angle glaucoma, African ancestry; PACG: primary angle closure glaucoma; PEXS/G: Pseudoexfoliation syndrome/glaucoma; IOP: intraocular pressure; CCT: central corneal thickness.

Analysis of GC regulation using gene expression profiling datasets discussed above, supports validity of 73% (19/26) of the top prioritized target genes. To identify any additional genes that might be GC-regulated, we searched on HaploReg v4.2 for all SNPs clustered at the 26 risk loci ^106^. Results are shown in Table S6. Compiling the genes newly-identified in this analysis with those already listed in Table 3 raised the total to 85% (22/26). Combining GC-regulation analyses with the other gene expression analyses discussed above provided support for 88% (23/26) of the top prioritized target genes.

### Independent replication

We tested for replication of individual SNPs that exceeded a threshold of *P*=5.0E-06 in the discovery cohort. Because most of our SNPs were rare, we also evaluated replication of the top 26 prioritized genes using a gene-based burden test that we have previously described^107^, which aggregates P values for all SNPs located within the specific gene tested. Complete results are shown in Table S7 and summarized in Table 4.

**Table 4.**
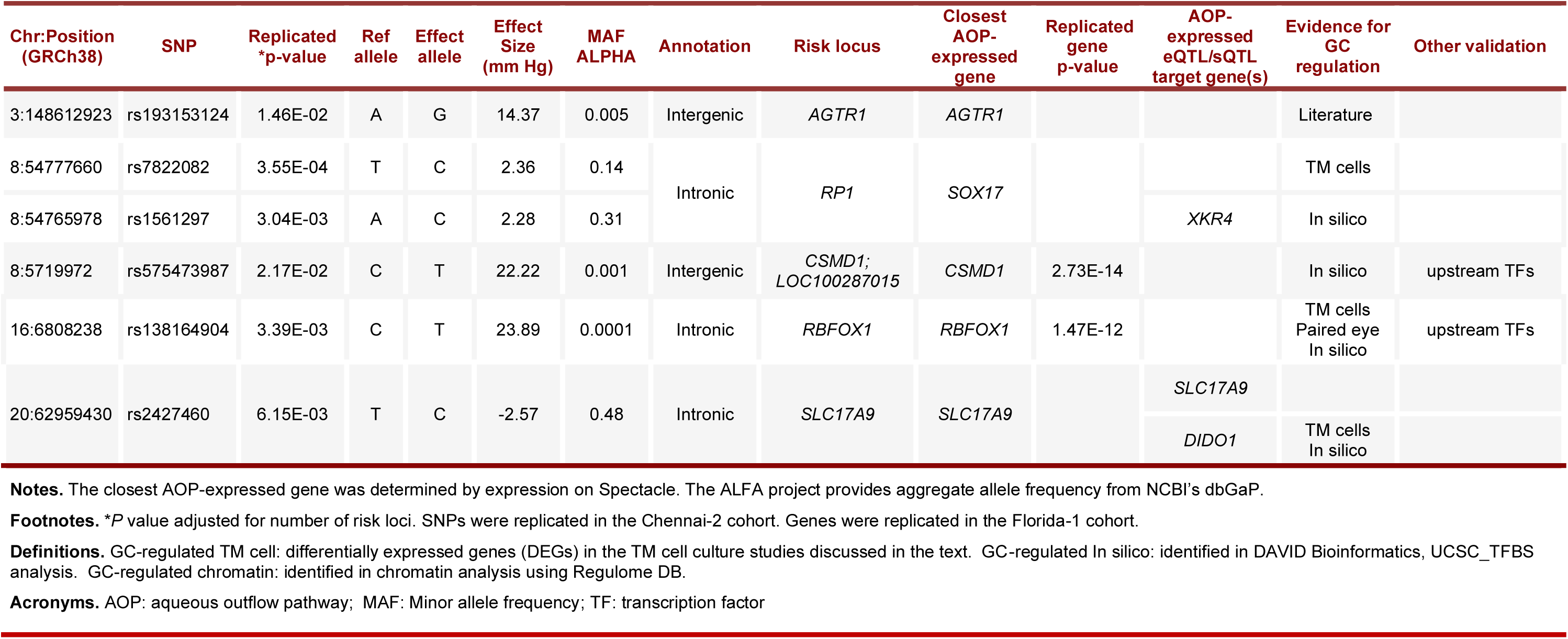
SNPs and prioritized target genes replicated.

| Chr:Position<br>(GRCh38) | SNP | Replicated<br>*p-value | Ref<br>allele | Effect<br>allele | Effect<br>Size<br>(mm Hg) | MAF<br>ALPHA | Annotation | Risk locus | Closest<br>AOP-expressed<br>gene | Replicated<br>gene<br>p-value | AOP-expressed<br>eQTL/sQTL<br>target gene(s) | Evidence for<br>GC<br>regulation | Other validation |
| --- | --- | --- | --- | --- | --- | --- | --- | --- | --- | --- | --- | --- | --- |
| 3:148612923 | rs193153124 | 1.46E-02 | A | G | 14.37 | 0.005 | Intergenic | *AGTR1* | *AGTR1* |  |  | Literature |  |
| 8:54777660 | rs7822082 | 3.55E-04 | T | C | 2.36 | 0.14 | Intronic | *RP1* | *SOX17* |  |  | TM cells |  |
| 8:54765978 | rs1561297 | 3.04E-03 | A | C | 2.28 | 0.31 |  |  |  |  | *XKR4* | In silico |  |
| 8:5719972 | rs575473987 | 2.17E-02 | C | T | 22.22 | 0.001 | Intergenic | *CSMD1*;<br>*LOC100287015* | *CSMD1* | 2.73E-14 |  | In silico | upstream TFs |
| 16:6808238 | rs138164904 | 3.39E-03 | C | T | 23.89 | 0.0001 | Intronic | *RBFOX1* | *RBFOX1* | 1.47E-12 |  | TM cells<br>Paired eye<br>In silico | upstream TFs |
| 20:62959430 | rs2427460 | 6.15E-03 | T | C | -2.57 | 0.48 | Intronic | *SLC17A9* | *SLC17A9* |  | *SLC17A9* |  |  |
|  |  |  |  |  |  |  |  |  |  |  | *DIDO1* | TM cells<br>In silico |  |

**Notes.** The closest AOP-expressed gene was determined by expression on Spectacle. The ALFA project provides aggregate allele frequency from NCBI's dbGaP.
**Footnotes.** \*P value adjusted for number of risk loci. SNPs were replicated in the Chennai-2 cohort. Genes were replicated in the Florida-1 cohort.
**Definitions.** GC-regulated TM cell: differentially expressed genes (DEGs) in the TM cell culture studies discussed in the text. GC-regulated In silico: identified in DAVID Bioinformatics, UCSC\_TFBS analysis. GC-regulated chromatin: identified in chromatin analysis using Regulome DB.
**Acronyms.** AOP: aqueous outflow pathway; MAF: Minor allele frequency; TF: transcription factor

Of 81 risk loci identified in the Indianapolis-2 replication cohort GWAS shown in Table S8, 15% (12/81) colocalized with risk loci identified in the Indianapolis-1 discovery cohort. However the SNPs associated with these loci were new; they did not replicate those found in the discovery GWAS performed with the Indianapolis-1 cohort. Two significant SNP replications were identified in the 12 month QT dataset, one each at risk loci *LUZP1* and *ANKRD18CP,* and 4 significant replications in the 3 month QT dataset, one each at risk loci *MYO1E, TMEM201, HAS2;SMLR*, and *TFPI*. All dropped below the threshold for replicative significance when adjusted for multiple comparisons.

The Florida-1 replication cohort yielded 1 nominally significant replication in the 12 month QT dataset at risk locus *PCP4*, and 7 nominally significant replications in the 3 month QT at risk loci *RP1*, *MACROD2*, *URB2* and *TBK1*. All dropped below the threshold for replicative significance when adjusted for multiple comparisons. However, when the gene-based burden test was applied for the top prioritized target genes identified in the Indianapolis-1 cohort, P values for 2/26 of these genes exceeded the statistical threshold for significance: *RBFOX1* and *CSMD1*. We found evidence for GC-regulation of both genes (Table 3). The risk loci encompassing both of these genes have also been linked to other high tension ocular phenotypes (Table S2).

The Chennai-1 replication cohort did not yield even nominally significant replications. However 4 SNPs were replicated in the Chennai-2 cohort with *P* values that exceeded the adjusted threshold for significance. Replicated SNP rs2427460 is located within an intron of *SLC17A9*; it is the only SNP discovered in this study with a protective effect (lowers IOP). Replicated SNPs rs7822082 and rs1561297 are located in an intron of *RP1*. All three are common in frequency. Replicated SNP rs193153124 is located intergenically, upstream of *AGTR1*. This SNP is rare. Although we did not identify *AGTR1* as a GC-regulated gene in validation analyses discussed above, a report in the literature documents GC stimulation of *AGTR1* expression^151^.

### Aqueous outflow pathway expression of top prioritized target genes from discovery and replication

Spectacle heat maps for relative AOP cell type expression of the top 26 prioritized target genes and the 3 prioritized target genes of replicated SNPs are shown in Figure S4. Findings are summarized in Table 5. Notably, 5 of these genes are not expressed in the cribriform JCT and 4 more genes are not expressed in either TM beam cells or cribriform JCT. All are expressed most strongly in other AOP cell types.

**Table 5.**
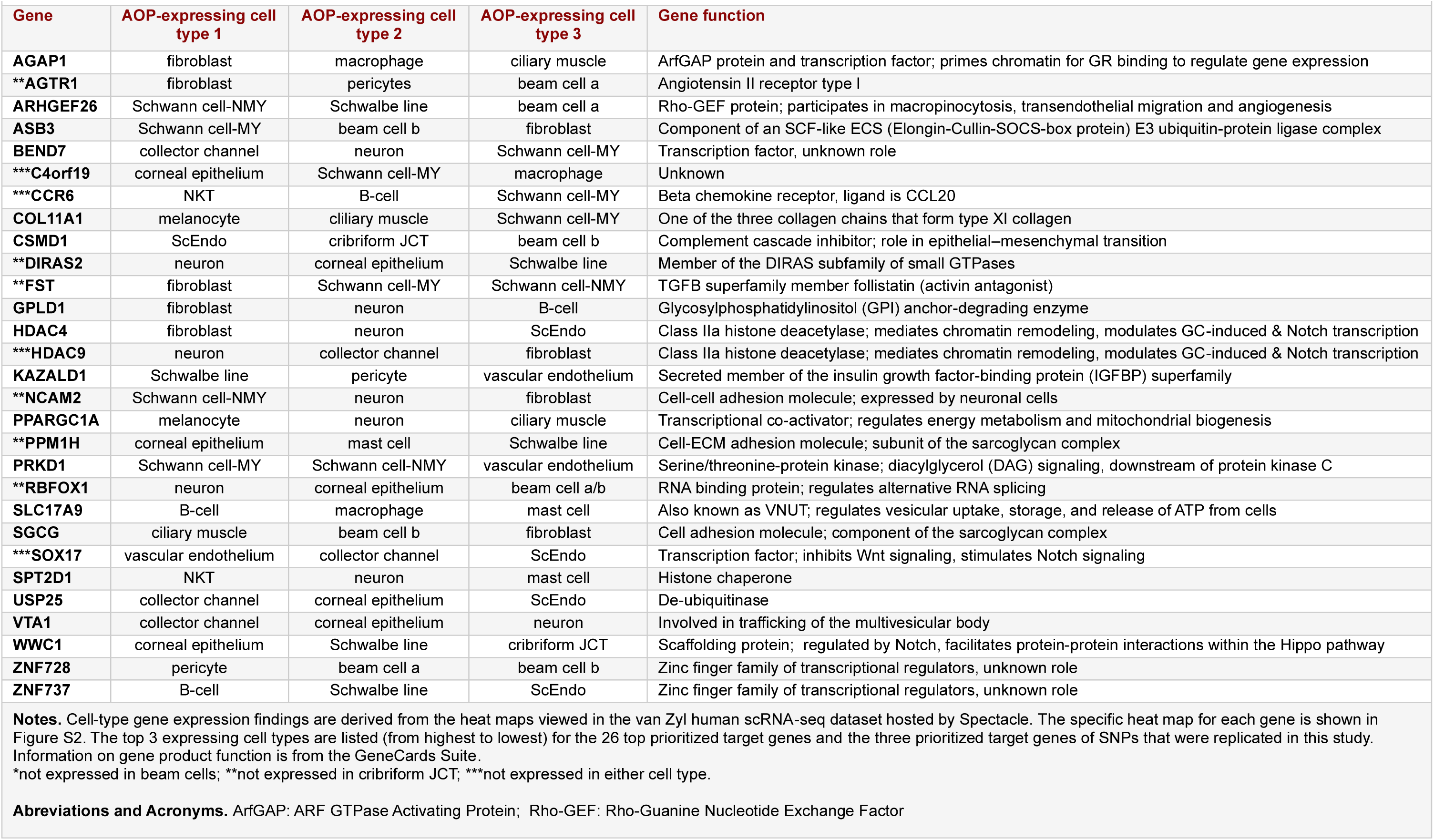
Aqueous outflow pathway (AOP) expression of top prioritized target genes and those of replicated SNPs.

### Functional annotation of SNPs clustered at the COL11A1 risk locus

We found the broad association of high-tension ocular phenotypes with different regulatory SNPs at risk locus *COL11A1* to be quite intriguing. To learn more about how the different SNPs might act to affect *COL11A1* gene expression, we searched multiple bioinformatics platforms, focusing in particular on data from the GTEx portal^91,108^. Findings are compiled in Table 4. Violin plots depicting SNP effects on *COL11A1* RNA splicing are shown in Figure S5.

**Table 6.** SNPs within and around the *COL11A1* gene associated with different high tension ocular phenotypes.

| High tension ocular phenotype | Genomic position (GRCh38) | SNP ID | MAC | Effect size (mm Hg) | MAF ALFA | P value | Location in *COL11A1* | Regulatory role | Regulatory process affected | Reference |
| --- | --- | --- | --- | --- | --- | --- | --- | --- | --- | --- |
| SIOH | 102337928 | rs140420703 | 5 | 16.49 | 0.002859 | 1.91E-06 | downstream | GR binding motif | Transcription by GCs |  |
| SIOH | 102400809 | rs563167766 | 3 | 19.42 | 0.001323 | 4.73E-06 | downstream |  |  |  |
| SIOH | 102496326 | rs77180278 | 24 | 7.34 | 0.009519 | 9.34E-08 | downstream | sQTL, negative effect | Intron 41 splicing |  |
| SIOH | 102754804 | rs112351653 | 24 | 7.42 | 0.017196 | 2.55E-08 | downstream | sQTL, negative effect | Intron 41 splicing |  |
| SIOH | 102760770 | rs180926150 | 3 | 19.42 | 0.000583 | 3.77E-06 | downstream |  |  |  |
| **Genomic position:102,876,473, exon 67 end** |  |  |  |  |  |  |  |  |  |  |
| PACG | 102888582 | rs1676486 |  |  | 0.187992 | 1.10E-05 | Exon 60 of 67 | Missense mutation | Protein translation | Vithana et al. (84) |
| PACG | 102914362 | rs3753841 |  |  | 0.397409 | 9.22E-10 | Exon 52 of 67 | Missense mutation | Protein translation | Vithana et al. (84) |
| PACG POAG | 102919817 | rs993471 |  |  | 0.457463 | 3.03E-11 | Intron 49 of 66 | Multiple TF binding sites (CHIP-seq) | Transcription | Han et al. (72)<br>Lo Faro et al. (83) |
| **Genomic position:102,946,956, intron 41 end** |  |  |  |  |  |  |  |  |  |  |
| SIOH | 102953612 | rs114413507 | 24 | 7.42 | 0.017561 | 2.57E-08 | Intron 41 of 66 | sQTL, negative effect | Intron 41 splicing |  |
| PEXG | 102957174 | rs3102053 |  |  | 0.283179 | 3.00E-04 | Intron 41 of 66 | sQTL, both negative and positive effects | Intron 41 splicing (neg)<br>intron 31 splicing (pos) | Krumbiegel et al. (87) |
| Stickler | 102961971 | None known |  |  | rare | Mendelian | Intron 41 of 66 | Splice site mutation | Intron 41 splicing | Kohmoto et al. (110) |
| **Genomic position:102,961,866, intron 41 start** |  |  |  |  |  |  |  |  |  |  |
| SIOH | 103007360 | rs116672066 | 23 | 8.05 | 0.01704 | 4.46E-09 | Intron 15 of 66 | sQTL, negative effect | Intron 41 splicing |  |
| **Genomic position:103,108,521, exon 1 start** |  |  |  |  |  |  |  |  |  |  |
| SIOH | 103168079 | rs111928960 | 22 | 8.60 | 0.016995 | 3.65E-09 | upstream | sQTL, negative effect | Intron 41 splicing |  |
| SIOH | 103288418 | rs113221952 | 19 | 7.92 | 0.016305 | 9.17E-07 | upstream | sQTL, negative effect | Intron 41 splicing |  |

**Notes.** The table compiles information obtained by examining genotype-phenotype association using the GTEx portal and the NIH Genome-Wide Repository of Associations Between SNPs and Phenotypes (GRASP) portal, performing *in silico* analyses using tools on the RegulomeDB and HaploReg v4.2 servers, extracting genomic position localization and sequence information from the Santa Cruz Genome Browser and the EMBL Nucleotide Sequence Database, and accessing disease phenotype information sourced on MedLine Plus. All effect sizes listed are positive. The *COL11A1* gene is located on chromosome 1 and has 66 introns and 67 exons.
**Headers, Abbreviations, Acronyms.** GC=glucocorticoid; GR=glucocorticoid receptor; MAC=minor allele count; MAF ALFA=minor allele frequency (allele frequency aggregator dataset from NCBI dbGAP); neg: negative; PACG=primary angle closure glaucoma; PEXG=pseudoexfoliation glaucoma; POAG=primary open angle glaucoma; pos: positive; SIOH=steroid-induced ocular hypertension; sQTL=splicing quantitative trait; Stickler=Type II Stickler Syndrome; TF=transcription factor

As already noted, the top 6 of 9 SNPs from our discovery GWAS clustered at the *COL11A1* risk locus co-localized with sQTLs identified in testis. While these SNPs are located within *COL11A1* intron 15, *COL11A1* intron 41 and intergenic DNA, they all interfere with splicing of intron 41. This could result in a longer, non-functional protein, or in nonsense-mediated decay of the elongating protein during translation. A 7^th^ hit SNP colocalizes with a GR-binding motif. Alternative splicing is reported to be coupled to transcription^109^, suggesting possible synergism between activate GR-binding at the regulatory motif and the 6 SNPs that affect splicing.

The Type II Stickler syndrome mutation localizes to a splice acceptor site of intron 41 that also interferes with its splicing^110^. A SNP associated with PEXS/G similarly interferes with intron 41 splicing. Interestingly, results indicate that the PEXS/G SNP can also positively affect alternative splicing within intron 31.

A SNP associated with both PACG and POAG was demonstrated by CHIP-seq to affect binding of multiple TFs. In addition, two SNPs associated with PACG are missense mutations that would interfere with mRNA translation and final protein production.

### Functional annotation of all prioritized target genes

While the p=5E-08 threshold for statistical significance is necessary to provide the stringency to limit false positives, a lesser threshold is acceptable for follow-ups, such as pathway enrichment analysis, for which the cost of a modestly greater set of false positives is low^111^. Alphabetical ordering of the full prioritized gene list in Table S3 highlighted functionally similar paralogues or gene family members by grouping them together. We counted 41 such paralogous groups. Based on information from GeneCards Suite^112^, we identified a set of 15 functional groups for the top prioritized genes; these are listed in Table S9.

In our final analysis, we used the full prioritized gene list to conduct pathway enrichment, pairing the NIH DAVID Bioinformatics functional annotation tool with the Reactome database^113^. The list of human paired eye study DEGs^96^, separated into steroid responders and non-responders, was put through the same analysis for comparison.

Results are summarized in Table S9. The top 5 Reactome events scored for the GWAS prioritized target gene list are: 1) “signaling by NOTCH”, 2) “diseases of signal transduction by growth factor receptors and second messengers”, 3) “cardiac conduction”, 4) “carbohydrate metabolism” and 5) “adipogenesis”. Pathways 2, 3, and 4 were also scored in analysis of the paired eye study DEG lists. Overlap between our prioritized target gene list and the paired eye study DEGs list support the validity of the genes.

As diagrammed in Figure 3, genes that clustered with “signaling by NOTCH” encompass the full range of Notch protein expression, maturation and signaling. Genes that clustered with “diseases of signal transduction by growth factor receptors and second messengers” included most of the same genes.

**Figure 2.**
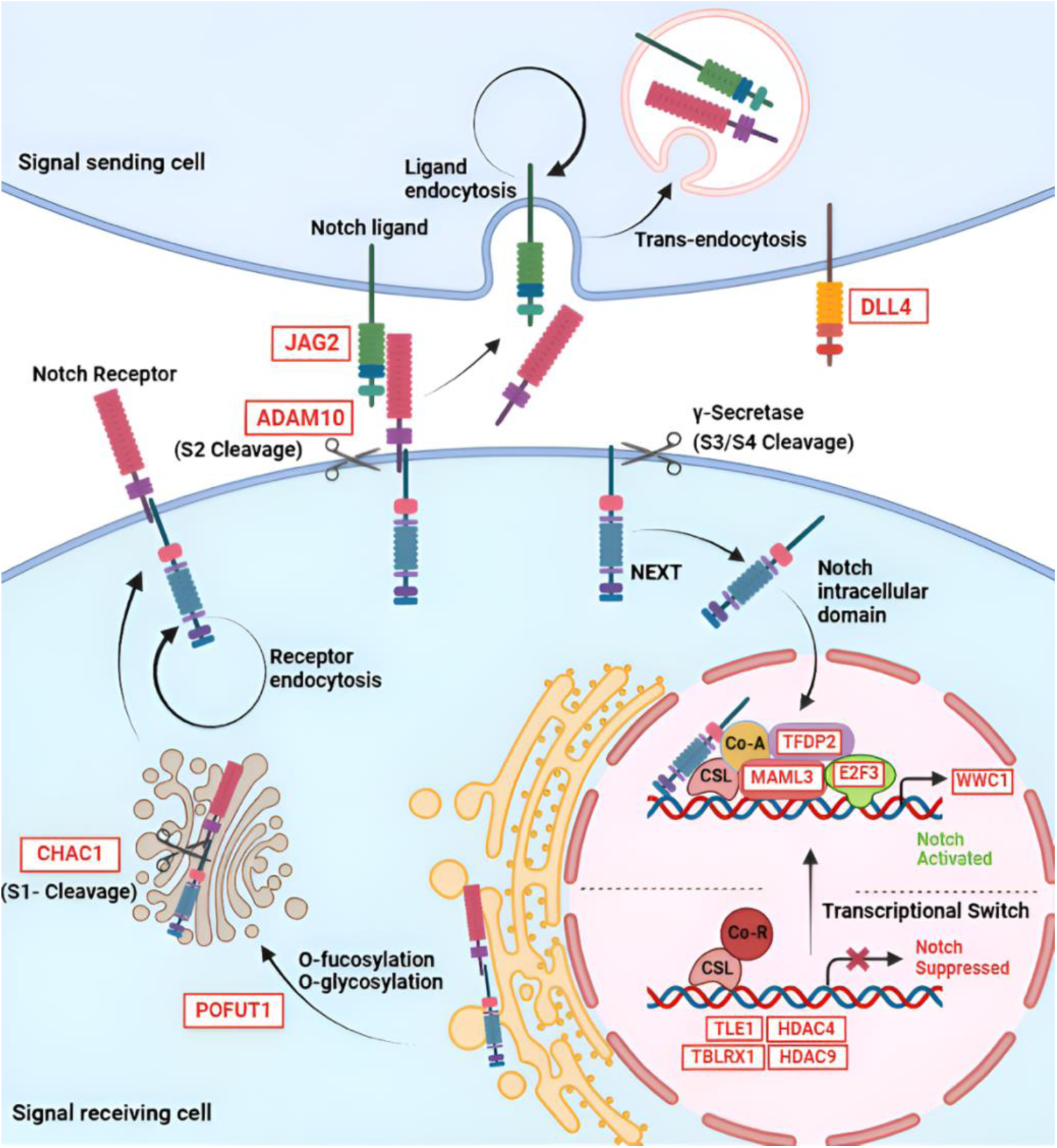
GWAS prioritized target genes that cluster with the pathway “signaling by NOTCH”. A schematic overview of the canonical Notch signaling pathway is shown, with white boxes indicating prioritized target genes of SNPs identified in this study. Notch receptors are synthesized as a single polypeptide that is O-fucosylated and glycosylated in the endoplasmic reticulum. *POFUT1* encodes a Golgi-localized fucosyltransferase that performs the first step in glycosylation of the Notch EGF-like domains. Loss of POFUT1 severely inhibits Notch signaling^156,157^. The Notch protein then moves to the trans-Golgi network, where it is cleaved by a Furin-like convertase(s) (S1), forming a heterodimer. *CHAC1* encodes an inhibitor of this cleavage^158^. The heterodimeric receptor is then transported to the cell membrane, where it binds with the ligand expressed in the neighboring cell. *DLL4* and *JAG2* encode members of the Notch ligand families Delta and Serrate, respectively. Receptor– ligand interaction initiates second cleavage (S2) within the extracellular domain. *ADAM10* encodes one of only two proteinases known to effect this cleavage step (the other is ADAM17) ^158^. After S2 cleavage, the Notch extracellular truncated fragment (NEXT), is ‘trans-endocytosed’ by the neighboring ligand-expressing cell, in a process that appears to be controlled by E3 ubiquitin ligases. This is followed by a 3^rd^ and 4^th^ cleavage (S3, S4) that occurs within the transmembrane domain and is mediated by a multi-protein γ-secretase. The intracellular domain of Notch (NICD) is then released and translocates into the nucleus and binds to the transcription factor CSL. This interaction leads to transcriptional activation by displacement of co-repressors (Co-R), including proteins encoded by *TLE1, HDAC4, HDAC9, and TBLRX1*. Simultaneous recruitment of co-activators (Co-A) occurs, such as members of the mastermind family, including the protein encoded by *MAML3*^159^. Transcription factors encoded by *TFDP2* and *E2F3* bind to the promoter regions of Notch target genes, thereby enhancing their transcription^160^. The protein encoded by *WWC1* (also known as Kibra) is a notch target gene and a key component of Hippo/YAP signaling^161^.

All prioritized target genes that clustered with “cardiac conduction” were part of phase I of the five conduction phases that occur in the sinoatrial node or “pacemaker” of the heart. The identified genes encode specific potassium channels and their modifiers: *KCND3, KCNIP1, KCNIP3, KCNIP4.* The paired eye study analysis provided an intriguing complement with “cardiac conduction” phase II receiving a strong score.

Genes that clustered with “carbohydrate metabolism”, were all part of the sub-pathway “glycosaminoglycan metabolism”. We note that metabolism was a major theme overall, with scored pathways also including “diseases of glycosylation” and “O-linked glycosylation”. Vesicle-mediated transport pathways involved in protein biosynthesis and recycling through the golgi and endoplasmic reticulum were also metabolism-related themes.

Genes that clustered with “adipogenesis”, were all part of the sub-pathway “transcriptional regulation of brown and beige adipocyte differentiation by EBF2”. These include the top prioritized gene *PPARGC1A*, encoding a coactivator of nuclear receptors (including the GR). The lower-ranked pathway “mitochondrial biogenesis” also includes *PPARGC1A.* Both are energy metabolism pathways.

## DISCUSSION

This pharmacogenomics study follows up on two pilot studies published by members of our team that identified significant genomic variants associated with steroid response in the eye for the first time^81,82^. These were the only published GWAS studies on SIOH up until now. For this new study, we built a substantially larger, but still uniform, discovery cohort by partnering with a single surgical practice. We then pooled our existing cohorts for replication. Our findings substantially expand on previous results, with 44 new SNPs of genome-level significance clustered at 26 risk loci in the discovery cohort, and a total of 531 SNPs of suggestive significance clustered at 262 risk loci. Similar to SNPs associated with complex disease^74–76^, essentially all SNPs discovered here were located in non-coding DNA, where they are predicted to exert regulatory effects on expression of nearby genes. However, in striking contrast to the predominantly common variants associated with complex disease^114^, including the various glaucoma subtypes^77^, most of the SNPs that we discovered here were rare or of low frequency.

We identified many more SNPs of genome-wide significance in our discovery cohort than would typically be expected for an *N* of only 439/422 unique genotypes^115–117^. However, drug response can exhibit quite large effect sizes and this feature increases the potential to reveal rare variants^79^. In disease, larger effect sizes are usually observed only when variants fall within protein-coding regions that alter the encoded protein’s structure/function^118^. These variants are typically under selective pressure of elimination, however drug response variants may not be under such pressure^78^. An additional consideration is that GCs act directly on the DNA through the GR; a single nucleotide change in a DNA binding site for the GC-activated GR could thus have a phenotypic effect similar to a GR protein mutation. Also important was our use of a QT for analysis, providing considerably more statistical power than a case-control study design. Maybe of most importance, the uniform data collection method we employed made it possible to use an early 3 month time point for calculating the QT. This proved much more effective than the 12 month time point, which may mask variants contributing to early steroid response. Our ability to isolate a relatively “pure” phenotype in this way is likely the key factor explaining our discovery of so many significant variants.

An important finding of this study is the substantial overlap among SIOH risk loci with those identified for high-tension ocular phenotypes, including almost 1/3^rd^ of the top 26 risk loci. The pathophysiology of SIOH is very similar to high-tension POAG ^9,10^, thus gene sharing is not unexpected. However, in no case did we find that individual SIOH SNPs were shared with high-tension POAG. Since POAG is a risk factor for SIOH, we chose to eliminate glaucoma patients or glaucoma suspects from our study, likely explaining this result. Our findings thus confirm the prediction that genes involved in the pathophysiology of SIOH and high-tension POAG overlap, but further show that the pathogenetic triggers differ. This is evidence that SIOH and POAG are distinct diseases.

More surprising was the overlap with other high-tension ocular phenotypes that are mechanistically different from SIOH or POAG. The best example was the risk locus that clustered SNPs located within and around *COL11A1*, as it was shared among six of the high-tension ocular phenotypes we examined here. *COL11A1* is one of our top prioritized target genes. It encodes one of the three chains of collagen type XI. Significantly, most of the SIOH SNPs clustered at this risk locus co-localize with sQTLs that interfere with splicing of intron 41, and thus production of a functional mRNA and protein product. SNPs associated with high tension ocular phenotypes were different from those we identified for SIOH, but they converged functionally with SIOH in this way.

*COL11A1* is expressed in the sclera and it has been proposed that alterations in expression might result in smaller hyperopic eyes predisposed to angle closure in PACG^125^. Alternatively, as *COL11A1* expression is also observed in cultured TM cells, a direct effect on aqueous outflow was suggested^125^. The analysis performed here on Spectacle confirmed that *Col11A1* is expressed in TM and JCT cells *in vivo*, however higher expression occurs in smooth muscle cells of the ciliary body. COL11A1 is known to regulate tendon fibril assembly and organization throughout the body. Significantly, the ciliary muscle and TM are biomechanically connected via tendon fibers. Contraction of the CM moves the scleral spur posteriorly and pulls the TM lamellae inward, leading to enlargement of the potential space within the TM and expansion of Schlemm’s canal^127–130^. This may be the most important mechanism for regulating IOP in primates; limited posterior movement capability has been linked to high-tension POAG^131^. Importantly, knockout of *COL11A1* in mice disrupted structure and function of the flexor digitorum longus tendon in the leg^126^. Based on these observations, we propose that *COL11A1* is essential for biomechanical coupling between the TM and CM to regulate aqueous outflow. Impaired tendon fiber remodeling in this region, due to insufficient COL11A1, may lead to uncoupling of the TM and CM, thereby reducing the capacity to dynamically regulate IOP. This is not an hypothesis that has been previously proposed to our knowledge, but can be feasibly tested using the existing knockout mouse^133^.

A key feature of SIOH and POAG pathogenesis is fibrotic changes in the TM and JCT^9,10^. Supporting this idea, upregulated expression of various collagen chains associated with fibrosis is typically reported in GC-regulated transcriptional profiling studies^119–123,124^. Yet fibrotic changes are not specific to steroid responders, thus fibrosis may not be enough to advance the pathology. In fact, our evaluation of SNPs associated with elevated IOP in the *COL11A1* gene indicate down-regulated expression of COL11A1 and *COL11A1* was specifically down-regulated in steroid responders in a paired eye study evaluated here. Taking these points together leads us to propose that the GC-induced fibrotic response in the TM and JCT must couple with a secondary mechanism to advance pathology.

This “advancing” mechanism appears to involve genes expressed in AOP tissues more broadly, as exemplified by *COL11A1*. Evidence for this was provided by our analysis of AOP cell-type expression of prioritized target genes on Spectacle; we found that many are not expressed in cells of the TM or the JCT. Moreover, for those genes that were found to be expressed in TM or JCT, expression was often stronger in other AOP cell types. Interestingly, a similar observation was recently made for genes involved in the pathophysiology of high-tension POAG using the same Spectacle datasets that we used here^134–136^.

Nevertheless, we also found evidence that pathology can involve exacerbation of the normal fibrotic response within the TM and JCT. Two of our other top prioritized target genes, FST and ARHGEF26, are both involved in the fibrotic response and expressed in the TM and JCT. FST is an inhibitor of fibrosis that is increased in the TM of high-tension glaucoma patient, perhaps as a compensatory response^148^. ARHGEF26 is a rho-guanine nucleotide exchange factor; knockdown ameliorated GC-induced myofibroblast transdifferentiation in TM cells and prevented the development of GC-induced OH in mice ^149^. The product of *AGTR1*, the prioritized target gene of a SNP replicated in this study, is implicated in the development of fibrosis in various tissues, including the heart, lungs, and kidneys^150^.

The remarkable number of SNPs discovered in this study predicting risk for SIOH, enabled us to identify a large number of potential target genes, misexpression of which could contribute to SIOH pathophysiology. Published results of gene expression profiling in cultured TM cells proved valuable for validation of our prioritized target gene list. We also employed computational analyses using CHIP-seq data accessed from the ENCODE project^95^. Upstream regulator analysis indicated the clustering of our prioritized target gene list with three TFs involved in eye development with a high enrichment score; the gene profiling list used for comparison also clustered with these TFs, but the score was much lower. This might be explained by gene expression changes due to culturing of the TM cells. The large number of prioritized target genes identified here in the discovery cohort provided sufficient input for pathway enrichment analysis.

The list of prioritized target genes identified in this study provided sufficient numbers to conduct pathway enrichment analysis. We uncovered a wealth of new hypothesis-generating information. Again, published results of gene expression profiling in cultured TM cells proved valuable for validation. The top pathway we identified was “signaling by NOTCH”. The Notch signaling pathway is known to modulate GR-dependent gene expression ^137^, but its role in high-tension ocular pathologies has only recently attracted attention. ECM production in GC-treated TM cells is inhibited via the Notch pathway^138^ and expression of Notch signaling pathway genes is increased in TM cells isolated from high-tension POAG patients^139^.

The next pathway identified was “cardiac conduction”, which clusters genes encoding a related group of potassium ion channels that serve as cardiac pacemakers: *KCND3, KCNIP1, KCNIP3, KCNIP4*. Their expression has not been previously reported in the AOPs to our knowledge. The resting membrane potential in human TM cells is maintained by a constitutive monovalent cation leak, but the responsible channels remain unidentified^140^; the cardiac pacemaker ion channels might be involved here. Another possibility is involvement in the so-called “ocular pulse”, i.e., the rhythmic wave of aqueous outflow that syncs with the cardiac pulse^21^. In high-tension glaucoma the tissues become less sensitive to the pulse, likely because of fibrotic pathophysiological processes^141^. Schlemm’s canal exhibits a hybrid lymphatic/blood vascular endothelial phenotype that responds to the ocular pulse similar to the TM, thus playing a crucial role in aqueous outflow and intraocular pressure regulation^142^.

The third pathway identified was “glycosaminoglycan metabolism”. These highly glycosylated molecules, including hyaluronic acid, chondroitin sulfates, keratan sulfates, and heparan sulfates, are crucial components of the TM, contributing to the filtration barrier, and affecting resistance to aqueous outflow ^143,144^. Defects in other biosynthetic and recycling pathways that we identified could synergize with GC-induced fibrosis to activate the UPR^58–61^, thus tipping the balance to pathology.

We also identified two significant pathways associated with energy metabolism that included the top prioritized gene *PPARGC1A*: “adipogenesis” and “mitochondrial biogenesis”. PPARGC1A, a coactivator of nuclear receptors including the GR, acts as a ‘molecular switch’ in multiple pathways controlling glucose homeostasis, and regulates mitochondrial biogenesis, hepatic gluconeogenesis, and muscle fiber-type switching ^145^. In glaucomatous optic neuropathy, alterations in lipid metabolism contribute to cellular injury and apoptosis, and mitochondrial dysfunction leads to impaired energy production that also compromises cellular viability ^146^. Disturbances in glucose metabolism, such as reduced glycolytic activity, affect energy availability and neurotrophic support that are vital for retinal ganglion cell survival ^147^. However, this may be the first evidence for disruption of energy metabolism in a high-tension ocular disorder involving the aqueous outflow pathways.

### Limitations of the study

The method used in this study to assess the QT was conservative. The timing of the follow-up visits may not have corresponded with the highest IOP attained. Steroid responders are treated with an IOP-lowering intervention once they respond, meaning these patients might never demonstrate their full potential. This likely leads to an under-estimate of the maximal change in IOP. Nevertheless, this study design proved to be quite successful for SNP discovery.

We used imputed data for rare variants, however, we were extremely conservative and only SNPs with a high quality score were retained in the analysis. Moreover, we validated our findings in several ways. Therefore, these rare variants imputed to the HRC panel with high quality appear to be trustworthy.

We prioritized genes based on their expression as detailed in scRNA datasets on Spectacle. However, the depth of single cell data is limited, so can miss some low expressing genes. Moreover, low expressing genes might be substantially upregulated in response to steroids. On the positive side, very few of the closest genes to SNPs were identified as not expressed, therefore this limitation would affect only a few of our determinations.

GC-regulated gene expression data used here for validation were obtained using primary cultured TM cell strains. This excludes other AOP tissues and likely introduces artifacts due to culture conditions. However we did have the use of AOP gene expression data from untreated eyes, curated on Spectacle, and will plan to use AOP tissues from GC-treated eyes to generate data for future studies.

The size of cohorts that we assembled for replication in this study were small and in some cases participants were of different ancestry than the discovery cohort. The Indianapolis-2 cohort was quite similar to the Indianapolis discovery cohort, however the percentage of steroid responders to non-responders was much smaller due to the method of enrollment. Participants in other cohorts were treated with different GCs, at different doses, delivered in different ways. Response to GCs in the eye has been shown to vary widely with differing parameters, likely because each type of treatment triggers different transcriptional response elements^9,10^. Despite these drawbacks, we replicated 4 common SNPs and one rare SNP in the Chennai-2 replication cohort. A feature shared by this cohort with the discovery cohort was long term delivery of GCs rather than administration by a single injection, suggesting this might be a key factor that led to success. However, while SNPs were not replicated in the other 3 cohorts, two prioritized target genes were replicated.

### Future perspectives

Similar to other high-tension ocular phenotypes, we observed overlap of risk loci between the Indianapolis-1 discovery cohort and the Indianapolis-2 replication cohort involving the same prioritized target genes, but entirely different SNPs. Thus mis-expression of specific genes may be determined by multiple SNPs, both common and rare, and suggests that many more of the rare SNPs remain to be found. Equally important, repeated identification of the same prioritized target genes suggests that overlapping gene sets will ultimately be defined across high-tension ocular phenotypes. In fact, our observation of numerous paralogous gene sets in our full prioritized gene list suggests that function will likely trump individual genes. The study reported here provides insight into how the different high-tension ocular phenotypes are related and a set of genes specifically defining SIOH has begun to take shape. Future studies will add to this.

GC use is widespread in ophthalmology and continues to be the mainstay of treatment for inflammatory eye diseases and post-surgery, where prolonged treatment is often required^9,10^. Furthermore, intravitreal steroid implants are increasingly being used for retinal vascular disease^152^. Both conditions are increasing in prevalence in our ageing society. SIOH represents a major clinical challenge and the ability to predict risk for individual patients is an attractive goal for delivery of personalized medicine. The large number of significant SNPs identified in this study set us on the road to begin using genomics for this purpose.

We began this report by introducing the concept of ADRs, a frequent cause of worldwide morbidity and mortality^1^. The SNPs and genes we discovered here for SIOH may also play a role in systemic steroid response, but more easily discoverable in SIOH because the QT is robust and a relatively pure phenotype can be isolated. Significantly, some of these genes are already pharmaceutical targets for stress-related diseases. One example is *AGTR1*, the target of replicated rare SNP rs193153124. *AGTR1* encodes angiotensin II receptor type 1, a protein implicated in blood pressure regulation, vascular tone, and fluid balance. Angiotensin-converting enzyme (ACE) inhibitors have shown efficacy for the treatment of pulmonary fibrosis in pre-clinical models^153^. Another example is SLC17A9, the target of replicated common SNP rs2427460. Also known as VNUT, SLC17A9 regulates vesicular uptake, storage and release of ATP from cells. Clodronic acid is a bisphosphonate that inhibits SLC17A9 activity. It is anti-inflammatory, approved in Australia, Canada, the UK and Italy for the prevention and treatment of osteoporosis in post-menopausal women, and in men to reduce vertebral fractures^154^. Clodronic acid has also been shown to be effective in preventing GC-induced bone loss in asthmatic patients^155^. We found that rs193153124 co-localizes with an eQTL that negatively regulates *SLC17A9* expression; here it was the only SNP we found associated with a reduction in IOP. Our results suggest that patients with the more common SLC17A9 variant might be most likely to benefit from this drug, whether used for the treatment of bone loss or for SIOH.

## SUPPLEMENTAL INFORMATION

**Figure S1.** Q-Q plots for the Indianapolis-1 discovery cohort

**Figure S2.** Q-Q and Manhattan plots for the Indianapolis-2 replication cohort

**Figure S3.** Aqueous outflow pathway (AOP) cell type expression of top prioritized target genes and prioritized target genes of replicated SNPs.

**Figure S4.** Effects of sQTLs identified in the *COL11A1* gene associated with SIOH or other high-tension ocular phenotypes.

**Table S1.** GWAS Results SNPs Indianapolis-1 Discovery Cohort

**Table S2.** GWAS Results Risk Loci Indianapolis-1 Discovery Cohort

**Table S3.** Co-localization with Other High-Tension Ocular Phenotypes

**Table S4.** Target Gene Prioritization

**Table S5.** Prioritized Target Gene Validation

**Table S6.** Top Prioritized Target Gene Additional Validation

**Table S7.** GWAS Results Indianapolis-2 Replication Cohort

**Table S8.** Independent Replication

**Table S9.** Functional Annotation Prioritized Genes

## ACKNOWLEDGEMENTS

Participant enrollment, sample management and genotyping for the Indianapolis-1 discovery cohort was funded by National Institutes of Health (NIH) project grant R01EY027315 (MEF) and grants from the Glaucoma Research Foundation (MEF) and the William E. Cross Foundation (SGS). This part of the project also received support from Research to Prevent Blindness to the New England Eye Center and the USC Keck School of Medicine.

- Statistical genetic analyses for discovery and replication were supported by a grant made to the New England Eye Center from the Massachusetts Lions Eye Research Fund and NIH grant UM1TR004398 awarded to the Tufts Clinical and Translational Science Institute. J.H. was supported by NIH training grant T32TR004417.
- The human paired eye study was supported by the Department of Biotechnology (DBT)-Wellcome Trust/India Alliance fellowship grant number IA/I/16/2/502694 (SS).
- Funding for enrollment and genotyping of the Indianapolis-2 replication cohort was provided by NIH grants R01EY023196, R01EY016482, R01EY016514, R01EY016835, R21EY015145 and P30EY011373, as well as the intramural research program of the National Human Genome Research Institute of NIH. Genotyping was performed by the Centre for Inherited Disease Research, Johns Hopkins University under NIH grant 1X01HG006619.
- Funding for the Florida-1 replication cohort was provided by grant number G2003-019 from the American Health Assistance Foundation National Glaucoma Research (SGS), an Investigator Award from Prevent Blindness America/Prevent Blindness Florida (SGS), NIH core center grant P30EY014801 (MEF) and an unrestricted grant from Research to Prevent Blindness to the University of Miami.
- Funding for the Chennai-1 and Chennai-2 replication cohorts was provided by the Indian Council of Medical Research [82/19/2012/PHGEN (TF)/BMS] (SC, RJG)
- The authors acknowledge the University of Miami Miller School of Medicine’s Hussman Institute for Human Genomics Biorepository Core Facility (RRID:SCR_017816) and Genotyping Core Facility (RRID:SCR_017820) for sample banking and genotyping services.
- The authors acknowledge the Tufts University High Performance Computer Cluster (https://it.tufts.edu/high-performance-computing) for data management.
- The content of this article is solely the responsibility of the authors and does not necessarily represent the official views of the NIH or the other funding agencies.

## AUTHOR CONTRIBUTIONS

Conceptualization, P.S., M.O.P. and M.E.F.

data curation, Z.S., S.P., A.G.M., S.G., T.I., J.H., E.M., P.H.B., S.S., S.C., A.J.G., M.O.P. and M.E.F.

formal analysis, Z.S., S.P., S.G., A.G.M., J.H., A.G., E.M., P.H.B. and M.E.F.

funding acquisition, C.E.W., S.S., R.J.G., S.C., S.K.I., S.G.S. and M.E.F.

investigation, S.P., T.I., M.O.P.

project administration, P.S., M.O.P. and M.E.F.

resources, F.W.P., C.E.W., S.S., R.J.G., S.C., J.H.L., S.K.I., S.G.S. and M.O.P.

supervision, W.D.S., F.W.P., C.E.W., S.S., R.J.G., S.C., J.H.L., S.K.I., S.G.S., P.S., M.O.P. and M.E.F.

validation, Z.S., S.P., S.G., J.H., E.M., A.J.G. and M.E.F.

visualization, Z.S., S.G., A.G.M., J.H. and M.E.F.

writing—original draft, Z.S. and M.E.F.

writing—review & editing, Z.S., S.P., S.G., T.I., A.G.M., J.H., A.G., E.M., P.H.B., W.D.S., F.W.P., C.E.W.,

S.S., R.J.G., S.C., J.H.L., S.K.I., S.G.S., A.J.G., P.S., M.O.P. and M.E.F.

## DECLARATION OF GENERATIVE AI AND AI-ASSISTED TECHNOLOGIES

During the preparation of this work, the author(s) used Google AI Overviews in order to rapidly find information on a subject, with hyperlinks to more information. After using this tool or service, the author(s) reviewed and edited the content as needed and take(s) full responsibility for the content of the publication.

## METHODS

### KEY RESOURCES TABLE

| REAGENT or RESOURCE | SOURCE | IDENTIFIER |
| --- | --- | --- |
| Critical commercial assays |  |  |
| Oragene DNA Self-Collection Kit | DNA Genotek, Inc.,<br>Ottawa, Ontario,<br>Canada | OG-500 |
| preplT L2P Kit | DNA Genotek, Inc.,<br>Ottawa, Ontario,<br>Canada | PT-L2P |
| Deposited data |  |  |
| Genotyping data, Indianapolis-1 cohort | This paper | In process of being submitted to NCBI dbGAP |
| Genotyping data, Florida-1 cohort | Gerzenstein et al. (32); Patel et al. (175); Jeong et al. (81); Fini et al. (9) | In process of being submitted to NCBI dbGAP |
| Genotyping data, Indianapolis-2 cohort | Igo et al. (162); Louttit et al. (163); Afshari et al. (174) | NCBI dbGaP accession code phs000421.v1.p1 |
| Genotyping data (whole genome sequencing), Chennai-1 cohort | Badrinarayanan et al. (82) | In process of being submitted to NCBI dbGAP |
| Genotyping data (whole genome sequencing), Chennai-2 cohort | This paper | In process of being submitted to NCBI dbGAP |
| Cultured trabecular meshwork cells (human), differentially-expressed gene lists (supplementary material in published paper) | Nehme et al. (92) |  |
| Cultured trabecular meshwork cells (human), paired eye study, differentially-expressed gene lists deposited in NCBI-SRA | Kathirvel et al. (96) | BioProject PRJNA729873 |
| Cultured trabecular meshwork cells (bovine), paired eye study, differentially-expressed gene lists (supplementary material in published paper) | Mao et al. (98) |  |
| EMBL Nucleotide Sequence Database | Venter et al. (183) | https://www.ebi.ac.uk/ |
| Encyclopedia of DNA Elements (ENCODE) | EP Consortium (95) | http://encodeproject.org |
| The GeneCards Suite | Stelzer et al. (112) | https://www.genecards.org/ |
| Genotype-Tissue Expression (GTEx) Portal | GT Consortium (91); GT Consortium (108) | https://gtexportal.org/home/ |
| HaploReg v4.2 | Ward et al. (106) | https://pubs.broadinstitute.org/mammals/haploreg/haploreg.php |
| Medline Plus |  | https://medlineplus.gov/ |
| NIH NCBI DAVID Bioinformatics | Huang da et al (93); Huang da et al (94) | https://davidbioinformatics.nih.gov/home.jsp |
| NIH NCBI dbSNP | Sherry et al. (180) | https://www.ncbi.nlm.nih.gov/snp/ |
| NIH NCBI Gene | Brown et al. (178) | https://www.ncbi.nlm.nih.gov/gene/ |
| NIH NCBI GRASP | Welter et al. (181) | https://grasp.nhlbi.nih.gov/Search.aspx |
| Reactome | Milacic et al. (113) | https://reactome.org/ |
| RegulomeDB | Boyle et al. (182) | https://regulomedb.org/regulome-search/ |
| Spectacle | Voigt et al. (90) | https://singlecell-eye.org/app/spectacle/ |
| UCSC Genome Browser | Perez et al. (177) | https://genome.ucsc.edu/index.html |
| Software and algorithms |  |  |
| BioRender Scientific Image and Illustration Software |  | https://www.biorender.com/ |
| Manhattan Plot Visualization | Core Team (173) | https://www.r-project.org/ |
| Michigan Imputation Reference Panel | Luo et al. (172) | https://imputationserver.sph.umich.edu/#! |
| *meta* R package | Hartung and Knapp (179) |  |
| nf-gwas-pipeline: A Nextflow Genome-Wide Association Study Pipeline | Song et al. (169) | https://genepi.github.io/nf-gwas/ |
| PLINK 2.0 | Chen et al. (176) | https://www.cog-genomics.org/plink/2.0/ |
| PC-AiR | Conomos et al. (170) | https://rdr.io/bioc/GENESIS/man/pcair.html |
| PC-Relate | Conomos et al. (171) | https://rdr.io/bioc/GENESIS/man/pcrelate.html |
| Quality control and quality assurance in genotypic data for genome-wide association studies | Laurie et al. (167);<br>Truong et al. (168) | https://currentprotocols.onlinelibrary.wiley.com/doi/full/10.1002/cpz1.373 |
| Manhattan Plot Visualization | Core Team (173) | https://www.r-project.org/ |

### RESOURCE AVAILABILITY

#### Lead contact

Further information and requests for resources and reagents should be directed to the lead contact, M. Elizabeth Fini

#### Materials availability

Saliva and DNA samples generated in this study are being banked at the University of Miami Miller School of Medicine’s Center for Genome Technology (CGT). Some samples may be available for collaborative studies. Please contact M Elizabeth Fini for more information.

#### Data availability

- New genotyping data are being deposited at dbGAP: the accession numbers (once obtained) will be publicly available as of the date of publication.
- Any additional information required to reanalyze the data reported in this paper is available from the lead contact upon request.

### STUDY PARTICIPANT DETAILS

Demographic and clinical data on the discovery cohort and the 4 replication cohorts included in this study are shown in Table 1.

#### Discovery cohort

All participants in the Indianapolis-1 discovery cohort were Fuch’s Endothelial Corneal Dystrophy (FECD) patients enrolled into the registry of the Cornea Research Foundation of America (CRFA, Indianapolis, IN, USA) following Descemet’s Membrane Endothelial Keratoplasty (DMEK) or Descemet’s Stripping Endothelial Keratoplasty (DSEK) at the Price Vision Group (PVG, Indianapolis, IN, USA) between the years 2003 and 2012. Patients were instructed to self-administer 1% prednisolone acetate eyedrops following surgery. Participants were enrolled from the registry into this study without regard to ancestry, however those selected for the GWAS (n=439) were all of European ancestry.

The Indianapolis-1 discovery cohort was compliant with the Declaration of Helsinki, the Health Insurance Portability and Accountability Act (HIPAA), and the Institutional Review Boards (IRBs) of the University of Southern California, Los Angeles, CA, USA and Tufts University, Boston, MA, USA.

#### Replication cohorts

Indianapolis-2 replication cohort participants were FECD patients enrolled into the registry of the CRFA following DMEK or DSEK at the PVG between the years 2006-2007 (these were different patients than enrolled into the Indianapolis-1 discovery cohort). The Indianapolis-2 cohort was a subset of patients originally enrolled in a larger multicenter study to map genes for FECD^162,163^. They all had corneal transplantation like the Indianapolis-1 discovery cohort, and were similarly instructed to self-administer 1% prednisolone acetate eyedrops following surgery. However, the percentage of steroid responders to non-responders was much smaller than for the Indianapolis-1 discovery cohort due to the method of enrollment. Those participants selected for GWAS were all of European ancestry (n=104).

Florida-1 replication cohort participants underwent a single intravitreal injection with triamcinolone acetate (IVTA; single injection of 4 mg) for various retina inflammatory diseases. This cohort was previously used for discovery in an SIOH candidate gene study^32^ and for replication in our Florida SIOH pilot study^81^. Participants selected for GWAS were all of European ancestry (n=50).

Chennai-1 replication cohort (n=52) and the Chennai-2 replication cohort (n=50) participants were treated for various retinal diseases by IVTA (single injection of 2 mg; Chennai-1) or an intravitreal dexamethasone slow-releasing implant (Oxurdex® intravitreal implant; Chennai-2). The Chennai-1 cohort was previously used as the discovery cohort in our Chennai pilot study^82^. The Chennai-2 cohort was used as the discovery cohort for a second study (manuscript in preparation). Participants in both cohorts were of Asian ancestry.

Replication cohorts were in compliance with the Declaration of Helsinki, HIPAA and the IRBs of the Case Western Reserve University, the University of Miami, Miami, FL, USA, the University of Southern California, Los Angeles, CA, USA, and the Vision Research Foundation, Chennai, Tamil Nadu, India. All subjects provided informed written consent under IRB-approved protocols.

### METHODS DETAIL

#### Study design for the discovery cohort

Qualified participants were selected from the CRFA registry with the aim of finding as many steroid responders as possible, with at least an equal number of non-responders. A steroid responder was defined as an individual that exhibited an IOP ≥ 6 mm Hg above baseline.

Patients were contacted for participation in the Indianapolis-1 discovery cohort if they met the following inclusion-exclusion criteria:

#### Inclusion Criteria

- Age 18 years or greater
- Normal baseline IOP (between 10 and 21 mm Hg)
- Had DMEK or DSEK surgery for FECD
- Had post-surgical treatment with 1% PA eye drops according to standard practice protocol
- Baseline IOP measurement and at least two IOP measurement in the study eye over 6-12 months by Goldman applanation tonometry

#### Exclusion Criteria

- Prior pars plana vitrectomy in the study eye
- Prior trabeculectomy or glaucoma tube shunt surgery in the study eye
- Anterior segment neovascularization in the study eye
- A history of glaucoma, suspected glaucoma, or OH in the study eye
- Use of any medication that could lower IOP at the time of study enrollment
- IOP spiking immediately following surgery due to surgical complications
- Any other eye surgery performed within the first year after DMEK or DSEK for FECD

Patients were instructed to instill topical prednisolone acetate 1% eye drops, 4 times daily for ≥4 months, then taper by 1 drop per month to once daily (the exact timing of the taper varied). This regimen was continued indefinitely unless the patient developed ocular hypertension, in which case the topical corticosteroid strength or dosing frequency was reduced and/or topical glaucoma medication was added as needed to maintain IOP within the target range. All IOP measurements were done with Goldmann applanation tonometry (Haag-Streit, Bern, Switzerland). The baseline IOP was recorded prior to surgery and at follow-up visits at ∼1, 3, 6, and 12 months after surgery.

FECD transplant patients are ideal for studying SIOH^164^. They seldom require medication after endothelial keratoplasty (EK) other than topical steroids. EK is relatively atraumatic to the cornea and angle structures. Clear corneal incisions are made that typically avoid the TM, and the incisions are small (2-5 mm). Almost all of the patients had cataract surgery before, or combined with EK; the intraocular lens was placed in the capsular bag, minimizing the risk of inflammation. Cataract surgery can be associated with a modest reduction in IOP (mean 1 mm Hg)^165^. A larger and more sustained reduction in IOP after cataract surgery may occur in patients with a history of open angle glaucoma, but that was a study exclusion. FECD affects only the cornea. This is important, as the confounding effects of other systemic or ocular phenotypes that could affect SIOH are minimized.

An increase in IOP can occur in the immediate period following corneal transplantation surgery^166^. Reasons for this include air bubble-induced pupillary block and (aqueous flow from the ciliary body through the pupil to the anterior chamber is blocked by the air bubble). These complications occur very infrequently at the practice from which participants in this study were derived. Synechiae formation that could alter aqueous outflow in these patients is also rarely seen. Any patients that did experience IOP increase in the immediate period following surgery were excluded from the study. Subsequent IOP increase was attributed to response to GC treatment^166^.

A total of 520 participants were consented and enrolled. Most subjects self-identified as white, non-hispanic, which is typical of FECD demographics. Participants were handed or mailed a saliva collection kit (OG-500, Oragene DNA Self-Collection Kit, DNA Genotek, Inc., Ottawa, Ontario, Canada) labeled with a coded identification number for confidentiality. Participants could fill and return the kit during an office visit or send it back in a prepaid mailing packet. Saliva was obtained from a total of 480 participants (93%) meeting study inclusion and exclusion criteria. A portion of DNA was extracted from each sample using the prepIT L2P Kit (DNA Genotek, Inc.). Extracted DNA was stored at −70°C and the remaining saliva was stored at ambient temperature as recommended by the kit manufacturer.

#### Genotyping and GWAS quality control for the discovery cohort

After enrollment of the Indianapolis-1 cohort was complete, DNA and saliva samples were transferred to the University of Miami Miller School of Medicine’s Center for Genome Technology (CGT). DNA samples were quantified by Nanodrop, and their quality was evaluated by Qubit (for concentration) and gel electrophoresis (for intactness). Any DNA samples that failed these quality control (QC) assessments were re-extracted from the banked saliva.

Final DNA samples from 471 subjects passed QC review. These samples were genotyped on the Illumina Infinium Global Screening Array-24 BeadChip (Illumina, Inc., San Diego, CA). Genotyping reproducibility was confirmed by including 5 HIHG controls and 1 blind duplicate. Sample call rates and SNP statistics were calculated. Of the 471 samples genotyped, 460 unique samples plus the 5 HIHG controls displayed >98% call rate.

QC was performed at both sample and SNP levels in accordance with Illumina protocols and published procedures^167,168^. Principal components of genetic ancestry and genetic relationship matrix (GRM) for cryptic relationships were inferred using PC-AiR and PC-Relate respectively, both implemented in our analysis pipeline^169–171^.

Of the 460 well-performing genotypes, 21 samples with mismatch genders/ DNA sample swap/missing age value and/or non-European ancestry were dropped. This left 439 samples for GWAS.

#### QT for the discovery cohort

Maximum change in IOP (deltaIOPmax) was used as the quantitative trait (QT). This was defined as the difference between the pre-surgical baseline IOP and the maximum IOP recorded in post-surgical follow-up. Two QTs were calculated: 1) deltaIOPmax across the entire 12 months of the study and 2) deltaIOPmax for the first 3 months post-surgery and initiation of GC treatment. All samples that passed QC were used in the 12 month analysis (*n*=439). Samples for the 3 month analysis included only those participants whose IOP was recorded for both the 1 month and 3 month visits (*n*=421).

#### GWAS for the discovery cohort

Linear regression was performed to determine association between QT and genotype. Genotype data were imputed using the HRC reference panel on the Michigan Imputation Server^172^. SNPs were filtered at R^2^=0.7 and a minor allele count (MAC) ≥ 3. The Manhattan plots are shown in Figure S2. The genomic control parameters were calculated to confirm no inflation introduced by population stratification and a quantile–quantile (Q–Q) probability plot was generated to visualize the distribution of the test statistics (Figure S1).

Single SNP association analysis was conducted using linear mixed-effects models, treating the QT as a continuous outcome variable and using the expected dosage of each SNP as the main covariate. The models were adjusted for baseline IOP, age, sex, the first four principal components of genetic ancestry and include GRM as a random effect to account for cryptic relationships among individuals. An additive genetic effects model was assumed.

Genotype data were imputed with the Haplotype Reference Consortium on the GRCh38 reference panel available on the Michigan Imputation Server^172^. Of the 481,134 SNPs successfully genotyped, 475,122 SNPs passed quality check on the server. After imputation, approximately 9.3 million SNPs were obtained. Imputed SNPs retained had an imputation R^2^ > 0.7 and a minor allele count (MAC) ≥ 3. Duplicated and ambiguous SNPs were removed.

Manhattan plots and Q-Q plots were constructed and Lambda statistics were determined using R^173^.

#### Study design, genotyping, QC, QT and GWAS for the replication cohorts

**Indianapolis-2 cohort.** Participants were selected for the Indianapolis-2 replication cohort without regard for their steroid-responder status. Participants were included in the Indianapolis-2 cohort if they met the same inclusion-exclusion criteria used for the Indianapolis-1 discovery cohort. As for the Indianapolis-1 cohort, the baseline IOP was recorded prior to surgery and at follow-up visits at ∼1, 3, 6, and 12 months after surgery.

DNA samples were genotyped on the Illumina 2.5M genotyping array (Illumina, Inc., San Diego, CA), as described^174^. Of the total original participants enrolled by the CRFA into the FECD study and genotyped, n=104 of them fit the inclusion-exclusion criteria used for the Indianapolis discovery cohort.

DeltaIOPmax was used as the QT, calculated at 12 months and 3 months post-surgery and initiation of GC treatment, as for the Indianapolis-1 cohort. However, the group of early 3 month responder participants identified in the Indianapolis-1 cohort were not apparent in the Indianapolis-2 cohort, likely because of the different method used for enrollment. All samples that passed QC were used in the 12-month analysis (*n*=104). Samples for the 3-month analysis included only those participants whose IOP was recorded for both the 1- and 3-month visits (*n*=102). GWAS QC, population stratification and linear regression analysis was performed as for the Indianapolis-1 cohort.

The Manhattan and Q-Q plots are shown in Figure S2. The list of top SNPs (p<5E-06) resulting from linear regression analyses are provided in Table S6, ordered by P value. None of the SNPs attained P-values exceeding the threshold for genome-wide significance (5E-08), although one SNP in the 12 month analysis came very close (rs74649788; p=5.53E-08). This is a rare intronic SNP within the gene for *L3MBTL4* on chromosome 18. The 12 month and 3 month QT results were merged in Table S6, then sorted by chromosomal position to reveal a total of 81 individual risk loci that clustered multiple top SNPs, with one overlapping between the two QT analyses (*PCBD1;UNC5B*).

**Florida-1 cohort.** Recruitment and inclusion-exclusion criteria for participants in the Florida-1 replication cohort were similar to the Indianapolis cohorts as previously reported^81^. IOP was measured in follow-up visits for up to 12 months.

DNA samples were genotyped in 2007 on the Affymetrix GeneChip Human Mapping 500K Array Set^175^. This was the first commercially-available genotyping platform, comprised of two chips with a total of only 1 million SNPs. DeltaIOPmax at 12 months post GC-treatment was used as the QT for those participants surviving QC (n=50). A GWAS was performed previously^81^. Imputation was not possible with the data as coded on the Affymetrix chips.

**Chennai-1 and Chennai-2 cohorts.** Recruitment and inclusion-exclusion for the 52 participants in the Chennai-1 cohort were previously described^82^. Inclusion-exclusion criteria for the 50 participants of the Chennai-2 cohort were the same (manuscript in preparation). IOP measurements were made at 1, 3, and 6 months after GC treatment was begun.

DNA samples were whole genome sequenced. Low pass 5 × Whole Genome Sequencing (WGS) (Illumina Hi Seq X10) was performed at Medgenome laboratories, Bangalore. Sequencing of the prepared DNA libraries was carried out in Hi Seq X10, which generates 2X150 bp sequence reads at 5X sequencing depths (∼15 GB). A minimum of 75% of the sequenced bases was of Q30 value. FASTQ files were generated from the sequence data and was used for further analysis.

DeltaIOPmax at 6 months post GC-treatment was used as the QT. The participants were then grouped into steroid responder (IOP ≥ 21mmHg), and steroid non-responder (IOP ≤ 21mmHg) categories for a case-control study design.

Quality control (QC) steps were performed using the FastQC tool. Case-control association testing was performed using the Chi-Square test implemented in PLINK^176^. Significant SNPs were identified using the Benjamini & Hochberg multiple corrections method with a p-value cut off of P<0.05^82^.

#### Risk locus overlap with other high tension ocular phenotypes

A risk locus was defined here as an ∼500 kb region of the genome that clusters a group of significant SNPs and was characterized by the gene(s) closest to most members of the cluster. Co-localization of SIOH risk loci with those discovered for other high-tension ocular phenotypes was investigated by cross-referencing with published reports^72,73,83–87^.

#### Target gene prioritization

To determine aqueous outflow pathway (AOP) expression, we searched the two relevant human scRNA-seq datasets on the Spectacle portal^90^. If the closest gene(s) to the hit SNPs was not AOP-expressed, we paired Spectacle with the UCSC Genome Browser^177^ to identify the next closest AOP-expressed gene(s), up and downstream within 500 KB of the SNP cluster. To identify additional target gene candidates, we searched SNPs at each risk locus on the Adult Genotype Tissue Expression (GTEx) Project portal^91^ to determine co-localization with expression or splicing quantitative trait loci (eQTLs/sQTLs). Target genes of expression QTLs were searched on Spectacle and if they were AOP-expressed, they were included in the prioritized gene list.

#### Target gene validation

We sought to validate our prioritized target genes by comparison to results of transcription profiling studies in cultured cells, and by computational analysis using available datasets.

We first determined whether prioritized target genes were GC-regulated, either directly by the activated GR, or as downstream targets of upstream effectors regulated by the activated GR by comparing our prioritized target gene list to lists of GC regulated genes in cultured TM cells. Since differentially-expressed genes (DEGs) vary according to the specific GCs used and treatment parameters^119–124^, we chose for this comparison a recent study co-authored by a member of our team that used RNA-seq to comprehensively profile GC-regulated DEGs, using three GCs with different chemical structures, treating primary TM cell strains isolated from two different individuals^92^.

To identify prioritized target genes that can be regulated directly by the GR, we used the NIH DAVID Bioinformatics functional annotation clustering tool ^93,94^ paired with the UCSC_TF Binding Site database that curates CHIP-seq data accessed from the ENCODE project^95^. This was run at low classification stringency, and we then determined prioritized target genes that clustered with the GR.

We also compared our prioritized target gene list to lists of DEGs associated with steroid response in paired eye studies. In this experimental design, one eye from an individual donor is exposed to GCs while being perfused in organ culture. IOP is then measured to determine if the eye is a steroid responder or non-responder and the fellow eye is used to establish a cultured primary TM cell strain to profile GC-regulated DEGs. For our analysis, we used the DEG list compiled in a human paired eye study conducted by members of our team^96^, as well as DEG lists from two studies conducted with bovine donor eyes^97,98^.

In our final analysis, we evaluated upstream regulators of our prioritized target gene list, comparing to human paired eye study DEGs expressed by cultured TM cells. team^96^. To do this, we ran the gene lists through the DAVID plus UCSC_TF binding site analysis as above, but this time at moderate stringency.

To identify any additional genes that might be GC-regulated among the top 26 prioritized target genes, we searched on HaploReg v4.2 for all SNPs clustered at the 26 risk loci ^106^.

#### SNP replication analysis

SNPs in the discovery dataset with a p<5.0E-06 were selected for replication testing. The association of these SNPs with the outcome was tested using the same linear mixed effect model adjusted for baseline IOP, age, sex, the first four principal components of genetic ancestry and include GRM as a random effect.

#### Gene-based burden testing

A gene-based SNP aggregation approach, as previously described^107^, was used for independent replication. Variants within each gene region were aggregated using an effect-signed MAF-weighted score. Gene boundaries (start and end positions) were defined based on the NCBI gene database^178^. Associations were tested using the same linear mixed-effects model as used in the GWAS analyses.

##### Annotation

We determined expression in the aqueous outflow pathways using data curated on Spectacle^90^. We obtained MAF ALPHA values by searching NCBI’s dbSNP^180^. We obtained gene product information from the GeneCards Suite^112^, examined genotype-phenotype association using the GTEx portal^91,108^ and the NIH Genome-Wide Repository of Associations Between SNPs and Phenotypes (GRASP) portal^181^. We performed *in silico* analyses using tools on the RegulomeDB^182^ and HaploReg v4.2^106^ servers. We extracted genomic position localization and sequence information from the University of California Santa Cruz (UCSF) Genome Browser^177^ and the EMBL Nucleotide Sequence Database^183^. We accessed disease phenotype information sourced on MedLine Plus. We performed transcription factor (TF) analysis analysis using the NIH DAVID Bioinformatics functional annotation tool paired with the UCSF_TF database and pathway enrichment analysis using the NIH DAVID Bioinformatics functional annotation tool paired with the Reactome database^113^.

