## Supplementary material for "Pharmacogenomics of steroid-induced ocular hypertension: relationship to high-tension glaucomas and new pathophysiologic insight": Suppl Figure S1-S4

**Supplementary Figure S1. Q-Q plots for the Indianapolis-1 discovery cohort.**

Quantile-quantile (Q-Q) plots showing the distribution of the observed P values from the logistic regression analysis for the GWAS scan against the expected distribution under the null hypothesis. The two quantitative traits (QTs) used are indicated. The x-axis of QQ-plot reports the expected −log10 (p-value) and the y-axis reports the observed −log10 ( p -value). The genomic control parameters (λ), which are close to 1, indicate that population stratification is addressed properly.

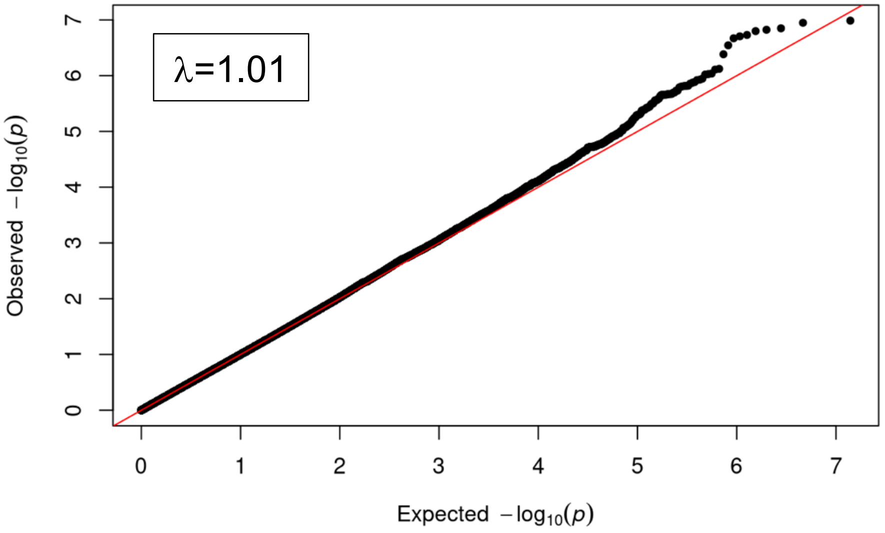


12 month QT

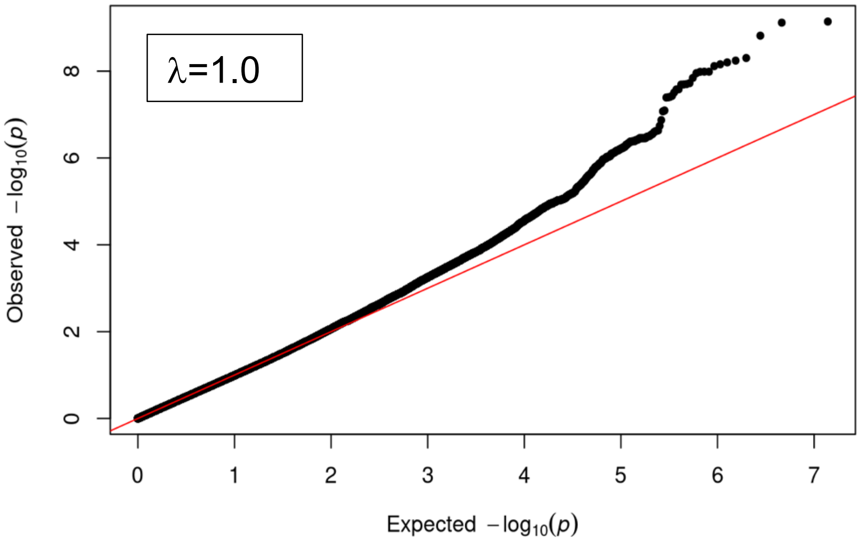


3 month QT

**Supplementary Figure S2. Q-Q and Manhattan plots for the Indianapolis-2 replication cohort.**

Quantile-quantile (Q-Q)-plot (left) and Manhattan plot (right) of the GWAS conducted for the Indianapolis-2 cohort with the quantitative trait (QT) as indicated. The x-axis of Q-Q-plot reports the expected −log10 (p-value) and the y-axis reports the observed −log10 ( p -value). The x-axis of Manhattan plot reports chromosomes and coordinates within chromosomes. The y-axis reports the −log10 ( p -value). The genomic control parameter (λ), which is close to 1, indicates that population stratification is addressed properly.

12 month QT

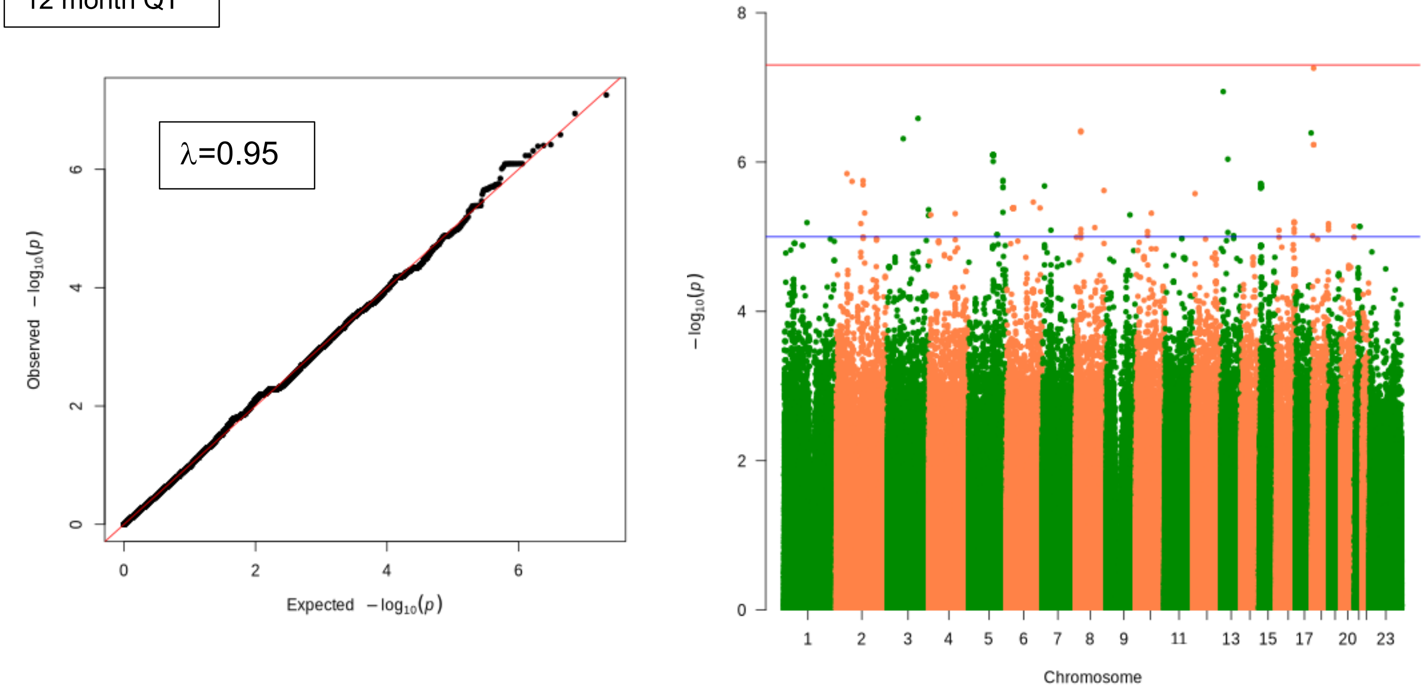


3 month QT

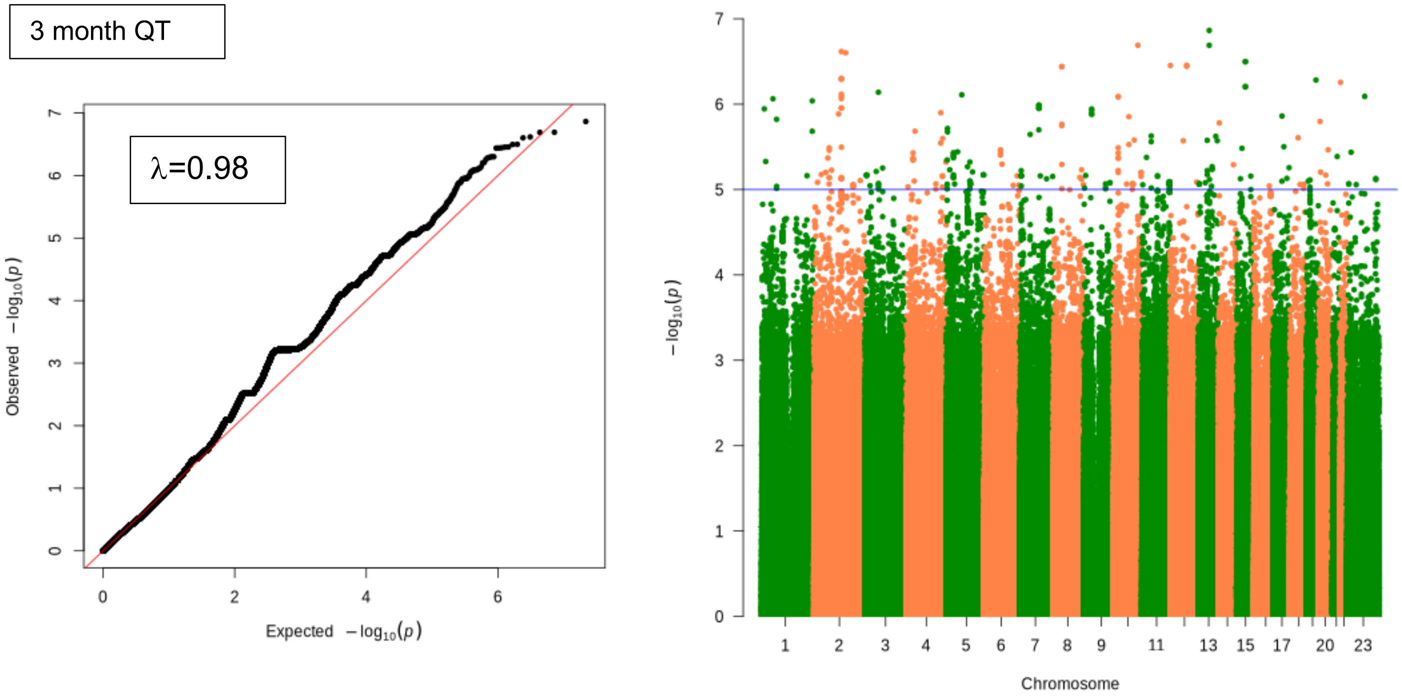

**Supplementary Figure S3. Aqueous outflow pathway (AOP) cell type expression of top prioritized target genes and prioritized target genes of replicated SNPs.**

Gene expression heat map as viewed in the van Zyl human scRNA-seq dataset hosted on Spectacle. The top prioritized target genes are in red. The 3 prioritized target genes of replicated SNPs are in blue.

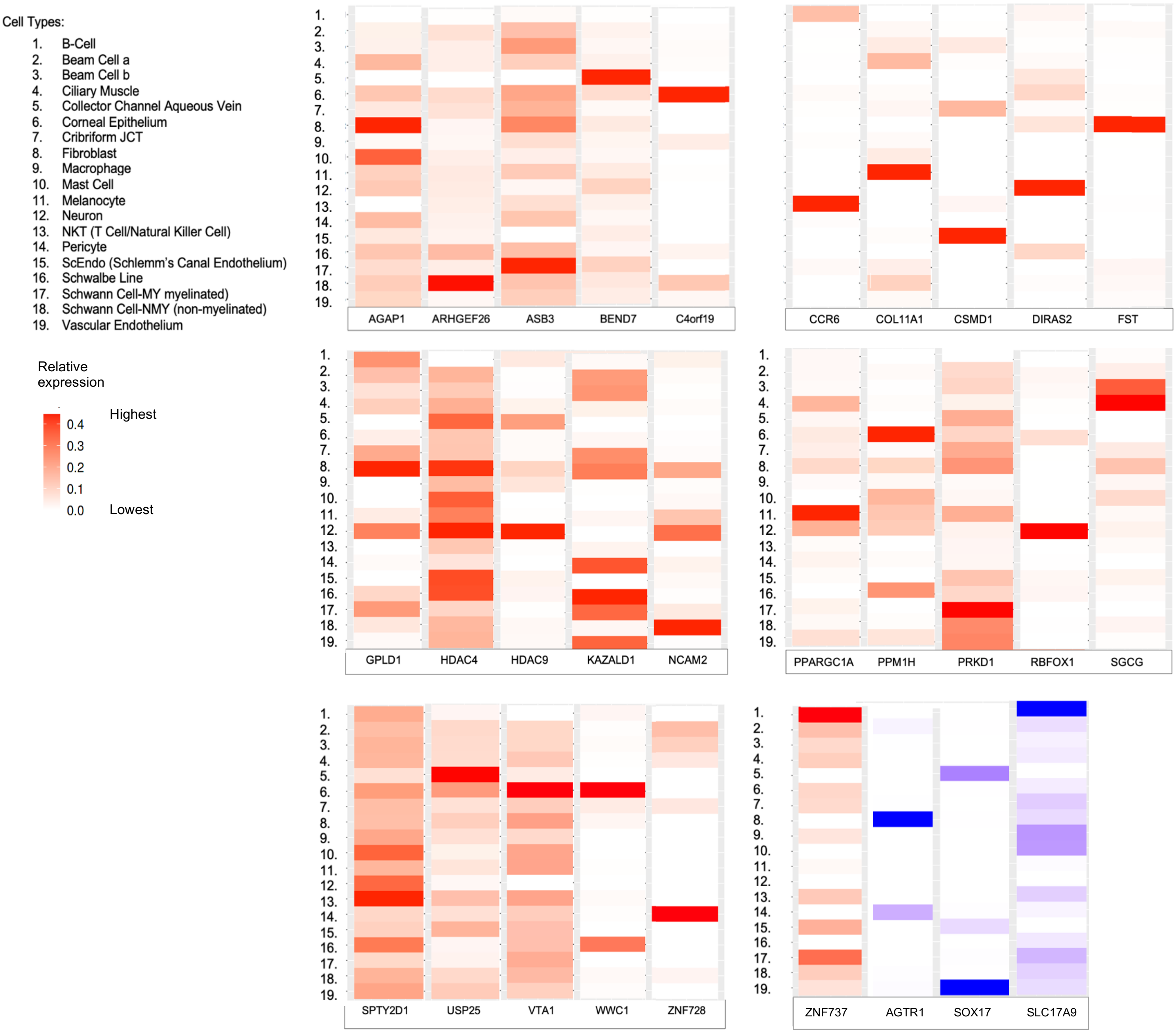

**Supplementary Figure S4. Effects of sQTLs identified in the *COL11A1* gene associated with SIOH or other high-tension ocular phenotypes.** Each of the indicated SNPs was searched on the GTEx portal. Shown are QTL violin plots depicting the effect of the minor allele on splicing (tissue type: testis).

*COL11A1* gene: 66 introns and 67 exons

Positional Information from Genome Build GRCh38:

- Chr1:102,876,473-103,108,521 = 232,049 bp
- Chr1: 102,979,612-102,984,138 = 8,885 bp = alternative splicing, intron 31
- Chr1: 102,946,956-102,961,866 = 14,911 bp = intron 41
- Chr1: 102,883,341-102,886,807 = 3,467 bp = intron 49
- Chr1: 102,888,311-102,886,807 = 3,497 bp = intron 62

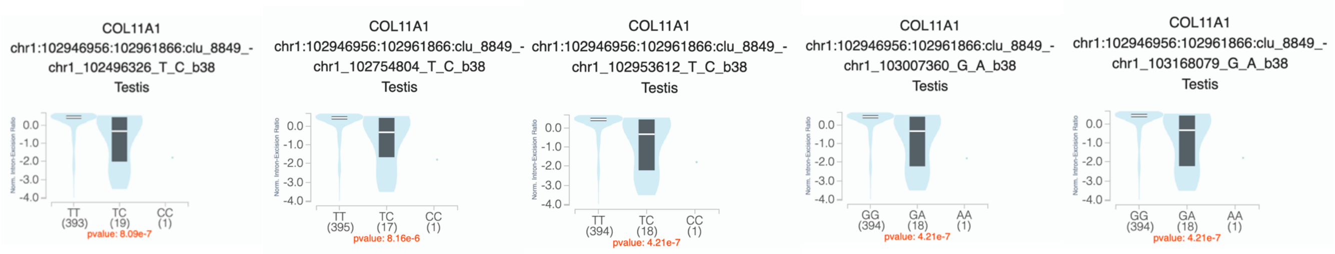


| | | | | |
|---|---|---|---|---|
| SIOH rs77180278<br>negative effect | SIOH rs112351653<br>negative effect | SIOH rs114413507<br>negative effect | SIOH rs116672066<br>negative effect | SIOH rs111928960<br>negative effect |

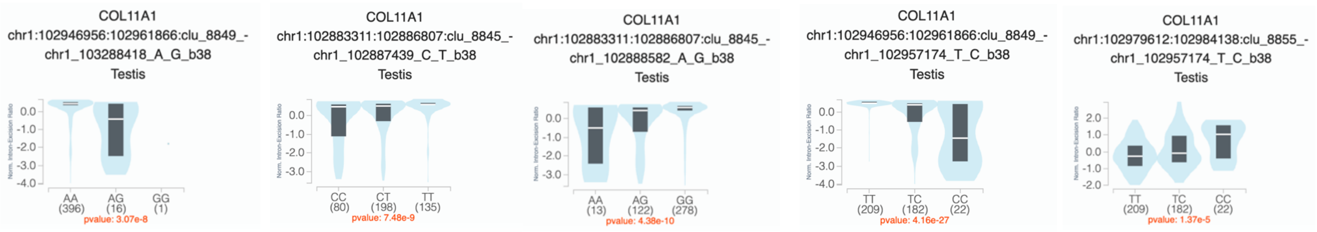


| | | | | |
|---|---|---|---|---|
| SIOH rs113221952<br>negative effect | PACG/POAG rs11164630<br>positive effect | PACG rs1676486<br>positive effect | PEXS rs3102053<br>negative effect | PEXS rs3102053<br>positive effect |
