## Supplementary material for "Pharmacogenomics of steroid-induced ocular hypertension: relationship to high-tension glaucomas and new pathophysiologic insight": Suppl Table S1

### Supplementary Table S1. GWAS Results SNPs Indianapolis-1 Discovery Cohort
### 12 month quantitative trait (QT)

**Headers**

QT: quantitiative trait; rsid: reference SNP cluster ID, chr: chromosome; pos_38, position of SNP on GRCh38 reference panel; Imputation_Rsq (if <0.8); REF and ALT, reference allele and alternate allele; n.obs: number of observations;
caf: common (major) allele frequency; MAC, minor allele count; Score: p-values from Score test; Score.SE; Score.Stat; Score.pval; EST; EST.SE; Func.refGene: SNP location with respect to nearest gene;
Gene.refGene; nearest gene upstream and downstream; GeneDetail.refGene: distance to nearest gene upstream and downstream; GeneDetail.refGene: distance to nearest gene upstream and downstream

**Notes**

Imputation Rsq listed only if less than 0.8

| QT | rsid | chr | pos_38 | Imputation_Rsq | REF | ALT | n.obs | caf | MAC | Score | Score.SE | Score.Stat | Score.pval | Est | Est.SE | Func.refGene | Gene.refGene | GeneDetail.refGene |
|---|---|---|---|---|---|---|---|---|---|---|---|---|---|---|---|---|---|---|
| 12 mo | rs142934021 | 5 | 53366842 | 0.721159995 | G | A | 439 | 0.0079727 | 7 | 1.70971028 | 0.301367945 | 5.673165672 | 1.40E-08 | 18.82471499 | 3.318202936 | intergenic | LOC257396;FST | dist=251716;dist=113787 |
| 12 mo | rs138164904 | 16 | 6808238 | | C | T | 439 | 0.0034169 | 3 | 1.270258009 | 0.230575889 | 5.509066939 | 3.61E-08 | 23.89264096 | 4.336966899 | intronic | RBFOX1 | . |
| 12 mo | rs138138661 | 3 | 147940027 | 0.736020029 | C | T | 439 | 0.0045558 | 4 | 1.27663259 | 0.23490951 | 5.434571772 | 5.49E-08 | 23.13474571 | 4.256958355 | intergenic | LOC440982;LINC02032 | dist=430117;dist=138132 |
| 12 mo | rs147669485 | 18 | 29023686 | | G | T | 439 | 0.0034169 | 4 | 1.284305415 | 0.241008458 | 5.3288811 | 9.88E-08 | 22.11076388 | 4.149231981 | intergenic | CDH2;MIR302F | dist=846556;dist=1275226 |
| 12 mo | rs148997617 | 3 | 148037789 | | C | T | 439 | 0.0045558 | 4 | 1.354154489 | 0.254680453 | 5.317072728 | 1.05E-07 | 20.87742767 | 3.926489017 | intergenic | LOC440982;LINC02032 | dist=527879;dist=40370 |
| 12 mo | rs74572772 | 22 | 34096660 | | A | G | 439 | 0.0182232 | 16 | 2.712607909 | 0.515127295 | 5.265898221 | 1.40E-07 | 10.22251833 | 1.941267739 | intergenic | LARGE1;LINC02885 | dist=173837;dist=660007 |
| 12 mo | rs148157126 | 20 | 32361036 | | C | T | 439 | 0.0045558 | 5 | 1.399252324 | 0.26592767 | 5.261777862 | 1.43E-07 | 19.78650011 | 3.760421027 | intronic | ASXL1 | . |
| 12 mo | rs80019988 | 22 | 34097658 | | G | A | 439 | 0.0182232 | 16 | 2.739120335 | 0.52174256 | 5.249946134 | 1.52E-07 | 10.06233062 | 1.916654069 | intergenic | LARGE1;LINC02885 | dist=174835;dist=659009 |
| 12 mo | rs186792608 | 2 | 42869559 | | A | G | 439 | 0.0034169 | 3 | 1.224622659 | 0.233550544 | 5.243501632 | 1.58E-07 | 22.45124992 | 4.281728413 | intergenic | HAAO;LINC01819 | dist=76976;dist=158293 |
| 12 mo | rs9974985 | 21 | 39935648 | 0.79768002 | G | A | 439 | 0.8861048 | 114 | -6.320045974 | 1.205919394 | -5.240852753 | 1.60E-07 | -4.345939521 | 0.829242821 | intergenic | PCP4;DSCAM | dist=6256;dist=75353 |
| 12 mo | rs113154814 | 2 | 68062365 | 0.753430009 | T | C | 439 | 0.0056948 | 5 | 1.38111384 | 0.263901723 | 5.233440026 | 1.66E-07 | 19.83101879 | 3.789289395 | intronic | C1D | . |
| 12 mo | rs1005412 | 21 | 39937023 | 0.782010019 | A | G | 439 | 0.8872437 | 112 | -6.127850017 | 1.173197527 | -5.223204001 | 1.76E-07 | -4.452109624 | 0.852371384 | intergenic | PCP4;DSCAM | dist=7631;dist=73978 |
| 12 mo | rs193041547 | 20 | 32184077 | | T | C | 439 | 0.0045558 | 4 | 1.384000945 | 0.265194908 | 5.218806627 | 1.80E-07 | 19.67913585 | 3.770811462 | intergenic | TM9SF4;TSPY26P | dist=16819;dist=5069 |
| 12 mo | rs77297738 | 22 | 34077554 | | C | T | 439 | 0.0182232 | 16 | 2.636152158 | 0.507182742 | 5.197637737 | 2.02E-07 | 10.24805718 | 1.971675922 | intergenic | LARGE1;LINC02885 | dist=154731;dist=679113 |
| 12 mo | rs146249289 | 20 | 32094428 | | C | T | 439 | 0.0045558 | 4 | 1.377562106 | 0.265036842 | 5.197624967 | 2.02E-07 | 19.6109527 | 3.773060354 | intronic | HCK | . |
| 12 mo | rs9636964 | 21 | 39932840 | 0.797720015 | A | G | 439 | 0.8849658 | 115 | -6.21374108 | 1.208541419 | -5.141520995 | 2.73E-07 | -4.254319226 | 0.827443714 | intergenic | PCP4;DSCAM | dist=3448;dist=78161 |
| 12 mo | rs76356799 | 3 | 179875980 | | G | A | 439 | 0.0045558 | 4 | 1.36392794 | 0.265929638 | 5.128905337 | 2.91E-07 | 19.28670071 | 3.760393191 | intronic | PEX5L | . |
| 12 mo | rs7275595 | 21 | 39935998 | 0.794510007 | G | A | 439 | 0.8883827 | 113 | -6.12484339 | 1.198927666 | -5.108601265 | 3.25E-07 | -4.260975379 | 0.834078676 | intergenic | PCP4;DSCAM | dist=6606;dist=75003 |
| 12 mo | rs192855100 | 20 | 32520938 | | G | A | 439 | 0.0045558 | 4 | 1.26803868 | 0.250556407 | 5.060891062 | 4.17E-07 | 20.19860967 | 3.99111726 | intronic | NOL4L | . |
| 12 mo | rs2427460 | 20 | 62959430 | 0.922999978 | T | C | 439 | 0.4863326 | 420 | -9.771088627 | 1.948334389 | -5.015098375 | 5.30E-07 | -2.574043965 | 0.513258918 | intronic | SLC17A9 | . |
| 12 mo | rs562831582 | 19 | 50731053 | 0.742709994 | A | C | 439 | 0.0034169 | 4 | 1.186136633 | 0.237177678 | 5.001046654 | 5.70E-07 | 21.08565484 | 4.216248377 | intergenic | CLEC11A;GPR32 | dist=5345;dist=39411 |
| 12 mo | rs149859280 | 20 | 32090367 | 0.777029991 | C | T | 439 | 0.0045558 | 5 | 1.36598448 | 0.273167403 | 5.000539835 | 5.72E-07 | 18.30577068 | 3.660758895 | intronic | HCK | . |
| 12 mo | rs138733283 | 20 | 32087884 | 0.776130021 | C | T | 439 | 0.0045558 | 5 | 1.365481855 | 0.273350907 | 4.995344152 | 5.87E-07 | 18.27447439 | 3.658301378 | intronic | HCK | . |
| 12 mo | rs80212581 | 16 | 6362966 | | C | T | 439 | 0.0034169 | 3 | 1.113691785 | 0.22359493 | 4.980845435 | 6.33E-07 | 22.27620028 | 4.472373329 | intronic | RBFOX1 | . |
| 12 mo | rs148248743 | 3 | 136415753 | 0.708679974 | C | T | 439 | 0.0022779 | 3 | 0.909706546 | 0.183031941 | 4.970206509 | 6.69E-07 | 27.15485873 | 5.463527255 | intronic | STAG1 | . |
| 12 mo | rs188353596 | 17 | 48001236 | | C | T | 439 | 0.0034169 | 3 | 1.130738257 | 0.227716806 | 4.965545927 | 6.85E-07 | 21.80579474 | 4.391419406 | intergenic | CDK5RAP3;COPZ2 | dist=19450;dist=24931 |
| 12 mo | rs113791989 | 13 | 61344755 | 0.787899971 | G | A | 439 | 0.0022779 | 3 | 0.975147098 | 0.196694436 | 4.957675046 | 7.13E-07 | 25.20495822 | 5.084027893 | intergenic | MIR3169;PCDH20 | dist=144874;dist=64931 |
| 12 mo | rs56324718 | 15 | 39987872 | | G | A | 439 | 0.0102506 | 9 | 1.956402255 | 0.395430594 | 4.947523752 | 7.52E-07 | 12.5117374 | 2.5288888 | intronic | EIF2AK4 | . |
| 12 mo | rs191298981 | 2 | 67882914 | 0.724789977 | C | T | 439 | 0.0045558 | 3 | 1.017681042 | 0.205887042 | 4.942909629 | 7.70E-07 | 24.00787141 | 4.857032236 | intergenic | LINC01812;C1D | dist=57352;dist=158216 |
| 12 mo | rs146919974 | 2 | 114495286 | 0.777989984 | T | C | 439 | 0.0045558 | 5 | 1.23575568 | 0.250278916 | 4.93751411 | 7.91E-07 | 19.72804656 | 3.995542316 | intronic | DPP10 | . |
| 12 mo | rs9305683 | 21 | 39933795 | 0.793929994 | G | A | 439 | 0.8895216 | 112 | -5.884975803 | 1.19568078 | -4.921862005 | 8.57E-07 | -4.116367918 | 0.836343627 | intergenic | PCP4;DSCAM | dist=4403;dist=77206 |
| 12 mo | rs145791959 | 20 | 32134626 | 0.783770025 | G | A | 439 | 0.0056948 | 5 | 1.351424877 | 0.274699531 | 4.919647552 | 8.67E-07 | 17.90919529 | 3.640341121 | intronic | TM9SF4 | . |
| 12 mo | rs142956968 | 10 | 23201236 | | C | T | 439 | 0.0159453 | 14 | 2.319717767 | 0.47190049 | 4.915692647 | 8.85E-07 | 10.41679921 | 2.119090829 | downstream | C10orf67 | dist=680 |
| 12 mo | rs532269430 | 13 | 73480315 | | T | C | 439 | 0.0045558 | 5 | 1.291806032 | 0.263414349 | 4.904083761 | 9.39E-07 | 18.61737516 | 3.796300403 | intergenic | KLF5;LINC00392 | dist=402772;dist=83929 |
| 12 mo | rs113063005 | 4 | 23507444 | 0.763409972 | T | C | 439 | 0.0068337 | 7 | 1.445039344 | 0.294886773 | 4.900319289 | 9.57E-07 | 16.61762999 | 3.391132089 | intergenic | GBA3;PPARGC1A | dist=687872;dist=284577 |
| 12 mo | rs147944608 | 10 | 108663951 | | C | T | 439 | 0.0125285 | 11 | 2.135371399 | 0.436244671 | 4.894893947 | 9.84E-07 | 11.22052434 | 2.292291613 | intergenic | LINC01435;XPNPEP1 | dist=594658;dist=1200815 |
| 12 mo | rs2606194 | 17 | 79214741 | | A | G | 439 | 0.9487472 | 44 | -4.201206996 | 0.858842579 | -4.891707862 | 1.00E-06 | -5.695697885 | 1.164357735 | intronic | RBFOX3 | . |
| 12 mo | rs113537164 | 2 | 68007798 | | A | C | 439 | 0.0045558 | 4 | 1.171984248 | 0.239752135 | 4.888316203 | 1.02E-06 | 20.38904135 | 4.17097432 | intergenic | LINC01812;C1D | dist=182236;dist=33332 |
| 12 mo | rs138055631 | 20 | 32192841 | | G | A | 439 | 0.0056948 | 5 | 1.347966833 | 0.276239343 | 4.879706185 | 1.06E-06 | 17.66477622 | 3.620049149 | UTR3 | PLAGL2 | NM_002657:c.*36110>0 |
| 12 mo | rs566724618 | 13 | 22889700 | | C | T | 439 | 0.0034169 | 3 | 1.064880575 | 0.218311246 | 4.877809059 | 1.07E-06 | 22.34337048 | 4.580616053 | ncRNA_intronic | LINC00621 | . |
| 12 mo | rs145764464 | 10 | 23312918 | | G | A | 439 | 0.0170843 | 15 | 2.47340447 | 0.507675165 | 4.872021796 | 1.10E-06 | 9.596730604 | 1.969763479 | intronic | C10orf67 | . |
| 12 mo | rs9981433 | 21 | 39937640 | 0.77809 | G | T | 439 | 0.881549 | 116 | -5.743602038 | 1.180422615 | -4.865716705 | 1.14E-06 | -4.122012439 | 0.84715422 | intergenic | PCP4;DSCAM | dist=8248;dist=73361 |
| 12 mo | rs139816293 | 20 | 32333540 | 0.77214998 | C | T | 439 | 0.0056948 | 6 | 1.378989447 | 0.283657989 | 4.861451108 | 1.17E-06 | 17.13842475 | 3.525372234 | UTR3 | KIF3B | NM_004798:c.*22210>0 |
| 12 mo | rs117699122 | 12 | 451096 | 0.758159995 | T | C | 439 | 0.0034169 | 4 | 1.10182785 | 0.226781885 | 4.858535554 | 1.18E-06 | 21.42382563 | 4.40952328 | intergenic | CCDC77;B4GALNT3 | dist=8456;dist=8843 |
| 12 mo | rs529110230 | 7 | 117535853 | | T | C | 439 | 0.0034169 | 3 | 1.126856399 | 0.232431066 | 4.848131618 | 1.25E-06 | 20.85836332 | 4.302350877 | intronic | CFTR | . |
| 12 mo | rs12114488 | 8 | 61760164 | | G | A | 439 | 0.3052392 | 266 | 8.910223585 | 1.843056308 | 4.834482563 | 1.33E-06 | 2.623079144 | 0.54257702 | intergenic | MIR4470;LINC02155 | dist=45305;dist=129675 |
| 12 mo | rs182868205 | 3 | 177994660 | | C | T | 439 | 0.0034169 | 3 | 1.020592047 | 0.21132014 | 4.829601415 | 1.37E-06 | 22.85443031 | 4.732156621 | intergenic | LINC02015;LINC01014 | dist=95436;dist=424541 |
| 12 mo | rs144138711 | 9 | 97160137 | | T | C | 439 | 0.0455581 | 40 | 3.827539823 | 0.793072242 | 4.826218371 | 1.39E-06 | 6.085471305 | 1.260919179 | ncRNA_intronic | ANKRD18CP | . |
| 12 mo | rs372194899 | 13 | 73783373 | | A | G | 439 | 0.0034169 | 3 | 1.092725742 | 0.226492304 | 4.824560146 | 1.40E-06 | 21.30121009 | 4.41516106 | intronic | KLF12 | . |
| 12 mo | rs368187808 | 13 | 73783371 | | A | C | 439 | 0.0034169 | 3 | 1.092690402 | 0.226499726 | 4.82424601 | 1.41E-06 | 21.2991251 | 4.415016369 | intronic | KLF12 | . |
| 12 mo | rs145804766 | 1 | 202153371 | | C | T | 439 | 0.0102506 | 9 | 1.845977123 | 0.382967209 | 4.820196299 | 1.43E-06 | 12.58644653 | 2.611189618 | intronic | PTPN7 | . |
| 12 mo | rs182211730 | 7 | 116363561 | | G | A | 439 | 0.0045558 | 4 | 1.236134718 | 0.256594603 | 4.817461878 | 1.45E-06 | 18.7746033 | 3.897198104 | intergenic | LOC102724434;CAV2 | dist=76834;dist=136177 |
| 12 mo | rs558784715 | 7 | 117403240 | | A | G | 439 | 0.0034169 | 3 | 1.113953973 | 0.231647992 | 4.80882206 | 1.52E-06 | 20.75917871 | 4.316894751 | intronic | ASZ1 | . |
| 12 mo | rs189709453 | 3 | 177856520 | | G | A | 439 | 0.0034169 | 3 | 0.994705176 | 0.207058897 | 4.803972151 | 1.56E-06 | 23.20099361 | 4.829543736 | ncRNA_intronic | LINC02015 | . |
| 12 mo | rs142721557 | 7 | 117578274 | | A | C | 439 | 0.0034169 | 3 | 1.1096866 | 0.231349797 | 4.796574767 | 1.61E-06 | 20.73299749 | 4.322458941 | intronic | CFTR | . |
| 12 mo | rs576047962 | 7 | 117468967 | | A | G | 439 | 0.0034169 | 3 | 1.109863163 | 0.231406391 | 4.796164697 | 1.62E-06 | 20.72615487 | 4.321401824 | intergenic | ASZ1;CFTR | dist=41474;dist=11058 |
| 12 mo | rs142215699 | 7 | 117559820 | | T | C | 439 | 0.0034169 | 3 | 1.109487618 | 0.231350082 | 4.795708764 | 1.62E-06 | 20.72922868 | 4.322453614 | ncRNA_intronic | CFTR-AS1 | . |

| | | | | | | | | | | | | | | | | | | |
|---|---|---|---|---|---|---|---|---|---|---|---|---|---|---|---|---|---|---|
| 12 mo | rs143281973 | 7 | 116367942 | | C | T | 439 | 0.0045558 | 4 | 1.258303619 | 0.262399562 | 4.795372412 | 1.62E-06 | 18.27507784 | 3.810981978 | intergenic | LOC102724434;CAV2 | dist=81215;dist=131796 |
| 12 mo | rs17127656 | 1 | 65477788 | | C | T | 439 | 0.0558087 | 49 | 4.531431604 | 0.945352558 | 4.793377416 | 1.64E-06 | 5.070465376 | 1.057806414 | intronic | LEPR | . |
| 12 mo | rs201355675 | 7 | 117585727 | | G | A | 439 | 0.0034169 | 3 | 1.108886281 | 0.231349354 | 4.7931246 | 1.64E-06 | 20.71812396 | 4.322467219 | intronic | CFTR | . |
| 12 mo | rs186767531 | 3 | 177921989 | | T | C | 439 | 0.0034169 | 3 | 0.994608422 | 0.207613329 | 4.790677095 | 1.66E-06 | 23.07499768 | 4.81664642 | intergenic | LINC02015;LINC01014 | dist=22765;dist=497212 |
| 12 mo | rs7518849 | 1 | 65483108 | | T | C | 439 | 0.0558087 | 49 | 4.51937809 | 0.943712555 | 4.788935003 | 1.68E-06 | 5.074569555 | 1.059644692 | intronic | LEPR | . |
| 12 mo | rs181132315 | 4 | 128726214 | 0.779959977 | C | T | 439 | 0.0034169 | 3 | 1.035727795 | 0.216325589 | 4.787819148 | 1.69E-06 | 22.13246791 | 4.62266164 | intergenic | LINC02615;JADE1 | dist=206818;dist=83486 |
| 12 mo | rs75082290 | 2 | 67604021 | | T | G | 439 | 0.0785877 | 69 | 4.880252144 | 1.019651405 | 4.786196655 | 1.70E-06 | 4.693953865 | 0.98072733 | intergenic | ETAA1;LINC01812 | dist=191932;dist=192033 |
| 12 mo | rs145421321 | 20 | 32274323 | | C | T | 439 | 0.0056948 | 6 | 1.344334892 | 0.281033355 | 4.783542121 | 1.72E-06 | 17.02126111 | 3.558296484 | intergenic | POFUT1;KIF3B | dist=35665;dist=3328 |
| 12 mo | rs143432612 | 20 | 32304972 | 0.798579991 | C | T | 439 | 0.0056948 | 6 | 1.349254459 | 0.282232222 | 4.780653489 | 1.75E-06 | 16.93872319 | 3.543181539 | intronic | KIF3B | . |
| 12 mo | rs140276610 | 16 | 6383734 | 0.729809999 | C | T | 439 | 0.0034169 | 4 | 1.029963075 | 0.215488262 | 4.779671375 | 1.76E-06 | 22.18065773 | 4.640624008 | intronic | RBFOX1 | . |
| 12 mo | rs559152067 | 18 | 68362375 | 0.792789996 | G | A | 439 | 0.0022779 | 3 | 0.920959359 | 0.192922126 | 4.773736308 | 1.81E-06 | 24.74436915 | 5.183438622 | intergenic | LOC643542;TMX3 | dist=462756;dist=311313 |
| 12 mo | rs147221953 | 13 | 22911583 | | G | A | 439 | 0.0034169 | 3 | 1.00672997 | 0.211147151 | 4.767906961 | 1.86E-06 | 22.58096755 | 4.736033596 | ncRNA_intronic | LINC00621 | . |
| 12 mo | rs556274646 | 11 | 439012 | | C | T | 439 | 0.0102506 | 9 | 1.757323369 | 0.369211146 | 4.759670415 | 1.94E-06 | 12.89145917 | 2.708477277 | intronic | ANO9 | . |
| 12 mo | rs190251199 | 14 | 105124240 | 0.736329973 | T | C | 439 | 0.0034169 | 3 | 0.998973364 | 0.210064697 | 4.755550928 | 1.98E-06 | 22.63850613 | 4.760438165 | intergenic | LINC02298;JAG2 | dist=24744;dist=16758 |
| 12 mo | rs188993522 | 7 | 117634677 | | T | C | 439 | 0.0034169 | 3 | 1.061817851 | 0.22328044 | 4.755534576 | 1.98E-06 | 21.29848267 | 4.478672657 | intronic | CFTR | . |
| 12 mo | rs568321148 | 2 | 29647482 | | T | G | 439 | 0.0034169 | 3 | 1.087769414 | 0.22901495 | 4.749774698 | 2.04E-06 | 20.74002026 | 4.366527169 | intronic | ALK | . |
| 12 mo | rs148815783 | 11 | 1013992 | | C | T | 439 | 0.0079727 | 8 | 1.656962032 | 0.349074609 | 4.746727463 | 2.07E-06 | 13.59803132 | 2.864717097 | exonic | MUC6 | . |
| 12 mo | rs76220567 | 2 | 12919819 | 0.745050013 | T | C | 439 | 0.0034169 | 4 | 1.089141003 | 0.22947558 | 4.746217455 | 2.07E-06 | 20.68288684 | 4.357762163 | intergenic | TRIB2;LOC100506474 | dist=177087;dist=46963 |
| 12 mo | rs35145334 | 1 | 23138629 | | A | G | 439 | 0.0170843 | 14 | 2.111856339 | 0.445194762 | 4.743668432 | 2.10E-06 | 10.65526559 | 2.246207918 | intronic | LUZP1 | . |
| 12 mo | rs11579567 | 1 | 65491458 | | C | A | 439 | 0.0558087 | 49 | 4.468235403 | 0.942530351 | 4.740680657 | 2.13E-06 | 5.029737931 | 1.060973792 | intronic | LEPR | . |
| 12 mo | rs114067899 | 1 | 84725361 | | G | A | 439 | 0.0079727 | 7 | 1.628650291 | 0.343835958 | 4.736707304 | 2.17E-06 | 13.77606734 | 2.908363649 | intergenic | SSX2IP;LPAR3 | dist=34893;dist=86241 |
| 12 mo | rs56821264 | 6 | 148382765 | | C | T | 439 | 0.0068337 | 6 | 1.520993492 | 0.321110203 | 4.736671324 | 2.17E-06 | 14.75092126 | 3.114195656 | intronic | SASH1 | . |
| 12 mo | rs536781978 | 2 | 29510815 | | A | G | 439 | 0.0034169 | 3 | 1.090050785 | 0.230277801 | 4.733633818 | 2.21E-06 | 20.55618824 | 4.342580992 | intronic | ALK | . |
| 12 mo | rs147267707 | 4 | 43024541 | | G | A | 439 | 0.0045558 | 4 | 1.241836175 | 0.262592348 | 4.729140754 | 2.25E-06 | 18.00943854 | 3.808184082 | intronic | GRXCR1 | . |
| 12 mo | rs77618729 | 4 | 43048951 | | T | C | 439 | 0.0045558 | 4 | 1.241458345 | 0.26252633 | 4.728890791 | 2.26E-06 | 18.01301525 | 3.809141731 | intergenic | GRXCR1;LINC02383 | dist=18293;dist=408583 |
| 12 mo | rs547186621 | 20 | 6126094 | 0.76117003 | A | G | 439 | 0.0034169 | 4 | 1.03230976 | 0.218862053 | 4.716714238 | 2.40E-06 | 21.55108289 | 4.569088099 | intergenic | FERMT1;CASC20 | dist=3064;dist=300638 |
| 12 mo | rs200198574 | 20 | 32358851 | | G | A | 439 | 0.0056948 | 6 | 1.343845487 | 0.285141824 | 4.712902058 | 2.44E-06 | 16.52827354 | 3.50702674 | intronic | ASXL1 | . |
| 12 mo | rs532464521 | 4 | 43569022 | | T | C | 439 | 0.0045558 | 4 | 1.233962678 | 0.261903215 | 4.711521696 | 2.46E-06 | 17.98955275 | 3.818204376 | intergenic | LINC02383;LINC02475 | dist=76479;dist=447839 |
| 12 mo | rs79252854 | 3 | 157805368 | | C | T | 439 | 0.0045558 | 5 | 1.264269707 | 0.26842653 | 4.709928283 | 2.48E-06 | 17.54643357 | 3.72541417 | intergenic | SLC66A1L;SHOX2 | dist=204274;dist=290537 |
| 12 mo | rs79935606 | 2 | 176473907 | | T | C | 439 | 0.0056948 | 5 | 1.354071441 | 0.2878717 | 4.703732401 | 2.55E-06 | 16.33968329 | 3.473769742 | intergenic | MTX2;MIR1246 | dist=135882;dist=127073 |
| 12 mo | rs556435089 | 4 | 43549016 | | G | A | 439 | 0.0045558 | 4 | 1.230811842 | 0.261936344 | 4.698896777 | 2.62E-06 | 17.93907904 | 3.817721456 | intergenic | LINC02383;LINC02475 | dist=56473;dist=467845 |
| 12 mo | rs62192733 | 20 | 1947511 | 0.746280015 | A | G | 439 | 0.0068337 | 7 | 1.395720916 | 0.297737289 | 4.687759871 | 2.76E-06 | 15.74461797 | 3.358665632 | ncRNA_exonic | PDYN-AS1 | . |
| 12 mo | rs7534177 | 1 | 65500037 | | A | G | 439 | 0.0569476 | 49 | 4.416632642 | 0.943519417 | 4.681020403 | 2.85E-06 | 4.961233778 | 1.059861601 | intronic | LEPR | . |
| 12 mo | rs139390630 | 1 | 84400513 | 0.718169987 | G | A | 439 | 0.0113895 | 13 | 1.872384943 | 0.40008221 | 4.680000507 | 2.87E-06 | 11.69759713 | 2.499486296 | intronic | DNASE2B | . |
| 12 mo | rs149706477 | 3 | 66162553 | | A | G | 439 | 0.0034169 | 3 | 1.043481199 | 0.223002578 | 4.679233802 | 2.88E-06 | 20.98286869 | 4.484253102 | intronic | SLC25A26 | . |
| 12 mo | rs189094663 | 4 | 11621826 | 0.791639984 | G | A | 439 | 0.0022779 | 3 | 0.91574426 | 0.195762636 | 4.677829629 | 2.90E-06 | 23.895416 | 5.108227082 | intergenic | HS3ST1;LINC02360 | dist=192932;dist=119125 |
| 12 mo | rs181182636 | 7 | 116014504 | | A | G | 439 | 0.0034169 | 3 | 0.996739233 | 0.213108408 | 4.677146454 | 2.91E-06 | 21.94726388 | 4.692447434 | intronic | TFEC | . |
| 12 mo | rs76098744 | 1 | 111806750 | | C | T | 439 | 0.0159453 | 14 | 2.284999945 | 0.488674498 | 4.675914038 | 2.93E-06 | 9.568565696 | 2.046351926 | intronic | KCND3 | . |
| 12 mo | rs4896997 | 6 | 148362788 | | C | T | 439 | 0.0068337 | 6 | 1.509673329 | 0.323427425 | 4.667734442 | 3.05E-06 | 14.43209229 | 3.091883755 | intronic | SASH1 | . |
| 12 mo | rs17078283 | 6 | 148375784 | | C | T | 439 | 0.0068337 | 6 | 1.509561069 | 0.323435684 | 4.667268152 | 3.05E-06 | 14.43028205 | 3.091804795 | intronic | SASH1 | . |
| 12 mo | rs73202425 | 7 | 109515659 | | T | C | 439 | 0.0125285 | 12 | 2.024115362 | 0.433684884 | 4.667249048 | 3.05E-06 | 10.76184396 | 2.305821662 | intergenic | C7orf66;EIF3IP1 | dist=631072;dist=443568 |
| 12 mo | rs4131286 | 6 | 148367827 | | G | T | 439 | 0.0068337 | 6 | 1.509448809 | 0.323443998 | 4.666801106 | 3.06E-06 | 14.42846716 | 3.091725323 | intronic | SASH1 | . |
| 12 mo | rs74683551 | 1 | 111811796 | | G | A | 439 | 0.0159453 | 14 | 2.279782265 | 0.488661396 | 4.665361915 | 3.08E-06 | 9.547228317 | 2.046406793 | intronic | KCND3 | . |
| 12 mo | rs139055031 | 3 | 158869060 | 0.75673002 | T | C | 439 | 0.0056948 | 6 | 1.332817863 | 0.286079347 | 4.658909756 | 3.18E-06 | 16.28537606 | 3.4955337 | intergenic | MFSD1;IQCJ | dist=39341;dist=200192 |
| 12 mo | rs138873576 | 7 | 116385305 | | G | T | 439 | 0.0045558 | 4 | 1.230673437 | 0.264318756 | 4.656020093 | 3.22E-06 | 17.61517106 | 3.78331079 | intergenic | LOC102724434;CAV2 | dist=98578;dist=114433 |
| 12 mo | rs148483098 | 3 | 157664262 | | C | A | 439 | 0.0056948 | 5 | 1.375665606 | 0.29567339 | 4.652652729 | 3.28E-06 | 15.73578443 | 3.382110237 | intergenic | SLC66A1L;SHOX2 | dist=63168;dist=431643 |
| 12 mo | rs187281112 | 15 | 32821186 | | C | T | 439 | 0.0056948 | 5 | 1.468174325 | 0.315662894 | 4.651083021 | 3.30E-06 | 14.73433563 | 3.167936491 | intronic | FMN1 | . |
| 12 mo | rs2902021 | 2 | 67598553 | | C | T | 439 | 0.0797267 | 71 | 4.743601835 | 1.019942721 | 4.650851208 | 3.31E-06 | 4.559914114 | 0.980447215 | intergenic | ETAA1;LINC01812 | dist=186464;dist=197501 |
| 12 mo | rs189912648 | 5 | 135429749 | | C | T | 439 | 0.0034169 | 4 | 1.088415104 | 0.234100744 | 4.649344921 | 3.33E-06 | 19.8604449 | 4.271665199 | intergenic | MACROH2A1;DCANP1 | dist=29862;dist=14465 |
| 12 mo | rs557276277 | 4 | 19012075 | 0.731580019 | T | C | 439 | 0.0056948 | 5 | 1.135923633 | 0.244416532 | 4.647491003 | 3.36E-06 | 19.01463443 | 4.091376275 | intergenic | LCORL;SLIT2 | dist=990200;dist=1239830 |
| 12 mo | rs147869916 | 1 | 227119951 | | A | G | 439 | 0.0056948 | 5 | 1.227452207 | 0.264112954 | 4.647451732 | 3.36E-06 | 17.59645507 | 3.78625881 | intronic | CDC42BPA | . |
| 12 mo | rs367732718 | 19 | 35427102 | 0.764100015 | G | A | 439 | 0.0022779 | 3 | 0.903832269 | 0.194554137 | 4.645659467 | 3.39E-06 | 23.87849231 | 5.139957519 | intergenic | LINC01531;FFAR2 | dist=10262;dist=21155 |
| 12 mo | rs62515436 | 8 | 56228644 | | G | T | 439 | 0.0170843 | 15 | 2.32339754 | 0.500488447 | 4.642260083 | 3.45E-06 | 9.275459023 | 1.998048118 | intergenic | CHCHD7;SDR16C5 | dist=9835;dist=71004 |
| 12 mo | rs189234695 | 3 | 147940371 | 0.726230025 | T | C | 439 | 0.0034169 | 3 | 0.898361941 | 0.193595051 | 4.640417897 | 3.48E-06 | 23.96971341 | 5.165421292 | intergenic | LOC440982;LINC02032 | dist=430461;dist=137788 |
| 12 mo | rs186893139 | 7 | 116456916 | | C | T | 439 | 0.0045558 | 4 | 1.155381378 | 0.249215977 | 4.636064639 | 3.55E-06 | 18.60259803 | 4.012583835 | intergenic | LOC102724434;CAV2 | dist=170189;dist=42822 |
| 12 mo | rs73384619 | 6 | 20457948 | | C | T | 439 | 0.022779 | 21 | 2.554663139 | 0.55175035 | 4.630106969 | 3.65E-06 | 8.391670204 | 1.812413894 | intronic | E2F3 | . |
| 12 mo | rs114280794 | 3 | 133123359 | | G | A | 439 | 0.0159453 | 14 | 2.436845188 | 0.527337114 | 4.621038659 | 3.82E-06 | 8.762968775 | 1.896320161 | intronic | TMEM108 | . |
| 12 mo | rs112148840 | 17 | 68237633 | 0.760460019 | C | T | 439 | 0.0022779 | 3 | 0.88865431 | 0.192328123 | 4.620511518 | 3.83E-06 | 24.02410753 | 5.199447601 | intronic | AMZ2 | . |
| 12 mo | rs147455971 | 4 | 140010558 | 0.772499979 | T | C | 439 | 0.0079727 | 5 | 1.238787979 | 0.268245339 | 4.618115576 | 3.87E-06 | 17.21601424 | 3.727930571 | intronic | MAML3 | . |
| 12 mo | rs144788248 | 3 | 19545253 | | T | C | 439 | 0.0113895 | 10 | 1.790726259 | 0.387778578 | 4.617909184 | 3.88E-06 | 11.90862374 | 2.578791237 | intergenic | KCNH8;EFHB | dist=9611;dist=334221 |
| 12 mo | rs111512950 | 5 | 154300867 | | C | T | 439 | 0.0091116 | 8 | 1.5632807 | 0.33944505 | 4.605401379 | 4.12E-06 | 13.56744305 | 2.945984928 | intronic | GALNT10 | . |
| 12 mo | rs149616342 | 12 | 98785035 | | C | T | 439 | 0.0113895 | 10 | 1.875050964 | 0.407388894 | 4.602606968 | 4.17E-06 | 11.2978214 | 2.454656997 | intronic | ANKS1B | . |
| 12 mo | rs17543620 | 4 | 169013574 | | T | C | 439 | 0.0501139 | 44 | 3.899464938 | 0.847586222 | 4.600670513 | 4.21E-06 | 5.427967555 | 1.179820972 | intergenic | CBR4;SH3RF1 | dist=3319;dist=80685 |
| 12 mo | rs190354334 | 7 | 116435792 | | G | A | 439 | 0.0045558 | 5 | 1.218759267 | 0.265120744 | 4.596997007 | 4.29E-06 | 17.33925809 | 3.7718663 | intergenic | LOC102724434;CAV2 | dist=149065;dist=63946 |
| 12 mo | rs17615362 | 4 | 169012936 | | G | A | 439 | 0.0501139 | 44 | 3.897763553 | 0.84804389 | 4.596181398 | 4.30E-06 | 5.419744721 | 1.179184251 | intergenic | CBR4;SH3RF1 | dist=2681;dist=81323 |
| 12 mo | rs117591241 | 3 | 190866890 | 0.774309993 | A | G | 439 | 0.0034169 | 4 | 1.059847527 | 0.230770266 | 4.592652012 | 4.38E-06 | 19.90140277 | 4.333313893 | intergenic | GMNC;SNAR-I | dist=4196;dist=11040 |
| 12 mo | rs61918041 | 12 | 6572702 | | C | T | 439 | 0.023918 | 21 | 2.644372798 | 0.575917252 | 4.591584621 | 4.40E-06 | 7.972646426 | 1.736360556 | intronic | CHD4 | . |
| 12 mo | rs113244573 | 12 | 6575219 | | G | A | 439 | 0.023918 | 21 | 2.639855196 | 0.575486058 | 4.587174889 | 4.49E-06 | 7.970957457 | 1.737661556 | intronic | CHD4 | . |
| 12 mo | rs62515405 | 8 | 56143419 | | G | A | 439 | 0.023918 | 21 | 2.59481628 | 0.56680103 | 4.578002058 | 4.69E-06 | 8.076912036 | 1.76428755 | intergenic | MOS;PLAG1 | dist=29437;dist=17490 |
| 12 mo | rs116897913 | 17 | 16935848 | | C | T | 439 | 0.0034169 | 3 | 1.038882394 | 0.22712082 | 4.574139844 | 4.78E-06 | 20.13967647 | 4.402942883 | intergenic | TBC1D27P;TNFRSF13B | dist=2672;dist=3233 |

| | | | | | | | | | | | | | | | | | | |
|---|---|---|---|---|---|---|---|---|---|---|---|---|---|---|---|---|---|---|
| 12 mo | rs143686474 | 7 | 16252374 | | A | C | 439 | 0.0125285 | 11 | 1.923890516 | 0.421029259 | 4.569493631 | 4.89E-06 | 10.85314984 | 2.375131845 | ncRNA_intronic | CRPPA-AS1 | . |
| 12 mo | rs143669489 | 3 | 159675096 | 0.739700019 | G | A | 439 | 0.0034169 | 3 | 0.861115457 | 0.188536714 | 4.567362191 | 4.94E-06 | 24.22531986 | 5.304006744 | intronic | IQCJ-SCHIP1;SCHIP1 | . |

### Supplementary Table S1. GWAS Results SNPs Indianapolis-1 Discovery Cohort
### 3 month quantitative trait (QT)

**Headers**

QT: quantitiative trait; rsid: reference SNP cluster ID, chr: chromosome; pos_38, position of SNP on GRCh38 reference panel; Imputation_Rsq (if <0.8); REF and ALT, reference allele and alternate allele; n.obs: number of observations;
caf: common (major) allele frequency; MAC, minor allele count; Score: p-values from Score test; Score.SE; Score.Stat; Score.pval; EST; EST.SE; Func.refGene: SNP location with respect to nearest gene;
Gene.refGene; nearest gene upstream and downstream; GeneDetail.refGene: distance to nearest gene upstream and downstream; GeneDetail.refGene: distance to nearest gene upstream and downstream

**Notes**

Imputation Rsq listed only if less than 0.8

| QT | rsid | chr | pos_38 | Imputation_Rsq | REF | ALT | n.obs | caf | MAC | Score | Score.SE | Score.Stat | Score.pval | Est | Est.SE | Func.refGene | Gene.refGene | GeneDetail.refGene |
|---|---|---|---|---|---|---|---|---|---|---|---|---|---|---|---|---|---|---|
| 3 month | rs113063005 | 4 | 23507444 | 0.763409972 | T | C | 421 | 0.005938 | 6 | 2.130262689 | 0.316937024 | 6.721406875 | 1.80E-11 | 21.2073894 | 3.155200957 | intergenic | GBA3;PPARGC1A | dist=687872;dist=284577 |
| 3 month | rs142106992 | 6 | 141948748 | | C | A | 421 | 0.003563 | 4 | 1.689946394 | 0.272044449 | 6.212023076 | 5.23E-10 | 22.83458862 | 3.675869896 | intergenic | MIR4465;NMBR | dist=1264865;dist=125736 |
| 3 month | rs181217257 | 2 | 239067423 | | C | T | 421 | 0.004751 | 4 | 1.788144205 | 0.287937999 | 6.210170973 | 5.29E-10 | 21.56773677 | 3.472969885 | intronic | HDAC4 | . |
| 3 month | rs188076929 | 2 | 239072023 | | T | C | 421 | 0.004751 | 4 | 1.865485168 | 0.305579814 | 6.104739516 | 1.03E-09 | 19.97756143 | 3.272467463 | intronic | HDAC4 | . |
| 3 month | rs117998251 | 10 | 13455976 | | C | T | 421 | 0.003563 | 3 | 1.610926128 | 0.264914355 | 6.080931816 | 1.19E-09 | 22.95433112 | 3.774804885 | intronic | BEND7 | . |
| 3 month | rs184458518 | 10 | 13429195 | | T | G | 421 | 0.003563 | 3 | 1.603233248 | 0.265164814 | 6.046176429 | 1.48E-09 | 22.80157891 | 3.771239424 | intergenic | SEPHS1;BEND7 | dist=80897;dist=9286 |
| 3 month | rs184425183 | 10 | 13415520 | | A | G | 421 | 0.003563 | 3 | 1.590271029 | 0.264865828 | 6.004062666 | 1.92E-09 | 22.66831744 | 3.775496477 | intergenic | SEPHS1;BEND7 | dist=67222;dist=22961 |
| 3 month | rs148153037 | 6 | 167087898 | | G | A | 421 | 0.008314 | 8 | 2.445317826 | 0.413004518 | 5.920801636 | 3.20E-09 | 14.33592462 | 2.421281019 | intergenic | CEP43;CCR6 | dist=35180;dist=23909 |
| 3 month | rs111285015 | 19 | 22940396 | 0.738849998 | G | A | 421 | 0.003563 | 3 | 1.280187265 | 0.216410877 | 5.915540303 | 3.31E-09 | 27.33476425 | 4.62083983 | intergenic | ZNF723;ZNF728 | dist=81729;dist=34487 |
| 3 month | rs111928960 | 1 | 103168079 | | G | A | 421 | 0.028504 | 22 | 4.045287237 | 0.68574164 | 5.899141891 | 3.65E-09 | 8.602572083 | 1.45827516 | intergenic | COL11A1;LOC101928436 | dist=59557;dist=325967 |
| 3 month | rs187520610 | 2 | 53132903 | | G | A | 421 | 0.004751 | 4 | 1.843352227 | 0.313121747 | 5.887014379 | 3.93E-09 | 18.80103964 | 3.193645952 | intergenic | MIR4431;ASB3 | dist=430288;dist=537076 |
| 3 month | rs545428520 | 5 | 168394261 | | T | C | 421 | 0.003563 | 4 | 1.606350972 | 0.272919814 | 5.885798274 | 3.96E-09 | 21.56603503 | 3.664079879 | intronic | WWC1 | . |
| 3 month | rs116672066 | 1 | 103007360 | | G | A | 421 | 0.027316 | 23 | 4.276024986 | 0.728927557 | 5.866186486 | 4.46E-09 | 8.047694764 | 1.37187844 | intronic | COL11A1 | . |
| 3 month | rs192134381 | 21 | 22078395 | | T | C | 421 | 0.003563 | 3 | 1.565034889 | 0.268006444 | 5.83954202 | 5.23E-09 | 21.78881202 | 3.731253572 | ncRNA_intronic | LINC01687 | . |
| 3 month | rs150586237 | 6 | 24491120 | | C | T | 421 | 0.003563 | 3 | 1.542166452 | 0.264483938 | 5.830851057 | 5.51E-09 | 22.04614425 | 3.780947933 | intergenic | GPLD1;ALDH5A1 | dist=1542;dist=3849 |
| 3 month | rs528404963 | 5 | 168425020 | | T | C | 421 | 0.003563 | 4 | 1.58410842 | 0.273744969 | 5.786803767 | 7.17E-09 | 21.13939767 | 3.653035168 | intronic | WWC1 | . |
| 3 month | rs76526501 | 7 | 18391487 | | G | A | 421 | 0.007126 | 6 | 2.233180137 | 0.387925975 | 5.75671721 | 8.58E-09 | 14.83973124 | 2.577811398 | intronic | HDAC9 | . |
| 3 month | rs74455595 | 7 | 18392161 | | A | G | 421 | 0.007126 | 6 | 2.233180137 | 0.387925975 | 5.75671721 | 8.58E-09 | 14.83973124 | 2.577811398 | intronic | HDAC9 | . |
| 3 month | rs75090694 | 7 | 18407813 | | A | G | 421 | 0.008314 | 7 | 2.37298061 | 0.41420374 | 5.729017827 | 1.01E-08 | 13.83140053 | 2.414270813 | intronic | HDAC9 | . |
| 3 month | rs575473987 | 8 | 5719972 | | C | T | 421 | 0.003563 | 3 | 1.471553256 | 0.257316966 | 5.718834929 | 1.07E-08 | 22.22486534 | 3.886257536 | intergenic | CSMD1;LOC100287015 | dist=725058;dist=683583 |
| 3 month | rs545690161 | 9 | 90268417 | 0.734099984 | G | A | 421 | 0.005938 | 6 | 1.900812423 | 0.332979511 | 5.708496652 | 1.14E-08 | 17.14368742 | 3.003187786 | intergenic | MIR4290HG;LINC01508 | dist=226918;dist=32479 |
| 3 month | rs77141817 | 4 | 37052137 | | T | C | 421 | 0.003563 | 3 | 1.553761456 | 0.274563203 | 5.659030198 | 1.52E-08 | 20.61102923 | 3.642148656 | intergenic | LINC02616;MIR4801 | dist=31431;dist=189773 |
| 3 month | rs190822761 | 4 | 37097734 | | G | T | 421 | 0.003563 | 3 | 1.550650175 | 0.274463961 | 5.649740562 | 1.61E-08 | 20.58463536 | 3.643465595 | intergenic | LINC02616;MIR4801 | dist=77028;dist=144176 |
| 3 month | rs10279777 | 7 | 18401966 | | G | A | 421 | 0.009501 | 8 | 2.476234721 | 0.438670193 | 5.644866606 | 1.65E-08 | 12.86813351 | 2.279616935 | intronic | HDAC9 | . |
| 3 month | rs10486295 | 7 | 18407184 | | G | A | 421 | 0.009501 | 8 | 2.488096085 | 0.441539202 | 5.635051363 | 1.75E-08 | 12.76229003 | 2.264804562 | intronic | HDAC9 | . |
| 3 month | rs147559909 | 2 | 236142879 | | T | C | 421 | 0.005938 | 5 | 1.858263542 | 0.329805301 | 5.634425932 | 1.76E-08 | 17.08409752 | 3.032091952 | intergenic | AGAP1;GBX2 | dist=11086;dist=22356 |
| 3 month | rs17169602 | 7 | 18407118 | | G | A | 421 | 0.009501 | 8 | 2.49198241 | 0.442489656 | 5.631730316 | 1.78E-08 | 12.72737168 | 2.259939835 | intronic | HDAC9 | . |
| 3 month | rs139598422 | 13 | 23312875 | 0.783299983 | A | G | 421 | 0.003563 | 4 | 1.544940466 | 0.274599144 | 5.626166351 | 1.84E-08 | 20.48865215 | 3.641671944 | intronic | SGCG | . |
| 3 month | rs74704551 | 14 | 29692681 | | C | T | 421 | 0.003563 | 3 | 1.302308212 | 0.231983741 | 5.613790894 | 1.98E-08 | 24.19907048 | 4.31064693 | intronic | PRKD1 | . |
| 3 month | rs77300464 | 7 | 18369138 | | A | G | 421 | 0.007126 | 6 | 2.141507953 | 0.382123995 | 5.604222657 | 2.09E-08 | 14.66597943 | 2.616951597 | intronic | HDAC9 | . |
| 3 month | rs75606013 | 7 | 18374990 | | G | A | 421 | 0.007126 | 6 | 2.140929626 | 0.382127995 | 5.602650559 | 2.11E-08 | 14.66171186 | 2.616924205 | intronic | HDAC9 | . |
| 3 month | rs541653703 | 11 | 18680239 | | G | A | 421 | 0.004751 | 4 | 1.710453669 | 0.305582922 | 5.597347058 | 2.18E-08 | 18.31694986 | 3.272434185 | intergenic | SPTY2D1;TMEM86A | dist=45897;dist=18540 |
| 3 month | rs117913371 | 10 | 101158729 | | G | A | 421 | 0.024941 | 21 | 3.968654489 | 0.710300363 | 5.587290525 | 2.31E-08 | 7.866095548 | 1.407855116 | intergenic | TLX1NB;LINC01514 | dist=17463;dist=17593 |
| 3 month | rs191053292 | 12 | 63051500 | 0.719120026 | T | C | 421 | 0.003563 | 3 | 1.354341299 | 0.243048963 | 5.572298207 | 2.51E-08 | 22.9266488 | 4.114397317 | intergenic | PPM1H;AVPR1A | dist=116350;dist=91259 |
| 3 month | rs112351653 | 1 | 102754804 | | T | C | 421 | 0.028504 | 24 | 4.178844629 | 0.750261996 | 5.569847131 | 2.55E-08 | 7.423869471 | 1.332867724 | intergenic | OLFM3;COL11A1 | dist=757570;dist=121663 |
| 3 month | rs114413507 | 1 | 102953612 | | T | C | 421 | 0.028504 | 24 | 4.177369718 | 0.750215949 | 5.568223023 | 2.57E-08 | 7.42216029 | 1.332949535 | intronic | COL11A1 | . |
| 3 month | rs138414342 | 11 | 18657851 | | G | A | 421 | 0.004751 | 5 | 1.685610798 | 0.304678389 | 5.532426519 | 3.16E-08 | 18.15825054 | 3.282149429 | intergenic | SPTY2D1;TMEM86A | dist=23509;dist=40928 |
| 3 month | rs16823323 | 3 | 153939413 | | G | A | 421 | 0.016627 | 14 | 3.214602568 | 0.582174532 | 5.521716236 | 3.36E-08 | 9.484640653 | 1.717697949 | intergenic | LINC02006;ARHGEF26-AS | dist=176887;dist=84988 |
| 3 month | rs183737367 | 9 | 90567765 | | T | C | 421 | 0.003563 | 3 | 1.295690386 | 0.235587115 | 5.499835526 | 3.80E-08 | 23.34523057 | 4.244714311 | ncRNA_intronic | LINC01501 | . |
| 3 month | rs560206697 | 19 | 20546292 | 0.700380027 | C | T | 421 | 0.002375 | 3 | 1.205675879 | 0.219610964 | 5.490053226 | 4.02E-08 | 24.99899431 | 4.553506728 | intronic | ZNF737 | . |
| 3 month | rs79486609 | 21 | 15872687 | | G | A | 421 | 0.003563 | 3 | 1.447442576 | 0.264672828 | 5.468799292 | 4.53E-08 | 20.66248857 | 3.77824957 | intronic | USP25 | . |
| 3 month | rs151115079 | 11 | 18634194 | 0.792460024 | T | C | 421 | 0.004751 | 5 | 1.669234911 | 0.305995633 | 5.455093905 | 4.89E-08 | 17.8273587 | 3.268020498 | intronic | SPTY2D1 | . |
| 3 month | rs185510569 | 2 | 222950143 | | G | A | 421 | 0.003563 | 3 | 1.507358682 | 0.277399453 | 5.433892049 | 5.51E-08 | 19.58869056 | 3.604909775 | intergenic | ACSL3;KCNE4 | dist=5505;dist=102047 |
| 3 month | rs187213609 | 9 | 90653183 | | C | T | 421 | 0.003563 | 3 | 1.269834136 | 0.233965931 | 5.427431802 | 5.72E-08 | 23.19753039 | 4.274126556 | intergenic | DIRAS2;SYK | dist=10359;dist=148417 |
| 3 month | rs117280553 | 21 | 15834844 | | T | C | 421 | 0.003563 | 3 | 1.465128514 | 0.27006967 | 5.425002059 | 5.80E-08 | 20.08741695 | 3.702748263 | intronic | USP25 | . |
| 3 month | rs183816745 | 5 | 92336071 | | A | G | 421 | 0.005938 | 5 | 1.692149037 | 0.312309144 | 5.418186024 | 6.02E-08 | 17.34879088 | 3.201955563 | intergenic | ARRDC3-AS1;NR2F1-AS1 | dist=915356;dist=1113174 |
| 3 month | rs180828621 | 10 | 122773893 | | G | A | 421 | 0.007126 | 6 | 1.931459198 | 0.357196912 | 5.407267342 | 6.40E-08 | 15.13805735 | 2.799576273 | ncRNA_intronic | DMBT1L1 | . |
| 3 month | rs142684595 | 3 | 55285273 | 0.77293998 | T | C | 421 | 0.005938 | 5 | 1.730096674 | 0.321166729 | 5.386911268 | 7.17E-08 | 16.77294307 | 3.113647549 | intergenic | LINC02017;WNT5A | dist=96059;dist=180442 |
| 3 month | rs184785969 | 21 | 15799112 | | C | A | 421 | 0.003563 | 3 | 1.446338922 | 0.269771603 | 5.361346058 | 8.26E-08 | 19.87364864 | 3.706839369 | intronic | USP25 | . |
| 3 month | rs77180278 | 1 | 102496326 | | T | C | 421 | 0.028504 | 24 | 3.885721783 | 0.727780259 | 5.339141503 | 9.34E-08 | 7.336199959 | 1.374041118 | intergenic | OLFM3;COL11A1 | dist=499092;dist=380141 |
| 3 month | rs189709453 | 3 | 177856520 | | G | A | 421 | 0.003563 | 3 | 1.314463975 | 0.246356303 | 5.335621454 | 9.52E-08 | 21.65814876 | 4.05916142 | ncRNA_intronic | LINC02015 | . |
| 3 month | rs111846247 | 5 | 95775939 | | T | C | 421 | 0.003563 | 3 | 1.3787786 | 0.259425442 | 5.314739335 | 1.07E-07 | 20.48657718 | 3.854672052 | intronic | RHOBTB3 | . |
| 3 month | rs145875128 | 4 | 32071514 | 0.719099998 | G | A | 421 | 0.003563 | 3 | 1.153690598 | 0.217089638 | 5.314351295 | 1.07E-07 | 24.47998599 | 4.606392132 | ncRNA_intronic | LINC02506 | . |
| 3 month | rs75848314 | 5 | 95762636 | | T | C | 421 | 0.003563 | 3 | 1.401525911 | 0.264060948 | 5.307584942 | 1.11E-07 | 20.0998481 | 3.787004507 | intronic | RHOBTB3 | . |
| 3 month | rs190193113 | 2 | 52858640 | | G | A | 421 | 0.003563 | 3 | 1.409901074 | 0.265819171 | 5.303985667 | 1.13E-07 | 19.95336019 | 3.761955902 | intergenic | MIR4431;ASB3 | dist=156025;dist=811339 |
| 3 month | rs183378658 | 2 | 52336233 | | C | T | 421 | 0.003563 | 3 | 1.421941101 | 0.268359175 | 5.298649105 | 1.17E-07 | 19.74461693 | 3.726349214 | ncRNA_intronic | LOC730100 | . |
| 3 month | rs143371352 | 13 | 46809699 | | C | T | 421 | 0.003563 | 3 | 1.364496992 | 0.257779154 | 5.29327904 | 1.20E-07 | 20.53416252 | 3.879289636 | intergenic | ESD;HTR2A | dist=12538;dist=21847 |
| 3 month | rs75186966 | 13 | 46821623 | | A | C | 421 | 0.003563 | 3 | 1.377358459 | 0.260942954 | 5.278389159 | 1.30E-07 | 20.22813447 | 3.832255231 | intergenic | ESD;HTR2A | dist=24462;dist=9923 |
| 3 month | rs532416695 | 2 | 176622062 | | G | A | 421 | 0.003563 | 3 | 1.400161077 | 0.265436681 | 5.274934396 | 1.33E-07 | 19.87266561 | 3.767376828 | intergenic | MIR1246;LINC01116 | dist=21010;dist=7519 |
| 3 month | rs145080832 | 2 | 48246004 | | G | A | 421 | 0.004751 | 4 | 1.617719031 | 0.30702207 | 5.269065159 | 1.37E-07 | 17.16184772 | 3.257095367 | intergenic | FBXO11;FOXN2 | dist=339506;dist=68295 |
| 3 month | rs182868205 | 3 | 177994660 | | C | T | 421 | 0.003563 | 3 | 1.323927501 | 0.251356167 | 5.267137522 | 1.39E-07 | 20.95487681 | 3.978418395 | intergenic | LINC02015;LINC01014 | dist=95436;dist=424541 |
| 3 month | rs143538552 | 18 | 31470299 | | A | G | 421 | 0.003563 | 3 | 1.44755014 | 0.274967228 | 5.264446053 | 1.41E-07 | 19.14572179 | 3.636797033 | intronic | DSG3 | . |

| | | | | | | | | | | | | | | | | | | |
|---|---|---|---|---|---|---|---|---|---|---|---|---|---|---|---|---|---|---|
| 3 month | rs7822082 | 8 | 54777660 | | T | C | 421 | 0.330166 | 277 | 11.70263936 | 2.228199285 | 5.252061357 | 1.50E-07 | 2.357087802 | 0.448792891 | intronic | RP1 | . |
| 3 month | rs72986533 | 6 | 142290121 | 0.786499977 | T | C | 421 | 0.008314 | 8 | 2.07386378 | 0.395157536 | 5.248194939 | 1.54E-07 | 13.28127256 | 2.530636288 | intergenic | VTA1;ADGRG6 | dist=65437;dist=11798 |
| 3 month | rs373746073 | 18 | 31478421 | | C | A | 421 | 0.003563 | 3 | 1.441999195 | 0.274877768 | 5.245965155 | 1.55E-07 | 19.08471968 | 3.637980642 | UTR3 | DSG3 | NM_001944:c.*21610>0 |
| 3 month | rs181193202 | 2 | 52300216 | | T | C | 421 | 0.003563 | 3 | 1.397666205 | 0.26682604 | 5.238117712 | 1.62E-07 | 19.63120884 | 3.747760153 | ncRNA_intronic | LOC730100 | . |
| 3 month | rs186767531 | 3 | 177921989 | | T | C | 421 | 0.003563 | 3 | 1.293967205 | 0.247047316 | 5.237730262 | 1.63E-07 | 21.20132426 | 4.047807581 | intergenic | LINC02015;LINC01014 | dist=22765;dist=497212 |
| 3 month | rs140797780 | 8 | 22230279 | | C | T | 421 | 0.003563 | 3 | 1.415632732 | 0.270894217 | 5.225776871 | 1.73E-07 | 19.29083956 | 3.691477848 | intronic | PHYHIP | . |
| 3 month | rs137880949 | 12 | 62912517 | 0.737010002 | T | C | 421 | 0.004751 | 3 | 1.263235635 | 0.24178583 | 5.224605748 | 1.75E-07 | 21.60840343 | 4.135891677 | intronic | PPM1H | . |
| 3 month | rs148532212 | 6 | 165086368 | | T | C | 421 | 0.003563 | 3 | 1.406500453 | 0.26958124 | 5.217352852 | 1.81E-07 | 19.35354569 | 3.709456929 | intergenic | MEAT6;C6orf118 | dist=264305;dist=193296 |
| 3 month | rs141754456 | 12 | 19998198 | 0.798650026 | T | C | 421 | 0.007126 | 7 | 2.240311418 | 0.429474084 | 5.21640654 | 1.82E-07 | 12.14603335 | 2.328429207 | intergenic | AEBP2;LINC02398 | dist=475971;dist=16487 |
| 3 month | rs117498042 | 6 | 165100792 | | C | T | 421 | 0.003563 | 3 | 1.405430664 | 0.269604623 | 5.212932361 | 1.86E-07 | 19.33547097 | 3.70913521 | intergenic | MEAT6;C6orf118 | dist=278729;dist=178872 |
| 3 month | rs184487573 | 6 | 167099983 | | A | G | 421 | 0.007126 | 7 | 2.008477086 | 0.385701261 | 5.207338654 | 1.92E-07 | 13.50096351 | 2.592680141 | intergenic | CEP43;CCR6 | dist=47265;dist=11824 |
| 3 month | rs149298750 | 4 | 126237546 | | A | C | 421 | 0.003563 | 3 | 1.219140188 | 0.234426329 | 5.200525875 | 1.99E-07 | 22.18405204 | 4.265732461 | intergenic | MIR2054;INTU | dist=730239;dist=1395411 |
| 3 month | rs140277951 | 10 | 80599344 | | G | A | 421 | 0.008314 | 6 | 1.967344821 | 0.378309358 | 5.200359916 | 1.99E-07 | 13.74631584 | 2.643339318 | intronic | SH2D4B | . |
| 3 month | rs558553658 | 2 | 15542600 | 0.761030018 | C | T | 421 | 0.004751 | 3 | 1.579685756 | 0.303851567 | 5.198873167 | 2.01E-07 | 17.10991069 | 3.291080613 | intronic | NBAS | . |
| 3 month | rs112679237 | 4 | 138219706 | 0.778339982 | T | C | 421 | 0.017815 | 18 | 3.098512369 | 0.596371661 | 5.195606321 | 2.04E-07 | 8.712027524 | 1.676806707 | intronic | SLC7A11 | . |
| 3 month | rs116862847 | 14 | 63674959 | 0.740740001 | C | T | 421 | 0.007126 | 7 | 1.757642639 | 0.338759972 | 5.188460225 | 2.12E-07 | 15.3160369 | 2.951942625 | intergenic | WDR89;SGPP1 | dist=33088;dist=9258 |
| 3 month | rs528809914 | 13 | 112386942 | | G | A | 421 | 0.003563 | 3 | 1.373302637 | 0.265345521 | 5.17552597 | 2.27E-07 | 19.50485519 | 3.768671108 | intronic | SPACA7 | . |
| 3 month | rs112007361 | 11 | 99317649 | | A | C | 421 | 0.030879 | 26 | 4.213518738 | 0.814161472 | 5.175286336 | 2.28E-07 | 6.356584678 | 1.228257582 | intronic | CNTN5 | . |
| 3 month | rs546144116 | 19 | 19452530 | 0.741270006 | C | T | 421 | 0.002375 | 3 | 1.157037115 | 0.22363222 | 5.173839072 | 2.29E-07 | 23.13548147 | 4.471627577 | intronic | GATAD2A | . |
| 3 month | rs9643828 | 8 | 54616513 | | C | T | 421 | 0.67696 | 279 | -11.04343772 | 2.134612444 | -5.173509483 | 2.30E-07 | -2.423629403 | 0.468469114 | intronic | RP1 | . |
| 3 month | rs17510814 | 12 | 28316036 | | A | C | 421 | 0.007126 | 6 | 1.905807603 | 0.368446684 | 5.17254649 | 2.31E-07 | 14.03879235 | 2.714096892 | intronic | CCDC91 | . |
| 3 month | rs113167689 | 12 | 28283029 | | C | T | 421 | 0.007126 | 6 | 1.892111617 | 0.365850586 | 5.171815182 | 2.32E-07 | 14.13641354 | 2.733356286 | intronic | CCDC91 | . |
| 3 month | rs140788628 | 20 | 15877856 | | C | A | 421 | 0.010689 | 8 | 2.265436379 | 0.438089306 | 5.171174799 | 2.33E-07 | 11.80392839 | 2.282639604 | intronic | MACROD2 | . |
| 3 month | rs141756120 | 12 | 28358163 | | A | C | 421 | 0.008314 | 6 | 1.868183897 | 0.361717894 | 5.164753879 | 2.41E-07 | 14.27840304 | 2.764585375 | intronic | CCDC91 | . |
| 3 month | rs117991215 | 12 | 28358540 | | T | C | 421 | 0.008314 | 6 | 1.855759228 | 0.359817411 | 5.15750259 | 2.50E-07 | 14.33366601 | 2.77918736 | intronic | CCDC91 | . |
| 3 month | rs191423619 | 4 | 125595472 | 0.741770029 | G | T | 421 | 0.002375 | 3 | 1.225954306 | 0.23771753 | 5.157189324 | 2.51E-07 | 21.69461097 | 4.206673365 | intergenic | MIR2054;INTU | dist=88165;dist=2037485 |
| 3 month | rs113651406 | 4 | 997179 | | C | T | 421 | 0.003563 | 3 | 1.369997433 | 0.265704278 | 5.156098506 | 2.52E-07 | 19.40540264 | 3.763582603 | intronic | IDUA | . |
| 3 month | rs187942235 | 18 | 29450465 | | C | T | 421 | 0.003563 | 3 | 1.294384565 | 0.251912631 | 5.138228126 | 2.77E-07 | 20.39686581 | 3.969630251 | intergenic | CDH2;MIR302F | dist=1273336;dist=848446 |
| 3 month | rs16920698 | 8 | 54765874 | | G | A | 421 | 0.330166 | 277 | 11.42894943 | 2.226765953 | 5.132532863 | 2.86E-07 | 2.304926953 | 0.449081772 | intronic | RP1 | . |
| 3 month | rs4737676 | 8 | 54766986 | | G | A | 421 | 0.330166 | 277 | 11.42891023 | 2.226765217 | 5.132516953 | 2.86E-07 | 2.304920569 | 0.44908192 | intronic | RP1 | . |
| 3 month | rs4737674 | 8 | 54749094 | | C | A | 421 | 0.330166 | 277 | 11.42523599 | 2.226081333 | 5.132443196 | 2.86E-07 | 2.305595541 | 0.449219885 | intronic | RP1 | . |
| 3 month | rs13277510 | 8 | 54761589 | | G | A | 421 | 0.330166 | 277 | 11.42815338 | 2.22667847 | 5.132377007 | 2.86E-07 | 2.304947516 | 0.449099416 | intronic | RP1 | . |
| 3 month | rs983248 | 8 | 54768232 | | C | T | 421 | 0.330166 | 277 | 11.43612477 | 2.228588003 | 5.131556283 | 2.87E-07 | 2.302604284 | 0.448714611 | intronic | RP1 | . |
| 3 month | rs1391463 | 8 | 54769316 | | T | G | 421 | 0.330166 | 277 | 11.43612548 | 2.228595636 | 5.131539026 | 2.87E-07 | 2.302588654 | 0.448713075 | intronic | RP1 | . |
| 3 month | rs4737201 | 8 | 54778898 | | C | T | 421 | 0.330166 | 277 | 11.43417402 | 2.228701352 | 5.130420012 | 2.89E-07 | 2.301977341 | 0.44869179 | intronic | RP1 | . |
| 3 month | rs549931083 | 12 | 20363352 | | A | C | 421 | 0.003563 | 3 | 1.829003959 | 0.357281438 | 5.119224688 | 3.07E-07 | 14.32826937 | 2.798913946 | intergenic | LINC02468;PDE3A | dist=235451;dist=5185 |
| 3 month | rs141281289 | 11 | 123822316 | 0.733070016 | A | G | 421 | 0.005938 | 6 | 1.657109443 | 0.323734372 | 5.118731856 | 3.08E-07 | 15.81151802 | 3.088952198 | intergenic | OR6M1;TMEM225 | dist=15967;dist=60603 |
| 3 month | rs184220112 | 2 | 48404604 | | C | A | 421 | 0.004751 | 4 | 1.587567329 | 0.310301123 | 5.116215225 | 3.12E-07 | 16.48790432 | 3.22267606 | intergenic | FOXN2;PPP1R21 | dist=25309;dist=36162 |
| 3 month | rs11987234 | 8 | 54757269 | | A | G | 421 | 0.328979 | 276 | 11.39664661 | 2.227939022 | 5.115331478 | 3.13E-07 | 2.295992587 | 0.448845319 | intronic | RP1 | . |
| 3 month | rs13276543 | 8 | 54775614 | | G | T | 421 | 0.328979 | 276 | 11.41118163 | 2.230992831 | 5.114844597 | 3.14E-07 | 2.29263157 | 0.448230934 | intronic | RP1 | . |
| 3 month | rs111838310 | 2 | 74446364 | 0.780420005 | C | A | 421 | 0.004751 | 5 | 1.465896314 | 0.287002463 | 5.107608836 | 3.26E-07 | 17.79639376 | 3.48429066 | intergenic | RTKN;INO80B-WBP1 | dist=4427;dist=8659 |
| 3 month | rs80156375 | 7 | 18403592 | | A | C | 421 | 0.009501 | 8 | 2.260074467 | 0.442534058 | 5.107119835 | 3.27E-07 | 11.5406255 | 2.25971308 | intronic | HDAC9 | . |
| 3 month | rs1561297 | 8 | 54765978 | | A | C | 421 | 0.332542 | 279 | 11.40464743 | 2.234979191 | 5.102798037 | 3.35E-07 | 2.28315237 | 0.447431459 | intronic | RP1 | . |
| 3 month | rs112983626 | 2 | 74470023 | 0.782959998 | G | A | 421 | 0.004751 | 4 | 1.464072657 | 0.286917293 | 5.102768952 | 3.35E-07 | 17.78480792 | 3.485324945 | intergenic | MOGS;MRPL53 | dist=4641;dist=1959 |
| 3 month | rs12548593 | 8 | 54762057 | | G | T | 421 | 0.332542 | 279 | 11.39766972 | 2.234516943 | 5.100730946 | 3.38E-07 | 2.282699606 | 0.447524018 | intronic | RP1 | . |
| 3 month | rs10105693 | 8 | 54727912 | | C | T | 421 | 0.328979 | 276 | 11.34015363 | 2.223938019 | 5.099132049 | 3.41E-07 | 2.292839101 | 0.449652819 | intronic | RP1 | . |
| 3 month | rs13278605 | 8 | 54775611 | | C | T | 421 | 0.328979 | 277 | 11.43432709 | 2.242496155 | 5.09892829 | 3.42E-07 | 2.273773481 | 0.445931645 | intronic | RP1 | . |
| 3 month | rs2083123 | 8 | 54767758 | | C | T | 421 | 0.332542 | 279 | 11.40261082 | 2.236761629 | 5.097821184 | 3.44E-07 | 2.279107938 | 0.447074908 | intronic | RP1 | . |
| 3 month | rs17017794 | 4 | 90904734 | | T | C | 421 | 0.039192 | 33 | 4.452559757 | 0.873737599 | 5.095991934 | 3.47E-07 | 5.83240545 | 1.144508375 | intronic | CCSER1 | . |
| 3 month | rs74521112 | 11 | 99218416 | | G | T | 421 | 0.032067 | 27 | 4.149638813 | 0.814414575 | 5.095241344 | 3.48E-07 | 6.256323868 | 1.227875864 | intronic | CNTN5 | . |
| 3 month | rs116651654 | 4 | 162317591 | 0.708450019 | C | T | 421 | 0.007126 | 6 | 1.661626207 | 0.326131851 | 5.094952243 | 3.49E-07 | 15.62236937 | 3.066244515 | intergenic | FSTL5;MIR4454 | dist=153557;dist=775983 |
| 3 month | rs565682685 | 9 | 90460046 | | T | C | 421 | 0.004751 | 4 | 1.368085345 | 0.268772597 | 5.090122138 | 3.58E-07 | 18.93839699 | 3.720617399 | intergenic | LINC01508;LINC01501 | dist=26557;dist=2386 |
| 3 month | rs144541665 | 1 | 103768107 | | G | A | 421 | 0.002375 | 3 | 1.181121027 | 0.232374488 | 5.082834329 | 3.72E-07 | 21.87346108 | 4.303398392 | intergenic | AMY1C;LOC100129138 | dist=9415;dist=304916 |
| 3 month | rs193253461 | 15 | 58937154 | 0.781300008 | A | G | 421 | 0.013064 | 12 | 2.449615619 | 0.482149005 | 5.080619466 | 3.76E-07 | 10.53744676 | 2.074047629 | intergenic | SLTM;RNF111 | dist=3475;dist=50509 |
| 3 month | rs77867199 | 7 | 18402652 | | G | T | 421 | 0.010689 | 9 | 2.377239679 | 0.468684022 | 5.072158572 | 3.93E-07 | 10.82212821 | 2.13363365 | intronic | HDAC9 | . |
| 3 month | rs189360484 | 12 | 1761344 | | A | G | 421 | 0.004751 | 4 | 1.530180886 | 0.301727521 | 5.071399785 | 3.95E-07 | 16.8078794 | 3.314248553 | intronic | ADIPOR2 | . |
| 3 month | rs79182806 | 7 | 18394204 | | T | C | 421 | 0.008314 | 7 | 2.11859485 | 0.418485528 | 5.062528352 | 4.14E-07 | 12.09726027 | 2.389568893 | intronic | HDAC9 | . |
| 3 month | rs189890455 | 2 | 48428258 | | C | T | 421 | 0.005938 | 5 | 1.690230255 | 0.334051451 | 5.059790188 | 4.20E-07 | 15.14673914 | 2.993550834 | intergenic | FOXN2;PPP1R21 | dist=48963;dist=12508 |
| 3 month | rs190294315 | 9 | 83330550 | | C | T | 421 | 0.005938 | 5 | 1.754632626 | 0.347069598 | 5.055564181 | 4.29E-07 | 14.56642764 | 2.881266486 | intronic | FRMD3 | . |
| 3 month | rs185819304 | 18 | 29421615 | | G | A | 421 | 0.003563 | 3 | 1.281236388 | 0.25348417 | 5.05450257 | 4.32E-07 | 19.94011134 | 3.945019527 | intergenic | CDH2;MIR302F | dist=1244486;dist=877296 |
| 3 month | rs148433854 | 19 | 30605571 | | G | A | 421 | 0.003563 | 3 | 1.369429098 | 0.271144709 | 5.050547011 | 4.41E-07 | 18.62675851 | 3.688067545 | intronic | ZNF536 | . |
| 3 month | rs576124203 | 3 | 142122354 | | T | G | 421 | 0.003563 | 3 | 1.313230925 | 0.260150702 | 5.047962263 | 4.47E-07 | 19.4039925 | 3.843925823 | intronic | TFDP2 | . |
| 3 month | rs143597860 | 1 | 103614521 | | A | G | 421 | 0.002375 | 3 | 1.179951068 | 0.233861351 | 5.045515482 | 4.52E-07 | 21.57481541 | 4.276037896 | intergenic | AMY2B;AMY2A | dist=34987;dist=2811 |
| 3 month | rs140782222 | 9 | 81413979 | 0.778829992 | T | C | 421 | 0.002375 | 3 | 1.1680569 | 0.231519821 | 5.045170192 | 4.53E-07 | 21.79152597 | 4.31928461 | intergenic | LINC01507;TLE1 | dist=1379424;dist=169704 |
| 3 month | rs1856085 | 1 | 103571923 | | G | A | 421 | 0.002375 | 3 | 1.181208876 | 0.234330387 | 5.040784048 | 4.64E-07 | 21.51143997 | 4.267478979 | intronic | AMY2B | . |
| 3 month | rs186768950 | 19 | 18695314 | | C | A | 421 | 0.002375 | 3 | 1.117877431 | 0.221839436 | 5.039128522 | 4.68E-07 | 22.71520611 | 4.507764788 | intronic | CRTC1 | . |
| 3 month | rs182303755 | 15 | 59342593 | | A | C | 421 | 0.013064 | 12 | 2.406496876 | 0.477647778 | 5.038224785 | 4.70E-07 | 10.54799166 | 2.093592905 | intronic | MYO1E | . |
| 3 month | rs545552231 | 3 | 142145508 | | C | T | 421 | 0.003563 | 3 | 1.275833022 | 0.253432385 | 5.034214646 | 4.80E-07 | 19.8641332 | 3.945825635 | intronic | TFDP2 | . |
| 3 month | rs78547898 | 22 | 32428291 | 0.730069995 | G | A | 421 | 0.003563 | 3 | 1.210956763 | 0.240651719 | 5.03198883 | 4.85E-07 | 20.90983951 | 4.155382737 | intronic | BPIFC | . |
| 3 month | rs78907958 | 7 | 18385394 | | T | G | 421 | 0.008314 | 7 | 2.090002735 | 0.415774551 | 5.026769271 | 4.99E-07 | 12.09013217 | 2.405149614 | intronic | HDAC9 | . |
| 3 month | rs113751774 | 4 | 23427231 | 0.70836997 | C | T | 421 | 0.007126 | 7 | 1.694246649 | 0.337089114 | 5.026109061 | 5.01E-07 | 14.91032743 | 2.966574592 | intergenic | GBA3;PPARGC1A | dist=607659;dist=364790 |
| 3 month | rs10958428 | 8 | 54773081 | | A | G | 421 | 0.333729 | 280 | 11.27671394 | 2.243728439 | 5.025881807 | 5.01E-07 | 2.239968848 | 0.445686734 | intronic | RP1 | . |
| 3 month | rs17776100 | 7 | 6386848 | | G | A | 421 | 0.029691 | 25 | 3.798893519 | 0.756560221 | 5.021270496 | 5.13E-07 | 6.63697397 | 1.321771845 | intronic | RAC1 | . |

| | | | | | | | | | | | | | | | | | | |
|---|---|---|---|---|---|---|---|---|---|---|---|---|---|---|---|---|---|---|
| 3 month | rs185464792 | 19 | 18686561 | | C | T | 421 | 0.002375 | 3 | 1.114587174 | 0.222127708 | 5.017776414 | 5.23E-07 | 22.58960154 | 4.501914727 | intronic | CRTC1 | . |
| 3 month | rs117025967 | 10 | 19918123 | | C | A | 421 | 0.013064 | 11 | 2.602875018 | 0.518823163 | 5.016882829 | 5.25E-07 | 9.669735635 | 1.927439002 | intronic | PLXDC2 | . |
| 3 month | rs188028357 | 7 | 101025144 | | C | T | 421 | 0.002375 | 3 | 1.150125852 | 0.229467369 | 5.012154263 | 5.38E-07 | 21.84255775 | 4.357918094 | intronic | MUC17 | . |
| 3 month | rs148556485 | 9 | 81408911 | 0.779910028 | A | C | 421 | 0.002375 | 3 | 1.17470554 | 0.234401563 | 5.011508991 | 5.40E-07 | 21.38001525 | 4.266183157 | intergenic | LINC01507;TLE1 | dist=1374356;dist=174772 |
| 3 month | rs184613584 | 17 | 50430860 | | A | C | 421 | 0.007126 | 6 | 1.77626107 | 0.354456155 | 5.011229298 | 5.41E-07 | 14.13779737 | 2.821223402 | intronic | ACSF2 | . |
| 3 month | rs528288879 | 2 | 65648796 | 0.785229981 | C | T | 421 | 0.005938 | 6 | 1.697856008 | 0.338843001 | 5.010745399 | 5.42E-07 | 14.78780847 | 2.951219288 | intergenic | SPRED2;MIR4778 | dist=216197;dist=709451 |
| 3 month | rs1437782 | 8 | 54720202 | | C | T | 421 | 0.330166 | 278 | 11.10161994 | 2.217354849 | 5.006695229 | 5.54E-07 | 2.2579585 | 0.450987807 | exonic | RP1 | . |
| 3 month | rs534845494 | 13 | 57639730 | | A | G | 421 | 0.004751 | 5 | 1.571808362 | 0.313987509 | 5.005958262 | 5.56E-07 | 15.94317648 | 3.184840074 | intronic | PCDH17 | . |
| 3 month | rs1008091735 | 19 | 30599192 | | T | C | 421 | 0.003563 | 3 | 1.355816116 | 0.270944036 | 5.004044884 | 5.61E-07 | 18.46892428 | 3.690799086 | intronic | ZNF536 | . |
| 3 month | rs567982164 | 5 | 25631188 | 0.711179972 | G | A | 421 | 0.002375 | 3 | 1.14494858 | 0.228986639 | 5.000067194 | 5.73E-07 | 21.8356286 | 4.367067031 | intergenic | LINC02211;CDH9 | dist=328908;dist=1249409 |
| 3 month | rs79213709 | 11 | 99222724 | | G | A | 421 | 0.032067 | 26 | 4.048879944 | 0.810020577 | 4.998490236 | 5.78E-07 | 6.17081884 | 1.23453654 | intronic | CNTN5 | . |
| 3 month | rs559008174 | 19 | 18765249 | 0.769429982 | C | T | 421 | 0.004751 | 5 | 1.594517191 | 0.319680825 | 4.987841211 | 6.11E-07 | 15.60256615 | 3.12812006 | intronic | CRTC1 | . |
| 3 month | rs541288561 | 19 | 18758635 | 0.77651 | T | G | 421 | 0.004751 | 5 | 1.591688559 | 0.31930164 | 4.984905675 | 6.20E-07 | 15.61190124 | 3.131834835 | intronic | CRTC1 | . |
| 3 month | rs190251199 | 14 | 105124240 | 0.736329973 | T | C | 421 | 0.003563 | 3 | 1.240668633 | 0.248987137 | 4.982862353 | 6.27E-07 | 20.01252919 | 4.016271726 | intergenic | LINC02298;JAG2 | dist=24744;dist=16758 |
| 3 month | rs138215817 | 14 | 22173619 | | A | G | 421 | 0.004751 | 4 | 1.521988034 | 0.305467544 | 4.982486896 | 6.28E-07 | 16.31101896 | 3.273670217 | intergenic | OR4E1;LOC105370401 | dist=502284;dist=206292 |
| 3 month | rs111407636 | 5 | 95744325 | | C | T | 421 | 0.003563 | 3 | 1.331610247 | 0.26725959 | 4.98246012 | 6.28E-07 | 18.64277396 | 3.741680518 | intronic | RHOBTB3 | . |
| 3 month | rs73085348 | 3 | 42669729 | | A | G | 421 | 0.011876 | 11 | 2.448275953 | 0.491389455 | 4.982353469 | 6.28E-07 | 10.13931704 | 2.035045707 | intergenic | ZBTB47;KLHL40 | dist=2149;dist=15808 |
| 3 month | rs2375537 | 8 | 54706948 | | C | T | 421 | 0.332542 | 280 | 11.08575521 | 2.225246111 | 4.981810848 | 6.30E-07 | 2.238768477 | 0.449388495 | intronic | RP1 | . |
| 3 month | rs1437781 | 8 | 54717292 | | T | C | 421 | 0.332542 | 280 | 11.08371192 | 2.225151228 | 4.981105007 | 6.32E-07 | 2.23854673 | 0.449407657 | intronic | RP1 | . |
| 3 month | rs2274997 | 1 | 229668899 | | A | G | 421 | 0.041568 | 35 | 4.42362483 | 0.889790308 | 4.971536315 | 6.64E-07 | 5.587312281 | 1.123860297 | intergenic | URB2;LINC01682 | dist=8699;dist=206651 |
| 3 month | rs147032554 | 1 | 186179732 | | T | G | 421 | 0.003563 | 3 | 1.360299196 | 0.273670575 | 4.970571635 | 6.68E-07 | 18.16260897 | 3.654028209 | intronic | HMCN1 | . |
| 3 month | rs111676272 | 5 | 95758594 | | C | A | 421 | 0.003563 | 3 | 1.327188035 | 0.267072797 | 4.969386812 | 6.72E-07 | 18.6068625 | 3.744297477 | intronic | RHOBTB3 | . |
| 3 month | rs2274996 | 1 | 229668791 | | C | T | 421 | 0.041568 | 35 | 4.420411647 | 0.88962935 | 4.968823976 | 6.74E-07 | 5.585274332 | 1.124063633 | intergenic | URB2;LINC01682 | dist=8591;dist=206759 |
| 3 month | rs2891865 | 1 | 229670621 | | A | G | 421 | 0.041568 | 35 | 4.419830983 | 0.88984663 | 4.966958165 | 6.80E-07 | 5.581813763 | 1.123789164 | intergenic | URB2;LINC01682 | dist=10421;dist=204929 |
| 3 month | rs2385790 | 1 | 229671745 | | C | T | 421 | 0.041568 | 35 | 4.418931852 | 0.88972149 | 4.966646197 | 6.81E-07 | 5.582248215 | 1.123947226 | intergenic | URB2;LINC01682 | dist=11545;dist=203805 |
| 3 month | rs12024557 | 1 | 229676610 | | A | C | 421 | 0.042755 | 35 | 4.42434228 | 0.891299524 | 4.963923079 | 6.91E-07 | 5.569309691 | 1.121957291 | intergenic | URB2;LINC01682 | dist=16410;dist=198940 |
| 3 month | rs78296164 | 9 | 132391328 | | C | T | 421 | 0.008314 | 7 | 2.121725249 | 0.427501046 | 4.963087859 | 6.94E-07 | 11.60953385 | 2.339175566 | intronic | TTF1 | . |
| 3 month | rs570407448 | 19 | 18769220 | 0.776769996 | G | A | 421 | 0.004751 | 5 | 1.587863075 | 0.320089569 | 4.960683605 | 7.02E-07 | 15.4977984 | 3.124125552 | intronic | CRTC1 | . |
| 3 month | rs76777840 | 2 | 48085811 | | G | A | 421 | 0.005938 | 5 | 1.714828291 | 0.345687108 | 4.960637102 | 7.03E-07 | 14.35007842 | 2.892789399 | intergenic | FBXO11;FOXN2 | dist=179313;dist=228488 |
| 3 month | rs528609331 | 11 | 125972300 | | C | T | 421 | 0.003563 | 3 | 1.256070429 | 0.253271825 | 4.959376858 | 7.07E-07 | 19.58124182 | 3.948327054 | intronic | CDON | . |
| 3 month | rs182437250 | 12 | 63214686 | 0.713500023 | T | C | 421 | 0.004751 | 4 | 1.307028829 | 0.263613284 | 4.958129612 | 7.12E-07 | 18.80834508 | 3.793435539 | intergenic | AVPR1A;DPY19L2 | dist=63485;dist=344227 |
| 3 month | rs147601511 | 8 | 22464806 | | A | G | 421 | 0.004751 | 4 | 1.494160188 | 0.301407236 | 4.957280417 | 7.15E-07 | 16.44711814 | 3.317770381 | intronic | PPP3CC | . |
| 3 month | rs858397 | 8 | 54702130 | | A | G | 421 | 0.331354 | 278 | 11.01774054 | 2.2226993 | 4.956919066 | 7.16E-07 | 2.230134803 | 0.449903412 | intronic | RP1 | . |
| 3 month | rs75334617 | 10 | 101196395 | | G | A | 421 | 0.038005 | 32 | 4.196585386 | 0.847261284 | 4.953118316 | 7.30E-07 | 5.846034048 | 1.180273451 | intergenic | LINC01514;LBX1 | dist=2248;dist=30581 |
| 3 month | rs4562666 | 1 | 229689023 | | T | C | 421 | 0.042755 | 36 | 4.44283225 | 0.897203314 | 4.951867853 | 7.35E-07 | 5.519225993 | 1.114574572 | intergenic | URB2;LINC01682 | dist=28823;dist=186527 |
| 3 month | rs382476 | 8 | 54678415 | | G | A | 421 | 0.666271 | 288 | -10.90475865 | 2.202374434 | -4.951364528 | 7.37E-07 | -2.24819379 | 0.454055398 | intronic | RP1 | . |
| 3 month | rs384543 | 8 | 54679049 | | G | A | 421 | 0.666271 | 288 | -10.90475865 | 2.202374434 | -4.951364528 | 7.37E-07 | -2.24819379 | 0.454055398 | intronic | RP1 | . |
| 3 month | rs446222 | 8 | 54662400 | | G | A | 421 | 0.666271 | 288 | -10.90656484 | 2.202740432 | -4.951361803 | 7.37E-07 | -2.247819003 | 0.453979954 | intronic | RP1 | . |
| 3 month | rs384127 | 8 | 54684929 | | G | A | 421 | 0.666271 | 288 | -10.90492469 | 2.202409773 | -4.951360471 | 7.37E-07 | -2.248155875 | 0.454048112 | intronic | RP1 | . |
| 3 month | rs12045643 | 1 | 229698303 | | C | T | 421 | 0.042755 | 36 | 4.472324416 | 0.903269355 | 4.951263308 | 7.37E-07 | 5.481491517 | 1.107089479 | intergenic | URB2;LINC01682 | dist=38103;dist=177247 |
| 3 month | rs147630370 | 4 | 86529522 | | T | C | 421 | 0.004751 | 4 | 1.530518883 | 0.309272178 | 4.948776482 | 7.47E-07 | 16.00136328 | 3.23339786 | intergenic | MAPK10;MIR4452 | dist=76327;dist=12960 |
| 3 month | rs532513136 | 8 | 134801505 | | C | A | 421 | 0.003563 | 3 | 1.227599638 | 0.248065954 | 4.948682463 | 7.47E-07 | 19.94905941 | 4.031185989 | upstream | MIR30B | dist=898 |
| 3 month | rs184265355 | 13 | 107359004 | | A | C | 421 | 0.005938 | 6 | 1.694033349 | 0.342665495 | 4.94369399 | 7.67E-07 | 14.42717187 | 2.918297916 | intronic | FAM155A | . |
| 3 month | rs148248743 | 3 | 136415753 | 0.708679974 | C | T | 421 | 0.002375 | 3 | 1.069833829 | 0.216429281 | 4.943110391 | 7.69E-07 | 22.83937904 | 4.620446891 | intronic | STAG1 | . |
| 3 month | rs72983831 | 6 | 141985963 | 0.75375998 | T | G | 421 | 0.005938 | 6 | 1.556807425 | 0.31516421 | 4.939670734 | 7.83E-07 | 15.67332386 | 3.17294911 | intergenic | MIR4465;NMBR | dist=1302080;dist=88521 |
| 3 month | rs80292573 | 15 | 59142887 | | T | G | 421 | 0.034442 | 30 | 3.721656224 | 0.753598905 | 4.938510655 | 7.87E-07 | 6.553234912 | 1.326965834 | intronic | MYO1E | . |
| 3 month | rs111927235 | 2 | 74256827 | | A | G | 421 | 0.004751 | 4 | 1.423024126 | 0.288923909 | 4.925255689 | 8.43E-07 | 17.04689553 | 3.461118894 | intronic | SLC4A5 | . |
| 3 month | rs369623 | 8 | 54659380 | | A | C | 421 | 0.666271 | 287 | -10.8407396 | 2.201888659 | -4.923382276 | 8.51E-07 | -2.235981486 | 0.45415557 | intronic | RP1 | . |
| 3 month | rs188720948 | 3 | 150352093 | | T | C | 421 | 0.003563 | 3 | 1.368098189 | 0.277991774 | 4.921362129 | 8.59E-07 | 17.7032653 | 3.597228742 | intergenic | LINC01214;TSC22D2 | dist=28346;dist=56205 |
| 3 month | rs148998974 | 22 | 40224526 | | A | G | 421 | 0.005938 | 5 | 1.667947442 | 0.339010604 | 4.920045046 | 8.65E-07 | 14.51295325 | 2.949760239 | intronic | TNRC6B | . |
| 3 month | rs144954214 | 16 | 76145464 | | A | G | 421 | 0.002375 | 3 | 1.106696587 | 0.225254382 | 4.913096823 | 8.96E-07 | 21.81132631 | 4.439425294 | intergenic | CPHXL;CNTNAP4 | dist=418974;dist=131937 |
| 3 month | rs145439370 | 15 | 58587566 | | T | C | 421 | 0.030879 | 26 | 3.500007735 | 0.712559708 | 4.911879937 | 9.02E-07 | 6.893288912 | 1.403391166 | intergenic | LIPC;ADAM10 | dist=17722;dist=1243 |
| 3 month | rs75689761 | 7 | 18366950 | | C | T | 421 | 0.008314 | 7 | 2.0345491 | 0.414387092 | 4.909779146 | 9.12E-07 | 11.84829172 | 2.413202584 | intronic | HDAC9 | . |
| 3 month | rs113221952 | 1 | 103288418 | | A | G | 421 | 0.021378 | 19 | 3.040841012 | 0.619495467 | 4.908576695 | 9.17E-07 | 7.923507043 | 1.614216816 | intergenic | COL11A1;LOC101928436 | dist=179896;dist=205628 |
| 3 month | rs61434999 | 7 | 18378728 | | A | G | 421 | 0.008314 | 7 | 2.033933624 | 0.414449316 | 4.907556958 | 9.22E-07 | 11.84115106 | 2.412840272 | intronic | HDAC9 | . |
| 3 month | rs433324 | 8 | 54652049 | | A | G | 421 | 0.666271 | 287 | -10.71178821 | 2.182726874 | -4.907525693 | 9.22E-07 | -2.248346209 | 0.458142524 | intronic | RP1 | . |
| 3 month | rs528140343 | 11 | 125849332 | | A | C | 421 | 0.003563 | 3 | 1.315991741 | 0.268284941 | 4.905201681 | 9.33E-07 | 18.28355207 | 3.727380292 | intergenic | PATE4;HYLS1 | dist=9260;dist=34282 |
| 3 month | rs34270375 | 1 | 88905019 | | G | A | 421 | 0.026128 | 21 | 3.119890277 | 0.63606437 | 4.904991419 | 9.34E-07 | 7.711470175 | 1.572167924 | intergenic | GTF2B;KYAT3 | dist=13452;dist=30754 |
| 3 month | rs75773869 | 7 | 18371222 | | G | T | 421 | 0.008314 | 7 | 2.02750066 | 0.413389996 | 4.904571175 | 9.36E-07 | 11.86427156 | 2.41902322 | intronic | HDAC9 | . |
| 3 month | rs79602997 | 7 | 18370627 | | G | A | 421 | 0.008314 | 7 | 2.026922666 | 0.413391185 | 4.903158896 | 9.43E-07 | 11.86082112 | 2.419016265 | intronic | HDAC9 | . |
| 3 month | rs56224400 | 6 | 97644799 | | T | C | 421 | 0.016627 | 14 | 2.845683986 | 0.580546566 | 4.901732531 | 9.50E-07 | 8.443306398 | 1.722514712 | ncRNA_intronic | LOC101927314 | . |
| 3 month | rs186532456 | 1 | 18621834 | | C | T | 421 | 0.003563 | 3 | 1.189953152 | 0.242873045 | 4.899486277 | 9.61E-07 | 20.17303433 | 4.117377453 | intergenic | KLHDC7A;PAX7 | dist=135848;dist=9012 |
| 3 month | rs118093638 | 11 | 18696777 | | C | T | 421 | 0.005938 | 5 | 1.681215257 | 0.343292823 | 4.897321308 | 9.72E-07 | 14.26572587 | 2.912965064 | intergenic | SPTY2D1;TMEM86A | dist=62435;dist=2002 |
| 3 month | rs77353774 | 12 | 28095919 | | G | A | 421 | 0.007126 | 6 | 1.82528489 | 0.373198242 | 4.890925749 | 1.00E-06 | 13.1054362 | 2.679541026 | intergenic | PTHLH;LOC729291 | dist=123186;dist=89706 |
| 3 month | rs532730683 | 9 | 1784492 | | G | T | 421 | 0.003563 | 3 | 1.275657724 | 0.260996603 | 4.887641107 | 1.02E-06 | 18.72683804 | 3.831467497 | intergenic | DMRT2;SMARCA2 | dist=726938;dist=230855 |
| 3 month | rs185139807 | 22 | 40198777 | | G | A | 421 | 0.005938 | 5 | 1.65498412 | 0.338624126 | 4.887379825 | 1.02E-06 | 14.43305263 | 2.953126858 | intronic | TNRC6B | . |
| 3 month | rs146333745 | 18 | 57830225 | 0.779890001 | C | T | 421 | 0.003563 | 4 | 1.368613476 | 0.280282935 | 4.882971117 | 1.04E-06 | 17.42157837 | 3.567823349 | intergenic | ATP8B1;NEDD4L | dist=26910;dist=214001 |
| 3 month | rs541680196 | 22 | 40132086 | | G | A | 421 | 0.005938 | 5 | 1.654258232 | 0.339093792 | 4.878468054 | 1.07E-06 | 14.38678079 | 2.949036589 | intronic | TNRC6B | . |
| 3 month | rs12036586 | 1 | 229690631 | | G | A | 421 | 0.047506 | 40 | 4.551491816 | 0.933583971 | 4.875289164 | 1.09E-06 | 5.222121756 | 1.071140927 | intergenic | URB2;LINC01682 | dist=30431;dist=184919 |
| 3 month | rs529523094 | 16 | 77681654 | | A | G | 421 | 0.003563 | 3 | 1.307832421 | 0.26837178 | 4.873211417 | 1.10E-06 | 18.15843462 | 3.726174193 | intergenic | ADAMTS18;NUDT7 | dist=246620;dist=40838 |
| 3 month | rs562032622 | 1 | 18632839 | | A | C | 421 | 0.003563 | 3 | 1.126818759 | 0.231494862 | 4.867575676 | 1.13E-06 | 21.0267115 | 4.319750303 | intronic | PAX7 | . |
| 3 month | rs78225611 | 7 | 18367841 | | A | C | 421 | 0.009501 | 8 | 2.147570798 | 0.441262518 | 4.866877908 | 1.13E-06 | 11.02943875 | 2.266224663 | intronic | HDAC9 | . |
| 3 month | rs77346868 | 7 | 18366976 | | A | G | 421 | 0.009501 | 8 | 2.148608551 | 0.441537806 | 4.866193838 | 1.14E-06 | 11.02101286 | 2.264811725 | intronic | HDAC9 | . |

| | | | | | | | | | | | | | | | | | | |
|---|---|---|---|---|---|---|---|---|---|---|---|---|---|---|---|---|---|---|
| 3 month | rs184098071 | 2 | 176251692 | | G | A | 421 | 0.003563 | 3 | 1.695892029 | 0.34869637 | 4.863520744 | 1.15E-06 | 13.94772404 | 2.86782452 | intergenic | HOXD1;MTX2 | dist=60785;dist=17750 |
| 3 month | rs118183140 | 21 | 34105187 | | C | T | 421 | 0.02019 | 17 | 3.113677552 | 0.640523765 | 4.861142898 | 1.17E-06 | 7.589324801 | 1.561222322 | UTR3 | SLC5A3 | NM_006933:c.*78320>0 |
| 3 month | rs113625788 | 22 | 19981659 | | C | T | 421 | 0.008314 | 7 | 2.000739147 | 0.411621625 | 4.860626906 | 1.17E-06 | 11.80848286 | 2.429415606 | exonic | ARVCF | . |
| 3 month | rs116189766 | 2 | 125636287 | 0.723290026 | T | C | 421 | 0.002375 | 3 | 1.114549479 | 0.229338184 | 4.85985133 | 1.17E-06 | 21.19076397 | 4.360372887 | intergenic | CNTNAP5;LINC01941 | dist=715069;dist=473813 |
| 3 month | rs76098744 | 1 | 111806750 | | C | T | 421 | 0.016627 | 14 | 2.811615503 | 0.579024995 | 4.855775707 | 1.20E-06 | 8.386124522 | 1.727041163 | intronic | KCND3 | . |
| 3 month | rs74683551 | 1 | 111811796 | | G | A | 421 | 0.016627 | 14 | 2.810351968 | 0.579019309 | 4.853641189 | 1.21E-06 | 8.38252043 | 1.727058121 | intronic | KCND3 | . |
| 3 month | rs185855183 | 12 | 101111348 | | C | T | 421 | 0.003563 | 3 | 1.256842073 | 0.258975211 | 4.853136594 | 1.22E-06 | 18.73977272 | 3.861373435 | intronic | ANO4 | . |
| 3 month | rs405226 | 8 | 54679776 | | A | G | 421 | 0.662708 | 291 | -10.75682372 | 2.217202771 | -4.851529081 | 1.23E-06 | -2.188130533 | 0.45101874 | intronic | RP1 | . |
| 3 month | rs3098298 | 8 | 54670278 | | C | T | 421 | 0.662708 | 291 | -10.75741494 | 2.217551047 | -4.851033737 | 1.23E-06 | -2.187563503 | 0.450947906 | intronic | RP1 | . |
| 3 month | rs367179 | 8 | 54675056 | | T | C | 421 | 0.662708 | 291 | -10.75741494 | 2.217551047 | -4.851033737 | 1.23E-06 | -2.187563503 | 0.450947906 | intronic | RP1 | . |
| 3 month | rs432393 | 8 | 54667738 | | C | T | 421 | 0.662708 | 291 | -10.75654812 | 2.217474543 | -4.850810194 | 1.23E-06 | -2.187538166 | 0.450963464 | intronic | RP1 | . |
| 3 month | rs117185941 | 21 | 37394182 | | G | A | 421 | 0.005938 | 5 | 1.529223098 | 0.31531516 | 4.849824212 | 1.24E-06 | 15.38087864 | 3.171430132 | intronic | DYRK1A | . |
| 3 month | rs139360368 | 5 | 74076284 | | A | C | 421 | 0.003563 | 4 | 1.280137576 | 0.264060375 | 4.847897286 | 1.25E-06 | 18.35904869 | 3.78701272 | intergenic | ARHGEF28;LINC01335 | dist=134291;dist=230126 |
| 3 month | rs184117160 | 15 | 59112107 | 0.770810008 | C | T | 421 | 0.014252 | 11 | 2.271648443 | 0.468677492 | 4.846933088 | 1.25E-06 | 10.34172362 | 2.133663377 | intronic | CCNB2 | . |
| 3 month | rs145676540 | 3 | 2002951 | | C | T | 421 | 0.005938 | 5 | 1.68032939 | 0.346683687 | 4.846866048 | 1.25E-06 | 13.98065798 | 2.884473769 | intergenic | CNTN6;CNTN4 | dist=598734;dist=95852 |
| 3 month | rs556293455 | 2 | 176085692 | | G | A | 421 | 0.003563 | 3 | 1.630922146 | 0.336702149 | 4.843812709 | 1.27E-06 | 14.38604633 | 2.969984018 | intergenic | EVX2;HOXD13 | dist=1730;dist=7029 |
| 3 month | rs150027952 | 9 | 100623134 | 0.704400003 | A | G | 421 | 0.002375 | 3 | 1.072594942 | 0.221501288 | 4.842386925 | 1.28E-06 | 21.86166485 | 4.514646431 | intergenic | CAVIN4;PLPPR1 | dist=34747;dist=405593 |
| 3 month | rs138217865 | 15 | 93849653 | | C | T | 421 | 0.004751 | 4 | 1.402299902 | 0.28969226 | 4.840653664 | 1.29E-06 | 16.70964097 | 3.451938959 | intergenic | LOC105370980;LINC02207 | dist=641605;dist=6907 |
| 3 month | rs112475378 | 3 | 1584977 | 0.749329984 | T | C | 421 | 0.016627 | 14 | 2.445371748 | 0.506000821 | 4.832742644 | 1.35E-06 | 9.550859286 | 1.976281377 | intergenic | CNTN6;CNTN4 | dist=180760;dist=513826 |
| 3 month | rs146007933 | 3 | 28185932 | | T | C | 421 | 0.021378 | 18 | 3.108745063 | 0.643323442 | 4.832320511 | 1.35E-06 | 7.511494525 | 1.554428045 | intergenic | LINC01980;CMC1 | dist=325607;dist=55687 |
| 3 month | rs192443987 | 3 | 135813503 | | G | A | 421 | 0.004751 | 4 | 1.31410558 | 0.27206124 | 4.830183004 | 1.36E-06 | 17.75402845 | 3.675643022 | intergenic | EPHB1;PPP2R3A | dist=553038;dist=152225 |
| 3 month | rs149493615 | 1 | 79414404 | | G | A | 421 | 0.008314 | 8 | 1.98260972 | 0.410759865 | 4.826688019 | 1.39E-06 | 11.750632 | 2.434512435 | intergenic | ADGRL4;LINC01781 | dist=407674;dist=1121351 |
| 3 month | rs140706881 | 10 | 94457778 | | G | A | 421 | 0.004751 | 4 | 1.226743807 | 0.254265992 | 4.824647601 | 1.40E-06 | 18.97480496 | 3.932889306 | intronic | TBC1D12 | . |
| 3 month | rs142894171 | 2 | 150581757 | | G | T | 421 | 0.003563 | 3 | 1.287888043 | 0.266960796 | 4.82425907 | 1.41E-06 | 18.07103941 | 3.74586836 | intergenic | LINC01920;LINC02612 | dist=9536;dist=47140 |
| 3 month | rs17746486 | 2 | 95056864 | 0.71280998 | C | T | 421 | 0.032067 | 29 | 3.355977065 | 0.696974974 | 4.815061071 | 1.47E-06 | 6.908513578 | 1.434771746 | intergenic | MAL;MRPS5 | dist=2872;dist=28507 |
| 3 month | rs180764936 | 12 | 101104843 | | T | C | 421 | 0.003563 | 3 | 1.197190428 | 0.248754746 | 4.812734014 | 1.49E-06 | 19.34730528 | 4.020023799 | intronic | ANO4 | . |
| 3 month | rs190806532 | 12 | 48468364 | 0.728540003 | G | T | 421 | 0.002375 | 3 | 1.082541559 | 0.225298273 | 4.804926128 | 1.55E-06 | 21.32695498 | 4.438560431 | intergenic | ZNF641;ANP32D | dist=117118;dist=4195 |
| 3 month | rs147393020 | 10 | 123019758 | | A | G | 421 | 0.005938 | 5 | 1.619245544 | 0.337859644 | 4.792657467 | 1.65E-06 | 14.1853505 | 2.959808958 | intronic | ACADSB | . |
| 3 month | rs79539453 | 11 | 125396457 | | C | T | 421 | 0.002375 | 3 | 1.102705134 | 0.230127137 | 4.791721423 | 1.65E-06 | 20.82206158 | 4.345424064 | intronic | PKNOX2 | . |
| 3 month | rs371245624 | 5 | 163453950 | | T | C | 421 | 0.003563 | 3 | 1.17843308 | 0.245958889 | 4.791179069 | 1.66E-06 | 19.47959308 | 4.065720108 | UTR3 | NUDCD2 | NM_001329991:c.*170>0;NM |
| 3 month | rs536803366 | 8 | 121989172 | 0.708639979 | T | C | 421 | 0.002375 | 3 | 1.194921552 | 0.249406377 | 4.791062547 | 1.66E-06 | 19.20986385 | 4.009520573 | intergenic | HAS2-AS1;SMILR | dist=343847;dist=425155 |
| 3 month | rs150077525 | 13 | 57405415 | 0.75563997 | A | G | 421 | 0.007126 | 7 | 1.765848692 | 0.368578785 | 4.790966716 | 1.66E-06 | 12.99848746 | 2.713124141 | intergenic | PRR20E;PCDH17 | dist=235197;dist=226329 |
| 3 month | rs371879555 | 12 | 22863028 | 0.786769986 | T | C | 421 | 0.003563 | 3 | 1.139634583 | 0.238093921 | 4.786491727 | 1.70E-06 | 20.10337646 | 4.200023233 | intergenic | ETNK1;LOC101928441 | dist=172363;dist=312608 |
| 3 month | rs571986619 | 6 | 84969114 | 0.767470002 | A | G | 421 | 0.002375 | 3 | 1.102004226 | 0.230254532 | 4.786026206 | 1.70E-06 | 20.78580673 | 4.343019831 | intergenic | TBX18;LINC02535 | dist=204516;dist=418105 |
| 3 month | rs183586634 | 21 | 37390730 | | G | A | 421 | 0.005938 | 5 | 1.598766374 | 0.334213375 | 4.783669629 | 1.72E-06 | 14.31322017 | 2.992100476 | intronic | DYRK1A | . |
| 3 month | rs143811231 | 1 | 79502446 | | T | C | 421 | 0.008314 | 8 | 1.975347907 | 0.413110429 | 4.78164619 | 1.74E-06 | 11.57474094 | 2.420660266 | intergenic | ADGRL4;LINC01781 | dist=495716;dist=1033309 |
| 3 month | rs2365739 | 1 | 62018790 | | G | A | 421 | 0.021378 | 18 | 3.317349453 | 0.694985007 | 4.773267649 | 1.81E-06 | 6.868159166 | 1.43887996 | intronic | PATJ | . |
| 3 month | rs146526206 | 4 | 90071867 | | T | C | 421 | 0.017815 | 15 | 2.815290022 | 0.58984771 | 4.772909977 | 1.82E-06 | 8.091766558 | 1.695352855 | intergenic | MMRN1;CCSER1 | dist=117257;dist=55527 |
| 3 month | rs567383525 | 8 | 114017265 | | C | T | 421 | 0.003563 | 3 | 1.282541719 | 0.268838431 | 4.770678479 | 1.84E-06 | 17.74552267 | 3.719706274 | intergenic | CSMD3;TRPS1 | dist=580326;dist=1391230 |
| 3 month | rs149425014 | 15 | 58659461 | 0.716620028 | T | C | 421 | 0.026128 | 22 | 3.013015463 | 0.631629727 | 4.770224288 | 1.84E-06 | 7.552247919 | 1.583206043 | intronic | ADAM10 | . |
| 3 month | rs559559983 | 1 | 246813830 | | C | A | 421 | 0.007126 | 5 | 1.645339674 | 0.345154188 | 4.76697004 | 1.87E-06 | 13.81113197 | 2.897255878 | intergenic | LINC01341;AHCTF1 | dist=22344;dist=25268 |
| 3 month | rs117166500 | 7 | 17013154 | | G | T | 421 | 0.007126 | 7 | 1.813506071 | 0.380670761 | 4.763975219 | 1.90E-06 | 12.51468647 | 2.626941975 | intergenic | AGR3;AHR | dist=131171;dist=285498 |
| 3 month | rs145766563 | 1 | 185160370 | | G | A | 421 | 0.003563 | 4 | 1.34508864 | 0.282384562 | 4.76332215 | 1.90E-06 | 16.86821019 | 3.541270076 | intronic | SWT1 | . |
| 3 month | rs540065886 | 9 | 2770228 | 0.797429979 | T | C | 421 | 0.003563 | 3 | 1.13736568 | 0.23877586 | 4.763319367 | 1.90E-06 | 19.94891509 | 4.188028045 | intergenic | KCNV2;PUM3 | dist=40191;dist=33927 |
| 3 month | rs118084887 | 21 | 37491518 | | T | C | 421 | 0.005938 | 5 | 1.518241868 | 0.31880157 | 4.762341255 | 1.91E-06 | 14.93826162 | 3.136747415 | intronic | DYRK1A | . |
| 3 month | rs140420703 | 1 | 102337928 | 0.700510025 | T | G | 421 | 0.004751 | 5 | 1.375529427 | 0.288838786 | 1.375529427 | 1.91E-06 | 16.48765399 | 3.462138915 | intergenic | OLFM3;COL11A1 | dist=340694;dist=538539 |
| 3 month | rs568321148 | 2 | 29647482 | | T | G | 421 | 0.003563 | 3 | 1.279867135 | 0.268808164 | 4.761265858 | 1.92E-06 | 17.71250463 | 3.720125102 | intronic | ALK | . |
| 3 month | rs55844051 | 7 | 23320744 | | T | C | 421 | 0.003563 | 3 | 1.232383167 | 0.25888714 | 4.760310483 | 1.93E-06 | 18.3875896 | 3.862687038 | intronic | IGF2BP3 | . |
| 3 month | rs187236873 | 5 | 92234630 | | G | A | 421 | 0.007126 | 6 | 1.81660653 | 0.381998785 | 4.755529603 | 1.98E-06 | 12.44906998 | 2.617809375 | intergenic | ARRDC3-AS1;NR2F1-AS1 | dist=813915;dist=1214615 |
| 3 month | rs1812506 | 8 | 54763541 | | A | G | 421 | 0.345606 | 291 | 10.64127813 | 2.239050947 | 4.752584188 | 2.01E-06 | 2.122588678 | 0.446617796 | intronic | RP1 | . |
| 3 month | rs539713344 | 7 | 100877165 | 0.772019982 | G | A | 421 | 0.002375 | 3 | 1.125805228 | 0.236990667 | 4.750420101 | 2.03E-06 | 20.04475604 | 4.219575451 | intronic | SRRT | . |
| 3 month | rs536781978 | 2 | 29510815 | | A | G | 421 | 0.003563 | 3 | 1.282866581 | 0.270080884 | 4.749934759 | 2.03E-06 | 17.58708236 | 3.702594509 | intronic | ALK | . |
| 3 month | rs76327548 | 12 | 100789188 | | G | A | 421 | 0.013064 | 11 | 2.458077916 | 0.517503975 | 4.749872531 | 2.04E-06 | 9.178427144 | 1.932352307 | intergenic | GAS2L3;ANO4 | dist=160900;dist=5588 |
| 3 month | rs149421869 | 2 | 53256291 | | G | T | 421 | 0.008314 | 8 | 1.92083422 | 0.404523583 | 4.748386252 | 2.05E-06 | 11.73821862 | 2.472043763 | intergenic | MIR4431;ASB3 | dist=553676;dist=413688 |
| 3 month | rs75024143 | 21 | 21784226 | | G | T | 421 | 0.014252 | 12 | 2.293452396 | 0.483452813 | 4.743901232 | 2.10E-06 | 9.812542411 | 2.068454196 | ncRNA_intronic | LINC01425 | . |
| 3 month | rs2375536 | 8 | 54728162 | | T | C | 421 | 0.347981 | 292 | 10.56754692 | 2.229301935 | 4.740294148 | 2.13E-06 | 2.126358065 | 0.448570911 | intronic | RP1 | . |
| 3 month | rs141326851 | 6 | 134511989 | | A | C | 421 | 0.016627 | 14 | 2.616968305 | 0.552323641 | 4.738106631 | 2.16E-06 | 8.578496884 | 1.810532677 | intergenic | LINC01010;LOC101928304 | dist=7969;dist=13329 |
| 3 month | rs187518659 | 1 | 98990189 | | T | G | 421 | 0.009501 | 8 | 1.921562765 | 0.405573328 | 4.737892339 | 2.16E-06 | 11.68196232 | 2.465645372 | intronic | PLPPR5 | . |
| 3 month | rs146479102 | 2 | 65598625 | 0.738789976 | G | A | 421 | 0.005938 | 5 | 1.448302352 | 0.305828916 | 4.735661917 | 2.18E-06 | 15.48467677 | 3.269801992 | intergenic | SPRED2;MIR4778 | dist=166026;dist=759622 |
| 3 month | rs16850124 | 1 | 229695584 | | T | C | 421 | 0.043943 | 37 | 4.326094288 | 0.91378287 | 4.73426941 | 2.20E-06 | 5.180956622 | 1.094351878 | intergenic | URB2;LINC01682 | dist=35384;dist=179966 |
| 3 month | rs115348382 | 1 | 9595845 | | G | A | 421 | 0.003563 | 3 | 1.230366386 | 0.259928152 | 4.733486451 | 2.21E-06 | 18.21074945 | 3.847216981 | intronic | TMEM201 | . |
| 3 month | rs184200893 | 2 | 69033781 | | C | T | 421 | 0.003563 | 3 | 1.121211249 | 0.237019909 | 4.730451775 | 2.24E-06 | 19.95803558 | 4.219054866 | intronic | ANTXR1 | . |
| 3 month | rs148781275 | 11 | 103769875 | 0.774079978 | A | G | 421 | 0.004751 | 5 | 1.423809766 | 0.301038131 | 4.729665848 | 2.25E-06 | 15.71118528 | 3.321838325 | intergenic | DYNC2H1;MIR4693 | dist=290012;dist=80031 |
| 3 month | rs139943877 | 3 | 155733500 | | G | A | 421 | 0.007126 | 7 | 1.713092922 | 0.362264246 | 4.728849011 | 2.26E-06 | 13.0535902 | 2.760415942 | intronic | PLCH1 | . |
| 3 month | rs546409459 | 11 | 125885094 | | A | G | 421 | 0.003563 | 3 | 1.180777655 | 0.249820856 | 4.726497517 | 2.28E-06 | 18.91954737 | 4.002868361 | intronic | HYLS1 | . |
| 3 month | rs186649043 | 3 | 175214503 | 0.769129992 | C | T | 421 | 0.003563 | 3 | 1.162234876 | 0.245948741 | 4.725516671 | 2.30E-06 | 19.21342086 | 4.065887859 | intronic | NAALADL2 | . |
| 3 month | rs80203220 | 3 | 123003484 | | C | T | 421 | 0.005938 | 6 | 1.662045235 | 0.351841788 | 4.723842626 | 2.31E-06 | 13.42604201 | 2.842186558 | intronic | SEMA5B | . |
| 3 month | rs541508507 | 2 | 169166755 | | G | A | 421 | 0.004751 | 4 | 1.442660176 | 0.305446461 | 4.723119638 | 2.32E-06 | 15.46300334 | 3.273896179 | intronic | LRP2 | . |
| 3 month | rs529011661 | 10 | 20083387 | | G | A | 421 | 0.004751 | 4 | 1.425272643 | 0.301883703 | 4.721263948 | 2.34E-06 | 15.63934688 | 3.312533901 | intronic | PLXDC2 | . |
| 3 month | rs182959028 | 22 | 45427152 | | T | C | 421 | 0.008314 | 7 | 1.863907765 | 0.395028955 | 4.718407959 | 2.38E-06 | 11.94446103 | 2.531460004 | intronic | RIBC2 | . |
| 3 month | rs146207930 | 9 | 126280057 | | A | G | 421 | 0.007126 | 6 | 1.87537986 | 0.397610416 | 4.716626584 | 2.40E-06 | 11.86243214 | 2.515024654 | intergenic | LOC101929116;MVB12B | dist=4142;dist=46772 |
| 3 month | rs290120 | 5 | 163841238 | | T | G | 421 | 0.007126 | 6 | 1.664921355 | 0.353031552 | 4.716069562 | 2.40E-06 | 13.3587764 | 2.832608007 | intergenic | MAT2B;LINC02143 | dist=321884;dist=607184 |
| 3 month | rs531769270 | 4 | 153489891 | 0.744459987 | T | C | 421 | 0.003563 | 3 | 1.339792194 | 0.284142497 | 4.715212285 | 2.41E-06 | 16.59453383 | 3.519360917 | intronic | TMEM131L | . |

| | | | | | | | | | | | | | | | | | | |
|---|---|---|---|---|---|---|---|---|---|---|---|---|---|---|---|---|---|---|
| 3 month | rs185620578 | 12 | 48175616 | 0.752129972 | C | T | 421 | 0.002375 | 3 | 1.072824946 | 0.227528468 | 4.715124023 | 2.42E-06 | 20.72322668 | 4.39505442 | intergenic | ASB8;CCDC184 | dist=18101;dist=8028 |
| 3 month | rs62447184 | 7 | 36534898 | 0.730589986 | G | A | 421 | 0.042755 | 43 | 4.329544629 | 0.918817438 | 4.712083651 | 2.45E-06 | 5.128422094 | 1.088355486 | intronic | AOAH | . |
| 3 month | rs118184666 | 12 | 20271815 | | G | A | 421 | 0.007126 | 6 | 2.026245701 | 0.430149631 | 4.710560123 | 2.47E-06 | 10.95098026 | 2.324772421 | intergenic | LINC02468;PDE3A | dist=143914;dist=96722 |
| 3 month | rs10494861 | 1 | 205362746 | | G | A | 421 | 0.003563 | 3 | 1.156337638 | 0.245524493 | 4.709663065 | 2.48E-06 | 19.18204982 | 4.072913404 | intergenic | KLHDC8A;LEMD1-AS1 | dist=5656;dist=10506 |
| 3 month | rs183817723 | 16 | 59268871 | 0.79569 | C | T | 421 | 0.003563 | 4 | 1.261174136 | 0.267795557 | 4.709466243 | 2.48E-06 | 17.5860507 | 3.734191901 | intergenic | GOT2;APOOP5 | dist=534555;dist=485270 |
| 3 month | rs12502861 | 4 | 2424578 | | T | C | 421 | 0.010689 | 9 | 1.987294963 | 0.422008794 | 4.709131639 | 2.49E-06 | 11.15884718 | 2.369618867 | intronic | CFAP99 | . |
| 3 month | rs191792521 | 3 | 195919734 | 0.757189989 | G | A | 421 | 0.008314 | 6 | 1.587589638 | 0.337332296 | 4.706307863 | 2.52E-06 | 13.95154842 | 2.964435992 | intergenic | TNK2-AS1;SDHAP1 | dist=6470;dist=40187 |
| 3 month | rs149949098 | 11 | 95366702 | | G | A | 421 | 0.016627 | 13 | 2.587327209 | 0.549803064 | 4.705916312 | 2.53E-06 | 8.559276254 | 1.818833079 | intergenic | LOC100129203;FAM76B | dist=132298;dist=402251 |
| 3 month | rs74343174 | 5 | 162066176 | 0.766919971 | C | A | 421 | 0.005938 | 5 | 1.498573008 | 0.318559187 | 4.704221604 | 2.55E-06 | 14.76718237 | 3.139134082 | intergenic | LINC01202;GABRG2 | dist=64980;dist=1289 |
| 3 month | rs189765693 | 4 | 4323066 | | T | C | 421 | 0.004751 | 4 | 1.425732565 | 0.30309955 | 4.70384257 | 2.55E-06 | 15.51913413 | 3.299246074 | intergenic | ZBTB49;NSG1 | dist=1283;dist=63466 |
| 3 month | rs189912648 | 5 | 135429749 | | C | T | 421 | 0.003563 | 4 | 1.303529678 | 0.277126356 | 4.703737665 | 2.55E-06 | 16.97325991 | 3.608462274 | intergenic | MACROH2A1;DCANP1 | dist=29862;dist=14465 |
| 3 month | rs151323346 | 12 | 20859090 | | T | C | 421 | 0.005938 | 4 | 1.39062978 | 0.295827471 | 4.700813538 | 2.59E-06 | 15.89038883 | 3.380348678 | intronic | SLCO1B3;SLCO1B3-SLCO1B7 | . |
| 3 month | rs536023430 | 7 | 147167980 | 0.788429976 | T | C | 421 | 0.003563 | 3 | 1.137129093 | 0.242081012 | 4.697308079 | 2.64E-06 | 19.40386834 | 4.130848564 | intronic | CNTNAP2 | . |
| 3 month | rs144026361 | 15 | 40956471 | 0.714779973 | C | T | 421 | 0.004751 | 5 | 1.365790883 | 0.290922074 | 4.694696631 | 2.67E-06 | 16.13729945 | 3.437346588 | UTR3 | CHAC1 | NM_001142776:c.*6970>0;NM |
| 3 month | rs186142189 | 2 | 67475575 | | G | A | 421 | 0.007126 | 6 | 1.7290309 | 0.36845255 | 4.692682681 | 2.70E-06 | 12.73619271 | 2.714053683 | intergenic | ETAA1;LINC01812 | dist=63486;dist=320479 |
| 3 month | rs142549310 | 2 | 169173996 | | C | T | 421 | 0.004751 | 4 | 1.443948619 | 0.307707803 | 4.692596694 | 2.70E-06 | 15.25017126 | 3.249836338 | exonic | LRP2 | . |
| 3 month | rs72832764 | 5 | 170577669 | | G | A | 421 | 0.003563 | 3 | 1.208783888 | 0.257601337 | 4.692459687 | 2.70E-06 | 18.21597569 | 3.881967434 | intronic | KCNIP1 | . |
| 3 month | rs183180157 | 1 | 181277985 | | A | C | 421 | 0.009501 | 7 | 1.744207622 | 0.371856251 | 4.690542692 | 2.72E-06 | 12.61385999 | 2.689211211 | intergenic | LINC01699;CACNA1E | dist=39381;dist=205532 |
| 3 month | rs73227413 | 21 | 21764653 | | G | A | 421 | 0.034442 | 28 | 3.687754962 | 0.78647158 | 4.688986931 | 2.75E-06 | 5.962055143 | 1.271501762 | ncRNA_intronic | LINC01425 | . |
| 3 month | rs185874707 | 8 | 18173165 | | C | T | 421 | 0.011876 | 10 | 2.405517016 | 0.513034555 | 4.688801158 | 2.75E-06 | 9.139347655 | 1.949186444 | intronic | NAT1 | . |
| 3 month | rs423841 | 8 | 54643509 | | G | A | 421 | 0.662708 | 291 | -10.22664529 | 2.181175145 | -4.688594272 | 2.75E-06 | -2.149572575 | 0.458468456 | intronic | RP1 | . |
| 3 month | rs191271637 | 17 | 54045899 | 0.762179971 | A | G | 421 | 0.003563 | 3 | 1.195597009 | 0.255047807 | 4.687736869 | 2.76E-06 | 18.379836 | 3.920833552 | intergenic | KIF2B;TOM1L1 | dist=220706;dist=854792 |
| 3 month | rs187978759 | 7 | 11672218 | | G | A | 421 | 0.003563 | 3 | 1.07420489 | 0.229159549 | 4.687585114 | 2.76E-06 | 20.4555522 | 4.3637719 | intronic | THSD7A | . |
| 3 month | rs193153124 | 3 | 148612923 | | A | G | 421 | 0.005938 | 5 | 1.529063164 | 0.32624874 | 4.686801743 | 2.78E-06 | 14.36573132 | 3.065145937 | intergenic | LINC02046;AGTR1 | dist=212967;dist=84948 |
| 3 month | rs1877768 | 6 | 16534692 | | C | T | 421 | 0.017815 | 15 | 2.650393922 | 0.565512508 | 4.686711405 | 2.78E-06 | 8.287546847 | 1.768307483 | intronic | ATXN1 | . |
| 3 month | rs76617932 | 1 | 180961288 | | T | C | 421 | 0.011876 | 9 | 2.08220138 | 0.444364012 | 4.685801107 | 2.79E-06 | 10.54496084 | 2.250407262 | intergenic | KIAA1614-AS1;STX6 | dist=6401;dist=11426 |
| 3 month | rs188034471 | 9 | 83322799 | | G | A | 421 | 0.004751 | 4 | 1.487832555 | 0.317535808 | 4.68555835 | 2.79E-06 | 14.75599992 | 3.149251127 | intronic | FRMD3 | . |
| 3 month | rs1595406 | 8 | 54718055 | | A | G | 421 | 0.345606 | 291 | 10.45008066 | 2.230645943 | 4.684777831 | 2.80E-06 | 2.100188892 | 0.448300638 | intronic | RP1 | . |
| 3 month | rs147627638 | 6 | 98725240 | | A | G | 421 | 0.007126 | 6 | 1.675781082 | 0.357955603 | 4.681533316 | 2.85E-06 | 13.07853062 | 2.793642539 | intergenic | MIR2113;PNKY | dist=700621;dist=104901 |
| 3 month | rs151272830 | 2 | 67455592 | | G | T | 421 | 0.007126 | 6 | 1.643645432 | 0.351116105 | 4.681202055 | 2.85E-06 | 13.33234787 | 2.848060758 | intergenic | ETAA1;LINC01812 | dist=43503;dist=340462 |
| 3 month | rs141169929 | 3 | 165090674 | 0.77117002 | A | G | 421 | 0.004751 | 5 | 1.473704308 | 0.315071386 | 4.677366376 | 2.91E-06 | 14.84541783 | 3.173883899 | intergenic | SI;SLITRK3 | dist=12178;dist=96046 |
| 3 month | rs720372 | 8 | 54716077 | | G | A | 421 | 0.346793 | 292 | 10.41667706 | 2.227853301 | 4.675656629 | 2.93E-06 | 2.098727338 | 0.448862589 | intronic | RP1 | . |
| 3 month | rs567080482 | 13 | 94423151 | | T | C | 421 | 0.004751 | 4 | 1.409440819 | 0.301549206 | 4.67399944 | 2.95E-06 | 15.49995607 | 3.316208371 | intergenic | GPC6;DCT | dist=15132;dist=13660 |
| 3 month | rs546286713 | 13 | 90758825 | | G | A | 421 | 0.004751 | 4 | 1.389978902 | 0.297519732 | 4.671888118 | 2.98E-06 | 15.70278409 | 3.361121605 | intergenic | LINC01049;LINC00410 | dist=223484;dist=132129 |
| 3 month | rs142311947 | 5 | 177957468 | 0.768029988 | G | A | 421 | 0.007126 | 8 | 1.813233127 | 0.388171459 | 4.671217019 | 2.99E-06 | 12.03390127 | 2.576181159 | ncRNA_intronic | LOC100128340 | . |
| 3 month | rs185155853 | 15 | 40951902 | 0.723249972 | C | T | 421 | 0.004751 | 5 | 1.370089738 | 0.293393972 | 4.669795111 | 3.02E-06 | 15.91646574 | 3.408386314 | intergenic | DLL4;CHAC1 | dist=12829;dist=1569 |
| 3 month | rs550763536 | 2 | 7312216 | | T | G | 421 | 0.005938 | 5 | 1.504113282 | 0.322217093 | 4.66801208 | 3.04E-06 | 14.48716465 | 3.103497679 | intergenic | LOC101929452;LOC1005062 | dist=234336;dist=109045 |
| 3 month | rs555040883 | 22 | 40235472 | | G | A | 421 | 0.004751 | 4 | 1.412000774 | 0.302532759 | 4.667265721 | 3.05E-06 | 15.42730692 | 3.305427169 | intronic | TNRC6B | . |
| 3 month | rs2375219 | 8 | 54785735 | | C | T | 421 | 0.393112 | 330 | 10.58980707 | 2.26895928 | 4.667253025 | 3.05E-06 | 2.057001668 | 0.440730695 | intronic | RP1 | . |
| 3 month | rs140062526 | 13 | 58711899 | | G | A | 421 | 0.005938 | 5 | 1.544633202 | 0.331067832 | 4.665609433 | 3.08E-06 | 14.09260875 | 3.020529034 | intergenic | LINC00374;DIAPH3 | dist=478782;dist=953688 |
| 3 month | rs572961122 | 13 | 107360637 | 0.793820024 | C | T | 421 | 0.005938 | 6 | 1.632302386 | 0.349861236 | 4.665570858 | 3.08E-06 | 13.33548956 | 2.858276075 | intronic | FAM155A | . |
| 3 month | rs544042801 | 11 | 68692775 | | G | A | 421 | 0.004751 | 4 | 1.370120555 | 0.293669353 | 4.665521075 | 3.08E-06 | 15.88698661 | 3.405190192 | intergenic | GAL;TESMIN | dist=1600;dist=14665 |
| 3 month | rs142993106 | 4 | 90036221 | | G | A | 421 | 0.017815 | 15 | 2.691736025 | 0.577013528 | 4.664944401 | 3.09E-06 | 8.084636111 | 1.733061622 | intergenic | MMRN1;CCSER1 | dist=81611;dist=91173 |
| 3 month | rs569916471 | 14 | 75424639 | 0.771009982 | G | A | 421 | 0.004751 | 5 | 1.390580812 | 0.298176949 | 4.663609368 | 3.11E-06 | 15.64040878 | 3.353713303 | intergenic | LINC01220;JDP2 | dist=128231;dist=3085 |
| 3 month | rs7104959 | 11 | 129976231 | | C | T | 421 | 0.003563 | 3 | 1.212990302 | 0.260168118 | 4.66233262 | 3.13E-06 | 17.92046105 | 3.843668505 | intronic | PRDM10 | . |
| 3 month | rs559228693 | 20 | 15982684 | 0.798879981 | G | A | 421 | 0.005938 | 5 | 1.512274875 | 0.324468321 | 4.660778189 | 3.15E-06 | 14.36435511 | 3.081964969 | ncRNA_intronic | LOC613266 | . |
| 3 month | rs187384541 | 8 | 1684075 | 0.760779977 | A | G | 421 | 0.035629 | 26 | 3.153891836 | 0.676696813 | 4.660716257 | 3.15E-06 | 6.887451174 | 1.477766677 | intronic | DLGAP2 | . |
| 3 month | rs141127122 | 22 | 40208435 | | G | A | 421 | 0.004751 | 4 | 1.411818313 | 0.302953602 | 4.660179993 | 3.16E-06 | 15.38248751 | 3.30083549 | intronic | TNRC6B | . |
| 3 month | rs193093906 | 10 | 125016920 | | G | A | 421 | 0.009501 | 9 | 2.062968829 | 0.442790343 | 4.659019469 | 3.18E-06 | 10.52195366 | 2.25840517 | intronic | CTBP2 | . |
| 3 month | rs77871739 | 9 | 135660463 | | G | A | 421 | 0.004751 | 4 | 1.439297375 | 0.309017583 | 4.657655271 | 3.20E-06 | 15.07246035 | 3.236061811 | intergenic | GLT6D1;LCN9 | dist=20923;dist=2859 |
| 3 month | rs117816016 | 8 | 102739034 | | C | T | 421 | 0.003563 | 3 | 1.090578449 | 0.234213862 | 4.656336053 | 3.22E-06 | 19.88070227 | 4.269602116 | intergenic | LOC101927245;GASAL1 | dist=52311;dist=67788 |
| 3 month | rs775626702 | 5 | 163203061 | | A | C | 421 | 0.003563 | 3 | 1.143758046 | 0.245686137 | 4.655362562 | 3.23E-06 | 18.94841366 | 4.070233717 | intergenic | GABRG2;CCNG1 | dist=1047522;dist=234510 |
| 3 month | rs146048121 | 6 | 141877818 | | G | A | 421 | 0.002375 | 3 | 1.082392071 | 0.232511623 | 4.655217038 | 3.24E-06 | 20.02143793 | 4.300860255 | intergenic | MIR4465;NMBR | dist=1193935;dist=196666 |
| 3 month | rs139493286 | 18 | 31236056 | | G | A | 421 | 0.003563 | 3 | 1.241116041 | 0.266612329 | 4.655133716 | 3.24E-06 | 17.46030927 | 3.750764282 | intergenic | DSC1;DSG1 | dist=73200;dist=82104 |
| 3 month | rs146442492 | 15 | 58689916 | 0.740549982 | C | T | 421 | 0.027316 | 24 | 3.06908876 | 0.65936392 | 4.654620409 | 3.25E-06 | 7.059258577 | 1.516613162 | intronic | ADAM10 | . |
| 3 month | rs181259864 | 7 | 97859511 | | C | A | 421 | 0.003563 | 3 | 1.263697226 | 0.271559968 | 4.653473908 | 3.26E-06 | 17.13608209 | 3.682427888 | ncRNA_intronic | CZ1P-ASNS | . |
| 3 month | rs150946694 | 22 | 46457283 | | T | C | 421 | 0.004751 | 4 | 1.413199859 | 0.303835874 | 4.651194872 | 3.30E-06 | 15.30824787 | 3.291250592 | intronic | CELSR1 | . |
| 3 month | rs181933850 | 5 | 92169830 | | A | G | 421 | 0.008314 | 7 | 1.957149723 | 0.420974257 | 4.64909597 | 3.33E-06 | 11.04365858 | 2.375442162 | intergenic | ARRDC3-AS1;NR2F1-AS1 | dist=749115;dist=1279415 |
| 3 month | rs113006316 | 2 | 74575233 | 0.706809998 | A | G | 421 | 0.002375 | 3 | 0.943984037 | 0.203082246 | 4.648284397 | 3.35E-06 | 22.88867923 | 4.924113346 | intronic | M1AP | . |
| 3 month | rs191006910 | 5 | 154836449 | | G | A | 421 | 0.003563 | 3 | 1.226165352 | 0.263817655 | 4.647775955 | 3.36E-06 | 17.61738031 | 3.790496891 | intronic | FAXDC2 | . |
| 3 month | rs176786 | 14 | 45813767 | | T | C | 421 | 0.32304 | 268 | 9.449422953 | 2.033680348 | 4.64646421 | 3.38E-06 | 2.284756409 | 0.49171936 | intergenic | LINC02303;LINC00871 | dist=98165;dist=250392 |
| 3 month | rs187047882 | 3 | 164567833 | | G | A | 421 | 0.003563 | 3 | 1.161853087 | 0.250171732 | 4.644222095 | 3.41E-06 | 18.56413613 | 3.997254169 | intergenic | MIR1263;LINC01324 | dist=396277;dist=146262 |
| 3 month | rs73057656 | 3 | 33899079 | 0.799390018 | A | G | 421 | 0.036817 | 33 | 3.638911487 | 0.783762258 | 4.642876649 | 3.44E-06 | 5.923832897 | 1.27589711 | intergenic | PDCD6IP;LOC101928135 | dist=29372;dist=976718 |
| 3 month | rs112557251 | 2 | 187510673 | 0.726930022 | T | C | 421 | 0.002375 | 3 | 1.043551353 | 0.224794821 | 4.642239298 | 3.45E-06 | 20.65100643 | 4.448501059 | intronic | TFPI | . |
| 3 month | rs190190051 | 5 | 92179668 | | G | A | 421 | 0.008314 | 7 | 1.947160785 | 0.419607459 | 4.640434158 | 3.48E-06 | 11.05898873 | 2.383179753 | intergenic | ARRDC3-AS1;NR2F1-AS1 | dist=758953;dist=1269577 |
| 3 month | rs180989936 | 1 | 193075048 | | A | G | 421 | 0.004751 | 4 | 1.253423246 | 0.270112952 | 4.640367065 | 3.48E-06 | 17.17935787 | 3.702154942 | intronic | RO60 | . |
| 3 month | rs188415494 | 8 | 25755782 | 0.735019982 | C | T | 421 | 0.003563 | 3 | 1.162251387 | 0.250533229 | 4.639110717 | 3.50E-06 | 18.51694779 | 3.991486498 | intergenic | CDCA2;EBF2 | dist=247865;dist=85943 |
| 3 month | rs1686289 | 14 | 45791779 | | G | A | 421 | 0.678147 | 277 | -9.537609426 | 2.056514006 | -4.637755637 | 3.52E-06 | -2.255153927 | 0.486259757 | intergenic | LINC02303;LINC00871 | dist=76177;dist=272380 |
| 3 month | rs76904423 | 12 | 100794966 | | G | A | 421 | 0.010689 | 10 | 2.190673587 | 0.472659243 | 4.634784191 | 3.57E-06 | 9.805762313 | 2.115689082 | UTR5 | ANO4 | NM_001286615:c.-1068200>0 |
| 3 month | rs183466664 | 12 | 26668754 | | A | G | 421 | 0.004751 | 4 | 1.341044738 | 0.289344304 | 4.634771518 | 3.57E-06 | 16.01818822 | 3.456090156 | intronic | ITPR2 | . |
| 3 month | rs999769259 | 4 | 61646247 | 0.702899992 | G | A | 421 | 0.003563 | 3 | 0.969956079 | 0.209381164 | 4.632489677 | 3.61E-06 | 22.12467252 | 4.775978806 | intronic | ADGRL3 | . |
| 3 month | rs553840536 | 16 | 25686574 | | A | G | 421 | 0.003563 | 3 | 1.093708141 | 0.236100775 | 4.632378455 | 3.61E-06 | 19.62034417 | 4.23547954 | intergenic | ZKSCAN2;HS3ST4 | dist=428729;dist=5385 |
| 3 month | rs183962155 | 4 | 21258020 | | A | C | 421 | 0.005938 | 6 | 1.557852883 | 0.336419694 | 4.630682783 | 3.64E-06 | 13.76460079 | 2.972477589 | intronic | KCNIP4 | . |

| | | | | | | | | | | | | | | | | | | |
|---|---|---|---|---|---|---|---|---|---|---|---|---|---|---|---|---|---|---|
| 3 month | rs138480898 | 1 | 184986525 | | C | T | 421 | 0.003563 | 3 | 1.261421145 | 0.272500077 | 4.629067106 | 3.67E-06 | 16.9873974 | 3.669723728 | intergenic | NIBAN1;LINC01633 | dist=12017;dist=15002 |
| 3 month | rs185158855 | 2 | 222785307 | 0.73951 | C | A | 421 | 0.004751 | 4 | 1.284200145 | 0.277572279 | 4.626543222 | 3.72E-06 | 16.66788644 | 3.602665239 | intergenic | MOGAT1;ACSL3 | dist=75377;dist=75728 |
| 3 month | rs180926150 | 1 | 102760770 | 0.792620003 | C | T | 421 | 0.002375 | 3 | 1.088498575 | 0.235408416 | 4.62387281 | 3.77E-06 | 19.64191801 | 4.247936485 | intergenic | OLFM3;COL11A1 | dist=763536;dist=115697 |
| 3 month | rs146728064 | 17 | 19362127 | | G | A | 421 | 0.007126 | 6 | 1.69020067 | 0.365674681 | 4.622143005 | 3.80E-06 | 12.64004111 | 2.734671147 | intronic | B9D1 | . |
| 3 month | rs111900874 | 7 | 89887355 | | G | A | 421 | 0.014252 | 12 | 2.536039096 | 0.548671755 | 4.622142612 | 3.80E-06 | 8.424240128 | 1.822583342 | ncRNA_intronic | STEAP2-AS1 | . |
| 3 month | rs574076561 | 7 | 49505151 | | A | G | 421 | 0.003563 | 3 | 1.086373837 | 0.235169216 | 4.619541003 | 3.85E-06 | 19.64347663 | 4.25225723 | intergenic | CDC14C;VWC2 | dist=577697;dist=268487 |
| 3 month | rs76554191 | 2 | 95301880 | | G | A | 421 | 0.04038 | 34 | 3.727552 | 0.807156314 | 4.618129021 | 3.87E-06 | 5.721480386 | 1.238917397 | intronic | KCNIP3 | . |
| 3 month | rs176783 | 14 | 45811710 | | A | G | 421 | 0.321853 | 267 | 9.35224324 | 2.025260115 | 4.617798558 | 3.88E-06 | 2.280101466 | 0.493763736 | intergenic | LINC02303;LINC00871 | dist=96108;dist=252449 |
| 3 month | rs140352232 | 2 | 107421656 | 0.797200024 | G | A | 421 | 0.002375 | 3 | 1.01264036 | 0.219291932 | 4.617772993 | 3.88E-06 | 21.05765113 | 4.560131294 | intergenic | MIR548AU;LINC01886 | dist=72131;dist=107763 |
| 3 month | rs139062456 | 8 | 13394482 | | C | T | 421 | 0.017815 | 15 | 2.721124769 | 0.589318868 | 4.617406495 | 3.89E-06 | 7.835158086 | 1.696874229 | intronic | DLC1 | . |
| 3 month | rs558614420 | 15 | 41518672 | 0.745689988 | C | T | 421 | 0.005938 | 6 | 1.524477016 | 0.330280733 | 4.615700711 | 3.92E-06 | 13.97508315 | 3.02772732 | intronic | RPAP1 | . |
| 3 month | rs12266995 | 10 | 24563854 | | T | C | 421 | 0.030879 | 26 | 3.568935699 | 0.773323681 | 4.615060663 | 3.93E-06 | 5.967825345 | 1.293119589 | intergenic | KIAA1217;ARHGAP21 | dist=16006;dist=19760 |
| 3 month | rs192750513 | 7 | 97948518 | | A | G | 421 | 0.003563 | 3 | 1.238714279 | 0.26844613 | 4.614386809 | 3.94E-06 | 17.18924692 | 3.725142178 | ncRNA_intronic | CZ1P-ASNS | . |
| 3 month | rs56302696 | 12 | 47899047 | 0.778989971 | G | A | 421 | 0.002375 | 3 | 1.068148373 | 0.231704225 | 4.60996503 | 4.03E-06 | 19.89590409 | 4.315847075 | intronic | VDR | . |
| 3 month | rs151015676 | 5 | 177963936 | 0.785820007 | T | G | 421 | 0.002375 | 3 | 1.054373875 | 0.228793957 | 4.608399149 | 4.06E-06 | 20.14213671 | 4.370744819 | intergenic | LOC100128340;PROP1 | dist=4136;dist=28299 |
| 3 month | rs12315614 | 12 | 64527177 | | C | A | 421 | 0.07601 | 64 | 5.614667617 | 1.219220913 | 4.605127386 | 4.12E-06 | 3.777106623 | 0.820195905 | intergenic | TBK1;RASSF3 | dist=25064;dist=83318 |
| 3 month | rs72837643 | 5 | 170761951 | 0.782980025 | T | C | 421 | 0.003563 | 4 | 1.251913633 | 0.271928683 | 4.60383075 | 4.15E-06 | 16.93028738 | 3.677434793 | intergenic | KCNIP1;GABRP | dist=25319;dist=21768 |
| 3 month | rs428110 | 14 | 45825457 | | A | C | 421 | 0.317102 | 265 | 9.275749143 | 2.014875505 | 4.603633881 | 4.15E-06 | 2.284822992 | 0.49630858 | intergenic | LINC02303;LINC00871 | dist=109855;dist=238702 |
| 3 month | rs111391231 | 7 | 89867731 | | T | C | 421 | 0.014252 | 12 | 2.546596314 | 0.553340001 | 4.602227036 | 4.18E-06 | 8.317177547 | 1.807207138 | intergenic | ZNF804B;STEAP2-AS1 | dist=529203;dist=14622 |
| 3 month | rs180765647 | 13 | 113724338 | | G | T | 421 | 0.002375 | 3 | 1.05190176 | 0.228624832 | 4.600995217 | 4.20E-06 | 20.12465213 | 4.373978059 | intronic | GRK1 | . |
| 3 month | rs568658857 | 12 | 49460215 | | G | A | 421 | 0.005938 | 5 | 1.486911933 | 0.32318174 | 4.600853791 | 4.21E-06 | 14.23611926 | 3.094234224 | intronic | SPATS2 | . |
| 3 month | rs182531466 | 5 | 92234256 | | C | A | 421 | 0.007126 | 6 | 1.668434681 | 0.362732486 | 4.599628498 | 4.23E-06 | 12.68049781 | 2.756852606 | intergenic | ARRDC3-AS1;NR2F1-AS1 | dist=813541;dist=1214989 |
| 3 month | rs181812512 | 11 | 66898258 | | C | T | 421 | 0.003563 | 3 | 1.08407707 | 0.235726848 | 4.598869755 | 4.25E-06 | 19.50931683 | 4.242198164 | intronic | PC | . |
| 3 month | rs556680896 | 13 | 100950161 | | C | T | 421 | 0.003563 | 3 | 1.24171677 | 0.270060937 | 4.597913279 | 4.27E-06 | 17.0254659 | 3.702867989 | ncRNA_intronic | NALCN-AS1 | . |
| 3 month | rs142928734 | 13 | 100948828 | | G | A | 421 | 0.003563 | 3 | 1.242180957 | 0.270237412 | 4.59662838 | 4.29E-06 | 17.00959296 | 3.700449886 | ncRNA_intronic | NALCN-AS1 | . |
| 3 month | rs143287889 | 4 | 35570658 | 0.79956001 | C | T | 421 | 0.002375 | 3 | 1.058108262 | 0.230199399 | 4.596485777 | 4.30E-06 | 19.96741 | 4.34406 | intergenic | LINC02484;ARAP2 | dist=1300911;dist=495346 |
| 3 month | rs543844012 | 9 | 30107025 | | C | T | 421 | 0.003563 | 3 | 1.164274175 | 0.253510467 | 4.592607905 | 4.38E-06 | 18.11604845 | 3.944610301 | intergenic | LINGO2;LINC01242 | dist=893424;dist=281910 |
| 3 month | rs566018180 | 10 | 84995296 | 0.773450017 | C | T | 421 | 0.003563 | 3 | 1.136146605 | 0.247392568 | 4.592484792 | 4.38E-06 | 18.56355199 | 4.04215862 | intergenic | CCSER2;LINC01519 | dist=476775;dist=198125 |
| 3 month | rs147171192 | 4 | 88214436 | | A | G | 421 | 0.007126 | 5 | 1.584612591 | 0.345114993 | 4.591549547 | 4.40E-06 | 13.30440472 | 2.89758492 | intronic | ABCG2 | . |
| 3 month | rs2876414 | 20 | 15833059 | | G | T | 421 | 0.022565 | 19 | 3.090964158 | 0.673260548 | 4.591037107 | 4.41E-06 | 6.819109065 | 1.485309072 | intronic | MACROD2 | . |
| 3 month | rs137873790 | 5 | 97751337 | | A | G | 421 | 0.013064 | 11 | 2.280553097 | 0.497276469 | 4.586086892 | 4.52E-06 | 9.222408811 | 2.010953789 | intergenic | LINC01340;LINC02234 | dist=80286;dist=89421 |
| 3 month | rs752259256 | 10 | 103165562 | 0.747950017 | T | C | 421 | 0.003563 | 3 | 1.029573618 | 0.224511445 | 4.58584023 | 4.52E-06 | 20.42586391 | 4.454115907 | intronic | NT5C2 | . |
| 3 month | rs185771987 | 5 | 73989664 | 0.738900006 | T | C | 421 | 0.003563 | 3 | 1.153019969 | 0.251461612 | 4.585272329 | 4.53E-06 | 18.23448239 | 3.976750144 | intergenic | ARHGEF28;LINC01335 | dist=47671;dist=316746 |
| 3 month | rs557092705 | 20 | 35601989 | | C | T | 421 | 0.003563 | 3 | 1.079320791 | 0.235404492 | 4.584962596 | 4.54E-06 | 19.47695457 | 4.248007298 | ncRNA_exonic | FER1L4 | . |
| 3 month | rs140642138 | 15 | 41832967 | | G | A | 421 | 0.005938 | 6 | 1.547446255 | 0.337525343 | 4.584681679 | 4.55E-06 | 13.58322205 | 2.962740492 | intronic | JMJD7;JMJD7-PLA2G4B | . |
| 3 month | rs139877408 | 2 | 128848699 | 0.79351002 | A | G | 421 | 0.002375 | 3 | 1.034497038 | 0.225706887 | 4.583364964 | 4.58E-06 | 20.30671294 | 4.430524974 | intergenic | HS6ST1;LOC101927881 | dist=529831;dist=15901 |
| 3 month | rs2327968 | 20 | 15832846 | | C | T | 421 | 0.024941 | 21 | 3.410130247 | 0.744098505 | 4.582901624 | 4.59E-06 | 6.158998564 | 1.343908089 | intronic | MACROD2 | . |
| 3 month | rs191986449 | 5 | 25745578 | | C | T | 421 | 0.004751 | 3 | 1.135135845 | 0.247721071 | 4.582314459 | 4.60E-06 | 18.49787925 | 4.036798308 | intergenic | LINC02211;CDH9 | dist=443298;dist=1135019 |
| 3 month | rs11690187 | 2 | 67338777 | | A | C | 421 | 0.005938 | 5 | 1.539986371 | 0.336119518 | 4.581663041 | 4.61E-06 | 13.63105325 | 2.975132201 | intergenic | LINC01828;ETAA1 | dist=49533;dist=58556 |
| 3 month | rs118040657 | 10 | 3430654 | 0.777450025 | C | T | 421 | 0.008314 | 8 | 1.732520372 | 0.378165204 | 4.581384943 | 4.62E-06 | 12.11477125 | 2.644346939 | ncRNA_intronic | LOC105376360 | . |
| 3 month | rs138109686 | 15 | 41759244 | | A | G | 421 | 0.005938 | 6 | 1.543778108 | 0.336970687 | 4.581342434 | 4.62E-06 | 13.59567051 | 2.96761718 | intronic | MGA | . |
| 3 month | rs563167766 | 1 | 102400809 | | G | A | 421 | 0.002375 | 3 | 1.07877255 | 0.235715413 | 1.07877255 | 4.73E-06 | 19.41573898 | 4.242403955 | intergenic | OLFM3;COL11A1 | dist=403575;dist=475658 |
| 3 month | rs545550279 | 8 | 114540200 | | G | T | 421 | 0.003563 | 3 | 1.091458604 | 0.238542228 | 4.57553622 | 4.75E-06 | 19.18124208 | 4.192129875 | intergenic | CSMD3;TRPS1 | dist=1103261;dist=868295 |
| 3 month | rs1498183 | 8 | 54804345 | | C | T | 421 | 0.394299 | 332 | 10.33728121 | 2.259624491 | 4.574778355 | 4.77E-06 | 2.024574602 | 0.442551408 | intronic | RP1 | . |
| 3 month | rs375790303 | 1 | 184561348 | | G | A | 421 | 0.008314 | 7 | 1.831959409 | 0.400449797 | 4.574754241 | 4.77E-06 | 11.42403934 | 2.497191924 | intronic | C1orf21 | . |
| 3 month | rs1396896 | 8 | 54782750 | | A | G | 421 | 0.397862 | 334 | 10.46979793 | 2.288829589 | 4.574302074 | 4.78E-06 | 1.998533266 | 0.436904523 | intronic | RP1 | . |
| 3 month | rs7843693 | 8 | 54779552 | | G | A | 421 | 0.397862 | 334 | 10.46760966 | 2.288570748 | 4.573863258 | 4.79E-06 | 1.998567561 | 0.436953938 | intronic | RP1 | . |
| 3 month | rs1391462 | 8 | 54787221 | | C | A | 421 | 0.397862 | 334 | 10.46683006 | 2.288537601 | 4.573588854 | 4.79E-06 | 1.998476605 | 0.436960267 | intronic | RP1 | . |
| 3 month | rs150539922 | 21 | 41856807 | | T | C | 421 | 0.003563 | 3 | 1.254834519 | 0.274365448 | 4.57358799 | 4.79E-06 | 16.66969372 | 3.64477381 | intronic | PRDM15 | . |
| 3 month | rs145896760 | 15 | 41827024 | | G | A | 421 | 0.005938 | 6 | 1.564221999 | 0.342059488 | 4.572953105 | 4.81E-06 | 13.36888249 | 2.923468092 | UTR3 | MAPKBP1 | NM_014994:c.*15880>0;NM_ |
| 3 month | rs181415102 | 4 | 111768110 | 0.716539979 | T | C | 421 | 0.003563 | 4 | 1.144945045 | 0.250383121 | 4.572772469 | 4.81E-06 | 18.26310193 | 3.993879436 | intergenic | MIR297;FAM241A | dist=907463;dist=377344 |
| 3 month | rs6080 | 15 | 58545734 | 0.79065001 | C | A | 421 | 0.043943 | 36 | 3.688174426 | 0.80689065 | 4.570847892 | 4.86E-06 | 5.664767454 | 1.239325304 | intronic | LIPC | . |
| 3 month | rs138249376 | 10 | 61756071 | | T | G | 421 | 0.004751 | 4 | 1.271516185 | 0.278189815 | 4.57067843 | 4.86E-06 | 16.43007109 | 3.594667912 | intronic | CABCOCO1 | . |
| 3 month | rs529345909 | 11 | 67343381 | | A | G | 421 | 0.003563 | 3 | 1.116845445 | 0.244375672 | 4.570198967 | 4.87E-06 | 18.70153001 | 4.092060355 | ncRNA_intronic | LOC100130987 | . |
| 3 month | rs191930622 | 12 | 47890872 | 0.790120006 | G | A | 421 | 0.002375 | 3 | 1.061255347 | 0.232228501 | 4.569875551 | 4.88E-06 | 19.67835791 | 4.306103677 | intronic | VDR | . |
| 3 month | rs145116559 | 4 | 111756440 | 0.70095998 | T | C | 421 | 0.003563 | 4 | 1.144113716 | 0.250422362 | 4.568736218 | 4.91E-06 | 18.24412236 | 3.993253602 | intergenic | MIR297;FAM241A | dist=895793;dist=389014 |
| 3 month | rs12678939 | 8 | 54792461 | | A | G | 421 | 0.394299 | 331 | 10.32905196 | 2.261417548 | 4.567512076 | 4.94E-06 | 2.019756183 | 0.442200513 | intronic | RP1 | . |
| 3 month | rs73586304 | 6 | 142518288 | | C | T | 421 | 0.003563 | 3 | 1.14329832 | 0.250357885 | 4.566655922 | 4.96E-06 | 18.24051164 | 3.99428202 | intergenic | ADGRG6;LOC153910 | dist=72022;dist=8167 |
| 3 month | rs113767990 | 14 | 81251219 | | G | A | 421 | 0.004751 | 4 | 1.429622227 | 0.313123218 | 4.565685793 | 4.98E-06 | 14.58111547 | 3.193630952 | intergenic | LOC101928504;STON2 | dist=27839;dist=9431 |

### Supplementary Table S1. GWAS Results SNPs Indianapolis-1 Discovery Cohort
### Overlap of 12 month and 3 month quantitative traits (QT)

**Headers**

QT: quantitiative trait; rsid: reference SNP cluster ID, chr: chromosome; pos_38, position of SNP on GRCh38 reference panel; REF and ALT, reference allele and alternate allele;
caf.12m: common (major) allele frequency, 12 month QT; Score.12m: p-values from Score test, 12 month QT; Score.SE.12m; Score.pval.12m
caf.3m: common (major) allele frequency, 3 month QT; Score.3m: p-values from Score test, 3 month QT; Score.SE.3m; Score.pval.3m

| rsid | chr | pos_38 | REF | ALT | | caf.12m | Score.12m | Score.SE.12m | Score.pval.12m | | caf.3m | Score.3m | Score.SE.3m | Score.pval.3m |
|---|---|---|---|---|---|---|---|---|---|---|---|---|---|---|
| rs76098744 | 1 | 111806750 | C | T | | 0.01594533 | 2.284999945 | 0.488674498 | 2.93E-06 | | 0.016627078 | 2.811615503 | 0.579024995 | 1.20E-06 |
| rs74683551 | 1 | 111811796 | G | A | | 0.01594533 | 2.279782265 | 0.488661396 | 3.08E-06 | | 0.016627078 | 2.810351968 | 0.579019309 | 1.21E-06 |
| rs536781978 | 2 | 29510815 | A | G | | 0.003416856 | 1.090050785 | 0.230277801 | 2.21E-06 | | 0.003562945 | 1.282866581 | 0.270080884 | 2.03E-06 |
| rs568321148 | 2 | 29647482 | T | G | | 0.003416856 | 1.087769414 | 0.22901495 | 2.04E-06 | | 0.003562945 | 1.279867135 | 0.268808164 | 1.92E-06 |
| rs148248743 | 3 | 136415753 | C | T | | 0.002277904 | 0.909706546 | 0.183031941 | 6.69E-07 | | 0.002375297 | 1.069833829 | 0.216429281 | 7.69E-07 |
| rs189709453 | 3 | 177856520 | G | A | | 0.003416856 | 0.994705176 | 0.207058897 | 1.56E-06 | | 0.003562945 | 1.314463975 | 0.246356303 | 9.52E-08 |
| rs186767531 | 3 | 177921989 | T | C | | 0.003416856 | 0.994608422 | 0.207613329 | 1.66E-06 | | 0.003562945 | 1.293967205 | 0.247047316 | 1.63E-07 |
| rs182868205 | 3 | 177994660 | C | T | | 0.003416856 | 1.020592047 | 0.21132014 | 1.37E-06 | | 0.003562945 | 1.323927501 | 0.251356167 | 1.39E-07 |
| rs113063005 | 4 | 23507444 | T | C | | 0.006833713 | 1.445039344 | 0.294886773 | 9.57E-07 | | 0.005938242 | 2.130262689 | 0.316937024 | 1.80E-11 |
| rs189912648 | 5 | 135429749 | C | T | | 0.003416856 | 1.088415104 | 0.234100744 | 3.33E-06 | | 0.003562945 | 1.303529678 | 0.277126356 | 2.55E-06 |
| rs190251199 | 14 | 105124240 | T | C | | 0.003416856 | 0.998973364 | 0.210064697 | 1.98E-06 | | 0.003562945 | 1.240668633 | 0.248987137 | 6.27E-07 |
