## Supplementary material for "Pharmacogenomics of steroid-induced ocular hypertension: relationship to high-tension glaucomas and new pathophysiologic insight": Suppl Table S2

**Supplementary Table S2. GWAS Results Risk Loci Indianapolis-1 Discovery Cohort**
**Merged loci count and quantitative trait (QT) overlap**

**Notes**
Top SNPs identified with the 12M and 3M QTs are merged, then clustered into risk loci by sorting by chromosomal position, then by chromosome
SNPs duplicated between 12 month and 3 month QT are boxed. Loci that cluster SNPs from both the 12 and 3 month QTs are boxed and highlighted in blue.
SNPs of genome-wide significance are highlighted in gray (column rsid) as are their p-values (column Score.pval).
Risk loci into which multiple SNPs cluster are boxed (column Gene.refGene) and those containing SNPs of genome-wide significance are shaded in gray
Count risk loci: columns adding up all risk loci broken down by p-value and QT, totals at bottom; 12+3 rep: risk loci clustering both 12M+3M QT SNPs that are duplicates;
12+3 diff: risk loci clustering both 12M+3M QT SNPs for which the 2 QTs identify different SNPs
**Headers**
QT: quantitative trait; rsid: reference SNP cluster ID, chr: chromosome; pos_38: position of SNP on GRCh38 reference panel; Score.pval: p-value; Est: effect size (mm Hg);
Func.refGene: SNP location with respect to nearest gene(s); Gene.refGene: nearest gene upstream and downstream; GeneDetail.refGene: distance to nearest gene upstream and downstream

| | | | | | | | | | | Count risk loci | | | | | |
|---|---|---|---|---|---|---|---|---|---|---|---|---|---|---|---|
| QT | rsid | chr | pos_38 | MAC | Score.pval | Est | Func.refGene | Gene.refGene | GeneDetail.refGene | 5E-08 12M | 5E-08 3M | 5E-06 12M | 5E-06 3M | 12+3 rep | 12+3 diff |
| 3M | rs115348382 | 1 | 9595845 | 3 | 2.20695702237544e-06 | 18.2107495 | intronic | TMEM201 | . | | | | 1 | | |
| 3M | rs186532456 | 1 | 18621834 | 3 | 9.60875608166652e-07 | 20.1730343 | intergenic | KLHDC7A;PAX7 | dist=135848;dist=9012 | | | | 1 | | |
| 3M | rs562032622 | 1 | 18632839 | 3 | 1.12975575994446e-06 | 21.0267115 | intronic | PAX7 | . | | | | | | |
| 12M | rs35145334 | 1 | 23138629 | 14 | 2.09882278975154e-06 | 10.6552656 | intronic | LUZP1 | . | | | 1 | | | |
| 3M | rs2365739 | 1 | 62018790 | 18 | 1.81260643542989e-06 | 6.86815917 | intronic | PATJ | . | | | | 1 | | |
| 12M | rs17127656 | 1 | 65477788 | 49 | 1.63996682822861e-06 | 5.07046538 | intronic | LEPR | . | | | 1 | | | |
| 12M | rs7518849 | 1 | 65483108 | 49 | 1.67668772877101e-06 | 5.07456956 | intronic | LEPR | . | | | | | | |
| 12M | rs11579567 | 1 | 65491458 | 49 | 2.1300143111827e-06 | 5.02973793 | intronic | LEPR | . | | | | | | |
| 12M | rs7534177 | 1 | 65500037 | 49 | 2.85450525546462e-06 | 4.96123378 | intronic | LEPR | . | | | | | | |
| 3M | rs149493615 | 1 | 79414404 | 8 | 1.38822331670427e-06 | 11.750632 | intergenic | ADGRL4;LINC01781 | dist=407674;dist=1121351 | | | | 1 | | |
| 3M | rs143811231 | 1 | 79502446 | 8 | 1.738654939087e-06 | 11.5747409 | intergenic | ADGRL4;LINC01781 | dist=495716;dist=1033309 | | | | | | |
| 12M | rs139390630 | 1 | 84400513 | 13 | 2.86874207705929e-06 | 11.6975971 | intronic | DNASE2B | . | | | 1 | | | |
| 12M | rs114067899 | 1 | 84725361 | 7 | 2.17218533160816e-06 | 13.7760673 | intergenic | SSX2IP;LPAR3 | dist=34893;dist=86241 | | | 1 | | | |
| 3M | rs34270375 | 1 | 88905019 | 21 | 9.34314238699532e-07 | 7.71147017 | intergenic | GTF2B;KYAT3 | dist=13452;dist=30754 | | | | 1 | | |
| 3M | rs187518659 | 1 | 98990189 | 8 | 2.15952482477188e-06 | 11.6819623 | intronic | PLPPR5 | . | | | | 1 | | |
| 3M | rs140420703 | 1 | 102337928 | 5 | 1.91423564672595e-06 | 16.487654 | intergenic | OLFM3;COL11A1 | dist=340694;dist=538539 | | | | | | |
| 3M | rs563167766 | 1 | 102400809 | 3 | 4.72618849175033e-06 | 19.415739 | intergenic | OLFM3;COL11A1 | dist=403575;dist=475658 | | | | | | |
| 3M | rs77180278 | 1 | 102496326 | 24 | 9.33877360643494e-08 | 7.33619996 | intergenic | OLFM3;COL11A1 | dist=499092;dist=380141 | | | | | | |
| 3M | rs112351653 | 1 | 102754804 | 24 | 2.54962936189828e-08 | 7.42386947 | intergenic | OLFM3;COL11A1 | dist=757570;dist=121663 | | 1 | | | | |
| 3M | rs180926150 | 1 | 102760770 | 3 | 3.76640865110522e-06 | 19.641918 | intergenic | OLFM3;COL11A1 | dist=763536;dist=115697 | | | | | | |
| 3M | rs114413507 | 1 | 102953612 | 24 | 2.57350301189632e-08 | 7.42216029 | intronic | COL11A1 | . | | | | | | |
| 3M | rs116672066 | 1 | 103007360 | 23 | 4.45932234807969e-09 | 8.04769476 | intronic | COL11A1 | . | | | | | | |
| 3M | rs111928960 | 1 | 103168079 | 22 | 3.65396910662882e-09 | 8.60257208 | intergenic | COL11A1;LOC101928436 | dist=59557;dist=325967 | | | | | | |
| 3M | rs113221952 | 1 | 103288418 | 19 | 9.173975220783e-07 | 7.92350704 | intergenic | COL11A1;LOC101928436 | dist=179896;dist=205628 | | | | | | |
| 3M | rs1856085 | 1 | 103571923 | 3 | 4.63628412408432e-07 | 21.51144 | intronic | AMY2B | . | | | | 1 | | |
| 3M | rs143597860 | 1 | 103614521 | 3 | 4.52300265269257e-07 | 21.5748154 | intergenic | AMY2B;AMY2A | dist=34987;dist=2811 | | | | | | |
| 3M | rs144541665 | 1 | 103768107 | 3 | 3.71844012409788e-07 | 21.8734611 | intergenic | AMY1C;LOC100129138 | dist=9415;dist=304916 | | | | | | |
| 3M | rs76098744 | 1 | 111806750 | 14 | 1.19916376071606e-06 | 8.38612452 | intronic | KCND3 | . | | | | 1 | 1 | |
| 12M | rs76098744 | 1 | 111806750 | 14 | 2.92647184366499E-06 | 9.5685657 | intronic | KCND3 | . | | | | | | |
| 3M | rs74683551 | 1 | 111811796 | 14 | 1.2121497054257e-06 | 8.38252043 | intronic | KCND3 | . | | | | | | |
| 12M | rs74683551 | 1 | 111811796 | 14 | 3.08074322019196E-06 | 9.54722832 | intronic | KCND3 | | | | | | | |
| 3M | rs76617932 | 1 | 180961288 | 9 | 2.78866978177701e-06 | 10.5449608 | intergenic | KIAA1614;STX6 | dist=6401;dist=11426 | | | | 1 | | |
| 3M | rs183180157 | 1 | 181277985 | 7 | 2.7248136592675E-06 | 12.61386 | intergenic | LINC01699;CACNA1E | dist=39381;dist=205532 | | | | | | |
| 3M | rs375790303 | 1 | 184561348 | 7 | 4.76779314562933e-06 | 11.4240393 | intronic | C1orf21 | . | | | | 1 | | |
| 3M | rs138480898 | 1 | 184986525 | 3 | 3.67316732025395e-06 | 16.9873974 | intergenic | NIBAN1;LINC01633 | dist=12017;dist=15002 | | | | | | |

| | | | | | | | | | | | | | | |
|---|---|---|---|---|---|---|---|---|---|---|---|---|---|---|
| 3M | rs145766563 | 1 | 185160370 | 4 | 1.9043145410504e-06 | 16.8682102 | intronic | SWT1 | . | | | | | |
| 3M | rs147032554 | 1 | 186179732 | 3 | 6.67557909700165e-07 | 18.162609 | intronic | HMCN1 | . | | | | | |
| 3M | rs180989936 | 1 | 193075048 | 4 | 3.47790847768381E-06 | 17.1793579 | intronic | RO60 | . | | | 1 | | |
| 12M | rs145804766 | 1 | 202153371 | 9 | 1.43417032447652e-06 | 12.5864465 | intronic | PTPN7 | . | | 1 | | | |
| 3M | rs10494861 | 1 | 205362746 | 3 | 2.48126631931009e-06 | 19.1820498 | intergenic | KLHDC8A;LEMD1 | dist=5656;dist=10506 | | | 1 | | |
| 12M | rs147869916 | 1 | 227119951 | 5 | 3.36060694276516e-06 | 17.5964551 | intronic | CDC42BPA | . | | 1 | | | |
| 3M | rs2274996 | 1 | 229668791 | 35 | 6.73601821695854e-07 | 5.58527433 | intergenic | URB2;LINC01682 | dist=8591;dist=206759 | | | 1 | | |
| 3M | rs2274997 | 1 | 229668899 | 35 | 6.64244186267036e-07 | 5.58731228 | intergenic | URB2;LINC01682 | dist=8699;dist=206651 | | | | | |
| 3M | rs2891865 | 1 | 229670621 | 35 | 6.80112526815562e-07 | 5.58181376 | intergenic | URB2;LINC01682 | dist=10421;dist=204929 | | | | | |
| 3M | rs2385790 | 1 | 229671745 | 35 | 6.81207031237396e-07 | 5.58224822 | intergenic | URB2;LINC01682 | dist=11545;dist=203805 | | | | | |
| 3M | rs12024557 | 1 | 229676610 | 35 | 6.90833131537333e-07 | 5.56930969 | intergenic | URB2;LINC01682 | dist=16410;dist=198940 | | | | | |
| 3M | rs4562666 | 1 | 229689023 | 36 | 7.35045143592774e-07 | 5.51922599 | intergenic | URB2;LINC01682 | dist=28823;dist=186527 | | | | | |
| 3M | rs12036586 | 1 | 229690631 | 40 | 1.08649183321501e-06 | 5.22212176 | intergenic | URB2;LINC01682 | dist=30431;dist=184919 | | | | | |
| 3M | rs16850124 | 1 | 229695584 | 37 | 2.19845550423288e-06 | 5.18095662 | intergenic | URB2;LINC01682 | dist=35384;dist=179966 | | | | | |
| 3M | rs12045643 | 1 | 229698303 | 36 | 7.37332600021845e-07 | 5.48149152 | intergenic | URB2;LINC01682 | dist=38103;dist=177247 | | | | | |
| 3M | rs559559983 | 1 | 246813830 | 5 | 1.8701710647004e-06 | 13.811132 | intergenic | LINC01341;AHCTF1 | dist=22344;dist=25268 | | | 1 | | |
| 3M | rs550763536 | 2 | 7312216 | 5 | 3.04128022440287e-06 | 14.4871647 | intergenic | LOC101929452;LOC100506274 | dist=234336;dist=109045 | | | 1 | | |
| 12M | rs76220567 | 2 | 12919819 | 4 | 2.07255893928856e-06 | 20.6828868 | intergenic | TRIB2;LOC100506474 | dist=177087;dist=46963 | | 1 | | | |
| 3M | rs558553658 | 2 | 15542600 | 3 | 2.00500271573415e-07 | 17.1099107 | intronic | NBAS | . | | | 1 | | |
| 3M | rs536781978 | 2 | 29510815 | 3 | 2.20535447064362e-06 | 20.5561882 | intronic | ALK | . | | | 1 | 1 | |
| 12M | rs536781978 | 2 | 29510815 | 3 | 2.20535447064362E-06 | 20.5561882 | intronic | ALK | | | | | | |
| 3M | rs568321148 | 2 | 29647482 | 3 | 2.03643401922156e-06 | 20.7400203 | intronic | ALK | . | | | | | |
| 12M | rs568321148 | 2 | 29647482 | 3 | 2.03643401922156E-06 | 20.7400203 | intronic | ALK | | | | | | |
| 12M | rs186792608 | 2 | 42869559 | 3 | 1.57557414570447e-07 | 22.4512499 | intergenic | HAAO;LINC01819 | dist=76976;dist=158293 | | 1 | | | |
| 3M | rs76777840 | 2 | 48085811 | 5 | 7.02623491851785e-07 | 14.3500784 | intergenic | FBXO11;FOXN2 | dist=179313;dist=228488 | | | 1 | | |
| 3M | rs145080832 | 2 | 48246004 | 4 | 1.37120287877638e-07 | 17.1618477 | intergenic | FBXO11;FOXN2 | dist=339506;dist=68295 | | | | | |
| 3M | rs184220112 | 2 | 48404604 | 4 | 3.11727344916156e-07 | 16.4879043 | intergenic | FOXN2;PPP1R21 | dist=25309;dist=36162 | | | | | |
| 3M | rs189890455 | 2 | 48428258 | 5 | 4.1971806049668e-07 | 15.1467391 | intergenic | FOXN2;PPP1R21 | dist=48963;dist=12508 | | | | | |
| 3M | rs181193202 | 2 | 52300216 | 3 | 1.62222587830176e-07 | 19.6312088 | ncRNA_intronic | LOC730100 | . | 1 | | | | |
| 3M | rs183378658 | 2 | 52336233 | 3 | 1.16662586966659e-07 | 19.7446169 | ncRNA_intronic | LOC730100 | . | | | | | |
| 3M | rs190193113 | 2 | 52858640 | 3 | 1.13301212531008e-07 | 19.9533602 | intergenic | MIR4431;ASB3 | dist=156025;dist=811339 | | | | | |
| 3M | rs187520610 | 2 | 53132903 | 4 | 3.93234514941546e-09 | 18.8010396 | intergenic | MIR4431;ASB3 | dist=430288;dist=537076 | | | | | |
| 3M | rs149421869 | 2 | 53256291 | 8 | 2.05046156638639e-06 | 11.7382186 | intergenic | MIR4431;ASB3 | dist=553676;dist=413688 | | | | | |
| 3M | rs146479102 | 2 | 65598625 | 5 | 2.18341304839895e-06 | 15.4846768 | intergenic | SPRED2;MIR4778 | dist=166026;dist=759622 | | | 1 | | |
| 3M | rs528288879 | 2 | 65648796 | 6 | 5.421960882917e-07 | 14.7878085 | intergenic | SPRED2;MIR4778 | dist=216197;dist=709451 | | | | | |
| 3M | rs11690187 | 2 | 67338777 | 5 | 4.61292753502872e-06 | 13.6310532 | intergenic | LINC01828;ETAA1 | dist=49533;dist=58556 | | 1 | 1 | | 1 |
| 3M | rs151272830 | 2 | 67455592 | 6 | 2.85197668301622e-06 | 13.3323479 | intergenic | ETAA1;LINC01812 | dist=43503;dist=340462 | | | | | |
| 3M | rs186142189 | 2 | 67475575 | 6 | 2.69645566407293e-06 | 12.7361927 | intergenic | ETAA1;LINC01812 | dist=63486;dist=320479 | | | | | |
| 12M | rs2902021 | 2 | 67598553 | 71 | 3.30567771883221e-06 | 4.55991411 | intergenic | ETAA1;LINC01812 | dist=186464;dist=197501 | | | | | |
| 12M | rs75082290 | 2 | 67604021 | 69 | 1.699715195267e-06 | 4.69395387 | intergenic | ETAA1;LINC01812 | dist=191932;dist=192033 | | | | | |
| 12M | rs191298981 | 2 | 67882914 | 3 | 7.69651662192921e-07 | 24.0078714 | intergenic | LINC01812;C1D | dist=57352;dist=158216 | | 1 | | | |
| 12M | rs113537164 | 2 | 68007798 | 4 | 1.017020860936e-06 | 20.3890413 | intergenic | LINC01812;C1D | dist=182236;dist=33332 | | | | | |
| 12M | rs113154814 | 2 | 68062365 | 5 | 1.66384008676077e-07 | 19.8310188 | intronic | C1D | . | | | | | |
| 3M | rs184200893 | 2 | 69033781 | 3 | 2.24020739771759E-06 | 19.9580356 | intronic | ANTXR1 | . | | | 1 | | |
| 3M | rs111927235 | 2 | 74256827 | 4 | 8.4250114725101e-07 | 17.0468955 | intronic | SLC4A5 | . | | | 1 | | |
| 3M | rs111838310 | 2 | 74446364 | 5 | 3.26261179053175e-07 | 17.7963938 | intergenic | RTKN;INO80B-WBP1 | dist=4427;dist=8659 | | | 1 | | |
| 3M | rs112983626 | 2 | 74470023 | 4 | 3.34719559082237e-07 | 17.7848079 | intergenic | MOGS;MRPL53 | dist=4641;dist=1959 | | | 1 | | |
| 3M | rs113006316 | 2 | 74575233 | 3 | 3.34707225210704e-06 | 22.8886792 | intronic | M1AP | . | | | 1 | | |
| 3M | rs17746486 | 2 | 95056864 | 29 | 1.47154878644706e-06 | 6.90851358 | intergenic | MAL;MRPS5 | dist=2872;dist=28507 | | | 1 | | |
| 3M | rs76554191 | 2 | 95301880 | 34 | 3.87215481208204e-06 | 5.72148039 | intronic | KCNIP3 | . | | | 1 | | |
| 3M | rs140352232 | 2 | 107421656 | 3 | 3.8788022972212e-06 | 21.0576511 | intergenic | MIR548AU;LINC01886 | dist=72131;dist=107763 | | | 1 | | |
| 12M | rs146919974 | 2 | 114495286 | 5 | 7.91246832646777e-07 | 19.7280466 | intronic | DPP10 | . | | 1 | | | |
| 3M | rs116189766 | 2 | 125636287 | 3 | 1.17473937061964e-06 | 21.190764 | intergenic | CNTNAP5;LINC01941 | dist=715069;dist=473813 | | | 1 | | |

| | | | | | | | | | | | | | | |
|---|---|---|---|---|---|---|---|---|---|---|---|---|---|---|
| 3M | rs139877408 | 2 | 128848699 | 3 | 4.57552408670287e-06 | 20.3067129 | intergenic | HS6ST1;LOC101927881 | dist=529831;dist=15901 | | | 1 | | |
| 3M | rs142894171 | 2 | 150581757 | 3 | 1.40524666465064e-06 | 18.0710394 | intergenic | LINC01920;LINC02612 | dist=9536;dist=47140 | | | 1 | | |
| 3M | rs541508507 | 2 | 169166755 | 4 | 2.32254126928461e-06 | 15.4630033 | intronic | LRP2 | . | | | 1 | | |
| 3M | rs142549310 | 2 | 169173996 | 4 | 2.69758962759472e-06 | 15.2501713 | exonic | LRP2 | . | | | | | |
| 3M | rs556293455 | 2 | 176085692 | 3 | 1.27371002978549e-06 | 14.3860463 | intergenic | EVX2;HOXD13 | dist=1730;dist=7029 | | | 1 | | |
| 3M | rs184098071 | 2 | 176251692 | 3 | 1.15315934031841e-06 | 13.947724 | intergenic | HOXD1;MTX2 | dist=60785;dist=17750 | | 1 | 1 | | 1 |
| 3M | rs79935606 | 2 | 176473907 | 5 | 2.55448017015286e-06 | 16.3396833 | intergenic | MTX2;MIR1246 | dist=135882;dist=127073 | | | | | |
| 12M | rs532416695 | 2 | 176622062 | 3 | 1.32803586650991e-07 | 19.8726656 | intergenic | MIR1246;LINC01116 | dist=21010;dist=7519 | | | | | |
| 3M | rs112557251 | 2 | 187510673 | 3 | 3.44653364518888e-06 | 20.6510064 | intronic | TFPI | . | | | 1 | | |
| 3M | rs185158855 | 2 | 222785307 | 4 | 3.71819301386344e-06 | 16.6678864 | intergenic | MOGAT1;ACSL3 | dist=75377;dist=75728 | | | 1 | | |
| 3M | rs185510569 | 2 | 222950143 | 3 | 5.51379458113093e-08 | 19.5886906 | intergenic | ACSL3;KCNE4 | dist=5505;dist=102047 | | | 1 | | |
| 3M | rs147559909 | 2 | 236142879 | 5 | 1.75642429015382e-08 | 17.0840975 | intergenic | AGAP1;GBX2 | dist=11086;dist=22356 | 1 | | | | |
| 3M | rs181217257 | 2 | 239067423 | 4 | 5.29269845950619e-10 | 21.5677368 | intronic | HDAC4 | . | 1 | | | | |
| 3M | rs188076929 | 2 | 239072023 | 4 | 1.0296847211524e-09 | 19.9775614 | intronic | HDAC4 | . | | | | | |
| 3M | rs112475378 | 3 | 1584977 | 14 | 1.34664824685029e-06 | 9.55085929 | intergenic | CNTN6;CNTN4 | dist=180760;dist=513826 | | | 1 | | |
| 3M | rs145676540 | 3 | 2002951 | 5 | 1.25427052978558e-06 | 13.980658 | intergenic | CNTN6;CNTN4 | dist=598734;dist=95852 | | | | | |
| 12M | rs144788248 | 3 | 19545253 | 10 | 3.87625815195203e-06 | 11.9086237 | intergenic | KCNH8;EFHB | dist=9611;dist=334221 | | 1 | | | |
| 3M | rs146007933 | 3 | 28185932 | 18 | 1.34950762676418e-06 | 7.51149453 | intergenic | LINC01980;CMC1 | dist=325607;dist=55687 | | | 1 | | |
| 3M | rs73057656 | 3 | 33899079 | 33 | 3.43591498795888e-06 | 5.9238329 | intergenic | PDCD6IP;LOC101928135 | dist=29372;dist=976718 | | | 1 | | |
| 3M | rs73085348 | 3 | 42669729 | 11 | 6.28155549798072e-07 | 10.139317 | intergenic | ZBTB47;KLHL40 | dist=2149;dist=15808 | | | 1 | | |
| 3M | rs142684595 | 3 | 55285273 | 5 | 7.16787983460343e-08 | 16.7729431 | intergenic | LINC02017;WNT5A | dist=96059;dist=180442 | | | 1 | | |
| 12M | rs149706477 | 3 | 66162553 | 3 | 2.87948941747911e-06 | 20.9828687 | intronic | SLC25A26 | . | | 1 | | | |
| 3M | rs80203220 | 3 | 123003484 | 6 | 2.31429533714517e-06 | 13.426042 | intronic | SEMA5B | . | | | 1 | | |
| 12M | rs114280794 | 3 | 133123359 | 14 | 3.81823609520308e-06 | 8.76296877 | intronic | TMEM108 | . | | | 1 | | |
| 3M | rs192443987 | 3 | 135813503 | 4 | 1.3640761999881e-06 | 17.7540285 | intergenic | EPHB1;PPP2R3A | dist=553038;dist=152225 | | | 1 | | |
| 3M | rs148248743 | 3 | 136415753 | 3 | 7.68859185089755e-07 | 22.839379 | intronic | STAG1 | . | | 1 | 1 | 1 | |
| 12M | rs148248743 | 3 | 136415753 | 3 | 6.68816287673531E-07 | 27.1548587 | intronic | STAG1 | . | | | | | |
| 3M | rs576124203 | 3 | 142122354 | 3 | 4.46547319819514e-07 | 19.4039925 | intronic | TFDP2 | . | | | 1 | | |
| 3M | rs545552231 | 3 | 142145508 | 3 | 4.79811814113124e-07 | 19.8641332 | intronic | TFDP2 | . | | | | | |
| 12M | rs138138661 | 3 | 147940027 | 4 | 5.4928185119564e-08 | 23.1347457 | intergenic | LOC440982;LINC02032 | dist=430117;dist=138132 | | 1 | | | |
| 12M | rs189234695 | 3 | 147940371 | 3 | 3.47705303907978e-06 | 23.9697134 | intergenic | LOC440982;LINC02032 | dist=430461;dist=137788 | | | | | |
| 12M | rs148997617 | 3 | 148037789 | 4 | 1.05449927842458e-07 | 20.8774277 | intergenic | LOC440982;LINC02032 | dist=527879;dist=40370 | | 1 | | | |
| 3M | rs193153124 | 3 | 148612923 | 5 | 2.77507554510464e-06 | 14.3657313 | intergenic | LINC02046;AGTR1 | dist=212967;dist=84948 | | | 1 | | |
| 3M | rs188720948 | 3 | 150352093 | 3 | 8.59439366670513e-07 | 17.7032653 | intergenic | LINC01214;TSC22D2 | dist=28346;dist=56205 | | | 1 | | |
| 3M | rs16823323 | 3 | 153939413 | 14 | 3.35704338797875e-08 | 9.48464065 | intergenic | LINC02006;ARHGEF26 | dist=176887;dist=84988 | 1 | | | | |
| 3M | rs139943877 | 3 | 155733500 | 7 | 2.25796218142194e-06 | 13.0535902 | intronic | PLCH1 | . | | | 1 | | |
| 12M | rs148483098 | 3 | 157664262 | 5 | 3.27691859100501e-06 | 15.7357844 | intergenic | SLC66A1L;SHOX2 | dist=63168;dist=431643 | | 1 | | | |
| 12M | rs79252854 | 3 | 157805368 | 5 | 2.4780397246083e-06 | 17.5464336 | intergenic | SLC66A1L;SHOX2 | dist=204274;dist=290537 | | | | | |
| 12M | rs139055031 | 3 | 158869060 | 6 | 3.17888515002842e-06 | 16.2853761 | intergenic | MFSD1;IQCJ | dist=39341;dist=200192 | | 1 | | | |
| 12M | rs143669489 | 3 | 159675096 | 3 | 4.93900179409246e-06 | 24.2253199 | intronic | IQCJ-SCHIP1;SCHIP1 | . | | | | | |
| 3M | rs187047882 | 3 | 164567833 | 3 | 3.41360195160902e-06 | 18.5641361 | intergenic | MIR1263;LINC01324 | dist=396277;dist=146262 | | | 1 | | |
| 3M | rs141169929 | 3 | 165090674 | 5 | 2.90582805983588e-06 | 14.8454178 | intergenic | SI;SLITRK3 | dist=12178;dist=96046 | | | | | |
| 3M | rs186649043 | 3 | 175214503 | 3 | 2.29531002487815e-06 | 19.2134209 | intronic | NAALADL2 | . | | | 1 | | |
| 3M | rs189709453 | 3 | 177856520 | 3 | 1.55548473782376e-06 | 23.2009936 | ncRNA_intronic | LINC02015 | . | | 1 | 1 | 1 | |
| 12M | rs189709453 | 3 | 177856520 | 3 | 1.55548473782376E-06 | 23.2009936 | ncRNA_intronic | LINC02015 | . | | | | | |
| 3M | rs186767531 | 3 | 177921989 | 3 | 1.62563417403378E-07 | 21.2013243 | intergenic | LINC02015;LINC01014 | dist=22765;dist=497212 | | | | | |
| 12M | rs186767531 | 3 | 177921989 | 3 | 1.66219440747609E-06 | 23.0749977 | intergenic | LINC02015;LINC01014 | dist=22765;dist=497212 | | | | | |
| 3M | rs182868205 | 3 | 177994660 | 3 | 1.36806623320862e-06 | 22.8544303 | intergenic | LINC02015;LINC01014 | dist=95436;dist=424541 | | | | | |
| 12M | rs182868205 | 3 | 177994660 | 3 | 1.36806623320862E-06 | 22.8544303 | intergenic | LINC02015;LINC01014 | dist=95436;dist=424541 | | | | | |
| 12M | rs76356799 | 3 | 179875980 | 4 | 2.9143183363234e-07 | 19.2867007 | intronic | PEX5L | . | | 1 | | | |
| 12M | rs117591241 | 3 | 190866890 | 4 | 4.37648501942309e-06 | 19.9014028 | intergenic | GMNC;SNAR-I | dist=4196;dist=11040 | | 1 | | | |
| 3M | rs191792521 | 3 | 195919734 | 6 | 2.52243498533689e-06 | 13.9515484 | intergenic | TNK2;SDHAP1 | dist=6470;dist=40187 | | | 1 | | |

| | | | | | | | | | | | | | | |
|---|---|---|---|---|---|---|---|---|---|---|---|---|---|---|
| 3M | rs113651406 | 4 | 997179 | 3 | 2.52148284014823e-07 | 19.4054026 | intronic | IDUA | . | | | | 1 | |
| 3M | rs12502861 | 4 | 2424578 | 9 | 2.48774370313747e-06 | 11.1588472 | intronic | CFAP99 | . | | | | 1 | |
| 3M | rs189765693 | 4 | 4323066 | 4 | 2.55310143124462e-06 | 15.5191341 | intergenic | ZBTB49;NSG1 | dist=1283;dist=63466 | | | | 1 | |
| 12M | rs189094663 | 4 | 11621826 | 3 | 2.89927274380224e-06 | 23.895416 | intergenic | HS3ST1;LINC02360 | dist=192932;dist=119125 | | | 1 | | |
| 12M | rs557276277 | 4 | 19012075 | 5 | 3.35996743882432e-06 | 19.0146344 | intergenic | LCORL;SLIT2 | dist=990200;dist=1239830 | | | 1 | | |
| 3M | rs183962155 | 4 | 21258020 | 6 | 3.64461882511902e-06 | 13.7646008 | intronic | KCNIP4 | . | | | | 1 | |
| 3M | rs113751774 | 4 | 23427231 | 7 | 5.00531306271799e-07 | 14.9103274 | intergenic | GBA3;PPARGC1A | dist=607659;dist=364790 | | 1 | 1 | | 1 |
| 3M | rs113063005 | 4 | 23507444 | 6 | 1.79978248668523e-11 | 21.2073894 | intergenic | GBA3;PPARGC1A | dist=687872;dist=284577 | | | | | |
| 12M | rs113063005 | 4 | 23507444 | 7 | 9.56810303773154E-07 | 16.61763 | intergenic | GBA3;PPARGC1A | | | | | | |
| 3M | rs145875128 | 4 | 32071514 | 3 | 1.07037949708586e-07 | 24.479986 | ncRNA_intronic | LINC02506 | . | | | | 1 | |
| 3M | rs143287889 | 4 | 35570658 | 3 | 4.29676290263636e-06 | 19.96741 | intergenic | LINC02484;ARAP2 | dist=1300911;dist=495346 | | | | | |
| 3M | rs77141817 | 4 | 37052137 | 3 | 1.52230770368993e-08 | 20.6110292 | intergenic | LINC02616;MIR4801 | dist=31431;dist=189773 | | 1 | | | |
| 3M | rs190822761 | 4 | 37097734 | 3 | 1.6069016808543e-08 | 20.5846354 | intergenic | LINC02616;MIR4801 | dist=77028;dist=144176 | | | | | |
| 12M | rs147267707 | 4 | 43024541 | 4 | 2.25472034099615e-06 | 18.0094385 | intronic | GRXCR1 | . | | | 1 | | |
| 12M | rs77618729 | 4 | 43048951 | 4 | 2.25749765325168e-06 | 18.0130153 | intergenic | GRXCR1;LINC02383 | dist=18293;dist=408583 | | | | | |
| 12M | rs556435089 | 4 | 43549016 | 4 | 2.61570608345655e-06 | 17.939079 | intergenic | LINC02383;LINC02475 | dist=56473;dist=467845 | | | | | |
| 12M | rs532464521 | 4 | 43569022 | 4 | 2.45873915171385e-06 | 17.9895528 | intergenic | LINC02383;LINC02475 | dist=76479;dist=447839 | | | | | |
| 3M | rs999769259 | 4 | 61646247 | 3 | 3.61294355616724e-06 | 22.1246725 | intronic | ADGRL3 | . | | | | 1 | |
| 3M | rs147630370 | 4 | 86529522 | 4 | 7.46814493303135e-07 | 16.0013633 | intergenic | MAPK10;MIR4452 | dist=76327;dist=12960 | | | | 1 | |
| 3M | rs147171192 | 4 | 88214436 | 5 | 4.39967162056646e-06 | 13.3044047 | intronic | ABCG2 | . | | | | 1 | |
| 3M | rs142993106 | 4 | 90036221 | 15 | 3.08700493831608e-06 | 8.08463611 | intergenic | MMRN1;CCSER1 | dist=81611;dist=91173 | | | | 1 | |
| 3M | rs146526206 | 4 | 90071867 | 15 | 1.81582969378392e-06 | 8.09176656 | intergenic | MMRN1;CCSER1 | dist=117257;dist=55527 | | | | | |
| 3M | rs17017794 | 4 | 90904734 | 33 | 3.46919898165651e-07 | 5.83240545 | intronic | CCSER1 | . | | | | | |
| 3M | rs145116559 | 4 | 111756440 | 4 | 4.90673849934313e-06 | 18.2441224 | intergenic | MIR297;FAM241A | dist=895793;dist=389014 | | | | 1 | |
| 3M | rs181415102 | 4 | 111768110 | 4 | 4.81312717634891e-06 | 18.2631019 | intergenic | MIR297;FAM241A | dist=907463;dist=377344 | | | | | |
| 3M | rs191423619 | 4 | 125595472 | 3 | 2.50684302750887e-07 | 21.694611 | intergenic | MIR2054;INTU | dist=88165;dist=2037485 | | | | 1 | |
| 3M | rs149298750 | 4 | 126237546 | 3 | 1.98725449883812e-07 | 22.184052 | intergenic | MIR2054;INTU | dist=730239;dist=1395411 | | | | | |
| 12M | rs181132315 | 4 | 128726214 | 3 | 1.68603480568751e-06 | 22.1324679 | intergenic | LINC02615;JADE1 | dist=206818;dist=83486 | | | 1 | | |
| 3M | rs112679237 | 4 | 138219706 | 18 | 2.04053672228041e-07 | 8.71202752 | intronic | SLC7A11 | . | | | | 1 | |
| 12M | rs147455971 | 4 | 140010558 | 5 | 3.87240564796723e-06 | 17.2160142 | intronic | MAML3 | . | | | 1 | | |
| 3M | rs531769270 | 4 | 153489891 | 3 | 2.4145887031028e-06 | 16.5945338 | intronic | TMEM131L | . | | | | 1 | |
| 3M | rs116651654 | 4 | 162317591 | 6 | 3.48829177722726e-07 | 15.6223694 | intergenic | FSTL5;MIR4454 | dist=153557;dist=775983 | | | | 1 | |
| 12M | rs17615362 | 4 | 169012936 | 44 | 4.30304118178725e-06 | 5.41974472 | intergenic | CBR4;SH3RF1 | dist=2681;dist=81323 | | | 1 | | |
| 12M | rs17543620 | 4 | 169013574 | 44 | 4.21133121953689e-06 | 5.42796755 | intergenic | CBR4;SH3RF1 | dist=3319;dist=80685 | | | | | |
| 3M | rs567982164 | 5 | 25631188 | 3 | 5.7310337989489e-07 | 21.8356286 | intergenic | LINC02211;CDH9 | dist=328908;dist=1249409 | | | | 1 | |
| 3M | rs191986449 | 5 | 25745578 | 3 | 4.59857674938651e-06 | 18.4978793 | intergenic | LINC02211;CDH9 | dist=443298;dist=1135019 | | | | | |
| 12M | rs142934021 | 5 | 53366842 | 7 | 1.40182538744286e-08 | 18.824715 | intergenic | LOC257396;FST | dist=251716;dist=113787 | 1 | | | | |
| 3M | rs185771987 | 5 | 73989664 | 3 | 4.53395085864185e-06 | 18.2344824 | intergenic | ARHGEF28;LINC01335 | dist=47671;dist=316746 | | | | 1 | |
| 3M | rs139360368 | 5 | 74076284 | 4 | 1.24776973080952e-06 | 18.3590487 | intergenic | ARHGEF28;LINC01335 | dist=134291;dist=230126 | | | | 1 | |
| 3M | rs181933850 | 5 | 92169830 | 7 | 3.33393070013435e-06 | 11.0436586 | intergenic | ARRDC3;NR2F1 | dist=749115;dist=1279415 | | | | 1 | |
| 3M | rs190190051 | 5 | 92179668 | 7 | 3.47677941958718e-06 | 11.0589887 | intergenic | ARRDC3;NR2F1 | dist=758953;dist=1269577 | | | | | |
| 3M | rs182531466 | 5 | 92234256 | 6 | 4.23245053141191e-06 | 12.6804978 | intergenic | ARRDC3;NR2F1 | dist=813541;dist=1214989 | | | | | |
| 3M | rs187236873 | 5 | 92234630 | 6 | 1.97926852689169e-06 | 12.44907 | intergenic | ARRDC3;NR2F1 | dist=813915;dist=1214615 | | | | | |
| 3M | rs183816745 | 5 | 92336071 | 5 | 6.02067560021534e-08 | 17.3487909 | intergenic | ARRDC3;NR2F1 | dist=915356;dist=1113174 | | | | | |
| 3M | rs111407636 | 5 | 95744325 | 3 | 6.27809322802765e-07 | 18.642774 | intronic | RHOBTB3 | . | | | | 1 | |
| 3M | rs111676272 | 5 | 95758594 | 3 | 6.71649637293761e-07 | 18.6068625 | intronic | RHOBTB3 | . | | | | | |
| 3M | rs75848314 | 5 | 95762636 | 3 | 1.11087241977704e-07 | 20.0998481 | intronic | RHOBTB3 | . | | | | | |
| 3M | rs111846247 | 5 | 95775939 | 3 | 1.06810112370498e-07 | 20.4865772 | intronic | RHOBTB3 | . | | | | | |
| 3M | rs137873790 | 5 | 97751337 | 11 | 4.51630701305597e-06 | 9.22240881 | intergenic | LINC01340;LINC02234 | dist=80286;dist=89421 | | | | 1 | |
| 3M | rs189912648 | 5 | 135429749 | 4 | 2.55441427995549e-06 | 16.9732599 | intergenic | MACROH2A1;DCANP1 | dist=29862;dist=14465 | | | 1 | 1 | 1 |
| 12M | rs189912648 | 5 | 135429749 | 4 | 3.32990944802665E-06 | 19.8604449 | intergenic | MACROH2A1;DCANP1 | dist=29862;dist=14465 | | | | | |
| 12M | rs111512950 | 5 | 154300867 | 8 | 4.11671051184833e-06 | 13.567443 | intronic | GALNT10 | . | | | 1 | | |

| | | | | | | | | | | | | |
|---|---|---|---|---|---|---|---|---|---|---|---|---|
| 3M | rs191006910 | 5 | 154836449 | 3 | 3.35533057793161e-06 | 17.6173803 | intronic | FAXDC2 | . | | | 1 |
| 3M | rs74343174 | 5 | 162066176 | 5 | 2.54836335504837e-06 | 14.7671824 | intergenic | LINC01202;GABRG2 | dist=64980;dist=1289 | | | 1 |
| 3M | rs775626702 | 5 | 163203061 | 3 | 3.23411085027866e-06 | 18.9484137 | intergenic | GABRG2;CCNG1 | dist=1047522;dist=234510 | | | |
| 3M | rs371245624 | 5 | 163453950 | 3 | 1.65804063735814e-06 | 19.4795931 | UTR3 | NUDCD2 | NM_001329991:c.*170>0;NM_145266:c.*170>0 | | | 1 |
| 3M | rs290120 | 5 | 163841238 | 6 | 2.40444258455957e-06 | 13.3587764 | intergenic | MAT2B;LINC02143 | dist=321884;dist=607184 | | | 1 |
| 3M | rs545428520 | 5 | 168394261 | 4 | 3.96137253437646e-09 | 21.566035 | intronic | WWC1 | . | 1 | | |
| 3M | rs528404963 | 5 | 168425020 | 4 | 7.1738299793335e-09 | 21.1393977 | intronic | WWC1 | . | | | |
| 3M | rs72832764 | 5 | 170577669 | 3 | 2.69939738977966e-06 | 18.2159757 | intronic | KCNIP1 | . | | | 1 |
| 3M | rs72837643 | 5 | 170761951 | 4 | 4.147895992623e-06 | 16.9302874 | intergenic | KCNIP1;GABRP | dist=25319;dist=21768 | | | |
| 3M | rs142311947 | 5 | 177957468 | 8 | 2.99420386474277e-06 | 12.0339013 | ncRNA_intronic | LOC100128340 | . | | | 1 |
| 3M | rs151015676 | 5 | 177963936 | 3 | 4.05781112465234e-06 | 20.1421367 | intergenic | LOC100128340;PROP1 | dist=4136;dist=28299 | | | |
| 3M | rs1877768 | 6 | 16534692 | 15 | 2.77630022916084e-06 | 8.28754685 | intronic | ATXN1 | . | | | 1 |
| 12M | rs73384619 | 6 | 20457948 | 21 | 3.65476878017977e-06 | 8.3916702 | intronic | E2F3 | . | | 1 | |
| 3M | rs150586237 | 6 | 24491120 | 3 | 5.5145390836127e-09 | 22.0461443 | intergenic | GPLD1;ALDH5A1 | dist=1542;dist=3849 | 1 | | |
| 3M | rs571986619 | 6 | 84969114 | 3 | 1.70115854486702e-06 | 20.7858067 | intergenic | TBX18;LINC02535 | dist=204516;dist=418105 | | | 1 |
| 3M | rs56224400 | 6 | 97644799 | 14 | 9.49951178557744e-07 | 8.4433064 | ncRNA_intronic | LOC101927314 | . | | | 1 |
| 3M | rs147627638 | 6 | 98725240 | 6 | 2.84737109324869e-06 | 13.0785306 | intergenic | MIR2113;PNKY | dist=700621;dist=104901 | | | |
| 3M | rs141326851 | 6 | 134511989 | 14 | 2.157242978017e-06 | 8.57849688 | intergenic | LINC01010;LOC101928304 | dist=7969;dist=13329 | | | 1 |
| 3M | rs146048121 | 6 | 141877818 | 3 | 3.23639603147815e-06 | 20.0214379 | intergenic | MIR4465;NMBR | dist=1193935;dist=196666 | 1 | | |
| 3M | rs142106992 | 6 | 141948748 | 4 | 5.23067431501407e-10 | 22.8345886 | intergenic | MIR4465;NMBR | dist=1264865;dist=125736 | | | |
| 3M | rs72983831 | 6 | 141985963 | 6 | 7.82545989102806e-07 | 15.6733239 | intergenic | MIR4465;NMBR | dist=1302080;dist=88521 | | | |
| 3M | rs72986533 | 6 | 142290121 | 8 | 1.53596722070896e-07 | 13.2812726 | intergenic | VTA1;ADGRG6 | dist=65437;dist=11798 | | | |
| 3M | rs73586304 | 6 | 142518288 | 3 | 4.95566451287885e-06 | 18.2405116 | intergenic | ADGRG6;LOC153910 | dist=72022;dist=8167 | | | |
| 12M | rs4896997 | 6 | 148362788 | 6 | 3.04539162547005e-06 | 14.4320923 | intronic | SASH1 | . | | 1 | |
| 12M | rs4131286 | 6 | 148367827 | 6 | 3.05925201460773e-06 | 14.4284672 | intronic | SASH1 | . | | | |
| 12M | rs17078283 | 6 | 148375784 | 6 | 3.05230865558584e-06 | 14.4302821 | intronic | SASH1 | . | | | |
| 12M | rs56821264 | 6 | 148382765 | 6 | 2.17257084724272e-06 | 14.7509213 | intronic | SASH1 | . | | | |
| 3M | rs148532212 | 6 | 165086368 | 3 | 1.8149830669263e-07 | 19.3535457 | intergenic | MEAT6;C6orf118 | dist=264305;dist=193296 | | | 1 |
| 3M | rs117498042 | 6 | 165100792 | 3 | 1.85878685547635e-07 | 19.335471 | intergenic | MEAT6;C6orf118 | dist=278729;dist=178872 | | | |
| 3M | rs148153037 | 6 | 167087898 | 8 | 3.20376100252789e-09 | 14.3359246 | intergenic | CEP43;CCR6 | dist=35180;dist=23909 | 1 | | |
| 3M | rs184487573 | 6 | 167099983 | 7 | 1.91568282565546e-07 | 13.5009635 | intergenic | CEP43;CCR6 | dist=47265;dist=11824 | | | |
| 3M | rs17776100 | 7 | 6386848 | 25 | 5.13308111425872e-07 | 6.63697397 | intronic | RAC1 | . | | | 1 |
| 3M | rs187978759 | 7 | 11672218 | 3 | 2.76447739355703e-06 | 20.4555522 | intronic | THSD7A | . | | | 1 |
| 12M | rs143686474 | 7 | 16252374 | 11 | 4.88904023817646e-06 | 10.8531498 | ncRNA_intronic | CRPPA | . | | 1 | |
| 3M | rs117166500 | 7 | 17013154 | 7 | 1.89815826168549e-06 | 12.5146865 | intergenic | AGR3;AHR | dist=131171;dist=285498 | | | 1 |
| 3M | rs75689761 | 7 | 18366950 | 7 | 9.11790229535791e-07 | 11.8482917 | intronic | HDAC9 | . | 1 | | |
| 3M | rs77346868 | 7 | 18366976 | 8 | 1.13767941006556e-06 | 11.0210129 | intronic | HDAC9 | . | | | |
| 3M | rs78225611 | 7 | 18367841 | 8 | 1.13375019479557e-06 | 11.0294387 | intronic | HDAC9 | . | | | |
| 3M | rs77300464 | 7 | 18369138 | 6 | 2.09191655072049e-08 | 14.6659794 | intronic | HDAC9 | . | | | |
| 3M | rs79602997 | 7 | 18370627 | 7 | 9.43076371109133e-07 | 11.8608211 | intronic | HDAC9 | . | | | |
| 3M | rs75773869 | 7 | 18371222 | 7 | 9.36316662267224e-07 | 11.8642716 | intronic | HDAC9 | . | | | |
| 3M | rs75606013 | 7 | 18374990 | 6 | 2.11098533218711e-08 | 14.6617119 | intronic | HDAC9 | . | | | |
| 3M | rs61434999 | 7 | 18378728 | 7 | 9.22178787748122e-07 | 11.8411511 | intronic | HDAC9 | . | | | |
| 3M | rs78907958 | 7 | 18385394 | 7 | 4.98811901973441e-07 | 12.0901322 | intronic | HDAC9 | . | | | |
| 3M | rs76526501 | 7 | 18391487 | 6 | 8.57654056311165e-09 | 14.8397312 | intronic | HDAC9 | . | | | |
| 3M | rs74455595 | 7 | 18392161 | 6 | 8.57654056311165e-09 | 14.8397312 | intronic | HDAC9 | . | | | |
| 3M | rs79182806 | 7 | 18394204 | 7 | 4.13732512224196e-07 | 12.0972603 | intronic | HDAC9 | . | | | |
| 3M | rs10279777 | 7 | 18401966 | 8 | 1.65309369490778e-08 | 12.8681335 | intronic | HDAC9 | . | | | |
| 3M | rs77867199 | 7 | 18402652 | 9 | 3.93328414300688e-07 | 10.8221282 | intronic | HDAC9 | . | | | |
| 3M | rs80156375 | 7 | 18403592 | 8 | 3.27106315853648e-07 | 11.5406255 | intronic | HDAC9 | . | | | |
| 3M | rs17169602 | 7 | 18407118 | 8 | 1.78410513894506e-08 | 12.7273717 | intronic | HDAC9 | . | | | |
| 3M | rs10486295 | 7 | 18407184 | 8 | 1.75006169477667e-08 | 12.76229 | intronic | HDAC9 | . | | | |

| | | | | | | | | | | | | |
|---|---|---|---|---|---|---|---|---|---|---|---|---|
| 3M | rs75090694 | 7 | 18407813 | 7 | 1.01013780102531e-08 | 13.8314005 | intronic | HDAC9 | . | | | |
| 3M | rs55844051 | 7 | 23320744 | 3 | 1.93295366566624e-06 | 18.3875896 | intronic | IGF2BP3 | . | | | 1 |
| 3M | rs62447184 | 7 | 36534898 | 43 | 2.45196683172423e-06 | 5.12842209 | intronic | AOAH | . | | | |
| 3M | rs574076561 | 7 | 49505151 | 3 | 3.8458987487847e-06 | 19.6434766 | intergenic | CDC14C;VWC2 | dist=577697;dist=268487 | | | 1 |
| 3M | rs111391231 | 7 | 89867731 | 12 | 4.17997193738163e-06 | 8.31717755 | intergenic | ZNF804B;STEAP2 | dist=529203;dist=14622 | | | 1 |
| 3M | rs111900874 | 7 | 89887355 | 12 | 3.79796761664437e-06 | 8.42424013 | ncRNA_intronic | STEAP2 | . | | | |
| 3M | rs181259864 | 7 | 97859511 | 3 | 3.26388920997597e-06 | 17.1360821 | ncRNA_intronic | CZ1P-ASNS | . | | | 1 |
| 3M | rs192750513 | 7 | 97948518 | 3 | 3.9425756746568e-06 | 17.1892469 | ncRNA_intronic | CZ1P-ASNS | . | | | |
| 3M | rs539713344 | 7 | 100877165 | 3 | 2.02994489815685e-06 | 20.044756 | intronic | SRRT | . | | | 1 |
| 3M | rs188028357 | 7 | 101025144 | 3 | 5.38240258932335e-07 | 21.8425578 | intronic | MUC17 | . | | | 1 |
| 12M | rs73202425 | 7 | 109515659 | 12 | 3.05259237348811e-06 | 10.761844 | intergenic | C7orf66;EIF3IP1 | dist=631072;dist=443568 | | 1 | |
| 12M | rs181182636 | 7 | 116014504 | 3 | 2.90894506747647e-06 | 21.9472639 | intronic | TFEC | . | | 1 | |
| 12M | rs182211730 | 7 | 116363561 | 4 | 1.45395858607717e-06 | 18.7746033 | intergenic | LOC102724434;CAV2 | dist=76834;dist=136177 | | 1 | |
| 12M | rs143281973 | 7 | 116367942 | 4 | 1.62372890091138e-06 | 18.2750778 | intergenic | LOC102724434;CAV2 | dist=81215;dist=131796 | | | |
| 12M | rs138873576 | 7 | 116385305 | 4 | 3.22380485845586e-06 | 17.6151711 | intergenic | LOC102724434;CAV2 | dist=98578;dist=114433 | | | |
| 12M | rs190354334 | 7 | 116435792 | 5 | 4.2862377340545e-06 | 17.3392581 | intergenic | LOC102724434;CAV2 | dist=149065;dist=63946 | | | |
| 12M | rs186893139 | 7 | 116456916 | 4 | 3.55104961576984e-06 | 18.602598 | intergenic | LOC102724434;CAV2 | dist=170189;dist=42822 | | | |
| 12M | rs558784715 | 7 | 117403240 | 3 | 1.5182227326648e-06 | 20.7591787 | intronic | ASZ1 | . | | 1 | |
| 12M | rs576047962 | 7 | 117468967 | 3 | 1.61732320840498e-06 | 20.7261549 | intergenic | ASZ1;CFTR | dist=41474;dist=11058 | | | |
| 12M | rs529110230 | 7 | 117535853 | 3 | 1.24629705452557e-06 | 20.8583633 | intronic | CFTR | . | | | |
| 12M | rs142215699 | 7 | 117559820 | 3 | 1.6210064953756e-06 | 20.7292287 | ncRNA_intronic | CFTR | . | | | |
| 12M | rs142721557 | 7 | 117578274 | 3 | 1.61401730397805e-06 | 20.7329975 | intronic | CFTR | . | | | |
| 12M | rs201355675 | 7 | 117585727 | 3 | 1.64203568676087e-06 | 20.718124 | intronic | CFTR | . | | | |
| 12M | rs188993522 | 7 | 117634677 | 3 | 1.9792198058251e-06 | 21.2984827 | intronic | CFTR | . | | | |
| 3M | rs536023430 | 7 | 147167980 | 3 | 2.63612685649085e-06 | 19.4038683 | intronic | CNTNAP2 | . | | | 1 |
| 3M | rs187384541 | 8 | 1684075 | 26 | 3.15110880283135e-06 | 6.88745117 | intronic | DLGAP2 | . | | | 1 |
| 3M | rs575473987 | 8 | 5719972 | 3 | 1.07256928472273e-08 | 22.2248653 | intergenic | CSMD1;LOC100287015 | dist=725058;dist=683583 | 1 | | |
| 3M | rs139062456 | 8 | 13394482 | 15 | 3.88565669511608e-06 | 7.83515809 | intronic | DLC1 | . | | | 1 |
| 3M | rs185874707 | 8 | 18173165 | 10 | 2.74810255839378e-06 | 9.13934765 | intronic | NAT1 | . | | | 1 |
| 3M | rs140797780 | 8 | 22230279 | 3 | 1.73425396214273e-07 | 19.2908396 | intronic | PHYHIP | . | | | 1 |
| 3M | rs147601511 | 8 | 22464806 | 4 | 7.14867637986866e-07 | 16.4471181 | intronic | PPP3CC | . | | | 1 |
| 3M | rs188415494 | 8 | 25755782 | 3 | 3.49911568838158e-06 | 18.5169478 | intergenic | CDCA2;EBF2 | dist=247865;dist=85943 | | | 1 |
| 3M | rs9643828 | 8 | 54616513 | 279 | 2.29737457097614e-07 | -2.4236294 | intronic | RP1 | . | | | 1 |
| 3M | rs423841 | 8 | 54643509 | 291 | 2.7508818251862e-06 | -2.1495726 | intronic | RP1 | . | | | |
| 3M | rs433324 | 8 | 54652049 | 287 | 9.22325760191311e-07 | -2.2483462 | intronic | RP1 | . | | | |
| 3M | rs369623 | 8 | 54659380 | 287 | 8.50610562336969e-07 | -2.2359815 | intronic | RP1 | . | | | |
| 3M | rs446222 | 8 | 54662400 | 288 | 7.36959451875675e-07 | -2.247819 | intronic | RP1 | . | | | |
| 3M | rs432393 | 8 | 54667738 | 291 | 1.22958170848882e-06 | -2.1875382 | intronic | RP1 | . | | | |
| 3M | rs3098298 | 8 | 54670278 | 291 | 1.22819650607779e-06 | -2.1875635 | intronic | RP1 | . | | | |
| 3M | rs367179 | 8 | 54675056 | 291 | 1.22819650607779e-06 | -2.1875635 | intronic | RP1 | . | | | |
| 3M | rs382476 | 8 | 54678415 | 288 | 7.36949131111935e-07 | -2.2481938 | intronic | RP1 | . | | | |
| 3M | rs384543 | 8 | 54679049 | 288 | 7.36949131111935e-07 | -2.2481938 | intronic | RP1 | . | | | |
| 3M | rs405226 | 8 | 54679776 | 291 | 1.2251324121268e-06 | -2.1881305 | intronic | RP1 | . | | | |
| 3M | rs384127 | 8 | 54684929 | 288 | 7.36964497256014e-07 | -2.2481559 | intronic | RP1 | . | | | |
| 3M | rs858397 | 8 | 54702130 | 278 | 7.161979275107e-07 | 2.2301348 | intronic | RP1 | . | | | |
| 3M | rs2375537 | 8 | 54706948 | 280 | 6.2991993988762e-07 | 2.23876848 | intronic | RP1 | . | | | |
| 3M | rs720372 | 8 | 54716077 | 292 | 2.93014534156507e-06 | 2.09872734 | intronic | RP1 | . | | | |
| 3M | rs1437781 | 8 | 54717292 | 280 | 6.32222203401203e-07 | 2.23854673 | intronic | RP1 | . | | | |
| 3M | rs1595406 | 8 | 54718055 | 291 | 2.80263765648492e-06 | 2.10018889 | intronic | RP1 | . | | | |
| 3M | rs1437782 | 8 | 54720202 | 278 | 5.53724964407086e-07 | 2.2579585 | exonic | RP1 | . | | | |
| 3M | rs10105693 | 8 | 54727912 | 276 | 3.41214459494352e-07 | 2.2928391 | intronic | RP1 | . | | | |
| 3M | rs2375536 | 8 | 54728162 | 292 | 2.13408172799341e-06 | 2.12635806 | intronic | RP1 | . | | | |
| 3M | rs4737674 | 8 | 54749094 | 277 | 2.86005059848221e-07 | 2.30559554 | intronic | RP1 | . | | | |

| | | | | | | | | | | | | |
|---|---|---|---|---|---|---|---|---|---|---|---|---|
| 3M | rs11987234 | 8 | 54757269 | 276 | 3.13190459972498e-07 | 2.29599259 | intronic | RP1 | . | | | |
| 3M | rs13277510 | 8 | 54761589 | 277 | 2.8610568625199e-07 | 2.30494752 | intronic | RP1 | . | | | |
| 3M | rs12548593 | 8 | 54762057 | 279 | 3.38344249342945e-07 | 2.28269961 | intronic | RP1 | . | | | |
| 3M | rs1812506 | 8 | 54763541 | 291 | 2.00833101672028e-06 | 2.12258868 | intronic | RP1 | . | | | |
| 3M | rs16920698 | 8 | 54765874 | 277 | 2.85868793626322e-07 | 2.30492695 | intronic | RP1 | . | | | |
| 3M | rs1561297 | 8 | 54765978 | 279 | 3.34668102178151e-07 | 2.28315237 | intronic | RP1 | . | | | |
| 3M | rs4737676 | 8 | 54766986 | 277 | 2.8589296796158e-07 | 2.30492057 | intronic | RP1 | . | | | |
| 3M | rs2083123 | 8 | 54767758 | 279 | 3.4358513898515e-07 | 2.27910794 | intronic | RP1 | . | | | |
| 3M | rs983248 | 8 | 54768232 | 277 | 2.87356270418993e-07 | 2.30260428 | intronic | RP1 | . | | | |
| 3M | rs1391463 | 8 | 54769316 | 277 | 2.87382622216009e-07 | 2.30258865 | intronic | RP1 | . | | | |
| 3M | rs10958428 | 8 | 54773081 | 280 | 5.0112447137886e-07 | 2.23996885 | intronic | RP1 | . | | | |
| 3M | rs13278605 | 8 | 54775611 | 277 | 3.41581916245031e-07 | 2.27377348 | intronic | RP1 | . | | | |
| 3M | rs13276543 | 8 | 54775614 | 276 | 3.13999361335922e-07 | 2.29263157 | intronic | RP1 | . | | | |
| 3M | rs7822082 | 8 | 54777660 | 277 | 1.5040633792954e-07 | 2.3570878 | intronic | RP1 | . | | | |
| 3M | rs4737201 | 8 | 54778898 | 277 | 2.89096388651053e-07 | 2.30197734 | intronic | RP1 | . | | | |
| 3M | rs7843693 | 8 | 54779552 | 334 | 4.78812399757755e-06 | 1.99856756 | intronic | RP1 | . | | | |
| 3M | rs1396896 | 8 | 54782750 | 334 | 4.77810054425197e-06 | 1.99853327 | intronic | RP1 | . | | | |
| 3M | rs2375219 | 8 | 54785735 | 330 | 3.05253330469401e-06 | 2.05700167 | intronic | RP1 | . | | | |
| 3M | rs1391462 | 8 | 54787221 | 334 | 4.79440218462524e-06 | 1.9984766 | intronic | RP1 | . | | | |
| 3M | rs12678939 | 8 | 54792461 | 331 | 4.93547251213243e-06 | 2.01975618 | intronic | RP1 | . | | | |
| 3M | rs1498183 | 8 | 54804345 | 332 | 4.76724404445351e-06 | 2.0245746 | intronic | RP1 | . | | | |
| 12M | rs62515405 | 8 | 56143419 | 21 | 4.69438076015372e-06 | 8.07691204 | intergenic | MOS;PLAG1 | dist=29437;dist=17490 | | 1 | |
| 12M | rs62515436 | 8 | 56228644 | 15 | 3.44618686083172e-06 | 9.27545902 | intergenic | CHCHD7;SDR16C5 | dist=9835;dist=71004 | | 1 | |
| 12M | rs12114488 | 8 | 61760164 | 266 | 1.33492404675942e-06 | 2.62307914 | intergenic | MIR4470;LINC02155 | dist=45305;dist=129675 | | 1 | |
| 3M | rs117816016 | 8 | 102739034 | 3 | 3.21886378098173e-06 | 19.8807023 | intergenic | LOC101927245;GASAL1 | dist=52311;dist=67788 | | | 1 |
| 3M | rs567383525 | 8 | 114017265 | 3 | 1.83606414606387e-06 | 17.7455227 | intergenic | CSMD3;TRPS1 | dist=580326;dist=1391230 | | | 1 |
| 3M | rs545550279 | 8 | 114540200 | 3 | 4.75001774825929e-06 | 19.1812421 | intergenic | CSMD3;TRPS1 | dist=1103261;dist=868295 | | | |
| 3M | rs536803366 | 8 | 121989172 | 3 | 1.65900394985684e-06 | 19.2098639 | intergenic | HAS2;SMILR | dist=343847;dist=425155 | | | 1 |
| 3M | rs532513136 | 8 | 134801505 | 3 | 7.47175269151994e-07 | 19.9490594 | upstream | MIR30B | dist=898 | | | 1 |
| 3M | rs532730683 | 9 | 1784492 | 3 | 1.02051348042702e-06 | 18.726838 | intergenic | DMRT2;SMARCA2 | dist=726938;dist=230855 | | | 1 |
| 3M | rs540065886 | 9 | 2770228 | 3 | 1.90434082268377e-06 | 19.9489151 | intergenic | KCNV2;PUM3 | dist=40191;dist=33927 | | | 1 |
| 3M | rs543844012 | 9 | 30107025 | 3 | 4.37741040782571e-06 | 18.1160485 | intergenic | LINGO2;LINC01242 | dist=893424;dist=281910 | | | 1 |
| 3M | rs148556485 | 9 | 81408911 | 3 | 5.40048598404711e-07 | 21.3800152 | intergenic | LINC01507;TLE1 | dist=1374356;dist=174772 | | | 1 |
| 3M | rs140782222 | 9 | 81413979 | 3 | 4.53117855536307e-07 | 21.791526 | intergenic | LINC01507;TLE1 | dist=1379424;dist=169704 | | | |
| 3M | rs188034471 | 9 | 83322799 | 4 | 2.79197739516285e-06 | 14.7559999 | intronic | FRMD3 | . | | | 1 |
| 3M | rs190294315 | 9 | 83330550 | 5 | 4.29120280825722e-07 | 14.5664276 | intronic | FRMD3 | . | | | |
| 3M | rs545690161 | 9 | 90268417 | 6 | 1.13978419923937e-08 | 17.1436874 | intergenic | MIR4290HG;LINC01508 | dist=226918;dist=32479 | 1 | | |
| 3M | rs565682685 | 9 | 90460046 | 4 | 3.57832955336558e-07 | 18.938397 | intergenic | LINC01508;LINC01501 | dist=26557;dist=2386 | | | |
| 3M | rs183737367 | 9 | 90567765 | 3 | 3.8014567877404e-08 | 23.3452306 | ncRNA_intronic | LINC01501 | . | | | |
| 3M | rs187213609 | 9 | 90653183 | 3 | 5.71707003010382e-08 | 23.1975304 | intergenic | DIRAS2;SYK | dist=10359;dist=148417 | | | |
| 12M | rs144138711 | 9 | 97160137 | 40 | 1.39149931661618e-06 | 6.08547131 | ncRNA_intronic | ANKRD18CP | . | | 1 | |
| 3M | rs150027952 | 9 | 100623134 | 3 | 1.2828864296083e-06 | 21.8616648 | intergenic | CAVIN4;PLPPR1 | dist=34747;dist=405593 | | | 1 |
| 3M | rs146207930 | 9 | 126280057 | 6 | 2.39787202381566e-06 | 11.8624321 | intergenic | LOC101929116;MVB12B | dist=4142;dist=46772 | | | 1 |
| 3M | rs78296164 | 9 | 132391328 | 7 | 6.93811773227571e-07 | 11.6095338 | intronic | TTF1 | . | | | 1 |
| 3M | rs77871739 | 9 | 135660463 | 4 | 3.19831184275799e-06 | 15.0724603 | intergenic | GLT6D1;LCN9 | dist=20923;dist=2859 | | | 1 |
| 3M | rs118040657 | 10 | 3430654 | 8 | 4.61906712585886e-06 | 12.1147712 | ncRNA_intronic | LOC105376360 | . | | | 1 |
| 3M | rs184425183 | 10 | 13415520 | 3 | 1.92440370103676e-09 | 22.6683174 | intergenic | SEPHS1;BEND7 | dist=67222;dist=22961 | 1 | | |
| 3M | rs184458518 | 10 | 13429195 | 3 | 1.48323670154931e-09 | 22.8015789 | intergenic | SEPHS1;BEND7 | dist=80897;dist=9286 | | | |
| 3M | rs117998251 | 10 | 13455976 | 3 | 1.19486099712372e-09 | 22.9543311 | intronic | BEND7 | . | | | |
| 3M | rs117025967 | 10 | 19918123 | 11 | 5.25165736443477e-07 | 9.66973563 | intronic | PLXDC2 | . | | | 1 |
| 3M | rs529011661 | 10 | 20083387 | 4 | 2.34383539239577e-06 | 15.6393469 | intronic | PLXDC2 | . | | | |
| 12M | rs142956968 | 10 | 23201236 | 14 | 8.84691036334147e-07 | 10.4167992 | downstream | C10orf67 | dist=680 | | 1 | |
| 12M | rs145764464 | 10 | 23312918 | 15 | 1.10461979254428e-06 | 9.5967306 | intronic | C10orf67 | . | | | |

| | | | | | | | | | | | | |
|---|---|---|---|---|---|---|---|---|---|---|---|---|
| 3M | rs12266995 | 10 | 24563854 | 26 | 3.92980514429013e-06 | 5.96782535 | intergenic | KIAA1217;ARHGAP21 | dist=16006;dist=19760 | | | 1 |
| 3M | rs138249376 | 10 | 61756071 | 4 | 4.86147789321271e-06 | 16.4300711 | intronic | CABCOCO1 | . | | | 1 |
| 3M | rs140277951 | 10 | 80599344 | 6 | 1.98902982335697e-07 | 13.7463158 | intronic | SH2D4B | . | | | 1 |
| 3M | rs566018180 | 10 | 84995296 | 3 | 4.3799943792317e-06 | 18.563552 | intergenic | CCSER2;LINC01519 | dist=476775;dist=198125 | | | 1 |
| 3M | rs140706881 | 10 | 94457778 | 4 | 1.40251021425162e-06 | 18.974805 | intronic | TBC1D12 | . | | | 1 |
| 3M | rs117913371 | 10 | 101158729 | 21 | 2.30639701253664e-08 | 7.86609555 | intergenic | TLX1NB;LINC01514 | dist=17463;dist=17593 | 1 | | |
| 3M | rs75334617 | 10 | 101196395 | 32 | 7.30335368747825e-07 | 5.84603405 | intergenic | LINC01514;LBX1 | dist=2248;dist=30581 | | | |
| 3M | rs752259256 | 10 | 103165562 | 3 | 4.52164287436037e-06 | 20.4258639 | intronic | NT5C2 | . | | | 1 |
| 12M | rs147944608 | 10 | 108663951 | 11 | 9.83587566974791e-07 | 11.2205243 | intergenic | LINC01435;XPNPEP1 | dist=594658;dist=1200815 | | 1 | |
| 3M | rs180828621 | 10 | 122773893 | 6 | 6.39935885478647e-08 | 15.1380574 | ncRNA_intronic | DMBT1L1 | . | | | 1 |
| 3M | rs147393020 | 10 | 123019758 | 5 | 1.64586496415273e-06 | 14.1853505 | intronic | ACADSB | . | | | 1 |
| 3M | rs193093906 | 10 | 125016920 | 9 | 3.17719154902332e-06 | 10.5219537 | intronic | CTBP2 | . | | | 1 |
| 12M | rs556274646 | 11 | 439012 | 9 | 1.93909342324625e-06 | 12.8914592 | intronic | ANO9 | . | | 1 | |
| 12M | rs148815783 | 11 | 1013992 | 8 | 2.06734210421034e-06 | 13.5980313 | exonic | MUC6 | . | | | 1 |
| 3M | rs151115079 | 11 | 18634194 | 5 | 4.89469717121508e-08 | 17.8273587 | intronic | SPTY2D1 | . | 1 | | |
| 3M | rs138414342 | 11 | 18657851 | 5 | 3.15830767684997e-08 | 18.1582505 | intergenic | SPTY2D1;TMEM86A | dist=23509;dist=40928 | | | |
| 3M | rs541653703 | 11 | 18680239 | 4 | 2.1765671670172e-08 | 18.3169499 | intergenic | SPTY2D1;TMEM86A | dist=45897;dist=18540 | | | |
| 3M | rs118093638 | 11 | 18696777 | 5 | 9.7151911784911e-07 | 14.2657259 | intergenic | SPTY2D1;TMEM86A | dist=62435;dist=2002 | | | |
| 3M | rs181812512 | 11 | 66898258 | 3 | 4.24789238469822e-06 | 19.5093168 | intronic | PC | . | | | 1 |
| 3M | rs529345909 | 11 | 67343381 | 3 | 4.87261379674454e-06 | 18.70153 | ncRNA_intronic | LOC100130987 | . | | | 1 |
| 3M | rs544042801 | 11 | 68692775 | 4 | 3.07835940417967e-06 | 15.8869866 | intergenic | GAL;TESMIN | dist=1600;dist=14665 | | | 1 |
| 3M | rs149949098 | 11 | 95366702 | 13 | 2.52728186016237e-06 | 8.55927625 | intergenic | LOC100129203;FAM76B | dist=132298;dist=402251 | | | 1 |
| 3M | rs74521112 | 11 | 99218416 | 27 | 3.48297260681022e-07 | 6.25632387 | intronic | CNTN5 | . | | | 1 |
| 3M | rs79213709 | 11 | 99222724 | 26 | 5.77809319512938e-07 | 6.17081884 | intronic | CNTN5 | . | | | |
| 3M | rs112007361 | 11 | 99317649 | 26 | 2.27561716225072e-07 | 6.35658468 | intronic | CNTN5 | . | | | |
| 3M | rs148781275 | 11 | 103769875 | 5 | 2.2488967712367e-06 | 15.7111853 | intergenic | DYNC2H1;MIR4693 | dist=290012;dist=80031 | | | 1 |
| 3M | rs141281289 | 11 | 123822316 | 6 | 3.07596924681855e-07 | 15.811518 | intergenic | OR6M1;TMEM225 | dist=15967;dist=60603 | | | 1 |
| 3M | rs79539453 | 11 | 125396457 | 3 | 1.65356394618456e-06 | 20.8220616 | intronic | PKNOX2 | . | | | 1 |
| 3M | rs528140343 | 11 | 125849332 | 3 | 9.33313906476653e-07 | 18.2835521 | intergenic | PATE4;HYLS1 | dist=9260;dist=34282 | | | 1 |
| 3M | rs546409459 | 11 | 125885094 | 3 | 2.2842558352674e-06 | 18.9195474 | intronic | HYLS1 | . | | | |
| 3M | rs528609331 | 11 | 125972300 | 3 | 7.07196590950882e-07 | 19.5812418 | intronic | CDON | . | | | 1 |
| 3M | rs7104959 | 11 | 129976231 | 3 | 3.12645341825951e-06 | 17.9204611 | intronic | PRDM10 | . | | | 1 |
| 12M | rs117699122 | 12 | 451096 | 4 | 1.18257175082115e-06 | 21.4238256 | intergenic | CCDC77;B4GALNT3 | dist=8456;dist=8843 | | 1 | |
| 3M | rs189360484 | 12 | 1761344 | 4 | 3.94900202113111e-07 | 16.8078794 | intronic | ADIPOR2 | . | | | 1 |
| 12M | rs61918041 | 12 | 6572702 | 21 | 4.39893214125342e-06 | 7.97264643 | intronic | CHD4 | . | | 1 | |
| 12M | rs113244573 | 12 | 6575219 | 21 | 4.49284300484012e-06 | 7.97095746 | intronic | CHD4 | . | | | |
| 3M | rs141754456 | 12 | 19998198 | 7 | 1.82427554715572e-07 | 12.1460333 | intergenic | AEBP2;LINC02398 | dist=475971;dist=16487 | | | 1 |
| 3M | rs118184666 | 12 | 20271815 | 6 | 2.47036908420995e-06 | 10.9509803 | intergenic | LINC02468;PDE3A | dist=143914;dist=96722 | | | 1 |
| 3M | rs549931083 | 12 | 20363352 | 3 | 3.06794275866128e-07 | 14.3282694 | intergenic | LINC02468;PDE3A | dist=235451;dist=5185 | | | |
| 3M | rs151323346 | 12 | 20859090 | 4 | 2.59127051015029e-06 | 15.8903888 | intronic | SLCO1B3;SLCO1B3-SLCO1B7 | . | | | |
| 3M | rs371879555 | 12 | 22863028 | 3 | 1.69721932161597e-06 | 20.1033765 | intergenic | ETNK1;LOC101928441 | dist=172363;dist=312608 | | | 1 |
| 3M | rs183466664 | 12 | 26668754 | 4 | 3.57331936763708e-06 | 16.0181882 | intronic | ITPR2 | . | | | 1 |
| 3M | rs77353774 | 12 | 28095919 | 6 | 1.00362822173041e-06 | 13.1054362 | intergenic | PTHLH;LOC729291 | dist=123186;dist=89706 | | | 1 |
| 3M | rs113167689 | 12 | 28283029 | 6 | 2.31830826822625e-07 | 14.1364135 | intronic | CCDC91 | . | | | 1 |
| 3M | rs17510814 | 12 | 28316036 | 6 | 2.30925017950923e-07 | 14.0387924 | intronic | CCDC91 | . | | | |
| 3M | rs141756120 | 12 | 28358163 | 6 | 2.4075549984719e-07 | 14.278403 | intronic | CCDC91 | . | | | |
| 3M | rs117991215 | 12 | 28358540 | 6 | 2.50265389686764e-07 | 14.333666 | intronic | CCDC91 | . | | | |
| 3M | rs191930622 | 12 | 47890872 | 3 | 4.88013920852721e-06 | 19.6783579 | intronic | VDR | . | | | 1 |
| 3M | rs56302696 | 12 | 47899047 | 3 | 4.02736708601484e-06 | 19.8959041 | intronic | VDR | . | | | |
| 3M | rs185620578 | 12 | 48175616 | 3 | 2.41563563938621e-06 | 20.7232267 | intergenic | ASB8;CCDC184 | dist=18101;dist=8028 | | | 1 |
| 3M | rs190806532 | 12 | 48468364 | 3 | 1.54808652729946e-06 | 21.326955 | intergenic | ZNF641;ANP32D | dist=117118;dist=4195 | | | 1 |
| 3M | rs568658857 | 12 | 49460215 | 5 | 4.20762702895843e-06 | 14.2361193 | intronic | SPATS2 | . | | | 1 |

| | | | | | | | | | | | | | | |
|---|---|---|---|---|---|---|---|---|---|---|---|---|---|---|
| 3M | rs137880949 | 12 | 62912517 | 3 | 1.74526564511423e-07 | 21.6084034 | intronic | PPM1H | . | 1 | | | | |
| 3M | rs191053292 | 12 | 63051500 | 3 | 2.51400610111527e-08 | 22.9266488 | intergenic | PPM1H;AVPR1A | dist=116350;dist=91259 | | | | | |
| 3M | rs182437250 | 12 | 63214686 | 4 | 7.11750755969757e-07 | 18.8083451 | intergenic | AVPR1A;DPY19L2 | dist=63485;dist=344227 | | | | | |
| 3M | rs12315614 | 12 | 64527177 | 64 | 4.12213451917224e-06 | 3.77710662 | intergenic | TBK1;RASSF3 | dist=25064;dist=83318 | | | 1 | | |
| 12M | rs149616342 | 12 | 98785035 | 10 | 4.17235147721501e-06 | 11.2978214 | intronic | ANKS1B | . | | 1 | | | |
| 3M | rs76327548 | 12 | 100789188 | 11 | 2.03544908278967e-06 | 9.17842714 | intergenic | GAS2L3;ANO4 | dist=160900;dist=5588 | | | 1 | | |
| 3M | rs76904423 | 12 | 100794966 | 10 | 3.57310045627432e-06 | 9.80576231 | UTR5 | ANO4 | NM_001286615:c.-1068200>0;NM_178826:c.-1068200>0 | | | | | |
| 3M | rs180764936 | 12 | 101104843 | 3 | 1.48879381343194e-06 | 19.3473053 | intronic | ANO4 | . | | | | | |
| 3M | rs185855183 | 12 | 101111348 | 3 | 1.21523926305084e-06 | 18.7397727 | intronic | ANO4 | . | | | | | |
| 12M | rs566724618 | 13 | 22889700 | 3 | 1.0727068950386e-06 | 22.3433705 | ncRNA_intronic | LINC00621 | . | 1 | 1 | | | 1 |
| 12M | rs147221953 | 13 | 22911583 | 3 | 1.86149707525764e-06 | 22.5809676 | ncRNA_intronic | LINC00621 | . | | | | | |
| 3M | rs139598422 | 13 | 23312875 | 4 | 1.84258662797026e-08 | 20.4886522 | intronic | SGCG | . | | | | | |
| 3M | rs143371352 | 13 | 46809699 | 3 | 1.20142384300403e-07 | 20.5341625 | intergenic | ESD;HTR2A | dist=12538;dist=21847 | | | 1 | | |
| 3M | rs75186966 | 13 | 46821623 | 3 | 1.30324459401267e-07 | 20.2281345 | intergenic | ESD;HTR2A | dist=24462;dist=9923 | | | | | |
| 3M | rs150077525 | 13 | 57405415 | 7 | 1.65979661476121e-06 | 12.9984875 | intergenic | PRR20E;PCDH17 | dist=235197;dist=226329 | | | 1 | | |
| 3M | rs534845494 | 13 | 57639730 | 5 | 5.5584801687088e-07 | 15.9431765 | intronic | PCDH17 | . | | | | | |
| 3M | rs140062526 | 13 | 58711899 | 5 | 3.07703679852045e-06 | 14.0926088 | intergenic | LINC00374;DIAPH3 | dist=478782;dist=953688 | | | 1 | | |
| 12M | rs113791989 | 13 | 61344755 | 3 | 7.13417559871126e-07 | 25.2049582 | intergenic | MIR3169;PCDH20 | dist=144874;dist=64931 | | 1 | | | |
| 12M | rs532269430 | 13 | 73480315 | 5 | 9.38644328296013e-07 | 18.6173752 | intergenic | KLF5;LINC00392 | dist=402772;dist=83929 | | 1 | | | |
| 12M | rs368187808 | 13 | 73783371 | 3 | 1.40533873966635e-06 | 21.2991251 | intronic | KLF12 | . | | 1 | | | |
| 12M | rs372194899 | 13 | 73783373 | 3 | 1.40312572084772e-06 | 21.3012101 | intronic | KLF12 | . | | | | | |
| 3M | rs546286713 | 13 | 90758825 | 4 | 2.98443520289082e-06 | 15.7027841 | intergenic | LINC01049;LINC00410 | dist=223484;dist=132129 | | | 1 | | |
| 3M | rs567080482 | 13 | 94423151 | 4 | 2.95390135279034e-06 | 15.4999561 | intergenic | GPC6;DCT | dist=15132;dist=13660 | | | 1 | | |
| 3M | rs142928734 | 13 | 100948828 | 3 | 4.29382451218382e-06 | 17.009593 | ncRNA_intronic | NALCN | . | | | 1 | | |
| 3M | rs556680896 | 13 | 100950161 | 3 | 4.26743541179837e-06 | 17.0254659 | ncRNA_intronic | NALCN | . | | | | | |
| 3M | rs184265355 | 13 | 107359004 | 6 | 7.66559973381936e-07 | 14.4271719 | intronic | FAM155A | . | | | 1 | | |
| 3M | rs572961122 | 13 | 107360637 | 6 | 3.07761414918354e-06 | 13.3354896 | intronic | FAM155A | . | | | | | |
| 3M | rs528809914 | 13 | 112386942 | 3 | 2.27269814316061e-07 | 19.5048552 | intronic | SPACA7 | . | | | 1 | | |
| 3M | rs180765647 | 13 | 113724338 | 3 | 4.20477084480749e-06 | 20.1246521 | intronic | GRK1 | . | | | 1 | | |
| 3M | rs138215817 | 14 | 22173619 | 4 | 6.27722428502198e-07 | 16.311019 | intergenic | OR4E1;LOC105370401 | dist=502284;dist=206292 | | | 1 | | |
| 3M | rs74704551 | 14 | 29692681 | 3 | 1.97941397505543e-08 | 24.1990705 | intronic | PRKD1 | . | 1 | | | | |
| 3M | rs1686289 | 14 | 45791779 | 277 | 3.52212847732742e-06 | -2.2551539 | intergenic | LINC02303;LINC00871 | dist=76177;dist=272380 | | | 1 | | |
| 3M | rs176783 | 14 | 45811710 | 267 | 3.87832460630423e-06 | 2.28010147 | intergenic | LINC02303;LINC00871 | dist=96108;dist=252449 | | | | | |
| 3M | rs176786 | 14 | 45813767 | 268 | 3.37672684888795e-06 | 2.28475641 | intergenic | LINC02303;LINC00871 | dist=98165;dist=250392 | | | | | |
| 3M | rs428110 | 14 | 45825457 | 265 | 4.15182083903445e-06 | 2.28482299 | intergenic | LINC02303;LINC00871 | dist=109855;dist=238702 | | | | | |
| 3M | rs116862847 | 14 | 63674959 | 7 | 2.1204003875388e-07 | 15.3160369 | intergenic | WDR89;SGPP1 | dist=33088;dist=9258 | | | 1 | | |
| 3M | rs569916471 | 14 | 75424639 | 5 | 3.10710931450174e-06 | 15.6404088 | intergenic | LINC01220;JDP2 | dist=128231;dist=3085 | | | 1 | | |
| 3M | rs113767990 | 14 | 81251219 | 4 | 4.97864018338844e-06 | 14.5811155 | intergenic | LOC101928504;STON2 | dist=27839;dist=9431 | | | 1 | | |
| 3M | rs190251199 | 14 | 105124240 | 3 | 6.2650518901636e-07 | 20.0125292 | intergenic | LINC02298;JAG2 | dist=24744;dist=16758 | | 1 | 1 | 1 | |
| 12M | rs190251199 | 14 | 105124240 | 3 | 1.9790595982044E-06 | 22.6385061 | intergenic | LINC02298;JAG2 | dist=24744;dist=16758 | | | | | |
| 12M | rs187281112 | 15 | 32821186 | 5 | 3.3019635783423e-06 | 14.7343356 | intronic | FMN1 | . | | 1 | | | |
| 12M | rs56324718 | 15 | 39987872 | 9 | 7.5163534768267e-07 | 12.5117374 | intronic | EIF2AK4 | . | | 1 | | | |
| 3M | rs185155853 | 15 | 40951902 | 5 | 3.01500290127566e-06 | 15.9164657 | intergenic | DLL4;CHAC1 | dist=12829;dist=1569 | | | 1 | | |
| 3M | rs144026361 | 15 | 40956471 | 5 | 2.67002672054584e-06 | 16.1372994 | UTR3 | CHAC1 | NM_001142776:c.*6970>0;NM_024111:c.*6970>0 | | | | | |
| 3M | rs558614420 | 15 | 41518672 | 6 | 3.91771199682616e-06 | 13.9750831 | intronic | RPAP1 | . | | | 1 | | |
| 3M | rs138109686 | 15 | 41759244 | 6 | 4.62000628076245e-06 | 13.5956705 | intronic | MGA | . | | | 1 | | |
| 3M | rs145896760 | 15 | 41827024 | 6 | 4.80897800219791e-06 | 13.3688825 | UTR3 | MAPKBP1 | NM_014994:c.*15880>0;NM_001128608:c.*15880> | | | 1 | | |
| 3M | rs140642138 | 15 | 41832967 | 6 | 4.54678591939158e-06 | 13.5832221 | intronic | JMJD7;JMJD7-PLA2G4B | . | | | 1 | | |
| 3M | rs6080 | 15 | 58545734 | 36 | 4.85754782149583e-06 | 5.66476745 | intronic | LIPC | . | | | 1 | | |
| 3M | rs145439370 | 15 | 58587566 | 26 | 9.02072868608363e-07 | 6.89328891 | intergenic | LIPC;ADAM10 | dist=17722;dist=1243 | | | | | |
| 3M | rs149425014 | 15 | 58659461 | 22 | 1.84020904634255e-06 | 7.55224792 | intronic | ADAM10 | . | | | | | |
| 3M | rs146442492 | 15 | 58689916 | 24 | 3.24578114232023e-06 | 7.05925858 | intronic | ADAM10 | . | | | | | |

| | | | | | | | | | | | | | | |
|---|---|---|---|---|---|---|---|---|---|---|---|---|---|---|
| 3M | rs193253461 | 15 | 58937154 | 12 | 3.76206055565965e-07 | 10.5374468 | intergenic | SLTM;RNF111 | dist=3475;dist=50509 | | | | 1 | |
| 3M | rs184117160 | 15 | 59112107 | 11 | 1.25384693259541e-06 | 10.3417236 | intronic | CCNB2 | . | | | | 1 | |
| 3M | rs80292573 | 15 | 59142887 | 30 | 7.87214777000235e-07 | 6.55323491 | intronic | MYO1E | . | | | | 1 | |
| 3M | rs182303755 | 15 | 59342593 | 12 | 4.69869428218951e-07 | 10.5479917 | intronic | MYO1E | . | | | | | |
| 3M | rs138217865 | 15 | 93849653 | 4 | 1.29412742467123e-06 | 16.709641 | intergenic | LOC105370980;LINC02207 | dist=641605;dist=6907 | | | | 1 | |
| 12M | rs80212581 | 16 | 6362966 | 3 | 6.33070894509269e-07 | 22.2762003 | intronic | RBFOX1 | . | 1 | | | | |
| 12M | rs140276610 | 16 | 6383734 | 4 | 1.75581952039286e-06 | 22.1806577 | intronic | RBFOX1 | . | | | | | |
| 12M | rs138164904 | 16 | 6808238 | 3 | 3.60740717035677e-08 | 23.892641 | intronic | RBFOX1 | . | | | | | |
| 3M | rs553840536 | 16 | 25686574 | 3 | 3.61488566678171e-06 | 19.6203442 | intergenic | ZKSCAN2;HS3ST4 | dist=428729;dist=5385 | | | | 1 | |
| 3M | rs183817723 | 16 | 59268871 | 4 | 2.48366342700424e-06 | 17.5860507 | intergenic | GOT2;APOOP5 | dist=534555;dist=485270 | | | | 1 | |
| 3M | rs144954214 | 16 | 76145464 | 3 | 8.96489764315735e-07 | 21.8113263 | intergenic | CPHXL;CNTNAP4 | dist=418974;dist=131937 | | | | 1 | |
| 3M | rs529523094 | 16 | 77681654 | 3 | 1.09798611725829e-06 | 18.1584346 | intergenic | ADAMTS18;NUDT7 | dist=246620;dist=40838 | | | | 1 | |
| 12M | rs116897913 | 17 | 16935848 | 3 | 4.78180386603079e-06 | 20.1396765 | intergenic | TBC1D27P;TNFRSF13B | dist=2672;dist=3233 | | | 1 | | |
| 3M | rs146728064 | 17 | 19362127 | 6 | 3.79796041699169e-06 | 12.6400411 | intronic | B9D1 | . | | | | 1 | |
| 12M | rs188353596 | 17 | 48001236 | 3 | 6.85080774831596e-07 | 21.8057947 | intergenic | CDK5RAP3;COPZ2 | dist=19450;dist=24931 | | | 1 | | |
| 3M | rs184613584 | 17 | 50430860 | 6 | 5.40834238559884e-07 | 14.1377974 | intronic | ACSF2 | . | | | | 1 | |
| 3M | rs191271637 | 17 | 54045899 | 3 | 2.76242881040307e-06 | 18.379836 | intergenic | KIF2B;TOM1L1 | dist=220706;dist=854792 | | | | 1 | |
| 12M | rs112148840 | 17 | 68237633 | 3 | 3.82795090389498e-06 | 24.0241075 | intronic | AMZ2 | . | | | 1 | | |
| 12M | rs2606194 | 17 | 79214741 | 44 | 9.99647458522552e-07 | -5.6956979 | intronic | RBFOX3 | . | | | 1 | | |
| 12M | rs147669485 | 18 | 29023686 | 4 | 9.8819660331789e-08 | 22.1107639 | intergenic | CDH2;MIR302F | dist=846556;dist=1275226 | | | 1 | 1 | 1 |
| 3M | rs185819304 | 18 | 29421615 | 3 | 4.31513948321026e-07 | 19.9401113 | intergenic | CDH2;MIR302F | dist=1244486;dist=877296 | | | | | |
| 3M | rs187942235 | 18 | 29450465 | 3 | 2.77341075652478e-07 | 20.3968658 | intergenic | CDH2;MIR302F | dist=1273336;dist=848446 | | | | | |
| 3M | rs139493286 | 18 | 31236056 | 3 | 3.23770514196012e-06 | 17.4603093 | intergenic | DSC1;DSG1 | dist=73200;dist=82104 | | | | 1 | |
| 3M | rs143538552 | 18 | 31470299 | 3 | 1.40612679764676e-07 | 19.1457218 | intronic | DSG3 | . | | | | | |
| 3M | rs373746073 | 18 | 31478421 | 3 | 1.55466285461099e-07 | 19.0847197 | UTR3 | DSG3 | NM_001944:c.*21610>0 | | | | | |
| 3M | rs146333745 | 18 | 57830225 | 4 | 1.04499182739545e-06 | 17.4215784 | intergenic | ATP8B1;NEDD4L | dist=26910;dist=214001 | | | | 1 | |
| 12M | rs559152067 | 18 | 68362375 | 3 | 1.80839131277764e-06 | 24.7443692 | intergenic | LOC643542;TMX3 | dist=462756;dist=311313 | | | 1 | | |
| 3M | rs185464792 | 19 | 18686561 | 3 | 5.22729611714582e-07 | 22.5896015 | intronic | CRTC1 | . | | | | 1 | |
| 3M | rs186768950 | 19 | 18695314 | 3 | 4.67656376928778e-07 | 22.7152061 | intronic | CRTC1 | . | | | | | |
| 3M | rs541288561 | 19 | 18758635 | 5 | 6.19920480169239e-07 | 15.6119012 | intronic | CRTC1 | . | | | | | |
| 3M | rs559008174 | 19 | 18765249 | 5 | 6.10577196408926e-07 | 15.6025661 | intronic | CRTC1 | . | | | | | |
| 3M | rs570407448 | 19 | 18769220 | 5 | 7.02455289936315e-07 | 15.4977984 | intronic | CRTC1 | . | | | | | |
| 3M | rs546144116 | 19 | 19452530 | 3 | 2.29332366104571e-07 | 23.1354815 | intronic | GATAD2A | . | | | | 1 | |
| 3M | rs560206697 | 19 | 20546292 | 3 | 4.01812636832912e-08 | 24.9989943 | intronic | ZNF737 | . | | 1 | | | |
| 3M | rs111285015 | 19 | 22940396 | 3 | 3.3078789506342e-09 | 27.3347642 | intergenic | ZNF723;ZNF728 | dist=81729;dist=34487 | | 1 | | | |
| 3M | rs1008091735 | 19 | 30599192 | 3 | 5.61396767442543e-07 | 18.4689243 | intronic | ZNF536 | . | | | | 1 | |
| 3M | rs148433854 | 19 | 30605571 | 3 | 4.40546670794092e-07 | 18.6267585 | intronic | ZNF536 | . | | | | | |
| 12M | rs367732718 | 19 | 35427102 | 3 | 3.38991794276911e-06 | 23.8784923 | intergenic | LINC01531;FFAR2 | dist=10262;dist=21155 | | | 1 | | |
| 12M | rs562831582 | 19 | 50731053 | 4 | 5.70199111188332e-07 | 21.0856548 | intergenic | CLEC11A;GPR32 | dist=5345;dist=39411 | | | 1 | | |
| 12M | rs62192733 | 20 | 1947511 | 7 | 2.76211842636971e-06 | 15.744618 | ncRNA_exonic | PDYN | . | | | 1 | | |
| 12M | rs547186621 | 20 | 6126094 | 4 | 2.39683963851735e-06 | 21.5510829 | intergenic | FERMT1;CASC20 | dist=3064;dist=300638 | | | 1 | | |
| 3M | rs2327968 | 20 | 15832846 | 21 | 4.58567809597369e-06 | 6.15899856 | intronic | MACROD2 | . | | | | 1 | |
| 3M | rs2876414 | 20 | 15833059 | 19 | 4.41048907235012e-06 | 6.81910907 | intronic | MACROD2 | . | | | | | |
| 3M | rs140788628 | 20 | 15877856 | 8 | 2.32626831948532e-07 | 11.8039284 | intronic | MACROD2 | . | | | | | |
| 3M | rs559228693 | 20 | 15982684 | 5 | 3.15016069009906e-06 | 14.3643551 | ncRNA_intronic | LOC613266 | . | | | | | |
| 12M | rs138733283 | 20 | 32087884 | 5 | 5.87309369634527e-07 | 18.2744744 | intronic | HCK | . | | | 1 | | |
| 12M | rs149859280 | 20 | 32090367 | 5 | 5.71700141373349e-07 | 18.3057707 | intronic | HCK | . | | | | | |
| 12M | rs146249289 | 20 | 32094428 | 4 | 2.01850841331451e-07 | 19.6109527 | intronic | HCK | . | | | | | |
| 12M | rs145791959 | 20 | 32134626 | 5 | 8.67001886662025e-07 | 17.9091953 | intronic | TM9SF4 | . | | | 1 | | |
| 12M | rs193041547 | 20 | 32184077 | 4 | 1.80079654856719e-07 | 19.6791358 | intergenic | TM9SF4;TSPY26P | dist=16819;dist=5069 | | | | | |
| 12M | rs138055631 | 20 | 32192841 | 5 | 1.06243999550455e-06 | 17.6647762 | UTR3 | PLAGL2 | NM_002657:c.*36110>0 | | | | | |
| 12M | rs145421321 | 20 | 32274323 | 6 | 1.72232779371331e-06 | 17.0212611 | intergenic | POFUT1;KIF3B | dist=35665;dist=3328 | | | | | |

| | | | | | | | | | | | | |
|---|---|---|---|---|---|---|---|---|---|---|---|---|
| 12M | rs143432612 | 20 | 32304972 | 6 | 1.74726298083525e-06 | 16.9387232 | intronic | KIF3B | . | | | |
| 12M | rs139816293 | 20 | 32333540 | 6 | 1.16528363587405e-06 | 17.1384248 | UTR3 | KIF3B | NM_004798:c.*22210>0 | | | |
| 12M | rs200198574 | 20 | 32358851 | 6 | 2.44213593266147e-06 | 16.5282735 | intronic | ASXL1 | . | | 1 | |
| 12M | rs148157126 | 20 | 32361036 | 5 | 1.42669073977956e-07 | 19.7865001 | intronic | ASXL1 | . | | | |
| 12M | rs192855100 | 20 | 32520938 | 4 | 4.17301607264236e-07 | 20.1986097 | intronic | NOL4L | . | | 1 | |
| 3M | rs557092705 | 20 | 35601989 | 3 | 4.54067714932956e-06 | 19.4769546 | ncRNA_exonic | FER1L4 | . | | | 1 |
| 12M | rs2427460 | 20 | 62959430 | 420 | 5.30063377285508e-07 | -2.574044 | intronic | SLC17A9 | . | | 1 | |
| 3M | rs184785969 | 21 | 15799112 | 3 | 8.26040996676763e-08 | 19.8736486 | intronic | USP25 | . | 1 | | |
| 3M | rs117280553 | 21 | 15834844 | 3 | 5.79538719308743e-08 | 20.087417 | intronic | USP25 | . | | | |
| 3M | rs79486609 | 21 | 15872687 | 3 | 4.53094466952756e-08 | 20.6624886 | intronic | USP25 | . | | | |
| 3M | rs73227413 | 21 | 21764653 | 28 | 2.74560920737872e-06 | 5.96205514 | ncRNA_intronic | LINC01425 | . | | | |
| 3M | rs75024143 | 21 | 21784226 | 12 | 2.0964109268482e-06 | 9.81254241 | ncRNA_intronic | LINC01425 | . | | | |
| 3M | rs192134381 | 21 | 22078395 | 3 | 5.23445134161758e-09 | 21.788812 | ncRNA_intronic | LINC01687 | . | 1 | | |
| 3M | rs118183140 | 21 | 34105187 | 17 | 1.16709964426479e-06 | 7.5893248 | UTR3 | SLC5A3 | NM_006933:c.*78320>0 | | | 1 |
| 3M | rs183586634 | 21 | 37390730 | 5 | 1.72123504106205e-06 | 14.3132202 | intronic | DYRK1A | . | | | 1 |
| 3M | rs117185941 | 21 | 37394182 | 5 | 1.2357093777619e-06 | 15.3808786 | intronic | DYRK1A | . | | | |
| 3M | rs118084887 | 21 | 37491518 | 5 | 1.91359719468201e-06 | 14.9382616 | intronic | DYRK1A | . | | | |
| 12M | rs9636964 | 21 | 39932840 | 115 | 2.72523192147064e-07 | -4.2543192 | intergenic | PCP4;DSCAM | dist=3448;dist=78161 | | 1 | |
| 12M | rs9305683 | 21 | 39933795 | 112 | 8.57246540213203e-07 | -4.1163679 | intergenic | PCP4;DSCAM | dist=4403;dist=77206 | | | |
| 12M | rs9974985 | 21 | 39935648 | 114 | 1.59836223443899e-07 | -4.3459395 | intergenic | PCP4;DSCAM | dist=6256;dist=75353 | | | |
| 12M | rs7275595 | 21 | 39935998 | 113 | 3.24552446398997e-07 | -4.2609754 | intergenic | PCP4;DSCAM | dist=6606;dist=75003 | | | |
| 12M | rs1005412 | 21 | 39937023 | 112 | 1.75853470115381e-07 | -4.4521096 | intergenic | PCP4;DSCAM | dist=7631;dist=73978 | | | |
| 12M | rs9981433 | 21 | 39937640 | 116 | 1.14042775714184e-06 | -4.1220124 | intergenic | PCP4;DSCAM | dist=8248;dist=73361 | | | |
| 3M | rs150539922 | 21 | 41856807 | 3 | 4.79442196264294e-06 | 16.6696937 | intronic | PRDM15 | . | | | 1 |
| 3M | rs113625788 | 22 | 19981659 | 7 | 1.17014602462676e-06 | 11.8084829 | exonic | ARVCF | . | | | 1 |
| 3M | rs78547898 | 22 | 32428291 | 3 | 4.85417555255089e-07 | 20.9098395 | intronic | BPIFC | . | | | 1 |
| 12M | rs77297738 | 22 | 34077554 | 16 | 2.01836979224915e-07 | 10.2480572 | intergenic | LARGE1;LINC02885 | dist=154731;dist=679113 | | 1 | |
| 12M | rs74572772 | 22 | 34096660 | 16 | 1.39505561568774e-07 | 10.2225183 | intergenic | LARGE1;LINC02885 | dist=173837;dist=660007 | | | |
| 12M | rs80019988 | 22 | 34097658 | 16 | 1.52143693387141e-07 | 10.0623306 | intergenic | LARGE1;LINC02885 | dist=174835;dist=659009 | | | |
| 3M | rs541680196 | 22 | 40132086 | 5 | 1.06912975962765e-06 | 14.3867808 | intronic | TNRC6B | . | | | 1 |
| 3M | rs185139807 | 22 | 40198777 | 5 | 1.02186831937043e-06 | 14.4330526 | intronic | TNRC6B | . | | | |
| 3M | rs141127122 | 22 | 40208435 | 4 | 3.1593298841889e-06 | 15.3824875 | intronic | TNRC6B | . | | | |
| 3M | rs148998974 | 22 | 40224526 | 5 | 8.65242968473446e-07 | 14.5129532 | intronic | TNRC6B | . | | | |
| 3M | rs555040883 | 22 | 40235472 | 4 | 3.05234476142884e-06 | 15.4273069 | intronic | TNRC6B | . | | | |
| 3M | rs182959028 | 22 | 45427152 | 7 | 2.37697466245549e-06 | 11.944461 | intronic | RIBC2 | . | | | 1 |
| 3M | rs150946694 | 22 | 46457283 | 4 | 3.3001729159907e-06 | 15.3082479 | intronic | CELSR1 | . | | | 1 |

| Combined Results | | | | |
|---|---|---|---|---|
| GENOME-WIDE RISK LOCI | 2 | 24 | | |
| SUGGESTIVE RISK LOCI (not genome-wide) | | | 74 | 200 |
| SUGGESTIVE RISK LOCI (including genome-wide) | | | 76 | 224 |
| TOTAL GENOME-WIDE & SUGGESTIVE RISK LOCI | | | | 274 |
| MERGED (Sum minus overlap) | | 26 | | 263 |

Overlapping Risk Loci

ALK, DCANP1, JAG2, KCND3, PPARGC1A, STAG1, TBLXR1 — OVERLAPPING RISK LOCI 7

AGTR1, CDH2, ETAA1, MTX2, SGCG — 4

| | |
|---|---|
| GENOME-WIDE SIGNIFICANCE SNPS | 45 |
| SUGGESTIVE SNPS | 498 |
| TOTAL SNPS | 543 |
| SNPS REPEATED 12&3 MO | 11 |
| Risk loci that cluster these 11 SNPs | 7 |
