## Supplementary material for "Pharmacogenomics of steroid-induced ocular hypertension: relationship to high-tension glaucomas and new pathophysiologic insight": Suppl Table S3

**Supplementary Table S3. Co-localization with Other High-Tension Ocular Phenotypes**

| Abbreviation or Acronym | | Reference number |
|---|---|---|
| POAG MA | primary open angle glaucoma, multi-ancestry | Han et al. (72); Lo Faro et al. (83) |
| POAG AF | primary open angle glaucoma, African ancestry | Verma et al. (73) |
| PACG | primary angle closure glaucoma | Vithana et al. (84) |
| PEXS/G | pseudoexfoliation syndrome/glaucoma | Zagajewska et al. (86); Krumbiegel et al. (87) |
| IOP | intraocular pressure | Han et al. (72) |
| CCT | central corneal thickness | Iglesias et al. (74); Choquet et al. (77); Igo et al. (162) |
| Stickler | Type II Stickler Syndrome | |

**Footnotes**

*subthreshold P value

**protective effect

**Count**

| | |
|---|---|
| 30.77% | risk loci containing SNPs of genome-wide significance |
| 18.70% | total risk loci |

| Gene | P value for top SNP of genome-wide significance in discovery cohort | High-tension ocular phenotype | | | | | Endophenotype | |
|---|---|---|---|---|---|---|---|---|
| | | POAG MA | POAG AF | PACG | PEXS/G | Stickler | IOP | CCT |
| *ADGRL3* | | X | *X | | | | *X | |
| *AHCTF1* | | | *X | | | | | |
| *ALK* | | X | | | | | X | |
| *ANO4* | | | *X | | | | | |
| *AP1AR* | | | *X | | | | | |
| *ARAP2* | | | *X | | | | | |
| *ARHGAP21* | | X | *X | | | | | |
| *ATXN1* | | | | | | | | X |
| *B4GALNT3* | | | *X | | | | | |
| *BEND7* | 1.19E-09 | | *X | | | | | |
| *CAV2* | | X | *X | | | | X | |
| *CCSER1* | | | *X | | | | | |
| *CFTR* | | X | | | | | X | |
| *CNTNAP2* | | | | | X | | | |
| *CNTN4* | | | *X | | | | | |
| *CNTN5* | | | *X | | | | | |
| *COL11A1* | 3.65E-09 | X | | X | *X | X | X | |
| *CRPPA* | | | *X | | | | | |
| *CSMD1* | 1.07E-08 | | *X | | | | | |
| *EIF2AK4* | | | *X | | | | | |
| *EYA2* | | X | | | | | | |
| *FST* | 1.40E-08 | | | | | | | X |
| *GMNC* | | | *X | | | | | |
| *HMCN1* | | X | | | | | | |
| *INTU* | | X | | | | | *X | |
| *ITPR2* | | X | | | | | | |
| *KCNIP4* | | | *X | | | | | |
| *KLF5* | | X | | | | | X | |
| *LARGE1* | | | *X | | | | | |
| *MACROD2* | | | *X | | | | | |
| *MAML3* | | | *X | | | | | |
| *MVB12B* | | X | | | | | X | |
| *NALCN* | | | *X | | | | | |
| *NCAM2* | | | *X | | | | | |
| *NIBAN1* | | | *X | | | | | |
| *NRXN1* | | | *X | | | | | |
| *PLXDC2* | | X | *X | | | | X | |
| *PPM1H* | 2.51E-08 | | *X | | | | | |
| *RBFOX1* | 3.60E-08 | | *X | | **X | | | |
| *RBFOX3* | | | *X | | | | | |
| *SCHIP1* | | | *X | | | | | |
| *SGCG* | 1.84E-08 | | | | | | | X |
| *SPRED2* | | X | | | | | X | |
| *STAG1* | | | | | | | | X |
| *RASSF3* | | | *X | | | | | |
| *TFEC* | | X | | | | | | |
| *THSD7A* | | X | *X | | | | X | |
| *TNARC6B* | | | *X | | | | | |
| *TRIB2* | | X | | | | | X | |
| *WWC1* | 3.96E-09 | | *X | | | | | |
