## Supplementary material for "Pharmacogenomics of steroid-induced ocular hypertension: relationship to high-tension glaucomas and new pathophysiologic insight": Suppl Table S4

**Supplementary Table S4. Target Gene Prioritization**

**SNP-associated genes that are aqueous outflow pathway-expressed**

### NOTES

Top SNPs identified with the 12M and 3M QTs are merged, then clustered into risk loci by sorting by chromosomal position, then by chromosome

SNPs duplicated between 12 month and 3 month QT are boxed. Loci that cluster SNPs from both the 12 and 3 month QTs are boxed and highlighted in blue

SNPs of genome-wide significance are highlighted in gray (column rsid) as are their p-values (column Score.pval)

Risk loci into which multiple SNPs cluster are boxed (column Gene.refGene) and those containing SNPs of genome-wide significance are shaded in gray

**HEADERS**

QT: quantitative trait; rsid: reference SNP cluster ID; chr: chromosome; pos_38: position of SNP on GRCh38 reference panel; Score.pval: p-value; Est: effect size (mm Hg);

Func.refGene: SNP location with respect to nearest gene(s); Gene.refGene: nearest gene upstream and downstream; GeneDetail.refGene: distance to nearest gene upstream and downstream;

Gene.refGene AOPe: Gene.refGenes that are aqueous outflow pathway (AOP)-expressed (as viewed on Spectacle);

up;downstream AOPe: next closest aqueous outflow pathway expressed gene(s) located upstream and downstream of the closest gene(s) were not AOPe;

e/sQTL AOPe: target gene(s) of an eQTL or sQTL if also AOPe; Count GC-regulated genes: columns adding up all AOPe genes for which evidence was found that they are GC-regulated, totals at bottom;

GC-DEG TM cells: glucocorticoid-regulated gene in TM cell dataset; GC-Regulated in silico: column adding up all glucocorticoid-regulated gene in DAVID dataset, total at bottom

Count new: Genes in DAVID dataset not already identified in TM cell dataset; Count overlap: column adding up all glucocorticoid-regulated genes in both TM cell and DAVID dataset, total at bottom

| QT | rsid | chr | pos_38 | Score.pval | Est | Func.refGene | Gene.refGene | GeneDetail.refGene | Gene.refGene AOP-expressed | up;downstream AOP-expressed |
|---|---|---|---|---|---|---|---|---|---|---|
| 3M | rs115348382 | 1 | 9595845 | 2.20695702237544e-06 | 18.2107495 | intronic | TMEM201 | . | TMEM201 | |
| 3M | rs186532456 | 1 | 18621834 | 9.60875608166652e-07 | 20.1730343 | intergenic | KLHDC7A;PAX7 | dist=135848;dist=9012 | KLHDC7A | |
| 3M | rs562032622 | 1 | 18632839 | 1.12975575994446e-06 | 21.0267115 | intronic | PAX7 | . | | |
| 12M | rs35145334 | 1 | 23138629 | 2.09882278975154e-06 | 10.6552656 | intronic | LUZP1 | . | LUZP1 | |
| 3M | rs2365739 | 1 | 62018790 | 1.81260643542989e-06 | 6.86815917 | intronic | PATJ | . | | TM2D1;L1TD1 |
| 12M | rs17127656 | 1 | 65477788 | 1.63996682822861e-06 | 5.07046538 | intronic | LEPR | . | LEPR | |
| 12M | rs7518849 | 1 | 65483108 | 1.67668772877101e-06 | 5.07456956 | intronic | LEPR | . | | |
| 12M | rs11579567 | 1 | 65491458 | 2.1300143111827e-06 | 5.02973793 | intronic | LEPR | . | | |
| 12M | rs7534177 | 1 | 65500037 | 2.85450525546462e-06 | 4.96123378 | intronic | LEPR | . | | |
| 3M | rs149493615 | 1 | 79414404 | 1.38822331670427e-06 | 11.750632 | intergenic | ADGRL4;LINC01781 | dist=407674;dist=1121351 | ADGRL4 | |
| 3M | rs143811231 | 1 | 79502446 | 1.738654939087e-06 | 11.5747409 | intergenic | ADGRL4;LINC01781 | dist=495716;dist=1033309 | | |
| 12M | rs139390630 | 1 | 84400513 | 2.86874207705929e-06 | 11.6975971 | intronic | DNASE2B | . | DNASE2B | |
| 12M | rs114067899 | 1 | 84725361 | 2.17218533160816e-06 | 13.7760673 | intergenic | SSX2IP;LPAR3 | dist=34893;dist=86241 | SSX2IP;LPAR3 | |
| 3M | rs34270375 | 1 | 88905019 | 9.34314238699532e-07 | 7.71147017 | intergenic | GTF2B;KYAT3 | dist=13452;dist=30754 | GTF2B | |
| 3M | rs187518659 | 1 | 98990189 | 2.15952482477188e-06 | 11.6819623 | intronic | PLPPR5 | . | PLPPR5 | |
| 3M | rs140420703 | 1 | 102337928 | 1.91423564672595e-06 | 16.487654 | intergenic | OLFM3;COL11A1 | dist=340694;dist=538539 | OLFM3;COL11A1 | |
| 3M | rs563167766 | 1 | 102400809 | 4.72618849175033e-06 | 19.415739 | intergenic | OLFM3;COL11A1 | dist=403575;dist=475658 | | |
| 3M | rs77180278 | 1 | 102496326 | 9.33877360643494e-08 | 7.33619996 | intergenic | OLFM3;COL11A1 | dist=499092;dist=380141 | | |
| 3M | rs112351653 | 1 | 102754804 | 2.54962936189828e-08 | 7.42386947 | intergenic | OLFM3;COL11A1 | dist=757570;dist=121663 | | |
| 3M | rs180926150 | 1 | 102760770 | 3.76640865110522e-06 | 19.641918 | intergenic | OLFM3;COL11A1 | dist=763536;dist=115697 | | |
| 3M | rs114413507 | 1 | 102953612 | 2.57350301189632e-08 | 7.42216029 | intronic | COL11A1 | . | | |
| 3M | rs116672066 | 1 | 103007360 | 4.45932234807969e-09 | 8.04769476 | intronic | COL11A1 | . | | |
| 3M | rs111928960 | 1 | 103168079 | 3.65396910662882e-09 | 8.60257208 | intergenic | COL11A1;LOC101928436 | dist=59557;dist=325967 | | |
| 3M | rs113221952 | 1 | 103288418 | 9.173975220783e-07 | 7.92350704 | intergenic | COL11A1;LOC101928436 | dist=179896;dist=205628 | | |
| 3M | rs1856085 | 1 | 103571923 | 4.63628412408432e-07 | 21.51144 | intronic | AMY2B | . | | |
| 3M | rs143597860 | 1 | 103614521 | 4.52300265269257e-07 | 21.5748154 | intergenic | AMY2B;AMY2A | dist=34987;dist=2811 | AMY2B;AMY2A | |
| 3M | rs144541665 | 1 | 103768107 | 3.71844012409788e-07 | 21.8734611 | intergenic | AMY1C;LOC100129138 | dist=9415;dist=304916 | | |
| 3M | rs76098744 | 1 | 111806750 | 1.19916376071606e-06 | 8.38612452 | intronic | KCND3 | . | KCND3 | |
| 12M | rs76098744 | 1 | 111806750 | 2.92647184366499E-06 | 9.5685657 | intronic | KCND3 | . | | |
| 3M | rs74683551 | 1 | 111811796 | 1.2121497054257e-06 | 8.38252043 | intronic | KCND3 | . | | |
| 12M | rs74683551 | 1 | 111811796 | 3.08074322019196E-06 | 9.54722832 | intronic | KCND3 | . | | |
| 3M | rs76617932 | 1 | 180961288 | 2.78866978177701e-06 | 10.5449608 | intergenic | KIAA1614;STX6 | dist=6401;dist=11426 | KIAA1614;STX6 | |
| 3M | rs183180157 | 1 | 181277985 | 2.7248136592675E-06 | 12.61386 | intergenic | LINC01699;CACNA1E | dist=39381;dist=205532 | | |
| 3M | rs375790303 | 1 | 184561348 | 4.76779314562933e-06 | 11.4240393 | intronic | C1orf21 | . | C1orf21 | |
| 3M | rs138480898 | 1 | 184986525 | 3.67316732025395e-06 | 16.9873974 | intergenic | NIBAN1;LINC01633 | dist=12017;dist=15002 | | EDEM3;RNF2 |
| 3M | rs145766563 | 1 | 185160370 | 1.9043145410504e-06 | 16.8682102 | intronic | SWT1 | . | SWT1 | |
| 3M | rs147032554 | 1 | 186179732 | 6.67557909700165e-07 | 18.162609 | intronic | HMCN1 | . | HMCN1 | |
| 3M | rs180989936 | 1 | 193075048 | 3.47790847768381E-06 | 17.1793579 | intronic | RO60 | . | | |
| 12M | rs145804766 | 1 | 202153371 | 1.43417032447652e-06 | 12.5864465 | intronic | PTPN7 | . | PTPN7 | |
| 3M | rs10494861 | 1 | 205362746 | 2.48126631931009e-06 | 19.1820498 | intergenic | KLHDC8A;LEMD1 | dist=5656;dist=10506 | KLHDC8A;LEMD1 | |
| 12M | rs147869916 | 1 | 227119951 | 3.36060694276516e-06 | 17.5964551 | intronic | CDC42BPA | . | CDC42BPA | |
| 3M | rs2274996 | 1 | 229668791 | 6.73601821695854e-07 | 5.58527433 | intergenic | URB2;LINC01682 | dist=8591;dist=206759 | URB2 | |
| 3M | rs2274997 | 1 | 229668899 | 6.64244186267036e-07 | 5.58731228 | intergenic | URB2;LINC01682 | dist=8699;dist=206651 | | |
| 3M | rs2891865 | 1 | 229670621 | 6.80112526815562e-07 | 5.58181376 | intergenic | URB2;LINC01682 | dist=10421;dist=204929 | | |
| 3M | rs2385790 | 1 | 229671745 | 6.81207031237396e-07 | 5.58224822 | intergenic | URB2;LINC01682 | dist=11545;dist=203805 | | |
| 3M | rs12024557 | 1 | 229676610 | 6.90833131537333e-07 | 5.56930969 | intergenic | URB2;LINC01682 | dist=16410;dist=198940 | | |
| 3M | rs4562666 | 1 | 229689023 | 7.35045143592774e-07 | 5.51922599 | intergenic | URB2;LINC01682 | dist=28823;dist=186527 | | |
| 3M | rs12036586 | 1 | 229690631 | 1.08649183321501e-06 | 5.22212176 | intergenic | URB2;LINC01682 | dist=30431;dist=184919 | | |
| 3M | rs16850124 | 1 | 229695584 | 2.19845550423288e-06 | 5.18095662 | intergenic | URB2;LINC01682 | dist=35384;dist=179966 | | |
| 3M | rs12045643 | 1 | 229698303 | 7.37332600021845e-07 | 5.48149152 | intergenic | URB2;LINC01682 | dist=38103;dist=177247 | | |
| 3M | rs559559983 | 1 | 246813830 | 1.8701710647004e-06 | 13.811132 | intergenic | LINC01341;AHCTF1 | dist=22344;dist=25268 | AHCTF1 | |
| 3M | rs550763536 | 2 | 7312216 | 3.04128022440287e-06 | 14.4871647 | intergenic | LOC101929452;LOC100506274 | dist=234336;dist=109045 | | RNF144A;ID2 |
| 12M | rs76220567 | 2 | 12919819 | 2.07255893928856e-06 | 20.6828868 | intergenic | TRIB2;LOC100506474 | dist=177087;dist=46963 | TRIB2 | |
| 3M | rs558553658 | 2 | 15542600 | 2.00500271573415e-07 | 17.1099107 | intronic | NBAS | . | NBAS | |
| 3M | rs536781978 | 2 | 29510815 | 2.20535447064362e-06 | 20.5561882 | intronic | ALK | . | ALK | |
| 12M | rs536781978 | 2 | 29510815 | 2.20535447064362E-06 | 20.5561882 | intronic | ALK | . | | |
| 3M | rs568321148 | 2 | 29647482 | 2.03643401922156e-06 | 20.7400203 | intronic | ALK | . | | |
| 12M | rs568321148 | 2 | 29647482 | 2.03643401922156E-06 | 20.7400203 | intronic | ALK | . | | |
| 12M | rs186792608 | 2 | 42869559 | 1.57557414570447e-07 | 22.4512499 | intergenic | HAAO;LINC01819 | dist=76976;dist=158293 | HAAO | |
| 3M | rs76777840 | 2 | 48085811 | 7.02623491851785e-07 | 14.3500784 | intergenic | FBXO11;FOXN2 | dist=179313;dist=228488 | FBXO11;FOXN2 | |
| 3M | rs145080832 | 2 | 48246004 | 1.37120287877638e-07 | 17.1618477 | intergenic | FBXO11;FOXN2 | dist=339506;dist=68295 | | |
| 3M | rs184220112 | 2 | 48404604 | 3.11727344916156e-07 | 16.4879043 | intergenic | FOXN2;PPP1R21 | dist=25309;dist=36162 | FOXN2;PPP1R21 | |

| | | | | | | | | | | |
|---|---|---|---|---|---|---|---|---|---|---|
| 3M | rs189890455 | 2 | 48428258 | 4.1971806049668e-07 | 15.1467391 | intergenic | FOXN2;PPP1R21 | dist=48963;dist=12508 | | |
| 3M | rs181193202 | 2 | 52300216 | 1.62222587830176e-07 | 19.6312088 | ncRNA_intronic | LOC730100 | . | | NRXN1;ASB3 |
| 3M | rs183378658 | 2 | 52336233 | 1.16662586966659e-07 | 19.7446169 | ncRNA_intronic | LOC730100 | . | | |
| 3M | rs190193113 | 2 | 52858640 | 1.13301212531008e-07 | 19.9533602 | intergenic | MIR4431;ASB3 | dist=156025;dist=811339 | | |
| 3M | rs187520610 | 2 | 53132903 | 3.93234514941546e-09 | 18.8010396 | intergenic | MIR4431;ASB3 | dist=430288;dist=537076 | ASB3 | |
| 3M | rs149421869 | 2 | 53256291 | 2.05046156636639e-06 | 11.7382186 | intergenic | MIR4431;ASB3 | dist=553676;dist=413688 | | |
| 3M | rs146479102 | 2 | 65598625 | 2.18341304839895e-06 | 15.4846768 | intergenic | SPRED2;MIR4778 | dist=166026;dist=759622 | SPRED2 | |
| 3M | rs528288879 | 2 | 65648796 | 5.421960882917e-07 | 14.7878085 | intergenic | SPRED2;MIR4778 | dist=216197;dist=709451 | | |
| 3M | rs11690187 | 2 | 67338777 | 4.61292753502872e-06 | 13.6310532 | intergenic | LINC01828;ETAA1 | dist=49533;dist=58556 | ETAA1 | |
| 3M | rs151272830 | 2 | 67455592 | 2.85197668301622e-06 | 13.3323479 | intergenic | ETAA1;LINC01812 | dist=43503;dist=340462 | | |
| 3M | rs186142189 | 2 | 67475575 | 2.69645566407293e-06 | 12.7361927 | intergenic | ETAA1;LINC01812 | dist=63486;dist=320479 | | |
| 12M | rs2902021 | 2 | 67598553 | 3.30567771883221e-06 | 4.55991411 | intergenic | ETAA1;LINC01812 | dist=186464;dist=197501 | | |
| 12M | rs75082290 | 2 | 67604021 | 1.699715195267e-06 | 4.69395387 | intergenic | ETAA1;LINC01812 | dist=191932;dist=192033 | | |
| 12M | rs191298981 | 2 | 67882914 | 7.69651662192921e-07 | 24.0078714 | intergenic | LINC01812;C1D | dist=57352;dist=158216 | C1D | |
| 12M | rs113537164 | 2 | 68007798 | 1.017020860936e-06 | 20.3890413 | intergenic | LINC01812;C1D | dist=182236;dist=33332 | | |
| 12M | rs113154814 | 2 | 68062365 | 1.66384008676077e-07 | 19.8310188 | intronic | C1D | . | | |
| 3M | rs184200893 | 2 | 69033781 | 2.24020739771759E-06 | 19.9580356 | intronic | ANTXR1 | . | | |
| 3M | rs111927235 | 2 | 74256827 | 8.4250114725101e-07 | 17.0468955 | intronic | SLC4A5 | . | SLC4A5 | |
| 3M | rs111838310 | 2 | 74446364 | 3.26261179053175e-07 | 17.7963938 | intergenic | RTKN;INO80B-WBP1 | dist=4427;dist=8659 | RTKN | |
| 3M | rs112983626 | 2 | 74470023 | 3.34719559082237e-07 | 17.7848079 | intergenic | MOGS;MRPL53 | dist=4641;dist=1959 | MOGS | |
| 3M | rs113006316 | 2 | 74575233 | 3.34707225210704e-06 | 22.8886792 | intronic | M1AP | . | M1AP | |
| 3M | rs17746486 | 2 | 95056864 | 1.47154878644706e-06 | 6.90851358 | intergenic | MAL;MRPS5 | dist=2872;dist=28507 | MAL;MRPS5 | |
| 3M | rs76554191 | 2 | 95301880 | 3.87215481208204e-06 | 5.72148039 | intronic | KCNIP3 | . | KCNIP3 | |
| 3M | rs140352232 | 2 | 107421656 | 3.8788022972212e-06 | 21.0576511 | intergenic | MIR548AU;LINC01886 | dist=72131;dist=107763 | | ST6GAL2;RGPD4 |
| 12M | rs146919974 | 2 | 114495286 | 7.91246832646777e-07 | 19.7280466 | intronic | DPP10 | . | DPP10 | |
| 3M | rs116189766 | 2 | 125636287 | 1.17473937061964e-06 | 21.190764 | intergenic | CNTNAP5;LINC01941 | dist=715069;dist=473813 | | TSN;GYPC |
| 3M | rs139877408 | 2 | 128848699 | 4.57552408670287e-06 | 20.3067129 | intergenic | HS6ST1;LOC101927881 | dist=529831;dist=15901 | HS6ST1 | |
| 3M | rs142894171 | 2 | 150581757 | 1.40524666465064e-06 | 18.0710394 | intergenic | LINC01920;LINC02612 | dist=9536;dist=47140 | | RND3; |
| 3M | rs541508507 | 2 | 169166755 | 2.32254126928461e-06 | 15.4630033 | intronic | LRP2 | . | LRP2 | |
| 3M | rs142549310 | 2 | 169173996 | 2.69758962759472e-06 | 15.2501713 | exonic | LRP2 | . | | |
| 3M | rs556293455 | 2 | 176085692 | 1.27371002978549e-06 | 14.3860463 | intergenic | EVX2;HOXD13 | dist=1730;dist=7029 | EVX2;HOXD13 | |
| 3M | rs184098071 | 2 | 176251692 | 1.15315934031841e-06 | 13.947724 | intergenic | HOXD1;MTX2 | dist=60785;dist=17750 | MTX2 | |
| 3M | rs79935606 | 2 | 176473907 | 2.55448017015286e-06 | 16.3396833 | intergenic | MTX2;MIR1246 | dist=135882;dist=127073 | | |
| 12M | rs532416695 | 2 | 176622062 | 1.32803586650991e-07 | 19.8726656 | intergenic | MIR1246;LINC01116 | dist=21010;dist=7519 | | |
| 3M | rs112557251 | 2 | 187510673 | 3.44653364518888e-06 | 20.6510064 | intronic | TFPI | . | TFPI | |
| 3M | rs185158855 | 2 | 222785307 | 3.71819301386344e-06 | 16.6678864 | intergenic | MOGAT1;ACSL3 | dist=75377;dist=75728 | MOGAT1;ACSL3 | |
| 3M | rs185510569 | 2 | 222950143 | 5.51379458113093e-08 | 19.5886906 | intergenic | ACSL3;KCNE4 | dist=5505;dist=102047 | ACSL3;KCNE4 | |
| 3M | rs147559909 | 2 | 236142879 | 1.75642429015382e-08 | 17.0840975 | intergenic | AGAP1;GBX2 | dist=11086;dist=22356 | AGAP1 | |
| 3M | rs181217257 | 2 | 239067423 | 5.29269845950619e-10 | 21.5677368 | intronic | HDAC4 | . | HDAC4 | |
| 3M | rs188076929 | 2 | 239072023 | 1.0296847211524e-09 | 19.9775614 | intronic | HDAC4 | . | | |
| 3M | rs112475378 | 3 | 1584977 | 1.34664824685029e-06 | 9.55085929 | intergenic | CNTN6;CNTN4 | dist=180760;dist=513826 | CNTN6;CNTN4 | |
| 3M | rs145676540 | 3 | 2002951 | 1.25427052978558e-06 | 13.980658 | intergenic | CNTN6;CNTN4 | dist=598734;dist=95852 | | |
| 12M | rs144788248 | 3 | 19545253 | 3.87625815195203e-06 | 11.9086237 | intergenic | KCNH8;EFHB | dist=9611;dist=334221 | KCNH8;EFHB | |
| 3M | rs146007933 | 3 | 28185932 | 1.34950762676418e-06 | 7.51149453 | intergenic | LINC01980;CMC1 | dist=325607;dist=55687 | CMC1 | |
| 3M | rs73057656 | 3 | 33899079 | 3.43591498795888e-06 | 5.9238329 | intergenic | PDCD6IP;LOC101928135 | dist=29372;dist=976718 | PDCD6IP | |
| 3M | rs73085348 | 3 | 42669729 | 6.28155549798072e-07 | 10.139317 | intergenic | ZBTB47;KLHL40 | dist=2149;dist=15808 | ZBTB47 | |
| 3M | rs142684595 | 3 | 55285273 | 7.16787983460343e-08 | 16.7729431 | intergenic | LINC02017;WNT5A | dist=96059;dist=180442 | WNT5A | |
| 12M | rs149706477 | 3 | 66162553 | 2.87948941747911e-06 | 20.9828687 | intronic | SLC25A26 | . | SLC25A26 | |
| 3M | rs80203220 | 3 | 123003484 | 2.31429533714517e-06 | 13.426042 | intronic | SEMA5B | . | SEMA5B | |
| 12M | rs114280794 | 3 | 133123359 | 3.81823609520308e-06 | 8.76296877 | intronic | TMEM108 | . | TMEM108 | |
| 3M | rs192443987 | 3 | 135813503 | 1.3640761999881e-06 | 17.7540285 | intergenic | EPHB1;PPP2R3A | dist=553038;dist=152225 | EPHB1;PPP2R3A | |
| 3M | rs148248743 | 3 | 136415753 | 7.68859185089755e-07 | 22.839379 | intronic | STAG1 | . | STAG1 | |
| 12M | rs148248743 | 3 | 136415753 | 6.68816287673531E-07 | 27.1548587 | intronic | STAG1 | . | | |
| 3M | rs576124203 | 3 | 142122354 | 4.46547319819514e-07 | 19.4039925 | intronic | TFDP2 | . | TFDP2 | |
| 3M | rs545552231 | 3 | 142145508 | 4.79811814113124e-07 | 19.8641332 | intronic | TFDP2 | . | | |
| 12M | rs138138661 | 3 | 147940027 | 5.4928185119564e-08 | 23.1347457 | intergenic | LOC440982;LINC02032 | dist=430117;dist=138132 | | |
| 12M | rs189234695 | 3 | 147940371 | 3.47705303907978e-06 | 23.9697134 | intergenic | LOC440982;LINC02032 | dist=430461;dist=137788 | | |
| 12M | rs148997617 | 3 | 148037789 | 1.05449927842458e-07 | 20.8774277 | intergenic | LOC440982;LINC02032 | dist=527879;dist=40370 | | |
| 3M | rs193153124 | 3 | 148612923 | 2.77507554510464e-06 | 14.3657173 | intergenic | LINC02046;AGTR1 | dist=212967;dist=84948 | AGTR1 | |
| 3M | rs188720948 | 3 | 150352093 | 8.59439366670513e-07 | 17.7032653 | intergenic | LINC01214;TSC22D2 | dist=28346;dist=56205 | TSC22D2 | |
| 3M | rs16823323 | 3 | 153939413 | 3.35704338797875e-08 | 9.48464065 | intergenic | LINC02006;ARHGEF26 | dist=176887;dist=84988 | ARHGEF26 | |
| 3M | rs139943877 | 3 | 155733500 | 2.25796218142194e-06 | 13.0535902 | intronic | PLCH1 | . | PLCH1 | |
| 12M | rs148483098 | 3 | 157664262 | 3.27691859100501e-06 | 15.7357844 | intergenic | SLC66A1L;SHOX2 | dist=63168;dist=431643 | SHOX2 | |
| 12M | rs79252854 | 3 | 157805368 | 2.4780397246083e-06 | 17.5464336 | intergenic | SLC66A1L;SHOX2 | dist=204274;dist=290537 | | |
| 12M | rs139055031 | 3 | 158869060 | 3.17888515002842e-06 | 16.2853761 | intergenic | MFSD1;IQCJ | dist=39341;dist=200192 | MFSD1 | |
| 12M | rs143669489 | 3 | 159675096 | 4.93900179409246e-06 | 24.2253199 | intronic | IQCJ-SCHIP1;SCHIP1 | . | IQCJ-SCHIP1;SCHIP1 | |
| 3M | rs187047882 | 3 | 164567833 | 3.41360195160902e-06 | 18.5641361 | intergenic | MIR1263;LINC01324 | dist=396277;dist=146262 | | |
| 3M | rs141169929 | 3 | 165090674 | 2.90582805983588e-06 | 14.8454178 | intergenic | SI;SLITRK3 | dist=12178;dist=96046 | SLITRK3 | |
| 3M | rs186649043 | 3 | 175214503 | 2.29531002487815e-06 | 19.2134209 | intronic | NAALADL2 | . | NAALADL2 | |
| 3M | rs189709453 | 3 | 177856520 | 1.55548473782376e-06 | 23.2009936 | ncRNA_intronic | LINC02015 | . | | TBL1XR1;ZMAT3 |
| 12M | rs189709453 | 3 | 177856520 | 1.55548473782376E-06 | 23.2009936 | ncRNA_intronic | LINC02015 | . | | |
| 3M | rs186767531 | 3 | 177921989 | 1.62563417403378E-07 | 21.2013243 | intergenic | LINC02015;LINC01014 | dist=22765;dist=497212 | | |
| 12M | rs186767531 | 3 | 177921989 | 1.66219440747609E-06 | 23.0749977 | intergenic | LINC02015;LINC01014 | dist=22765;dist=497212 | | |
| 3M | rs182868205 | 3 | 177994660 | 1.36806623320862e-06 | 22.8544303 | intergenic | LINC02015;LINC01014 | dist=95436;dist=424541 | | |
| 12M | rs182868205 | 3 | 177994660 | 1.36806623320862E-06 | 22.8544303 | intergenic | LINC02015;LINC01014 | dist=95436;dist=424541 | | |
| 12M | rs76356799 | 3 | 179875980 | 2.9143183363234e-07 | 19.2867007 | intronic | PEX5L | . | PEX5L | |
| 12M | rs117591241 | 3 | 190866890 | 4.37648501942309e-06 | 19.9014028 | intergenic | GMNC;SNAR-I | dist=4196;dist=11040 | GMNC | |
| 3M | rs191792521 | 3 | 195919734 | 2.52243498533689e-06 | 13.9515484 | intergenic | TNK2;SDHAP1 | dist=6470;dist=40187 | TNK2 | |
| 3M | rs113651406 | 4 | 997179 | 2.52148284014823e-07 | 19.4054026 | intronic | IDUA | . | IDUA | |
| 3M | rs12502861 | 4 | 2424578 | 2.48774370313747e-06 | 11.1588472 | intronic | CFAP99 | . | CFAP99 | |
| 3M | rs189765693 | 4 | 4323066 | 2.55310143124462e-06 | 15.5191341 | intergenic | ZBTB49;NSG1 | dist=1283;dist=63466 | ZBTB49;NSG1 | |
| 12M | rs189094663 | 4 | 11621826 | 2.89927274380224e-06 | 23.895416 | intergenic | HS3ST1;LINC02360 | dist=192932;dist=119125 | HS3ST1 | |
| 12M | rs557276277 | 4 | 19012075 | 3.35996743882432e-06 | 19.0146344 | intergenic | LCORL;SLIT2 | dist=990200;dist=1239830 | LCORL;SLIT2 | |

| | | | | | | | | | | |
|---|---|---|---|---|---|---|---|---|---|---|
| 3M | rs183962155 | 4 | 21258020 | 3.64461882511902e-06 | 13.7646008 | intronic | KCNIP4 | . | KCNIP4 | |
| 3M | rs113751774 | 4 | 23427231 | 5.00531306271799e-07 | 14.9103274 | intergenic | GBA3;PPARGC1A | dist=607659;dist=364790 | | |
| 3M | rs113063005 | 4 | 23507444 | 1.79978248668523e-11 | 21.2073894 | intergenic | GBA3;PPARGC1A | dist=687872;dist=284577 | PPARGC1A | |
| 12M | rs113063005 | 4 | 23507444 | 9.56810303773154E-07 | 16.61763 | intergenic | GBA3;PPARGC1A | | | |
| 3M | rs145875128 | 4 | 32071514 | 1.07037949708586e-07 | 24.479986 | ncRNA_intronic | LINC02506 | . | ARAP2 | |
| 3M | rs143287889 | 4 | 35570658 | 4.29676290263636e-06 | 19.96741 | intergenic | LINC02484;ARAP2 | dist=1300911;dist=495346 | | |
| 3M | rs77141817 | 4 | 37052137 | 1.52230770368993e-08 | 20.6110292 | intergenic | LINC02616;MIR4801 | dist=31431;dist=189773 | | ;C4orf19,RELL1 |
| 3M | rs190822761 | 4 | 37097734 | 1.6069016808543e-08 | 20.5846354 | intergenic | LINC02616;MIR4801 | dist=77028;dist=144176 | | |
| 12M | rs147267707 | 4 | 43024541 | 2.25472034099615e-06 | 18.0094385 | intronic | GRXCR1 | . | | ATP8A1;KCTD8 |
| 12M | rs77618729 | 4 | 43048951 | 2.25749765325168e-06 | 18.0130153 | intergenic | GRXCR1;LINC02383 | dist=18293;dist=408583 | | |
| 12M | rs556435089 | 4 | 43549016 | 2.61570608345655e-06 | 17.939079 | intergenic | LINC02383;LINC02475 | dist=56473;dist=467845 | | |
| 12M | rs532464521 | 4 | 43569022 | 2.45873915171385e-06 | 17.9895528 | intergenic | LINC02383;LINC02475 | dist=76479;dist=447839 | | |
| 3M | rs999769259 | 4 | 61646247 | 3.61294355616724e-06 | 22.1246725 | intronic | ADGRL3 | . | ADGRL3 | |
| 3M | rs147630370 | 4 | 86529522 | 7.46814493303135e-07 | 16.0013633 | intergenic | MAPK10;MIR4452 | dist=76327;dist=12960 | MAPK10 | |
| 3M | rs147171192 | 4 | 88214436 | 4.39967162056646e-06 | 13.3044047 | intronic | ABCG2 | . | ABCG2 | |
| 3M | rs142993106 | 4 | 90036221 | 3.08700493831608e-06 | 8.08463611 | intergenic | MMRN1;CCSER1 | dist=81611;dist=91173 | MMRN1;CCSER1 | |
| 3M | rs146526206 | 4 | 90071867 | 1.81582969378392e-06 | 8.09176656 | intergenic | MMRN1;CCSER1 | dist=117257;dist=55527 | | |
| 3M | rs17017794 | 4 | 90904734 | 3.46919898165651e-07 | 5.83240545 | intronic | CCSER1 | . | | |
| 3M | rs145116559 | 4 | 111756440 | 4.90673849934313e-06 | 18.2441224 | intergenic | MIR297;FAM241A | dist=895793;dist=389014 | | PITX2;AP1AR |
| 3M | rs181415102 | 4 | 111768110 | 4.81312717634891e-06 | 18.2631019 | intergenic | MIR297;FAM241A | dist=907463;dist=377344 | | |
| 3M | rs191423619 | 4 | 125595472 | 2.50684302750887e-07 | 21.694611 | intergenic | MIR2054;INTU | dist=88165;dist=2037485 | INTU | |
| 3M | rs149298750 | 4 | 126237546 | 1.98725449883812e-07 | 22.184052 | intergenic | MIR2054;INTU | dist=730239;dist=1395411 | | |
| 12M | rs181132315 | 4 | 128726214 | 1.68603480568751e-06 | 22.1324679 | intergenic | LINC02615;JADE1 | dist=206818;dist=83486 | JADE1 | |
| 3M | rs112679237 | 4 | 138219706 | 2.04053672228041e-07 | 8.71202752 | intronic | SLC7A11 | . | SLC7A11 | |
| 12M | rs147455971 | 4 | 140010558 | 3.87240564796723e-06 | 17.2160142 | intronic | MAML3 | . | MAML3 | |
| 3M | rs531769270 | 4 | 153489891 | 2.4145887031028e-06 | 16.5945338 | intronic | TMEM131L | . | | MND1;TLR2 |
| 3M | rs116651654 | 4 | 162317591 | 3.48829177722726e-07 | 15.6223694 | intergenic | FSTL5;MIR4454 | dist=153557;dist=775983 | FSTL5 | |
| 12M | rs17615362 | 4 | 169012936 | 4.30304118178725e-06 | 5.41974472 | intergenic | CBR4;SH3RF1 | dist=2681;dist=81323 | CBR4;SH3RF1 | |
| 12M | rs17543620 | 4 | 169013574 | 4.21133121953689e-06 | 5.42796755 | intergenic | CBR4;SH3RF1 | dist=3319;dist=80685 | | |
| 3M | rs567982164 | 5 | 25631188 | 5.7310337989489e-07 | 21.8356286 | intergenic | LINC02211;CDH9 | dist=328908;dist=1249409 | CDH9 | |
| 3M | rs191986449 | 5 | 25745578 | 4.59857674938651e-06 | 18.4978793 | intergenic | LINC02211;CDH9 | dist=443298;dist=1135019 | | |
| 12M | rs142934021 | 5 | 53366842 | 1.40182538744286e-08 | 18.824715 | intergenic | LOC257396;FST | dist=251716;dist=113787 | FST | |
| 3M | rs185771987 | 5 | 73989664 | 4.53395085864185e-06 | 18.2344824 | intergenic | ARHGEF28;LINC01335 | dist=47671;dist=316746 | ARHGEF28 | |
| 3M | rs139360368 | 5 | 74076284 | 1.24776973080952e-06 | 18.3590487 | intergenic | ARHGEF28;LINC01335 | dist=134291;dist=230126 | | |
| 3M | rs181933850 | 5 | 92169830 | 3.33393070013435e-06 | 11.0436586 | intergenic | ARRDC3;NR2F1 | dist=749115;dist=1279415 | ARRDC3;NR2F1 | |
| 3M | rs190190051 | 5 | 92179668 | 3.47677941958718e-06 | 11.0589887 | intergenic | ARRDC3;NR2F1 | dist=758953;dist=1269577 | | |
| 3M | rs182531466 | 5 | 92234256 | 4.23245053141191e-06 | 12.6804978 | intergenic | ARRDC3;NR2F1 | dist=813541;dist=1214989 | | |
| 3M | rs187236873 | 5 | 92234630 | 1.97926852689169e-06 | 12.44907 | intergenic | ARRDC3;NR2F1 | dist=813915;dist=1214615 | | |
| 3M | rs183816745 | 5 | 92336071 | 6.02067560021534e-08 | 17.3487909 | intergenic | ARRDC3;NR2F1 | dist=915356;dist=1113174 | | |
| 3M | rs111407636 | 5 | 95744325 | 6.27809322802765e-07 | 18.642774 | intronic | RHOBTB3 | . | RHOBTB3 | |
| 3M | rs111676272 | 5 | 95758594 | 6.71649637293761e-07 | 18.6068625 | intronic | RHOBTB3 | . | | |
| 3M | rs75848314 | 5 | 95762636 | 1.11087241977704e-07 | 20.0998481 | intronic | RHOBTB3 | . | | |
| 3M | rs111846247 | 5 | 95775939 | 1.06810112370498e-07 | 20.4865772 | intronic | RHOBTB3 | . | | |
| 3M | rs137873790 | 5 | 97751337 | 4.51630701305597e-06 | 9.22240881 | intergenic | LINC01340;LINC02234 | dist=80286;dist=89421 | | RIOK2;RGMB |
| 3M | rs189912648 | 5 | 135429749 | 2.55441427995549e-06 | 16.9732599 | intergenic | MACROH2A1;DCANP1 | dist=29862;dist=14465 | MACROH2A1;DCANP1 | |
| 12M | rs189912648 | 5 | 135429749 | 3.32990944802665E-06 | 19.8604449 | intergenic | MACROH2A1;DCANP1 | dist=29862;dist=14465 | | |
| 12M | rs111512950 | 5 | 154300867 | 4.11671051184833e-06 | 13.567443 | intronic | GALNT10 | . | GALNT10 | |
| 3M | rs191006910 | 5 | 154836449 | 3.35533057793161e-06 | 17.6173803 | intronic | FAXDC2 | . | FAXDC2 | |
| 3M | rs74343174 | 5 | 162066176 | 2.54836335504837e-06 | 14.7671824 | intergenic | LINC01202;GABRG2 | dist=64980;dist=1289 | GABRG2 | |
| 3M | rs775626702 | 5 | 163203061 | 3.23411085027866e-06 | 18.9484137 | intergenic | GABRG2;CCNG1 | dist=1047522;dist=234510 | GABRG2;CCNG1 | |
| 3M | rs371245624 | 5 | 163453950 | 1.65804063735814e-06 | 19.4795931 | UTR3 | NUDCD2 | NM_001329991:c.*170>0;NM_ | NUDCD2 | |
| 3M | rs290120 | 5 | 163841238 | 2.40444258455957e-06 | 13.3587764 | intergenic | MAT2B;LINC02143 | dist=321884;dist=607184 | MAT2B | |
| 3M | rs545428520 | 5 | 168394261 | 3.96137253437646e-09 | 21.566035 | intronic | WWC1 | . | WWC1 | |
| 3M | rs528404963 | 5 | 168425020 | 7.1738299793335e-09 | 21.1393977 | intronic | WWC1 | . | | |
| 3M | rs72832764 | 5 | 170577669 | 2.69939738977966e-06 | 18.2159757 | intronic | KCNIP1 | . | KCNIP1 | |
| 3M | rs72837643 | 5 | 170761951 | 4.147895992623e-06 | 16.9302874 | intergenic | KCNIP1;GABRP | dist=25319;dist=21768 | GABRP | |
| 3M | rs142311947 | 5 | 177957468 | 2.99420386474277e-06 | 12.0339013 | ncRNA_intronic | LOC100128340 | . | | FAM153A;FAM153C |
| 3M | rs151015676 | 5 | 177963936 | 4.05781112465234e-06 | 20.1421367 | intergenic | LOC100128340;PROP1 | dist=4136;dist=28299 | | |
| 3M | rs1877768 | 6 | 16534692 | 2.77630022916084e-06 | 8.28754685 | intronic | ATXN1 | . | ATXN1 | |
| 12M | rs73384619 | 6 | 20457948 | 3.65476878017977e-06 | 8.3916702 | intronic | E2F3 | . | E2F3 | |
| 3M | rs150586237 | 6 | 24491120 | 5.5145390836127e-09 | 22.0461443 | intergenic | GPLD1;ALDH5A1 | dist=1542;dist=3849 | GPLD1;ALDH5A1 | |
| 3M | rs571986619 | 6 | 84969114 | 1.70115854486702e-06 | 20.7858067 | intergenic | TBX18;LINC02535 | dist=204516;dist=418105 | TBX18 | |
| 3M | rs56224400 | 6 | 97644799 | 9.49951178557744e-07 | 8.4433064 | ncRNA_intronic | LOC101927314 | . | | MMS22L;POU3F2 |
| 3M | rs147627638 | 6 | 98725240 | 2.84737109324869e-06 | 13.0785306 | intergenic | MIR2113;PNKY | dist=700621;dist=104901 | | POU3F2 |
| 3M | rs141326851 | 6 | 134511989 | 2.157242978017e-06 | 8.57849688 | intergenic | LINC01010;LOC101928304 | dist=7969;dist=13329 | | SGK1;ALDH8A1 |
| 3M | rs146048121 | 6 | 141877818 | 3.23639603147815e-06 | 20.0214379 | intergenic | MIR4465;NMBR | dist=1193935;dist=196666 | | |
| 3M | rs142106992 | 6 | 141948748 | 5.23067431501407e-10 | 22.8345886 | intergenic | MIR4465;NMBR | dist=1264865;dist=125736 | | |
| 3M | rs72983831 | 6 | 141985963 | 7.82545989102806e-07 | 15.6733239 | intergenic | MIR4465;NMBR | dist=1302080;dist=88521 | | |
| 3M | rs72986533 | 6 | 142290121 | 1.53596722070896e-07 | 13.2812726 | intergenic | VTA1;ADGRG6 | dist=65437;dist=11798 | VTA1;AGRG6 | |
| 3M | rs73586304 | 6 | 142518288 | 4.95566451287885e-06 | 18.2405116 | intergenic | ADGRG6;LOC153910 | dist=72022;dist=8167 | | |
| 12M | rs4896997 | 6 | 148362788 | 3.04539162547005e-06 | 14.4320923 | intronic | SASH1 | . | | |
| 12M | rs4131286 | 6 | 148367827 | 3.05925201460773e-06 | 14.4284672 | intronic | SASH1 | . | SASH1 | |
| 12M | rs17078283 | 6 | 148375784 | 3.05230865558584e-06 | 14.4302821 | intronic | SASH1 | . | | |
| 12M | rs56821264 | 6 | 148382765 | 2.17257084724272e-06 | 14.7509213 | intronic | SASH1 | . | | |
| 3M | rs148532212 | 6 | 165086368 | 1.8149830669263e-07 | 19.3535457 | intergenic | MEAT6;C6orf118 | dist=264305;dist=193296 | C6orf118 | |
| 3M | rs117498042 | 6 | 165100792 | 1.85878685547635e-07 | 19.335471 | intergenic | MEAT6;C6orf118 | dist=278729;dist=178872 | | |
| 3M | rs148153037 | 6 | 167087898 | 3.20376100252789e-09 | 14.3359246 | intergenic | CEP43;CCR6 | dist=35180;dist=23909 | CCR6 | |
| 3M | rs184487573 | 6 | 167099983 | 1.91568282565546e-07 | 13.5009635 | intergenic | CEP43;CCR6 | dist=47265;dist=11824 | | |
| 3M | rs17776100 | 7 | 6386848 | 5.13308111425872e-07 | 6.63697397 | intronic | RAC1 | . | RAC1 | |
| 3M | rs187978759 | 7 | 11672218 | 2.76447739355703e-06 | 20.4555522 | intronic | THSD7A | . | THSD7A | |
| 12M | rs143686474 | 7 | 16252374 | 4.88904023817646e-06 | 10.8531498 | ncRNA_intronic | CRPPA | . | | MEOX2;SOSTDC1 |
| 3M | rs117166500 | 7 | 17013154 | 1.89815826168549e-06 | 12.5146865 | intergenic | AGR3;AHR | dist=131171;dist=285498 | AGR3;AHR | |

| | | | | | | | | | | |
|---|---|---|---|---|---|---|---|---|---|---|
| 3M | rs75689761 | 7 | 18366950 | 9.11790229535791e-07 | 11.8482917 | intronic | HDAC9 | . | | |
| 3M | rs77346868 | 7 | 18366976 | 1.13767941006556e-06 | 11.0210129 | intronic | HDAC9 | . | | |
| 3M | rs78225611 | 7 | 18367841 | 1.13375019479557e-06 | 11.0294387 | intronic | HDAC9 | . | | |
| 3M | rs77300464 | 7 | 18369138 | 2.09191655072049e-08 | 14.6659794 | intronic | HDAC9 | . | HDAC9 | |
| 3M | rs79602997 | 7 | 18370627 | 9.43076371109133e-07 | 11.8608211 | intronic | HDAC9 | . | | |
| 3M | rs75773869 | 7 | 18371222 | 9.36316662267224e-07 | 11.8642716 | intronic | HDAC9 | . | | |
| 3M | rs75606013 | 7 | 18374990 | 2.11098533218711e-08 | 14.6617119 | intronic | HDAC9 | . | | |
| 3M | rs61434999 | 7 | 18378728 | 9.22178787748122e-07 | 11.8411511 | intronic | HDAC9 | . | | |
| 3M | rs78907958 | 7 | 18385394 | 4.98811901973441e-07 | 12.0901322 | intronic | HDAC9 | . | | |
| 3M | rs76526501 | 7 | 18391487 | 8.57654056311165e-09 | 14.8397312 | intronic | HDAC9 | . | | |
| 3M | rs74455595 | 7 | 18392161 | 8.57654056311165e-09 | 14.8397312 | intronic | HDAC9 | . | | |
| 3M | rs79182806 | 7 | 18394204 | 4.13732512224196e-07 | 12.0972603 | intronic | HDAC9 | . | | |
| 3M | rs10279777 | 7 | 18401966 | 1.65309369490778e-08 | 12.8681335 | intronic | HDAC9 | . | | |
| 3M | rs77867199 | 7 | 18402652 | 3.93328414300688e-07 | 10.8221282 | intronic | HDAC9 | . | | |
| 3M | rs80156375 | 7 | 18403592 | 3.27106315853648e-07 | 11.5406255 | intronic | HDAC9 | . | | |
| 3M | rs17169602 | 7 | 18407118 | 1.78410513894506e-08 | 12.7273717 | intronic | HDAC9 | . | | |
| 3M | rs10486295 | 7 | 18407184 | 1.75006169477667e-08 | 12.76229 | intronic | HDAC9 | . | | |
| 3M | rs75090694 | 7 | 18407813 | 1.01013780102531e-08 | 13.8314005 | intronic | HDAC9 | . | | |
| 3M | rs55844051 | 7 | 23320744 | 1.93295366566624e-06 | 18.3875896 | intronic | IGF2BP3 | . | IGF2BP3 | |
| 3M | rs62447184 | 7 | 36534898 | 2.45196683172423e-06 | 5.12842209 | intronic | AOAH | . | | |
| 3M | rs574076561 | 7 | 49505151 | 3.8458987487847e-06 | 19.6434766 | intergenic | CDC14C;VWC2 | dist=577697;dist=268487 | VWC2 | |
| 3M | rs111391231 | 7 | 89867731 | 4.17997193738163e-06 | 8.31717755 | intergenic | ZNF804B;STEAP2 | dist=529203;dist=14622 | ZNF804B;STEAP2 | |
| 3M | rs111900874 | 7 | 89887355 | 3.79796761664437e-06 | 8.42424013 | ncRNA_intronic | STEAP2 | . | | |
| 3M | rs181259864 | 7 | 97859511 | 3.26388920997597e-06 | 17.1360821 | ncRNA_intronic | CZ1P-ASNS | . | CZ1P-ASNS | ASNS;TAC1 |
| 3M | rs192750513 | 7 | 97948518 | 3.9425756746568e-06 | 17.1892469 | ncRNA_intronic | CZ1P-ASNS | . | | |
| 3M | rs539713344 | 7 | 100877165 | 2.02994489815685e-06 | 20.044756 | intronic | SRRT | . | SRRT | |
| 3M | rs188028357 | 7 | 101025144 | 5.38240258932335e-07 | 21.8425578 | intronic | MUC17 | . | | MUC12;TRIM56 |
| 12M | rs73202425 | 7 | 109515659 | 3.05259237348811e-06 | 10.761844 | intergenic | C7orf66;EIF3IP1 | dist=631072;dist=443568 | EIF3IP1 | |
| 12M | rs181182636 | 7 | 116014504 | 2.90894506747647e-06 | 21.9472639 | intronic | TFEC | . | TFEC | |
| 12M | rs182211730 | 7 | 116363561 | 1.45395858607717e-06 | 18.7746033 | intergenic | LOC102724434;CAV2 | dist=76834;dist=136177 | CAV2 | |
| 12M | rs143281973 | 7 | 116367942 | 1.62372890091138e-06 | 18.2750778 | intergenic | LOC102724434;CAV2 | dist=81215;dist=131796 | | |
| 12M | rs138873576 | 7 | 116385305 | 3.22380485845586e-06 | 17.6151711 | intergenic | LOC102724434;CAV2 | dist=98578;dist=114433 | | |
| 12M | rs190354334 | 7 | 116435792 | 4.2862377340545e-06 | 17.3392581 | intergenic | LOC102724434;CAV2 | dist=149065;dist=63946 | | |
| 12M | rs186893139 | 7 | 116456916 | 3.55104961576984e-06 | 18.602598 | intergenic | LOC102724434;CAV2 | dist=170189;dist=42822 | | |
| 12M | rs558784715 | 7 | 117403240 | 1.5182227326648e-06 | 20.7591787 | intronic | ASZ1 | . | | |
| 12M | rs576047962 | 7 | 117468967 | 1.61732320840498e-06 | 20.7261549 | intergenic | ASZ1;CFTR | dist=41474;dist=11058 | CFTR | |
| 12M | rs529110230 | 7 | 117535853 | 1.24629705452557e-06 | 20.8583633 | intronic | CFTR | . | | |
| 12M | rs142215699 | 7 | 117559820 | 1.6210064953756e-06 | 20.7292287 | ncRNA_intronic | CFTR | . | | |
| 12M | rs142721557 | 7 | 117578274 | 1.61401730397805e-06 | 20.7329975 | intronic | CFTR | . | | |
| 12M | rs201355675 | 7 | 117585727 | 1.64203568676087e-06 | 20.718124 | intronic | CFTR | . | | |
| 12M | rs188993522 | 7 | 117634677 | 1.9792198058251e-06 | 21.2984827 | intronic | CFTR | . | | |
| 3M | rs536023430 | 7 | 147167980 | 2.63612685649085e-06 | 19.4038683 | intronic | CNTNAP2 | . | CNTNAP2 | |
| 3M | rs187384541 | 8 | 1684075 | 3.15110880283135e-06 | 6.88745117 | intronic | DLGAP2 | . | DLGAP2 | |
| 3M | rs575473987 | 8 | 5719972 | 1.07256928472273e-08 | 22.2248653 | intergenic | CSMD1;LOC100287015 | dist=725058;dist=683583 | CSMD1 | |
| 3M | rs139062456 | 8 | 13394482 | 3.88565669511608e-06 | 7.83515809 | intronic | DLC1 | . | DLC1 | |
| 3M | rs185874707 | 8 | 18173165 | 2.74810255839378e-06 | 9.13934765 | intronic | NAT1 | . | NAT1 | |
| 3M | rs140797780 | 8 | 22230279 | 1.73425396214273e-07 | 19.2908396 | intronic | PHYHIP | . | PHYHIP | |
| 3M | rs147601511 | 8 | 22464806 | 7.14867637986866e-07 | 16.4471181 | intronic | PPP3CC | . | PPP3CC | |
| 3M | rs188415494 | 8 | 25755782 | 3.49911568838158e-06 | 18.5169478 | intergenic | CDCA2;EBF2 | dist=247865;dist=85943 | CDCA2;EBF2 | |
| 3M | rs9643828 | 8 | 54616513 | 2.29737457097614e-07 | -2.4236294 | intronic | RP1 | . | | SOX17;XKR4 |
| 3M | rs423841 | 8 | 54643509 | 2.7508818251862e-06 | -2.1495726 | intronic | RP1 | . | | |
| 3M | rs433324 | 8 | 54652049 | 9.22325760191311e-07 | -2.2483462 | intronic | RP1 | . | | |
| 3M | rs369623 | 8 | 54659380 | 8.50610562336969e-07 | -2.2359815 | intronic | RP1 | . | | |
| 3M | rs446222 | 8 | 54662400 | 7.36959451875675e-07 | -2.247819 | intronic | RP1 | . | | |
| 3M | rs432393 | 8 | 54667738 | 1.22958170848882e-06 | -2.1875382 | intronic | RP1 | . | | |
| 3M | rs3098298 | 8 | 54670278 | 1.22819650607779e-06 | -2.1875635 | intronic | RP1 | . | | |
| 3M | rs367179 | 8 | 54675056 | 1.22819650607779e-06 | -2.1875635 | intronic | RP1 | . | | |
| 3M | rs382476 | 8 | 54678415 | 7.36949131111935e-07 | -2.2481938 | intronic | RP1 | . | | |
| 3M | rs384543 | 8 | 54679049 | 7.36949131111935e-07 | -2.2481938 | intronic | RP1 | . | | |
| 3M | rs405226 | 8 | 54679776 | 1.2251324121268e-06 | -2.1881305 | intronic | RP1 | . | | |
| 3M | rs384127 | 8 | 54684929 | 7.36964497256014e-07 | -2.2481559 | intronic | RP1 | . | | |
| 3M | rs858397 | 8 | 54702130 | 7.161979275107e-07 | 2.2301348 | intronic | RP1 | . | | |
| 3M | rs2375537 | 8 | 54706948 | 6.2991993988762e-07 | 2.23876848 | intronic | RP1 | . | | |
| 3M | rs720372 | 8 | 54716077 | 2.93014534156507e-06 | 2.09872734 | intronic | RP1 | . | | |
| 3M | rs1437781 | 8 | 54717292 | 6.32222203401203e-07 | 2.23854673 | intronic | RP1 | . | | |
| 3M | rs1595406 | 8 | 54718055 | 2.80263765648492e-06 | 2.10018889 | intronic | RP1 | . | | |
| 3M | rs1437782 | 8 | 54720202 | 5.53724964407086e-07 | 2.2579585 | exonic | RP1 | . | | |
| 3M | rs10105693 | 8 | 54727912 | 3.41214459494352e-07 | 2.2928391 | intronic | RP1 | . | | |
| 3M | rs2375536 | 8 | 54728162 | 2.13408172799341e-06 | 2.12635806 | intronic | RP1 | . | | |
| 3M | rs4737674 | 8 | 54749094 | 2.86005059848221e-07 | 2.30559554 | intronic | RP1 | . | | |
| 3M | rs11987234 | 8 | 54757269 | 3.13190459972498e-07 | 2.29599259 | intronic | RP1 | . | | |
| 3M | rs13277510 | 8 | 54761589 | 2.8610568625199e-07 | 2.30494752 | intronic | RP1 | . | | |
| 3M | rs12548593 | 8 | 54762057 | 3.38344249342945e-07 | 2.28269961 | intronic | RP1 | . | | |
| 3M | rs1812506 | 8 | 54763541 | 2.00833101672028e-06 | 2.12258868 | intronic | RP1 | . | | |
| 3M | rs16920698 | 8 | 54765874 | 2.85868793626322e-07 | 2.30492695 | intronic | RP1 | . | | |
| 3M | rs1561297 | 8 | 54765978 | 3.34668102178151e-07 | 2.28315237 | intronic | RP1 | . | | |
| 3M | rs4737676 | 8 | 54766986 | 2.8589296796158e-07 | 2.30492057 | intronic | RP1 | . | | |
| 3M | rs2083123 | 8 | 54767758 | 3.4358513898515e-07 | 2.27910794 | intronic | RP1 | . | | |
| 3M | rs983248 | 8 | 54768232 | 2.87356270418993e-07 | 2.30260428 | intronic | RP1 | . | | |
| 3M | rs1391463 | 8 | 54769316 | 2.87382622216009e-07 | 2.30258865 | intronic | RP1 | . | | |
| 3M | rs10958428 | 8 | 54773081 | 5.0112447137886e-07 | 2.23996885 | intronic | RP1 | . | | |
| 3M | rs13278605 | 8 | 54775611 | 3.41581916245031e-07 | 2.27377348 | intronic | RP1 | . | | |
| 3M | rs13276543 | 8 | 54775614 | 3.13999361335922e-07 | 2.29263157 | intronic | RP1 | . | | |

| | | | | | | | | | | |
|---|---|---|---|---|---|---|---|---|---|---|
| 3M | rs7822082 | 8 | 54777660 | 1.5040633792954e-07 | 2.3570878 | intronic | RP1 | . | | |
| 3M | rs4737201 | 8 | 54778898 | 2.89096388651053e-07 | 2.30197734 | intronic | RP1 | . | | |
| 3M | rs7843693 | 8 | 54779552 | 4.78812399757755e-06 | 1.99856756 | intronic | RP1 | . | | |
| 3M | rs1396896 | 8 | 54782750 | 4.77810054425197e-06 | 1.99853327 | intronic | RP1 | . | | |
| 3M | rs2375219 | 8 | 54785735 | 3.05253330469401e-06 | 2.05700167 | intronic | RP1 | . | | |
| 3M | rs1391462 | 8 | 54787221 | 4.79440218462524e-06 | 1.9984766 | intronic | RP1 | . | | |
| 3M | rs12678939 | 8 | 54792461 | 4.93547251213243e-06 | 2.01975618 | intronic | RP1 | . | | |
| 3M | rs1498183 | 8 | 54804345 | 4.76724404445351e-06 | 2.0245746 | intronic | RP1 | . | | |
| 12M | rs62515405 | 8 | 56143419 | 4.69438076015372e-06 | 8.07691204 | intergenic | MOS;PLAG1 | dist=29437;dist=17490 | MOS;PLAG1 | |
| 12M | rs62515436 | 8 | 56228644 | 3.44618686083172e-06 | 9.27545902 | intergenic | CHCHD7;SDR16C5 | dist=9835;dist=71004 | CHCHD7;SDR16C5 | |
| 12M | rs12114488 | 8 | 61760164 | 1.33492404675942e-06 | 2.62307914 | intergenic | MIR4470;LINC02155 | dist=45305;dist=129675 | | ASPH;NKAIN3 |
| 3M | rs117816016 | 8 | 102739034 | 3.21886378098173e-06 | 19.8807023 | intergenic | LOC101927245;GASAL1 | dist=52311;dist=67788 | | KLF10;AZIN1 |
| 3M | rs567383525 | 8 | 114017265 | 1.83606414606387e-06 | 17.7455227 | intergenic | CSMD3;TRPS1 | dist=580326;dist=1391230 | CSMD3 | |
| 3M | rs545550279 | 8 | 114540200 | 4.75001774825929e-06 | 19.1812421 | intergenic | CSMD3;TRPS1 | dist=1103261;dist=868295 | TRPS1 | |
| 3M | rs536803366 | 8 | 121989172 | 1.65900394985684e-06 | 19.2098639 | intergenic | HAS2;SMILR | dist=343847;dist=425155 | | |
| 3M | rs532513136 | 8 | 134801505 | 7.47175269151994e-07 | 19.9490594 | upstream | MIR30B | dist=898 | | ZFAT |
| 3M | rs532730683 | 9 | 1784492 | 1.02051348042702e-06 | 18.726838 | intergenic | DMRT2;SMARCA2 | dist=726938;dist=230855 | DMRT2;SMARCA2 | |
| 3M | rs540065886 | 9 | 2770228 | 1.90434082268377e-06 | 19.9489151 | intergenic | KCNV2;PUM3 | dist=40191;dist=33927 | PUM3 | |
| 3M | rs543844012 | 9 | 30107025 | 4.37741040782571e-06 | 18.1160485 | intergenic | LINGO2;LINC01242 | dist=893424;dist=281910 | LINGO2 | |
| 3M | rs148556485 | 9 | 81408911 | 5.40048598404711e-07 | 21.3800152 | intergenic | LINC01507;TLE1 | dist=1374356;dist=174772 | TLE1 | |
| 3M | rs140782222 | 9 | 81413979 | 4.53117855536307e-07 | 21.791526 | intergenic | LINC01507;TLE1 | dist=1379424;dist=169704 | | |
| 3M | rs188034471 | 9 | 83322799 | 2.79197739516285e-06 | 14.7559999 | intronic | FRMD3 | . | | |
| 3M | rs190294315 | 9 | 83330550 | 4.29120280825722e-07 | 14.5664276 | intronic | FRMD3 | . | FRMD3 | |
| 3M | rs545690161 | 9 | 90268417 | 1.13978419923937e-08 | 17.1436874 | intergenic | MIR4290HG;LINC01508 | dist=226918;dist=32479 | | |
| 3M | rs565682685 | 9 | 90460046 | 3.57832955336558e-07 | 18.938397 | intergenic | LINC01508;LINC01501 | dist=26557;dist=2386 | | |
| 3M | rs183737367 | 9 | 90567765 | 3.8014567877404e-08 | 23.3452306 | ncRNA_intronic | LINC01501 | . | | |
| 3M | rs187213609 | 9 | 90653183 | 5.71707003010382e-08 | 23.1975304 | intergenic | DIRAS2;SYK | dist=10359;dist=148417 | DIRAS2;SYK | |
| 12M | rs144138711 | 9 | 97160137 | 1.39149931661618e-06 | 6.08547131 | ncRNA_intronic | ANKRD18CP | . | | CTSV;CCDC180 |
| 3M | rs150027952 | 9 | 100623134 | 1.2828864296083e-06 | 21.8616648 | intergenic | CAVIN4;PLPPR1 | dist=34747;dist=405593 | PLPPR1 | |
| 3M | rs146207930 | 9 | 126280057 | 2.39787202381566e-06 | 11.8624321 | intergenic | LOC101929116;MVB12B | dist=4142;dist=46772 | MVB12B | |
| 3M | rs78296164 | 9 | 132391328 | 6.93811773227571e-07 | 11.6095338 | intronic | TTF1 | . | TTF1 | |
| 3M | rs77871739 | 9 | 135660463 | 3.19831184275799e-06 | 15.0724603 | intergenic | GLT6D1;LCN9 | dist=20923;dist=2859 | | MRPS2;CAMSAP1 |
| 3M | rs118040657 | 10 | 3430654 | 4.61906712585886e-06 | 12.1147712 | ncRNA_intronic | LOC105376360 | . | | PITRM1;KLF6 |
| 3M | rs184425183 | 10 | 13415520 | 1.92440370103676e-09 | 22.6683174 | intergenic | SEPHS1;BEND7 | dist=67222;dist=22961 | SEPHS1;BEND7 | |
| 3M | rs184458518 | 10 | 13429195 | 1.48323670154931e-09 | 22.8015789 | intergenic | SEPHS1;BEND7 | dist=80897;dist=9286 | | |
| 3M | rs117998251 | 10 | 13455976 | 1.19486099712372e-09 | 22.9543311 | intronic | BEND7 | . | | |
| 3M | rs117025967 | 10 | 19918123 | 5.25165736443477e-07 | 9.66973563 | intronic | PLXDC2 | . | | MALRD1;NEBL |
| 3M | rs529011661 | 10 | 20083387 | 2.34383539239577e-06 | 15.6393469 | intronic | PLXDC2 | . | | |
| 12M | rs142956968 | 10 | 23201236 | 8.84691036334147e-07 | 10.4167992 | downstream | C10orf67 | dist=680 | | MSRB2;OTUD1 |
| 12M | rs145764464 | 10 | 23312918 | 1.10461979254428e-06 | 9.5967306 | intronic | C10orf67 | . | | |
| 3M | rs12266995 | 10 | 24563854 | 3.92980514429013e-06 | 5.96782535 | intergenic | KIAA1217;ARHGAP21 | dist=16006;dist=19760 | ARHGAP21 | |
| 3M | rs138249376 | 10 | 61756071 | 4.86147789321271e-06 | 16.4300711 | intronic | CABCOCO1 | . | | TMEM26;ARID5B |
| 3M | rs140277951 | 10 | 80599344 | 1.98902982335697e-07 | 13.7463158 | intronic | SH2D4B | . | | TSPAN14;NRG3 |
| 3M | rs566018180 | 10 | 84995296 | 4.3799943792317e-06 | 18.563552 | intergenic | CCSER2;LINC01519 | dist=476775;dist=198125 | CCSER2 | |
| 3M | rs140706881 | 10 | 94457778 | 1.40251021425162e-06 | 18.974805 | intronic | TBC1D12 | . | TBC1D12 | |
| 3M | rs117913371 | 10 | 101158729 | 2.30639701253664e-08 | 7.86609555 | intergenic | TLX1NB;LINC01514 | dist=17463;dist=17593 | | KAZALD1;BTRC |
| 3M | rs75334617 | 10 | 101196395 | 7.30335368747825e-07 | 5.84603405 | intergenic | LINC01514;LBX1 | dist=2248;dist=30581 | | |
| 3M | rs752259256 | 10 | 103165562 | 4.52164287436037e-06 | 20.4258639 | intronic | NT5C2 | . | NT5C2 | |
| 12M | rs147944608 | 10 | 108663951 | 9.83587566974791e-07 | 11.2205243 | intergenic | LINC01435;XPNPEP1 | dist=594658;dist=1200815 | rs144138711 | |
| 3M | rs180828621 | 10 | 122773893 | 6.39935885478647e-08 | 15.1380574 | ncRNA_intronic | DMBT1L1 | . | | CUZD1;FAM24B |
| 3M | rs147393020 | 10 | 123019758 | 1.64586496415273e-06 | 14.1853505 | intronic | ACADSB | . | ACADSB | |
| 3M | rs193093906 | 10 | 125016920 | 3.17719154902332e-06 | 10.5219537 | intronic | CTBP2 | . | CTBP2 | |
| 12M | rs556274646 | 11 | 439012 | 1.93909342324625e-06 | 12.8914592 | intronic | ANO9 | . | ANO9 | |
| 12M | rs148815783 | 11 | 1013992 | 2.06734210421034e-06 | 13.5980313 | exonic | MUC6 | . | | AP2A2; |
| 3M | rs151115079 | 11 | 18634194 | 4.89469717121508e-08 | 17.8273587 | intronic | SPTY2D1 | . | | |
| 3M | rs138414342 | 11 | 18657851 | 3.15830767684997e-08 | 18.1582505 | intergenic | SPTY2D1;TMEM86A | dist=23509;dist=40928 | SPTY2D1;TMEM86A | |
| 3M | rs541653703 | 11 | 18680239 | 2.1765671670172e-08 | 18.3169499 | intergenic | SPTY2D1;TMEM86A | dist=45897;dist=18540 | | |
| 3M | rs118093638 | 11 | 18696777 | 9.7151911784911e-07 | 14.2657259 | intergenic | SPTY2D1;TMEM86A | dist=62435;dist=2002 | | |
| 3M | rs181812512 | 11 | 66898258 | 4.24789238469822e-06 | 19.5093168 | intronic | PC | . | PC | |
| 3M | rs529345909 | 11 | 67343381 | 4.87261379674454e-06 | 18.70153 | ncRNA_intronic | LOC100130987 | . | LOC100130987 | SSH3;POLD4 |
| 3M | rs544042801 | 11 | 68692775 | 3.07835940417967e-06 | 15.8869866 | intergenic | GAL;TESMIN | dist=1600;dist=14665 | GAL | |
| 3M | rs149949098 | 11 | 95366702 | 2.52728186016237e-06 | 8.55927625 | intergenic | LOC100129203;FAM76B | dist=132298;dist=402251 | FAM76B | |
| 3M | rs74521112 | 11 | 99218416 | 3.48297260681022e-07 | 6.25632387 | intronic | CNTN5 | . | CNTN5 | |
| 3M | rs79213709 | 11 | 99222724 | 5.77809319512938e-07 | 6.17081884 | intronic | CNTN5 | . | | |
| 3M | rs112007361 | 11 | 99317649 | 2.27561716225072e-07 | 6.35658468 | intronic | CNTN5 | . | | |
| 3M | rs148781275 | 11 | 103769875 | 2.2488967712367e-06 | 15.7111853 | intergenic | DYNC2H1;MIR4693 | dist=290012;dist=80031 | DYNC2H1 | |
| 3M | rs141281289 | 11 | 123822316 | 3.07596924681855e-07 | 15.811518 | intergenic | OR6M1;TMEM225 | dist=15967;dist=60603 | | No gene nearby |
| 3M | rs79539453 | 11 | 125396457 | 1.65356394618456e-06 | 20.8220616 | intronic | PKNOX2 | . | PKNOX2 | |
| 3M | rs528140343 | 11 | 125849332 | 9.33313906476653e-07 | 18.2835521 | intergenic | PATE4;HYLS1 | dist=9260;dist=34282 | | |
| 3M | rs546409459 | 11 | 125885094 | 2.2842558352674e-06 | 18.9195474 | intronic | HYLS1 | . | HYLS1 | |
| 3M | rs528609331 | 11 | 125972300 | 7.07196590950882e-07 | 19.5812418 | intronic | CDON | . | CDON | |
| 3M | rs7104959 | 11 | 129976231 | 3.12645341825951e-06 | 17.9204611 | intronic | PRDM10 | . | PRDM10 | |
| 12M | rs117699122 | 12 | 451096 | 1.18257175082115e-06 | 21.4238256 | intergenic | CCDC77;B4GALNT3 | dist=8456;dist=8843 | CCDC77;B4GALNT3 | |
| 3M | rs189360484 | 12 | 1761344 | 3.94900202113111e-07 | 16.8078794 | intronic | ADIPOR2 | . | ADIPOR2 | |
| 12M | rs61918041 | 12 | 6572702 | 4.39893214125342e-06 | 7.97264643 | intronic | CHD4 | . | CHD4 | |
| 12M | rs113244573 | 12 | 6575219 | 4.49284300484012e-06 | 7.97095746 | intronic | CHD4 | . | | |
| 3M | rs141754456 | 12 | 19998198 | 1.82427554715572e-07 | 12.1460333 | intergenic | AEBP2;LINC02398 | dist=475971;dist=16487 | AEBP2 | |
| 3M | rs118184666 | 12 | 20271815 | 2.47036908420995e-06 | 10.9509803 | intergenic | LINC02468;PDE3A | dist=143914;dist=96722 | | |
| 3M | rs549931083 | 12 | 20363352 | 3.06794275866128e-07 | 14.3282694 | intergenic | LINC02468;PDE3A | dist=235451;dist=5185 | PDE3A | |
| 3M | rs151323346 | 12 | 20859090 | 2.59127051015029e-06 | 15.8903888 | intronic | SLCO1B3;SLCO1B3-SLCO1B7 | . | | |
| 3M | rs371879555 | 12 | 22863028 | 1.69721932161597e-06 | 20.1033765 | intergenic | ETNK1;LOC101928441 | dist=172363;dist=312608 | ETNK1 | |
| 3M | rs183466664 | 12 | 26668754 | 3.57331936763708e-06 | 16.0181882 | intronic | ITPR2 | . | ITPR2 | |
| 3M | rs77353774 | 12 | 28095919 | 1.00362822173041e-06 | 13.1054362 | intergenic | PTHLH;LOC729291 | dist=123186;dist=89706 | PTHLH | |

| | | | | | | | | | | |
|---|---|---|---|---|---|---|---|---|---|---|
| 3M | rs113167689 | 12 | 28283029 | 2.31830826822625e-07 | 14.1364135 | intronic | CCDC91 | . | | |
| 3M | rs17510814 | 12 | 28316036 | 2.30925017950923e-07 | 14.0387924 | intronic | CCDC91 | . | CCDC91 | |
| 3M | rs141756120 | 12 | 28358163 | 2.4075549984719e-07 | 14.278403 | intronic | CCDC91 | . | | |
| 3M | rs117991215 | 12 | 28358540 | 2.50265389686764e-07 | 14.333666 | intronic | CCDC91 | . | | |
| 3M | rs191930622 | 12 | 47890872 | 4.88013920852721e-06 | 19.6783579 | intronic | VDR | . | VDR | |
| 3M | rs56302696 | 12 | 47899047 | 4.02736708601484e-06 | 19.8959041 | intronic | VDR | . | | |
| 3M | rs185620578 | 12 | 48175616 | 2.41563563938621e-06 | 20.7232267 | intergenic | ASB8;CCDC184 | dist=18101;dist=8028 | ASB8;CCDC184 | |
| 3M | rs190806532 | 12 | 48468364 | 1.54808652729946e-06 | 21.326955 | intergenic | ZNF641;ANP32D | dist=117118;dist=4195 | ZNF641;ANP32D | |
| 3M | rs568658857 | 12 | 49460215 | 4.20762702895843e-06 | 14.2361193 | intronic | SPATS2 | . | SPATS2 | |
| 3M | rs137880949 | 12 | 62912517 | 1.74526564511423e-07 | 21.6084034 | intronic | PPM1H | . | | |
| 3M | rs191053292 | 12 | 63051500 | 2.51400610111527e-08 | 22.9266488 | intergenic | PPM1H;AVPR1A | dist=116350;dist=91259 | PPM1H;AVPR1A | |
| 3M | rs182437250 | 12 | 63214686 | 7.11750755969757e-07 | 18.8083451 | intergenic | AVPR1A;DPY19L2 | dist=63485;dist=344227 | | |
| 3M | rs12315614 | 12 | 64527177 | 4.12213451917224e-06 | 3.77710662 | intergenic | TBK1;RASSF3 | dist=25064;dist=83318 | TBK1,RASSF3 | |
| 12M | rs149616342 | 12 | 98785035 | 4.17235147721501e-06 | 11.2978214 | intronic | ANKS1B | . | ANKS1B | |
| 3M | rs76327548 | 12 | 100789188 | 2.03544908278967e-06 | 9.17842714 | intergenic | GAS2L3;ANO4 | dist=160900;dist=5588 | | |
| 3M | rs76904423 | 12 | 100794966 | 3.57310045627432e-06 | 9.80576231 | UTR5 | ANO4 | NM_001286615:c.-1068200>0;I | GAS2L3;ANO4 | |
| 3M | rs180764936 | 12 | 101104843 | 1.48879381343194e-06 | 19.3473053 | intronic | ANO4 | . | | |
| 3M | rs185855183 | 12 | 101111348 | 1.21523926305084e-06 | 18.7397727 | intronic | ANO4 | . | | |
| 12M | rs566724618 | 13 | 22889700 | 1.0727068950386e-06 | 22.3433705 | ncRNA_intronic | LINC00621 | . | SGCG | |
| 12M | rs147221953 | 13 | 22911583 | 1.86149707525764e-06 | 22.5809676 | ncRNA_intronic | LINC00621 | . | | |
| 3M | rs139598422 | 13 | 23312875 | 1.84258662797026e-08 | 20.4886522 | intronic | SGCG | . | | |
| 3M | rs143371352 | 13 | 46809699 | 1.20142384300403e-07 | 20.5341625 | intergenic | ESD;HTR2A | dist=12538;dist=21847 | ESD;HTR2A | |
| 3M | rs75186966 | 13 | 46821623 | 1.30324459401267e-07 | 20.2281345 | intergenic | ESD;HTR2A | dist=24462;dist=9923 | | |
| 3M | rs150077525 | 13 | 57405415 | 1.65979661476121e-06 | 12.9984875 | intergenic | PRR20E;PCDH17 | dist=235197;dist=226329 | PCDH17 | |
| 3M | rs534845494 | 13 | 57639730 | 5.5584801687088e-07 | 15.9431765 | intronic | PCDH17 | . | | |
| 3M | rs140062526 | 13 | 58711899 | 3.07703679852045e-06 | 14.0926088 | intergenic | LINC00374;DIAPH3 | dist=478782;dist=953688 | DIAPH3 | |
| 12M | rs113791989 | 13 | 61344755 | 7.13417559871126e-07 | 25.2049582 | intergenic | MIR3169;PCDH20 | dist=144874;dist=64931 | PCDH20 | |
| 12M | rs532269430 | 13 | 73480315 | 9.38644328296013e-07 | 18.6173752 | intergenic | KLF5;LINC00392 | dist=402772;dist=83929 | KLF5 | |
| 12M | rs368187808 | 13 | 73783371 | 1.40533873966635e-06 | 21.2991251 | intronic | KLF12 | . | KLF12 | |
| 12M | rs372194899 | 13 | 73783373 | 1.40312572084772e-06 | 21.3012101 | intronic | KLF12 | . | | |
| 3M | rs546286713 | 13 | 90758825 | 2.98443520289082e-06 | 15.7027841 | intergenic | LINC01049;LINC00410 | dist=223484;dist=132129 | | SLITRK5;GPC5 |
| 3M | rs567080482 | 13 | 94423151 | 2.95390135279034e-06 | 15.4999561 | intergenic | GPC6;DCT | dist=15132;dist=13660 | GPC6;DCT | |
| 3M | rs142928734 | 13 | 100948828 | 4.29382451218382e-06 | 17.009593 | ncRNA_intronic | NALCN | . | NALCN | |
| 3M | rs556680896 | 13 | 100950161 | 4.26743541179837e-06 | 17.0254659 | ncRNA_intronic | NALCN | . | | |
| 3M | rs184265355 | 13 | 107359004 | 7.66559973381936e-07 | 14.4271719 | intronic | FAM155A | . | FAM155A | |
| 3M | rs572961122 | 13 | 107360637 | 3.07761414918354e-06 | 13.3354896 | intronic | FAM155A | . | | |
| 3M | rs528809914 | 13 | 112386942 | 2.27269814316061e-07 | 19.5048552 | intronic | SPACA7 | . | | TUBGCP3 |
| 3M | rs180765647 | 13 | 113724338 | 4.20477084480749e-06 | 20.1246521 | intronic | GRK1 | . | | ATP4B;TMEM255B |
| 3M | rs138215817 | 14 | 22173619 | 6.27722428502198e-07 | 16.311019 | intergenic | OR4E1;LOC105370401 | dist=502284;dist=206292 | | SALL2; |
| 3M | rs74704551 | 14 | 29692681 | 1.97941397505543e-08 | 24.1990705 | intronic | PRKD1 | . | PRKD1 | |
| 3M | rs1686289 | 14 | 45791779 | 3.52212847732742e-06 | -2.2551539 | intergenic | LINC02303;LINC00871 | dist=76177;dist=272380 | | MIS18BP1;RPL10L |
| 3M | rs176783 | 14 | 45811710 | 3.87832460630423e-06 | 2.28010147 | intergenic | LINC02303;LINC00871 | dist=96108;dist=252449 | | |
| 3M | rs176786 | 14 | 45813767 | 3.37672684888795e-06 | 2.28475641 | intergenic | LINC02303;LINC00871 | dist=98165;dist=250392 | | |
| 3M | rs428110 | 14 | 45825457 | 4.15182083903445e-06 | 2.28482299 | intergenic | LINC02303;LINC00871 | dist=109855;dist=238702 | | |
| 3M | rs116862847 | 14 | 63674959 | 2.1204003875388e-07 | 15.3160369 | intergenic | WDR89;SGPP1 | dist=33088;dist=9258 | WDR89;SGPP1 | |
| 3M | rs569916471 | 14 | 75424639 | 3.10710931450174e-06 | 15.6404088 | intergenic | LINC01220;JDP2 | dist=128231;dist=3085 | JDP2 | |
| 3M | rs113767990 | 14 | 81251219 | 4.97864018338844e-06 | 14.5811155 | intergenic | LOC101928504;STON2 | dist=27839;dist=9431 | STON2 | |
| 3M | rs190251199 | 14 | 105124240 | 6.2650518901636e-07 | 20.0125292 | intergenic | LINC02298;JAG2 | dist=24744;dist=16758 | JAG2 | |
| 12M | rs190251199 | 14 | 105124240 | 1.9790595982044E-06 | 22.6385061 | intergenic | LINC02298;JAG2 | dist=24744;dist=16758 | | |
| 12M | rs187281112 | 15 | 32821186 | 3.3019635783423e-06 | 14.7343356 | intronic | FMN1 | . | FMN1 | |
| 12M | rs56324718 | 15 | 39987872 | 7.5163534768267e-07 | 12.5117374 | intronic | EIF2AK4 | . | EIF2AK4 | |
| 3M | rs185155853 | 15 | 40951902 | 3.01500290127566e-06 | 15.9164657 | intergenic | DLL4;CHAC1 | dist=12829;dist=1569 | | |
| 3M | rs144026361 | 15 | 40956471 | 2.67002672054584e-06 | 16.1372994 | UTR3 | CHAC1 | NM_001142776:c.*6970>0;NM | DLL4;CHAC1 | |
| 3M | rs558614420 | 15 | 41518672 | 3.91771199682616e-06 | 13.9750831 | intronic | RPAP1 | . | | LTK;TYRO3 |
| 3M | rs138109686 | 15 | 41759244 | 4.62000628076245e-06 | 13.5956705 | intronic | MGA | . | MGA | |
| 3M | rs145896760 | 15 | 41827024 | 4.80897800219791e-06 | 13.3688825 | UTR3 | MAPKBP1 | NM_014994:c.*15880>0;NM_0 | MAPKBP1 | |
| 3M | rs140642138 | 15 | 41832967 | 4.54678591939158e-06 | 13.5832221 | intronic | JMJD7;JMJD7-PLA2G4B | . | JMJD7;JMJD7-PLA2G4B | |
| 3M | rs6080 | 15 | 58545734 | 4.85754782149583e-06 | 5.66476745 | intronic | LIPC | . | | |
| 3M | rs145439370 | 15 | 58587566 | 9.02072868608363e-07 | 6.89328891 | intergenic | LIPC;ADAM10 | dist=17722;dist=1243 | LIPC;ADAM10 | |
| 3M | rs149425014 | 15 | 58659461 | 1.84020904634255e-06 | 7.55224792 | intronic | ADAM10 | . | | |
| 3M | rs146442492 | 15 | 58689916 | 3.24578114232023e-06 | 7.05925858 | intronic | ADAM10 | . | | |
| 3M | rs193253461 | 15 | 58937154 | 3.76206055565965e-07 | 10.5374468 | intergenic | SLTM;RNF111 | dist=3475;dist=50509 | SLTM;RNF111 | |
| 3M | rs184117160 | 15 | 59112107 | 1.25384693259541e-06 | 10.3417236 | intronic | CCNB2 | . | CCNB2 | |
| 3M | rs80292573 | 15 | 59142887 | 7.87214777000235e-07 | 6.55323491 | intronic | MYO1E | . | MYO1E | |
| 3M | rs182303755 | 15 | 59342593 | 4.69869428218951e-07 | 10.5479917 | intronic | MYO1E | . | | |
| 3M | rs138217865 | 15 | 93849653 | 1.29412742467123e-06 | 16.709641 | intergenic | LOC105370980;LINC02207 | dist=641605;dist=6907 | | RGMA;MCTP2 |
| 12M | rs80212581 | 16 | 6362966 | 6.33070894509269e-07 | 22.2762003 | intronic | RBFOX1 | . | | |
| 12M | rs140276610 | 16 | 6383734 | 1.75581952039286e-06 | 22.1806577 | intronic | RBFOX1 | . | | |
| 12M | rs138164904 | 16 | 6808238 | 3.60740717035677e-08 | 23.892641 | intronic | RBFOX1 | . | RBFOX1 | |
| 3M | rs553840536 | 16 | 25686574 | 3.61488566678171e-06 | 19.6203442 | intergenic | ZKSCAN2;HS3ST4 | dist=428729;dist=5385 | ZKSCAN2;HS3ST4 | |
| 3M | rs183817723 | 16 | 59268871 | 2.48366342700424e-06 | 17.5860507 | intergenic | GOT2;APOOP5 | dist=534555;dist=485270 | GOT2 | |
| 3M | rs144954214 | 16 | 76145464 | 8.96489764315735e-07 | 21.8113263 | intergenic | CPHXL;CNTNAP4 | dist=418974;dist=131937 | CNTNAP4 | |
| 3M | rs529523094 | 16 | 77681654 | 1.09798611725829e-06 | 18.1584346 | intergenic | ADAMTS18;NUDT7 | dist=246620;dist=40838 | NUDT7 | |
| 12M | rs116897913 | 17 | 16935848 | 4.78180386603079e-06 | 20.1396765 | intergenic | TBC1D27P;TNFRSF13B | dist=2672;dist=3233 | TNFRSF13B | |
| 3M | rs146728064 | 17 | 19362127 | 3.79796041699169e-06 | 12.6400411 | intronic | B9D1 | . | B9D1 | |
| 12M | rs188353596 | 17 | 48001236 | 6.85080774831596e-07 | 21.8057947 | intergenic | CDK5RAP3;COPZ2 | dist=19450;dist=24931 | CDK5RAP3;COPZ2 | |
| 3M | rs184613584 | 17 | 50430860 | 5.40834238559884e-07 | 14.1377974 | intronic | ACSF2 | . | ACSF2 | |
| 3M | rs191271637 | 17 | 54045899 | 2.76242881040307e-06 | 18.379836 | intergenic | KIF2B;TOM1L1 | dist=220706;dist=854792 | KIF2B;TOM1L1 | |
| 12M | rs112148840 | 17 | 68237633 | 3.82795090389498e-06 | 24.0241075 | intronic | AMZ2 | . | AMZ2 | |
| 12M | rs2606194 | 17 | 79214741 | 9.99647458252552e-07 | -5.6956979 | intronic | RBFOX3 | . | RBFOX3 | |
| 12M | rs147669485 | 18 | 29023686 | 9.8819660331789e-08 | 22.1107639 | intergenic | CDH2;MIR302F | dist=846556;dist=1275226 | CDH2 | |
| 3M | rs185819304 | 18 | 29421615 | 4.31513948321026e-07 | 19.9401113 | intergenic | CDH2;MIR302F | dist=1244486;dist=877296 | | |
| 3M | rs187942235 | 18 | 29450465 | 2.77341075652478e-07 | 20.3968658 | intergenic | CDH2;MIR302F | dist=1273336;dist=848446 | CDH2 | |

| | | | | | | | | | | |
|---|---|---|---|---|---|---|---|---|---|---|
| 3M | rs139493286 | 18 | 31236056 | 3.23770514196012e-06 | 17.4603093 | intergenic | DSC1;DSG1 | dist=73200;dist=82104 | DSG1 | |
| 3M | rs143538552 | 18 | 31470299 | 1.40612679764676e-07 | 19.1457218 | intronic | DSG3 | . | DSG1;DSG3 | |
| 3M | rs373746073 | 18 | 31478421 | 1.55466285461099e-07 | 19.0847197 | UTR3 | DSG3 | NM_001944:c.*21610>0 | | |
| 3M | rs146333745 | 18 | 57830225 | 1.04499182739545e-06 | 17.4215784 | intergenic | ATP8B1;NEDD4L | dist=26910;dist=214001 | ATP8B1;NEDD4L | |
| 12M | rs559152067 | 18 | 68362375 | 1.80839131277764e-06 | 24.7443692 | intergenic | LOC643542;TMX3 | dist=462756;dist=311313 | TMX3 | |
| 3M | rs185464792 | 19 | 18686561 | 5.22729611714582e-07 | 22.5896015 | intronic | CRTC1 | . | CRTC1 | |
| 3M | rs186768950 | 19 | 18695314 | 4.67656376928778e-07 | 22.7152061 | intronic | CRTC1 | . | | |
| 3M | rs541288561 | 19 | 18758635 | 6.19920480169239e-07 | 15.6119012 | intronic | CRTC1 | . | | |
| 3M | rs559008174 | 19 | 18765249 | 6.10577196408926e-07 | 15.6025661 | intronic | CRTC1 | . | | |
| 3M | rs570407448 | 19 | 18769220 | 7.02455289936315e-07 | 15.4977984 | intronic | CRTC1 | . | | |
| 3M | rs546144116 | 19 | 19452530 | 2.29332366104571e-07 | 23.1354815 | intronic | GATAD2A | . | GATAD2A | |
| 3M | rs560206697 | 19 | 20546292 | 4.01812636832912e-08 | 24.9989943 | intronic | ZNF737 | . | ZNF737 | |
| 3M | rs111285015 | 19 | 22940396 | 3.3078789506342e-09 | 27.3347642 | intergenic | ZNF723;ZNF728 | dist=81729;dist=34487 | ZNF728 | |
| 3M | rs1008091735 | 19 | 30599192 | 5.61396767442543e-07 | 18.4689243 | intronic | ZNF536 | . | ZNF536 | |
| 3M | rs148433854 | 19 | 30605571 | 4.40546670794092e-07 | 18.6267585 | intronic | ZNF536 | . | | |
| 12M | rs367732718 | 19 | 35427102 | 3.38991794276911e-06 | 23.8784923 | intergenic | LINC01531;FFAR2 | dist=10262;dist=21155 | FFAR2 | |
| 12M | rs562831582 | 19 | 50731053 | 5.70199111188332e-07 | 21.0856548 | intergenic | CLEC11A;GPR32 | dist=5345;dist=39411 | CLEC11A | |
| 12M | rs62192733 | 20 | 1947511 | 2.76211842636971e-06 | 15.744618 | ncRNA_exonic | PDYN | . | | SIRPA;STK35 |
| 12M | rs547186621 | 20 | 6126094 | 2.39683963851735e-06 | 21.5510829 | intergenic | FERMT1;CASC20 | dist=3064;dist=300638 | FERMT1 | |
| 3M | rs2327968 | 20 | 15832846 | 4.58567809597369e-06 | 6.15899856 | intronic | MACROD2 | . | MACROD2 | |
| 3M | rs2876414 | 20 | 15833059 | 4.41048907235012e-06 | 6.81910907 | intronic | MACROD2 | . | | |
| 3M | rs140788628 | 20 | 15877856 | 2.32626831948532e-07 | 11.8039284 | intronic | MACROD2 | . | | |
| 3M | rs559228693 | 20 | 15982684 | 3.15016069009906e-06 | 14.3643551 | ncRNA_intronic | LOC613266 | . | | |
| 12M | rs138733283 | 20 | 32087884 | 5.87309369634527e-07 | 18.2744744 | intronic | HCK | . | HCK | |
| 12M | rs149859280 | 20 | 32090367 | 5.71700141373349e-07 | 18.3057707 | intronic | HCK | . | | |
| 12M | rs146249289 | 20 | 32094428 | 2.01850841331451e-07 | 19.6109527 | intronic | HCK | . | | |
| 12M | rs145791959 | 20 | 32134626 | 8.67001886662025e-07 | 17.9091953 | intronic | TM9SF4 | . | TM9SF4 | |
| 12M | rs193041547 | 20 | 32184077 | 1.80079654856719e-07 | 19.6791358 | intergenic | TM9SF4;TSPY26P | dist=16819;dist=5069 | | |
| 12M | rs138055631 | 20 | 32192841 | 1.06243999550455e-06 | 17.6647762 | UTR3 | PLAGL2 | NM_002657:c.*36110>0 | PLAGL2 | |
| 12M | rs145421321 | 20 | 32274323 | 1.72232779371331e-06 | 17.0212611 | intergenic | POFUT1;KIF3B | dist=35665;dist=3328 | POFUT1;KIF3B | |
| 12M | rs143432612 | 20 | 32304972 | 1.74726298083525e-06 | 16.9387232 | intronic | KIF3B | . | | |
| 12M | rs139816293 | 20 | 32333540 | 1.16528363587405e-06 | 17.1384248 | UTR3 | KIF3B | NM_004798:c.*22210>0 | | |
| 12M | rs200198574 | 20 | 32358851 | 2.44213593266147e-06 | 16.5282735 | intronic | ASXL1 | . | ASXL1 | |
| 12M | rs148157126 | 20 | 32361036 | 1.42669073977956e-07 | 19.7865001 | intronic | ASXL1 | . | | |
| 12M | rs192855100 | 20 | 32520938 | 4.17301607264236e-07 | 20.1986097 | intronic | NOL4L | . | NOL4L | |
| 3M | rs557092705 | 20 | 35601989 | 4.54067714932956e-06 | 19.4769546 | ncRNA_exonic | FER1L4 | . | | ERGIC3;SPAG4 |
| 12M | rs2427460 | 20 | 62959430 | 5.30063377285508e-07 | -2.574044 | intronic | SLC17A9 | . | SLC17A9 | |
| 3M | rs184785969 | 21 | 15799112 | 8.26040996676763e-08 | 19.8736486 | intronic | USP25 | . | USP25 | |
| 3M | rs117280553 | 21 | 15834844 | 5.79538719308743e-08 | 20.087417 | intronic | USP25 | . | | |
| 3M | rs79486609 | 21 | 15872687 | 4.53094466952756e-08 | 20.6624886 | intronic | USP25 | . | | |
| 3M | rs73227413 | 21 | 21764653 | 2.74560920737872e-06 | 5.96205514 | ncRNA_intronic | LINC01425 | . | | |
| 3M | rs75024143 | 21 | 21784226 | 2.0964109268482e-06 | 9.81254241 | ncRNA_intronic | LINC01425 | . | | |
| 3M | rs192134381 | 21 | 22078395 | 5.23445134161758e-09 | 21.788812 | ncRNA_intronic | LINC01687 | . | | NCAM2;MRPL39 |
| 3M | rs118183140 | 21 | 34105187 | 1.16709964426479e-06 | 7.5893248 | UTR3 | SLC5A3 | NM_006933:c.*78320>0 | SLC5A3 | |
| 3M | rs183586634 | 21 | 37390730 | 1.72123504106205e-06 | 14.3132202 | intronic | DYRK1A | . | DYRK1A | |
| 3M | rs117185941 | 21 | 37394182 | 1.2357093777619e-06 | 15.3808786 | intronic | DYRK1A | . | | |
| 3M | rs118084887 | 21 | 37491518 | 1.91359719468201e-06 | 14.9382616 | intronic | DYRK1A | . | | |
| 12M | rs9636964 | 21 | 39932840 | 2.72523192147064e-07 | -4.2543192 | intergenic | PCP4;DSCAM | dist=3448;dist=78161 | PCP4;DSCAM | |
| 12M | rs9305683 | 21 | 39933795 | 8.57246540213203e-07 | -4.1163679 | intergenic | PCP4;DSCAM | dist=4403;dist=77206 | | |
| 12M | rs9974985 | 21 | 39935648 | 1.59836223443899e-07 | -4.3459395 | intergenic | PCP4;DSCAM | dist=6256;dist=75353 | | |
| 12M | rs7275595 | 21 | 39935998 | 3.24552446398997e-07 | -4.2609754 | intergenic | PCP4;DSCAM | dist=6606;dist=75003 | | |
| 12M | rs1005412 | 21 | 39937023 | 1.75853470115381e-07 | -4.4521096 | intergenic | PCP4;DSCAM | dist=7631;dist=73978 | | |
| 12M | rs9981433 | 21 | 39937640 | 1.14042775714184e-06 | -4.1220124 | intergenic | PCP4;DSCAM | dist=8248;dist=73361 | | |
| 3M | rs150539922 | 21 | 41856807 | 4.79442196264294e-06 | 16.6696937 | intronic | PRDM15 | . | PRDM15 | |
| 3M | rs113625788 | 22 | 19981659 | 1.17014602462676e-06 | 11.8084829 | exonic | ARVCF | . | ARVCF | |
| 3M | rs78547898 | 22 | 32428291 | 4.85417555255089e-07 | 20.9098395 | intronic | BPIFC | . | | RTCB;FBXO7 |
| 12M | rs77297738 | 22 | 34077554 | 2.01836979224915e-07 | 10.2480572 | intergenic | LARGE1;LINC02885 | dist=154731;dist=679113 | LARGE1 | |
| 12M | rs74572772 | 22 | 34096660 | 1.39505561568774e-07 | 10.2225183 | intergenic | LARGE1;LINC02885 | dist=173837;dist=660007 | | |
| 12M | rs80019988 | 22 | 34097658 | 1.52143693387141e-07 | 10.0623306 | intergenic | LARGE1;LINC02885 | dist=174835;dist=659009 | | |
| 3M | rs541680196 | 22 | 40132086 | 1.06912975962765e-06 | 14.3867808 | intronic | TNRC6B | . | TNRC6B | |
| 3M | rs185139807 | 22 | 40198777 | 1.02186831937043e-06 | 14.4330526 | intronic | TNRC6B | . | | |
| 3M | rs141127122 | 22 | 40208435 | 3.1593298841889e-06 | 15.3824875 | intronic | TNRC6B | . | | |
| 3M | rs148998974 | 22 | 40224526 | 8.65242968473446e-07 | 14.5129532 | intronic | TNRC6B | . | | |
| 3M | rs555040883 | 22 | 40235472 | 3.05234476142884e-06 | 15.4273069 | intronic | TNRC6B | . | | |
| 3M | rs182959028 | 22 | 45427152 | 2.37697466245549e-06 | 11.944461 | intronic | RIBC2 | . | RIBC2 | |
| 3M | rs150946694 | 22 | 46457283 | 3.3001729159907e-06 | 15.3082479 | intronic | CELSR1 | . | CELSR1 | |

Combined Results

7 Risk loci cluster these 11 SNPs

GENOME-WIDE RISK LOCI

SUGGESTIVE RISK LOCI (not genome-wide)

SUGGESTIVE RISK LOCI (including genome-wide)

TOTAL GENOME-WIDE & SUGGESTIVE

MERGED (Sum minus overlap)

Overlapping Risk Loci

ALK, DCANP1, JAG2, KCND3, PPARGC1A, STAG1, TBLXR1

GENOME-LEVEL SNPS

AGTR1, CDH2, ETAA1, MTX2, SGCG

SUGGESTIVE SNPS

TOTAL SNPS

SNPS REPEATED 12&3 MO

**Supplementary Table S4. Target Gene Prioritization**

**Co-localization of associated SNPs with eQTLs or sQTLs and their target genes**

**NOTES**

SNPs are grouped by risk locus and ordered by chromosomal position; risk loci are boxed; the one risk locus with SNPs of genome-wide signficance is highlighted in gray

**HEADERS**

QT: quantitiative trait; MAF ALPHA: minor allele frequency, dbSNP ALPHA project; rsid: reference SNP cluster ID, chr: chromosome;
pos_38, position of SNP on GRCh38 reference panel; REF and ALT, reference allele and alternative allele; Func.refGene: SNP location with respect to nearest gene;
Gene.refGene; nearest gene upstream and downstream; GeneDetail.refGene: distance to nearest gene upstream and downstream;
AOP: aqueous outflow pathway; QTL: quantitative trait locus; eQTL: expression quantitative trait locus; sQTL: slicing quantitative trait locus;

**Gray Key for MAF ALPHA**

light gray shaded: common SNP; mid-gray shaded: low frequency SNP; dark gray shaded: rare SNP

**Counts**

Loci and SNP counts are at bottom

| QT | MAF ALPHA | rsid | chr | pos_38 | REF | ALT | Func.refGene | Gene.refGene | GeneDetail.refGene | Closest gene(s) AOP-expressed | QTL driven gene, AOP-expressed | QTL type | Effect | QTL driven gene, AOP-expressed | QTL type | Effect | QTL driven AOP-expressed gene | QTL type | Effect |
|---|---|---|---|---|---|---|---|---|---|---|---|---|---|---|---|---|---|---|---|
| 12 mo | 0.093415 | rs17127656 | 1 | 65477788 | C | T | intronic | LEPR | . | LEPR | | | | | | | | | |
| 12 mo | 0.08334 | rs7518849 | 1 | 65483108 | T | C | intronic | LEPR | . | | LEPROT | eQTL | Positive | | | | | | |
| 12 mo | 0.069508 | rs11579567 | 1 | 65491458 | C | A | intronic | LEPR | . | | LEPROT | eQTL | Positive | | | | | | |
| 12 mo | 0.083589 | rs7534177 | 1 | 65500037 | A | G | intronic | LEPR | . | | LEPROT | eQTL | Positive | | | | | | |
| 3 mo | 0.002859 | rs140420703 | 1 | 102337928 | T | G | intergenic | OLFM3;COL11A1 | dist=340694;dist=538539 | COL11A1 | | | | | | | | | |
| 3 mo | 0.001323 | rs563167766 | 1 | 102400809 | G | A | intergenic | OLFM3;COL11A1 | dist=403575;dist=475658 | | | | | | | | | | |
| 3 mo | 0.009519 | rs77180278 | 1 | 102496326 | T | C | intergenic | OLFM3;COL11A1 | dist=499092;dist=380141 | | COL11A1 | sQTL | Negative | | | | | | |
| 3 mo | 0.017196 | rs112351653 | 1 | 102754804 | T | C | intergenic | OLFM3;COL11A1 | dist=757570;dist=121663 | | COL11A1 | sQTL | Negative | | | | | | |
| 3 mo | 0.000583 | rs180926150 | 1 | 102760770 | C | T | intergenic | OLFM3;COL11A1 | dist=763536;dist=115697 | | | | | | | | | | |
| 3 mo | 0.017561 | rs114413507 | 1 | 102953612 | T | C | intronic | COL11A1 | intron 41 of 66 | | COL11A1 | sQTL | Negative | | | | | | |
| 3 mo | 0.01704 | rs116672066 | 1 | 103007360 | G | A | intronic | COL11A1 | intron 15 of 66 | | COL11A1 | sQTL | Negative | | | | | | |
| 3 mo | 0.016995 | rs111928960 | 1 | 103168079 | G | A | intergenic | COL11A1;LOC101928436 | dist=59557;dist=325967 | | COL11A1 | sQTL | Negative | | | | | | |
| 3 mo | 0.016305 | rs113221952 | 1 | 103288418 | A | G | intergenic | COL11A1;LOC101928436 | dist=179896;dist=205628 | | COL11A1 | sQTL | Negative | | | | | | |
| 3&12 mo | 0.017673 | rs76098744 | 1 | 111806750 | C | T | intronic | KCND3 | . | KCND3 | | | | | | | | | |
| 3&12 mo | 0.009033 | rs74683551 | 1 | 111811796 | G | A | intronic | KCND3 | . | | WNT2B | eQTL | Positive | | | | | | |
| 3 mo | 0.059723 | rs2274996 | 1 | 229668791 | C | T | intergenic | URB2;LINC01682 | dist=8591;dist=206759 | URB2 | TAF5L | sQTL | Negative | | | | | | |
| 3 mo | 0.096148 | rs2274997 | 1 | 229668899 | A | G | intergenic | URB2;LINC01682 | dist=8699;dist=206651 | | TAF5L | sQTL | Negative | | | | | | |
| 3 mo | 0.05264 | rs2891865 | 1 | 229670621 | A | G | intergenic | URB2;LINC01682 | dist=10421;dist=204929 | | | | | | | | | | |
| 3 mo | 0.069533 | rs2385790 | 1 | 229671745 | C | T | intergenic | URB2;LINC01682 | dist=11545;dist=203805 | | TAF5L | sQTL | Negative | | | | | | |
| 3 mo | 0.025802 | rs12024557 | 1 | 229676610 | A | C | intergenic | URB2;LINC01682 | dist=16410;dist=198940 | | | | | | | | | | |
| 3 mo | 0.052214 | rs4562666 | 1 | 229689023 | T | C | intergenic | URB2;LINC01682 | dist=28823;dist=186527 | | | | | | | | | | |
| 3 mo | 0.090471 | rs12036586 | 1 | 229690631 | G | A | intergenic | URB2;LINC01682 | dist=30431;dist=184919 | | not AOP expressed | sQTL | Negative | | | | | | |
| 3 mo | 0.084159 | rs16850124 | 1 | 229695584 | T | C | intergenic | URB2;LINC01682 | dist=35384;dist=179966 | | TAF5L | sQTL | Negative | | | | | | |
| 3 mo | 0.060487 | rs12045643 | 1 | 229698303 | C | T | intergenic | URB2;LINC01682 | dist=38103;dist=177247 | | | | | | | | | | |
| 3 mo | unknown | rs17746486 | 2 | 95056864 | C | T | intergenic | MAL;MRPS5 | dist=2872;dist=28507 | MAL;MRPS5 | GPAT2 | eQTL | Positive | ZNF514 | eQTL | Positive | ZNF514 | sQTL | Positive |
| 3 mo | 0.015 | rs73085348 | 3 | 42669729 | A | G | intergenic | ZBTB47;KLHL40 | dist=2149;dist=15808 | ZBTB47 | NKTR | eQTL | Negative | KRBOX1 | eQTL | Negative | | | |
| 3 mo | 0.007 | rs139943877 | 3 | 155733500 | G | A | intronic | PLCH1 | . | PLCH1 | GMPS | eQTL | Positive | PLCH1 | eQTL | Negative | | | |
| 12 mo | 0.014 | rs76356799 | 3 | 179875980 | G | A | intronic | PEX5L | . | PEX5L | USP13 | eQTL | Positive | | | | | | |
| 12 mo | 0.069 | rs17615362 | 4 | 169012936 | G | A | intergenic | CBR4;SH3RF1 | dist=2681;dist=81323 | SH3RF1 | CBR4 | eQTL | Positive | CBR4 | sQTL | Pos/Neg | PALLD | eQTL | Negative |
| 12 mo | 0.023 | rs17543620 | 4 | 169013574 | T | C | intergenic | CBR4;SH3RF1 | dist=3319;dist=80685 | | CBR4 | eQTL | Positive | CBR4 | sQTL | Pos/Neg | PALLD | eQTL | Negative |
| 3 mo | 0.002 | rs111407636 | 5 | 95744325 | C | T | intronic | RHOBTB3 | . | RHOBTB3 | | | | | | | | | |
| 3 mo | unknown | rs111676272 | 5 | 95758594 | C | A | intronic | RHOBTB3 | . | | | | | | | | | | |
| 3 mo | 0.008602 | rs75848314 | 5 | 95762636 | T | C | intronic | RHOBTB3 | . | | PCSK1 | eQTL | Negative | | | | | | |
| 3 mo | 0.012388 | rs111846247 | 5 | 95775939 | T | C | intronic | RHOBTB3 | . | | GLRX | eQTL | Positive | | | | | | |
| 3 mo | 0.030 | rs17776100 | 7 | 6386848 | G | A | intronic | RAC1 | . | RAC1 | CCZ1 | eQTL | Negative | CCZ1 | sQTL | Pos/Neg | CCZ1B | eQTL | Positive |
| 3 mo | 0.28 | rs9643828 | 8 | 54616513 | C | T | intronic | RP1 | . | SOX17;XKR4 | not AOP expressed | eQTL | Positive | | | | | | |
| 3 mo | 0.365421 | rs423841 | 8 | 54643509 | G | A | intronic | RP1 | . | | XKR4 | eQTL | Positive | | | | | | |
| 3 mo | 0.24537 | rs433324 | 8 | 54652049 | A | G | intronic | RP1 | . | | XKR4 | eQTL | Positive | | | | | | |
| 3 mo | 0.31998 | rs369623 | 8 | 54659380 | A | C | intronic | RP1 | . | | XKR4 | eQTL | Positive | | | | | | |
| 3 mo | 0.292979 | rs446222 | 8 | 54662400 | G | A | intronic | RP1 | . | | not AOP expressed | eQTL | Positive | | | | | | |
| 3 mo | 0.499229 | rs432393 | 8 | 54667738 | C | T | intronic | RP1 | . | | XKR4 | eQTL | Positive | | | | | | |
| 3 mo | 0.380572 | rs3098298 | 8 | 54670278 | C | T | intronic | RP1 | . | | XKR4 | eQTL | Positive | | | | | | |
| 3 mo | 0.380413 | rs367179 | 8 | 54675056 | T | C | intronic | RP1 | . | | XKR4 | eQTL | Positive | | | | | | |
| 3 mo | 0.2961237 | rs382476 | 8 | 54678415 | G | A | intronic | RP1 | . | | XKR4 | eQTL | Positive | | | | | | |
| 3 mo | 0.320148 | rs384543 | 8 | 54679049 | G | A | intronic | RP1 | . | | XKR4 | eQTL | Positive | | | | | | |
| 3 mo | 0.392869 | rs405226 | 8 | 54679776 | A | G | intronic | RP1 | . | | XKR4 | eQTL | Positive | | | | | | |
| 3 mo | 0.313393 | rs384127 | 8 | 54684929 | G | A | intronic | RP1 | . | | XKR4 | eQTL | Positive | | | | | | |
| 3 mo | 0.296631 | rs858397 | 8 | 54702130 | A | G | intronic | RP1 | . | | XKR4 | eQTL | Negative | | | | | | |
| 3 mo | 0.345139 | rs2375537 | 8 | 54706948 | C | T | intronic | RP1 | . | | XKR4 | eQTL | Negative | | | | | | |
| 3 mo | 0.393862 | rs720372 | 8 | 54716077 | G | A | intronic | RP1 | . | | XKR4 | eQTL | Negative | | | | | | |
| 3 mo | 0.311297 | rs1437781 | 8 | 54717292 | T | C | intronic | RP1 | . | | XKR4 | eQTL | Negative | | | | | | |
| 3 mo | 0.339565 | rs1595406 | 8 | 54718055 | A | G | intronic | RP1 | . | | XKR4 | eQTL | Negative | | | | | | |
| 3 mo | 0.2992666 | rs1437782 | 8 | 54720202 | C | T | exonic | RP1 | . | | XKR4 | eQTL | Negative | | | | | | |
| 3 mo | 0.304288 | rs10105693 | 8 | 54727912 | C | T | intronic | RP1 | . | | XKR4 | eQTL | Negative | | | | | | |
| 3 mo | 0.34109 | rs2375536 | 8 | 54728162 | T | C | intronic | RP1 | . | | XKR4 | eQTL | Negative | | | | | | |
| 3 mo | 0.31424 | rs4737674 | 8 | 54749094 | C | A | intronic | RP1 | . | | XKR4 | eQTL | Negative | | | | | | |
| 3 mo | 0.306432 | rs11987234 | 8 | 54757269 | A | G | intronic | RP1 | . | | XKR4 | eQTL | Negative | | | | | | |
| 3 mo | 0.309582 | rs13277510 | 8 | 54761589 | G | A | intronic | RP1 | . | | XKR4 | eQTL | Negative | | | | | | |
| 3 mo | 0.372041 | rs12548593 | 8 | 54762057 | G | T | intronic | RP1 | . | | XKR4 | eQTL | Negative | | | | | | |
| 3 mo | 0.394494 | rs1812506 | 8 | 54763541 | A | G | intronic | RP1 | . | | XKR4 | eQTL | Negative | | | | | | |
| 3 mo | 0.313335 | rs16920698 | 8 | 54765874 | G | A | intronic | RP1 | . | | XKR4 | eQTL | Negative | | | | | | |
| 3 mo | 0.313298 | rs1561297 | 8 | 54765978 | A | C | intronic | RP1 | . | | XKR4 | eQTL | Negative | | | | | | |
| 3 mo | 0.309423 | rs4737676 | 8 | 54766986 | G | A | intronic | RP1 | . | | XKR4 | eQTL | Negative | | | | | | |
| 3 mo | 0.350238 | rs2083123 | 8 | 54767758 | C | T | intronic | RP1 | . | | XKR4 | eQTL | Negative | | | | | | |
| 3 mo | 0.302059 | rs983248 | 8 | 54768232 | C | T | intronic | RP1 | . | | XKR4 | eQTL | Negative | | | | | | |
| 3 mo | 0.312123 | rs1391463 | 8 | 54769316 | T | G | intronic | RP1 | . | | XKR4 | eQTL | Negative | | | | | | |
| 3 mo | 0.331682 | rs10958428 | 8 | 54773081 | A | G | intronic | RP1 | . | | XKR4 | eQTL | Negative | | | | | | |
| 3 mo | 0.290454 | rs13278605 | 8 | 54775611 | C | T | intronic | RP1 | . | | XKR4 | eQTL | Negative | | | | | | |
| 3 mo | 0.178507 | rs13276543 | 8 | 54775614 | G | T | intronic | RP1 | . | | XKR4 | eQTL | Negative | | | | | | |
| 3 mo | 0.141896 | rs7822082 | 8 | 54777660 | T | C | intronic | RP1 | . | | XKR4 | eQTL | Negative | | | | | | |
| 3 mo | 0.313945 | rs4737201 | 8 | 54778898 | C | T | intronic | RP1 | . | | XKR4 | eQTL | Negative | | | | | | |
| 3 mo | 0.420064 | rs7843693 | 8 | 54779552 | G | A | intronic | RP1 | . | | XKR4 | eQTL | Negative | | | | | | |
| 3 mo | 0.222222 | rs1396896 | 8 | 54782750 | A | G | intronic | RP1 | . | | XKR4 | eQTL | Negative | | | | | | |
| 3 mo | 0.224434 | rs2375219 | 8 | 54785735 | C | T | intronic | RP1 | . | | | | | | | | | | |
| 3 mo | 0.261909 | rs1391462 | 8 | 54787221 | C | A | intronic | RP1 | . | | XKR4 | eQTL | Negative | | | | | | |
| 3 mo | 0.354923 | rs12678939 | 8 | 54792461 | A | G | intronic | RP1 | . | | | | | | | | | | |
| 3 mo | 0.364267 | rs1498183 | 8 | 54804345 | C | T | intronic | RP1 | . | | | | | | | | | | |
| 12 mo | unknown | rs12114488 | 8 | 61760164 | G | A | intergenic | MIR4470;LINC02155 | dist=45305;dist=129675 | ASPH | ASPH | eQTL | Negative | | | | | | |
| 12 mo | 0.023 | rs144138711 | 9 | 97160137 | T | C | ncRNA_intronic | ANKRD18CP | . | CTSV;CCDC180 | CCDC180 | sQTL | Pos/Neg | | | | | | |
| 3 mo | 0.017 | rs149949098 | 11 | 95366702 | G | A | intergenic | LOC100129203;FAM76B | dist=132298;dist=402251 | FAM76B | ENDOD1 | eQTL | Positive | | | | | | |
| 12 mo | 0.007191 | rs145791959 | 20 | 32134626 | G | A | intronic | TM9SF4 | . | TM9SF4 | LINC00028 | eQTL | Negative | | | | | | |
| 12 mo | 0.004852 | rs193041547 | 20 | 32184077 | T | C | intergenic | TM9SF4;TSPY26P | dist=16819;dist=5069 | | | | | | | | | | |
| 12 mo | 0.007191 | rs138055631 | 20 | 32192841 | G | A | UTR3 | PLAGL2 | NM_002657:c.*36110>0 | PLAGL2 | LINC00028 | eQTL | Negative | | | | | | |
| 12 mo | 0.007191 | rs145421321 | 20 | 32274323 | C | T | intergenic | POFUT1;KIF3B | dist=35665;dist=3328 | POFUT1;KIF3B | LINC00028 | eQTL | Negative | | | | | | |
| 12 mo | 0.00667 | rs143432612 | 20 | 32304972 | C | T | intronic | KIF3B | . | | LINC00028 | eQTL | Negative | | | | | | |
| 12 mo | 0.006569 | rs139816293 | 20 | 32333540 | C | T | UTR3 | KIF3B | NM_004798:c.*22210>0 | | LINC00028 | eQTL | Negative | | | | | | |
| 12 mo | 0.481741 | rs2427460 | 20 | 62959430 | T | C | intronic | SLC17A9 | . | SLC17A9 | SLC17A9 | eQTL | Negative | SLC17A9 | sQTL | Positive | DIDO1 | sQTL | Positive |
| 12 mo | 0.108413 | rs9636964 | 21 | 39932840 | A | G | intergenic | PCP4;DSCAM | dist=3448;dist=78161 | PCP4;DSCAM | PCP4 | eQTL | Negative | | | | | | |
| 12 mo | 0.08756 | rs9305683 | 21 | 39933795 | G | A | intergenic | PCP4;DSCAM | dist=4403;dist=77206 | | | | | | | | | | |
| 12 mo | 0.116287 | rs9974985 | 21 | 39935648 | G | A | intergenic | PCP4;DSCAM | dist=6256;dist=75353 | | | | | | | | | | |
| 12 mo | 0.092658 | rs7275595 | 21 | 39935998 | G | A | intergenic | PCP4;DSCAM | dist=6606;dist=75003 | | | | | | | | | | |
| 12 mo | 0.096064 | rs1005412 | 21 | 39937023 | A | G | intergenic | PCP4;DSCAM | dist=7631;dist=73978 | | | | | | | | | | |
| 12 mo | 0.122456 | rs9981433 | 21 | 39937640 | G | T | intergenic | PCP4;DSCAM | dist=8248;dist=73361 | | | | | | | | | | |

18 risk loci cluster SNPs that co-localize with QTLs: 6.87%
Total SNPs: 93
common SNPs: 67.74%
15 rare, 13 low frequency, 63 common, 3 unknown

**Supplementary Table S4. Target Gene Prioritization**
**Prioritized target gene lists from the Indianapolis-1 discovery cohort analysis**

**NOTES**
Alphabetical ordering of the full prioritized gene list highlights functionally similar paralogues or gene family members by grouping them together. We counted 41 such paralogous groups with shared function.
Information on gene product function is from the GeneCards Suite
**HEADERS**
AOP: aqueous outflow pathway; DEG: differentially expressed gene; eQTL: expression quantitative trait locus; TM: trabecular meshwork

| Indianapolis-1 discovery cohort | | | | | |
|---|---|---|---|---|---|
| AOP-expressed | AOP-expressed up, downstream | AOP-expressed eQTLs | Combined | Paralogue groups | Gene product function |
| ABCG2 | AGTR1 | ASPH | ABCG2 | | Superfamily of ATP-binding cassette (ABC) transporters, white subfamily |
| ACADSB | ALDH8A1 | CBR4 | ACADSB | | Acyl-CoA dehydrogenase family of enzymes that catalyze the dehydrogenation of acyl-CoA derivatives in the metabolism of fatty acids or branch chained amino acids |
| ACSF2 | AP1AR | CCDC180 | ACSF2 | ACSF2 | Acyl-CoA synthase family, catalyzes the initial reaction in fatty acid metabolism, by forming a thioester with CoA |
| ACSL3 | AP2A2 | CCZ1 | ACSL3 | ACSL3 | " |
| ADAM10 | ARAP2 | CCZ1B | ADAM10 | | ADAM family transmembrane metalloprotease which mediates the ectodomain shedding of a myriad of transmembrane proteins |
| ADGRG6 | ARID5B | COL11A1 | ADGRG6 | | G-protein coupled receptor activated by type IV collagen |
| ADGRL3 | ASB3 | DIDO1 | ADGRL3 | ADGRL3 | Latrophilin subfamily of G-protein coupled receptors |
| ADGRL4 | ASPH | ENDOD1 | ADGRL4 | ADGRL4 | " |
| ADIPOR2 | ATP4B | GLRX | ADIPOR2 | | Adiponectin receptor for ADIPOQ, an essential hormone secreted by adipocytes that regulates glucose and lipid metabolism |
| AEBP2 | ATP8A1 | GMPS | AEBP2 | | Acts as an accessory subunit for the core Polycomb repressive complex 2 (PRC2), which mediates histone H3K27 (H3K27me3) trimethylation on chromatin leading to transcriptional repression of the affected target gene |
| AGAP1 | AZIN1 | GPAT2 | AGAP1 | | GTPase-activating protein for ARF1 and ARF5 |
| AGR3 | BTRC | KRBOX1 | AGR3 | | Disulfide isomerase (PDI) family of endoplasmic reticulum (ER) proteins that catalyze protein folding and thiol-disulfide interchange reactions |
| AGTR1 | C4orf19 | LEPROT | AGTR1 | | Receptor for angiotensin II, a potent vasopressor hormone and a primary regulator of aldosterone secretion |
| AHCTF1 | CAMSAP1 | LINC00028 | AHCTF1 | | Involved in nuclear pore complex assembly and regulation of cytokinesis. |
| AHR | CCDC180 | LINC00342 | AHR | | ligand-activated helix-loop-helix transcription factor involved in the regulation of biological responses to planar aromatic hydrocarbons. |
| ALDH5A1 | CTSV | NKTR | ALDH5A1 | ALDH5A1 | Aldehyde dehydrogenase family |
| ALK | CUZD1 | PALLD | ALDH8A1 | ALDH8A1 | " |
| AMY2A | DIRAS2 | PCP4 | ALK | | receptor tyrosine kinase of the insulin receptor superfamily |
| AMY2B | EDEM3 | PCSK1 | AMY2A | AMY2A | Alpha-amylase family, catalyze starch catabolism |
| AMZ2 | ERGIC3 | PLCH1 | AMY2B | AMY2B | " |
| ANKS1B | FAM24B | SLC17A9 | AMZ2 | | Zinc metalloprotease that displays some activity against angiotensin-3 |
| ANO4 | FBX07 | TAF5L | ANKS1B | | Unknown |
| ANO9 | GPC5 | TEXT4 | ANO4 | ANO4 | Anotamin family (Ca2+-activated Cl− channel) |
| ANP32D | GYPC | USP13 | ANO9 | ANO9 | " |
| AOAH | ID2 | WNT2B | ANP32D | | Phosphoprotein 32 (PP32) |
| ARAP2 | IQCM | XKR4 | AOAH | | Catalyzes the hydrolysis of acyloxyacyl-linked fatty acyl chains from bacterial lipopolysaccharides, effectively detoxifying these molecules |
| ARHGAP21 | KAZALD1 | ZNF514 | AP1AR | | Enables AP-1 adaptor complex binding activity and kinesin binding activity |
| ARHGEF26 | KCTD8 | | AP2A2 | | Subunit of the AP-2 adaptor protein complex, which is involved in linking lipid and protein membrane components with the clathrin lattice |
| ARHGEF28 | KLF10 | | ARAP2 | | Phosphatidylinositol 3,4,5-trisphosphate-dependent GTPase-activating protein that modulates actin cytoskeleton remodeling by regulating ARF and RHO family members. |
| ARRDC3 | KLF6 | | ARHGAP21 | | Functions as a GTPase-activating protein (GAP) for RHOA and CDC42. |
| ARVCF | L1TD1 | | ARHGEF26 | ARHGEF26 | Rho-guanine nucleotide exchange factor (Rho-GEF) family |
| ASB3 | LTK | | ARHGEF28 | ARHGEF28 | " |
| ASB8 | MALRD1 | | ARID5B | | The encoded protein forms a histone H3K9Me2 demethylase complex with PHD finger protein 2 and regulates the transcription of target genes involved in adipogenesis and liver development |
| ASNS | MCTP2 | | ARRDC3 | | arrestin family of proteins, which regulate G protein-mediated signaling |
| ASXL1 | MEOX2 | | ARVCF | | Catenin family member that plays an important role in the formation of adherens junction complexes |
| ATP8B1 | MIS18BP1 | | ASB3 | ASB3 | Family of substrate-recognition components of a SCF-like ECS (Elongin-Cullin-SOCS-box protein) E3 ubiquitin-protein ligase complex |
| ATXN1 | MMS22L | | ASB8 | ASB8 | " |
| AVPR1A | MND1 | | ASNS | | Involved in the synthesis of asparagine |
| B4GALNT3 | MRPL39 | | ASPH | | Role in calcium homeostasis |
| B9D1 | MRPS2 | | ASXL1 | | Probable Polycomb group (PcG) protein involved in transcriptional regulation mediated by ligand-bound nuclear hormone receptors, such as retinoic acid receptors (RARs) and peroxisome proliferator-activated receptor gamma (PPARG) |
| BEND7 | MSRB2 | | ATP4B | | P-type cation-transporting ATPase |
| C1D | NCAM2 | | ATP8A1 | ATP8A1 | Catalytic component of a P4-ATPase flippase complex |
| C1orf21 | NEBL | | ATP8B1 | ATP8B1 | " |
| C6orf118 | NKAIN3 | | ATXN1 | | Chromatin-binding factor that repress Notch signaling in the absence of Notch intracellular domain by acting as a CBF1 corepressor. |
| CAV2 | NRC32 | | AVPR1A | | Receptor for arginine vasopressin |
| CBR4 | NRG3 | | AZIN1 | | Antizyme inhibitor family |
| CCDC184 | NRXN1 | | B4GALNT3 | | Catalyzes transfer of N-acetylgalactosamine (GalNAc) in N-linked glycans and probably O-linked glycans |
| CCDC77 | OTUD1 | | B9D1 | | Component of the tectonic-like complex in primary cilia, required for ciliogenesis and sonic hedgehog/SHH signaling |
| CCDC91 | PITRM1 | | BEND7 | | Transcription factor, unknown role |
| CCNB2 | PITX2 | | BTRC | | F-box protein, one of the four subunits of ubiquitin protein ligase complex SCF (SKP1-cullin-F-box), which functions in phosphorylation-dependent ubiquitination |
| CCNG1 | POLD4 | | C1D | | Nuclear Receptor Corepressor, binds RAC3, promotes apoptosis |
| CCR6 | POU3F2 | | C1orf21 | | Unknown |
| CCSER1 | RELL1 | | C4orf19 | | Unknown |
| CCSER2 | RGMA | | C6orf118 | | Unknown |
| CDC42BPA | RGMB | | CAMSAP1 | | Key microtubule-organizing protein that specifically binds the minus-end of non-centrosomal microtubules and regulates their dynamics and organization |
| CDCA2 | RGPD4 | | CAV2 | | Caveolin, a scaffolding protein within caveolar membranes |
| CDH2 | RIOK2 | | CBR4 | | Carbonyl Reductase, forms part of the mitochondrial fatty acid synthase |
| CDH9 | RND3 | | CCDC180 | | Coiled-Coil Domain-Containing Protein, unknown function |
| CDK5RAP3 | RNF144A | | CCDC184 | | Coiled-Coil Domain-Containing Protein, unknown function |
| CDON | RNF2 | | CCDC77 | | Coiled-Coil Domain-Containing Protein, unknown function |
| CELSR1 | RPL10L | | CCDC91 | | Coiled-Coil Domain-Containing Protein, involved in the regulation of membrane traffic through the trans-Golgi network |
| CFAP99 | RTCB | | CCNB2 | CCNB2 | Cyclin (cell cycle regulator) |
| CFTR | SALL2 | | CCNG1 | CCNG1 | " |
| CHAC1 | SGK1 | | CCR6 | | Beta chemokine receptor family, ligand is CCL20 |
| CHCHD7 | SIRPA | | CCSER1 | CCSER1 | Microtubule binding protein family, mediates bundle formation |
| CHD4 | SLITRK5 | | CCSER2 | CCSER2 | " |
| CLEC11A | SOSTDC1 | | CCZ1 | CCZ1 | Enables guanyl-nucleotide exchange factor activity, predicted to be involved in vesicle-mediated transport. |
| CMC1 | SOX17 | | CCZ1B | CCZ1B | " |
| CNTN4 | SPAG4 | | CDC42BPA | | Serine/threonine-protein kinase which is an important downstream effector of CDC42 and plays a role in the regulation of cytoskeleton reorganization and cell migratio |
| CNTN5 | SSH3 | | CDCA2 | | Targeting subunit of the cell-cycle associated protein, protein phosphatase 1, with a role in targeting this protein to chromatin during anaphase. |
| CNTN6 | ST6GAL2 | | CDH2 | CDH2 | Cadherin family (calcium-dependent cell adhesion protein) |
| CNTNAP2 | STK35 | | CDH9 | CDH9 | " |
| CNTNAP4 | TBL1XR1 | | CDK5RAP3 | | Substrate adapter of E3 ligase complexes mediating ufmylation, the covalent attachment of the ubiquitin-like modifier UFM1 to substrate proteins, and which is involved in various processes, such as ribosome recycling and reticulophagy (also called ER-phagy) |
| COL11A1 | TLR2 | | CDON | | Component of a cell-surface receptor complex that mediates cell-cell interactions between muscle precursor cells. |
| COPZ2 | TM2D1 | | CELSR1 | | cadherin superfamily, flamingo subfamily |
| CRTC1 | TMEM255B | | CFAP99 | | Cilia And Flagella Associated Protein |
| CSMD1 | TMEM26 | | CFTR | | Superfamily of ATP-binding cassette (ABC) transporters, functions as a chloride channel; mutations in this gene cause cystic fibrosis |
| CSMD3 | TSN | | CHAC1 | | Gamma-glutamylcyclotransferase family; deglycinates the Notch receptor, which prevents receptor maturation and inhibits Notch signaling |
| CTBP2 | TSPAN14 | | CHCHD7 | | Predicted to be involved in mitochondrial respiratory chain complex assembly |
| DCANP1 | TUBGCP3 | | CHD4 | | CHD family (modifies chromatin structure) |
| DCT | TYRO3 | | CLEC11A | | C-type lectin superfamily; functions as a growth factor for primitive hematopoietic progenitor cells |
| DIAPH3 | XKR4 | | CMC1 | | Component of the MITRAC (mitochondrial translation regulation assembly intermediate of cytochrome c oxidase complex) complex, that regulates cytochrome c oxidase assembly |
| DIRAS2 | ZFAT | | CNTN4 | CNTN4 | Contactin (axon-associated cell adhesion molecule) |
| DLC1 | ZMAT3 | | CNTN5 | CNTN5 | " |
| DLGAP2 | | | CNTN6 | CNTN6 | " |
| DLL4 | | | CNTNAP2 | CNTNAP2 | Contactin-associated protein family |
| DMRT2 | | | CNTNAP4 | CNTNAP4 | " |
| DNASE2B | | | COL11A1 | | One of the three collagen chains that form type XI collagen |
| DPP10 | | | COPZ2 | | subunit of the coatomer protein complex, a seven-subunit complex that functions in the formation of COPI-type, non-clathrin-coated vesicles. COPI vesicles function in the retrograde Golgi-to-ER transport of dilysine-tagged proteins |
| DSCAM | | | CRTC1 | | CREB-regulated transcription coactivator protein; involved in energy metabolism |
| DSG1 | | | CSMD1 | CSMD1 | CUB And Sushi Multiple Domains 1 family, complement cascade inhibitor, epithelial-mesenchymal transition |
| DSG3 | | | CSMD3 | CSMD3 | " |
| DYNC2H1 | | | CTBP2 | | Corepressor targeting diverse transcription regulators. Functions in brown adipose tissue (BAT) differentiation |
| DYRK1A | | | CTSV | | Cathepsin V, a lysosomal cysteine proteinase |
| E2F3 | | | CUZD1 | | Localized to zymogen granules, where it functions in trypsinogen activation |
| EBF2 | | | DCANP1 | | Binds with and transactivates the corticotropin-releasing hormone (CRH) promoter |
| EFHB | | | DCT | | Dopachrome Tautomerase, role in melanin biosynthesis, also involved in energy metabolism, particularly related to adipose tissue function and lipid metabolism |
| EIF2AK4 | | | DIAPH3 | | Diaphanous subfamily of the formin family; actin nucleation and elongation factor required for the assembly of F-actin structures, such as actin cables and stress fibers |
| EIF3IP1 | | | DIDO1 | | Death inducer-obliterator-1 gene, upregulated by apoptotic signals |
| EPHB1 | | | DIRAS2 | | DIRAS subfamily of small GTPases |
| ESD | | | DLC1 | | Rho GTPase Activating Protein |
| ETAA1 | | | DLGAP2 | | May play a role in the molecular organization of synapses and neuronal cell signaling. |
| ETNK1 | | | DLL4 | | Ligand in Notch signaling, Delta subfamily, characterized by a DSL domain, EGF repeats, and a transmembrane domain |
| EVX2 | | | DMRT2 | | DMRT family transcriptional activator that directly regulates early activation of the myogenic determination |
| FAM155A | | | DNASE2B | | Hydrolyzes DNA under acidic conditions |
| FAM76B | | | DPP10 | | S9B family in clan SC of the serine proteases, promotes cell surface expression of the potassium channel KCND2 and modulates its activity and gating characteristics |
| FAXDC2 | | | DSCAM | | immunoglobulin superfamily of cell adhesion molecules, role in neuronal self-avoidance |
| FBXO11 | | | DSG1 | DSG1 | Desmoglein protein subfamily, components of desmosome |
| FERMT1 | | | DSG3 | DSG3 | " |
| FFAR2 | | | DYNC2H1 | | Dynein, may function as a motor for intraflagellar retrograde transport. Functions in cilia biogenesis. |
| FMN1 | | | DYRK1A | | Dual-specificity tyrosine phosphorylation-regulated kinase (DYRK) family |
| FOXN2 | | | E2F3 | | Transcription factor, inhibits adipogenesis |
| FRMD3 | | | EBF2 | | Transcription factor that acts in synergy with the Wnt-responsive LEF1/CTNNB1 pathway |
| FST | | | EDEM3 | | Involved in endoplasmic reticulum-associated degradation (ERAD), a quality control mechanism |
| FSTL5 | | | EFHB | | Microtubule inner protein (MIP) part of the dynein-decorated doublet microtubules (DMTs) in cilia axoneme; involved in regulation of calcineurin-NFAT signaling cascade and regulation of store-operated calcium entry |
| GABRG2 | | | EIF2AK4 | | Member of a family of kinases that phosphorylate the alpha subunit of eukaryotic translation initiation factor-2 (EIF2), downregulating protein synthesis, involved in the integrated stress response |
| GABRP | | | EIF3IP1 | | Eukaryotic Translation Initiation Factor 3 Subunit I Pseudogene 1 |
| GAL | | | ENDOD1 | | Plays a role in the modulation of innate immune signaling through the cGAS-STING pathway by interacting with RNF26 |
| GALNT10 | | | EPHB1 | | Receptor tyrosine kinase which binds promiscuously transmembrane ephrin-B family ligands residing on adjacent cells, leading to contact-dependent bidirectional signaling into neighboring cells. |
| GAS2L3 | | | ERGIC3 | | Involved in endoplasmic reticulum to Golgi vesicle-mediated transport and positive regulation of intracellular protein transport. |
| GATAD2A | | | ESD | | Serine hydrolase that belongs to the esterase D family, may be involved in the recycling of sialic acids |

| | | | |
|---|---|---|---|
| GMNC | ETAA1 | | Replication stress response protein that accumulates at DNA damage sites and promotes replication fork progression and integrity |
| GOT2 | ETNK1 | | An ethanolamine kinase, may be a rate-controlling step in phosphatidylethanolamine biosynthesis |
| GPC6 | EVX2 | | Homeobox transcription factor |
| GPLD1 | FAM155A | | Also known as NALF1; auxillary component of the NALCN sodium channel complex, a channel that regulates the resting membrane potential and controls neuronal excitability |
| GTF2B | FAM24B | | Unknown |
| HAAO | FAM76B | | Gene Ontology (GO) annotations related to this gene include deNEDDylase activity |
| HAS2 | FAXDC2 | | Promotes megakaryocyte differentiation |
| HCK | FBXO11 | FBXO11 | Substrate recognition component of a SCF (SKP1-CUL1-F-box protein) E3 ubiquitin-protein ligase complex |
| HDAC4 | FBXO7 | FBXO7 | " |
| HDAC9 | FERMT1 | | Fermitin family member, involved in integrin signaling and linkage of the actin cytoskeleton to the extracellular matrix |
| HMCN1 | FFAR2 | | GP40 family of G protein-coupled receptors, regulates whole-body energy homeostasis, adipogenesis |
| HOXD13 | FMN1 | | Formin homology family, plays a role in the formation of adherens junction and the polymerization of linear actin cables |
| HS3ST1 | FOXN2 | | Transcription factor |
| HS3ST4 | FRMD3 | | Unknown |
| HS6ST1 | FST | FST | TGFB superfamily member follistatin (activin antagonist) |
| HTR2A | FSTL5 | FSTL5 | " |
| HYLS1 | GABRG2 | GABRG2 | Subunit of the GABA-A receptor, a major inhibitory neurotransmitter in the brain |
| IDUA | GABRP | GABRP | " |
| IGF2BP3 | GAL | | Neuroendocrine hormone of the central and peripheral nervous systems, role in contraction of smooth muscle |
| INTU | GALNT10 | | Catalyzes the first step in the synthesis of mucin-type oligosaccharides |
| IQCJ-SCHIP1 | GAS2L3 | | Cytoskeletal linker protein, may promote and stabilize the formation of the actin and microtubule network |
| ITPR2 | GATAD2A | | Transcriptional repressor, acts as a component of the histone deacetylase NuRD complex which participates in the remodeling of chromatin |
| JADE1 | GLRX | | Glutaredoxin family, antioxidant defense system |
| JAG2 | GMNC | | Regulator of DNA replication |
| JDP2 | GMPS | | Catalyzes the conversion of xanthine monophosphate (XMP) to GMP |
| JMJD7 | GOT2 | | Glutamic-oxaloacetic transaminase, role in the intracellular NAD(H) redox balance, facilitates cellular uptake of long-chain free fatty acids |
| JMJD7-PLA2G4B | GPAT2 | | Glycerol-3-phosphate O-acyltransferase |
| KCND3 | GPC5 | GPC5 | Glypican family, act as co-receptors on the cell surface, playing crucial roles in regulating various signaling pathways by modifying the availability of growth factors through their heparan sulfate chain |
| KCND3 | GPC6 | GPC6 | " |
| KCNE4 | GPLD1 | | Glycosylphosphatidylinositol (GPI) anchor-degrading enzyme |
| KCNH8 | GTF2B | | General transcription factor IIB, one of the ubiquitous factors required for transcription initiation by RNA polymerase II |
| KCNIP1 | GYPC | | Integral membrane glycoprotein, regulates the mechanical stability of red cells |
| KCNIP3 | HAAO | | Catalyzes the synthesis of quinolinic acid (QUIN) from 3-hydroxyanthranilic acid, increased cerebral levels of QUIN may participate in the pathogenesis of neurologic and inflammatory disorders |
| KCNIP4 | HAS2 | | Hyaluronic acid synthetase |
| KIAA1614 | HCK | | Src family of tyrosine kinases, regulates innate immune responses |
| KIF2B | HDAC4 | HDAC4 | Class IIa histone deacetylase |
| KIF3B | HDAC9 | HDAC9 | " |
| KLF12 | HMCN1 | | Hemicentin, multifunctional |
| KLF5 | HOXD13 | | Homeobox family transcription factors |
| KLHDC7A | HS3ST1 | HS3ST1 | Heparan sulfate biosynthetic enzyme family |
| KLHDC8A | HS3ST4 | HS3ST4 | " |
| LARGE1 | HS6ST1 | HS6ST1 | " |
| LCORL | HTR2A | | Seratonin receptor; stimulating these receptors increases IOP by increasing blood flow to the ciliary body, which increases aqueous humor production |
| LEMD1 | HYLS1 | | Centriolar and ciliogenesis associated protein |
| LEPR | ID2 | | Transcriptional regulator, regulates the circadian clock, contributes to the regulation of a variety of liver clock-controlled genes involved in lipid metabolism |
| LINGO2 | IDUA | | Enzyme that catalyzes hydrolysis of the terminal alpha-L-iduronic acid residues of two glycosaminoglycans, dermatan sulfate and heparan sulfate, required for their degradation |
| LIPC | IGF2BP3 | | Binds to the 5' UTR of the insulin-like growth factor II leader 3 mRNA and may repress translation of insulin-like growth factor II |
| LOC100130987 | INTU | | Plays a key role in ciliogenesis |
| LPAR3 | IQCJ-SCHIP1 | | Read-through protein; may play a role in action potential conduction in myelinated cells through the organization of molecular complexes at nodes of Ranvier and axon initial segments |
| LRP2 | IQCM | | Unknown |
| LUZP1 | ITPR2 | | Inositol 1,4,5-triphosphate receptor family, whose members are second messenger intracellular calcium release channels |
| M1AP | JADE1 | | Enables transcription coactivator activity. Contributes to histone acetyltransferase activity. Involved in several processes, including negative regulation of canonical Wnt signaling pathway |
| MACROD2 | JAG2 | | Notch ligand of the serrate family |
| MACROH2A1 | JDP2 | | Component of the AP-1 transcription factor that represses transactivation mediated by the Jun family of protein |
| MAL | JMJD7 | | Endopeptidase that cleaves histones N-terminal tails at the carboxyl side of methylated arginine or lysine residues, to generate 'tailless nucleosomes', which may trigger transcription elongation |
| MAML3 | JMJD7-PLA2G4B | | Read-through protein; unknown function |
| MAPK10 | KAZALD1 | | Secreted member of the insulin growth factor-binding protein (IGFBP) superfamily |
| MAPKBP1 | KCND3 | KCND3 | Voltage-gated potassium (Kv) channel |
| MAT2B | KCNE4 | KCNE4 | " |
| MFSD1 | KCNH8 | KCNH8 | " |
| MGA | KCNIP1 | KCNIP1 | Voltage-gated potassium (Kv) channel interacting protein |
| MMRN1 | KCNIP3 | KCNIP3 | " |
| MOGAT1 | KCNIP4 | KCNIP4 | " |
| MOGS | KCTD8 | | Auxiliary subunit of GABA-B receptors that determine the pharmacology and kinetics of the receptor response |
| MOS | KIAA1614 | | Predicted to be involved in centrosome cycle; establishment or maintenance of cell polarity; and regulation of cellular localization |
| MRPS5 | KIF2B | KIF2B | Kinesin family (plus end microtubule-dependent motor) |
| MTX2 | KIF3B | KIF3B | " |
| MTX2 | KLF10 | KLF10 | Kruppel-like factor subfamily of zinc finger proteins (transcriptional regulators) |
| MUC12 | KLF12 | KLF12 | " |
| MVB12B | KLF5 | KLF5 | " |
| MYO1E | KLF6 | KLF6 | " |
| NAALADL2 | KLHDC7A | KLHDC7A | Kelch domain-containing family, unknown function |
| NALCN | KLHDC8A | KLHDC8A | " |
| NAT1 | KRBOX1 | | Predicted to be involved in regulation of DNA-templated transcription |
| NBAS | L1TD1 | | Predicted to enable single-stranded RNA binding activity. Predicted to be involved in retrotransposition |
| NEDD4L | LARGE1 | | N-acetylglucosaminyltransferase gene family, participates in glycosylation of alpha-dystroglycan, and may carry out the synthesis of glycoprotein and glycosphingolipid sugar chains |
| NOL4L | LCORL | | Ligand dependent nuclear receptor corepressor |
| NR2F1 | LEMD1 | | Unknown |
| NSG1 | LEPR | | Receptor for leptin, an adipocyte-specific hormone that regulates body weight, and is involved in the regulation of fat metabolism |
| NT5C2 | LEPROT | | Negatively regulates leptin receptor (LEPR) cell surface expression, and thus decreases response to leptin. |
| NUDCD2 | LINC00028 | | Unknown |
| NUDT7 | LINC00342 | | Unknown |
| OLFM3 | LINGO2 | | Predicted to act upstream of or within positive regulation of synapse assembly. |
| PC | LIPC | | Catalyzes the hydrolysis of triglycerides and phospholipids |
| PCDH17 | LOC100130987 | | Unknown |
| PCDH20 | LPAR3 | | Receptor for lysophosphatidic acid (LPA), a mediator of diverse cellular activities |
| PCP4 | LRP2 | | Megalin, a multi-ligand endocytic receptor |
| PDCD6IP | LTK | | Ros/insulin receptor family of tyrosine kinases |
| PDE3A | LUZP1 | | Unknown |
| PEX5L | M1AP | | Required for meiosis I progression during spermatogenesis |
| PHYHIP | MACROD2 | | Chromatin modifier; deacetylates O-acetyl-ADP ribose, a signaling molecule generated by the deacetylation of acetylated lysine residues in histones and other proteins |
| PKNOX2 | MACROH2A1 | | Chromatin modifier; variant histone H2A which replaces conventional H2A in a subset of nucleosomes where it represses transcription |
| PLAG1 | MAL | | Integral membrane protein, could be an important component in vesicular trafficking cycling between the Golgi complex and the apical plasma membrane. Could be involved in myelin biogenesis and/or myelin function |
| PLAGL2 | MALRD1 | | Enhances production and/or transport of FGF19 and thus has a role in regulation of bile acid synthesis |
| PLCH1 | MAML3 | | Mastermind Like Transcriptional Coactivator, involved in Notch signaling pathway and positive regulation of transcription by RNA polymerase II |
| PLPPR1 | MAPK10 | | MAP kinase family of signal transduction proteins |
| PLPPR5 | MAPKBP1 | | MAP kinase signaling, scaffold protein |
| POFUT1 | MAT2B | | Regulatory subunit of S-adenosylmethionine synthetase |
| PPARGC1A | MCTP2 | | Predicted to be involved in regulation of neurotransmitter secretion |
| PPM1H | MEOX2 | | Homeobox transcription factor, acts as a negative regulator of angiogenesis |
| PPP1R21 | MFSD1 | | Lysosomal dipeptide uniporter |
| PPP2R3A | MGA | | Functions as a dual-specificity transcription factor, regulating the expression of both MAX-network and T-box family target genes. Functions as a repressor or an activator. |
| PPP3CC | MIS18BP1 | | Required for recruitment of CENPA to centromeres and normal chromosome segregation during mitosis |
| PRDM10 | MMRN1 | | Multimerin, a massive, soluble protein found in platelets and in the endothelium of blood vessels; carrier protein for platelet (but not plasma) factor V/Va |
| PRDM15 | MMS22L | | Forms a complex with tonsoku-like, DNA repair protein (TONSL), and this complex recognizes and repairs DNA double-strand breaks at sites of stalled or collapsed replication forks |
| PRKD1 | MND1 | | Required for proper homologous chromosome pairing and efficient cross-over and intragenic recombination during meiosis |
| PTHLH | MOGAT1 | | Acyl-CoA:monoacylglycerol acyltransferase; involved in glycerolipid synthesis and lipid metabolism |
| PTPN7 | MOGS | | This gene encodes the first enzyme in the N-linked oligosaccharide processing pathway |
| PUM3 | MOS | | Serine/threonine kinase that activates the MAP kinase cascade through direct phosphorylation of the MAP kinase activator MEK |
| RAC1 | MRPL39 | MRPL39 | Mitochondrial ribosomal protein |
| RASSF3 | MRPS2 | MRPS2 | " |
| RBFOX1 | MRPS5 | MRPS5 | " |
| RBFOX3 | MSRB2 | | Methionine-sulfoxide reductase; upon oxidative stress, may play a role in the preservation of mitochondrial integrity |
| RCBTB1 | MTX2 | | Involved in transport of proteins into the mitochondrion |
| RHOBTB3 | MUC12 | | Membrane-associated mucin |
| RIBC2 | MVB12B | | Component of the ESCRT-I complex, a regulator of vesicular trafficking process. |
| RNF111 | MYO1E | | Nonmuscle class I myosin of the unconventional myosin protein family, actin-based molecular motors; involved in clathrin-mediated endocytosis |
| RTKN | NAALADL2 | | N-acetylated alpha-linked acidic dipeptidase (NAALADase) gene family |
| SASH1 | NALCN | | Voltage-independent, nonselective cation channel which belongs to a family that regulates the resting membrane potential and excitability of neurons |
| SCHIP1 | NAT1 | | One of two arylamine N-acetyltransferase (NAT) genes in the human genome, participates in the detoxification of a plethora of hydrazine and arylamine drugs |
| SDR16C5 | NBAS | | Involved in Golgi-to-endoplasmic reticulum (ER) retrograde transport |
| SEMA5B | NCAM2 | | Neural cell adhesion molecule |
| SEPHS1 | NEBL | | Binds to actin and plays an important role in the assembly of the Z-disk in cardiac muscle |
| SGCG | NEDD4L | | Member of Nedd4 family of HECT domain E3 ubiquitin ligases |
| SGPP1 | NIFK | | May bind RNA and may play a role in mitosis and cell cycle progression |
| SH3RF1 | NKAIN3 | | Interacts with the beta subunit of Na,K-ATPase |
| SHOX2 | NKTR | | PPIase that catalyzes the cis-trans isomerization of proline imidic peptide bonds in oligopeptides and may therefore assist protein folding |
| SLC17A9 | NOL4L | | Unknown |
| SLC25A26 | NR2F1 | NR2F1 | Nuclear hormone receptor and transcriptional regulator (Coup transcription factor) |
| SLC4A5 | NRC32 | NRC32 | " (mineralocorticoid receptor) |
| SLC5A3 | NRG3 | | Neuregulin gene family, direct ligand for the ERBB4 tyrosine kinase receptor |
| SLC7A11 | NRXN1 | | Neurexin-1, binds neuroligins |
| SLIT2 | NSG1 | | Role in the recycling mechanism in neurons of multiple receptors, acts at the level of early endosomes to promote sorting of receptors toward a recycling pathway |
| SLITRK3 | NT5C2 | | Hydrolase that serves as an important role in cellular purine metabolism |

| | | | |
|---|---|---|---|
| SLTM | NUDCD2 | | May regulate the LIS1/dynein pathway by stabilizing LIS1 with Hsp90 chaperone |
| SMARCA2 | NUDT7 | | Nudix hydrolase involved in eliminating potentially toxic nucleotide metabolites from the cell |
| SPATS2 | OLFM3 | | Olfactomedin Related ER Localized Protein |
| SPRED2 | OTUD1 | | Deubiquitinating enzyme |
| SPTY2D1 | PALLD | | Cytoskeletal protein required for organizing the actin cytoskeleton |
| SRRT | PC | | Pyruvate carboxylase |
| SSX2IP | PCDH17 | PCDH17 | Protocadherin gene family, a subfamily of the cadherin superfamily |
| STAG1 | PCDH20 | PCDH20 | " |
| STEAP2 | PCP4 | | Modulator of calcium-binding by calmodulin |
| STON2 | PCSK1 | | Subtilisin-like proprotein convertase family, process protein and peptide precursors trafficking through regulated or constitutive branches of the secretory pathway |
| STX6 | PDCD6IP | | Multifunctional protein involved in endocytosis, multivesicular body biogenesis, membrane repair, cytokinesis, apoptosis and maintenance of tight junction integrity |
| SWT1 | PDE3A | | Member of the cGMP-inhibited cyclic nucleotide phosphodiesterase (cGI-PDE) family |
| SYK | PEX5L | | Accessory subunit of hyperpolarization-activated cyclic nucleotide-gated (HCN) channels, regulating their cell-surface expression and cyclic nucleotide dependence |
| TAC1 | PHYHIP | | Interacts with PHYH, a peroxisomal protein involved in the alpha-oxidation of 3-methyl branched fatty acids |
| TBC1D12 | PITRM1 | | ATP-dependent metalloprotease that degrades post-cleavage mitochondrial transit peptides |
| TBK1 | PITX2 | | RIEG/PITX homeobox family of transcription factors |
| TBX18 | PKNOX2 | | Homeodomain transcription factor |
| TFDP2 | PLAG1 | PLAG1 | Transcription factor |
| TFEC | PLAGL2 | PLAGL2 | " |
| TFPI | PLCH1 | | PLC-eta family of the phosphoinositide-specific phospholipase C |
| THSD7A | PLPPR1 | PLPPR1 | Plasticity-related gene (PRG) family, mediates lipid phosphate phosphatase activity in neurons |
| TLE1 | PLPPR5 | PLPPR5 | " |
| TM9SF4 | POFUT1 | | Enzyme that O-fucosylates EGF-like repeats, including in the Notch protein; essential for Notch activity |
| TMEM108 | POLD4 | | Component of the tetrameric DNA polymerase delta complex (Pol-delta4), plays a role in high fidelity genome replication and repair |
| TMEM201 | POU3F2 | | POU-III class of neural transcription factors. The encoded protein is involved in neuronal differentiation and enhances the activation of corticotropin-releasing hormone regulated genes |
| TMEM86A | PPARGC1A | | transcriptional coactivator that regulates the genes involved in energy metabolism |
| TMX3 | PPM1H | | Enables identical protein binding activity and phosphoprotein phosphatase activity |
| TNFRSF13B | PPP1R21 | | Component of the FERRY complex (Five-subunit Endosomal Rab5 and RNA/ribosome intermediary), functions as a RAB5A effector involved in the localization and the distribution of specific mRNAs most likely by mediating their endosomal transport. |
| TNK2 | PPP2R3A | | Regulatory subunits of the protein phosphatase 2 |
| TNRC6B | PPP3CC | | Calcineurin, a calcium-dependent, calmodulin-stimulated protein phosphatase involved in the downstream regulation of dopaminergic signal transduction |
| TOM1L1 | PRDM10 | PRDM10 | Transcription factor that contains C2H2-type zinc-finger, unknown role |
| TRIB2 | PRDM15 | PRDM15 | " |
| TRIM56 | PRKD1 | | Serine/threonine-protein kinase that converts transient diacylglycerol (DAG) signals into prolonged physiological effects downstream of PKC |
| TRPS1 | PTHLH | | Parathyroid hormone-like hormone, a neuroendocrine peptide |
| TSC22D2 | PTPN7 | | protein tyrosine phosphatase (PTP) family. PTPs are known to be signaling molecules that regulate a variety of cellular processes including cell growth, differentiation, mitotic cycle, and oncogenic transformation |
| TTF1 | PUM3 | | Inhibits the poly(ADP-ribosyl)ation activity of PARP1 and the degradation of PARP1 by CASP3 following genotoxic stress |
| URB2 | RAC1 | | Rac family GTPase |
| USP25 | RASSF3 | | A member of a subfamily of the RAS superfamily, plasma membrane GTP-binding proteins that modulate intracellular signal transduction pathways. |
| VDR | RBFOX1 | RBFOX1 | Fox-1 family of RNA-binding proteins, regulate tissue-specific alternative splicing |
| VTA1 | RBFOX3 | RBFOX3 | " |
| VWC2 | RCBTB1 | | Encodes a protein that regulates angiogenesis; plays a role in maintaining healthy blood vessel development in the eye |
| WDR89 | RELL1 | | Involved in positive regulation of p38MAPK cascade |
| WNT5A | RGMA | RGMA | Member of the repulsive guidance molecule (RGM) family |
| WWC1 | RGMB | RGMB | " |
| XPNPEP1 | RGPD4 | | Predicted to contribute to GTPase activator activity. Predicted to be involved in NLS-bearing protein import into nucleus. Predicted to be part of nuclear pore. |
| ZBTB47 | RHOBTB3 | | Rab9-regulated ATPase required for endosome to Golgi transport. |
| ZBTB49 | RIBC2 | | Predicted to be involved in flagellated sperm motility. Located in axonemal microtubule. |
| ZKSCAN2 | RIOK2 | | Serine/threonine-protein kinase involved in the final steps of cytoplasmic maturation of the 40S ribosomal subunit. |
| ZNF536 | RND3 | | Rho Family GTPase |
| ZNF641 | RNF111 | RNF111 | RING finger domain-containing E3 ubiquitin ligase family, modifies transcription |
| ZNF728 | RNF144A | RNF144A | " |
| ZNF737 | RNF2 | RNF2 | " |
| ZNF804B | RPL10L | | Ribosome protein L10 |
| | RTCB | | Catalytic subunit of the tRNA-splicing ligase complex |
| | RTKN | | Scaffold protein that interacts with GTP-bound Rho proteins to inhibit their GTPase activity |
| | SALL2 | | Transcription factor that plays a role in eye development |
| | SASH1 | | Scaffold protein involved in the TLR4 signaling pathway |
| | SCHIP1 | | Predicted to be involved in positive regulation of hippo signaling. |
| | SDR16C5 | | Short-chain alcohol dehydrogenase/reductase superfamily of proteins and is involved in the oxidation of retinol to retinaldehyde |
| | SEMA5B | | Semaphorin protein family which regulates axon growth during development of the nervous system |
| | SEPHS1 | | Enzyme that synthesizes selenophosphate from selenide and ATP |
| | SGCG | | Component of the sarcoglycan complex, a subcomplex of the dystrophin-glycoprotein complex which forms a link between the F-actin cytoskeleton and the extracellular matrix of muscle |
| | SGK1 | | Serum/Glucocorticoid Regulated Kinase, plays an important role in cellular stress response |
| | SGPP1 | | Sphingosine-1-phosphate (S1P) is a bioactive sphingolipid metabolite that regulates diverse biologic processes; SGPP1 catalyzes the degradation of S1P via salvage and recycling of sphingosine into long-chain ceramides |
| | SH3RF1 | | Ubiquitin-protein ligase involved in protein sorting at the trans-Golgi network |
| | SHOX2 | | Homeodomain transcription factor |
| | SIRPA | | Signal-regulatory-protein (SIRP) family, multifunctional |
| | SLC17A9 | | Participates in the vesicular uptake, storage, and secretion of adenoside triphosphate (ATP) and other nucleotides |
| | SLC25A26 | | Transport of S-adenosylmethionine (SAM) into the mitochondria |
| | SLC4A5 | | Mediates sodium- and bicarbonate-dependent electrogenic sodium bicarbonate cotransport |
| | SLC5A3 | | Electrogenic Na(+)-coupled sugar symporter that actively transports myo-inositol and its stereoisomer scyllo-inositol across the plasma membrane |
| | SLC7A11 | | Heterodimer with SLC3A2, that functions as an antiporter by mediating the exchange of extracellular anionic L-cystine and intracellular L-glutamate across the cellular plasma membrane |
| | SLIT2 | | Slit family of secreted glycoproteins, roles in axon guidance and neuronal migration, ligand of ROBO receptors |
| | SLITRK3 | SLITRK3 | SLITRK family of structurally related transmembrane proteins that are involved in controlling neurite outgrowth |
| | SLITRK5 | SLITRK5 | " |
| | SLTM | | When overexpressed, acts as a general inhibitor of transcription that eventually leads to apoptosis |
| | SMARCA2 | | SWI/SNF family of proteins with helicase and ATPase activities and are thought to regulate transcription of certain genes by altering the chromatin structure around those genes |
| | SOSTDC1 | | Sclerostin family member, functions as a bone morphogenetic protein (BMP) antagonist |
| | SOX17 | | Transcription factor that Inhibits Wnt signaling, activates Notch signaling |
| | SPAG4 | | Involved in spermatogenesis |
| | SPATS2 | | Unknown |
| | SPRED2 | | Negatively regulates Ras signaling pathways and downstream activation of MAP kinases |
| | SPTY2D1 | | Histone chaperone |
| | SRRT | | Acts as a mediator between the cap-binding complex (CBC) and the primary microRNAs (miRNAs) processing machinery during cell proliferation |
| | SSH3 | | Protein phosphatase which may play a role in the regulation of actin filament dynamics |
| | SSX2IP | | Belongs to an adhesion system, which plays a role in the organization of homotypic, interneuronal and heterotypic cell-cell adherens junctions (AJs). |
| | ST6GAL2 | | Sialyltransferase, transfers sialic acid from the donor of substrate CMP-sialic acid to galactose containing acceptor substrate |
| | STAG1 | | Component of cohesin complex, a complex required for the cohesion of sister chromatids after DNA replication |
| | STEAP2 | | Integral membrane protein that functions as a NADPH-dependent ferric-chelate reductase |
| | STK35 | | Serine/threonine protein kinase |
| | STON2 | | Adapter protein involved in endocytic machinery |
| | STX6 | | Targets endosomes to the trans-Golgi network, and may therefore function in retrograde trafficking |
| | SWT1 | | Transcription factor |
| | SYK | | Non-receptor type Tyr protein kinase |
| | TAC1 | | Encodes four products of the tachykinin peptide hormone family, substance P and neurokinin A, as well as the related peptides, neuropeptide K and neuropeptide gamma |
| | TAF5L | | Functions as a component of the PCAF complex, capable of efficiently acetylating histones in a nucleosomal context |
| | TBC1D12 | | RAB11A-binding protein that plays a role in neurite outgrowth; enables GTPase activity |
| | TBK1 | | Serine/threonine kinase that plays an essential role in regulating inflammatory responses to foreign agents |
| | TBL1XR1 | | F-box-like protein involved in the recruitment of the ubiquitin/19S proteasome complex to nuclear receptor-regulated transcription units; plays an essential role in transcription activation mediated by nuclear receptors. |
| | TBX18 | | Tbx1 sub-family, acts as a transcriptional repressor by antagonizing transcriptional activators in the T-box family |
| | TEKT4 | | Microtubule inner protein (MIP) part of the dynein-decorated doublet microtubules (DMTs) in cilia and flagellar axoneme |
| | TFDP2 | | Binds DNA cooperatively with E2F family members to stimulate transcription |
| | TFEC | | Microphthalmia (MiT) family of basic helix-loop-helix leucine zipper transcription factors |
| | TFPI | | Kunitz-type serine protease inhibitor that regulates the tissue factor (TF)-dependent pathway of blood coagulation |
| | THSD7A | | Plays a role in actin cytoskeleton rearrangement; soluble form promotes endothelial cell migration and filopodia formation during sprouting angiogenesis |
| | TLE1 | | Transcriptional corepressor involved in WNT and Notch signaling |
| | TLR2 | | Toll-like receptor (TLR) family, role in pathogen recognition and activation of innate immunity |
| | TM2D1 | | May participate in amyloid-beta-induced apoptosis via its interaction with beta-APP42 |
| | TM9SF4 | | Multifunctional protein located in Golgi apparatus and early endosome |
| | TMEM108 | | Multifunctional protein located in endosomes |
| | TMEM201 | | Proposed to be involved in actin-dependent nuclear movement via association with transmembrane actin-associated nuclear (TAN) lines which are bound to F-actin cables and couple the nucleus to retrograde actin flow |
| | TMEM255B | | Unknown |
| | TMEM26 | | Selective surface protein marker of beige adipocytes, which may coexist with classical brown adipocytes in brown adipose tissue |
| | TMEM86A | | Enzyme involved in lysoplasmalogen metabolism in the adipocyte tissue and macrophages |
| | TMX3 | | Disulfide isomerase (PDI) family of endoplasmic reticulum (ER) proteins that catalyze protein folding and thiol-disulfide interchange reactions |
| | TNFRSF13B | | Tumor necrosis factor (TNF) receptor superfamily; mediates calcineurin-dependent activation of NF-AT, as well as activation of NF-kappa-B and AP-1; involved in the stimulation of B- and T-cell function and the regulation of humoral immunity |
| | TNK2 | | Non-receptor tyrosine-protein and serine/threonine-protein kinase that is implicated in cell spreading and migration, cell survival, cell growth and proliferatio |
| | TNRC6B | | Plays a role in RNA-mediated gene silencing by both micro-RNAs (miRNAs) and short interfering RNAs (siRNAs) |
| | TOM1L1 | | Probable adapter protein involved in signaling pathways. Interacts with the SH2 and SH3 domains of various signaling proteins when it is phosphorylated |
| | TRIB2 | | Tribbles family, interacts with MAPK kinases and regulates activation of MAP kinases |
| | TRIM56 | | Tripartite motif (TRIM) family, E3 ubiquitin-protein ligase |
| | TRPS1 | | Transcriptional repressor. Binds specifically to GATA sequences and represses expression of GATA-regulated genes |
| | TSC22D2 | | Involved in negative regulation of cell cycle |
| | TSN | | DNA-binding protein that specifically recognizes consensus sequences at the breakpoint junctions in chromosomal translocation |
| | TSPAN14 | | Required for ADAM10 exit from the endoplasmic reticulum and for enzymatic maturation and trafficking to the cell surface as well as substrate specificity; negatively regulates ADAM10-mediated cleavage of GP6 |
| | TTF1 | | Transcription termination factor that is localized to the nucleolus and plays a critical role in ribosomal gene transcription |
| | TUBGCP3 | | Gamma-tubulin complex necessary for microtubule nucleation at the centrosome |
| | TYRO3 | | Receptor tyrosine kinase that transduces signals from the extracellular matrix into the cytoplasm by binding to several ligands including TULP1 or GAS6 |
| | URB2 | | Involved in regulation of signal transduction by p53 class mediator and ribosome biogenesis |
| | USP13 | USP13 | De-ubiquitinase |
| | USP25 | USP25 | " |
| | VDR | | Vitamin D3 receptor, a member of the nuclear hormone receptor superfamily |
| | VTA1 | | Involved in trafficking of the multivesicular body |
| | VWC2 | | BMP antagonist |

| | | |
|---|---|---|
| WDR89 | | Unknown |
| WNT2B | WNT2B | Wingless-type MMTV integration site (WNT) family of highly conserved, secreted signaling factors, function in the Wnt signaling pathway |
| WNT5A | WNT5A | " |
| WWC1 | | Acts as a scaffolding protein, facilitating protein-protein interactions within the Hippo pathway, regulated by Notch |
| XKR4 | | Phospholipid scramblase that promotes phosphatidylserine exposure on apoptotic cell surface |
| XPNPEP1 | | Metalloaminopeptidase that plays a role in degradation and maturation of tachykinins, neuropeptides, and peptide hormones |
| ZBTB47 | ZBTB47 | Zinc Finger And BTB Domain Containing transcription factor |
| ZBTB49 | ZBTB49 | " |
| ZFAT | | Transcriptional regulator involved in apoptosis and cell survival |
| ZKSCAN2 | | Transcriptional regulator |
| ZMAT3 | | Target gene of p53/TP53 |
| ZNF514 | | Zinc finger family of transcriptional regulators, unknown role |
| ZNF536 | | Zinc finger family of transcriptional regulators, transcriptional repressor that negatively regulates neuron differentiation by repressing retinoic acid-induced gene transcription |
| ZNF641 | | Zinc finger family of transcriptional regulators, unknown role |
| ZNF728 | | Zinc finger family of transcriptional regulators, unknown role |
| ZNF737 | | Zinc finger family of transcriptional regulators, unknown role |
| ZNF804B | | Zinc finger family of transcriptional regulators, unknown role |

408 total genes

41 paralogous groups
