## Supplementary material for "Pharmacogenomics of steroid-induced ocular hypertension: relationship to high-tension glaucomas and new pathophysiologic insight": Suppl Table S5

### Supplementary Table S5. Prioritized Target Gene Validation
### GC-regulation of prioritized target genes

**NOTES**

Alphabetical ordering of the full prioritized gene list highlights functionally similar paralogues or gene family members by grouping them together.
We counted 41 such paralogous groups with shared function.

**HEADERS**

AOP: aqueous outflow pathway; DEG: differentially expressed gene;
eQTL: expression quantitative trait locus; TM: trabecular meshwork
DEGs in TM cells: column adding up all glucocorticoid-regulated genes in TM cell dataset;
Count GC-DEG: column adding up all such genes, total at bottom;
GC-Regulated in silico: column adding up all GC-regulated genes in DAVID dataset, total at bottom; New DEGs in silico: column adding up all genes in DAVID dataset not already identified in TM cell dataset, total at bottom;
DEGs overlap: column adding up all GC-regulated genes in both TM cell and DAVID dataset, total at bottom

| | GC-regulated count | | | | |
|---|---|---|---|---|---|
| | DEGs in TM cells | New DEGs paired eye | Paired eye overlap | New DEGs in silico | In silico overlap |
| Prioritized genes | | | | | |
| ABCG2 | 1 | | | | 1 |
| ACADSB | | | | 1 | |
| ACSF2 | | | | | |
| ACSL3 | | | | 1 | |
| ADAM10 | 1 | | | | |
| ADGRG6 | | | | 1 | |
| ADGRL3 | | | | 1 | |
| ADGRL4 | | | | | |
| ADIPOR2 | | | | 1 | |
| AEBP2 | | | | 1 | |
| AGAP1 | | | | 1 | |
| AGR3 | | | | | |
| AGTR1 | | | | | |
| AHCTF1 | 1 | | | | 1 |
| AHR | | | | | |
| ALDH5A1 | 1 | | | | 1 |
| ALDH8A1 | | | | | |
| ALK | | | | 1 | |
| AMY2A | | | | | |
| AMY2B | | | | | |
| AMZ2 | | | | | |
| ANKS1B | | | | 1 | |
| ANO4 | | | | 1 | |
| ANO9 | | | | | |
| ANP32D | 1 | | | | |
| AOAH | | | | | |
| AP1AR | | | | | |
| AP2A2 | | | | 1 | |
| ARAP2 | | | | | |
| ARHGAP21 | | 1 | | | |
| ARHGEF26 | 1 | | | | 1 |
| ARHGEF28 | | | | | |
| ARID5B | | | | 1 | |
| ARRDC3 | 1 | | | | 1 |
| ARVCF | 1 | | | | 1 |
| ASB3 | | | | | |
| ASB8 | 1 | | | | 1 |
| ASNS | 1 | | | | 1 |
| ASPH | 1 | | | | 1 |
| ASXL1 | | | | 1 | |
| ATP4B | | | | 1 | |
| ATP8A1 | | | | | |
| ATP8B1 | 1 | | | | 1 |
| ATXN1 | 1 | | | | 1 |
| AVPR1A | | | | | |
| AZIN1 | | | | 1 | |
| B4GALNT3 | | | | | |
| B9D1 | | 1 | | | |
| BEND7 | 1 | | | | 1 |

| | | | | | |
|---|---|---|---|---|---|
| BTRC | 1 | | | | 1 |
| C1D | | | | | |
| C1orf21 | 1 | | | | 1 |
| C4orf19 | | | | 1 | |
| C6orf118 | | | | | |
| CAMSAP1 | | | | | |
| CAV2 | 1 | | | | 1 |
| CBR4 | | | | | |
| CCDC180 | | | | | |
| CCDC184 | | | | | |
| CCDC77 | | | | 1 | |
| CCDC91 | 1 | | | | 1 |
| CCNB2 | 1 | | | | |
| CCNG1 | 1 | | | | |
| CCR6 | | | | | |
| CCSER1 | | | | 1 | |
| CCSER2 | | | | 1 | |
| CCZ1 | | | | | |
| CCZ1B | | | | | |
| CDC42BPA | 1 | | | | 1 |
| CDCA2 | 1 | | 1 | | 1 |
| CDH2 | 1 | | | | 1 |
| CDH9 | | | | 1 | |
| CDK5RAP3 | | | | | |
| CDON | | | | 1 | |
| CELSR1 | | | | 1 | |
| CFAP99 | | | | | |
| CFTR | | 1 | | | |
| CHAC1 | 1 | | | | |
| CHCHD7 | | | | | |
| CHD4 | | | | 1 | |
| CLEC11A | | | | 1 | |
| CMC1 | | | | | |
| CNTN4 | | | | 1 | |
| CNTN5 | | | | 1 | |
| CNTN6 | 1 | | | | |
| CNTNAP2 | 1 | | | | 1 |
| CNTNAP4 | | 1 | | | |
| COL11A1 | 1 | | 1 | | 1 |
| COPZ2 | 1 | | | | 1 |
| CRTC1 | | | | 1 | |
| CSMD1 | | | | 1 | |
| CSMD3 | | | | 1 | |
| CTBP2 | | | | 1 | |
| CTSV | | | | | |
| CUZD1 | | | | | |
| DCANP1 | | | | | |
| DCT | | | 1 | 1 | |
| DIAPH3 | 1 | | | | 1 |
| DIDO1 | 1 | | | | 1 |
| DIRAS2 | | | | | |
| DLC1 | 1 | | | | 1 |
| DLGAP2 | | | | | |
| DLL4 | | | | | |
| DMRT2 | | | | | |
| DNASE2B | | | | 1 | |
| DPP10 | | | | 1 | |
| DSCAM | | | | 1 | |
| DSG1 | | | | | |
| DSG3 | | | | | |
| DYNC2H1 | 1 | | | | 1 |
| DYRK1A | | | | 1 | |
| E2F3 | | | | | |
| EBF2 | | | | 2 | |
| EDEM3 | 1 | | | | |
| EFHB | | | | | |
| EIF2AK4 | | | | 2 | |
| EIF3IP1 | | | | | |
| ENDOD1 | | | | | |
| EPHB1 | | | | 1 | |
| ERGIC3 | 1 | | | | 1 |
| ESD | | | | | |
| ETAA1 | | | | | |

| | | | | | |
|---|---|---|---|---|---|
| ETNK1 | 1 | | | | |
| EVX2 | | | | 2 | |
| FAM155A | 1 | | | | |
| FAM24B | | | | | |
| FAM76B | | | | | |
| FAXDC2 | | | | | |
| FBXO11 | | | | 1 | |
| FBXO7 | 1 | | | | 1 |
| FERMT1 | | | | | |
| FFAR2 | | | | | |
| FMN1 | | | | | |
| FOXN2 | 1 | | 1 | | 1 |
| FRMD3 | | 1 | | | |
| FST | 1 | | | | |
| FSTL5 | 1 | | | | 1 |
| GABRG2 | | | | 1 | |
| GABRP | | 1 | | | |
| GAL | | | | | |
| GALNT10 | 1 | | | | |
| GAS2L3 | 1 | | | | 1 |
| GATAD2A | | | | 1 | |
| GLRX | | | | | |
| GMNC | 1 | | | | |
| GMPS | | | | 1 | |
| GOT2 | 1 | | | | |
| GPAT2 | | | | 1 | |
| GPC5 | 1 | | | | 1 |
| GPC6 | | | | 1 | |
| GPLD1 | | | | 1 | |
| GTF2B | | | | 1 | |
| GYPC | | | | | |
| HAAO | | | | | |
| HAS2 | | | | 1 | |
| HCK | 1 | | | | |
| HDAC4 | 1 | | | | |
| HDAC9 | | | 1 | 1 | |
| HMCN1 | | | | 1 | |
| HOXD13 | | | | 1 | |
| HS3ST1 | 1 | | | | 1 |
| HS3ST4 | | | 1 | 1 | |
| HS6ST1 | 1 | | 1 | | |
| HTR2A | | | | 1 | |
| HYLS1 | 1 | | | | |
| ID2 | 1 | | | | |
| IDUA | 1 | | | | 1 |
| IGF2BP3 | | | | 1 | |
| INTU | 1 | | | | |
| IQCJ-SCHIP1 | 1 | | | | 1 |
| IQCM | | | | 1 | |
| ITPR2 | | | | | |
| JADE1 | | | | | |
| JAG2 | | | | 2 | |
| JDP2 | | | | | |
| JMJD7 | 1 | | | | |
| JMJD7-PLA2G4B | 1 | | | | 1 |
| KAZALD1 | | | | 1 | |
| KCND3 | | | 1 | 1 | |
| KCNE4 | 1 | | | | 1 |
| KCNH8 | | | | 1 | |
| KCNIP1 | 1 | | | | 1 |
| KCNIP3 | | | | 1 | |
| KCNIP4 | | | | 1 | |
| KCTD8 | | | | 1 | |
| KIAA1614 | | | | 1 | |
| KIF2B | | | | | |
| KIF3B | 1 | | | | 1 |
| KLF10 | | | | 1 | |
| KLF12 | 1 | | | | |
| KLF5 | 1 | | | | |
| KLF6 | | | 1 | 1 | |
| KLHDC7A | 1 | | | | 1 |
| KLHDC8A | 1 | | | | 1 |
| KRBOX1 | 1 | | | | |

| | | | | | |
|---|---|---|---|---|---|
| L1TD1 | | | | | |
| LARGE1 | | | | 1 | |
| LCORL | | | | | |
| LEMD1 | | | | 1 | |
| LEPR | | | | | |
| LEPROT | | | | | |
| LINC00028 | | | | 1 | |
| LINC00342 | 1 | | | | 1 |
| LINGO2 | | | | | |
| LIPC | 1 | | | | |
| LOC100130987 | | | | 1 | |
| LPAR3 | | | | 1 | |
| LRP2 | | | | 1 | |
| LTK | | | | | |
| LUZP1 | | | | 1 | |
| M1AP | | | | | |
| MACROD2 | | | | 1 | |
| MACROH2A1 | | | | 1 | |
| MAL | | | | 1 | |
| MALRD1 | | | | 1 | |
| MAML3 | | | | 1 | |
| MAPK10 | | | | | |
| MAPKBP1 | | | | 1 | |
| MAT2B | 1 | | | | 1 |
| MCTP2 | 1 | | | | 1 |
| MEOX2 | 1 | | 1 | | 1 |
| MFSD1 | | | | 1 | |
| MGA | | | | | |
| MIS18BP1 | 1 | | | | |
| MMRN1 | 1 | | | | 1 |
| MMS22L | | | | | |
| MND1 | | | | | |
| MOGAT1 | | | | 1 | |
| MOGS | 1 | | | | |
| MOS | | | | | |
| MRPL39 | | | | | |
| MRPS2 | | | | | |
| MRPS5 | | | | | |
| MSRB2 | | | | | |
| MTX2 | | | | | |
| MUC12 | | 1 | | | |
| MVB12B | | | | 1 | |
| MYO1E | | | | | |
| NAALADL2 | | | | | |
| NALCN | 1 | | | | 1 |
| NAT1 | 1 | | | | 1 |
| NBAS | | | | | |
| NCAM2 | | | | | |
| NEBL | | | | | |
| NEDD4L | | | | 1 | |
| NIFK | | | | 1 | |
| NKAIN3 | | | | 1 | |
| NKTR | 1 | | | | 1 |
| NOL4L | | | | | |
| NR2F1 | | | | 1 | |
| NRC32 | 1 | | | | 1 |
| NRG3 | | | | 1 | |
| NRXN1 | | | | | |
| NSG1 | | | | | |
| NT5C2 | | | | 1 | |
| NUDCD2 | | | | 1 | |
| NUDT7 | | | | 1 | |
| OLFM3 | | | | 1 | |
| OTUD1 | 1 | | | | |
| PALLD | 1 | | 1 | | |
| PC | | | | | |
| PCDH17 | | | | 1 | |
| PCDH20 | 1 | | 1 | | 1 |
| PCP4 | | | | 1 | |
| PCSK1 | | | | 1 | |
| PDCD6IP | 1 | | | | 1 |
| PDE3A | | | | | |
| PEX5L | 1 | | | | |

| | | | | | |
|---|---|---|---|---|---|
| PHYHIP | | | | 1 | |
| PITRM1 | | | | 1 | |
| PITX2 | 1 | | | | 1 |
| PKNOX2 | | | | 1 | |
| PLAG1 | | | | 1 | |
| PLAGL2 | 1 | | | | 1 |
| PLCH1 | 1 | | | | |
| PLPPR1 | | | | | |
| PLPPR5 | | | | | |
| POFUT1 | | | | 1 | |
| POLD4 | | | | | |
| POU3F2 | | | | 1 | |
| PPARGC1A | 1 | | | | 1 |
| PPM1H | | | | 1 | |
| PPP1R21 | 1 | | | | 1 |
| PPP2R3A | 1 | | | | 1 |
| PPP3CC | | | | 1 | |
| PRDM10 | | | | | |
| PRDM15 | | | | 1 | |
| PRKD1 | | | 1 | 1 | |
| PTHLH | | | | 1 | |
| PTPN7 | 1 | | | | 1 |
| PUM3 | 1 | | | | 1 |
| RAC1 | | | | | |
| RASSF3 | | 1 | | | |
| RBFOX1 | 1 | | 1 | | 1 |
| RBFOX3 | | | | | |
| RCBTB1 | 1 | | | | 1 |
| RELL1 | 1 | | | | 1 |
| RGMA | | | | 1 | |
| RGMB | | | | | |
| RGPD4 | 1 | | | | 1 |
| RHOBTB3 | 1 | | | | |
| RIBC2 | | | | 1 | |
| RIOK2 | 1 | | | | 1 |
| RND3 | | | | 1 | |
| RNF111 | | | | | |
| RNF144A | 1 | | | | 1 |
| RNF2 | | | | | |
| RPL10L | | | | 1 | |
| RTCB | | | | 1 | |
| RTKN | | | | 1 | |
| SALL2 | | | | | |
| SASH1 | | | | 1 | |
| SCHIP1 | | | | | |
| SDR16C5 | 1 | | | | 1 |
| SEMA5B | 1 | | | | 1 |
| SEPHS1 | | | | | |
| SGCG | | | | 1 | |
| SGK1 | | | | | |
| SGPP1 | 1 | | | | 1 |
| SH3RF1 | | | | 1 | |
| SHOX2 | | | | | |
| SIRPA | 1 | | | | 1 |
| SLC17A9 | | | | | |
| SLC25A26 | 1 | | | | |
| SLC4A5 | | | | | |
| SLC5A3 | | | | 1 | |
| SLC7A11 | | | | 1 | |
| SLIT2 | 1 | | | | 1 |
| SLITRK3 | | | | | |
| SLITRK5 | 1 | | | | 1 |
| SLTM | | | | | |
| SMARCA2 | | | | | |
| SOSTDC1 | | | | | |
| SOX17 | 1 | | | | 1 |
| SPAG4 | | | | 1 | |
| SPATS2 | 1 | | | | |
| SPRED2 | | | | 1 | |
| SPTY2D1 | | | | 1 | |
| SRRT | | | | | |
| SSH3 | 1 | | | | |
| SSX2IP | | | | | |

| | | | | | |
|---|---|---|---|---|---|
| ST6GAL2 | | | | | |
| STAG1 | | 1 | | | |
| STEAP2 | | | | | |
| STK35 | 1 | | 1 | | 1 |
| STON2 | 1 | | | | 1 |
| STX6 | | | | 1 | |
| SWT1 | 1 | | | | 1 |
| SYK | 1 | | | | 1 |
| TAC1 | | | | | |
| TAF5L | | | | | |
| TBC1D12 | | | | | |
| TBK1 | | | | | |
| TBL1XR1 | 1 | | 1 | | |
| TBX18 | | | | | |
| TEKT4 | 1 | | | | |
| TFDP2 | 1 | | | | |
| TFEC | 1 | | | | |
| TFPI | | | | 1 | |
| THSD7A | | | | | |
| TLE1 | 1 | | | | |
| TLR2 | | | | 1 | |
| TM2D1 | 1 | | | | 1 |
| TM9SF4 | 1 | | | | |
| TMEM108 | | | | | |
| TMEM201 | 1 | | | | |
| TMEM255B | | | | | |
| TMEM26 | 1 | | | | 1 |
| TMEM86A | | | | | |
| TMX3 | | | | | |
| TNFRSF13B | 1 | | 1 | | |
| TNK2 | | | | 1 | |
| TNRC6B | | | | 1 | |
| TOM1L1 | | | | 1 | |
| TRIB2 | 1 | | | | 1 |
| TRIM56 | 1 | | | | 1 |
| TRPS1 | 1 | | | | |
| TSC22D2 | 1 | | | | 1 |
| TSN | 1 | | | | |
| TSPAN14 | | | | 1 | |
| TTF1 | | | | 1 | |
| TUBGCP3 | | | | | |
| TYRO3 | | | | | |
| URB2 | | | | | |
| USP13 | | | | | |
| USP25 | 1 | | | | 1 |
| VDR | | | | | |
| VTA1 | 1 | | | | 1 |
| VWC2 | | | | 1 | |
| WDR89 | 1 | | | | 1 |
| WNT2B | | | | | |
| WNT5A | | | | | |
| WWC1 | | | | | |
| XKR4 | 1 | | | | 1 |
| XPNPEP1 | 1 | | | | |
| ZBTB47 | 1 | | | | 1 |
| ZBTB49 | | | | 1 | |
| ZFAT | 1 | | | | 1 |
| ZKSCAN2 | | | | 1 | |
| ZMAT3 | | | | | |
| ZNF514 | | | | | |
| ZNF536 | | | | | |
| ZNF641 | 1 | | | | 1 |
| ZNF728 | 1 | | | | |
| ZNF737 | | | | | |
| ZNF804B | | | | | |
| | 134 | 9 | 17 | 133 | 88 |
| GC-regulated count: | 33% | 2% | 4% | 33% | 22% |
| Percent of total: | red eye: | 26 | | | |

| | |
|---|---|
| Sum GC-regulated in silico: | 221 |
| Percent GC-regulated in silico | 54% |

| | |
|---|---|
| Total GC-regulated: | 276 |
| Percent GC-regulated total: | 68% |
| Total prioritized target genes: | 406 |

**Supplementary Table S5. Prioritized Target Gene Validation**

**GC-regulated differentially expressed genes (DEGs) from a human paired eye study**

From Kathirvel et al. (96)

| Responders all DEGs | Responders down-regulated DEGs | Responders up-regulated DEGs | Non-responders all DEGs | Non-responders down-regulated DEGs | Non-responders up-regulated DEGs | All DEGs that match GWAS prioritized genes | Responder DEGs that match GWAS prioritized genes |
|---|---|---|---|---|---|---|---|
| AATK | AATK | ABCA6 | ABCA6 | ADM2 | ABCA6 | ARHGEF26 | BEND7 |
| ABCA6 | ABCB1 | ABCA9 | ACA59 | ADRA2A | ACA59 | BEND7 | COL11A1 |
| ABCA9 | ABI3 | ABRA | ADH1A | AQP1 | ADH1A | CDH2 | COPZ2 |
| ABCB1 | ACP5 | ACA59 | ADH1B | ARHGAP9 | ADH1B | CHAC1 | DIAPH3 |
| ABI3 | ACPP | ACTG1P3 | ADH4 | ARSI | ADH4 | COL11A1 | FRMD3 |
| ABRA | ADAMTS9-AS1 | ADH1A | ADM2 | ASPN | ADRA1B | COPZ2 | FST |
| ACA59 | ADRA2A | ADH1B | ADRA1B | ATP1A3 | AFAP1L1 | DIAPH3 | HMCN1 |
| ACP5 | ADRB1 | ADH4 | ADRA2A | BEX2 | AIM1 | FRMD3 | KCNE4 |
| ACPP | AGR2 | ADRA1B | AFAP1L1 | BST2 | AKR1B15 | FST | KLHDC7A |
| ACTG1P3 | ANO7 | AFF2 | AIM1 | C1orf87 | ALDH1L1-AS2 | GAL | MFSD1 |
| ADAMTS9-AS1 | AOC1 | ANGPTL1 | AKR1B15 | C2orf40 | ALOX15B | HMCN1 | MVB12B |
| ADH1A | AOC3 | ANGPTL5 | ALDH1L1-AS2 | CA3 | ANGPTL1 | HS6ST1 | PC |
| ADH1B | AOC4P | ANGPTL7 | ALOX15B | CADM1 | ANGPTL5 | HTR2A | PCP4 |
| ADH4 | AP1M2 | ANO3 | ANGPTL1 | CCND2 | ANGPTL7 | KCNE4 | RBFOX3 |
| ADRA1B | AQP1 | AOX1 | ANGPTL5 | CCND2-AS1 | ANKRD2 | KLHDC7A | STEAP2 |
| ADRA2A | AQP3 | APOD | ANGPTL7 | CCND2-AS2 | AOX1 | MFSD1 | STON2 |
| ADRB1 | AQP5 | B3GALT2 | ANKRD2 | CERS1 | APCDD1 | MVB12B | TNK2 |
| AFF2 | ARSI | BCRP1 | AOX1 | CHAC1 | APOD | PC | |
| AGR2 | ASCL2 | BHLHE22 | APCDD1 | CILP2 | ARHGEF26 | PCP4 | |
| ANGPTL1 | ATAD3C | C1QTNF7 | APOD | CLDN1 | B3GNT7 | PTHLH | |
| ANGPTL5 | ATP1A2 | C5AR2 | AQP1 | CNGA1 | C3 | RBFOX1 | |
| ANGPTL7 | ATP8B4 | CCDC54 | ARHGAP9 | CNTN6 | CHI3L2 | RBFOX3 | |
| ANO3 | AZGP1 | CHRM2 | ARHGEF26 | COL19A1 | CHRNA5 | STEAP2 | |
| ANO7 | B3GAT1 | CPM | ARSI | CPA4 | CLSTN2 | STON2 | |
| AOC1 | B4GALNT3 | CYP51A1P2 | ASPN | CTNNA3 | CNTN1 | TBX18 | |
| AOC3 | BCAS1 | CYP7B1 | ATP1A3 | ELOVL2-AS1 | CPM | TNK2 | |
| AOC4P | BCL11B | DKK2 | B3GNT7 | EPHA3 | CRISPLD2 | | |
| AOX1 | BCL6B | FAM46B | BEX2 | ERICH2 | DGKG | | |
| AP1M2 | BMP7 | FAM65C | BST2 | FNDC1 | DKK2 | | |
| APOD | BST2 | FGD4 | C1orf87 | FST | DUSP5 | | |
| AQP1 | BTC | FGF14 | C2orf40 | GRIA2 | EDNRB | | |
| AQP3 | C10orf128 | FGFR4 | C3 | GRID2 | FAM107A | | |
| AQP5 | C1orf115 | FHL5 | CA3 | GRM5 | FAM150B | | |
| ARSI | C1orf116 | FKBP5 | CADM1 | IGFL2 | FAM46B | | |
| ASCL2 | C1QC | FMO2 | CCND2 | IL32 | FGFR4 | | |
| ATAD3C | C1QTNF2 | FPR1 | CCND2-AS1 | INA | FHL5 | | |
| ATP1A2 | C2orf40 | FRG2C | CCND2-AS2 | IPCEF1 | FKBP5 | | |
| ATP8B4 | C2orf54 | FRMD3 | CERS1 | KAL1 | FMO2 | | |
| AZGP1 | C3orf80 | GALNT15 | CHAC1 | KCNMB2 | FPR1 | | |
| B3GALT2 | C5orf38 | GIP | CHI3L2 | KCNS1 | GALNT15 | | |
| B3GAT1 | C7 | GJA5 | CHRNA5 | KCTD16 | GGT5 | | |
| B4GALNT3 | CACNG4 | GPX3 | CILP2 | KIAA1211 | GPM6B | | |
| BCAS1 | CALML3 | H19 | CLDN1 | KLHDC7B | GPRC5B | | |
| BCL11B | CAMK2A | HIF3A | CLSTN2 | KRT17 | GRK5-IT1 | | |
| BCL6B | CAMK2B | HNRNPA3P11 | CNGA1 | KRT17P1 | H19 | | |
| BCRP1 | CAMSAP3 | HSPD1P11 | CNTN1 | KRT23 | HEYL | | |
| BHLHE22 | CAPN11 | IGF2 | CNTN6 | LAMP3 | HIF3A | | |
| BMP7 | CASKIN1 | ITGA10 | COL19A1 | LFNG | HLX | | |
| BST2 | CBFA2T3 | KCNE1 | CPA4 | LINC01133 | HMGN2P15 | | |
| BTC | CBLC | KIAA1456 | CPM | LPHN3 | IGF2 | | |
| C10orf128 | CCDC64B | KLF15 | CRISPLD2 | LRRC15 | IGF2-AS | | |
| C1orf115 | CCDC88C | KRT18P62 | CTNNA3 | LRRN4CL | INHBB | | |
| C1orf116 | CCL5 | LDHAL6B | DGKG | LTK | ISM1 | | |

| | | | | | |
|---|---|---|---|---|---|
| C1QC | CCM2L | LEP | DKK2 | MXRA5Y | KIF5C |
| C1QTNF2 | CCR1 | LINC00547 | DUSP5 | MYHAS | LGI3 |
| C1QTNF7 | CD177 | LINC00664 | EDNRB | NGEF | LINC00525 |
| C2orf40 | CD34 | LINC00702 | ELOVL2-AS1 | NGFR | LINC00702 |
| C2orf54 | CD38 | LINC01088 | EPHA3 | NMNAT2 | LINC00704 |
| C3orf80 | CD7 | LOC100421166 | ERICH2 | NPY6R | LINC00968 |
| C5AR2 | CD74 | LSP1 | FAM107A | NTM | LINC01088 |
| C5orf38 | CD79B | MAOA | FAM150B | PADI2 | LSP1 |
| C7 | CDH1 | MAP1LC3C | FAM46B | PI16 | MAOA |
| CACNG4 | CDH22 | MARCH10 | FGFR4 | PLXNC1 | METTL7A |
| CALML3 | CDH3 | MIR5690 | FHL5 | PRSS35 | MIR5685 |
| CAMK2A | CDH5 | MOB3B | FKBP5 | RAB39B | MIRLET7D |
| CAMK2B | CEBPA | MRO | FMO2 | RARRES2 | MOB3B |
| CAMSAP3 | CEMIP | MYOC | FNDC1 | RBFOX1 | MREG |
| CAPN11 | CERS1 | NEDD9 | FPR1 | RDH12 | MRO |
| CASKIN1 | CFD | NKAIN2 | FST | RGS16 | MTFP1 |
| CBFA2T3 | CHGB | NPSR1-AS1 | GALNT15 | RGS7BP | MTSS1 |
| CBLC | CHI3L1 | NTRK2 | GGT5 | RIMS2 | MYBPHL |
| CCDC54 | CHIT1 | OCA2 | GPM6B | RIMS3 | MYOC |
| CCDC64B | CHODL | OLAH | GPRC5B | SEMA3D | NCAM1-AS1 |
| CCDC88C | CHRDL2 | P2RY14 | GRIA2 | SEMA6B | NEDD9 |
| CCL5 | CHRM1 | PDK4 | GRID2 | SLC14A1 | NR0B1 |
| CCM2L | CHRNA2 | PDLIM1P4 | GRK5-IT1 | SLC24A2 | NRCAM |
| CCR1 | CHRNA4 | PER1 | GRM5 | SLC7A5 | OCA2 |
| CD177 | CILP2 | PLCE1-AS1 | H19 | TAC3 | OLAH |
| CD34 | CITED1 | PPP1R14A | HEYL | TMEM63C | P2RY14 |
| CD38 | CLCA2 | PRODH | HIF3A | TNFSF15 | PDK4 |
| CD7 | CLDN3 | PRR33 | HLX | TNFSF18 | PER1 |
| CD74 | CLDN4 | PTK2B | HMGN2P15 | TNNT2 | PLIN5 |
| CD79B | CLEC14A | RAPGEF5 | IGF2 | UNC5B | PMEL |
| CDH1 | CNFN | RN7SKP69 | IGF2-AS | VCAN | PNMT |
| CDH22 | COL14A1 | RN7SL608P | IGFL2 | VCAN-AS1 | POM121L9P |
| CDH3 | COL15A1 | RNA5SP111 | IL32 | VNN1 | PPP1R14A |
| CDH5 | COL17A1 | RPL7P57 | INA | WNT2 | PRODH |
| CEBPA | COL9A1 | SAA1 | INHBB | | PRR33 |
| CEMIP | COL9A3 | SAA2 | IPCEF1 | | PTGDR2 |
| CERS1 | CPA3 | SAA4 | ISM1 | | PTHLH |
| CFD | CPA4 | SAMHD1 | KAL1 | | RAMP2 |
| CHGB | CPAMD8 | SCN3A | KCNMB2 | | RAMP2-AS1 |
| CHI3L1 | CPLX1 | SEMG1 | KCNS1 | | RGCC |
| CHIT1 | CPNE4 | SLC16A10 | KCTD16 | | RN7SKP97 |
| CHODL | CPXM1 | SLC16A12 | KIAA1211 | | RPA4 |
| CHRDL2 | CRABP2 | SLC38A11 | KIF5C | | RPL23AP81 |
| CHRM1 | CTSH | ST7-AS2 | KLHDC7B | | SAA1 |
| CHRM2 | CUX2 | SYNDIG1 | KRT17 | | SAA2 |
| CHRNA2 | CWH43 | TIMP4 | KRT17P1 | | SAMHD1 |
| CHRNA4 | CX3CL1 | TLDC2 | KRT23 | | SCARA5 |
| CILP2 | CXADR | TNFAIP8L3 | LAMP3 | | SCN3A |
| CITED1 | CXCL13 | TSC22D3 | LFNG | | SFTPC |
| CLCA2 | CXCL14 | TUSC5 | LGI3 | | SIX2 |
| CLDN3 | CXorf36 | UBE2CP1 | LINC00525 | | SLC16A10 |
| CLDN4 | CYBB | USP2 | LINC00702 | | SLC16A12 |
| CLEC14A | CYP24A1 | XRCC6P2 | LINC00704 | | SOAT2 |
| CNFN | CYP26A1 | ZBTB16 | LINC00968 | | SOX13 |
| COL14A1 | CYTH4 | | LINC01088 | | SPP1 |
| COL15A1 | DACT2 | | LINC01133 | | STAR |
| COL17A1 | DENND1C | | LPHN3 | | STEAP4 |
| COL9A1 | DES | | LRRC15 | | STOX1 |
| COL9A3 | DIO3 | | LRRN4CL | | SYN2 |
| CPA3 | DIO3OS | | LSP1 | | SYTL4 |
| CPA4 | DIRAS3 | | LTK | | TBXAS1 |
| CPAMD8 | DOC2B | | MAOA | | TIMP4 |
| CPLX1 | DRAXIN | | METTL7A | | TLDC2 |

| | | | |
|---|---|---|---|
| CPM | DSC2 | MIR5685 | TLE2 |
| CPNE4 | DTX1 | MIRLET7D | TLE6 |
| CPXM1 | ECSCR | MOB3B | TMOD1 |
| CRABP2 | EDN3 | MREG | TNNT3 |
| CTSH | EFCC1 | MRO | TRAV39 |
| CUX2 | EFNA1 | MTFP1 | TRPC3 |
| CWH43 | EHF | MTSS1 | TRPV6 |
| CX3CL1 | ELF3 | MXRA5Y | TXNRD1 |
| CXADR | ELFN1 | MYBPHL | USP2 |
| CXCL13 | ELFN2 | MYHAS | VAV3 |
| CXCL14 | ELOVL2 | MYOC | WSCD1 |
| CXorf36 | ELOVL7 | NCAM1-AS1 | XRCC6P2 |
| CYBB | EMID1 | NEDD9 | ZBTB16 |
| CYP24A1 | ENPP6 | NGEF | |
| CYP26A1 | EPCAM | NGFR | |
| CYP51A1P2 | ESPN | NMNAT2 | |
| CYP7B1 | ESRP1 | NPY6R | |
| CYTH4 | EVPL | NR0B1 | |
| DACT2 | EVX2 | NRCAM | |
| DENND1C | EXOC3L4 | NTM | |
| DES | FAM110D | OCA2 | |
| DIO3 | FAM19A3 | OLAH | |
| DIO3OS | FAM201A | P2RY14 | |
| DIRAS3 | FAM3B | PADI2 | |
| DKK2 | FAM3D | PDK4 | |
| DOC2B | FAM46C | PER1 | |
| DRAXIN | FAR2P1 | PI16 | |
| DSC2 | FAR2P2 | PLIN5 | |
| DTX1 | FBN3 | PLXNC1 | |
| ECSCR | FGR | PMEL | |
| EDN3 | FLT4 | PNMT | |
| EFCC1 | FNDC1 | POM121L9P | |
| EFNA1 | FOLH1 | PPP1R14A | |
| EHF | FOXA1 | PRODH | |
| ELF3 | FOXQ1 | PRR33 | |
| ELFN1 | FST | PRSS35 | |
| ELFN2 | FXYD2 | PTGDR2 | |
| ELOVL2 | FXYD3 | PTHLH | |
| ELOVL7 | FXYD6 | RAB39B | |
| EMID1 | FZD10 | RAMP2 | |
| ENPP6 | FZD10-AS1 | RAMP2-AS1 | |
| EPCAM | G0S2 | RARRES2 | |
| ESPN | GALNT16 | RBFOX1 | |
| ESRP1 | GAP43 | RDH12 | |
| EVPL | GATA5 | RGCC | |
| EVX2 | GGT6 | RGS16 | |
| EXOC3L4 | GIMAP1 | RGS7BP | |
| FAM110D | GIMAP4 | RIMS2 | |
| FAM19A3 | GIMAP5 | RIMS3 | |
| FAM201A | GIMAP6 | RN7SKP97 | |
| FAM3B | GIMAP7 | RPA4 | |
| FAM3D | GIMAP8 | RPL23AP81 | |
| FAM46B | GJB1 | SAA1 | |
| FAM46C | GJB2 | SAA2 | |
| FAM65C | GLB1L2 | SAMHD1 | |
| FAR2P1 | GNA15 | SCARA5 | |
| FAR2P2 | GNG4 | SCN3A | |
| FBN3 | GOLT1A | SEMA3D | |
| FGD4 | GP2 | SEMA6B | |
| FGF14 | GPR143 | SFTPC | |
| FGFR4 | GPR20 | SIX2 | |
| FGR | GPR56 | SLC14A1 | |
| FHL5 | GRAMD4P2 | SLC16A10 | |

| | | |
|---|---|---|
| FKBP5 | GREB1 | SLC16A12 |
| FLT4 | GREM2 | SLC24A2 |
| FMO2 | GRHL2 | SLC7A5 |
| FNDC1 | GRIK4 | SOAT2 |
| FOLH1 | GRIN1 | SOX13 |
| FOXA1 | GYLTL1B | SPP1 |
| FOXQ1 | HES2 | STAR |
| FPR1 | HGD | STEAP4 |
| FRG2C | HID1 | STOX1 |
| FRMD3 | HLA-DMB | SYN2 |
| FST | HLA-DOA | SYTL4 |
| FXYD2 | HLA-DPA1 | TAC3 |
| FXYD3 | HLA-DQA1 | TBXAS1 |
| FXYD6 | HLA-DQA2 | TIMP4 |
| FZD10 | HLA-DQB1 | TLDC2 |
| FZD10-AS1 | HLA-DRA | TLE2 |
| G0S2 | HLA-DRB1 | TLE6 |
| GALNT15 | HMCN2 | TMEM63C |
| GALNT16 | HMGCS2 | TMOD1 |
| GAP43 | HMGN2P46 | TNFSF15 |
| GATA5 | HOTTIP | TNFSF18 |
| GGT6 | HOXA10 | TNNT2 |
| GIMAP1 | HOXA11 | TNNT3 |
| GIMAP4 | HOXA11-AS | TRAV39 |
| GIMAP5 | HOXA13 | TRPC3 |
| GIMAP6 | HOXA3 | TRPV6 |
| GIMAP7 | HOXA5 | TXNRD1 |
| GIMAP8 | HOXA7 | UNC5B |
| GIP | HOXA9 | USP2 |
| GJA5 | HOXB13 | VAV3 |
| GJB1 | HOXD10 | VCAN |
| GJB2 | HOXD13 | VCAN-AS1 |
| GLB1L2 | HOXD3 | VNN1 |
| GNA15 | HOXD8 | WNT2 |
| GNG4 | HOXD9 | WSCD1 |
| GOLT1A | HPD | XRCC6P2 |
| GP2 | HPN | ZBTB16 |
| GPR143 | HPSE2 | |
| GPR20 | HSH2D | |
| GPR56 | HSPA12B | |
| GPX3 | HSPA6 | |
| GRAMD4P2 | ICAM2 | |
| GREB1 | IGF1 | |
| GREM2 | IGHG3 | |
| GRHL2 | IGHM | |
| GRIK4 | IGHV4-34 | |
| GRIN1 | IGHV4-39 | |
| GYLTL1B | IGJ | |
| H19 | IGKV1-5 | |
| HES2 | IGKV3-11 | |
| HGD | IGLC3 | |
| HID1 | IGLV1-40 | |
| HIF3A | IGSF9 | |
| HLA-DMB | IRF8 | |
| HLA-DOA | IRX4 | |
| HLA-DPA1 | ISL1 | |
| HLA-DQA1 | ITGAX | |
| HLA-DQA2 | ITM2A | |
| HLA-DQB1 | JPH3 | |
| HLA-DRA | JPH4 | |
| HLA-DRB1 | KCNH2 | |
| HMCN2 | KCNH6 | |
| HMGCS2 | KCNK5 | |

| | |
|---|---|
| HMGN2P46 | KCNN3 |
| HNRNPA3P11 | KCNQ1 |
| HOTTIP | KIAA1210 |
| HOXA10 | KIAA1211L |
| HOXA11 | KIAA1324 |
| HOXA11-AS | KIF12 |
| HOXA13 | KIF1A |
| HOXA3 | KL |
| HOXA5 | KLK11 |
| HOXA7 | KLK2 |
| HOXA9 | KLK3 |
| HOXB13 | KLK4 |
| HOXD10 | KLK7 |
| HOXD13 | KLKP1 |
| HOXD3 | KREMEN2 |
| HOXD8 | KRT13 |
| HOXD9 | KRT14 |
| HPD | KRT15 |
| HPN | KRT18 |
| HPSE2 | KRT23 |
| HSH2D | KRT5 |
| HSPA12B | KRT6A |
| HSPA6 | KRT79 |
| HSPD1P11 | KRT8 |
| ICAM2 | LAD1 |
| IGF1 | LAMP3 |
| IGF2 | LAMP5 |
| IGHG3 | LCK |
| IGHM | LCN2 |
| IGHV4-34 | LCN6 |
| IGHV4-39 | LCP1 |
| IGJ | LGALS7B |
| IGKV1-5 | LGR6 |
| IGKV3-11 | LINC00086 |
| IGLC3 | LINC00261 |
| IGLV1-40 | LINC00668 |
| IGSF9 | LINC00890 |
| IRF8 | LINC00964 |
| IRX4 | LINC01018 |
| ISL1 | LINC01297 |
| ITGA10 | LINC01315 |
| ITGAX | LMAN1L |
| ITM2A | LMO2 |
| JPH3 | LRG1 |
| JPH4 | LRRC15 |
| KCNE1 | LRRC26 |
| KCNH2 | LYPD3 |
| KCNH6 | LYZ |
| KCNK5 | MAL2 |
| KCNN3 | MALL |
| KCNQ1 | MAOB |
| KIAA1210 | MARVELD3 |
| KIAA1211L | MB |
| KIAA1324 | MCF2L |
| KIAA1456 | MCHR1 |
| KIF12 | MEOX1 |
| KIF1A | MEST |
| KL | MIR200A |
| KLF15 | MIR205HG |
| KLK11 | MIR3189 |
| KLK2 | MIR429 |
| KLK3 | MLC1 |
| KLK4 | MMP7 |

| | |
|---|---|
| KLK7 | MMP9 |
| KLKP1 | MPZL2 |
| KREMEN2 | MS4A6A |
| KRT13 | MSI1 |
| KRT14 | MSMB |
| KRT15 | MT1G |
| KRT18 | MT1H |
| KRT18P62 | MXRA5 |
| KRT23 | MXRA5Y |
| KRT5 | MYBPC1 |
| KRT6A | MYCT1 |
| KRT79 | MYH14 |
| KRT8 | MYL4 |
| LAD1 | MYOZ3 |
| LAMP3 | MZB1 |
| LAMP5 | NAT8L |
| LCK | NDP |
| LCN2 | NEFH |
| LCN6 | NELL2 |
| LCP1 | NFAM1 |
| LDHAL6B | NGEF |
| LEP | NGFR |
| LGALS7B | NKD2 |
| LGR6 | NKX3-1 |
| LINC00086 | NKX3-2 |
| LINC00261 | NOS3 |
| LINC00547 | NOSTRIN |
| LINC00664 | NOVA2 |
| LINC00668 | NPR1 |
| LINC00702 | NPY |
| LINC00890 | NTF4 |
| LINC00964 | NWD1 |
| LINC01018 | NYX |
| LINC01088 | OGDHL |
| LINC01297 | OR2I1P |
| LINC01315 | OVOL2 |
| LMAN1L | P2RX1 |
| LMO2 | P2RX2 |
| LOC100421166 | PAGE4 |
| LRG1 | PALD1 |
| LRRC15 | PARVG |
| LRRC26 | PCAT18 |
| LSP1 | PCAT4 |
| LYPD3 | PCDH17 |
| LYZ | PCGEM1 |
| MAL2 | PCP4 |
| MALL | PDE2A |
| MAOA | PDE3B |
| MAOB | PDE9A |
| MAP1LC3C | PDZK1IP1 |
| MARCH10 | PECAM1 |
| MARVELD3 | PGF |
| MB | PGM5-AS1 |
| MCF2L | PHF21B |
| MCHR1 | PI15 |
| MEOX1 | PI16 |
| MEST | PIGR |
| MIR200A | PKP1 |
| MIR205HG | PKP3 |
| MIR3189 | PLA2G2A |
| MIR429 | PLA2G4F |
| MIR5690 | PLA2G7 |
| MLC1 | PLCH2 |

| | |
|---|---|
| MMP7 | PLVAP |
| MMP9 | PMCH |
| MOB3B | POTEH |
| MPZL2 | PRAC1 |
| MRO | PROK1 |
| MS4A6A | PRR15L |
| MSI1 | PRSS16 |
| MSMB | PRSS22 |
| MT1G | PRSS35 |
| MT1H | PRSS8 |
| MXRA5 | PTCH2 |
| MXRA5Y | PTGER1 |
| MYBPC1 | PTPN6 |
| MYCT1 | RAB11FIP4 |
| MYH14 | RAB25 |
| MYL4 | RAI2 |
| MYOC | RAMP3 |
| MYOZ3 | RAP1GAP |
| MZB1 | RARRES2 |
| NAT8L | RASAL3 |
| NDP | RBBP8NL |
| NEDD9 | RBFOX3 |
| NEFH | RBM47 |
| NELL2 | RELN |
| NFAM1 | REM1 |
| NGEF | RGS16 |
| NGFR | RGS7BP |
| NKAIN2 | RIC3 |
| NKD2 | RLN1 |
| NKX3-1 | RNASE1 |
| NKX3-2 | RNF165 |
| NOS3 | RNF43 |
| NOSTRIN | ROBO4 |
| NOVA2 | RORC |
| NPR1 | RPLP0P2 |
| NPSR1-AS1 | RTN4RL1 |
| NPY | RUFY4 |
| NTF4 | S100A14 |
| NTRK2 | S1PR1 |
| NWD1 | SALL3 |
| NYX | SCGB3A1 |
| OCA2 | SDK2 |
| OGDHL | SELE |
| OLAH | SELP |
| OR2I1P | SEMA6B |
| OVOL2 | SERPINB11 |
| P2RX1 | SFN |
| P2RX2 | SFRP2 |
| P2RY14 | SFRP4 |
| PAGE4 | SH2D3C |
| PALD1 | SHH |
| PARVG | SHISA6 |
| PCAT18 | SLC14A1 |
| PCAT4 | SLC2A5 |
| PCDH17 | SLC44A4 |
| PCGEM1 | SLC45A3 |
| PCP4 | SLC52A3 |
| PDE2A | SLC7A14 |
| PDE3B | SLCO2A1 |
| PDE9A | SMOC1 |
| PDK4 | SMR3B |
| PDLIM1P4 | SORL1 |
| PDZK1IP1 | SOX18 |

| | |
|---|---|
| PECAM1 | SP5 |
| PER1 | SP8 |
| PGF | SPDEF |
| PGM5-AS1 | SPINK5 |
| PHF21B | SPINT1 |
| PI15 | SPNS2 |
| PI16 | SPOCK3 |
| PIGR | SPRR1B |
| PKP1 | SRD5A2 |
| PKP3 | SSTR1 |
| PLA2G2A | SSTR2 |
| PLA2G4F | ST14 |
| PLA2G7 | STAB1 |
| PLCE1-AS1 | STAC2 |
| PLCH2 | STC1 |
| PLVAP | SULT1C4 |
| PMCH | SULT2B1 |
| POTEH | SYNDIG1 |
| PPP1R14A | SYT13 |
| PRAC1 | SYT17 |
| PRODH | SYT7 |
| PROK1 | SYTL1 |
| PRR15L | TAC3 |
| PRR33 | TAL1 |
| PRSS16 | TBX1 |
| PRSS22 | TBX4 |
| PRSS35 | TBX5-AS1 |
| PRSS8 | TCEAL2 |
| PTCH2 | TENM1 |
| PTGER1 | TFCP2L1 |
| PTK2B | TFF1 |
| PTPN6 | TIE1 |
| RAB11FIP4 | TMC5 |
| RAB25 | TMC6 |
| RAI2 | TMC8 |
| RAMP3 | TMEFF2 |
| RAP1GAP | TMEM125 |
| RAPGEF5 | TMEM150C |
| RARRES2 | TMEM179 |
| RASAL3 | TMEM63C |
| RBBP8NL | TMPRSS2 |
| RBFOX3 | TNFSF15 |
| RBM47 | TNFSF18 |
| RELN | TNNT2 |
| REM1 | TNRC6C-AS1 |
| RGS16 | TNS4 |
| RGS7BP | TP63 |
| RIC3 | TPD52 |
| RLN1 | TPSAB1 |
| RN7SKP69 | TPSB2 |
| RN7SL608P | TPSD1 |
| RNA5SP111 | TRGC1 |
| RNASE1 | TRIM29 |
| RNF165 | TRPM8 |
| RNF43 | TRPV6 |
| ROBO4 | TSPAN1 |
| RORC | TSPAN7 |
| RPL7P57 | TTC22 |
| RPLP0P2 | TYROBP |
| RTN4RL1 | UPK3A |
| RUFY4 | VAMP8 |
| S100A14 | VENTX |
| S1PR1 | VIPR1 |

SAA1
SAA2
SAA4
SALL3
SAMHD1
SCGB3A1
SCN3A
SDK2
SELE
SELP
SEMA6B
SEMG1
SERPINB11
SFN
SFRP2
SFRP4
SH2D3C
SHH
SHISA6
SLC14A1
SLC16A10
SLC16A12
SLC2A5
SLC38A11
SLC44A4
SLC45A3
SLC52A3
SLC7A14
SLCO2A1
SMOC1
SMR3B
SORL1
SOX18
SP5
SP8
SPDEF
SPINK5
SPINT1
SPNS2
SPOCK3
SPRR1B
SRD5A2
SSTR1
SSTR2
ST14
ST7-AS2
STAB1
STAC2
STC1
SULT1C4
SULT2B1
SYNDIG1
SYT13
SYT17
SYT7
SYTL1
TAC3
TAL1
TBX1
TBX4
TBX5-AS1
TCEAL2
TENM1
VSTM2A
VWA1
VWF
WFDC2
WNK2
WNT10A
WNT10B
WNT11
WNT2
WNT4
WNT6
WNT7B
WSCD2
ZDHHC8P1
ZMYND15
ZNF385C

TFCP2L1
TFF1
TIE1
TIMP4
TLDC2
TMC5
TMC6
TMC8
TMEFF2
TMEM125
TMEM150C
TMEM179
TMEM63C
TMPRSS2
TNFAIP8L3
TNFSF15
TNFSF18
TNNT2
TNRC6C-AS1
TNS4
TP63
TPD52
TPSAB1
TPSB2
TPSD1
TRGC1
TRIM29
TRPM8
TRPV6
TSC22D3
TSPAN1
TSPAN7
TTC22
TUSC5
TYROBP
UBE2CP1
UPK3A
USP2
VAMP8
VENTX
VIPR1
VSTM2A
VWA1
VWF
WFDC2
WNK2
WNT10A
WNT10B
WNT11
WNT2
WNT4
WNT6
WNT7B
WSCD2
XRCC6P2
ZBTB16
ZDHHC8P1
ZMYND15
ZNF385C

**Supplementary Table S5. Prioritized Target Gene Validation**
**Paired eye study matches with prioritized target genes**

These studies used paired human donor eyes, or paired eyes from bovine donors as described in the text.
Gene functions are from GeneCards.
*Genes associated with a SNP identified in this study of genome-wide significance

| Species | Gene | Log2 Fold change | Function |
|---|---|---|---|
| **Genes differentially regulated in TM cells of steroid responders** | | | |
| Bovine | *BEND7 | -1.11 | Transcription factor |
| Bovine | *COL11A1 | -1.63 | Collagen XI subunit involved in tendon fibrillogenesis |
| Bovine | COPZ2 | -0.83 | Adaptor for COPI-1 mediated Golgi-ER transport |
| Bovine | DIAPH3 | 1.34 | Assembly of F-actin structures (Formin family) |
| Human | FRMD3 | 2.21 | Unknown |
| Human | *FST | -2.08 | Activin antagonist (TGFB superfamily member) |
| Bovine | HMCN1 | -1.54 | Multifunctional (Hemicentin) |
| Bovine | KCNE4 | -1.83 | Voltage-gated potassium channel, delayed rectifier (regulates KCNQ1) |
| Bovine | KLHDC7A | 2.16 | Unknown |
| Bovine | MFSD1 | -0.72 | Recycles lysosomal proteolysis products |
| Bovine | MVB12B | -0.82 | Component of ESCRT-I complex that regulates vesicular trafficking |
| Bovine | PC | -0.77 | Pyruvate carboxylase |
| Human | PCP4 | -4.30 | Calmodulin regulation |
| Bovine | PTHLH | -0.52 | Parathyroid hormone-like hormone |
| Human | RBFOX3 | -2.41 | Regulates alternative RNA splicing |
| Bovine | STEAP2 | 0.77 | Metalloreductase |
| Bovine | STON2 | -1.55 | Regulates vesicle-mediated transport |
| Bovine | TNK2 | -0.83 | Non-receptor protein kinase, downstream effector of CDC42 |
| **Genes differentially regulated in TM cells of steroid non-responders** | | | |
| Human | *ARHGEF26 | 2.08 | Rho-guanine nucleotide exchange factor (RhoG) |
| Bovine | CDH2 | -0.95 | Cell-cell adhesion (N-cadherin) |
| Human | CHAC1 | -2.43 | Enzymatic inhibition of Notch (g-glutamylcyclotransferase family) |
| Human | *FST | -2.00 | TGFB superfamily inhibitor (follistatin) |
| Bovine | GAL | -2.26 | Neuroendocrine peptide that controls smooth muscle contraction |
| Bovine | HS6ST1 | -0.92 | Enzyme that modifies heparan sulfate |
| Bovine | HTR2A | 1.93 | Serotonin receptor that reduces IOP when activated |
| Bovine | PTHLH | -2.50 | Parathyroid hormone-like hormone |
| Human | PTHLH | 2.42 | Parathyroid hormone-like hormone |
| Human | *RBFOX1 | -3.80 | Regulates alternative RNA splicing |
| Bovine | TBX18 | 1.12 | Transcriptional repressor |

**Supplementary Table S5. Prioritized Target Gene Validation**
**GC-regulation in silico**

As analyzed using NIH Database for Annotation, Visualization and Integrated Discovery (DAVID) Bioinformatics functional annotation clustering tool

**GWAS: 406 genes submitted** **402 DAVID IDs** UCSF_TFBS

Classification stringency: Lowest
Annotation Cluster 1 (top score) Enrichment score: 9.76

| Transcription Factor | Gene # | % of DAVID IDs | P value | Bonferroni-corrected | Notes |
|---|---|---|---|---|---|
| GR | 222 | 0.58 | 4.7E-08 | 8.3E-06 | Glucocorticoid receptor |

Classification stringency: Medium
Single Annotation Cluster Enrichment score: 24.16

| Transcription Factor | Gene # | % DAVID IDs | P value | Bonferroni-corrected | Notes |
|---|---|---|---|---|---|
| S8 | 252 | 0.63 | 4.00E-28 | 7.00E-26 | Paired domain TF |
| LHX3 | 220 | 0.55 | 9.10E-28 | 1.60E-25 | LIM domain TF |
| CHX10 | 211 | 0.52 | 1.30E-17 | 6.80E-17 | Homeobox TF |

**Paired Eye: 618 genes submitted** **590 DAVID IDs** **Responders** UCSF_TFBS

Classification stringency: Lowest
Annotation Cluster 1 (top score) Enrichment score: 3.42

| TF | Gene # | % DAVID IDs | P value | Bonferroni-corrected | Notes |
|---|---|---|---|---|---|
| GR | 264 | 0.45 | 8.01E-03 | 7.60E-01 | Glucocorticoid receptor |

Classification stringency: Medium
Single Annotation Cluster Enrichment score: 1.46

| Transcription Factor | Gene # | % DAVID IDs | P value | Bonferroni-corrected | Notes |
|---|---|---|---|---|---|
| S8 | 244 | 0.41 | 2.00E-04 | 3.40E-02 | |
| LHX3 | 182 | 0.31 | 4.72E-02 | 1.00E+00 | |
| CHX10 | 196 | 0.33 | 1.80E-01 | 1.00E+00 | |
| CART1 | 200 | 0.34 | 9.00E-01 | 1.00E+00 | Encoded by *ALX4,* a paired-like homeodomain TF |
