## Supplementary material for "Pharmacogenomics of steroid-induced ocular hypertension: relationship to high-tension glaucomas and new pathophysiologic insight": Suppl Table S6

**Supplementary Table S6. Top Prioritized Target Genes Additional Validation**
**Summary of chromatin analysis for SNPs that cluster in the top 26 risk loci**

The analysis was performed on RegulomeDB

| Prioritized Gene | Hit SNP | SNP Significance | Genome-level hit SNP in LD at $R^2$=0.4 | SNP Significance | Co-localizes | AOP cell types where transcription factors listed are primarily expressed |
|---|---|---|---|---|---|---|
| **Features of active chromatin** | | | | | | |
| HDAC9 | rs74455595 | Genome-level | | | caQTL | |
| NCAM2 | rs75024143 | Suggestive | | | SiPhy cons | |
| SPTY2D1 | rs151115079 | Genome-wide | | | SiPhy cons | |
| **Chip-identified binding proteins indicating active transcription** | | | | | | |
| BEND7 | rs184425183 | Genome-wide | | | CCNT2 | All cell types |
| GPLD1 | rs150586237 | Genome-wide | | | CTCF | All cell types |
| HDAC4 | rs188076929 | Genome-wide | | | RNA pol II | All cell types |
| SPTY2D1 | rs151115079 | Genome-wide | | | RNA pol II, TBP, TAF1 | All cell types |
| **TF binding motif** | | | | | | |
| AGAP1 | rs188848340 | Suggestive | rs147559909 | Genome-wide | GR binding motif | All cell types |
| COL11A1 | rs140420703 | Suggestive | | | GR binding motif | All cell types |
| CCR6 | rs184487573 | Suggestive | rs148153037 | Genome-wide | GR binding motif | All cell types |
| GPLD1 | Multiple non-hits | | rs150586237 | Genome-wide | GR binding motif | All cell types |
| HDAC4 | rs188076929 | | | | GR binding motif | All cell types |
| PPM1H | rs182437250 | Suggestive | | | GR binding motif | All cell types |
| **Chip-identified binding proteins known to interact with the GR** | | | | | | |
| HDAC4 | rs181217257 | Suggestive | | | JUND | All cell types |
| HDAC9 | rs10279777 | Genome-level | | | Pu-1, encoded by *SPI1* | Macrophages, ciliary muscle cells, neurons |
| SPTY2D1 | rs138414342 | Genome-level | | | STAT1, STAT3 | All cell types |
| SPTY2D1 | rs151115079 | Genome-level | | | Oct-2, encoded by *POU2F2* | macrophages, ciliary muscle cells, neurons |
| SPTY2D1 | rs151115079 | Genome-level | | | HEY1 | TM, vascular endothelium, Schwann cells |

**KEY**

| | |
|---|---|
| caQTL: | "Chromatin accessibility" QTL as measured by the Assay for Transposase-Accessible Chromatin (ATAC-seq). Chromatin accessibility is a reliable indicator of local cis-regulatory activity (1). |
| CCNT2: | A transcription factor that is a regulatory subunit of the positive transcription elongation factor B (P-TEFb) complex. P-TEFb is essential for RNA polymerase II's elongation of transcription and co-transcriptional processing. |
| CTCF: | An eleven zinc finger (ZF), multivalent transcriptional regulator that organizes chromatin. |
| JUND: | A subunit of AP1, which can act as a pioneer factor to prime GR binding. |
| HEY1: | A TF target of the NOTCH signaling pathway. GCs inhibit HEY1 expression (2). |
| Oct-2: | Encoded by POU2F2. The GR interacts physically with POU2F2 to modulate transcription (3). |
| Pu-1: | Also known as Pu.1, a tissue-specific TF of the hematopoietic lineage (4) encoded by SPI1. GR binding elements in DNA are primed by Pu-1, which interacts with the GR (5). |
| RNA pol II: | RNA polymerase II, an enzyme responsible for transcribing genes that encode proteins, as well as some non-coding RNA genes (6). |
| SiPhy Cons: | A measure of evolutionary conservation based on alignment of 17 vertebrate species used by HaploregV.2. Conservation suggests function importance of these loci. |
| STAT1: | A transcription factor. the GR does not physically bind STAT1, however an indirect mechanism of cross-modulation has been demonstrated that integrates STAT1 and the GR with PU.1 (7). |
| STAT3: | A transcription factor. STAT3 interacts with the GR reciprocally by tethering, i.e., GR tethering to DNA-bound STAT3 results in transcriptional repression, whereas STAT3 tethering to GR results in synergism (8). |
| TBP: | Also called TATA binding protein, a component of the transcription initiation machinery. |
| TAF1: | A TBP-associated factor. |
