## Supplementary material for "Pharmacogenomics of steroid-induced ocular hypertension: relationship to high-tension glaucomas and new pathophysiologic insight": Suppl Table S7

**Supplementary Table S7. Independent Replication**

Indianpolis-2 cohort, 12M quantitative trait (QT) data, comparing to discovery 12M QT data

| | |
|---|---|
| Threshold for replicative significance: | 5.00E-02 |
| Total SNPs found: | 92 |
| Total risk loci found: | 61 |

Gray shading = SNPs and target genes of genome-wide significance in the Indianapolis-1 cohort prior to adjustment for multiple comparisons
NA: not applicable

| ID | chr | POS_37 | freq | MAC | Score | Score.SE | Score.Stat | Score.pval | Func.refGene | Gene.refGene | GeneDetail.refGene | rsID | gnomAD_genome_ALL | gnomAD_genome_NFE | Rsq | hwe |
|---|---|---|---|---|---|---|---|---|---|---|---|---|---|---|---|---|
| 1:23465122:A:G | 1 | 23465122 | 0.004136 | 1 | 0.272297 | 0.126064 | 2.159983 | 3.08E-02 | intronic | LUZP1 | . | rs35145334 | 0.0093 | 0.016 | 0.847376 | 1 |
| 9:99922419:T:C | 9 | 99922419 | 0.020922 | 4 | 0.611973 | 0.283548 | 2.158269 | 3.09E-02 | ncRNA_intronic | ANKRD18CP | . | rs144138711 | 0.0219 | 0.0313 | 0.851063 | 1 |
| 3:179593768:G:A | 3 | 1.8E+08 | 0.005053 | 1 | -0.237 | 0.148742 | -1.59336 | 1.11E-01 | intronic | PEX5L | . | rs76356799 | 0.0159 | 0.0043 | 0.943713 | 1 |
| 8:62672723:G:A | 8 | 62672723 | 0.270408 | 56 | 1.388749 | 0.884022 | 1.570944 | 1.16E-01 | intergenic | MIR4470;NKAIN3 | dist=45305;dist=488778 | rs12114488 | 0.2842 | 0.3161 | 0.983174 | 0.315736 |
| 16:6412967:C:T | 16 | 6412967 | 0.009704 | 2 | -0.32659 | 0.210726 | -1.54983 | 1.21E-01 | intronic | RBFOX1 | . | rs80212581 | 0.0053 | 0.0049 | 0.980445 | 1 |
| 16:6433735:C:T | 16 | 6433735 | 0.009495 | 2 | -0.32103 | 0.208076 | -1.54287 | 1.23E-01 | intronic | RBFOX1 | . | rs140276610 | 0.0049 | 0.0053 | 0.976824 | 1 |
| 5:52662672:G:A | 5 | 52662672 | 0.010612 | 2 | 0.318972 | 0.208004 | 1.53349 | 1.25E-01 | intergenic | LOC257396;FST | dist=251716;dist=113592 | rs142934021 | 0.0054 | 0.0073 | 0.873041 | 1 |
| 20:61590782:T:C | 20 | 61590782 | 0.436893 | 90 | -1.66492 | 1.092258 | -1.52429 | 1.27E-01 | intronic | SLC17A9 | . | rs2427460 | 0.4464 | 0.502 | 1 | 0.843783 |
| 3:136134595:C:T | 3 | 1.36E+08 | 0.003675 | 1 | -0.1502 | 0.108093 | -1.38952 | 1.65E-01 | intronic | STAG1 | . | rs148248743 | 0.0018 | 0.0025 | 0.67579 | 1 |
| 2:67825685:C:T | 2 | 67825685 | 0.118481 | 24 | 0.992877 | 0.716431 | 1.385865 | 1.66E-01 | intergenic | ETAA1;LOC101927701 | dist=188152;dist=197501 | rs2902021 | 0.1449 | 0.0918 | 0.896322 | 0.6091 |
| 21:41307923:G:A | 21 | 41307923 | 0.891942 | 22 | 0.884584 | 0.662312 | 1.3356 | 1.82E-01 | intergenic | PCP4;DSCAM | dist=6601;dist=76420 | rs7275595 | 0.9016 | 0.9021 | 0.944606 | 1 |
| 21:41307573:G:A | 21 | 41307573 | 0.894325 | 22 | 0.881903 | 0.660646 | 1.33491 | 1.82E-01 | intergenic | PCP4;DSCAM | dist=6251;dist=76770 | rs9974985 | 0.8689 | 0.9016 | 0.955895 | 1 |
| 7:109155716:T:C | 7 | 1.09E+08 | 0.012738 | 3 | 0.281003 | 0.211468 | 1.328821 | 1.84E-01 | intergenic | C7orf66;EIF3IP1 | dist=631072;dist=443568 | rs73202425 | 0.0101 | 0.0152 | 0.773926 | 1 |
| 21:41304765:A:G | 21 | 41304765 | 0.894335 | 22 | 0.874736 | 0.658912 | 1.327545 | 1.84E-01 | intergenic | PCP4;DSCAM | dist=3443;dist=79578 | rs9636964 | 0.8763 | 0.9031 | 0.954712 | 1 |
| 21:41305720:G:A | 21 | 41305720 | 0.894335 | 22 | 0.874386 | 0.658989 | 1.326859 | 1.85E-01 | intergenic | PCP4;DSCAM | dist=4398;dist=78623 | rs9305683 | 0.9114 | 0.9025 | 0.954916 | 1 |
| 16:6858239:C:T | 16 | 6858239 | 0.001539 | 0 | 0.06332 | 0.049039 | 1.291222 | 1.97E-01 | intronic | RBFOX1 | . | rs138164904 | 0.0006 | 0.0009 | 0.313951 | 1 |
| 10:110423709:C:T | 10 | 1.1E+08 | 0.023029 | 5 | -0.46784 | 0.374512 | -1.2492 | 2.12E-01 | intergenic | LINC01435;XPNPEP1 | dist=594658;dist=1200815 | rs147944608 | 0.0105 | 0.0143 | 0.927213 | 1 |
| 11:439012:C:T | 11 | 439012 | 0.00417 | 1 | 0.159245 | 0.128995 | 1.234507 | 2.17E-01 | intronic | ANO9 | . | rs556274646 | 0.0033 | 0.006 | 0.85841 | 1 |
| 7:16291999:A:C | 7 | 16291999 | 0.00518 | 1 | 0.183439 | 0.150082 | 1.22226 | 2.22E-01 | ncRNA_intronic | ISPD-AS1 | . | rs143686474 | 0.0064 | 0.0088 | 0.935964 | 1 |
| 13:74357508:A:C | 13 | 74357508 | 0.009981 | 2 | -0.23228 | 0.205459 | -1.13056 | 2.58E-01 | intronic | KLF12 | . | rs368187808 | 0.0015 | 0.0025 | 0.90129 | 1 |
| 13:74357510:A:G | 13 | 74357510 | 0.009976 | 2 | -0.23215 | 0.205448 | -1.12998 | 2.58E-01 | intronic | KLF12 | . | rs372194899 | 0.0015 | 0.0025 | 0.90161 | 1 |
| 21:41309565:G:T | 21 | 41309565 | 0.873786 | 26 | 0.828474 | 0.746251 | 1.110182 | 2.67E-01 | intergenic | PCP4;DSCAM | dist=8243;dist=74778 | rs9981433 | 0.8718 | 0.8747 | 1 | 0.657884 |
| 3:158586849:T:C | 3 | 1.59E+08 | 0.005316 | 1 | 0.117913 | 0.111428 | 1.058203 | 2.90E-01 | intergenic | MFSD1;IQCJ | dist=39341;dist=200192 | rs139055031 | 0.0034 | 0.0053 | 0.484725 | 1 |
| 1:65965720:A:G | 1 | 65965720 | 0.073262 | 15 | 0.565301 | 0.545915 | 1.035511 | 3.00E-01 | intronic | LEPR | . | rs7534177 | 0.0979 | 0.0654 | 0.986327 | 1 |
| 4:169934087:G:A | 4 | 1.7E+08 | 0.087379 | 18 | 0.629008 | 0.621674 | 1.011797 | 3.12E-01 | intergenic | CBR4;SH3RF1 | dist=2619;dist=81320 | rs17615362 | 0.0556 | 0.0641 | 1 | 0.556401 |
| 4:169934725:T:C | 4 | 1.7E+08 | 0.087379 | 18 | 0.626322 | 0.620036 | 1.010139 | 3.12E-01 | intergenic | CBR4;SH3RF1 | dist=3257;dist=80682 | rs17543620 | 0.0621 | 0.0702 | 0.995146 | 1 |
| 1:65943471:C:T | 1 | 65943471 | 0.072874 | 15 | 0.552048 | 0.546971 | 1.009281 | 3.13E-01 | intronic | LEPR | . | rs17127656 | 0.1088 | 0.0656 | 0.992874 | 1 |
| 1:65957141:C:A | 1 | 65957141 | 0.072816 | 15 | 0.550472 | 0.548981 | 1.002716 | 3.16E-01 | intronic | LEPR | . | rs11579567 | 0.0982 | 0.0657 | 1 | 1 |
| 1:65948791:T:C | 1 | 65948791 | 0.072762 | 15 | 0.547758 | 0.546973 | 1.001434 | 3.17E-01 | intronic | LEPR | . | rs7518849 | 0.098 | 0.0656 | 0.993624 | 1 |
| 21:41308948:A:G | 21 | 41308948 | 0.883495 | 24 | 0.709232 | 0.720103 | 0.984903 | 3.25E-01 | intergenic | PCP4;DSCAM | dist=7626;dist=75395 | rs1005412 | 0.872 | 0.8867 | 1 | 0.621408 |
| 17:77210823:A:G | 17 | 77210823 | 0.935019 | 13 | -0.44428 | 0.461435 | -0.96281 | 3.36E-01 | intronic | RBFOX3 | . | rs2606194 | 0.8833 | 0.9459 | 0.8207 | 1 |
| 22:34473543:C:T | 22 | 34473543 | 0.024262 | 5 | 0.317336 | 0.332644 | 0.953978 | 3.40E-01 | intergenic | LARGE;ISX | dist=157127;dist=988586 | rs77297738 | 0.0124 | 0.0161 | 0.99959 | 1 |
| 17:46078602:C:T | 17 | 46078602 | 0.009699 | 2 | -0.20219 | 0.216503 | -0.93387 | 3.50E-01 | intergenic | CDK5RAP3;COPZ2 | dist=19450;dist=24931 | rs188353596 | 0.0024 | 0.0041 | 0.99899 | 1 |
| 20:30948839:C:T | 20 | 30948839 | 0.005403 | 1 | 0.133812 | 0.148013 | 0.904051 | 3.66E-01 | intronic | ASXL1 | . | rs148157126 | 0.0048 | 0.0067 | 0.890955 | 1 |
| 20:30946654:G:A | 20 | 30946654 | 0.005393 | 1 | 0.133515 | 0.147933 | 0.902538 | 3.67E-01 | intronic | ASXL1 | . | rs200198574 | 0.0051 | 0.0072 | 0.891725 | 1 |
| 20:30921343:C:T | 20 | 30921343 | 0.005325 | 1 | 0.132942 | 0.147636 | 0.900467 | 3.68E-01 | UTR3 | KIF3B | NM_004798:c.*2221C>T | rs139816293 | 0.0052 | 0.0072 | 0.899871 | 1 |
| 20:30892775:C:T | 20 | 30892775 | 0.005316 | 1 | 0.13291 | 0.147611 | 0.900412 | 3.68E-01 | intronic | KIF3B | . | rs143432612 | 0.005 | 0.0071 | 0.901 | 1 |
| 20:30862126:C:T | 20 | 30862126 | 0.005733 | 1 | 0.129444 | 0.147579 | 0.877116 | 3.80E-01 | intergenic | POFUT1;KIF3B | dist=35659;dist=3328 | rs145421321 | 0.0052 | 0.0075 | 0.840335 | 1 |
| 20:30771880:T:C | 20 | 30771880 | 0.006121 | 1 | 0.123868 | 0.151109 | 0.819724 | 4.12E-01 | intergenic | TM9SF4;TSPY26P | dist=16819;dist=5069 | rs193041547 | 0.0034 | 0.0046 | 0.830953 | 1 |
| 20:30780644:G:A | 20 | 30780644 | 0.00665 | 1 | 0.124148 | 0.152056 | 0.816465 | 4.14E-01 | UTR3 | PLAGL2 | NM_002657:c.*3611C>T | rs138055631 | 0.0052 | 0.0075 | 0.775332 | 1 |
| 20:30722429:G:A | 20 | 30722429 | 0.006835 | 1 | 0.123006 | 0.153898 | 0.79927 | 4.24E-01 | intronic | TM9SF4 | . | rs145791959 | 0.0052 | 0.0074 | 0.773327 | 1 |
| 1:202122499:C:T | 1 | 2.02E+08 | 0.009515 | 2 | 0.169365 | 0.213461 | 0.793423 | 4.28E-01 | intronic | PTPN7 | . | rs145804766 | 0.0059 | 0.008 | 0.98004 | 1 |
| 2:67831153:T:G | 2 | 67831153 | 0.068092 | 14 | 0.452063 | 0.578871 | 0.780939 | 4.35E-01 | intergenic | ETAA1;LOC101927701 | dist=193620;dist=192033 | rs75082290 | 0.0671 | 0.0909 | 0.993697 | 0.380032 |
| 20:30682231:C:T | 20 | 30682231 | 0.006354 | 1 | 0.117001 | 0.151263 | 0.773494 | 4.39E-01 | intronic | HCK | . | rs146249289 | 0.0032 | 0.0045 | 0.805469 | 1 |
| 20:6106741:A:G | 20 | 6106741 | 0.006544 | 1 | 0.123424 | 0.159657 | 0.773058 | 4.39E-01 | intergenic | FERMT1;CASC20 | dist=2550;dist=300638 | rs547186621 | 0.0023 | 0.0028 | 0.816709 | 1 |
| 20:30678170:C:T | 20 | 30678170 | 0.007146 | 1 | 0.114355 | 0.153477 | 0.745095 | 4.56E-01 | intronic | HCK | . | rs149859280 | 0.0049 | 0.0072 | 0.738256 | 1 |

| | | | | | | | | | | | | | | | | |
|---|---|---|---|---|---|---|---|---|---|---|---|---|---|---|---|---|
| 20:30675687:C:T | 20 | 30675687 | 0.007228 | 1 | 0.113145 | 0.153721 | 0.736045 | 4.62E-01 | intronic | HCK | . | rs138733283 | 0.0052 | 0.0074 | 0.732554 | 1 |
| 3:19586745:T:C | 3 | 19586745 | 0.006107 | 1 | -0.09553 | 0.135182 | -0.70668 | 4.80E-01 | intergenic | KCNH8;EFHB | dist=9610;dist=334221 | rs144788248 | 0.0068 | 0.0095 | 0.646941 | 1 |
| 13:61918888:G:A | 13 | 61918888 | 0.003325 | 1 | 0.070028 | 0.1011 | 0.69266 | 4.89E-01 | intergenic | MIR3169;PCDH20 | dist=144874;dist=64931 | rs113791989 | 0.0034 | 0.0052 | 0.633334 | 1 |
| 14:105590577:T:C | 14 | 1.06E+08 | 0.004782 | 1 | -0.06932 | 0.101416 | -0.68353 | 4.94E-01 | intergenic | LOC102723354;JAG2 | dist=24744;dist=16741 | rs190251199 | 0.0007 | 0.0012 | 0.479557 | 1 |
| 18:66029612:G:A | 18 | 66029612 | 0.003282 | 1 | -0.06389 | 0.095487 | -0.66911 | 5.03E-01 | intergenic | LOC643542;TMX3 | dist=462756;dist=311313 | rs559152067 | 0.0016 | 0.0026 | 0.635695 | 1 |
| 2:68110046:C:T | 2 | 68110046 | 0.005029 | 1 | 0.101998 | 0.153105 | 0.6662 | 5.05E-01 | intergenic | LOC101927701;C1D | dist=57352;dist=159286 | rs191298981 | 0.0082 | 0.0125 | 0.96377 | 1 |
| 7:117175907:T:C | 7 | 1.17E+08 | 0.004854 | 1 | -0.10071 | 0.153134 | -0.65764 | 5.11E-01 | intronic | CFTR | . | rs529110230 | 0.0019 | 0.0031 | 1 | 1 |
| 7:117199874:T:C | 7 | 1.17E+08 | 0.004854 | 1 | -0.10071 | 0.153134 | -0.65764 | 5.11E-01 | intronic | CFTR | . | rs142215699 | 0.0019 | 0.0031 | 1 | 1 |
| 7:117218328:A:C | 7 | 1.17E+08 | 0.004854 | 1 | -0.10071 | 0.153134 | -0.65764 | 5.11E-01 | intronic | CFTR | . | rs142721557 | 0.0019 | 0.0031 | 1 | 1 |
| 7:117225781:G:A | 7 | 1.17E+08 | 0.004854 | 1 | -0.10071 | 0.153134 | -0.65764 | 5.11E-01 | intronic | CFTR | . | rs201355675 | 0.0019 | 0.0031 | 1 | 1 |
| 7:117109021:A:G | 7 | 1.17E+08 | 0.004864 | 1 | -0.10065 | 0.153135 | -0.65728 | 5.11E-01 | intergenic | ASZ1;CFTR | dist=41444;dist=10996 | rs576047962 | 0.0017 | 0.0028 | 0.997998 | 1 |
| 13:74054452:T:C | 13 | 74054452 | 0.00485 | 1 | -0.09932 | 0.153747 | -0.64602 | 5.18E-01 | intergenic | KLF5;LINC00392 | dist=402772;dist=83929 | rs532269430 | 0.0015 | 0.0027 | 0.998995 | 1 |
| 7:117043294:A:G | 7 | 1.17E+08 | 0.004893 | 1 | -0.09876 | 0.152972 | -0.64563 | 5.19E-01 | intronic | ASZ1 | . | rs558784715 | 0.0016 | 0.0026 | 0.990072 | 1 |
| 2:68289497:T:C | 2 | 68289497 | 0.004859 | 1 | 0.097766 | 0.1532 | 0.638158 | 5.23E-01 | intronic | C1D | . | rs113154814 | 0.0106 | 0.0165 | 0.996991 | 1 |
| 2:68234930:A:C | 2 | 68234930 | 0.004864 | 1 | 0.097713 | 0.153361 | 0.637145 | 5.24E-01 | intergenic | LOC101927701;C1D | dist=182236;dist=34402 | rs113537164 | 0.0075 | 0.0113 | 0.997998 | 1 |
| 22:34492649:A:G | 22 | 34492649 | 0.019971 | 4 | 0.173113 | 0.277568 | 0.623678 | 5.33E-01 | intergenic | LARGE;ISX | dist=176233;dist=969480 | rs74572772 | 0.013 | 0.0159 | 0.632536 | 1 |
| 12:6681868:C:T | 12 | 6681868 | 0.03383 | 7 | -0.21438 | 0.367327 | -0.58361 | 5.59E-01 | intronic | CHD4 | . | rs61918041 | 0.0236 | 0.0328 | 0.98866 | 1 |
| 12:6684385:G:A | 12 | 6684385 | 0.033092 | 7 | -0.20101 | 0.360436 | -0.5577 | 5.77E-01 | intronic | CHD4 | . | rs113244573 | 0.0235 | 0.0322 | 0.969714 | 1 |
| 1:84866196:G:A | 1 | 84866196 | 0.013515 | 3 | -0.12062 | 0.230011 | -0.52442 | 6.00E-01 | intronic | DNASE2B | . | rs139390630 | 0.0097 | 0.0138 | 0.823131 | 1 |
| 4:11623450:G:A | 4 | 11623450 | 0.009558 | 2 | 0.110272 | 0.211241 | 0.522021 | 6.02E-01 | intergenic | HS3ST1;LOC101929019 | dist=192913;dist=601625 | rs189094663 | 0.0041 | 0.0072 | 0.979654 | 1 |
| 4:19013698:T:C | 4 | 19013698 | 0.003825 | 1 | -0.0443 | 0.086384 | -0.51281 | 6.08E-01 | intergenic | LCORL;SLIT2 | dist=990215;dist=1239830 | rs557276277 | 0.0083 | 0.0145 | 0.408399 | 1 |
| 3:177574308:G:A | 3 | 1.78E+08 | 0.004859 | 1 | 0.074678 | 0.153016 | 0.488038 | 6.26E-01 | ncRNA_intronic | KCCAT211 | . | rs189709453 | 0.002 | 0.0034 | 0.998997 | 1 |
| 3:177639777:T:C | 3 | 1.78E+08 | 0.004854 | 1 | 0.074358 | 0.153014 | 0.485954 | 6.27E-01 | intergenic | KCCAT211;LINC01014 | dist=22765;dist=497212 | rs186767531 | 0.0027 | 0.0044 | 1 | 1 |
| 3:177712448:C:T | 3 | 1.78E+08 | 0.004854 | 1 | 0.074358 | 0.153014 | 0.485954 | 6.27E-01 | intergenic | KCCAT211;LINC01014 | dist=95436;dist=424541 | rs182868205 | 0.0026 | 0.0045 | 1 | 1 |
| 15:40280073:G:A | 15 | 40280073 | 0.009694 | 2 | 0.101464 | 0.208912 | 0.485679 | 6.27E-01 | intronic | EIF2AK4 | . | rs56324718 | 0.0071 | 0.0074 | 0.998486 | 1 |
| 6:20458179:C:T | 6 | 20458179 | 0.014573 | 3 | 0.117291 | 0.247031 | 0.474803 | 6.35E-01 | intronic | E2F3 | . | rs73384619 | 0.0596 | 0.0208 | 0.999325 | 1 |
| 15:33113387:C:T | 15 | 33113387 | 0.004772 | 1 | -0.0691 | 0.149659 | -0.4617 | 6.44E-01 | intronic | FMN1 | . | rs187281112 | 0.0022 | 0.0039 | 0.982918 | 1 |
| 2:13059945:T:C | 2 | 13059945 | 0.003388 | 1 | -0.04453 | 0.098359 | -0.45274 | 6.51E-01 | intergenic | TRIB2;LOC100506474 | dist=177087;dist=46963 | rs76220567 | 0.0046 | 0.0056 | 0.634926 | 1 |
| 7:117274731:T:C | 7 | 1.17E+08 | 0.009432 | 2 | -0.08504 | 0.20862 | -0.40762 | 6.84E-01 | intronic | CFTR | . | rs188993522 | 0.0017 | 0.003 | 0.971904 | 1 |
| 22:34493647:G:A | 22 | 34493647 | 0.018714 | 4 | 0.107941 | 0.264895 | 0.407485 | 6.84E-01 | intergenic | LARGE;ISX | dist=177231;dist=968482 | rs80019988 | 0.013 | 0.016 | 0.588641 | 1 |
| 3:190584679:A:G | 3 | 1.91E+08 | 0.006 | 1 | -0.06018 | 0.150285 | -0.40046 | 6.89E-01 | intergenic | GMNC;SNAR-I | dist=4214;dist=11040 | rs117591241 | 0.0085 | 0.0102 | 0.813408 | 1 |
| 4:23509067:T:C | 4 | 23509067 | 0.00517 | 1 | 0.051794 | 0.145762 | 0.355335 | 7.22E-01 | intergenic | MIR548AJ2;PPARGC1A | dist=44341;dist=284577 | rs113063005 | 0.0035 | 0.0047 | 0.828627 | 1 |
| 1:85191044:G:A | 1 | 85191044 | 0.009714 | 2 | -0.0613 | 0.204456 | -0.29982 | 7.64E-01 | intergenic | SSX2IP;LPAR3 | dist=34804;dist=88042 | rs114067899 | 0.006 | 0.0099 | 0.999496 | 1 |
| 2:43096699:A:G | 2 | 43096699 | 0.00484 | 1 | -0.04328 | 0.153475 | -0.28203 | 7.78E-01 | intergenic | HAAO;LOC102723854 | dist=76948;dist=158293 | rs186792608 | 0.0013 | 0.0022 | 0.996985 | 1 |
| 20:1928157:A:G | 20 | 1928157 | 0.020388 | 4 | 0.072971 | 0.368343 | 0.198106 | 8.43E-01 | intergenic | SIRPA;PDYN | dist=7617;dist=31245 | rs62192733 | 0.0101 | 0.0133 | 0.933047 | 1 |
| 3:132842203:G:A | 3 | 1.33E+08 | 0.004782 | 1 | -0.02836 | 0.150192 | -0.18881 | 8.50E-01 | intronic | TMEM108 | . | rs114280794 | 0.0059 | 0.0089 | 0.984928 | 1 |
| 17:16839162:C:T | 17 | 16839162 | 0.004714 | 1 | 0.025523 | 0.144513 | 0.176614 | 8.60E-01 | intergenic | KRT16P2;TNFRSF13B | dist=103015;dist=3236 | rs116897913 | 0.0122 | 0.0155 | 0.918062 | 1 |
| 1:112349372:C:T | 1 | 1.12E+08 | 0.014718 | 3 | -0.04239 | 0.252199 | -0.16808 | 8.67E-01 | intronic | KCND3 | . | rs76098744 | 0.0239 | 0.0167 | 0.988834 | 1 |
| 1:112354418:G:A | 1 | 1.12E+08 | 0.01468 | 3 | -0.04122 | 0.252178 | -0.16346 | 8.70E-01 | intronic | KCND3 | . | rs74683551 | 0.0237 | 0.0165 | 0.991424 | 1 |
| 8:57055978:G:A | 8 | 57055978 | 0.015505 | 3 | -0.03569 | 0.25758 | -0.13855 | 8.90E-01 | intergenic | MOS;PLAG1 | dist=29437;dist=17490 | rs62515405 | 0.0194 | 0.0231 | 0.875866 | 1 |
| 10:23490165:C:T | 10 | 23490165 | 0.009024 | 2 | 0.022892 | 0.177334 | 0.129089 | 8.97E-01 | intergenic | PTF1A;LINC01552 | dist=6984;dist=2580 | rs142956968 | 0.0083 | 0.0142 | 0.711695 | 1 |
| 18:26603650:G:T | 18 | 26603650 | 0.014316 | 3 | -0.02055 | 0.226207 | -0.09086 | 9.28E-01 | intergenic | CDH2;MIR302F | dist=846240;dist=1275226 | rs147669485 | 0.0046 | 0.0075 | 0.684042 | 1 |
| 8:57141203:G:T | 8 | 57141203 | 0.006277 | 1 | -0.01312 | 0.146099 | -0.0898 | 9.28E-01 | intergenic | CHCHD7;SDR16C5 | dist=10027;dist=71367 | rs62515436 | 0.0114 | 0.0136 | 0.695945 | 1 |
| 10:23601847:G:A | 10 | 23601847 | 0.009451 | 2 | 0.015327 | 0.177616 | 0.086294 | 9.31E-01 | intergenic | LINC01552;C10orf67 | dist=73126;dist=3673 | rs145764464 | 0.0088 | 0.0145 | 0.685335 | 1 |
| 4:129647369:C:T | 4 | 1.3E+08 | 0.008466 | 2 | 0.013773 | 0.182927 | 0.075292 | 9.40E-01 | intergenic | LOC100507487;JADE1 | dist=206818;dist=83409 | rs181132315 | 0.0051 | 0.0056 | 0.803697 | 1 |
| 3:159392885:G:A | 3 | 1.59E+08 | 0.00367 | 1 | 0.006371 | 0.093544 | 0.068105 | 9.46E-01 | intronic | IQCJ-SCHIP1;SCHIP1 | . | rs143669489 | 0.0016 | 0.0029 | 0.705905 | 1 |

**Supplementary Table S7. Independent Replication**

Indianpolis-2 cohort, 3M quantitative trait (QT) data, comparing to discovery 3M QT data

| | |
|---|---|
| Threshold for replicative significance: | 5.00E-02 |
| Total SNPs found: | 290 |
| Total risk loci found: | 164 |

Gray shading = SNPs and target genes of genome-wide significance in the Indianapolis-1 cohort prior to adjustment for multiple comparisons
NA: not applicable

| ID | chr | POS_37 | freq | MAC | Score | Score.SE | Score.Stat | Score.pval | Func.refGene | Gene.refGene | GeneDetail.refGene | rsID | gnomAD_genome_ALL | gnomAD_genome_NFE | Rsq | hwe |
|---|---|---|---|---|---|---|---|---|---|---|---|---|---|---|---|---|
| 15:59634792: | 15 | 59634792 | 0.00785437 | 2 | 0.72549628 | 0.23800588 | 3.04822843 | 2.30E-03 | intronic | MYO1E | . | rs182303755 | 0.0048 | 0.008 | 0.815149 | 1 |
| 1:9655903:G: | 1 | 9655903 | 0.0048835 | 1 | 0.44239391 | 0.2092153 | 2.11453898 | 3.45E-02 | intronic | TMEM201 | . | rs115348382 | 0.003 | 0.0038 | 0.988049 | 1 |
| 8:123001411: | 8 | 123001411 | 0.01012621 | 2 | -0.5208585 | 0.25910027 | -2.01025844 | 4.44E-02 | intergenic | HAS2;SMILR | dist=343847;dist=425155 | rs536803366 | 0.0019 | 0.0033 | 0.829081 | 1 |
| 2:188375400: | 2 | 188375400 | 0.00799029 | 2 | 0.44447255 | 0.22253499 | 1.99731537 | 4.58E-02 | intronic | TFPI | . | rs112557251 | 0.0071 | 0.0095 | 0.735086 | 1 |
| 4:91825885:T | 4 | 91825885 | 0.03020388 | 6 | -0.93114505 | 0.48910792 | -1.90376194 | 5.69E-02 | intronic | CCSER1 | . | rs17017794 | 0.0466 | 0.0424 | 0.96782 | 1 |
| 4:163238743: | 4 | 163238743 | 0.01746117 | 4 | -0.60422933 | 0.32428843 | -1.86324663 | 6.24E-02 | intergenic | FSTL5;MIR4454 | dist=153557;dist=775983 | rs116651654 | 0.0122 | 0.0151 | 0.707424 | 1 |
| 8:18030674:C | 8 | 18030674 | 0.00499515 | 1 | -0.33601812 | 0.19641112 | -1.71078969 | 8.71E-02 | intronic | NAT1 | . | rs185874707 | 0.005 | 0.0077 | 0.968653 | 1 |
| 13:58213864: | 13 | 58213864 | 0.00693689 | 1 | 0.3593686 | 0.21068655 | 1.70570263 | 8.81E-02 | intronic | PCDH17 | . | rs534845494 | 0.0028 | 0.0046 | 0.795123 | 1 |
| 8:13251991:C | 8 | 13251991 | 0.02897087 | 6 | 0.81880035 | 0.48330222 | 1.69417875 | 9.02E-02 | intronic | DLC1 | . | rs139062456 | 0.0108 | 0.012 | 0.994566 | 1 |
| 1:180930424: | 1 | 180930424 | 0.00915534 | 2 | -0.45705544 | 0.27028043 | -1.69104159 | 9.08E-02 | intergenic | KIAA1614;STX6 | dist=15185;dist=11426 | rs76617932 | 0.007 | 0.0085 | 0.944101 | 1 |
| 13:57979549: | 13 | 57979549 | 0.0086068 | 2 | 0.39361307 | 0.24115519 | 1.63219821 | 1.03E-01 | intergenic | PRR20E;PCDH17 | dist=235197;dist=226240 | rs150077525 | 0.0043 | 0.0069 | 0.811421 | 1 |
| 6:16534923:C | 6 | 16534923 | 0.02279612 | 5 | 0.66818429 | 0.42178265 | 1.58419103 | 1.13E-01 | intronic | ATXN1 | . | rs1877768 | 0.0351 | 0.0251 | 0.946874 | 1 |
| 10:20372316: | 10 | 20372316 | 0.00485922 | 1 | -0.31534037 | 0.2019935 | -1.56114122 | 1.18E-01 | intronic | PLXDC2 | . | rs529011661 | 0.0024 | 0.004 | 0.998997 | 1 |
| 7:18414613:G | 7 | 18414613 | 0.00750971 | 2 | 0.35381562 | 0.22837308 | 1.54928774 | 1.21E-01 | intronic | HDAC9 | . | rs75606013 | 0.0176 | 0.0088 | 0.837192 | 1 |
| 7:18410845:G | 7 | 18410845 | 0.0075 | 2 | 0.35371015 | 0.22837492 | 1.54881349 | 1.21E-01 | intronic | HDAC9 | . | rs75773869 | 0.0169 | 0.0089 | 0.838286 | 1 |
| 7:18408761:A | 7 | 18408761 | 0.00750485 | 2 | 0.35353624 | 0.22837328 | 1.54806305 | 1.22E-01 | intronic | HDAC9 | . | rs77300464 | 0.0194 | 0.0089 | 0.837738 | 1 |
| 7:18410250:G | 7 | 18410250 | 0.00750485 | 2 | 0.35353624 | 0.22837328 | 1.54806305 | 1.22E-01 | intronic | HDAC9 | . | rs79602997 | 0.0172 | 0.0091 | 0.837738 | 1 |
| 7:18406573:C | 7 | 18406573 | 0.00750971 | 2 | 0.35365957 | 0.22847274 | 1.54792894 | 1.22E-01 | intronic | HDAC9 | . | rs75689761 | 0.0199 | 0.009 | 0.837902 | 1 |
| 8:55692112:G | 8 | 55692112 | 0.46905825 | 97 | -2.28157298 | 1.48522301 | -1.53618209 | 1.24E-01 | intergenic | RP1;XKR4 | dist=148718;dist=322905 | rs7843693 | 0.4432 | 0.388 | 0.990415 | 0.690082 |
| 8:55695310:A | 8 | 55695310 | 0.46908253 | 97 | -2.28105744 | 1.48521945 | -1.53583866 | 1.25E-01 | intergenic | RP1;XKR4 | dist=151916;dist=319707 | rs1396896 | 0.4436 | 0.3881 | 0.990319 | 0.690082 |
| 16:59302775: | 16 | 59302775 | 0.00819903 | 2 | 0.3450539 | 0.22555602 | 1.52979244 | 1.26E-01 | intergenic | GOT2;APOOP5 | dist=534514;dist=485270 | rs183817723 | 0.0044 | 0.0032 | 0.723856 | 1 |
| 10:24852783: | 10 | 24852783 | 0.02349515 | 5 | -0.67863561 | 0.44451742 | -1.52667945 | 1.27E-01 | intergenic | KIAA1217;ARHGAP21 | dist=16006;dist=19755 | rs12266995 | 0.1372 | 0.0298 | 0.965512 | 1 |
| 12:64920957: | 12 | 64920957 | 0.06796117 | 14 | -1.28781283 | 0.84420877 | -1.52546725 | 1.27E-01 | intergenic | TBK1;RASSF3 | dist=25058;dist=83336 | rs12315614 | 0.0961 | 0.0987 | 1 | 0.0619479 |
| 8:55699781:C | 8 | 55699781 | 0.46989321 | 97 | -2.25231369 | 1.48620762 | -1.51547715 | 1.30E-01 | intergenic | RP1;XKR4 | dist=156387;dist=315236 | rs1391462 | 0.4423 | 0.3872 | 0.985264 | 0.55697 |
| 10:12670548 | 10 | 126705489 | 0.00558252 | 1 | -0.30609897 | 0.20590453 | -1.48660628 | 1.37E-01 | intronic | CTBP2 | . | rs193093906 | 0.0079 | 0.0128 | 0.884528 | 1 |
| 10:13471195: | 10 | 13471195 | 0.00971359 | 2 | 0.42523176 | 0.2870612 | 1.48132789 | 1.39E-01 | intergenic | SEPHS1;BEND7 | dist=80897;dist=9289 | rs184458518 | 0.0051 | 0.0091 | 0.953876 | 1 |
| 10:13497976: | 10 | 13497976 | 0.00968447 | 2 | 0.4241555 | 0.28706533 | 1.47755736 | 1.40E-01 | intronic | BEND7 | . | rs117998251 | 0.0045 | 0.0079 | 0.956758 | 1 |
| 21:38863820: | 21 | 38863820 | 0.00485437 | 1 | -0.29925684 | 0.20380794 | -1.46832769 | 1.42E-01 | intronic | DYRK1A | . | rs118084887 | 0.0034 | 0.0057 | 0.993983 | 1 |
| 8:55705021:A | 8 | 55705021 | 0.46419418 | 96 | -2.19012891 | 1.49158643 | -1.46832183 | 1.42E-01 | intergenic | RP1;XKR4 | dist=161627;dist=309996 | rs12678939 | 0.3477 | 0.3842 | 0.987656 | 0.549725 |
| 21:38766484: | 21 | 38766484 | 0.00491748 | 1 | -0.30034437 | 0.20455787 | -1.46826114 | 1.42E-01 | intronic | DYRK1A | . | rs117185941 | 0.0044 | 0.0069 | 0.987225 | 1 |
| 21:38763032: | 21 | 38763032 | 0.00491748 | 1 | -0.30034437 | 0.20455787 | -1.46826114 | 1.42E-01 | intronic | DYRK1A | . | rs183586634 | 0.0035 | 0.0057 | 0.987225 | 1 |
| 8:55716905:C | 8 | 55716905 | 0.46564078 | 96 | -2.17879595 | 1.48634229 | -1.46587765 | 1.43E-01 | intergenic | RP1;XKR4 | dist=173511;dist=298112 | rs1498183 | 0.3488 | 0.3861 | 0.983748 | 0.690082 |
| 13:59286033: | 13 | 59286033 | 0.00468932 | 1 | 0.27744904 | 0.19035668 | 1.45752193 | 1.45E-01 | intergenic | LINC00374;DIAPH3 | dist=478782;dist=953688 | rs140062526 | 0.0029 | 0.005 | 0.934134 | 1 |
| 8:55698295:C | 8 | 55698295 | 0.46430583 | 96 | -2.17232303 | 1.49332116 | -1.45469246 | 1.46E-01 | intergenic | RP1;XKR4 | dist=154901;dist=316722 | rs2375219 | 0.3341 | 0.3847 | 0.990078 | 0.549725 |
| 10:13457520: | 10 | 13457520 | 0.00958252 | 2 | 0.41311337 | 0.28412944 | 1.45396183 | 1.46E-01 | intergenic | SEPHS1;BEND7 | dist=67222;dist=22964 | rs184425183 | 0.0037 | 0.0066 | 0.946776 | 1 |
| 8:55529073:C | 8 | 55529073 | 0.66018932 | 70 | 1.95385328 | 1.3487623 | 1.44862685 | 1.47E-01 | intronic | RP1 | . | rs9643828 | 0.698 | 0.6852 | 0.999978 | 0.826165 |
| 7:18406599:A | 7 | 18406599 | 0.0096068 | 2 | 0.40693621 | 0.28244996 | 1.44073739 | 1.50E-01 | intronic | HDAC9 | . | rs77346868 | 0.0255 | 0.009 | 0.987516 | 1 |
| 7:18407464:A | 7 | 18407464 | 0.00961165 | 2 | 0.40687836 | 0.2824402 | 1.44058233 | 1.50E-01 | intronic | HDAC9 | . | rs78225611 | 0.0266 | 0.0093 | 0.987013 | 1 |
| 7:18441589:G | 7 | 18441589 | 0.00975728 | 2 | 0.41068288 | 0.2856315 | 1.4378067 | 1.50E-01 | intronic | HDAC9 | . | rs10279777 | 0.0875 | 0.0105 | 0.993989 | 1 |
| 7:18425017:T | 7 | 18425017 | 0.00968447 | 2 | 0.40865008 | 0.28449932 | 1.43638331 | 1.51E-01 | intronic | HDAC9 | . | rs78907958 | 0.0305 | 0.0097 | 0.993463 | 1 |
| 7:18418351:A | 7 | 18418351 | 0.00967961 | 2 | 0.40862411 | 0.28450667 | 1.43625493 | 1.51E-01 | intronic | HDAC9 | . | rs61434999 | 0.0296 | 0.0096 | 0.993966 | 1 |
| 7:18447436:A | 7 | 18447436 | 0.00998544 | 2 | 0.41019612 | 0.28563996 | 1.43606001 | 1.51E-01 | intronic | HDAC9 | . | rs75090694 | 0.0289 | 0.0089 | 0.971757 | 1 |
| 7:18431110:G | 7 | 18431110 | 0.00970874 | 2 | 0.40971334 | 0.28568154 | 1.43416107 | 1.52E-01 | intronic | HDAC9 | . | rs76526501 | 0.0299 | 0.009 | 0.998991 | 1 |
| 7:18431784:A | 7 | 18431784 | 0.00970874 | 2 | 0.40971334 | 0.28568154 | 1.43416107 | 1.52E-01 | intronic | HDAC9 | . | rs74455595 | 0.0207 | 0.0091 | 0.998991 | 1 |
| 7:18433827:T | 7 | 18433827 | 0.00970874 | 2 | 0.40971334 | 0.28568154 | 1.43416107 | 1.52E-01 | intronic | HDAC9 | . | rs79182806 | 0.0192 | 0.0093 | 0.998991 | 1 |
| 7:18446741:G | 7 | 18446741 | 0.00971359 | 2 | 0.40983667 | 0.28582926 | 1.43385134 | 1.52E-01 | intronic | HDAC9 | . | rs17169602 | 0.0606 | 0.0097 | 0.999496 | 1 |
| 7:18443215:A | 7 | 18443215 | 0.00970874 | 2 | 0.40982622 | 0.28583174 | 1.43380237 | 1.52E-01 | intronic | HDAC9 | . | rs80156375 | 0.0293 | 0.0095 | 1 | 1 |
| 7:18446807:G | 7 | 18446807 | 0.00970874 | 2 | 0.40939242 | 0.28553735 | 1.43376139 | 1.52E-01 | intronic | HDAC9 | . | rs10486295 | 0.0603 | 0.0093 | 0.997983 | 1 |
| 7:18442275:G | 7 | 18442275 | 0.00971845 | 2 | 0.40947561 | 0.28568053 | 1.43333399 | 1.52E-01 | intronic | HDAC9 | . | rs77867199 | 0.1233 | 0.0109 | 0.997984 | 1 |
| 11:12526635 | 11 | 125266353 | 0.00484952 | 1 | 0.2862077 | 0.20021915 | 1.42947215 | 1.53E-01 | intronic | PKNOX2 | . | rs79539453 | 0.0389 | 0.0025 | 0.998995 | 1 |

| | | | | | | | | | | | | | | | | |
|---|---|---|---|---|---|---|---|---|---|---|---|---|---|---|---|---|
| 9:93415465:C | 9 | 93415465 | 0.00476214 | 1 | 0.2780232 | 0.19538179 | 1.42297394 | 1.55E-01 | intergenic | DIRAS2;SYK | dist=10078;dist=148497 | rs187213609 | 0.0016 | 0.0021 | 0.968915 | 1 |
| 8:1632241:A: | 8 | 1632241 | 0.03656311 | 8 | 0.7855068 | 0.55347195 | 1.41923506 | 1.56E-01 | intronic | DLGAP2 | . | rs187384541 | 0.0001 | 7.43E-05 | 0.733414 | 1 |
| 7:36574504:G | 7 | 36574504 | 0.0497233 | 10 | -0.80335714 | 0.56735516 | -1.41596869 | 1.57E-01 | intronic | AOAH | . | rs62447184 | 0.0457 | 0.0637 | 0.770789 | 1 |
| 3:155451289: | 3 | 155451289 | 0.0125 | 3 | -0.42971876 | 0.30457365 | -1.4108862 | 1.58E-01 | intergenic | PLCH1;C3orf33 | dist=29292;dist=29112 | rs139943877 | 0.0066 | 0.0089 | 0.872873 | 1 |
| 16:77715551: | 16 | 77715551 | 0.00730583 | 2 | 0.30227684 | 0.22038314 | 1.37159694 | 1.70E-01 | intergenic | ADAMTS18;NUDT7 | dist=246540;dist=40838 | rs529523094 | 0.002 | 0.003 | 0.754817 | 1 |
| 8:55556069:G | 8 | 55556069 | 0.63747088 | 75 | 1.84802295 | 1.36717244 | 1.35171168 | 1.76E-01 | intergenic | RP1;XKR4 | dist=12675;dist=458948 | rs423841 | 0.5779 | 0.6682 | 0.980476 | 0.669247 |
| 3:122722331: | 3 | 122722331 | 0.01008738 | 2 | 0.36512822 | 0.2705252 | 1.34970132 | 1.77E-01 | intronic | SEMA5B | . | rs80203220 | 0.0061 | 0.0094 | 0.95657 | 1 |
| 14:22641516: | 14 | 22641516 | 0.00970874 | 2 | 0.3901669 | 0.29078665 | 1.34176346 | 1.80E-01 | intergenic | OR4E1;DAD1 | dist=502284;dist=392291 | rs138215817 | 0.0013 | 0.002 | 0.981013 | 1 |
| 8:55676101:A | 8 | 55676101 | 0.37233981 | 77 | -1.84721403 | 1.39730572 | -1.32198273 | 1.86E-01 | intergenic | RP1;XKR4 | dist=132707;dist=338916 | rs1812506 | 0.4385 | 0.3406 | 0.99112 | 1 |
| 8:55630615:A | 8 | 55630615 | 0.37378641 | 77 | -1.85238189 | 1.40315023 | -1.32015935 | 1.87E-01 | intergenic | RP1;XKR4 | dist=87221;dist=384402 | rs1595406 | 0.4595 | 0.3453 | 1 | 1 |
| 8:55628637:G | 8 | 55628637 | 0.37276214 | 77 | -1.84311115 | 1.39647 | -1.31983584 | 1.87E-01 | intergenic | RP1;XKR4 | dist=85243;dist=386380 | rs720372 | 0.4592 | 0.3452 | 0.99107 | 1 |
| 8:55564609:A | 8 | 55564609 | 0.64240777 | 74 | 1.79482003 | 1.37005262 | 1.3100373 | 1.90E-01 | intergenic | RP1;XKR4 | dist=21215;dist=450408 | rs433324 | 0.5758 | 0.6715 | 0.98104 | 0.829223 |
| 8:55640722:T | 8 | 55640722 | 0.36893689 | 76 | -1.79459071 | 1.41295153 | -1.27010068 | 2.04E-01 | intergenic | RP1;XKR4 | dist=97328;dist=374295 | rs2375536 | 0.4555 | 0.3417 | 0.999979 | 1 |
| 9:93330047:T | 9 | 93330047 | 0.00485437 | 1 | 0.21871848 | 0.17295374 | 1.26460683 | 2.06E-01 | ncRNA_intronic | LINC01501 | . | rs183737367 | 0.0016 | 0.002 | 0.751636 | 1 |
| 22:19969182: | 22 | 19969182 | 0.00311651 | 1 | -0.1179241 | 0.09397486 | -1.25484732 | 2.10E-01 | exonic | ARVCF | . | rs113625788 | 0.0017 | 0.0025 | 0.324346 | 1 |
| 2:48655397:C | 2 | 48655397 | 0.01459709 | 3 | 0.44891532 | 0.35836978 | 1.25265953 | 2.10E-01 | intergenic | FOXN2;PPP1R21 | dist=48963;dist=12511 | rs189890455 | 0.0021 | 0.0032 | 0.996316 | 1 |
| 3:153657202: | 3 | 153657202 | 0.03714078 | 8 | 0.78752596 | 0.62917763 | 1.25167508 | 2.11E-01 | intergenic | C3orf79;ARHGEF26-AS1 | dist=436716;dist=84988 | rs16823323 | 0.0473 | 0.0194 | 0.961634 | 0.100888 |
| 9:93030699:G | 9 | 93030699 | 0.0043932 | 1 | -0.17792576 | 0.14232613 | -1.25012711 | 2.11E-01 | intergenic | MIR4290HG;LINC01508 | dist=226918;dist=32479 | rs545690161 | 0.0048 | 0.0067 | 0.588805 | 1 |
| 15:58879765: | 15 | 58879765 | 0.0559466 | 12 | 0.71250471 | 0.58898954 | 1.20970689 | 2.26E-01 | intergenic | LIPC;ADAM10 | dist=18692;dist=7638 | rs145439370 | 0.0125 | 0.0206 | 0.857914 | 1 |
| 21:23136973: | 21 | 23136973 | 0.04752427 | 10 | -0.69443215 | 0.57532419 | -1.20702755 | 2.27E-01 | ncRNA_intronic | LINC01425 | . | rs73227413 | 0.0173 | 0.0277 | 0.954032 | 1 |
| 6:167501386: | 6 | 167501386 | 0.00606796 | 1 | -0.2461956 | 0.20397669 | -1.20697911 | 2.27E-01 | intergenic | FGFR1OP;CCR6 | dist=45480;dist=23909 | rs148153037 | 0.0107 | 0.0154 | 0.844667 | 1 |
| 8:55592336:A | 8 | 55592336 | 0.63262136 | 76 | 1.58213184 | 1.33625472 | 1.18400468 | 2.36E-01 | intergenic | RP1;XKR4 | dist=48942;dist=422681 | rs405226 | 0.5785 | 0.6707 | 0.981155 | 0.526796 |
| 8:55582838:C | 8 | 55582838 | 0.63262136 | 76 | 1.58213184 | 1.33625472 | 1.18400468 | 2.36E-01 | intergenic | RP1;XKR4 | dist=39444;dist=432179 | rs3098298 | 0.579 | 0.6705 | 0.981155 | 0.526796 |
| 8:55587616:T | 8 | 55587616 | 0.63262136 | 76 | 1.58213184 | 1.33625472 | 1.18400468 | 2.36E-01 | intergenic | RP1;XKR4 | dist=44222;dist=427401 | rs367179 | 0.5779 | 0.6699 | 0.981155 | 0.526796 |
| 8:55580298:C | 8 | 55580298 | 0.63255825 | 76 | 1.57973282 | 1.33643121 | 1.18205323 | 2.37E-01 | intergenic | RP1;XKR4 | dist=36904;dist=434719 | rs432393 | 0.5785 | 0.6703 | 0.980895 | 0.526796 |
| 8:55680318:C | 8 | 55680318 | 0.36259224 | 75 | -1.58182122 | 1.34109432 | -1.17950035 | 2.38E-01 | intergenic | RP1;XKR4 | dist=136924;dist=334699 | rs2083123 | 0.3732 | 0.326 | 0.991313 | 0.518201 |
| 8:55674617:G | 8 | 55674617 | 0.3626602 | 75 | -1.58167598 | 1.34115013 | -1.17934297 | 2.38E-01 | intergenic | RP1;XKR4 | dist=131223;dist=340400 | rs12548593 | 0.401 | 0.3263 | 0.990983 | 0.518201 |
| 8:55685641:A | 8 | 55685641 | 0.36258738 | 75 | -1.58094881 | 1.34102424 | -1.17891143 | 2.38E-01 | intergenic | RP1;XKR4 | dist=142247;dist=329376 | rs10958428 | 0.4012 | 0.3258 | 0.991337 | 0.518201 |
| 8:55619508:C | 8 | 55619508 | 0.36306311 | 75 | -1.57981267 | 1.34073667 | -1.17831689 | 2.39E-01 | intergenic | RP1;XKR4 | dist=76114;dist=395509 | rs2375537 | 0.4222 | 0.3305 | 0.991097 | 0.518201 |
| 8:55678538:A | 8 | 55678538 | 0.36407767 | 75 | -1.58650703 | 1.34702313 | -1.17778752 | 2.39E-01 | intergenic | RP1;XKR4 | dist=135144;dist=336479 | rs1561297 | 0.4011 | 0.3261 | 1 | 0.53138 |
| 8:55629852:T | 8 | 55629852 | 0.36407767 | 75 | -1.58650703 | 1.34702313 | -1.17778752 | 2.39E-01 | intergenic | RP1;XKR4 | dist=86458;dist=385165 | rs1437781 | 0.4243 | 0.3302 | 1 | 0.53138 |
| 7:97577830:A | 7 | 97577830 | 0.00496602 | 1 | -0.22977371 | 0.19710528 | -1.16574102 | 2.44E-01 | intergenic | ASNS;MIR5692A1 | dist=75976;dist=15140 | rs192750513 | 0.0017 | 0.0021 | 0.977693 | 1 |
| 14:10559057 | 14 | 105590577 | 0.00478155 | 1 | -0.15912936 | 0.13727357 | -1.15921342 | 2.46E-01 | intergenic | LOC102723354;JAG2 | dist=24744;dist=16741 | rs190251199 | 0.0007 | 0.0012 | 0.479557 | 1 |
| 6:167513471: | 6 | 167513471 | 0.00788835 | 2 | -0.2748431 | 0.23718786 | -1.15875705 | 2.47E-01 | intergenic | FGFR1OP;CCR6 | dist=57565;dist=11824 | rs184487573 | 0.0111 | 0.0159 | 0.851684 | 1 |
| 7:97488823:C | 7 | 97488823 | 0.00485437 | 1 | -0.2274835 | 0.19712797 | -1.15398893 | 2.49E-01 | intronic | ASNS | . | rs181259864 | 0.0017 | 0.0021 | 1 | 1 |
| 17:48508221: | 17 | 48508221 | 0.02424757 | 5 | 0.51565366 | 0.45058422 | 1.14441129 | 2.52E-01 | intronic | ACSF2 | . | rs184613584 | 0.0079 | 0.012 | 0.992901 | 1 |
| 3:28227423:T | 3 | 28227423 | 0.02367961 | 5 | -0.43930097 | 0.39074283 | -1.12427134 | 2.61E-01 | intergenic | LOC100996624;CMC1 | dist=351796;dist=55701 | rs146007933 | 0.0187 | 0.0233 | 0.830561 | 1 |
| 15:58951660: | 15 | 58951660 | 0.04635437 | 10 | 0.56667404 | 0.50817586 | 1.11511404 | 2.65E-01 | intronic | ADAM10 | . | rs149425014 | 0.0102 | 0.0173 | 0.759352 | 1 |
| 8:55691458:C | 8 | 55691458 | 0.35292233 | 73 | -1.50681641 | 1.35767381 | -1.10985157 | 2.67E-01 | intergenic | RP1;XKR4 | dist=148064;dist=323559 | rs4737201 | 0.3056 | 0.323 | 0.990899 | 0.825756 |
| 8:55690220:T | 8 | 55690220 | 0.3529466 | 73 | -1.50258745 | 1.35814603 | -1.10635191 | 2.69E-01 | intergenic | RP1;XKR4 | dist=146826;dist=324797 | rs7822082 | 0.292 | 0.3259 | 0.991442 | 0.825756 |
| 8:55590975:G | 8 | 55590975 | 0.63752427 | 75 | 1.47054953 | 1.33573729 | 1.10092721 | 2.71E-01 | intergenic | RP1;XKR4 | dist=47581;dist=424042 | rs382476 | 0.6871 | 0.6731 | 0.981044 | 0.669247 |
| 8:55591609:G | 8 | 55591609 | 0.63752427 | 75 | 1.47054953 | 1.33573729 | 1.10092721 | 2.71E-01 | intergenic | RP1;XKR4 | dist=48215;dist=423408 | rs384543 | 0.6874 | 0.6732 | 0.981044 | 0.669247 |
| 8:55571940:A | 8 | 55571940 | 0.63752913 | 75 | 1.46958173 | 1.33575453 | 1.10018847 | 2.71E-01 | intergenic | RP1;XKR4 | dist=28546;dist=443077 | rs369623 | 0.6874 | 0.6729 | 0.981058 | 0.669247 |
| 8:55681876:T | 8 | 55681876 | 0.35778156 | 74 | -1.47304892 | 1.3405121 | -1.09887029 | 2.72E-01 | intergenic | RP1;XKR4 | dist=138482;dist=333141 | rs1391463 | 0.3108 | 0.3238 | 0.991203 | 0.662691 |
| 8:55597489:G | 8 | 55597489 | 0.63731068 | 75 | 1.46895537 | 1.33738957 | 1.09837508 | 2.72E-01 | intergenic | RP1;XKR4 | dist=54095;dist=417528 | rs384127 | 0.6871 | 0.6739 | 0.980706 | 0.669247 |
| 8:55679546:G | 8 | 55679546 | 0.35778156 | 74 | -1.47207174 | 1.34069826 | -1.09798884 | 2.72E-01 | intergenic | RP1;XKR4 | dist=136152;dist=335471 | rs4737676 | 0.3059 | 0.3233 | 0.991451 | 0.662691 |
| 8:55680792:C | 8 | 55680792 | 0.35778156 | 74 | -1.47207174 | 1.34069826 | -1.09798884 | 2.72E-01 | intergenic | RP1;XKR4 | dist=137398;dist=334225 | rs983248 | 0.3066 | 0.3238 | 0.991451 | 0.662691 |
| 8:55669829:A | 8 | 55669829 | 0.35778641 | 74 | -1.4718121 | 1.34072426 | -1.0977739 | 2.72E-01 | intergenic | RP1;XKR4 | dist=126435;dist=345188 | rs11987234 | 0.2969 | 0.3231 | 0.991224 | 0.662691 |
| 8:55688174:G | 8 | 55688174 | 0.35774757 | 74 | -1.47128038 | 1.3406321 | -1.09745274 | 2.72E-01 | intergenic | RP1;XKR4 | dist=144780;dist=326843 | rs13276543 | 0.2881 | 0.3202 | 0.991558 | 0.662691 |
| 8:55674149:G | 8 | 55674149 | 0.3578301 | 74 | -1.47115141 | 1.34056307 | -1.09741305 | 2.72E-01 | intergenic | RP1;XKR4 | dist=130755;dist=340868 | rs13277510 | 0.306 | 0.323 | 0.991119 | 0.662691 |
| 8:55678434:G | 8 | 55678434 | 0.35777185 | 74 | -1.47134764 | 1.34078154 | -1.09738059 | 2.72E-01 | intergenic | RP1;XKR4 | dist=135040;dist=336583 | rs16920698 | 0.3061 | 0.3233 | 0.991493 | 0.662691 |
| 8:55614690:A | 8 | 55614690 | 0.35821845 | 74 | -1.47023155 | 1.3401521 | -1.0970632 | 2.73E-01 | intergenic | RP1;XKR4 | dist=71296;dist=400327 | rs858397 | 0.2987 | 0.3262 | 0.991085 | 0.662691 |
| 8:55688171:C | 8 | 55688171 | 0.3592233 | 74 | -1.47572798 | 1.34660514 | -1.09588768 | 2.73E-01 | intergenic | RP1;XKR4 | dist=144777;dist=326846 | rs13278605 | 0.2879 | 0.32 | 1 | 0.671646 |
| 8:55632762:C | 8 | 55632762 | 0.3592233 | 74 | -1.47572798 | 1.34660514 | -1.09588768 | 2.73E-01 | intergenic | RP1;XKR4 | dist=89368;dist=382255 | rs1437782 | 0.3139 | 0.3281 | 1 | 0.671646 |
| 8:55574960:G | 8 | 55574960 | 0.6407767 | 74 | 1.47572798 | 1.34660514 | 1.09588768 | 2.73E-01 | intergenic | RP1;XKR4 | dist=31566;dist=440057 | rs446222 | 0.6836 | 0.6727 | 1 | 0.671646 |
| 8:55661654:C | 8 | 55661654 | 0.35807282 | 74 | -1.46115439 | 1.33994925 | -1.09045502 | 2.76E-01 | intergenic | RP1;XKR4 | dist=118260;dist=353363 | rs4737674 | 0.3142 | 0.3243 | 0.990315 | 0.662691 |
| 3:148330710: | 3 | 148330710 | 0.00986893 | 2 | 0.31483129 | 0.29092366 | 1.08217836 | 2.79E-01 | intergenic | LOC440982;AGTR1 | dist=1103013;dist=84948 | rs193153124 | 0.0046 | 0.0071 | 0.978098 | 1 |
| 10:3472846:C | 10 | 3472846 | 0.00759709 | 2 | 0.23535612 | 0.21966033 | 1.07145481 | 2.84E-01 | ncRNA_intronic | LOC105376360 | . | rs118040657 | 0.0038 | 0.0061 | 0.75025 | 1 |
| 4:90957372:G | 4 | 90957372 | 0.01477185 | 3 | 0.3764391 | 0.35152562 | 1.07087246 | 2.84E-01 | intergenic | MMRN1;CCSER1 | dist=81592;dist=91312 | rs142993106 | 0.0141 | 0.0158 | 0.985319 | 1 |
| 4:35572280:C | 4 | 35572280 | 0.00960194 | 2 | 0.30233113 | 0.28236665 | 1.0707041 | 2.84E-01 | intergenic | LOC101928622;ARAP2 | dist=1530765;dist=495340 | rs143287889 | 0.003 | 0.0051 | 0.988019 | 1 |
| 15:58837933: | 15 | 58837933 | 0.06800486 | 14 | 0.7150937 | 0.66861064 | 1.06952186 | 2.85E-01 | intronic | LIPC | . | rs6080 | 0.0208 | 0.0325 | 0.926378 | 1 |

| | | | | | | | | | | | | | | | | |
|---|---|---|---|---|---|---|---|---|---|---|---|---|---|---|---|---|
| 5:177390937: | 5 | 177390937 | 0.00842233 | 2 | -0.26338408 | 0.24665417 | -1.06782741 | 2.86E-01 | intergenic | LOC728554;PROP1 | dist=79668;dist=28299 | rs151015676 | 0.0028 | 0.0041 | 0.793454 | 1 |
| 4:87450675:T | 4 | 87450675 | 0.01436893 | 3 | -0.35002974 | 0.32934748 | -1.06279767 | 2.88E-01 | intergenic | MAPK10;MIR4452 | dist=76392;dist=12960 | rs147630370 | 0.0042 | 0.0069 | 0.846233 | 1 |
| 16:76179362: | 16 | 76179362 | 0.00514078 | 1 | 0.21213859 | 0.19985807 | 1.06144616 | 2.88E-01 | intergenic | TERF2IP;CNTNAP4 | dist=488021;dist=131814 | rs144954214 | 0.012 | 0.0091 | 0.86118 | 1 |
| 8:55640472:C | 8 | 55640472 | 0.35365049 | 73 | -1.4121439 | 1.3487482 | -1.04700336 | 2.95E-01 | intergenic | RP1;XKR4 | dist=97078;dist=374545 | rs10105693 | 0.2971 | 0.3231 | 0.991111 | 0.825756 |
| 2:48312950:G | 2 | 48312950 | 0.01345146 | 3 | 0.34321802 | 0.33187281 | 1.03418543 | 3.01E-01 | intergenic | FBXO11;FOXN2 | dist=180018;dist=228845 | rs76777840 | 0.0027 | 0.0042 | 0.92873 | 1 |
| 15:58982115: | 15 | 58982115 | 0.04498058 | 9 | 0.5128583 | 0.49958433 | 1.02657003 | 3.05E-01 | intronic | ADAM10 | . | rs146442492 | 0.0093 | 0.0159 | 0.750996 | 1 |
| 10:96217535: | 10 | 96217535 | 0.00973786 | 2 | -0.29443974 | 0.29332166 | -1.00381179 | 3.15E-01 | intronic | TBC1D12 | . | rs140706881 | 0.0099 | 0.0125 | 0.989983 | 1 |
| 1:103753974: | 1 | 103753974 | 0.01480097 | 3 | 0.24859719 | 0.25571623 | 0.97216041 | 3.31E-01 | intergenic | COL11A1;LOC101928436 | dist=179922;dist=206727 | rs113221952 | 0.0148 | 0.0207 | 0.596434 | 1 |
| 4:154411043: | 4 | 154411043 | 0.00691748 | 1 | -0.1809627 | 0.18830796 | -0.96099339 | 3.37E-01 | intronic | KIAA0922 | . | rs531769270 | 0.0009 | 0.0013 | 0.587973 | 1 |
| 2:177116420: | 2 | 177116420 | 0.0049466 | 1 | 0.1849834 | 0.19319589 | 0.95749136 | 3.38E-01 | intergenic | HOXD1;MTX2 | dist=60785;dist=17703 | rs184098071 | 0.0024 | 0.0035 | 0.909749 | 1 |
| 2:126393864: | 2 | 126393864 | 0.0048835 | 1 | -0.1835457 | 0.19343847 | -0.94885831 | 3.43E-01 | intergenic | CNTNAP5;GYPC | dist=720910;dist=1019647 | rs116189766 | 0.004 | 0.006 | 0.992027 | 1 |
| 4:89135588:A | 4 | 89135588 | 0.01553884 | 3 | -0.30708339 | 0.32653729 | -0.94042364 | 3.47E-01 | intronic | ABCG2 | . | rs147171192 | 0.0053 | 0.0061 | 0.870597 | 1 |
| 12:49853998: | 12 | 49853998 | 0.00226214 | 0 | -0.08861555 | 0.09615902 | -0.92155213 | 3.57E-01 | intronic | SPATS2 | . | rs568658857 | 0.0019 | 0.0036 | 0.464789 | 1 |
| 13:10801298 | 13 | 108012985 | 0.00766019 | 2 | -0.20567337 | 0.23037633 | -0.89277127 | 3.72E-01 | intronic | FAM155A | . | rs572961122 | 0.0029 | 0.0042 | 0.822166 | 1 |
| 4:90993018:T | 4 | 90993018 | 0.01207282 | 2 | 0.2641875 | 0.29872262 | 0.8843907 | 3.76E-01 | intergenic | MMRN1;CCSER1 | dist=117238;dist=55666 | rs146526206 | 0.0145 | 0.0161 | 0.885605 | 1 |
| 1:205331874: | 1 | 205331874 | 0.00485437 | 1 | 0.18816409 | 0.21525414 | 0.87414854 | 3.82E-01 | intergenic | KLHDC8A;LEMD1-AS1 | dist=5656;dist=10506 | rs10494861 | 0.0305 | 0.0033 | 1 | 1 |
| 3:177574308: | 3 | 177574308 | 0.00485922 | 1 | 0.18000965 | 0.20700024 | 0.86961084 | 3.85E-01 | ncRNA_intronic | KCCAT211 | . | rs189709453 | 0.002 | 0.0034 | 0.998997 | 1 |
| 12:28248852: | 12 | 28248852 | 0.0097233 | 2 | -0.25519409 | 0.29372871 | -0.86880879 | 3.85E-01 | intergenic | PTHLH;CCDC91 | dist=123936;dist=161281 | rs77353774 | 0.0047 | 0.0069 | 0.99849 | 1 |
| 3:177712448: | 3 | 177712448 | 0.00485437 | 1 | 0.17982452 | 0.20699733 | 0.86872868 | 3.85E-01 | intergenic | KCCAT211;LINC01014 | dist=95436;dist=424541 | rs182868205 | 0.0026 | 0.0045 | 1 | 1 |
| 3:177639777: | 3 | 177639777 | 0.00485437 | 1 | 0.17982452 | 0.20699733 | 0.86872868 | 3.85E-01 | intergenic | KCCAT211;LINC01014 | dist=22765;dist=497212 | rs186767531 | 0.0027 | 0.0044 | 1 | 1 |
| 12:26821687: | 12 | 26821687 | 0.00485437 | 1 | 0.17982452 | 0.20699733 | 0.86872868 | 3.85E-01 | intronic | ITPR2 | . | rs183466664 | 0.0039 | 0.0046 | 1 | 1 |
| 13:10801135 | 13 | 108011352 | 0.00770388 | 2 | -0.19963877 | 0.23042389 | -0.86639788 | 3.86E-01 | intronic | FAM155A | . | rs184265355 | 0.0023 | 0.0033 | 0.817644 | 1 |
| 1:185129502: | 1 | 185129502 | 0.00303398 | 1 | -0.08542469 | 0.0996576 | -0.85718185 | 3.91E-01 | intronic | SWT1 | . | rs145766563 | 0.0007 | 0.0009 | 0.364085 | 1 |
| 1:184955657: | 1 | 184955657 | 0.0019466 | 0 | -0.06342383 | 0.07405374 | -0.85645693 | 3.92E-01 | intergenic | FAM129A;RNF2 | dist=11939;dist=58894 | rs138480898 | 0.0008 | 0.001 | 0.31212 | 1 |
| 12:20424749: | 12 | 20424749 | 0.00976214 | 2 | -0.24642395 | 0.28890319 | -0.85296376 | 3.94E-01 | intergenic | LOC100506393;PDE3A | dist=172947;dist=97430 | rs118184666 | 0.005 | 0.0075 | 0.982689 | 1 |
| 12:28511473: | 12 | 28511473 | 0.00980583 | 2 | -0.25000717 | 0.29372451 | -0.8511621 | 3.95E-01 | intronic | CCDC91 | . | rs117991215 | 0.0044 | 0.0065 | 0.990095 | 1 |
| 12:28435962: | 12 | 28435962 | 0.00981553 | 2 | -0.24986877 | 0.29370617 | -0.85074401 | 3.95E-01 | intronic | CCDC91 | . | rs113167689 | 0.0047 | 0.0069 | 0.989132 | 1 |
| 7:89497045:T | 7 | 89497045 | 0.02540777 | 5 | -0.37731499 | 0.4449349 | -0.8480229 | 3.96E-01 | intergenic | ZNF804B;STEAP2-AS1 | dist=530674;dist=14622 | rs111391231 | 0.0077 | 0.0123 | 0.929489 | 1 |
| 12:28468969: | 12 | 28468969 | 0.0098301 | 2 | -0.24869443 | 0.29371655 | -0.84671574 | 3.97E-01 | intronic | CCDC91 | . | rs17510814 | 0.005 | 0.007 | 0.98766 | 1 |
| 12:28511096: | 12 | 28511096 | 0.0098301 | 2 | -0.24817368 | 0.29372682 | -0.84491322 | 3.98E-01 | intronic | CCDC91 | . | rs141756120 | 0.005 | 0.0071 | 0.987665 | 1 |
| 7:89516669:G | 7 | 89516669 | 0.02492719 | 5 | -0.36944585 | 0.43781414 | -0.84384174 | 3.99E-01 | ncRNA_intronic | STEAP2-AS1 | . | rs111900874 | 0.0077 | 0.0123 | 0.916811 | 1 |
| 13:23887014: | 13 | 23887014 | 0.00415534 | 1 | -0.14213336 | 0.16933337 | -0.83937004 | 4.01E-01 | intronic | SGCG | . | rs139598422 | 0.003 | 0.0047 | 0.764294 | 1 |
| 4:990967:C:T | 4 | 990967 | 0.00427185 | 1 | -0.14854758 | 0.17859567 | -0.83175355 | 4.06E-01 | intronic | IDUA | . | rs113651406 | 0.0025 | 0.0041 | 0.847816 | 1 |
| 11:99188380: | 11 | 99188380 | 0.02467476 | 5 | 0.36739406 | 0.44665973 | 0.82253678 | 4.11E-01 | intronic | CNTN5 | . | rs112007361 | 0.0222 | 0.0289 | 0.976846 | 1 |
| 18:55497457: | 18 | 55497457 | 0.00826699 | 2 | -0.19784548 | 0.24068482 | -0.82201064 | 4.11E-01 | intergenic | ATP8B1;NEDD4L | dist=27130;dist=214153 | rs146333745 | 0.0026 | 0.0042 | 0.804458 | 1 |
| 20:15858501: | 20 | 15858501 | 0.0188932 | 4 | -0.31907799 | 0.38923887 | -0.81974853 | 4.12E-01 | intronic | MACROD2 | . | rs140788628 | 0.0068 | 0.0107 | 0.930617 | 1 |
| 1:79880089:G | 1 | 79880089 | 0.00569418 | 1 | -0.11994275 | 0.14942111 | -0.80271621 | 4.22E-01 | intergenic | ADGRL4;LOC101927412 | dist=407594;dist=1121351 | rs149493615 | 0.0081 | 0.012 | 0.311201 | 1 |
| 12:20151132: | 12 | 20151132 | 0.01435922 | 3 | -0.28274382 | 0.35330677 | -0.80027854 | 4.24E-01 | intergenic | AEBP2;LOC100506393 | dist=475959;dist=16487 | rs141754456 | 0.0061 | 0.0088 | 0.984648 | 1 |
| 11:99089147: | 11 | 99089147 | 0.02435437 | 5 | 0.35240798 | 0.44618974 | 0.78981641 | 4.30E-01 | intronic | CNTN5 | . | rs74521112 | 0.0221 | 0.0288 | 0.986931 | 1 |
| 11:99093455: | 11 | 99093455 | 0.02435437 | 5 | 0.35229432 | 0.44618837 | 0.78956412 | 4.30E-01 | intronic | CNTN5 | . | rs79213709 | 0.0223 | 0.0291 | 0.986921 | 1 |
| 1:18948328:C | 1 | 18948328 | 0.00481553 | 1 | 0.1618529 | 0.20583153 | 0.7863368 | 4.32E-01 | intergenic | KLHDC7A;PAX7 | dist=135848;dist=9172 | rs186532456 | 0.0043 | 0.0063 | 0.991961 | 1 |
| 1:18959333:A | 1 | 18959333 | 0.00481553 | 1 | 0.1618529 | 0.20583153 | 0.7863368 | 4.32E-01 | intronic | PAX7 | . | rs562032622 | 0.0049 | 0.007 | 0.991961 | 1 |
| 3:136134595: | 3 | 136134595 | 0.00367476 | 1 | -0.11370888 | 0.14607079 | -0.77845055 | 4.36E-01 | intronic | STAG1 | . | rs148248743 | 0.0018 | 0.0025 | 0.67579 | 1 |
| 2:223814861: | 2 | 223814861 | 0.00459709 | 1 | 0.14317546 | 0.18417463 | 0.77738969 | 4.37E-01 | intergenic | ACSL3;KCNE4 | dist=6742;dist=101787 | rs185510569 | 0.0033 | 0.0052 | 0.882334 | 1 |
| 11:95099866: | 11 | 95099866 | 0.01479126 | 3 | -0.2745327 | 0.35316263 | -0.7773549 | 4.37E-01 | intergenic | LOC100129203;FAM76B | dist=132298;dist=402240 | rs149949098 | 0.0154 | 0.0228 | 0.947369 | 1 |
| 14:46294660: | 14 | 46294660 | 0.27903398 | 57 | 1.01329024 | 1.31694435 | 0.76942525 | 4.42E-01 | intergenic | MIS18BP1;LINC00871 | dist=572055;dist=238702 | rs428110 | 0.2639 | 0.3041 | 0.993976 | 0.624668 |
| 3:135532345: | 3 | 135532345 | 0.00469418 | 1 | -0.14923903 | 0.19399589 | -0.76928965 | 4.42E-01 | intergenic | EPHB1;PPP2R3A | dist=553038;dist=152170 | rs192443987 | 0.0019 | 0.0025 | 0.933075 | 1 |
| 1:89370702:G | 1 | 89370702 | 0.02508738 | 5 | -0.29457778 | 0.38461758 | -0.76589785 | 4.44E-01 | intergenic | GTF2B;CCBL2 | dist=13401;dist=30754 | rs34270375 | 0.0157 | 0.0211 | 0.688983 | 1 |
| 9:129042336: | 9 | 129042336 | 0.00452427 | 1 | 0.14328556 | 0.18753726 | 0.7640378 | 4.45E-01 | intergenic | LOC101929116;MVB12B | dist=4142;dist=46787 | rs146207930 | 0.0174 | 0.024 | 0.931691 | 1 |
| 18:27001580: | 18 | 27001580 | 0.00237864 | 0 | 0.07277788 | 0.09580724 | 0.75962812 | 4.47E-01 | intergenic | CDH2;MIR302F | dist=1244170;dist=877296 | rs185819304 | 0.0025 | 0.0021 | 0.46127 | 1 |
| 18:27030430: | 18 | 27030430 | 0.0019466 | 0 | 0.05910881 | 0.07809365 | 0.75689647 | 4.49E-01 | intergenic | CDH2;MIR302F | dist=1273020;dist=848446 | rs187942235 | 0.0023 | 0.002 | 0.374316 | 1 |
| 8:25613298:C | 8 | 25613298 | 0.00600971 | 1 | -0.1326355 | 0.17748976 | -0.74728537 | 4.55E-01 | intergenic | CDCA2;EBF2 | dist=247873;dist=85948 | rs188415494 | 0.0019 | 0.0033 | 0.644528 | 1 |
| 4:23509067:T | 4 | 23509067 | 0.0051699 | 1 | 0.14590427 | 0.19775652 | 0.73779751 | 4.61E-01 | intergenic | MIR548AJ2;PPARGC1A | dist=44341;dist=284577 | rs113063005 | 0.0035 | 0.0047 | 0.828627 | 1 |
| 7:17052778:G | 7 | 17052778 | 0.00495146 | 1 | -0.15159745 | 0.20607567 | -0.73563974 | 4.62E-01 | intergenic | AGR3;AHR | dist=131165;dist=285498 | rs117166500 | 0.0057 | 0.0054 | 0.980476 | 1 |
| 14:46280913: | 14 | 46280913 | 0.27882524 | 57 | 0.94398138 | 1.30478252 | 0.72347795 | 4.69E-01 | intergenic | MIS18BP1;LINC00871 | dist=558308;dist=252449 | rs176783 | 0.2781 | 0.3092 | 0.982613 | 0.803044 |
| 12:1870510:A | 12 | 1870510 | 0.00532524 | 1 | 0.14689221 | 0.20312563 | 0.72315942 | 4.70E-01 | intronic | ADIPOR2 | . | rs189360484 | 0.0003 | 0.0005 | 0.907353 | 1 |
| 14:46282970: | 14 | 46282970 | 0.2815534 | 58 | 0.94410239 | 1.31947222 | 0.71551517 | 4.74E-01 | intergenic | MIS18BP1;LINC00871 | dist=560365;dist=250392 | rs176786 | 0.2792 | 0.3091 | 1 | 0.634039 |
| 2:95967628:G | 2 | 95967628 | 0.0575534 | 12 | 0.55813521 | 0.78126016 | 0.71440377 | 4.75E-01 | intronic | KCNIP3 | . | rs76554191 | 0.0321 | 0.044 | 0.983166 | 0.169814 |
| 1:181247121: | 1 | 181247121 | 0.01397087 | 3 | -0.2247907 | 0.31663714 | -0.70993157 | 4.78E-01 | intergenic | GM140;CACNA1E | dist=39381;dist=205565 | rs183180157 | 0.0045 | 0.0053 | 0.835566 | 1 |
| 20:15813704: | 20 | 15813704 | 0.0290534 | 6 | 0.34861589 | 0.49308591 | 0.70700841 | 4.80E-01 | intronic | MACROD2 | . | rs2876414 | 0.0327 | 0.0266 | 0.996403 | 1 |
| 14:46260982: | 14 | 46260982 | 0.71485437 | 59 | -0.92281342 | 1.30945611 | -0.70473032 | 4.81E-01 | intergenic | MIS18BP1;LINC00871 | dist=538377;dist=272380 | rs1686289 | 0.6998 | 0.6934 | 0.977943 | 0.807255 |

| | | | | | | | | | | | | | | | | |
|---|---|---|---|---|---|---|---|---|---|---|---|---|---|---|---|---|
| 20:15813491: | 20 | 15813491 | 0.02942718 | 6 | 0.34556664 | 0.49252048 | 0.70162899 | 4.83E-01 | intronic | MACROD2 | . | rs2327968 | 0.0388 | 0.0336 | 0.983503 | 1 |
| 1:104114545: | 1 | 104114545 | 0.00949515 | 2 | 0.1690099 | 0.24351883 | 0.69403217 | 4.88E-01 | intronic | AMY2B | . | rs1856085 | 0.0047 | 0.0013 | 0.930312 | 1 |
| 1:104157143: | 1 | 104157143 | 0.00958738 | 2 | 0.17118256 | 0.2471166 | 0.69271977 | 4.88E-01 | intergenic | AMY2B;AMY2A | dist=34987;dist=2811 | rs143597860 | 0.0048 | 0.0014 | 0.948665 | 1 |
| 1:104310729: | 1 | 104310729 | 0.00925728 | 2 | 0.16424125 | 0.23779794 | 0.69067566 | 4.90E-01 | intergenic | AMY1A;LOC100129138 | dist=9418;dist=304916 | rs144541665 | 0.0048 | 0.0013 | 0.909477 | 1 |
| 1:79968131:T | 1 | 79968131 | 0.00691748 | 1 | -0.1243414 | 0.18038128 | -0.68932542 | 4.91E-01 | intergenic | ADGRL4;LOC101927412 | dist=495636;dist=1033309 | rs143811231 | 0.0073 | 0.0122 | 0.488277 | 1 |
| 7:100668425: | 7 | 100668425 | 0.00264078 | 1 | -0.07336753 | 0.107454 | -0.68278085 | 4.95E-01 | intronic | MUC17 | . | rs188028357 | 0.0012 | 0.0005 | 0.492675 | 1 |
| 22:45823032: | 22 | 45823032 | 0.00218932 | 0 | 0.06223473 | 0.09281172 | 0.67054821 | 5.03E-01 | intronic | RIBC2 | . | rs182959028 | 0.0055 | 0.0093 | 0.447795 | 1 |
| 6:134833127: | 6 | 134833127 | 0.00968932 | 2 | 0.19262125 | 0.28806153 | 0.66868094 | 5.04E-01 | intergenic | LINC01010;LOC101928304 | dist=7969;dist=13329 | rs141326851 | 0.0152 | 0.0166 | 0.993965 | 1 |
| 4:21259643:A | 4 | 21259643 | 0.00308738 | 1 | -0.07272364 | 0.10889196 | -0.66785139 | 5.04E-01 | intronic | KCNIP4 | . | rs183962155 | 0.0027 | 0.004 | 0.43349 | 1 |
| 3:1626661:T: | 3 | 1626661 | 0.00864078 | 2 | 0.14649408 | 0.21943009 | 0.66761162 | 5.04E-01 | intergenic | CNTN6;CNTN4 | dist=181369;dist=513889 | rs112475378 | 0.0133 | 0.0218 | 0.650295 | 1 |
| 10:82359100: | 10 | 82359100 | 0.00753398 | 2 | -0.15711058 | 0.23632408 | -0.66480988 | 5.06E-01 | intronic | SH2D4B | . | rs140277951 | 0.0046 | 0.0063 | 0.833841 | 1 |
| 2:48473143:G | 2 | 48473143 | 0.00489806 | 1 | -0.13344564 | 0.20718978 | -0.64407443 | 5.20E-01 | intergenic | FBXO11;FOXN2 | dist=340211;dist=68652 | rs145080832 | 0.0023 | 0.0035 | 0.991117 | 1 |
| 2:48631743:C | 2 | 48631743 | 0.00489806 | 1 | -0.13344564 | 0.20718978 | -0.64407443 | 5.20E-01 | intergenic | FOXN2;PPP1R21 | dist=25309;dist=36165 | rs184220112 | 0.0021 | 0.0032 | 0.991117 | 1 |
| 12:21012024: | 12 | 21012024 | 0.00468447 | 1 | -0.12737119 | 0.19849718 | -0.64167754 | 5.21E-01 | intronic | SLCO1B3 | . | rs151323346 | 0.0013 | 0.0021 | 0.950859 | 1 |
| 10:10295615 | 10 | 102956152 | 0.04854369 | 10 | -0.38863869 | 0.61565434 | -0.63126119 | 5.28E-01 | intergenic | LINC01514;LBX1 | dist=2248;dist=30581 | rs75334617 | 0.0323 | 0.0466 | 0.99979 | 1 |
| 2:223650026: | 2 | 223650026 | 0.00842718 | 2 | -0.15458772 | 0.2482031 | -0.62282751 | 5.33E-01 | intergenic | MOGAT1;ACSL3 | dist=75377;dist=75706 | rs185158855 | 0.0044 | 0.0069 | 0.854385 | 1 |
| 2:15682724:C | 2 | 15682724 | 0.00928155 | 2 | -0.16218645 | 0.2646395 | -0.61285806 | 5.40E-01 | intronic | NBAS | . | rs558553658 | 0.003 | 0.0053 | 0.953147 | 1 |
| 3:164808462: | 3 | 164808462 | 0.00857282 | 2 | -0.14648565 | 0.23949598 | -0.61164138 | 5.41E-01 | intergenic | SI;SLITRK3 | dist=12179;dist=96046 | rs141169929 | 0.0036 | 0.0053 | 0.774857 | 1 |
| 5:73285489:T | 5 | 73285489 | 0.00504854 | 1 | -0.11957494 | 0.1957184 | -0.61095399 | 5.41E-01 | intergenic | ARHGEF28;LINC01335 | dist=47671;dist=316746 | rs185771987 | 0.0027 | 0.0041 | 0.85912 | 1 |
| 22:46853180: | 22 | 46853180 | 0.00681553 | 1 | -0.11713689 | 0.19236142 | -0.60894169 | 5.43E-01 | intronic | CELSR1 | . | rs150946694 | 0.0051 | 0.0062 | 0.629526 | 1 |
| 13:91411079: | 13 | 91411079 | 0.0158932 | 3 | 0.21782246 | 0.36676046 | 0.59390933 | 5.53E-01 | intergenic | LINC01049;LINC00410 | dist=223484;dist=132129 | rs546286713 | 0.0038 | 0.0055 | 0.935869 | 1 |
| 2:53483429:G | 2 | 53483429 | 0.01595631 | 3 | -0.19860931 | 0.34469792 | -0.57618367 | 5.64E-01 | intergenic | MIR4431;ASB3 | dist=553676;dist=413688 | rs149421869 | 0.0032 | 0.0047 | 0.902614 | 1 |
| 15:94392882: | 15 | 94392882 | 0.00485437 | 1 | 0.12054613 | 0.21073279 | 0.57203309 | 5.67E-01 | intergenic | RGMA;LOC101927153 | dist=760439;dist=6907 | rs138217865 | 0.0013 | 0.0021 | 1 | 1 |
| 5:73372109:A | 5 | 73372109 | 0.00487864 | 1 | -0.11618831 | 0.20721267 | -0.56072008 | 5.75E-01 | intergenic | ARHGEF28;LINC01335 | dist=134291;dist=230126 | rs139360368 | 0.0031 | 0.0041 | 0.995013 | 1 |
| 11:66665729: | 11 | 66665729 | 0.00576214 | 1 | -0.09669872 | 0.17269163 | -0.55995021 | 5.76E-01 | intronic | PC | . | rs181812512 | 0.0026 | 0.0045 | 0.831451 | 1 |
| 8:22322319:A | 8 | 22322319 | 0.00464563 | 1 | 0.10981454 | 0.19724 | 0.55675591 | 5.78E-01 | intronic | PPP3CC | . | rs147601511 | 0.002 | 0.0036 | 0.915529 | 1 |
| 2:108038112: | 2 | 108038112 | 0.00713592 | 1 | 0.11343718 | 0.20593234 | 0.55084683 | 5.82E-01 | intergenic | MIR548AU;GACAT1 | dist=72131;dist=332456 | rs140352232 | 0.0021 | 0.0032 | 0.724891 | 1 |
| 1:99455745:T | 1 | 99455745 | 0.01333981 | 3 | -0.17828798 | 0.32573131 | -0.54734677 | 5.84E-01 | intronic | PLPPR5 | . | rs187518659 | 0.0043 | 0.0068 | 0.924329 | 1 |
| 1:193044178: | 1 | 193044178 | 0.00491262 | 1 | -0.103734 | 0.1980264 | -0.52383928 | 6.00E-01 | intronic | TROVE2 | . | rs180989936 | 0.0017 | 0.0029 | 0.880828 | 1 |
| 12:10118874 | 12 | 101188744 | 0.01456311 | 3 | -0.17403446 | 0.33240476 | -0.52356189 | 6.01E-01 | UTR5 | ANO4 | NM_178826:c.-106820G>A;NM_ | rs76904423 | 0.0146 | 0.0193 | 1 | 1 |
| 12:10118296 | 12 | 101182966 | 0.01448544 | 3 | -0.17284956 | 0.3309544 | -0.52227606 | 6.01E-01 | intergenic | GAS2L3;ANO4 | dist=160900;dist=5408 | rs76327548 | 0.0293 | 0.0223 | 0.994618 | 1 |
| 15:59435086: | 15 | 59435086 | 0.03308738 | 7 | 0.24283277 | 0.48819428 | 0.49741011 | 6.19E-01 | intronic | MYO1E | . | rs80292573 | 0.0289 | 0.025 | 0.830242 | 1 |
| 4:2426305:T: | 4 | 2426305 | 0.00950971 | 2 | -0.13017179 | 0.26223206 | -0.49639921 | 6.20E-01 | intronic | CFAP99 | . | rs12502861 | 0.0205 | 0.0183 | 0.855507 | 1 |
| 12:63306297: | 12 | 63306297 | 0.00448544 | 1 | -0.08561271 | 0.18168687 | -0.47121024 | 6.37E-01 | intronic | PPM1H | . | rs137880949 | 0.0011 | 0.0019 | 0.834481 | 1 |
| 1:112354418: | 1 | 112354418 | 0.01467961 | 3 | 0.16005174 | 0.34143383 | 0.46876357 | 6.39E-01 | intronic | KCND3 | . | rs74683551 | 0.0237 | 0.0165 | 0.991424 | 1 |
| 1:112349372: | 1 | 112349372 | 0.01471845 | 3 | 0.15899613 | 0.34146237 | 0.46563294 | 6.41E-01 | intronic | KCND3 | . | rs76098744 | 0.0239 | 0.0167 | 0.988834 | 1 |
| 10:12453340 | 10 | 124533409 | 0.00937379 | 2 | -0.12917376 | 0.27778761 | -0.46500908 | 6.42E-01 | ncRNA_intronic | DMBT1P1 | . | rs180828621 | 0.0022 | 0.0035 | 0.959645 | 1 |
| 9:85945465:C | 9 | 85945465 | 0.00970874 | 2 | -0.13617591 | 0.29512332 | -0.46142038 | 6.44E-01 | intronic | FRMD3 | . | rs190294315 | 0.0073 | 0.0119 | 1 | 1 |
| 9:85937714:G | 9 | 85937714 | 0.00960194 | 2 | -0.13458944 | 0.2917304 | -0.46134869 | 6.45E-01 | intronic | FRMD3 | . | rs188034471 | 0.0061 | 0.0101 | 0.987885 | 1 |
| 2:95722609:C | 2 | 95722609 | 0.05342233 | 11 | 0.3575444 | 0.77672804 | 0.46032122 | 6.45E-01 | intergenic | MAL;MRPS5 | dist=2872;dist=30343 | rs17746486 | 0.0239 | 0.0341 | 0.999522 | 0.0225883 |
| 2:176950420: | 2 | 176950420 | 0.00808252 | 2 | 0.10253581 | 0.23091352 | 0.44404422 | 6.57E-01 | intergenic | EVX2;HOXD13 | dist=1730;dist=7112 | rs556293455 | 0.0025 | 0.0034 | 0.832379 | 1 |
| 5:163268244: | 5 | 163268244 | 0.0051068 | 1 | -0.08900504 | 0.20103246 | -0.44273967 | 6.58E-01 | intergenic | MAT2B;LOC101927835 | dist=321885;dist=607184 | rs290120 | 0.0026 | 0.0041 | 0.951037 | 1 |
| 18:29050262: | 18 | 29050262 | 0.00488835 | 1 | 0.09200424 | 0.20908917 | 0.44002393 | 6.60E-01 | intronic | DSG3 | . | rs143538552 | 0.0034 | 0.0051 | 0.993025 | 1 |
| 18:29058384: | 18 | 29058384 | 0.00489806 | 1 | 0.09185895 | 0.20908607 | 0.43933555 | 6.60E-01 | UTR3 | DSG3 | NM_001944:c.*2161C>A | rs373746073 | 0.0034 | 0.0051 | 0.991053 | 1 |
| 7:100474786: | 7 | 100474786 | 0.00286893 | 1 | -0.04219979 | 0.09659454 | -0.43687546 | 6.62E-01 | intronic | SRRT | . | rs539713344 | 0.0017 | 0.0016 | 0.366221 | 1 |
| 11:67110852: | 11 | 67110852 | 0.00853398 | 2 | -0.09278968 | 0.21503911 | -0.43150141 | 6.66E-01 | ncRNA_intronic | LOC100130987 | . | rs529345909 | 0.0027 | 0.0046 | 0.784094 | 1 |
| 7:49544747:A | 7 | 49544747 | 0.00566019 | 1 | -0.08265789 | 0.20216681 | -0.40885986 | 6.83E-01 | intergenic | CDC14C;VWC2 | dist=577698;dist=268510 | rs574076561 | 0.0012 | 0.0023 | 0.74363 | 1 |
| 18:28816019: | 18 | 28816019 | 0.00424757 | 1 | 0.07214139 | 0.1775047 | 0.40641962 | 6.84E-01 | intergenic | DSC1;DSG1 | dist=73200;dist=82033 | rs139493286 | 0.0022 | 0.0029 | 0.823631 | 1 |
| 3:33940571:A | 3 | 33940571 | 0.06219417 | 13 | 0.25709647 | 0.65361904 | 0.393343 | 6.94E-01 | intergenic | PDCD6IP;LOC101928135 | dist=29372;dist=976718 | rs73057656 | 0.0413 | 0.0633 | 0.850362 | 1 |
| 22:32824278: | 22 | 32824278 | 0.00672816 | 1 | -0.0796672 | 0.21324038 | -0.37360276 | 7.09E-01 | intronic | BPIFC | . | rs78547898 | 0.0862 | 0.0418 | 0.735175 | 1 |
| 14:81717563: | 14 | 81717563 | 0.00485922 | 1 | -0.07380478 | 0.19779175 | -0.37314389 | 7.09E-01 | intergenic | GTF2A1;STON2 | dist=29988;dist=9431 | rs113767990 | 0.0061 | 0.0091 | 0.994994 | 1 |
| 10:12477927 | 10 | 124779274 | 0.0145534 | 3 | 0.12543817 | 0.34800648 | 0.36044779 | 7.19E-01 | intronic | ACADSB | . | rs147393020 | 0.0038 | 0.0058 | 0.999324 | 1 |
| 1:62484462:G | 1 | 62484462 | 0.01630097 | 3 | 0.11681659 | 0.32574933 | 0.35860884 | 7.20E-01 | intronic | INADL | . | rs2365739 | 0.0268 | 0.0259 | 0.907808 | 1 |
| 5:161493182: | 5 | 161493182 | 0.00951456 | 2 | 0.08538779 | 0.23837656 | 0.35820547 | 7.20E-01 | intergenic | LINC01202;GABRG2 | dist=64980;dist=1466 | rs74343174 | 0.0055 | 0.008 | 0.676915 | 1 |
| 2:67702707:G | 2 | 67702707 | 0.00943204 | 2 | 0.09036869 | 0.26393369 | 0.34239166 | 7.32E-01 | intergenic | ETAA1;LOC101927701 | dist=65174;dist=320479 | rs186142189 | 0.0034 | 0.0047 | 0.971958 | 1 |
| 2:67682724:G | 2 | 67682724 | 0.00944175 | 2 | 0.08898472 | 0.26330544 | 0.33795246 | 7.35E-01 | intergenic | ETAA1;LOC101927701 | dist=45191;dist=340462 | rs151272830 | 0.0031 | 0.0044 | 0.967052 | 1 |
| 2:151438271: | 2 | 151438271 | 0.01 | 2 | -0.09249531 | 0.294759 | -0.31379979 | 7.54E-01 | intergenic | LOC101929260;LOC101929 | dist=9536;dist=47140 | rs142894171 | 0.0041 | 0.0067 | 0.969366 | 1 |
| 1:229826378: | 1 | 229826378 | 0.0314466 | 6 | -0.1482821 | 0.49541331 | -0.29930988 | 7.65E-01 | intergenic | URB2;GALNT2 | dist=30431;dist=367158 | rs12036586 | 0.1365 | 0.0478 | 0.954664 | 1 |
| 10:20207052: | 10 | 20207052 | 0.01941748 | 4 | -0.12169638 | 0.41089326 | -0.29617517 | 7.67E-01 | intronic | PLXDC2 | . | rs117025967 | 0.0094 | 0.0101 | 1 | 1 |
| 1:102803484: | 1 | 102803484 | 0.00400971 | 1 | -0.04914759 | 0.1667589 | -0.29472244 | 7.68E-01 | intergenic | OLFM3;COL11A1 | dist=340694;dist=538539 | rs140420703 | 0.0042 | 0.0066 | 0.785826 | 1 |
| 7:11711845:G | 7 | 11711845 | 0.00747087 | 2 | -0.0681348 | 0.23499403 | -0.28994267 | 7.72E-01 | intronic | THSD7A | . | rs187978759 | 0.0012 | 0.0022 | 0.828633 | 1 |

| | | | | | | | | | | | | | | | |
|---|---|---|---|---|---|---|---|---|---|---|---|---|---|---|---|
| 7:6426479:G: | 7 | 6426479 | 0.02857282 | 6 | -0.12434427 | 0.44149559 | -0.28164329 | 7.78E-01 | intronic | RAC1 | . | rs17776100 | 0.028 | 0.0338 | 0.781514 | 1 |
| 6:99173116:A | 6 | 99173116 | 0.00428155 | 1 | -0.04769256 | 0.17560257 | -0.27159375 | 7.86E-01 | intergenic | MIR2113;POU3F2 | dist=700621;dist=109464 | rs147627638 | 0.0055 | 0.0081 | 0.81131 | 1 |
| 4:112677596: | 4 | 112677596 | 0.0119466 | 2 | -0.06454508 | 0.25063805 | -0.25752307 | 7.97E-01 | intergenic | PITX2;C4orf32 | dist=1114317;dist=388957 | rs145116559 | 0.0023 | 0.0041 | 0.636852 | 1 |
| 19:23123198: | 19 | 23123198 | 0.01480097 | 3 | 0.08187372 | 0.31929389 | 0.25642121 | 7.98E-01 | intergenic | ZNF99;ZNF728 | dist=156225;dist=34487 | rs111285015 | 0.0062 | 0.0102 | 0.724747 | 1 |
| 3:195646605: | 3 | 195646605 | 0.01941748 | 4 | 0.0978037 | 0.38203395 | 0.25600787 | 7.98E-01 | intergenic | TNK2;SDHAP1 | dist=10725;dist=40187 | rs191792521 | 0.0062 | 0.0105 | 0.863354 | 1 |
| 4:112689266: | 4 | 112689266 | 0.0119466 | 2 | -0.0609985 | 0.25169008 | -0.24235559 | 8.09E-01 | intergenic | PITX2;C4orf32 | dist=1125987;dist=377287 | rs181415102 | 0.0019 | 0.0033 | 0.642191 | 1 |
| 11:12369302 | 11 | 123693024 | 0.01457282 | 3 | 0.08589684 | 0.36200104 | 0.2372834 | 8.12E-01 | intergenic | OR6M1;TMEM225 | dist=15967;dist=60609 | rs141281289 | 0.0053 | 0.0089 | 0.994625 | 1 |
| 6:142307100: | 6 | 142307100 | 0.01361165 | 3 | 0.05884836 | 0.25543344 | 0.23038627 | 8.18E-01 | intergenic | MIR4465;NMBR | dist=1302080;dist=89645 | rs72983831 | 0.0225 | 0.0173 | 0.660998 | 1 |
| 4:139140860: | 4 | 139140860 | 0.03605825 | 7 | -0.11040444 | 0.49326313 | -0.22382462 | 8.23E-01 | intronic | SLC7A11 | . | rs112679237 | 0.0142 | 0.0215 | 0.854571 | 1 |
| 6:165514281: | 6 | 165514281 | 0.00637379 | 1 | -0.04360284 | 0.19925364 | -0.21883085 | 8.27E-01 | intergenic | MEAT6;C6orf118 | dist=278729;dist=178872 | rs117498042 | 0.0042 | 0.0043 | 0.737781 | 1 |
| 15:59229353: | 15 | 59229353 | 0.01822816 | 4 | 0.07070707 | 0.326751 | 0.21639435 | 8.29E-01 | intergenic | SLTM;RNF111 | dist=3501;dist=50512 | rs193253461 | 0.0039 | 0.0067 | 0.779464 | 1 |
| 6:98092675:T | 6 | 98092675 | 0.00759223 | 2 | -0.04473159 | 0.21551523 | -0.20755653 | 8.36E-01 | ncRNA_intronic | LOC101927314 | . | rs56224400 | 0.0082 | 0.0137 | 0.681669 | 1 |
| 12:63608466: | 12 | 63608466 | 0.00901456 | 2 | -0.04938488 | 0.24082995 | -0.20506121 | 8.38E-01 | intergenic | AVPR1A;DPY19L2 | dist=61876;dist=344227 | rs182437250 | 0.002 | 0.0033 | 0.739218 | 1 |
| 15:59404306: | 15 | 59404306 | 0.01714078 | 4 | 0.06477319 | 0.32705492 | 0.19804989 | 8.43E-01 | intronic | CCNB2 | . | rs184117160 | 0.0036 | 0.0062 | 0.821999 | 1 |
| 12:63445280: | 12 | 63445280 | 0.01001456 | 2 | -0.05165032 | 0.26703588 | -0.1934209 | 8.47E-01 | intergenic | PPM1H;AVPR1A | dist=116615;dist=91259 | rs191053292 | 0.002 | 0.0031 | 0.818462 | 1 |
| 21:35477486: | 21 | 35477486 | 0.03398058 | 7 | 0.09515843 | 0.53639433 | 0.17740386 | 8.59E-01 | UTR3 | SLC5A3 | NM_006933:c.*7832C>T | rs118183140 | 0.0133 | 0.0202 | 1 | 1 |
| 6:165499857: | 6 | 165499857 | 0.00646602 | 1 | -0.03576208 | 0.20752178 | -0.17232927 | 8.63E-01 | intergenic | MEAT6;C6orf118 | dist=264305;dist=193296 | rs148532212 | 0.0029 | 0.0035 | 0.785224 | 1 |
| 6:142611258: | 6 | 142611258 | 0.01621359 | 3 | -0.05521243 | 0.32412283 | -0.17034416 | 8.65E-01 | intergenic | VTA1;ADGRG6 | dist=69173;dist=11798 | rs72986533 | 0.0083 | 0.014 | 0.839327 | 1 |
| 3:141841196: | 3 | 141841196 | 0.00939806 | 2 | -0.04205967 | 0.26798504 | -0.15694783 | 8.75E-01 | intronic | TFDP2 | . | rs576124203 | 0.0017 | 0.0028 | 0.88872 | 1 |
| 5:91631888:A | 5 | 91631888 | 0.00402427 | 1 | 0.02563369 | 0.16856192 | 0.15207283 | 8.79E-01 | intergenic | ARRDC3-AS1;NR2F1-AS1 | dist=915356;dist=1113174 | rs183816745 | 0.0085 | 0.0153 | 0.828309 | 1 |
| 5:91530447:G | 5 | 91530447 | 0.00445631 | 1 | 0.02838568 | 0.18665843 | 0.15207283 | 8.79E-01 | intergenic | ARRDC3-AS1;NR2F1-AS1 | dist=813915;dist=1214615 | rs187236873 | 0.0085 | 0.0153 | 0.917633 | 1 |
| 5:91465647:A | 5 | 91465647 | 0.00482524 | 1 | 0.03073569 | 0.20211164 | 0.15207283 | 8.79E-01 | intergenic | ARRDC3-AS1;NR2F1-AS1 | dist=749115;dist=1279415 | rs181933850 | 0.0085 | 0.0151 | 0.993971 | 1 |
| 5:91475485:G | 5 | 91475485 | 0.00477185 | 1 | 0.03039555 | 0.19987499 | 0.15207283 | 8.79E-01 | intergenic | ARRDC3-AS1;NR2F1-AS1 | dist=758953;dist=1269577 | rs190190051 | 0.0084 | 0.0151 | 0.982918 | 1 |
| 5:91530073:C | 5 | 91530073 | 0.00445146 | 1 | 0.02835476 | 0.1864551 | 0.15207283 | 8.79E-01 | intergenic | ARRDC3-AS1;NR2F1-AS1 | dist=813541;dist=1214989 | rs182531466 | 0.0079 | 0.0143 | 0.916629 | 1 |
| 3:141864350: | 3 | 141864350 | 0.00941262 | 2 | -0.04067729 | 0.27007401 | -0.15061534 | 8.80E-01 | intronic | TFDP2 | . | rs545552231 | 0.0017 | 0.0029 | 0.904348 | 1 |
| 6:142269885: | 6 | 142269885 | 0.01472816 | 3 | -0.0499046 | 0.331489 | -0.15054679 | 8.80E-01 | intergenic | MIR4465;NMBR | dist=1264865;dist=126860 | rs142106992 | 0.0068 | 0.0116 | 0.974922 | 1 |
| 1:229804538: | 1 | 229804538 | 0.01941748 | 4 | 0.05619014 | 0.39697883 | 0.14154444 | 8.87E-01 | intergenic | URB2;GALNT2 | dist=8591;dist=388998 | rs2274996 | 0.119 | 0.0431 | 1 | 1 |
| 1:229806368: | 1 | 229806368 | 0.01941748 | 4 | 0.05619014 | 0.39697883 | 0.14154444 | 8.87E-01 | intergenic | URB2;GALNT2 | dist=10421;dist=387168 | rs2891865 | 0.0843 | 0.0424 | 1 | 1 |
| 1:229824770: | 1 | 229824770 | 0.01941748 | 4 | 0.05619014 | 0.39697883 | 0.14154444 | 8.87E-01 | intergenic | URB2;GALNT2 | dist=28823;dist=368766 | rs4562666 | 0.1094 | 0.043 | 1 | 1 |
| 1:229831331: | 1 | 229831331 | 0.01941748 | 4 | 0.05619014 | 0.39697883 | 0.14154444 | 8.87E-01 | intergenic | URB2;GALNT2 | dist=35384;dist=362205 | rs16850124 | 0.1613 | 0.0447 | 1 | 1 |
| 21:23156546: | 21 | 23156546 | 0.01758738 | 4 | 0.0449786 | 0.32176516 | 0.13978704 | 8.89E-01 | ncRNA_intronic | LINC01425 | . | rs75024143 | 0.007 | 0.0122 | 0.702913 | 1 |
| 2:52563371:C | 2 | 52563371 | 0.00220874 | 0 | -0.01305165 | 0.09516368 | -0.1371495 | 8.91E-01 | intergenic | NRXN1;MIR4431 | dist=1303697;dist=366289 | rs183378658 | 0.0017 | 0.0025 | 0.422723 | 1 |
| 1:229807492: | 1 | 229807492 | 0.01933981 | 4 | 0.05291944 | 0.39489101 | 0.13401025 | 8.93E-01 | intergenic | URB2;GALNT2 | dist=11545;dist=386044 | rs2385790 | 0.0661 | 0.0417 | 0.993425 | 1 |
| 2:52527354:T | 2 | 52527354 | 0.00207767 | 0 | -0.01200578 | 0.08995596 | -0.13346282 | 8.94E-01 | intergenic | NRXN1;MIR4431 | dist=1267680;dist=402306 | rs181193202 | 0.0018 | 0.0027 | 0.401439 | 1 |
| 1:229812357: | 1 | 229812357 | 0.01934466 | 4 | 0.05261465 | 0.39487567 | 0.13324359 | 8.94E-01 | intergenic | URB2;GALNT2 | dist=16410;dist=381179 | rs12024557 | 0.1093 | 0.0426 | 0.993171 | 1 |
| 1:229804646: | 1 | 229804646 | 0.01936408 | 4 | 0.05234459 | 0.39487456 | 0.13256005 | 8.95E-01 | intergenic | URB2;GALNT2 | dist=8699;dist=388890 | rs2274997 | 0.1186 | 0.043 | 0.992157 | 1 |
| 2:67565909:A | 2 | 67565909 | 0.00786408 | 2 | 0.02790624 | 0.21524991 | 0.12964579 | 8.97E-01 | intergenic | LOC102800447;ETAA1 | dist=49533;dist=58533 | rs11690187 | 0.0032 | 0.0047 | 0.837388 | 1 |
| 21:43276916: | 21 | 43276916 | 0.00510194 | 1 | -0.02342163 | 0.20678399 | -0.11326615 | 9.10E-01 | intronic | PRDM15 | . | rs150539922 | 0.0026 | 0.0035 | 0.918546 | 1 |
| 2:53085778:G | 2 | 53085778 | 0.00436408 | 1 | -0.02145063 | 0.19447952 | -0.11029764 | 9.12E-01 | intergenic | MIR4431;ASB3 | dist=156025;dist=811339 | rs190193113 | 0.0016 | 0.0024 | 0.894549 | 1 |
| 1:103633635: | 1 | 103633635 | 0.01638835 | 3 | 0.03086612 | 0.28612599 | 0.10787596 | 9.14E-01 | intergenic | COL11A1;LOC101928436 | dist=59583;dist=327066 | rs111928960 | 0.015 | 0.0213 | 0.591355 | 1 |
| 22:40528090: | 22 | 40528090 | 0.00556311 | 1 | -0.01974371 | 0.19432826 | -0.1015998 | 9.19E-01 | intronic | TNRC6B | . | rs541680196 | 0.0007 | 0.0013 | 0.747052 | 1 |
| 1:103419168: | 1 | 103419168 | 0.01709709 | 4 | -0.02811392 | 0.29796789 | -0.09435217 | 9.25E-01 | intronic | COL11A1 | . | rs114413507 | 0.0156 | 0.0224 | 0.530043 | 1 |
| 3:164285621: | 3 | 164285621 | 0.00413592 | 1 | 0.01403123 | 0.17131386 | 0.08190364 | 9.35E-01 | intergenic | MIR1263;LINC01324 | dist=396277;dist=146262 | rs187047882 | 0.0026 | 0.004 | 0.827625 | 1 |
| 3:42711221:A | 3 | 42711221 | 0.01973786 | 4 | -0.03198562 | 0.40445891 | -0.0790825 | 9.37E-01 | intergenic | ZBTB47;KLHL40 | dist=2149;dist=15790 | rs73085348 | 0.0132 | 0.0192 | 0.962032 | 1 |
| 22:40620530: | 22 | 40620530 | 0.00540777 | 1 | 0.01375367 | 0.20531176 | 0.0669892 | 9.47E-01 | intronic | TNRC6B | . | rs148998974 | 0.0007 | 0.0013 | 0.851831 | 1 |
| 7:146865072: | 7 | 146865072 | 0.00787379 | 2 | 0.01517629 | 0.24014534 | 0.06319627 | 9.50E-01 | intronic | CNTNAP2 | . | rs536023430 | 0.0025 | 0.0031 | 0.812462 | 1 |
| 22:40604439: | 22 | 40604439 | 0.00533495 | 1 | 0.01161262 | 0.20487662 | 0.05668105 | 9.55E-01 | intronic | TNRC6B | . | rs141127122 | 0.0004 | 0.0006 | 0.859782 | 1 |
| 1:102961882: | 1 | 102961882 | 0.01406311 | 3 | 0.01428562 | 0.25538907 | 0.05593671 | 9.55E-01 | intergenic | OLFM3;COL11A1 | dist=499092;dist=380141 | rs77180278 | 0.0151 | 0.0218 | 0.587391 | 1 |
| 2:65875930:C | 2 | 65875930 | 0.00468447 | 1 | 0.01016422 | 0.18533674 | 0.05484192 | 9.56E-01 | intergenic | SPRED2;MIR4778 | dist=216274;dist=709451 | rs528288879 | 0.0043 | 0.0063 | 0.81586 | 1 |
| 22:40631476: | 22 | 40631476 | 0.00598544 | 1 | -0.00987839 | 0.20679901 | -0.04776807 | 9.62E-01 | intronic | TNRC6B | . | rs555040883 | 0.0003 | 0.0005 | 0.785825 | 1 |
| 1:103220360: | 1 | 103220360 | 0.01623787 | 3 | -0.013522 | 0.29931658 | -0.04517624 | 9.64E-01 | intergenic | OLFM3;COL11A1 | dist=757570;dist=121663 | rs112351653 | 0.0156 | 0.0224 | 0.659632 | 1 |
| 22:40594781: | 22 | 40594781 | 0.00522816 | 1 | 0.00894748 | 0.19987571 | 0.04476521 | 9.64E-01 | intronic | TNRC6B | . | rs185139807 | 0.0007 | 0.0013 | 0.834478 | 1 |
| 1:103472916: | 1 | 103472916 | 0.01778641 | 4 | 0.01007671 | 0.31422553 | 0.03206841 | 9.74E-01 | intronic | COL11A1 | . | rs116672066 | 0.0151 | 0.0219 | 0.573939 | 1 |
| 6:85678832:A | 6 | 85678832 | 0.00214078 | 0 | 0.00222023 | 0.08048978 | 0.02758402 | 9.78E-01 | intergenic | TBX18;NT5E | dist=204878;dist=480470 | rs571986619 | 0.0004 | 0.0005 | 0.356816 | 1 |
| 10:10291848 | 10 | 102918486 | 0.02915534 | 6 | 0.01274117 | 0.49073343 | 0.02596352 | 9.79E-01 | intergenic | TLX1NB;LINC01514 | dist=17463;dist=17593 | rs117913371 | 0.0141 | 0.0239 | 0.987133 | 1 |
| 9:135266715: | 9 | 135266715 | 0.00491262 | 1 | 0.00344492 | 0.20028125 | 0.01720042 | 9.86E-01 | intronic | TTF1 | . | rs78296164 | 0.0047 | 0.0078 | 0.98038 | 1 |
| 1:229834050: | 1 | 229834050 | 0.01748544 | 4 | -0.00570229 | 0.36438542 | -0.01564905 | 9.88E-01 | intergenic | URB2;GALNT2 | dist=38103;dist=359486 | rs12045643 | 0.1113 | 0.0429 | 0.928351 | 1 |

**Supplementary Table S7. Independent Replication**

Florida-1 cohort, 12M quantitative trait (QT) data, comparing to discovery 12M QT

| | |
|---|---|
| Threshold for replicative significance: | 5.00E-02 |
| Total SNPs found: | 1 |
| Total risk loci found: | 1 |

NA: not applicable

| rsid | chr | pos_37 | REF | ALT | caf | MAC | Est | Est.SE | Score.pval | caf.sch | Est.sch | Est.SE.sch | pval.sch | adj pval SNPs | adj pval loci | Closest gene | Prioritized gene |
|---|---|---|---|---|---|---|---|---|---|---|---|---|---|---|---|---|---|
| rs9636964 | 21 | 41304765 | A | G | 0.884965831 | 115 | -4.254319226 | 0.827443714 | 2.73E-07 | 0.06 | 2.37989208 | 2.5889182 | 3.58E-01 | 3.58E-01 | 3.58E-01 | PCP4 | PCP4 |
| rs142934021 | 5 | 52662672 | G | A | 0.007972665 | 7 | 18.82471499 | 3.318202936 | 1.40E-08 | NA | NA | NA | NA | | | | |
| rs138164904 | 16 | 6858239 | C | T | 0.003416856 | 3 | 23.89264096 | 4.336966899 | 3.61E-08 | NA | NA | NA | NA | | | | |
| rs138138661 | 3 | 147657814 | C | T | 0.004555809 | 4 | 23.13474571 | 4.256958355 | 5.49E-08 | NA | NA | NA | NA | | | | |
| rs147669485 | 18 | 26603650 | G | T | 0.003416856 | 4 | 22.11076388 | 4.149231981 | 9.88E-08 | NA | NA | NA | NA | | | | |
| rs148997617 | 3 | 147755576 | C | T | 0.004555809 | 4 | 20.87742767 | 3.926489017 | 1.05E-07 | NA | NA | NA | NA | | | | |
| rs74572772 | 22 | 34492649 | A | G | 0.018223235 | 16 | 10.22251833 | 1.941267739 | 1.40E-07 | NA | NA | NA | NA | | | | |
| rs148157126 | 20 | 30948839 | C | T | 0.004555809 | 5 | 19.78650011 | 3.760421027 | 1.43E-07 | NA | NA | NA | NA | | | | |
| rs80019988 | 22 | 34493647 | G | A | 0.018223235 | 16 | 10.06233062 | 1.916654069 | 1.52E-07 | NA | NA | NA | NA | | | | |
| rs186792608 | 2 | 43096699 | A | G | 0.003416856 | 3 | 22.45124992 | 4.281728413 | 1.58E-07 | NA | NA | NA | NA | | | | |
| rs9974985 | 21 | 41307573 | G | A | 0.886104784 | 114 | -4.345939521 | 0.829242821 | 1.60E-07 | NA | NA | NA | NA | | | | |
| rs113154814 | 2 | 68289497 | T | C | 0.005694761 | 5 | 19.83101879 | 3.789289395 | 1.66E-07 | NA | NA | NA | NA | | | | |
| rs1005412 | 21 | 41308948 | A | G | 0.887243736 | 112 | -4.452109624 | 0.852371384 | 1.76E-07 | NA | NA | NA | NA | | | | |
| rs386818616 | 21 | 41308948 | A | G | 0.887243736 | 112 | -4.452109624 | 0.852371384 | 1.76E-07 | NA | NA | NA | NA | | | | |
| rs193041547 | 20 | 30771880 | T | C | 0.004555809 | 4 | 19.67913585 | 3.770811462 | 1.80E-07 | NA | NA | NA | NA | | | | |
| rs77297738 | 22 | 34473543 | C | T | 0.018223235 | 16 | 10.24805718 | 1.971675922 | 2.02E-07 | NA | NA | NA | NA | | | | |
| rs146249289 | 20 | 30682231 | C | T | 0.004555809 | 4 | 19.6109527 | 3.773060354 | 2.02E-07 | NA | NA | NA | NA | | | | |
| rs76356799 | 3 | 179593768 | G | A | 0.004555809 | 4 | 19.28670071 | 3.760393191 | 2.91E-07 | NA | NA | NA | NA | | | | |
| rs7275595 | 21 | 41307923 | G | A | 0.888382688 | 113 | -4.260975379 | 0.834078676 | 3.25E-07 | NA | NA | NA | NA | | | | |
| rs192855100 | 20 | 31108741 | G | A | 0.004555809 | 4 | 20.19860967 | 3.99111726 | 4.17E-07 | NA | NA | NA | NA | | | | |
| rs2427460 | 20 | 61590782 | T | C | 0.486332574 | 420 | -2.574043965 | 0.513258918 | 5.30E-07 | NA | NA | NA | NA | | | | |
| rs562831582 | 19 | 51234310 | A | C | 0.003416856 | 4 | 21.08565484 | 4.216248377 | 5.70E-07 | NA | NA | NA | NA | | | | |
| rs149859280 | 20 | 30678170 | C | T | 0.004555809 | 5 | 18.30577068 | 3.660758895 | 5.72E-07 | NA | NA | NA | NA | | | | |
| rs138733283 | 20 | 30675687 | C | T | 0.004555809 | 5 | 18.27447439 | 3.658301378 | 5.87E-07 | NA | NA | NA | NA | | | | |
| rs80212581 | 16 | 6412967 | C | T | 0.003416856 | 3 | 22.27620028 | 4.472373329 | 6.33E-07 | NA | NA | NA | NA | | | | |
| rs148248743 | 3 | 136134595 | C | T | 0.002277904 | 3 | 27.15485873 | 5.463527255 | 6.69E-07 | NA | NA | NA | NA | | | | |
| rs188353596 | 17 | 46078602 | C | T | 0.003416856 | 3 | 21.80579474 | 4.391419406 | 6.85E-07 | NA | NA | NA | NA | | | | |
| rs113791989 | 13 | 61918888 | G | A | 0.002277904 | 3 | 25.20495822 | 5.084027893 | 7.13E-07 | NA | NA | NA | NA | | | | |
| rs56324718 | 15 | 40280073 | G | A | 0.010250569 | 9 | 12.5117374 | 2.5288888 | 7.52E-07 | NA | NA | NA | NA | | | | |
| rs191298981 | 2 | 68110046 | C | T | 0.004555809 | 3 | 24.00787141 | 4.857032236 | 7.70E-07 | NA | NA | NA | NA | | | | |
| rs146919974 | 2 | 115252863 | T | C | 0.004555809 | 5 | 19.72804656 | 3.995542316 | 7.91E-07 | NA | NA | NA | NA | | | | |
| rs9305683 | 21 | 41305720 | G | A | 0.88952164 | 112 | -4.116367918 | 0.836343627 | 8.57E-07 | NA | NA | NA | NA | | | | |
| rs145791959 | 20 | 30722429 | G | A | 0.005694761 | 5 | 17.90919529 | 3.640341121 | 8.67E-07 | NA | NA | NA | NA | | | | |
| rs142956968 | 10 | 23490165 | C | T | 0.01594533 | 14 | 10.41679921 | 2.119090829 | 8.85E-07 | NA | NA | NA | NA | | | | |
| rs532269430 | 13 | 74054452 | T | C | 0.004555809 | 5 | 18.61737516 | 3.796300403 | 9.39E-07 | NA | NA | NA | NA | | | | |
| rs113063005 | 4 | 23509067 | T | C | 0.006833713 | 7 | 16.61762999 | 3.391132089 | 9.57E-07 | NA | NA | NA | NA | | | | |
| rs147944608 | 10 | 110423709 | C | T | 0.012528474 | 11 | 11.22052434 | 2.292291613 | 9.84E-07 | NA | NA | NA | NA | | | | |
| rs2606194 | 17 | 77210823 | A | G | 0.948747153 | 44 | -5.695697885 | 1.164357735 | 1.00E-06 | NA | NA | NA | NA | | | | |
| rs113537164 | 2 | 68234930 | A | C | 0.004555809 | 4 | 20.38904135 | 4.17097432 | 1.02E-06 | NA | NA | NA | NA | | | | |
| rs138055631 | 20 | 30780644 | G | A | 0.005694761 | 5 | 17.66477622 | 3.620049149 | 1.06E-06 | NA | NA | NA | NA | | | | |
| rs566724618 | 13 | 23463839 | C | T | 0.003416856 | 3 | 22.34337048 | 4.580616053 | 1.07E-06 | NA | NA | NA | NA | | | | |
| rs145764464 | 10 | 23601847 | G | A | 0.017084282 | 15 | 9.596730604 | 1.969763479 | 1.10E-06 | NA | NA | NA | NA | | | | |
| rs9981433 | 21 | 41309565 | G | T | 0.881548975 | 116 | -4.122012439 | 0.84715422 | 1.14E-06 | NA | NA | NA | NA | | | | |
| rs139816293 | 20 | 30921343 | C | T | 0.005694761 | 6 | 17.13842475 | 3.525372234 | 1.17E-06 | NA | NA | NA | NA | | | | |
| rs117699122 | 12 | 560262 | T | C | 0.003416856 | 4 | 21.42382563 | 4.40952328 | 1.18E-06 | NA | NA | NA | NA | | | | |
| rs529110230 | 7 | 117175907 | T | C | 0.003416856 | 3 | 20.85836332 | 4.302350877 | 1.25E-06 | NA | NA | NA | NA | | | | |
| rs12114488 | 8 | 62672723 | G | A | 0.30523918 | 266 | 2.623079144 | 0.54257702 | 1.33E-06 | NA | NA | NA | NA | | | | |
| rs182868205 | 3 | 177712448 | C | T | 0.003416856 | 3 | 22.85443031 | 4.732156621 | 1.37E-06 | NA | NA | NA | NA | | | | |

| | | | | | | | | | | | | | |
|---|---|---|---|---|---|---|---|---|---|---|---|---|---|
| rs144138711 | 9 | 99922419 | T | C | 0.045558087 | 40 | 6.085471305 | 1.260919179 | 1.39E-06 | NA | NA | NA | NA |
| rs372194899 | 13 | 74357510 | A | G | 0.003416856 | 3 | 21.30121009 | 4.41516106 | 1.40E-06 | NA | NA | NA | NA |
| rs368187808 | 13 | 74357508 | A | C | 0.003416856 | 3 | 21.2991251 | 4.415016369 | 1.41E-06 | NA | NA | NA | NA |
| rs145804766 | 1 | 202122499 | C | T | 0.010250569 | 9 | 12.58644653 | 2.611189618 | 1.43E-06 | NA | NA | NA | NA |
| rs182211730 | 7 | 116003615 | G | A | 0.004555809 | 4 | 18.7746033 | 3.897198104 | 1.45E-06 | NA | NA | NA | NA |
| rs558784715 | 7 | 117043294 | A | G | 0.003416856 | 3 | 20.75917871 | 4.316894751 | 1.52E-06 | NA | NA | NA | NA |
| rs189709453 | 3 | 177574308 | G | A | 0.003416856 | 3 | 23.20099361 | 4.829543736 | 1.56E-06 | NA | NA | NA | NA |
| rs142721557 | 7 | 117218328 | A | C | 0.003416856 | 3 | 20.73299749 | 4.322458941 | 1.61E-06 | NA | NA | NA | NA |
| rs576047962 | 7 | 117109021 | A | G | 0.003416856 | 3 | 20.72615487 | 4.321401824 | 1.62E-06 | NA | NA | NA | NA |
| rs142215699 | 7 | 117199874 | T | C | 0.003416856 | 3 | 20.72922868 | 4.322453614 | 1.62E-06 | NA | NA | NA | NA |
| rs143281973 | 7 | 116007996 | C | T | 0.004555809 | 4 | 18.27507784 | 3.810981978 | 1.62E-06 | NA | NA | NA | NA |
| rs17127656 | 1 | 65943471 | C | T | 0.055808656 | 49 | 5.070465376 | 1.057806414 | 1.64E-06 | NA | NA | NA | NA |
| rs201355675 | 7 | 117225781 | G | A | 0.003416856 | 3 | 20.71812396 | 4.322467219 | 1.64E-06 | NA | NA | NA | NA |
| rs186767531 | 3 | 177639777 | T | C | 0.003416856 | 3 | 23.07499768 | 4.81664642 | 1.66E-06 | NA | NA | NA | NA |
| rs7518849 | 1 | 65948791 | T | C | 0.055808656 | 49 | 5.074569555 | 1.059644692 | 1.68E-06 | NA | NA | NA | NA |
| rs181132315 | 4 | 129647369 | C | T | 0.003416856 | 3 | 22.13246791 | 4.62266164 | 1.69E-06 | NA | NA | NA | NA |
| rs75082290 | 2 | 67831153 | T | G | 0.078587699 | 69 | 4.693953865 | 0.98072733 | 1.70E-06 | NA | NA | NA | NA |
| rs145421321 | 20 | 30862126 | C | T | 0.005694761 | 6 | 17.02126111 | 3.558296484 | 1.72E-06 | NA | NA | NA | NA |
| rs143432612 | 20 | 30892775 | C | T | 0.005694761 | 6 | 16.93872319 | 3.543181539 | 1.75E-06 | NA | NA | NA | NA |
| rs140276610 | 16 | 6433735 | C | T | 0.003416856 | 4 | 22.18065773 | 4.640624008 | 1.76E-06 | NA | NA | NA | NA |
| rs559152067 | 18 | 66029612 | G | A | 0.002277904 | 3 | 24.74436915 | 5.183438622 | 1.81E-06 | NA | NA | NA | NA |
| rs147221953 | 13 | 23485722 | G | A | 0.003416856 | 3 | 22.58096755 | 4.736033596 | 1.86E-06 | NA | NA | NA | NA |
| rs556274646 | 11 | 439012 | C | T | 0.010250569 | 9 | 12.89145917 | 2.708477277 | 1.94E-06 | NA | NA | NA | NA |
| rs190251199 | 14 | 105590577 | T | C | 0.003416856 | 3 | 22.63850613 | 4.760438165 | 1.98E-06 | NA | NA | NA | NA |
| rs188993522 | 7 | 117274731 | T | C | 0.003416856 | 3 | 21.29848267 | 4.478672657 | 1.98E-06 | NA | NA | NA | NA |
| rs568321148 | 2 | 29870348 | T | G | 0.003416856 | 3 | 20.74002026 | 4.366527169 | 2.04E-06 | NA | NA | NA | NA |
| rs148815783 | 11 | 1013992 | C | T | 0.007972665 | 8 | 13.59803132 | 2.864717097 | 2.07E-06 | NA | NA | NA | NA |
| rs76220567 | 2 | 13059945 | T | C | 0.003416856 | 4 | 20.68288684 | 4.357762163 | 2.07E-06 | NA | NA | NA | NA |
| rs35145334 | 1 | 23465122 | A | G | 0.017084282 | 14 | 10.65526559 | 2.246207918 | 2.10E-06 | NA | NA | NA | NA |
| rs11579567 | 1 | 65957141 | C | A | 0.055808656 | 49 | 5.029737931 | 1.060973792 | 2.13E-06 | NA | NA | NA | NA |
| rs114067899 | 1 | 85191044 | G | A | 0.007972665 | 7 | 13.77606734 | 2.908363649 | 2.17E-06 | NA | NA | NA | NA |
| rs56821264 | 6 | 148703901 | C | T | 0.006833713 | 6 | 14.75092126 | 3.114195656 | 2.17E-06 | NA | NA | NA | NA |
| rs917335559 | 6 | 148703901 | C | T | 0.006833713 | 6 | 14.75092126 | 3.114195656 | 2.17E-06 | NA | NA | NA | NA |
| rs536781978 | 2 | 29733681 | A | G | 0.003416856 | 3 | 20.55618824 | 4.342580992 | 2.21E-06 | NA | NA | NA | NA |
| rs147267707 | 4 | 43026558 | G | A | 0.004555809 | 4 | 18.00943854 | 3.808184082 | 2.25E-06 | NA | NA | NA | NA |
| rs77618729 | 4 | 43050968 | T | C | 0.004555809 | 4 | 18.01301525 | 3.809141731 | 2.26E-06 | NA | NA | NA | NA |
| rs547186621 | 20 | 6106741 | A | G | 0.003416856 | 4 | 21.55108289 | 4.569088099 | 2.40E-06 | NA | NA | NA | NA |
| rs200198574 | 20 | 30946654 | G | A | 0.005694761 | 6 | 16.52827354 | 3.50702674 | 2.44E-06 | NA | NA | NA | NA |
| rs532464521 | 4 | 43571039 | T | C | 0.004555809 | 4 | 17.98955275 | 3.818204376 | 2.46E-06 | NA | NA | NA | NA |
| rs79252854 | 3 | 157523157 | C | T | 0.004555809 | 5 | 17.54643357 | 3.72541417 | 2.48E-06 | NA | NA | NA | NA |
| rs79935606 | 2 | 177338635 | T | C | 0.005694761 | 5 | 16.33968329 | 3.473769742 | 2.55E-06 | NA | NA | NA | NA |
| rs556435089 | 4 | 43551033 | G | A | 0.004555809 | 4 | 17.93907904 | 3.817721456 | 2.62E-06 | NA | NA | NA | NA |
| rs62192733 | 20 | 1928157 | A | G | 0.006833713 | 7 | 15.74461797 | 3.358665632 | 2.76E-06 | NA | NA | NA | NA |
| rs7534177 | 1 | 65965720 | A | G | 0.056947608 | 49 | 4.961233778 | 1.059861601 | 2.85E-06 | NA | NA | NA | NA |
| rs139390630 | 1 | 84866196 | G | A | 0.011389522 | 13 | 11.69759713 | 2.499486296 | 2.87E-06 | NA | NA | NA | NA |
| rs149706477 | 3 | 66148228 | A | G | 0.003416856 | 3 | 20.98286869 | 4.484253102 | 2.88E-06 | NA | NA | NA | NA |
| rs189094663 | 4 | 11623450 | G | A | 0.002277904 | 3 | 23.895416 | 5.108227082 | 2.90E-06 | NA | NA | NA | NA |
| rs181182636 | 7 | 115654558 | A | G | 0.003416856 | 3 | 21.94726388 | 4.692447434 | 2.91E-06 | NA | NA | NA | NA |
| rs76098744 | 1 | 112349372 | C | T | 0.01594533 | 14 | 9.568565696 | 2.046351926 | 2.93E-06 | NA | NA | NA | NA |
| rs4896997 | 6 | 148683924 | C | T | 0.006833713 | 6 | 14.43209229 | 3.091883755 | 3.05E-06 | NA | NA | NA | NA |
| rs17078283 | 6 | 148696920 | C | T | 0.006833713 | 6 | 14.43028205 | 3.091804795 | 3.05E-06 | NA | NA | NA | NA |
| rs73202425 | 7 | 109155716 | T | C | 0.012528474 | 12 | 10.76184396 | 2.305821662 | 3.05E-06 | NA | NA | NA | NA |
| rs4131286 | 6 | 148688963 | G | T | 0.006833713 | 6 | 14.42846716 | 3.091725323 | 3.06E-06 | NA | NA | NA | NA |
| rs74683551 | 1 | 112354418 | G | A | 0.01594533 | 14 | 9.547228317 | 2.046406793 | 3.08E-06 | NA | NA | NA | NA |
| rs139055031 | 3 | 158586849 | T | C | 0.005694761 | 6 | 16.28537606 | 3.4955337 | 3.18E-06 | NA | NA | NA | NA |
| rs138873576 | 7 | 116025359 | G | T | 0.004555809 | 4 | 17.61517106 | 3.78331079 | 3.22E-06 | NA | NA | NA | NA |
| rs148483098 | 3 | 157382051 | C | A | 0.005694761 | 5 | 15.73578443 | 3.382110237 | 3.28E-06 | NA | NA | NA | NA |
| rs187281112 | 15 | 33113387 | C | T | 0.005694761 | 5 | 14.73433563 | 3.167936491 | 3.30E-06 | NA | NA | NA | NA |

| | | | | | | | | | | | | | |
|---|---|---|---|---|---|---|---|---|---|---|---|---|---|
| rs1162141034 | 2 | 67825685 | C | T | 0.079726651 | 71 | 4.559914114 | 0.980447215 | 3.31E-06 | NA | NA | NA | NA |
| rs146465493 | 2 | 67825685 | C | T | 0.079726651 | 71 | 4.559914114 | 0.980447215 | 3.31E-06 | NA | NA | NA | NA |
| rs2902021 | 2 | 67825685 | C | T | 0.079726651 | 71 | 4.559914114 | 0.980447215 | 3.31E-06 | NA | NA | NA | NA |
| rs189912648 | 5 | 134765439 | C | T | 0.003416856 | 4 | 19.8604449 | 4.271665199 | 3.33E-06 | NA | NA | NA | NA |
| rs557276277 | 4 | 19013698 | T | C | 0.005694761 | 5 | 19.01463443 | 4.091376275 | 3.36E-06 | NA | NA | NA | NA |
| rs147869916 | 1 | 227307652 | A | G | 0.005694761 | 5 | 17.59645507 | 3.78625881 | 3.36E-06 | NA | NA | NA | NA |
| rs367732718 | 19 | 35918004 | G | A | 0.002277904 | 3 | 23.87849231 | 5.139957519 | 3.39E-06 | NA | NA | NA | NA |
| rs62515436 | 8 | 57141203 | G | T | 0.017084282 | 15 | 9.275459023 | 1.998048118 | 3.45E-06 | NA | NA | NA | NA |
| rs189234695 | 3 | 147658158 | T | C | 0.003416856 | 3 | 23.96971341 | 5.165421292 | 3.48E-06 | NA | NA | NA | NA |
| rs186893139 | 7 | 116096970 | C | T | 0.004555809 | 4 | 18.60259803 | 4.012583835 | 3.55E-06 | NA | NA | NA | NA |
| rs73384619 | 6 | 20458179 | C | T | 0.022779043 | 21 | 8.391670204 | 1.812413894 | 3.65E-06 | NA | NA | NA | NA |
| rs114280794 | 3 | 132842203 | G | A | 0.01594533 | 14 | 8.762968775 | 1.896320161 | 3.82E-06 | NA | NA | NA | NA |
| rs112148840 | 17 | 66233774 | C | T | 0.002277904 | 3 | 24.02410753 | 5.199447601 | 3.83E-06 | NA | NA | NA | NA |
| rs147455971 | 4 | 140931712 | T | C | 0.007972665 | 5 | 17.21601424 | 3.727930571 | 3.87E-06 | NA | NA | NA | NA |
| rs144788248 | 3 | 19586745 | T | C | 0.011389522 | 10 | 11.90862374 | 2.578791237 | 3.88E-06 | NA | NA | NA | NA |
| rs111512950 | 5 | 153680427 | C | T | 0.009111617 | 8 | 13.56744305 | 2.945984928 | 4.12E-06 | NA | NA | NA | NA |
| rs149616342 | 12 | 99178813 | C | T | 0.011389522 | 10 | 11.2978214 | 2.454656997 | 4.17E-06 | NA | NA | NA | NA |
| rs17543620 | 4 | 169934725 | T | C | 0.050113895 | 44 | 5.427967555 | 1.179820972 | 4.21E-06 | NA | NA | NA | NA |
| rs190354334 | 7 | 116075846 | G | A | 0.004555809 | 5 | 17.33925809 | 3.7718663 | 4.29E-06 | NA | NA | NA | NA |
| rs17615362 | 4 | 169934087 | G | A | 0.050113895 | 44 | 5.419744721 | 1.179184251 | 4.30E-06 | NA | NA | NA | NA |
| rs117591241 | 3 | 190584679 | A | G | 0.003416856 | 4 | 19.90140277 | 4.333313893 | 4.38E-06 | NA | NA | NA | NA |
| rs1472243465 | 3 | 190584679 | A | G | 0.003416856 | 4 | 19.90140277 | 4.333313893 | 4.38E-06 | NA | NA | NA | NA |
| rs61918041 | 12 | 6681868 | C | T | 0.023917995 | 21 | 7.972646426 | 1.736360556 | 4.40E-06 | NA | NA | NA | NA |
| rs113244573 | 12 | 6684385 | G | A | 0.023917995 | 21 | 7.970957457 | 1.737661556 | 4.49E-06 | NA | NA | NA | NA |
| rs62515405 | 8 | 57055978 | G | A | 0.023917995 | 21 | 8.076912036 | 1.76428755 | 4.69E-06 | NA | NA | NA | NA |
| rs116897913 | 17 | 16839162 | C | T | 0.003416856 | 3 | 20.13967647 | 4.402942883 | 4.78E-06 | NA | NA | NA | NA |
| rs143686474 | 7 | 16291999 | A | C | 0.012528474 | 11 | 10.85314984 | 2.375131845 | 4.89E-06 | NA | NA | NA | NA |
| rs143669489 | 3 | 159392885 | G | A | 0.003416856 | 3 | 24.22531986 | 5.304006744 | 4.94E-06 | NA | NA | NA | NA |

**Supplementary Table S7. Independent Replication**

Florida-1 cohort, 12M quantitative trait (QT) data, comparing to discovery 3M QT

| | |
|---|---|
| Threshold for replicative significance = | 5.00E-02 |
| Total SNPs found: | 7 |
| Total risk loci found: | 4 |

NA: not applicable

| MAF ALPHA | rsid | chr | pos_37 | REF | ALT | caf | MAC | Est | Est.SE | Score.pval | caf.sch | Est.sch | Est.SE.sch | pval.sch | adj pval SNPs | adj pval loci | Closest gene | Prioritized gene |
|---|---|---|---|---|---|---|---|---|---|---|---|---|---|---|---|---|---|---|
| 0.345139 | rs2375537 | 8 | 55619508 | C | T | 0.332541568 | 280 | 2.23876848 | 0.44938849 | 6.30E-07 | 0.37 | 1.57403151 | 1.34444327 | 2.42E-01 | 1.00E+00 | 9.67E-01 | RP1 | SOX17 |
| 0.042426 | rs2327968 | 20 | 15813491 | C | T | 0.024940618 | 21 | 6.15899856 | 1.34390809 | 4.59E-06 | 0.05 | 3.76518574 | 2.97243008 | 2.05E-01 | 1.00E+00 | 8.21E-01 | MACROD2 | MACROD2 |
| 0.036625 | rs2876414 | 20 | 15813704 | G | T | 0.022565321 | 19 | 6.81910907 | 1.48530907 | 4.41E-06 | 0.05 | 3.76518574 | 2.97243008 | 2.05E-01 | 1.00E+00 | 8.21E-01 | MACROD2 | MACROD2 |
| 0.365421 | rs423841 | 8 | 55556069 | G | A | 0.662707838 | 291 | -2.1495726 | 0.45846846 | 2.75E-06 | 0.62 | -1.324842 | 1.28913487 | 3.04E-01 | 1.00E+00 | 1.00E+00 | RP1 | SOX17 |
| 0.096148 | rs2274997 | 1 | 229804646 | A | G | 0.041567696 | 35 | 5.58731228 | 1.1238603 | 6.64E-07 | 0.05 | 2.30579163 | 3.86926843 | 5.51E-01 | 1.00E+00 | 1.00E+00 | URB2 | URB2 |
| 0.283534 | rs9643828 | 8 | 55529073 | C | T | 0.67695962 | 279 | -2.4236294 | 0.46846911 | 2.30E-07 | 0.69 | -0.6795104 | 1.3833319 | 6.23E-01 | 1.00E+00 | 1.00E+00 | RP1 | SOX17 |
| 0.097988 | rs12315614 | 12 | 64920957 | C | A | 0.076009501 | 64 | 3.77710662 | 0.8201959 | 4.12E-06 | 0.08 | -0.1411232 | 2.16890982 | 9.48E-01 | 1.00E+00 | 1.00E+00 | TBK1 | TBK1 |
| | rs113063005 | 4 | 23509067 | T | C | 0.005938242 | 6 | 21.2073894 | 3.15520096 | 1.80E-11 | NA | NA | NA | NA | | | | |
| | rs142106992 | 6 | 142269885 | C | A | 0.003562945 | 4 | 22.8345886 | 3.6758699 | 5.23E-10 | NA | NA | NA | NA | | | | |
| | rs181217257 | 2 | 239989119 | C | T | 0.004750594 | 4 | 21.5677368 | 3.47296989 | 5.29E-10 | NA | NA | NA | NA | | | | |
| | rs188076929 | 2 | 239993719 | T | C | 0.004750594 | 4 | 19.9775614 | 3.27246746 | 1.03E-09 | NA | NA | NA | NA | | | | |
| | rs117998251 | 10 | 13497976 | C | T | 0.003562945 | 3 | 22.9543311 | 3.77480488 | 1.19E-09 | NA | NA | NA | NA | | | | |
| | rs184458518 | 10 | 13471195 | T | G | 0.003562945 | 3 | 22.8015789 | 3.77123942 | 1.48E-09 | NA | NA | NA | NA | | | | |
| | rs184425183 | 10 | 13457520 | A | G | 0.003562945 | 3 | 22.6683174 | 3.77549648 | 1.92E-09 | NA | NA | NA | NA | | | | |
| | rs148153037 | 6 | 167501386 | G | A | 0.008313539 | 8 | 14.3359246 | 2.42128102 | 3.20E-09 | NA | NA | NA | NA | | | | |
| | rs111285015 | 19 | 23123198 | G | A | 0.003562945 | 3 | 27.3347642 | 4.62083983 | 3.31E-09 | NA | NA | NA | NA | | | | |
| | rs111928960 | 1 | 103633635 | G | A | 0.028503563 | 22 | 8.60257208 | 1.45827516 | 3.65E-09 | NA | NA | NA | NA | | | | |
| | rs187520610 | 2 | 53360041 | G | A | 0.004750594 | 4 | 18.8010396 | 3.19364595 | 3.93E-09 | NA | NA | NA | NA | | | | |
| | rs545428520 | 5 | 167821266 | T | C | 0.003562945 | 4 | 21.566035 | 3.66407988 | 3.96E-09 | NA | NA | NA | NA | | | | |
| | rs116672066 | 1 | 103472916 | G | A | 0.027315914 | 23 | 8.04769476 | 1.37187844 | 4.46E-09 | NA | NA | NA | NA | | | | |
| | rs192134381 | 21 | 23450714 | T | C | 0.003562945 | 3 | 21.788812 | 3.73125357 | 5.23E-09 | NA | NA | NA | NA | | | | |
| | rs397836601 | 21 | 23450714 | T | C | 0.003562945 | 3 | 21.788812 | 3.73125357 | 5.23E-09 | NA | NA | NA | NA | | | | |
| | rs150586237 | 6 | 24491348 | C | T | 0.003562945 | 3 | 22.0461443 | 3.78094793 | 5.51E-09 | NA | NA | NA | NA | | | | |
| | rs528404963 | 5 | 167852025 | T | C | 0.003562945 | 4 | 21.1393977 | 3.65303517 | 7.17E-09 | NA | NA | NA | NA | | | | |
| | rs76526501 | 7 | 18431110 | G | A | 0.007125891 | 6 | 14.8397312 | 2.5778114 | 8.58E-09 | NA | NA | NA | NA | | | | |
| | rs74455595 | 7 | 18431784 | A | G | 0.007125891 | 6 | 14.8397312 | 2.5778114 | 8.58E-09 | NA | NA | NA | NA | | | | |
| | rs75090694 | 7 | 18447436 | A | G | 0.008313539 | 7 | 13.8314005 | 2.41427081 | 1.01E-08 | NA | NA | NA | NA | | | | |
| | rs575473987 | 8 | 5577494 | C | T | 0.003562945 | 3 | 22.2248653 | 3.88625754 | 1.07E-08 | NA | NA | NA | NA | | | | |
| | rs545690161 | 9 | 93030699 | G | A | 0.005938242 | 6 | 17.1436874 | 3.00318779 | 1.14E-08 | NA | NA | NA | NA | | | | |
| | rs77141817 | 4 | 37053759 | T | C | 0.003562945 | 3 | 20.6110292 | 3.64214866 | 1.52E-08 | NA | NA | NA | NA | | | | |
| | rs190822761 | 4 | 37099356 | G | T | 0.003562945 | 3 | 20.5846354 | 3.64346559 | 1.61E-08 | NA | NA | NA | NA | | | | |
| | rs10279777 | 7 | 18441589 | G | A | 0.009501188 | 8 | 12.8681335 | 2.27961694 | 1.65E-08 | NA | NA | NA | NA | | | | |
| | rs10486295 | 7 | 18446807 | G | A | 0.009501188 | 8 | 12.76229 | 2.26480456 | 1.75E-08 | NA | NA | NA | NA | | | | |
| | rs147559909 | 2 | 237051523 | T | C | 0.005938242 | 5 | 17.0840975 | 3.03209195 | 1.76E-08 | NA | NA | NA | NA | | | | |
| | rs17169602 | 7 | 18446741 | G | A | 0.009501188 | 8 | 12.7273717 | 2.25993984 | 1.78E-08 | NA | NA | NA | NA | | | | |
| | rs139598422 | 13 | 23887014 | A | G | 0.003562945 | 4 | 20.4886522 | 3.64167194 | 1.84E-08 | NA | NA | NA | NA | | | | |
| | rs74704551 | 14 | 30161887 | C | T | 0.003562945 | 3 | 24.1990705 | 4.31064693 | 1.98E-08 | NA | NA | NA | NA | | | | |
| | rs77300464 | 7 | 18408761 | A | G | 0.007125891 | 6 | 14.6659794 | 2.6169516 | 2.09E-08 | NA | NA | NA | NA | | | | |
| | rs75606013 | 7 | 18414613 | G | A | 0.007125891 | 6 | 14.6617119 | 2.61692421 | 2.11E-08 | NA | NA | NA | NA | | | | |
| | rs541653703 | 11 | 18701786 | G | A | 0.004750594 | 4 | 18.3169499 | 3.27243418 | 2.18E-08 | NA | NA | NA | NA | | | | |
| | rs117913371 | 10 | 102918486 | G | A | 0.024940618 | 21 | 7.86609555 | 1.40785512 | 2.31E-08 | NA | NA | NA | NA | | | | |
| | rs191053292 | 12 | 63445280 | T | C | 0.003562945 | 3 | 22.9266488 | 4.11439732 | 2.51E-08 | NA | NA | NA | NA | | | | |
| | rs112351653 | 1 | 103220360 | T | C | 0.028503563 | 24 | 7.42386947 | 1.33286772 | 2.55E-08 | NA | NA | NA | NA | | | | |
| | rs114413507 | 1 | 103419168 | T | C | 0.028503563 | 24 | 7.42216029 | 1.33294954 | 2.57E-08 | NA | NA | NA | NA | | | | |
| | rs138414342 | 11 | 18679398 | G | A | 0.004750594 | 5 | 18.1582505 | 3.28214943 | 3.16E-08 | NA | NA | NA | NA | | | | |
| | rs16823323 | 3 | 153657202 | G | A | 0.016627078 | 14 | 9.48464065 | 1.71769795 | 3.36E-08 | NA | NA | NA | NA | | | | |
| | rs183737367 | 9 | 93330047 | T | C | 0.003562945 | 3 | 23.3452306 | 4.24471431 | 3.80E-08 | NA | NA | NA | NA | | | | |
| | rs560206697 | 19 | 20729098 | C | T | 0.002375297 | 3 | 24.9989943 | 4.55350673 | 4.02E-08 | NA | NA | NA | NA | | | | |
| | rs79486609 | 21 | 17245006 | G | A | 0.003562945 | 3 | 20.6624886 | 3.77824957 | 4.53E-08 | NA | NA | NA | NA | | | | |

| | | | | | | | | | | | | | |
|---|---|---|---|---|---|---|---|---|---|---|---|---|---|
| rs151115079 | 11 | 18655741 | T | C | 0.004750594 | 5 | 17.8273587 | 3.2680205 | 4.89E-08 | NA | NA | NA | NA |
| rs185510569 | 2 | 223814861 | G | A | 0.003562945 | 3 | 19.5886906 | 3.60490977 | 5.51E-08 | NA | NA | NA | NA |
| rs187213609 | 9 | 93415465 | C | T | 0.003562945 | 3 | 23.1975304 | 4.27412656 | 5.72E-08 | NA | NA | NA | NA |
| rs117280553 | 21 | 17207163 | T | C | 0.003562945 | 3 | 20.087417 | 3.70274826 | 5.80E-08 | NA | NA | NA | NA |
| rs183816745 | 5 | 91631888 | A | G | 0.005938242 | 5 | 17.3487909 | 3.20195556 | 6.02E-08 | NA | NA | NA | NA |
| rs180828621 | 10 | 124533409 | G | A | 0.007125891 | 6 | 15.1380574 | 2.79957627 | 6.40E-08 | NA | NA | NA | NA |
| rs142684595 | 3 | 55319301 | T | C | 0.005938242 | 5 | 16.7729431 | 3.11364755 | 7.17E-08 | NA | NA | NA | NA |
| rs184785969 | 21 | 17171431 | C | A | 0.003562945 | 3 | 19.8736486 | 3.70683937 | 8.26E-08 | NA | NA | NA | NA |
| rs77180278 | 1 | 102961882 | T | C | 0.028503563 | 24 | 7.33619996 | 1.37404112 | 9.34E-08 | NA | NA | NA | NA |
| rs189709453 | 3 | 177574308 | G | A | 0.003562945 | 3 | 21.6581488 | 4.05916142 | 9.52E-08 | NA | NA | NA | NA |
| rs111846247 | 5 | 95111643 | T | C | 0.003562945 | 3 | 20.4865772 | 3.85467205 | 1.07E-07 | NA | NA | NA | NA |
| rs145875128 | 4 | 32073136 | G | A | 0.003562945 | 3 | 24.479986 | 4.60639213 | 1.07E-07 | NA | NA | NA | NA |
| rs75848314 | 5 | 95098340 | T | C | 0.003562945 | 3 | 20.0998481 | 3.78700451 | 1.11E-07 | NA | NA | NA | NA |
| rs190193113 | 2 | 53085778 | G | A | 0.003562945 | 3 | 19.9533602 | 3.7619559 | 1.13E-07 | NA | NA | NA | NA |
| rs183378658 | 2 | 52563371 | C | T | 0.003562945 | 3 | 19.7446169 | 3.72634921 | 1.17E-07 | NA | NA | NA | NA |
| rs143371352 | 13 | 47383834 | C | T | 0.003562945 | 3 | 20.5341625 | 3.87928964 | 1.20E-07 | NA | NA | NA | NA |
| rs75186966 | 13 | 47395758 | A | C | 0.003562945 | 3 | 20.2281345 | 3.83225523 | 1.30E-07 | NA | NA | NA | NA |
| rs532416695 | 2 | 177486790 | G | A | 0.003562945 | 3 | 19.8726656 | 3.76737683 | 1.33E-07 | NA | NA | NA | NA |
| rs145080832 | 2 | 48473143 | G | A | 0.004750594 | 4 | 17.1618477 | 3.25709537 | 1.37E-07 | NA | NA | NA | NA |
| rs182868205 | 3 | 177712448 | C | T | 0.003562945 | 3 | 20.9548768 | 3.97841839 | 1.39E-07 | NA | NA | NA | NA |
| rs143538552 | 18 | 29050262 | A | G | 0.003562945 | 3 | 19.1457218 | 3.63679703 | 1.41E-07 | NA | NA | NA | NA |
| rs1221830047 | 8 | 55690220 | T | C | 0.330166271 | 277 | 2.3570878 | 0.44879289 | 1.50E-07 | NA | NA | NA | NA |
| rs7822082 | 8 | 55690220 | T | C | 0.330166271 | 277 | 2.3570878 | 0.44879289 | 1.50E-07 | NA | NA | NA | NA |
| rs72986533 | 6 | 142611258 | T | C | 0.008313539 | 8 | 13.2812726 | 2.53063629 | 1.54E-07 | NA | NA | NA | NA |
| rs373746073 | 18 | 29058384 | C | A | 0.003562945 | 3 | 19.0847197 | 3.63798064 | 1.55E-07 | NA | NA | NA | NA |
| rs181193202 | 2 | 52527354 | T | C | 0.003562945 | 3 | 19.6312088 | 3.74776015 | 1.62E-07 | NA | NA | NA | NA |
| rs186767531 | 3 | 177639777 | T | C | 0.003562945 | 3 | 21.2013243 | 4.04780758 | 1.63E-07 | NA | NA | NA | NA |
| rs140797780 | 8 | 22087792 | C | T | 0.003562945 | 3 | 19.2908396 | 3.69147785 | 1.73E-07 | NA | NA | NA | NA |
| rs137880949 | 12 | 63306297 | T | C | 0.004750594 | 3 | 21.6084034 | 4.13589168 | 1.75E-07 | NA | NA | NA | NA |
| rs148532212 | 6 | 165499857 | T | C | 0.003562945 | 3 | 19.3535457 | 3.70945693 | 1.81E-07 | NA | NA | NA | NA |
| rs141754456 | 12 | 20151132 | T | C | 0.007125891 | 7 | 12.1460333 | 2.32842921 | 1.82E-07 | NA | NA | NA | NA |
| rs117498042 | 6 | 165514281 | C | T | 0.003562945 | 3 | 19.335471 | 3.70913521 | 1.86E-07 | NA | NA | NA | NA |
| rs184487573 | 6 | 167513471 | A | G | 0.007125891 | 7 | 13.5009635 | 2.59268014 | 1.92E-07 | NA | NA | NA | NA |
| rs149298750 | 4 | 127158701 | A | C | 0.003562945 | 3 | 22.184052 | 4.26573246 | 1.99E-07 | NA | NA | NA | NA |
| rs140277951 | 10 | 82359100 | G | A | 0.008313539 | 6 | 13.7463158 | 2.64333932 | 1.99E-07 | NA | NA | NA | NA |
| rs558553658 | 2 | 15682724 | C | T | 0.004750594 | 3 | 17.1099107 | 3.29108061 | 2.01E-07 | NA | NA | NA | NA |
| rs112679237 | 4 | 139140860 | T | C | 0.017814727 | 18 | 8.71202752 | 1.67680671 | 2.04E-07 | NA | NA | NA | NA |
| rs116862847 | 14 | 64141677 | C | T | 0.007125891 | 7 | 15.3160369 | 2.95194262 | 2.12E-07 | NA | NA | NA | NA |
| rs528809914 | 13 | 113041256 | G | A | 0.003562945 | 3 | 19.5048552 | 3.76867111 | 2.27E-07 | NA | NA | NA | NA |
| rs112007361 | 11 | 99188380 | A | C | 0.03087886 | 26 | 6.35658468 | 1.22825758 | 2.28E-07 | NA | NA | NA | NA |
| rs546144116 | 19 | 19563339 | C | T | 0.002375297 | 3 | 23.1354815 | 4.47162758 | 2.29E-07 | NA | NA | NA | NA |
| rs17510814 | 12 | 28468969 | A | C | 0.007125891 | 6 | 14.0387924 | 2.71409689 | 2.31E-07 | NA | NA | NA | NA |
| rs113167689 | 12 | 28435962 | C | T | 0.007125891 | 6 | 14.1364135 | 2.73335629 | 2.32E-07 | NA | NA | NA | NA |
| rs140788628 | 20 | 15858501 | C | A | 0.010688836 | 8 | 11.8039284 | 2.2826396 | 2.33E-07 | NA | NA | NA | NA |
| rs141756120 | 12 | 28511096 | A | C | 0.008313539 | 6 | 14.278403 | 2.76458538 | 2.41E-07 | NA | NA | NA | NA |
| rs117991215 | 12 | 28511473 | T | C | 0.008313539 | 6 | 14.333666 | 2.77918736 | 2.50E-07 | NA | NA | NA | NA |
| rs191423619 | 4 | 126516627 | G | T | 0.002375297 | 3 | 21.694611 | 4.20667337 | 2.51E-07 | NA | NA | NA | NA |
| rs113651406 | 4 | 990967 | C | T | 0.003562945 | 3 | 19.4054026 | 3.7635826 | 2.52E-07 | NA | NA | NA | NA |
| rs187942235 | 18 | 27030430 | C | T | 0.003562945 | 3 | 20.3968658 | 3.96963025 | 2.77E-07 | NA | NA | NA | NA |
| rs16920698 | 8 | 55678434 | G | A | 0.330166271 | 277 | 2.30492695 | 0.44908177 | 2.86E-07 | NA | NA | NA | NA |
| rs4737676 | 8 | 55679546 | G | A | 0.330166271 | 277 | 2.30492057 | 0.44908192 | 2.86E-07 | NA | NA | NA | NA |
| rs4737674 | 8 | 55661654 | C | A | 0.330166271 | 277 | 2.30559554 | 0.44921988 | 2.86E-07 | NA | NA | NA | NA |
| rs13277510 | 8 | 55674149 | G | A | 0.330166271 | 277 | 2.30494752 | 0.44909942 | 2.86E-07 | NA | NA | NA | NA |
| rs983248 | 8 | 55680792 | C | T | 0.330166271 | 277 | 2.30260428 | 0.44871461 | 2.87E-07 | NA | NA | NA | NA |
| rs1391463 | 8 | 55681876 | T | G | 0.330166271 | 277 | 2.30258865 | 0.44871307 | 2.87E-07 | NA | NA | NA | NA |
| rs4737201 | 8 | 55691458 | C | T | 0.330166271 | 277 | 2.30197734 | 0.44869179 | 2.89E-07 | NA | NA | NA | NA |
| rs549931083 | 12 | 20516286 | A | C | 0.003562945 | 3 | 14.3282694 | 2.79891395 | 3.07E-07 | NA | NA | NA | NA |
| rs141281289 | 11 | 123693024 | A | G | 0.005938242 | 6 | 15.811518 | 3.0889522 | 3.08E-07 | NA | NA | NA | NA |

| | | | | | | | | | | | | | |
|---|---|---|---|---|---|---|---|---|---|---|---|---|---|
| rs184220112 | 2 | 48631743 | C | A | 0.004750594 | 4 | 16.4879043 | 3.22267606 | 3.12E-07 | NA | NA | NA | NA |
| rs11987234 | 8 | 55669829 | A | G | 0.328978622 | 276 | 2.29599259 | 0.44884532 | 3.13E-07 | NA | NA | NA | NA |
| rs13276543 | 8 | 55688174 | G | T | 0.328978622 | 276 | 2.29263157 | 0.44823093 | 3.14E-07 | NA | NA | NA | NA |
| rs111838310 | 2 | 74673491 | C | A | 0.004750594 | 5 | 17.7963938 | 3.48429066 | 3.26E-07 | NA | NA | NA | NA |
| rs80156375 | 7 | 18443215 | A | C | 0.009501188 | 8 | 11.5406255 | 2.25971308 | 3.27E-07 | NA | NA | NA | NA |
| rs1561297 | 8 | 55678538 | A | C | 0.332541568 | 279 | 2.28315237 | 0.44743146 | 3.35E-07 | NA | NA | NA | NA |
| rs112983626 | 2 | 74697150 | G | A | 0.004750594 | 4 | 17.7848079 | 3.48532494 | 3.35E-07 | NA | NA | NA | NA |
| rs12548593 | 8 | 55674617 | G | T | 0.332541568 | 279 | 2.28269961 | 0.44752402 | 3.38E-07 | NA | NA | NA | NA |
| rs10105693 | 8 | 55640472 | C | T | 0.328978622 | 276 | 2.2928391 | 0.44965282 | 3.41E-07 | NA | NA | NA | NA |
| rs13278605 | 8 | 55688171 | C | T | 0.328978622 | 277 | 2.27377348 | 0.44593165 | 3.42E-07 | NA | NA | NA | NA |
| rs2083123 | 8 | 55680318 | C | T | 0.332541568 | 279 | 2.27910794 | 0.44707491 | 3.44E-07 | NA | NA | NA | NA |
| rs17017794 | 4 | 91825885 | T | C | 0.039192399 | 33 | 5.83240545 | 1.14450838 | 3.47E-07 | NA | NA | NA | NA |
| rs74521112 | 11 | 99089147 | G | T | 0.032066508 | 27 | 6.25632387 | 1.22787586 | 3.48E-07 | NA | NA | NA | NA |
| rs116651654 | 4 | 163238743 | C | T | 0.007125891 | 6 | 15.6223694 | 3.06624451 | 3.49E-07 | NA | NA | NA | NA |
| rs565682685 | 9 | 93222328 | T | C | 0.004750594 | 4 | 18.938397 | 3.7206174 | 3.58E-07 | NA | NA | NA | NA |
| rs144541665 | 1 | 104310729 | G | A | 0.002375297 | 3 | 21.8734611 | 4.30339839 | 3.72E-07 | NA | NA | NA | NA |
| rs193253461 | 15 | 59229353 | A | G | 0.013064133 | 12 | 10.5374468 | 2.07404763 | 3.76E-07 | NA | NA | NA | NA |
| rs77867199 | 7 | 18442275 | G | T | 0.010688836 | 9 | 10.8221282 | 2.13363365 | 3.93E-07 | NA | NA | NA | NA |
| rs189360484 | 12 | 1870510 | A | G | 0.004750594 | 4 | 16.8078794 | 3.31424855 | 3.95E-07 | NA | NA | NA | NA |
| rs79182806 | 7 | 18433827 | T | C | 0.008313539 | 7 | 12.0972603 | 2.38956889 | 4.14E-07 | NA | NA | NA | NA |
| rs189890455 | 2 | 48655397 | C | T | 0.005938242 | 5 | 15.1467391 | 2.99355083 | 4.20E-07 | NA | NA | NA | NA |
| rs190294315 | 9 | 85945465 | C | T | 0.005938242 | 5 | 14.5664276 | 2.88126649 | 4.29E-07 | NA | NA | NA | NA |
| rs185819304 | 18 | 27001580 | G | A | 0.003562945 | 3 | 19.9401113 | 3.94501953 | 4.32E-07 | NA | NA | NA | NA |
| rs148433854 | 19 | 31096478 | G | A | 0.003562945 | 3 | 18.6267585 | 3.68806754 | 4.41E-07 | NA | NA | NA | NA |
| rs576124203 | 3 | 141841196 | T | G | 0.003562945 | 3 | 19.4039925 | 3.84392582 | 4.47E-07 | NA | NA | NA | NA |
| rs143597860 | 1 | 104157143 | A | G | 0.002375297 | 3 | 21.5748154 | 4.2760379 | 4.52E-07 | NA | NA | NA | NA |
| rs140782222 | 9 | 84028894 | T | C | 0.002375297 | 3 | 21.791526 | 4.31928461 | 4.53E-07 | NA | NA | NA | NA |
| rs1856085 | 1 | 104114545 | G | A | 0.002375297 | 3 | 21.51144 | 4.26747898 | 4.64E-07 | NA | NA | NA | NA |
| rs186768950 | 19 | 18806124 | C | A | 0.002375297 | 3 | 22.7152061 | 4.50776479 | 4.68E-07 | NA | NA | NA | NA |
| rs182303755 | 15 | 59634792 | A | C | 0.013064133 | 12 | 10.5479917 | 2.09359291 | 4.70E-07 | NA | NA | NA | NA |
| rs545552231 | 3 | 141864350 | C | T | 0.003562945 | 3 | 19.8641332 | 3.94582563 | 4.80E-07 | NA | NA | NA | NA |
| rs78547898 | 22 | 32824278 | G | A | 0.003562945 | 3 | 20.9098395 | 4.15538274 | 4.85E-07 | NA | NA | NA | NA |
| rs796777817 | 22 | 32824278 | G | A | 0.003562945 | 3 | 20.9098395 | 4.15538274 | 4.85E-07 | NA | NA | NA | NA |
| rs78907958 | 7 | 18425017 | T | G | 0.008313539 | 7 | 12.0901322 | 2.40514961 | 4.99E-07 | NA | NA | NA | NA |
| rs113751774 | 4 | 23428854 | C | T | 0.007125891 | 7 | 14.9103274 | 2.96657459 | 5.01E-07 | NA | NA | NA | NA |
| rs10958428 | 8 | 55685641 | A | G | 0.333729216 | 280 | 2.23996885 | 0.44568673 | 5.01E-07 | NA | NA | NA | NA |
| rs17776100 | 7 | 6426479 | G | A | 0.029691211 | 25 | 6.63697397 | 1.32177185 | 5.13E-07 | NA | NA | NA | NA |
| rs185464792 | 19 | 18797371 | C | T | 0.002375297 | 3 | 22.5896015 | 4.50191473 | 5.23E-07 | NA | NA | NA | NA |
| rs117025967 | 10 | 20207052 | C | A | 0.013064133 | 11 | 9.66973563 | 1.927439 | 5.25E-07 | NA | NA | NA | NA |
| rs188028357 | 7 | 100668425 | C | T | 0.002375297 | 3 | 21.8425578 | 4.35791809 | 5.38E-07 | NA | NA | NA | NA |
| rs148556485 | 9 | 84023826 | A | C | 0.002375297 | 3 | 21.3800152 | 4.26618316 | 5.40E-07 | NA | NA | NA | NA |
| rs184613584 | 17 | 48508221 | A | C | 0.007125891 | 6 | 14.1377974 | 2.8212234 | 5.41E-07 | NA | NA | NA | NA |
| rs528288879 | 2 | 65875930 | C | T | 0.005938242 | 6 | 14.7878085 | 2.95121929 | 5.42E-07 | NA | NA | NA | NA |
| rs1437782 | 8 | 55632762 | C | T | 0.330166271 | 278 | 2.2579585 | 0.45098781 | 5.54E-07 | NA | NA | NA | NA |
| rs534845494 | 13 | 58213864 | A | G | 0.004750594 | 5 | 15.9431765 | 3.18484007 | 5.56E-07 | NA | NA | NA | NA |
| rs1008091735 | 19 | 31090099 | T | C | 0.003562945 | 3 | 18.4689243 | 3.69079909 | 5.61E-07 | NA | NA | NA | NA |
| rs567982164 | 5 | 25631297 | G | A | 0.002375297 | 3 | 21.8356286 | 4.36706703 | 5.73E-07 | NA | NA | NA | NA |
| rs79213709 | 11 | 99093455 | G | A | 0.032066508 | 26 | 6.17081884 | 1.23453654 | 5.78E-07 | NA | NA | NA | NA |
| rs559008174 | 19 | 18876059 | C | T | 0.004750594 | 5 | 15.6025661 | 3.12812006 | 6.11E-07 | NA | NA | NA | NA |
| rs541288561 | 19 | 18869445 | T | G | 0.004750594 | 5 | 15.6119012 | 3.13183483 | 6.20E-07 | NA | NA | NA | NA |
| rs190251199 | 14 | 105590577 | T | C | 0.003562945 | 3 | 20.0125292 | 4.01627173 | 6.27E-07 | NA | NA | NA | NA |
| rs138215817 | 14 | 22641516 | A | G | 0.004750594 | 4 | 16.311019 | 3.27367022 | 6.28E-07 | NA | NA | NA | NA |
| rs111407636 | 5 | 95080029 | C | T | 0.003562945 | 3 | 18.642774 | 3.74168052 | 6.28E-07 | NA | NA | NA | NA |
| rs73085348 | 3 | 42711221 | A | G | 0.011876485 | 11 | 10.139317 | 2.03504571 | 6.28E-07 | NA | NA | NA | NA |
| rs1437781 | 8 | 55629852 | T | C | 0.332541568 | 280 | 2.23854673 | 0.44940766 | 6.32E-07 | NA | NA | NA | NA |
| rs147032554 | 1 | 186148864 | T | G | 0.003562945 | 3 | 18.162609 | 3.65402821 | 6.68E-07 | NA | NA | NA | NA |
| rs111676272 | 5 | 95094298 | C | A | 0.003562945 | 3 | 18.6068625 | 3.74429748 | 6.72E-07 | NA | NA | NA | NA |
| rs2274996 | 1 | 229804538 | C | T | 0.041567696 | 35 | 5.58527433 | 1.12406363 | 6.74E-07 | NA | NA | NA | NA |

| | | | | | | | | | | | | | |
|---|---|---|---|---|---|---|---|---|---|---|---|---|---|
| rs2891865 | 1 | 229806368 | A | G | 0.041567696 | 35 | 5.58181376 | 1.12378916 | 6.80E-07 | NA | NA | NA | NA |
| rs2385790 | 1 | 229807492 | C | T | 0.041567696 | 35 | 5.58224822 | 1.12394723 | 6.81E-07 | NA | NA | NA | NA |
| rs12024557 | 1 | 229812357 | A | C | 0.042755344 | 35 | 5.56930969 | 1.12195729 | 6.91E-07 | NA | NA | NA | NA |
| rs78296164 | 9 | 135266715 | C | T | 0.008313539 | 7 | 11.6095338 | 2.33917557 | 6.94E-07 | NA | NA | NA | NA |
| rs570407448 | 19 | 18880030 | G | A | 0.004750594 | 5 | 15.4977984 | 3.12412555 | 7.02E-07 | NA | NA | NA | NA |
| rs76777840 | 2 | 48312950 | G | A | 0.005938242 | 5 | 14.3500784 | 2.8927894 | 7.03E-07 | NA | NA | NA | NA |
| rs528609331 | 11 | 125842195 | C | T | 0.003562945 | 3 | 19.5812418 | 3.94832705 | 7.07E-07 | NA | NA | NA | NA |
| rs182437250 | 12 | 63608466 | T | C | 0.004750594 | 4 | 18.8083451 | 3.79343554 | 7.12E-07 | NA | NA | NA | NA |
| rs147601511 | 8 | 22322319 | A | G | 0.004750594 | 4 | 16.4471181 | 3.31777038 | 7.15E-07 | NA | NA | NA | NA |
| rs858397 | 8 | 55614690 | A | G | 0.331353919 | 278 | 2.2301348 | 0.44990341 | 7.16E-07 | NA | NA | NA | NA |
| rs75334617 | 10 | 102956152 | G | A | 0.038004751 | 32 | 5.84603405 | 1.18027345 | 7.30E-07 | NA | NA | NA | NA |
| rs4562666 | 1 | 229824770 | T | C | 0.042755344 | 36 | 5.51922599 | 1.11457457 | 7.35E-07 | NA | NA | NA | NA |
| rs382476 | 8 | 55590975 | G | A | 0.666270784 | 288 | -2.2481938 | 0.4540554 | 7.37E-07 | NA | NA | NA | NA |
| rs384543 | 8 | 55591609 | G | A | 0.666270784 | 288 | -2.2481938 | 0.4540554 | 7.37E-07 | NA | NA | NA | NA |
| rs446222 | 8 | 55574960 | G | A | 0.666270784 | 288 | -2.247819 | 0.45397995 | 7.37E-07 | NA | NA | NA | NA |
| rs384127 | 8 | 55597489 | G | A | 0.666270784 | 288 | -2.2481559 | 0.45404811 | 7.37E-07 | NA | NA | NA | NA |
| rs12045643 | 1 | 229834050 | C | T | 0.042755344 | 36 | 5.48149152 | 1.10708948 | 7.37E-07 | NA | NA | NA | NA |
| rs147630370 | 4 | 87450675 | T | C | 0.004750594 | 4 | 16.0013633 | 3.23339786 | 7.47E-07 | NA | NA | NA | NA |
| rs532513136 | 8 | 135813748 | C | A | 0.003562945 | 3 | 19.9490594 | 4.03118599 | 7.47E-07 | NA | NA | NA | NA |
| rs184265355 | 13 | 108011352 | A | C | 0.005938242 | 6 | 14.4271719 | 2.91829792 | 7.67E-07 | NA | NA | NA | NA |
| rs148248743 | 3 | 136134595 | C | T | 0.002375297 | 3 | 22.839379 | 4.62044689 | 7.69E-07 | NA | NA | NA | NA |
| rs72983831 | 6 | 142307100 | T | G | 0.005938242 | 6 | 15.6733239 | 3.17294911 | 7.83E-07 | NA | NA | NA | NA |
| rs80292573 | 15 | 59435086 | T | G | 0.034441805 | 30 | 6.55323491 | 1.32696583 | 7.87E-07 | NA | NA | NA | NA |
| rs111927235 | 2 | 74483954 | A | G | 0.004750594 | 4 | 17.0468955 | 3.46111889 | 8.43E-07 | NA | NA | NA | NA |
| rs369623 | 8 | 55571940 | A | C | 0.666270784 | 287 | -2.2359815 | 0.45415557 | 8.51E-07 | NA | NA | NA | NA |
| rs188720948 | 3 | 150069880 | T | C | 0.003562945 | 3 | 17.7032653 | 3.59722874 | 8.59E-07 | NA | NA | NA | NA |
| rs148998974 | 22 | 40620530 | A | G | 0.005938242 | 5 | 14.5129532 | 2.94976024 | 8.65E-07 | NA | NA | NA | NA |
| rs144954214 | 16 | 76179362 | A | G | 0.002375297 | 3 | 21.8113263 | 4.43942529 | 8.96E-07 | NA | NA | NA | NA |
| rs145439370 | 15 | 58879765 | T | C | 0.03087886 | 26 | 6.89328891 | 1.40339117 | 9.02E-07 | NA | NA | NA | NA |
| rs75689761 | 7 | 18406573 | C | T | 0.008313539 | 7 | 11.8482917 | 2.41320258 | 9.12E-07 | NA | NA | NA | NA |
| rs113221952 | 1 | 103753974 | A | G | 0.021377672 | 19 | 7.92350704 | 1.61421682 | 9.17E-07 | NA | NA | NA | NA |
| rs61434999 | 7 | 18418351 | A | G | 0.008313539 | 7 | 11.8411511 | 2.41284027 | 9.22E-07 | NA | NA | NA | NA |
| rs433324 | 8 | 55564609 | A | G | 0.666270784 | 287 | -2.2483462 | 0.45814252 | 9.22E-07 | NA | NA | NA | NA |
| rs528140343 | 11 | 125719227 | A | C | 0.003562945 | 3 | 18.2835521 | 3.72738029 | 9.33E-07 | NA | NA | NA | NA |
| rs34270375 | 1 | 89370702 | G | A | 0.026128266 | 21 | 7.71147017 | 1.57216792 | 9.34E-07 | NA | NA | NA | NA |
| rs75773869 | 7 | 18410845 | G | T | 0.008313539 | 7 | 11.8642716 | 2.41902322 | 9.36E-07 | NA | NA | NA | NA |
| rs79602997 | 7 | 18410250 | G | A | 0.008313539 | 7 | 11.8608211 | 2.41901626 | 9.43E-07 | NA | NA | NA | NA |
| rs56224400 | 6 | 98092675 | T | C | 0.016627078 | 14 | 8.4433064 | 1.72251471 | 9.50E-07 | NA | NA | NA | NA |
| rs186532456 | 1 | 18948328 | C | T | 0.003562945 | 3 | 20.1730343 | 4.11737745 | 9.61E-07 | NA | NA | NA | NA |
| rs118093638 | 11 | 18718324 | C | T | 0.005938242 | 5 | 14.2657259 | 2.91296506 | 9.72E-07 | NA | NA | NA | NA |
| rs77353774 | 12 | 28248852 | G | A | 0.007125891 | 6 | 13.1054362 | 2.67954103 | 1.00E-06 | NA | NA | NA | NA |
| rs532730683 | 9 | 1784492 | G | T | 0.003562945 | 3 | 18.726838 | 3.8314675 | 1.02E-06 | NA | NA | NA | NA |
| rs185139807 | 22 | 40594781 | G | A | 0.005938242 | 5 | 14.4330526 | 2.95312686 | 1.02E-06 | NA | NA | NA | NA |
| rs146333745 | 18 | 55497457 | C | T | 0.003562945 | 4 | 17.4215784 | 3.56782335 | 1.04E-06 | NA | NA | NA | NA |
| rs541680196 | 22 | 40528090 | G | A | 0.005938242 | 5 | 14.3867808 | 2.94903659 | 1.07E-06 | NA | NA | NA | NA |
| rs12036586 | 1 | 229826378 | G | A | 0.047505938 | 40 | 5.22212176 | 1.07114093 | 1.09E-06 | NA | NA | NA | NA |
| rs529523094 | 16 | 77715551 | A | G | 0.003562945 | 3 | 18.1584346 | 3.72617419 | 1.10E-06 | NA | NA | NA | NA |
| rs562032622 | 1 | 18959333 | A | C | 0.003562945 | 3 | 21.0267115 | 4.3197503 | 1.13E-06 | NA | NA | NA | NA |
| rs78225611 | 7 | 18407464 | A | C | 0.009501188 | 8 | 11.0294387 | 2.26622466 | 1.13E-06 | NA | NA | NA | NA |
| rs77346868 | 7 | 18406599 | A | G | 0.009501188 | 8 | 11.0210129 | 2.26481172 | 1.14E-06 | NA | NA | NA | NA |
| rs184098071 | 2 | 177116420 | G | A | 0.003562945 | 3 | 13.947724 | 2.86782452 | 1.15E-06 | NA | NA | NA | NA |
| rs118183140 | 21 | 35477486 | C | T | 0.020190024 | 17 | 7.5893248 | 1.56122232 | 1.17E-06 | NA | NA | NA | NA |
| rs113625788 | 22 | 19969182 | C | T | 0.008313539 | 7 | 11.8084829 | 2.42941561 | 1.17E-06 | NA | NA | NA | NA |
| rs116189766 | 2 | 126393864 | T | C | 0.002375297 | 3 | 21.190764 | 4.36037289 | 1.17E-06 | NA | NA | NA | NA |
| rs76098744 | 1 | 112349372 | C | T | 0.016627078 | 14 | 8.38612452 | 1.72704116 | 1.20E-06 | NA | NA | NA | NA |
| rs74683551 | 1 | 112354418 | G | A | 0.016627078 | 14 | 8.38252043 | 1.72705812 | 1.21E-06 | NA | NA | NA | NA |
| rs185855183 | 12 | 101505126 | C | T | 0.003562945 | 3 | 18.7397727 | 3.86137343 | 1.22E-06 | NA | NA | NA | NA |
| rs405226 | 8 | 55592336 | A | G | 0.662707838 | 291 | -2.1881305 | 0.45101874 | 1.23E-06 | NA | NA | NA | NA |

| | | | | | | | | | | | | | |
|---|---|---|---|---|---|---|---|---|---|---|---|---|---|
| rs3098298 | 8 | 55582838 | C | T | 0.662707838 | 291 | -2.1875635 | 0.45094791 | 1.23E-06 | NA | NA | NA | NA |
| rs367179 | 8 | 55587616 | T | C | 0.662707838 | 291 | -2.1875635 | 0.45094791 | 1.23E-06 | NA | NA | NA | NA |
| rs432393 | 8 | 55580298 | C | T | 0.662707838 | 291 | -2.1875382 | 0.45096346 | 1.23E-06 | NA | NA | NA | NA |
| rs117185941 | 21 | 38766484 | G | A | 0.005938242 | 5 | 15.3808786 | 3.17143013 | 1.24E-06 | NA | NA | NA | NA |
| rs1329159859 | 21 | 38766484 | G | A | 0.005938242 | 5 | 15.3808786 | 3.17143013 | 1.24E-06 | NA | NA | NA | NA |
| rs139360368 | 5 | 73372109 | A | C | 0.003562945 | 4 | 18.3590487 | 3.78701272 | 1.25E-06 | NA | NA | NA | NA |
| rs184117160 | 15 | 59404306 | C | T | 0.014251781 | 11 | 10.3417236 | 2.13366338 | 1.25E-06 | NA | NA | NA | NA |
| rs145676540 | 3 | 2044635 | C | T | 0.005938242 | 5 | 13.980658 | 2.88447377 | 1.25E-06 | NA | NA | NA | NA |
| rs556293455 | 2 | 176950420 | G | A | 0.003562945 | 3 | 14.3860463 | 2.96998402 | 1.27E-06 | NA | NA | NA | NA |
| rs150027952 | 9 | 103385416 | A | G | 0.002375297 | 3 | 21.8616648 | 4.51464643 | 1.28E-06 | NA | NA | NA | NA |
| rs138217865 | 15 | 94392882 | C | T | 0.004750594 | 4 | 16.709641 | 3.45193896 | 1.29E-06 | NA | NA | NA | NA |
| rs112475378 | 3 | 1626661 | T | C | 0.016627078 | 14 | 9.55085929 | 1.97628138 | 1.35E-06 | NA | NA | NA | NA |
| rs146007933 | 3 | 28227423 | T | C | 0.021377672 | 18 | 7.51149453 | 1.55442805 | 1.35E-06 | NA | NA | NA | NA |
| rs192443987 | 3 | 135532345 | G | A | 0.004750594 | 4 | 17.7540285 | 3.67564302 | 1.36E-06 | NA | NA | NA | NA |
| rs149493615 | 1 | 79880089 | G | A | 0.008313539 | 8 | 11.750632 | 2.43451244 | 1.39E-06 | NA | NA | NA | NA |
| rs140706881 | 10 | 96217535 | G | A | 0.004750594 | 4 | 18.974805 | 3.93288931 | 1.40E-06 | NA | NA | NA | NA |
| rs142894171 | 2 | 151438271 | G | T | 0.003562945 | 3 | 18.0710394 | 3.74586836 | 1.41E-06 | NA | NA | NA | NA |
| rs17746486 | 2 | 95722609 | C | T | 0.032066508 | 29 | 6.90851358 | 1.43477175 | 1.47E-06 | NA | NA | NA | NA |
| rs180764936 | 12 | 101498621 | T | C | 0.003562945 | 3 | 19.3473053 | 4.0200238 | 1.49E-06 | NA | NA | NA | NA |
| rs190806532 | 12 | 48862147 | G | T | 0.002375297 | 3 | 21.326955 | 4.43856043 | 1.55E-06 | NA | NA | NA | NA |
| rs147393020 | 10 | 124779274 | A | G | 0.005938242 | 5 | 14.1853505 | 2.95980896 | 1.65E-06 | NA | NA | NA | NA |
| rs79539453 | 11 | 125266353 | C | T | 0.002375297 | 3 | 20.8220616 | 4.34542406 | 1.65E-06 | NA | NA | NA | NA |
| rs371245624 | 5 | 162880956 | T | C | 0.003562945 | 3 | 19.4795931 | 4.06572011 | 1.66E-06 | NA | NA | NA | NA |
| rs536803366 | 8 | 123001411 | T | C | 0.002375297 | 3 | 19.2098639 | 4.00952057 | 1.66E-06 | NA | NA | NA | NA |
| rs555249476 | 8 | 123001411 | T | C | 0.002375297 | 3 | 19.2098639 | 4.00952057 | 1.66E-06 | NA | NA | NA | NA |
| rs150077525 | 13 | 57979549 | A | G | 0.007125891 | 7 | 12.9984875 | 2.71312414 | 1.66E-06 | NA | NA | NA | NA |
| rs371879555 | 12 | 23015962 | T | C | 0.003562945 | 3 | 20.1033765 | 4.20002323 | 1.70E-06 | NA | NA | NA | NA |
| rs571986619 | 6 | 85678832 | A | G | 0.002375297 | 3 | 20.7858067 | 4.34301983 | 1.70E-06 | NA | NA | NA | NA |
| rs183586634 | 21 | 38763032 | G | A | 0.005938242 | 5 | 14.3132202 | 2.99210048 | 1.72E-06 | NA | NA | NA | NA |
| rs143811231 | 1 | 79968131 | T | C | 0.008313539 | 8 | 11.5747409 | 2.42066027 | 1.74E-06 | NA | NA | NA | NA |
| rs2365739 | 1 | 62484462 | G | A | 0.021377672 | 18 | 6.86815917 | 1.43887996 | 1.81E-06 | NA | NA | NA | NA |
| rs146526206 | 4 | 90993018 | T | C | 0.017814727 | 15 | 8.09176656 | 1.69535286 | 1.82E-06 | NA | NA | NA | NA |
| rs567383525 | 8 | 115029494 | C | T | 0.003562945 | 3 | 17.7455227 | 3.71970627 | 1.84E-06 | NA | NA | NA | NA |
| rs149425014 | 15 | 58951660 | T | C | 0.026128266 | 22 | 7.55224792 | 1.58320604 | 1.84E-06 | NA | NA | NA | NA |
| rs559559983 | 1 | 246977132 | C | A | 0.007125891 | 5 | 13.811132 | 2.89725588 | 1.87E-06 | NA | NA | NA | NA |
| rs117166500 | 7 | 17052778 | G | T | 0.007125891 | 7 | 12.5146865 | 2.62694198 | 1.90E-06 | NA | NA | NA | NA |
| rs145766563 | 1 | 185129502 | G | A | 0.003562945 | 4 | 16.8682102 | 3.54127008 | 1.90E-06 | NA | NA | NA | NA |
| rs540065886 | 9 | 2770228 | T | C | 0.003562945 | 3 | 19.9489151 | 4.18802805 | 1.90E-06 | NA | NA | NA | NA |
| rs118084887 | 21 | 38863820 | T | C | 0.005938242 | 5 | 14.9382616 | 3.13674741 | 1.91E-06 | NA | NA | NA | NA |
| rs140420703 | 1 | 102803484 | T | G | 0.004750594 | 5 | 16.487654 | 3.46213891 | 1.91E-06 | NA | NA | NA | NA |
| rs568321148 | 2 | 29870348 | T | G | 0.003562945 | 3 | 17.7125046 | 3.7201251 | 1.92E-06 | NA | NA | NA | NA |
| rs55844051 | 7 | 23360363 | T | C | 0.003562945 | 3 | 18.3875896 | 3.86268704 | 1.93E-06 | NA | NA | NA | NA |
| rs187236873 | 5 | 91530447 | G | A | 0.007125891 | 6 | 12.44907 | 2.61780937 | 1.98E-06 | NA | NA | NA | NA |
| rs1812506 | 8 | 55676101 | A | G | 0.345605701 | 291 | 2.12258868 | 0.4466178 | 2.01E-06 | NA | NA | NA | NA |
| rs539713344 | 7 | 100474786 | G | A | 0.002375297 | 3 | 20.044756 | 4.21957545 | 2.03E-06 | NA | NA | NA | NA |
| rs536781978 | 2 | 29733681 | A | G | 0.003562945 | 3 | 17.5870824 | 3.70259451 | 2.03E-06 | NA | NA | NA | NA |
| rs76327548 | 12 | 101182966 | G | A | 0.013064133 | 11 | 9.17842714 | 1.93235231 | 2.04E-06 | NA | NA | NA | NA |
| rs149421869 | 2 | 53483429 | G | T | 0.008313539 | 8 | 11.7382186 | 2.47204376 | 2.05E-06 | NA | NA | NA | NA |
| rs75024143 | 21 | 23156546 | G | T | 0.014251781 | 12 | 9.81254241 | 2.0684542 | 2.10E-06 | NA | NA | NA | NA |
| rs2375536 | 8 | 55640722 | T | C | 0.347980998 | 292 | 2.12635806 | 0.44857091 | 2.13E-06 | NA | NA | NA | NA |
| rs141326851 | 6 | 134833127 | A | C | 0.016627078 | 14 | 8.57849688 | 1.81053268 | 2.16E-06 | NA | NA | NA | NA |
| rs187518659 | 1 | 99455745 | T | G | 0.009501188 | 8 | 11.6819623 | 2.46564537 | 2.16E-06 | NA | NA | NA | NA |
| rs146479102 | 2 | 65825759 | G | A | 0.005938242 | 5 | 15.4846768 | 3.26980199 | 2.18E-06 | NA | NA | NA | NA |
| rs16850124 | 1 | 229831331 | T | C | 0.043942993 | 37 | 5.18095662 | 1.09435188 | 2.20E-06 | NA | NA | NA | NA |
| rs115348382 | 1 | 9655903 | G | A | 0.003562945 | 3 | 18.2107495 | 3.84721698 | 2.21E-06 | NA | NA | NA | NA |
| rs184200893 | 2 | 69260913 | C | T | 0.003562945 | 3 | 19.9580356 | 4.21905487 | 2.24E-06 | NA | NA | NA | NA |
| rs148781275 | 11 | 103640603 | A | G | 0.004750594 | 5 | 15.7111853 | 3.32183832 | 2.25E-06 | NA | NA | NA | NA |
| rs139943877 | 3 | 155451289 | G | A | 0.007125891 | 7 | 13.0535902 | 2.76041594 | 2.26E-06 | NA | NA | NA | NA |

| | | | | | | | | | | | | | |
|---|---|---|---|---|---|---|---|---|---|---|---|---|---|
| rs546409459 | 11 | 125754989 | A | G | 0.003562945 | 3 | 18.9195474 | 4.00286836 | 2.28E-06 | NA | NA | NA | NA |
| rs186649043 | 3 | 174932293 | C | T | 0.003562945 | 3 | 19.2134209 | 4.06588786 | 2.30E-06 | NA | NA | NA | NA |
| rs80203220 | 3 | 122722331 | C | T | 0.005938242 | 6 | 13.426042 | 2.84218656 | 2.31E-06 | NA | NA | NA | NA |
| rs541508507 | 2 | 170023265 | G | A | 0.004750594 | 4 | 15.4630033 | 3.27389618 | 2.32E-06 | NA | NA | NA | NA |
| rs529011661 | 10 | 20372316 | G | A | 0.004750594 | 4 | 15.6393469 | 3.3125339 | 2.34E-06 | NA | NA | NA | NA |
| rs182959028 | 22 | 45823032 | T | C | 0.008313539 | 7 | 11.944461 | 2.53146 | 2.38E-06 | NA | NA | NA | NA |
| rs146207930 | 9 | 129042336 | A | G | 0.007125891 | 6 | 11.8624321 | 2.51502465 | 2.40E-06 | NA | NA | NA | NA |
| rs290120 | 5 | 163268244 | T | G | 0.007125891 | 6 | 13.3587764 | 2.83260801 | 2.40E-06 | NA | NA | NA | NA |
| rs531769270 | 4 | 154411043 | T | C | 0.003562945 | 3 | 16.5945338 | 3.51936092 | 2.41E-06 | NA | NA | NA | NA |
| rs185620578 | 12 | 48569399 | C | T | 0.002375297 | 3 | 20.7232267 | 4.39505442 | 2.42E-06 | NA | NA | NA | NA |
| rs1301444047 | 7 | 36574504 | G | A | 0.042755344 | 43 | 5.12842209 | 1.08835549 | 2.45E-06 | NA | NA | NA | NA |
| rs62447184 | 7 | 36574504 | G | A | 0.042755344 | 43 | 5.12842209 | 1.08835549 | 2.45E-06 | NA | NA | NA | NA |
| rs118184666 | 12 | 20424749 | G | A | 0.007125891 | 6 | 10.9509803 | 2.32477242 | 2.47E-06 | NA | NA | NA | NA |
| rs10494861 | 1 | 205331874 | G | A | 0.003562945 | 3 | 19.1820498 | 4.0729134 | 2.48E-06 | NA | NA | NA | NA |
| rs183817723 | 16 | 59302775 | C | T | 0.003562945 | 4 | 17.5860507 | 3.7341919 | 2.48E-06 | NA | NA | NA | NA |
| rs12502861 | 4 | 2426305 | T | C | 0.010688836 | 9 | 11.1588472 | 2.36961887 | 2.49E-06 | NA | NA | NA | NA |
| rs191792521 | 3 | 195646605 | G | A | 0.008313539 | 6 | 13.9515484 | 2.96443599 | 2.52E-06 | NA | NA | NA | NA |
| rs149949098 | 11 | 95099866 | G | A | 0.016627078 | 13 | 8.55927625 | 1.81883308 | 2.53E-06 | NA | NA | NA | NA |
| rs74343174 | 5 | 161493182 | C | A | 0.005938242 | 5 | 14.7671824 | 3.13913408 | 2.55E-06 | NA | NA | NA | NA |
| rs189765693 | 4 | 4324793 | T | C | 0.004750594 | 4 | 15.5191341 | 3.29924607 | 2.55E-06 | NA | NA | NA | NA |
| rs189912648 | 5 | 134765439 | C | T | 0.003562945 | 4 | 16.9732599 | 3.60846227 | 2.55E-06 | NA | NA | NA | NA |
| rs151323346 | 12 | 21012024 | T | C | 0.005938242 | 4 | 15.8903888 | 3.38034868 | 2.59E-06 | NA | NA | NA | NA |
| rs536023430 | 7 | 146865072 | T | C | 0.003562945 | 3 | 19.4038683 | 4.13084856 | 2.64E-06 | NA | NA | NA | NA |
| rs144026361 | 15 | 41248669 | C | T | 0.004750594 | 5 | 16.1372994 | 3.43734659 | 2.67E-06 | NA | NA | NA | NA |
| rs186142189 | 2 | 67702707 | G | A | 0.007125891 | 6 | 12.7361927 | 2.71405368 | 2.70E-06 | NA | NA | NA | NA |
| rs142549310 | 2 | 170030506 | C | T | 0.004750594 | 4 | 15.2501713 | 3.24983634 | 2.70E-06 | NA | NA | NA | NA |
| rs72832764 | 5 | 170004673 | G | A | 0.003562945 | 3 | 18.2159757 | 3.88196743 | 2.70E-06 | NA | NA | NA | NA |
| rs183180157 | 1 | 181247121 | A | C | 0.009501188 | 7 | 12.61386 | 2.68921121 | 2.72E-06 | NA | NA | NA | NA |
| rs73227413 | 21 | 23136973 | G | A | 0.034441805 | 28 | 5.96205514 | 1.27150176 | 2.75E-06 | NA | NA | NA | NA |
| rs185874707 | 8 | 18030674 | C | T | 0.011876485 | 10 | 9.13934765 | 1.94918644 | 2.75E-06 | NA | NA | NA | NA |
| rs191271637 | 17 | 52123260 | A | G | 0.003562945 | 3 | 18.379836 | 3.92083355 | 2.76E-06 | NA | NA | NA | NA |
| rs187978759 | 7 | 11711845 | G | A | 0.003562945 | 3 | 20.4555522 | 4.3637719 | 2.76E-06 | NA | NA | NA | NA |
| rs193153124 | 3 | 148330710 | A | G | 0.005938242 | 5 | 14.3657313 | 3.06514594 | 2.78E-06 | NA | NA | NA | NA |
| rs1877768 | 6 | 16534923 | C | T | 0.017814727 | 15 | 8.28754685 | 1.76830748 | 2.78E-06 | NA | NA | NA | NA |
| rs76617932 | 1 | 180930424 | T | C | 0.011876485 | 9 | 10.5449608 | 2.25040726 | 2.79E-06 | NA | NA | NA | NA |
| rs188034471 | 9 | 85937714 | G | A | 0.004750594 | 4 | 14.7559999 | 3.14925113 | 2.79E-06 | NA | NA | NA | NA |
| rs1595406 | 8 | 55630615 | A | G | 0.345605701 | 291 | 2.10018889 | 0.44830064 | 2.80E-06 | NA | NA | NA | NA |
| rs147627638 | 6 | 99173116 | A | G | 0.007125891 | 6 | 13.0785306 | 2.79364254 | 2.85E-06 | NA | NA | NA | NA |
| rs151272830 | 2 | 67682724 | G | T | 0.007125891 | 6 | 13.3323479 | 2.84806076 | 2.85E-06 | NA | NA | NA | NA |
| rs141169929 | 3 | 164808462 | A | G | 0.004750594 | 5 | 14.8454178 | 3.1738839 | 2.91E-06 | NA | NA | NA | NA |
| rs720372 | 8 | 55628637 | G | A | 0.346793349 | 292 | 2.09872734 | 0.44886259 | 2.93E-06 | NA | NA | NA | NA |
| rs567080482 | 13 | 95075405 | T | C | 0.004750594 | 4 | 15.4999561 | 3.31620837 | 2.95E-06 | NA | NA | NA | NA |
| rs546286713 | 13 | 91411079 | G | A | 0.004750594 | 4 | 15.7027841 | 3.3611216 | 2.98E-06 | NA | NA | NA | NA |
| rs142311947 | 5 | 177384469 | G | A | 0.007125891 | 8 | 12.0339013 | 2.57618116 | 2.99E-06 | NA | NA | NA | NA |
| rs185155853 | 15 | 41244100 | C | T | 0.004750594 | 5 | 15.9164657 | 3.40838631 | 3.02E-06 | NA | NA | NA | NA |
| rs550763536 | 2 | 7452347 | T | G | 0.005938242 | 5 | 14.4871647 | 3.10349768 | 3.04E-06 | NA | NA | NA | NA |
| rs555040883 | 22 | 40631476 | G | A | 0.004750594 | 4 | 15.4273069 | 3.30542717 | 3.05E-06 | NA | NA | NA | NA |
| rs2375219 | 8 | 55698295 | C | T | 0.393111639 | 330 | 2.05700167 | 0.44073069 | 3.05E-06 | NA | NA | NA | NA |
| rs140062526 | 13 | 59286033 | G | A | 0.005938242 | 5 | 14.0926088 | 3.02052903 | 3.08E-06 | NA | NA | NA | NA |
| rs572961122 | 13 | 108012985 | C | T | 0.005938242 | 6 | 13.3354896 | 2.85827607 | 3.08E-06 | NA | NA | NA | NA |
| rs544042801 | 11 | 68460243 | G | A | 0.004750594 | 4 | 15.8869866 | 3.40519019 | 3.08E-06 | NA | NA | NA | NA |
| rs142993106 | 4 | 90957372 | G | A | 0.017814727 | 15 | 8.08463611 | 1.73306162 | 3.09E-06 | NA | NA | NA | NA |
| rs569916471 | 14 | 75891342 | G | A | 0.004750594 | 5 | 15.6404088 | 3.3537133 | 3.11E-06 | NA | NA | NA | NA |
| rs7104959 | 11 | 129846126 | C | T | 0.003562945 | 3 | 17.9204611 | 3.84366851 | 3.13E-06 | NA | NA | NA | NA |
| rs559228693 | 20 | 15963329 | G | A | 0.005938242 | 5 | 14.3643551 | 3.08196497 | 3.15E-06 | NA | NA | NA | NA |
| rs187384541 | 8 | 1632241 | A | G | 0.035629454 | 26 | 6.88745117 | 1.47776668 | 3.15E-06 | NA | NA | NA | NA |
| rs141127122 | 22 | 40604439 | G | A | 0.004750594 | 4 | 15.3824875 | 3.30083549 | 3.16E-06 | NA | NA | NA | NA |
| rs193093906 | 10 | 126705489 | G | A | 0.009501188 | 9 | 10.5219537 | 2.25840517 | 3.18E-06 | NA | NA | NA | NA |

| | | | | | | | | | | | | | |
|---|---|---|---|---|---|---|---|---|---|---|---|---|---|
| rs77871739 | 9 | 138552309 | G | A | 0.004750594 | 4 | 15.0724603 | 3.23606181 | 3.20E-06 | NA | NA | NA | NA |
| rs117816016 | 8 | 103751262 | C | T | 0.003562945 | 3 | 19.8807023 | 4.26960212 | 3.22E-06 | NA | NA | NA | NA |
| rs775626702 | 5 | 162630067 | A | C | 0.003562945 | 3 | 18.9484137 | 4.07023372 | 3.23E-06 | NA | NA | NA | NA |
| rs146048121 | 6 | 142198955 | G | A | 0.002375297 | 3 | 20.0214379 | 4.30086025 | 3.24E-06 | NA | NA | NA | NA |
| rs139493286 | 18 | 28816019 | G | A | 0.003562945 | 3 | 17.4603093 | 3.75076428 | 3.24E-06 | NA | NA | NA | NA |
| rs146442492 | 15 | 58982115 | C | T | 0.027315914 | 24 | 7.05925858 | 1.51661316 | 3.25E-06 | NA | NA | NA | NA |
| rs181259864 | 7 | 97488823 | C | A | 0.003562945 | 3 | 17.1360821 | 3.68242789 | 3.26E-06 | NA | NA | NA | NA |
| rs150946694 | 22 | 46853180 | T | C | 0.004750594 | 4 | 15.3082479 | 3.29125059 | 3.30E-06 | NA | NA | NA | NA |
| rs181933850 | 5 | 91465647 | A | G | 0.008313539 | 7 | 11.0436586 | 2.37544216 | 3.33E-06 | NA | NA | NA | NA |
| rs113006316 | 2 | 74802360 | A | G | 0.002375297 | 3 | 22.8886792 | 4.92411335 | 3.35E-06 | NA | NA | NA | NA |
| rs191006910 | 5 | 154216009 | G | A | 0.003562945 | 3 | 17.6173803 | 3.79049689 | 3.36E-06 | NA | NA | NA | NA |
| rs176786 | 14 | 46282970 | T | C | 0.32304038 | 268 | 2.28475641 | 0.49171936 | 3.38E-06 | NA | NA | NA | NA |
| rs187047882 | 3 | 164285621 | G | A | 0.003562945 | 3 | 18.5641361 | 3.99725417 | 3.41E-06 | NA | NA | NA | NA |
| rs73057656 | 3 | 33940571 | A | G | 0.036817102 | 33 | 5.9238329 | 1.27589711 | 3.44E-06 | NA | NA | NA | NA |
| rs112557251 | 2 | 188375400 | T | C | 0.002375297 | 3 | 20.6510064 | 4.44850106 | 3.45E-06 | NA | NA | NA | NA |
| rs190190051 | 5 | 91475485 | G | A | 0.008313539 | 7 | 11.0589887 | 2.38317975 | 3.48E-06 | NA | NA | NA | NA |
| rs180989936 | 1 | 193044178 | A | G | 0.004750594 | 4 | 17.1793579 | 3.70215494 | 3.48E-06 | NA | NA | NA | NA |
| rs188415494 | 8 | 25613298 | C | T | 0.003562945 | 3 | 18.5169478 | 3.9914865 | 3.50E-06 | NA | NA | NA | NA |
| rs1686289 | 14 | 46260982 | G | A | 0.678147268 | 277 | -2.2551539 | 0.48625976 | 3.52E-06 | NA | NA | NA | NA |
| rs76904423 | 12 | 101188744 | G | A | 0.010688836 | 10 | 9.80576231 | 2.11568908 | 3.57E-06 | NA | NA | NA | NA |
| rs183466664 | 12 | 26821687 | A | G | 0.004750594 | 4 | 16.0181882 | 3.45609016 | 3.57E-06 | NA | NA | NA | NA |
| rs999769259 | 4 | 62511965 | G | A | 0.003562945 | 3 | 22.1246725 | 4.77597881 | 3.61E-06 | NA | NA | NA | NA |
| rs553840536 | 16 | 25697895 | A | G | 0.003562945 | 3 | 19.6203442 | 4.23547954 | 3.61E-06 | NA | NA | NA | NA |
| rs183962155 | 4 | 21259643 | A | C | 0.005938242 | 6 | 13.7646008 | 2.97247759 | 3.64E-06 | NA | NA | NA | NA |
| rs138480898 | 1 | 184955657 | C | T | 0.003562945 | 3 | 16.9873974 | 3.66972373 | 3.67E-06 | NA | NA | NA | NA |
| rs185158855 | 2 | 223650026 | C | A | 0.004750594 | 4 | 16.6678864 | 3.60266524 | 3.72E-06 | NA | NA | NA | NA |
| rs180926150 | 1 | 103226326 | C | T | 0.002375297 | 3 | 19.641918 | 4.24793649 | 3.77E-06 | NA | NA | NA | NA |
| rs146728064 | 17 | 19265440 | G | A | 0.007125891 | 6 | 12.6400411 | 2.73467115 | 3.80E-06 | NA | NA | NA | NA |
| rs111900874 | 7 | 89516669 | G | A | 0.014251781 | 12 | 8.42424013 | 1.82258334 | 3.80E-06 | NA | NA | NA | NA |
| rs574076561 | 7 | 49544747 | A | G | 0.003562945 | 3 | 19.6434766 | 4.25225723 | 3.85E-06 | NA | NA | NA | NA |
| rs76554191 | 2 | 95967628 | G | A | 0.040380048 | 34 | 5.72148039 | 1.2389174 | 3.87E-06 | NA | NA | NA | NA |
| rs176783 | 14 | 46280913 | A | G | 0.321852732 | 267 | 2.28010147 | 0.49376374 | 3.88E-06 | NA | NA | NA | NA |
| rs140352232 | 2 | 108038112 | G | A | 0.002375297 | 3 | 21.0576511 | 4.56013129 | 3.88E-06 | NA | NA | NA | NA |
| rs139062456 | 8 | 13251991 | C | T | 0.017814727 | 15 | 7.83515809 | 1.69687423 | 3.89E-06 | NA | NA | NA | NA |
| rs558614420 | 15 | 41810870 | C | T | 0.005938242 | 6 | 13.9750831 | 3.02772732 | 3.92E-06 | NA | NA | NA | NA |
| rs12266995 | 10 | 24852783 | T | C | 0.03087886 | 26 | 5.96782535 | 1.29311959 | 3.93E-06 | NA | NA | NA | NA |
| rs192750513 | 7 | 97577830 | A | G | 0.003562945 | 3 | 17.1892469 | 3.72514218 | 3.94E-06 | NA | NA | NA | NA |
| rs56302696 | 12 | 48292830 | G | A | 0.002375297 | 3 | 19.8959041 | 4.31584707 | 4.03E-06 | NA | NA | NA | NA |
| rs151015676 | 5 | 177390937 | T | G | 0.002375297 | 3 | 20.1421367 | 4.37074482 | 4.06E-06 | NA | NA | NA | NA |
| rs72837643 | 5 | 170188955 | T | C | 0.003562945 | 4 | 16.9302874 | 3.67743479 | 4.15E-06 | NA | NA | NA | NA |
| rs143048774 | 14 | 46294660 | A | C | 0.317102138 | 265 | 2.28482299 | 0.49630858 | 4.15E-06 | NA | NA | NA | NA |
| rs428110 | 14 | 46294660 | A | C | 0.317102138 | 265 | 2.28482299 | 0.49630858 | 4.15E-06 | NA | NA | NA | NA |
| rs111391231 | 7 | 89497045 | T | C | 0.014251781 | 12 | 8.31717755 | 1.80720714 | 4.18E-06 | NA | NA | NA | NA |
| rs180765647 | 13 | 114427311 | G | T | 0.002375297 | 3 | 20.1246521 | 4.37397806 | 4.20E-06 | NA | NA | NA | NA |
| rs568658857 | 12 | 49853998 | G | A | 0.005938242 | 5 | 14.2361193 | 3.09423422 | 4.21E-06 | NA | NA | NA | NA |
| rs182531466 | 5 | 91530073 | C | A | 0.007125891 | 6 | 12.6804978 | 2.75685261 | 4.23E-06 | NA | NA | NA | NA |
| rs181812512 | 11 | 66665729 | C | T | 0.003562945 | 3 | 19.5093168 | 4.24219816 | 4.25E-06 | NA | NA | NA | NA |
| rs556680896 | 13 | 101602415 | C | T | 0.003562945 | 3 | 17.0254659 | 3.70286799 | 4.27E-06 | NA | NA | NA | NA |
| rs142928734 | 13 | 101601082 | G | A | 0.003562945 | 3 | 17.009593 | 3.70044989 | 4.29E-06 | NA | NA | NA | NA |
| rs143287889 | 4 | 35572280 | C | T | 0.002375297 | 3 | 19.96741 | 4.34406 | 4.30E-06 | NA | NA | NA | NA |
| rs543844012 | 9 | 30107023 | C | T | 0.003562945 | 3 | 18.1160485 | 3.9446103 | 4.38E-06 | NA | NA | NA | NA |
| rs566018180 | 10 | 86755052 | C | T | 0.003562945 | 3 | 18.563552 | 4.04215862 | 4.38E-06 | NA | NA | NA | NA |
| rs147171192 | 4 | 89135588 | A | G | 0.007125891 | 5 | 13.3044047 | 2.89758492 | 4.40E-06 | NA | NA | NA | NA |
| rs137873790 | 5 | 97087041 | A | G | 0.013064133 | 11 | 9.22240881 | 2.01095379 | 4.52E-06 | NA | NA | NA | NA |
| rs752259256 | 10 | 104925319 | T | C | 0.003562945 | 3 | 20.4258639 | 4.45411591 | 4.52E-06 | NA | NA | NA | NA |
| rs185771987 | 5 | 73285489 | T | C | 0.003562945 | 3 | 18.2344824 | 3.97675014 | 4.53E-06 | NA | NA | NA | NA |
| rs557092705 | 20 | 34189911 | C | T | 0.003562945 | 3 | 19.4769546 | 4.2480073 | 4.54E-06 | NA | NA | NA | NA |
| rs140642138 | 15 | 42125165 | G | A | 0.005938242 | 6 | 13.5832221 | 2.96274049 | 4.55E-06 | NA | NA | NA | NA |

| | | | | | | | | | | | | | |
|---|---|---|---|---|---|---|---|---|---|---|---|---|---|
| rs139877408 | 2 | 129606273 | A | G | 0.002375297 | 3 | 20.3067129 | 4.43052497 | 4.58E-06 | NA | NA | NA | NA |
| rs191986449 | 5 | 25745687 | C | T | 0.004750594 | 3 | 18.4978793 | 4.03679831 | 4.60E-06 | NA | NA | NA | NA |
| rs11690187 | 2 | 67565909 | A | C | 0.005938242 | 5 | 13.6310532 | 2.9751322 | 4.61E-06 | NA | NA | NA | NA |
| rs118040657 | 10 | 3472846 | C | T | 0.008313539 | 8 | 12.1147712 | 2.64434694 | 4.62E-06 | NA | NA | NA | NA |
| rs138109686 | 15 | 42051442 | A | G | 0.005938242 | 6 | 13.5956705 | 2.96761718 | 4.62E-06 | NA | NA | NA | NA |
| rs563167766 | 1 | 102866365 | G | A | 0.002375297 | 3 | 19.415739 | 4.24240395 | 4.73E-06 | NA | NA | NA | NA |
| rs545550279 | 8 | 115552429 | G | T | 0.003562945 | 3 | 19.1812421 | 4.19212988 | 4.75E-06 | NA | NA | NA | NA |
| rs1498183 | 8 | 55716905 | C | T | 0.394299287 | 332 | 2.0245746 | 0.44255141 | 4.77E-06 | NA | NA | NA | NA |
| rs375790303 | 1 | 184530482 | G | A | 0.008313539 | 7 | 11.4240393 | 2.49719192 | 4.77E-06 | NA | NA | NA | NA |
| rs1396896 | 8 | 55695310 | A | G | 0.397862233 | 334 | 1.99853327 | 0.43690452 | 4.78E-06 | NA | NA | NA | NA |
| rs7843693 | 8 | 55692112 | G | A | 0.397862233 | 334 | 1.99856756 | 0.43695394 | 4.79E-06 | NA | NA | NA | NA |
| rs1391462 | 8 | 55699781 | C | A | 0.397862233 | 334 | 1.9984766 | 0.43696027 | 4.79E-06 | NA | NA | NA | NA |
| rs150539922 | 21 | 43276916 | T | C | 0.003562945 | 3 | 16.6696937 | 3.64477381 | 4.79E-06 | NA | NA | NA | NA |
| rs145896760 | 15 | 42119222 | G | A | 0.005938242 | 6 | 13.3688825 | 2.92346809 | 4.81E-06 | NA | NA | NA | NA |
| rs181415102 | 4 | 112689266 | T | C | 0.003562945 | 4 | 18.2631019 | 3.99387944 | 4.81E-06 | NA | NA | NA | NA |
| rs6080 | 15 | 58837933 | C | A | 0.043942993 | 36 | 5.66476745 | 1.2393253 | 4.86E-06 | NA | NA | NA | NA |
| rs138249376 | 10 | 63515829 | T | G | 0.004750594 | 4 | 16.4300711 | 3.59466791 | 4.86E-06 | NA | NA | NA | NA |
| rs529345909 | 11 | 67110852 | A | G | 0.003562945 | 3 | 18.70153 | 4.09206035 | 4.87E-06 | NA | NA | NA | NA |
| rs191930622 | 12 | 48284655 | G | A | 0.002375297 | 3 | 19.6783579 | 4.30610368 | 4.88E-06 | NA | NA | NA | NA |
| rs145116559 | 4 | 112677596 | T | C | 0.003562945 | 4 | 18.2441224 | 3.9932536 | 4.91E-06 | NA | NA | NA | NA |
| rs12678939 | 8 | 55705021 | A | G | 0.394299287 | 331 | 2.01975618 | 0.44220051 | 4.94E-06 | NA | NA | NA | NA |
| rs73586304 | 6 | 142839425 | C | T | 0.003562945 | 3 | 18.2405116 | 3.99428202 | 4.96E-06 | NA | NA | NA | NA |
| rs113767990 | 14 | 81717563 | G | A | 0.004750594 | 4 | 14.5811155 | 3.19363095 | 4.98E-06 | NA | NA | NA | NA |

**Supplementary Table S7. Independent Replication**

Florida-1 cohort, gene-based replication of top 26 prioritized genes

| Gene | Chr | Start | End | No.SNPs | Pvalue | Pvalue_adjusted |
|---|---|---|---|---|---|---|
| RBFOX1 | 16 | 6069095 | 7763340 | 54 | 0.00026108 | 0.003394054 |
| CSMD1 | 8 | 2792875 | 4852494 | 50 | 0.00333806 | 0.021697393 |
| HDAC9 | 7 | 18126572 | 19042039 | 13 | 0.02338453 | 0.101332968 |
| PRKD1 | 14 | 30045687 | 30661104 | 5 | 0.03541744 | 0.115106687 |
| NCAM2 | 21 | 22370633 | 22915650 | 5 | 0.05728018 | 0.139142201 |
| PPM1H | 12 | 63037762 | 63328817 | 4 | 0.06421948 | 0.139142201 |
| SPTY2D1 | 11 | 18627948 | 18656338 | 1 | 0.07862637 | 0.146020404 |
| AGAP1 | 2 | 236402733 | 237040444 | 2 | 0.09194578 | 0.149411895 |
| SGCG | 13 | 23755091 | 23899304 | 1 | 0.12415358 | 0.179332944 |
| HDAC4 | 2 | 239969864 | 240323348 | 1 | 0.27330078 | 0.35529101 |
| GPLD1 | 6 | 24424793 | 24495433 | 2 | 0.5232773 | 0.618418633 |
| VTA1 | 6 | 142468367 | 142545826 | 1 | 0.65065227 | 0.704873297 |
| WWC1 | 5 | 167718656 | 167899308 | 1 | 0.90507783 | 0.90507783 |
| ARHGEF26 | 3 | 153838792 | 153975616 | 0 | | |
| ASB3 | 2 | 53897117 | 54014090 | 0 | | |
| BEND7 | 10 | 13480484 | 13570974 | 0 | | |
| C4orf19 | 4 | 37455563 | 37625117 | 0 | | |
| CCR6 | 6 | 167525295 | 167553184 | 0 | | |
| COL11A1 | 1 | 103342023 | 103574052 | 0 | | |
| DIRAS2 | 9 | 93372114 | 93405386 | 0 | | |
| FST | 5 | 52776239 | 52782964 | 0 | | |
| KAZALD1 | 10 | 102821598 | 102827888 | 0 | | |
| PPARGC1A | 4 | 23756664 | 23905712 | 0 | | |
| USP25 | 21 | 17102344 | 17252377 | 0 | | |
| ZNF728 | 19 | 23158270 | 23185978 | 0 | | |
| ZNF737 | 19 | 20718631 | 20748615 | 0 | | |

*signed MB score test that collapse SNPs with MAF < 0.01

**Supplementary Table S7. Independent Replication**

Chennai-1 cohort, case-control data, comparing to discovery 12M quantitative trait (QT) data

Threshold for replicative significance: 5.00E-02
Total SNPs found: 37
Total risk loci found: 21

NA: not applicable

| rsid | chr | pos_37 | REF | ALT | n.obs | caf | MAC | Est | Est.SE | Score.pval | pval.chennai | adj pval SNPs | adj pval loci | Closest gene | Prioritized gene |
|---|---|---|---|---|---|---|---|---|---|---|---|---|---|---|---|
| rs76098744 | 1 | 112349372 | C | T | 439 | 0.01594533 | 14 | 9.5685657 | 2.04635193 | 2.93E-06 | 6.78E-02 | 1.00E+00 | 1.00E+00 | KCND3 | KCND3 |
| rs74683551 | 1 | 112354418 | G | A | 439 | 0.01594533 | 14 | 9.54722832 | 2.04640679 | 3.08E-06 | 6.78E-02 | 1.00E+00 | 1.00E+00 | KCND3 | KCND3 |
| rs2606194 | 17 | 77210823 | A | G | 439 | 0.948747153 | 44 | -5.6956979 | 1.16435773 | 1.00E-06 | 9.08E-02 | 1.00E+00 | 1.00E+00 | RBFOX3 | RBFOX3 |
| rs7534177 | 1 | 65965720 | A | G | 439 | 0.056947608 | 49 | 4.96123378 | 1.0598616 | 2.85E-06 | 1.44E-01 | 1.00E+00 | 1.00E+00 | LEPR | LEPR |
| rs2427460 | 20 | 61590782 | T | C | 439 | 0.486332574 | 420 | -2.574044 | 0.51325892 | 5.30E-07 | 1.72E-01 | 1.00E+00 | 1.00E+00 | SLC17A9 | SLC17A9 |
| rs77297738 | 22 | 34473543 | C | T | 439 | 0.018223235 | 16 | 10.2480572 | 1.97167592 | 2.02E-07 | 1.98E-01 | 1.00E+00 | 1.00E+00 | LARGE1 | LARGE1 |
| rs74572772 | 22 | 34492649 | A | G | 439 | 0.018223235 | 16 | 10.2225183 | 1.94126774 | 1.40E-07 | 1.98E-01 | 1.00E+00 | 1.00E+00 | LARGE1 | LARGE1 |
| rs80019988 | 22 | 34493647 | G | A | 439 | 0.018223235 | 16 | 10.0623306 | 1.91665407 | 1.52E-07 | 1.98E-01 | 1.00E+00 | 1.00E+00 | LARGE1 | LARGE1 |
| rs17127656 | 1 | 65943471 | C | T | 439 | 0.055808656 | 49 | 5.07046538 | 1.05780641 | 1.64E-06 | 2.75E-01 | 1.00E+00 | 1.00E+00 | LEPR | LEPR |
| rs7518849 | 1 | 65948791 | T | C | 439 | 0.055808656 | 49 | 5.07456956 | 1.05964469 | 1.68E-06 | 2.75E-01 | 1.00E+00 | 1.00E+00 | LEPR | LEPR |
| rs11579567 | 1 | 65957141 | C | A | 439 | 0.055808656 | 49 | 5.02973793 | 1.06097379 | 2.13E-06 | 2.75E-01 | 1.00E+00 | 1.00E+00 | LEPR | LEPR |
| rs35145334 | 1 | 23465122 | A | G | 439 | 0.017084282 | 14 | 10.6552656 | 2.24620792 | 2.10E-06 | 2.96E-01 | 1.00E+00 | 1.00E+00 | LUZP1 | LUZP1 |
| rs142956968 | 10 | 23490165 | C | T | 439 | 0.01594533 | 14 | 10.4167992 | 2.11909083 | 8.85E-07 | 2.96E-01 | 1.00E+00 | 1.00E+00 | C10orf67 | OTUD1 |
| rs145764464 | 10 | 23601847 | G | A | 439 | 0.017084282 | 15 | 9.5967306 | 1.96976348 | 1.10E-06 | 2.96E-01 | 1.00E+00 | 1.00E+00 | C10orf67 | OTUD1 |
| rs117699122 | 12 | 560262 | T | C | 439 | 0.003416856 | 4 | 21.4238256 | 4.40952328 | 1.18E-06 | 2.96E-01 | 1.00E+00 | 1.00E+00 | CCDC77 | CCDC77 |
| rs149616342 | 12 | 99178813 | C | T | 439 | 0.011389522 | 10 | 11.2978214 | 2.454657 | 4.17E-06 | 2.96E-01 | 1.00E+00 | 1.00E+00 | ANKS1B | ANKS1B |
| rs62192733 | 20 | 1928157 | A | G | 439 | 0.006833713 | 7 | 15.744618 | 3.35866563 | 2.76E-06 | 2.96E-01 | 1.00E+00 | 1.00E+00 | PDYN | SIRPA |
| rs117591241 | 3 | 190584679 | A | G | 439 | 0.003416856 | 4 | 19.9014028 | 4.33331389 | 4.38E-06 | 3.34E-01 | 1.00E+00 | 1.00E+00 | GMNC | GMNC |
| rs17615362 | 4 | 169934087 | G | A | 439 | 0.050113895 | 44 | 5.41974472 | 1.17918425 | 4.30E-06 | 3.34E-01 | 1.00E+00 | 1.00E+00 | CBR4 | CBR4 |
| rs17543620 | 4 | 169934725 | T | C | 439 | 0.050113895 | 44 | 5.42796755 | 1.17982097 | 4.21E-06 | 3.34E-01 | 1.00E+00 | 1.00E+00 | CBR4 | CBR4 |
| rs62515405 | 8 | 57055978 | G | A | 439 | 0.023917995 | 21 | 8.07691204 | 1.76428755 | 4.69E-06 | 3.34E-01 | 1.00E+00 | 1.00E+00 | PLAG1 | PLAG1 |
| rs113244573 | 12 | 6684385 | G | A | 439 | 0.023917995 | 21 | 7.97095746 | 1.73766156 | 4.49E-06 | 3.34E-01 | 1.00E+00 | 1.00E+00 | CHD4 | CHD4 |
| rs139816293 | 20 | 30921343 | C | T | 439 | 0.005694761 | 6 | 17.1384248 | 3.52537223 | 1.17E-06 | 3.34E-01 | 1.00E+00 | 1.00E+00 | KIF3B | KIF3B |
| rs76356799 | 3 | 179593768 | G | A | 439 | 0.004555809 | 4 | 19.2867007 | 3.76039319 | 2.91E-07 | 5.13E-01 | 1.00E+00 | 1.00E+00 | PEX5L | PEX5L |
| rs147944608 | 10 | 110423709 | C | T | 439 | 0.012528474 | 11 | 11.2205243 | 2.29229161 | 9.84E-07 | 5.84E-01 | 1.00E+00 | 1.00E+00 | XPNPEP1 | XPNPEP1 |
| rs4896997 | 6 | 148683924 | C | T | 439 | 0.006833713 | 6 | 14.4320923 | 3.09188376 | 3.05E-06 | 6.04E-01 | 1.00E+00 | 1.00E+00 | SASH1 | SASH1 |
| rs17078283 | 6 | 148696920 | C | T | 439 | 0.006833713 | 6 | 14.4302821 | 3.0918048 | 3.05E-06 | 6.04E-01 | 1.00E+00 | 1.00E+00 | SASH1 | SASH1 |
| rs56821264 | 6 | 148703901 | C | T | 439 | 0.006833713 | 6 | 14.7509213 | 3.11419566 | 2.17E-06 | 6.04E-01 | 1.00E+00 | 1.00E+00 | SASH1 | SASH1 |
| rs9981433 | 21 | 41309565 | G | T | 439 | 0.881548975 | 116 | -4.1220124 | 0.84715422 | 1.14E-06 | 7.75E-01 | 1.00E+00 | 1.00E+00 | PCP4 | PCP4 |
| rs12114488 | 8 | 62672723 | G | A | 439 | 0.30523918 | 266 | 2.62307914 | 0.54257702 | 1.33E-06 | 8.99E-01 | 1.00E+00 | 1.00E+00 | MIR4470 | ASPH |
| rs9636964 | 21 | 41304765 | A | G | 439 | 0.884965831 | 115 | -4.2543192 | 0.82744371 | 2.73E-07 | 9.23E-01 | 1.00E+00 | 1.00E+00 | PCP4 | PCP4 |
| rs9305683 | 21 | 41305720 | G | A | 439 | 0.88952164 | 112 | -4.1163679 | 0.83634363 | 8.57E-07 | 9.23E-01 | 1.00E+00 | 1.00E+00 | PCP4 | PCP4 |
| rs9974985 | 21 | 41307573 | G | A | 439 | 0.886104784 | 114 | -4.3459395 | 0.82924282 | 1.60E-07 | 9.23E-01 | 1.00E+00 | 1.00E+00 | PCP4 | PCP4 |
| rs7275595 | 21 | 41307923 | G | A | 439 | 0.888382688 | 113 | -4.2609754 | 0.83407868 | 3.25E-07 | 9.23E-01 | 1.00E+00 | 1.00E+00 | PCP4 | PCP4 |
| rs1005412 | 21 | 41308948 | A | G | 439 | 0.887243736 | 112 | -4.4521096 | 0.85237138 | 1.76E-07 | 9.23E-01 | 1.00E+00 | 1.00E+00 | PCP4 | PCP4 |
| rs75082290 | 2 | 67831153 | T | G | 439 | 0.078587699 | 69 | 4.69395387 | 0.98072733 | 1.70E-06 | 9.56E-01 | 1.00E+00 | 1.00E+00 | ETAA1 | ETAA1 |
| rs61918041 | 12 | 6681868 | C | T | 439 | 0.023917995 | 21 | 7.97264643 | 1.73636056 | 4.40E-06 | 9.56E-01 | 1.00E+00 | 1.00E+00 | CHD4 | CHD4 |
| rs139390630 | 1 | 84866196 | G | A | 439 | 0.011389522 | 13 | 11.6975971 | 2.4994863 | 2.87E-06 | NA | | | | |
| rs114067899 | 1 | 85191044 | G | A | 439 | 0.007972665 | 7 | 13.7760673 | 2.90836365 | 2.17E-06 | NA | | | | |
| rs145804766 | 1 | 202122499 | C | T | 439 | 0.010250569 | 9 | 12.5864465 | 2.61118962 | 1.43E-06 | NA | | | | |
| rs147869916 | 1 | 227307652 | A | G | 439 | 0.005694761 | 5 | 17.5964551 | 3.78625881 | 3.36E-06 | NA | | | | |
| rs76220567 | 2 | 13059945 | T | C | 439 | 0.003416856 | 4 | 20.6828868 | 4.35776216 | 2.07E-06 | NA | | | | |
| rs536781978 | 2 | 29733681 | A | G | 439 | 0.003416856 | 3 | 20.5561882 | 4.34258099 | 2.21E-06 | NA | | | | |

| | | | | | | | | | | | |
|---|---|---|---|---|---|---|---|---|---|---|---|
| rs568321148 | 2 | 29870348 | T | G | 439 | 0.003416856 | 3 | 20.7400203 | 4.36652717 | 2.04E-06 | NA |
| rs186792608 | 2 | 43096699 | A | G | 439 | 0.003416856 | 3 | 22.4512499 | 4.28172841 | 1.58E-07 | NA |
| rs1162141034 | 2 | 67825685 | C | T | 439 | 0.079726651 | 71 | 4.55991411 | 0.98044721 | 3.31E-06 | NA |
| rs146465493 | 2 | 67825685 | C | T | 439 | 0.079726651 | 71 | 4.55991411 | 0.98044721 | 3.31E-06 | NA |
| rs2902021 | 2 | 67825685 | C | T | 439 | 0.079726651 | 71 | 4.55991411 | 0.98044721 | 3.31E-06 | NA |
| rs191298981 | 2 | 68110046 | C | T | 439 | 0.004555809 | 3 | 24.0078714 | 4.85703224 | 7.70E-07 | NA |
| rs113537164 | 2 | 68234930 | A | C | 439 | 0.004555809 | 4 | 20.3890413 | 4.17097432 | 1.02E-06 | NA |
| rs113154814 | 2 | 68289497 | T | C | 439 | 0.005694761 | 5 | 19.8310188 | 3.7892894 | 1.66E-07 | NA |
| rs146919974 | 2 | 115252863 | T | C | 439 | 0.004555809 | 5 | 19.7280466 | 3.99554232 | 7.91E-07 | NA |
| rs79935606 | 2 | 177338635 | T | C | 439 | 0.005694761 | 5 | 16.3396833 | 3.47376974 | 2.55E-06 | NA |
| rs144788248 | 3 | 19586745 | T | C | 439 | 0.011389522 | 10 | 11.9086237 | 2.57879124 | 3.88E-06 | NA |
| rs149706477 | 3 | 66148228 | A | G | 439 | 0.003416856 | 3 | 20.9828687 | 4.4842531 | 2.88E-06 | NA |
| rs114280794 | 3 | 132842203 | G | A | 439 | 0.01594533 | 14 | 8.76296877 | 1.89632016 | 3.82E-06 | NA |
| rs148248743 | 3 | 136134595 | C | T | 439 | 0.002277904 | 3 | 27.1548587 | 5.46352726 | 6.69E-07 | NA |
| rs138138661 | 3 | 147657814 | C | T | 439 | 0.004555809 | 4 | 23.1347457 | 4.25695836 | 5.49E-08 | NA |
| rs189234695 | 3 | 147658158 | T | C | 439 | 0.003416856 | 3 | 23.9697134 | 5.16542129 | 3.48E-06 | NA |
| rs148997617 | 3 | 147755576 | C | T | 439 | 0.004555809 | 4 | 20.8774277 | 3.92648902 | 1.05E-07 | NA |
| rs148483098 | 3 | 157382051 | C | A | 439 | 0.005694761 | 5 | 15.7357844 | 3.38211024 | 3.28E-06 | NA |
| rs79252854 | 3 | 157523157 | C | T | 439 | 0.004555809 | 5 | 17.5464336 | 3.72541417 | 2.48E-06 | NA |
| rs139055031 | 3 | 158586849 | T | C | 439 | 0.005694761 | 6 | 16.2853761 | 3.4955337 | 3.18E-06 | NA |
| rs143669489 | 3 | 159392885 | G | A | 439 | 0.003416856 | 3 | 24.2253199 | 5.30400674 | 4.94E-06 | NA |
| rs189709453 | 3 | 177574308 | G | A | 439 | 0.003416856 | 3 | 23.2009936 | 4.82954374 | 1.56E-06 | NA |
| rs186767531 | 3 | 177639777 | T | C | 439 | 0.003416856 | 3 | 23.0749977 | 4.81664642 | 1.66E-06 | NA |
| rs182868205 | 3 | 177712448 | C | T | 439 | 0.003416856 | 3 | 22.8544303 | 4.73215662 | 1.37E-06 | NA |
| rs1472243465 | 3 | 190584679 | A | G | 439 | 0.003416856 | 4 | 19.9014028 | 4.33331389 | 4.38E-06 | NA |
| rs189094663 | 4 | 11623450 | G | A | 439 | 0.002277904 | 3 | 23.895416 | 5.10822708 | 2.90E-06 | NA |
| rs557276277 | 4 | 19013698 | T | C | 439 | 0.005694761 | 5 | 19.0146344 | 4.09137627 | 3.36E-06 | NA |
| rs113063005 | 4 | 23509067 | T | C | 439 | 0.006833713 | 7 | 16.61763 | 3.39113209 | 9.57E-07 | NA |
| rs147267707 | 4 | 43026558 | G | A | 439 | 0.004555809 | 4 | 18.0094385 | 3.80818408 | 2.25E-06 | NA |
| rs77618729 | 4 | 43050968 | T | C | 439 | 0.004555809 | 4 | 18.0130153 | 3.80914173 | 2.26E-06 | NA |
| rs556435089 | 4 | 43551033 | G | A | 439 | 0.004555809 | 4 | 17.939079 | 3.81772146 | 2.62E-06 | NA |
| rs532464521 | 4 | 43571039 | T | C | 439 | 0.004555809 | 4 | 17.9895528 | 3.81820438 | 2.46E-06 | NA |
| rs181132315 | 4 | 129647369 | C | T | 439 | 0.003416856 | 3 | 22.1324679 | 4.62266164 | 1.69E-06 | NA |
| rs147455971 | 4 | 140931712 | T | C | 439 | 0.007972665 | 5 | 17.2160142 | 3.72793057 | 3.87E-06 | NA |
| rs142934021 | 5 | 52662672 | G | A | 439 | 0.007972665 | 7 | 18.824715 | 3.31820294 | 1.40E-08 | NA |
| rs189912648 | 5 | 134765439 | C | T | 439 | 0.003416856 | 4 | 19.8604449 | 4.2716652 | 3.33E-06 | NA |
| rs111512950 | 5 | 153680427 | C | T | 439 | 0.009111617 | 8 | 13.567443 | 2.94598493 | 4.12E-06 | NA |
| rs73384619 | 6 | 20458179 | C | T | 439 | 0.022779043 | 21 | 8.3916702 | 1.81241389 | 3.65E-06 | NA |
| rs4131286 | 6 | 148688963 | G | T | 439 | 0.006833713 | 6 | 14.4284672 | 3.09172532 | 3.06E-06 | NA |
| rs917335559 | 6 | 148703901 | C | T | 439 | 0.006833713 | 6 | 14.7509213 | 3.11419566 | 2.17E-06 | NA |
| rs143686474 | 7 | 16291999 | A | C | 439 | 0.012528474 | 11 | 10.8531498 | 2.37513184 | 4.89E-06 | NA |
| rs73202425 | 7 | 109155716 | T | C | 439 | 0.012528474 | 12 | 10.761844 | 2.30582166 | 3.05E-06 | NA |
| rs181182636 | 7 | 115654558 | A | G | 439 | 0.003416856 | 3 | 21.9472639 | 4.69244743 | 2.91E-06 | NA |
| rs182211730 | 7 | 116003615 | G | A | 439 | 0.004555809 | 4 | 18.7746033 | 3.8971981 | 1.45E-06 | NA |
| rs143281973 | 7 | 116007996 | C | T | 439 | 0.004555809 | 4 | 18.2750778 | 3.81098198 | 1.62E-06 | NA |
| rs138873576 | 7 | 116025359 | G | T | 439 | 0.004555809 | 4 | 17.6151711 | 3.78331079 | 3.22E-06 | NA |
| rs190354334 | 7 | 116075846 | G | A | 439 | 0.004555809 | 5 | 17.3392581 | 3.7718663 | 4.29E-06 | NA |
| rs186893139 | 7 | 116096970 | C | T | 439 | 0.004555809 | 4 | 18.602598 | 4.01258383 | 3.55E-06 | NA |
| rs558784715 | 7 | 117043294 | A | G | 439 | 0.003416856 | 3 | 20.7591787 | 4.31689475 | 1.52E-06 | NA |
| rs576047962 | 7 | 117109021 | A | G | 439 | 0.003416856 | 3 | 20.7261549 | 4.32140182 | 1.62E-06 | NA |
| rs529110230 | 7 | 117175907 | T | C | 439 | 0.003416856 | 3 | 20.8583633 | 4.30235088 | 1.25E-06 | NA |
| rs142215699 | 7 | 117199874 | T | C | 439 | 0.003416856 | 3 | 20.7292287 | 4.32245361 | 1.62E-06 | NA |

| | | | | | | | | | | | |
|---|---|---|---|---|---|---|---|---|---|---|---|
| rs142721557 | 7 | 117218328 | A | C | 439 | 0.003416856 | 3 | 20.7329975 | 4.32245894 | 1.61E-06 | NA |
| rs201355675 | 7 | 117225781 | G | A | 439 | 0.003416856 | 3 | 20.718124 | 4.32246722 | 1.64E-06 | NA |
| rs188993522 | 7 | 117274731 | T | C | 439 | 0.003416856 | 3 | 21.2984827 | 4.47867266 | 1.98E-06 | NA |
| rs62515436 | 8 | 57141203 | G | T | 439 | 0.017084282 | 15 | 9.27545902 | 1.99804812 | 3.45E-06 | NA |
| rs144138711 | 9 | 99922419 | T | C | 439 | 0.045558087 | 40 | 6.08547131 | 1.26091918 | 1.39E-06 | NA |
| rs556274646 | 11 | 439012 | C | T | 439 | 0.010250569 | 9 | 12.8914592 | 2.70847728 | 1.94E-06 | NA |
| rs148815783 | 11 | 1013992 | C | T | 439 | 0.007972665 | 8 | 13.5980313 | 2.8647171 | 2.07E-06 | NA |
| rs566724618 | 13 | 23463839 | C | T | 439 | 0.003416856 | 3 | 22.3433705 | 4.58061605 | 1.07E-06 | NA |
| rs147221953 | 13 | 23485722 | G | A | 439 | 0.003416856 | 3 | 22.5809676 | 4.7360336 | 1.86E-06 | NA |
| rs113791989 | 13 | 61918888 | G | A | 439 | 0.002277904 | 3 | 25.2049582 | 5.08402789 | 7.13E-07 | NA |
| rs532269430 | 13 | 74054452 | T | C | 439 | 0.004555809 | 5 | 18.6173752 | 3.7963004 | 9.39E-07 | NA |
| rs368187808 | 13 | 74357508 | A | C | 439 | 0.003416856 | 3 | 21.2991251 | 4.41501637 | 1.41E-06 | NA |
| rs372194899 | 13 | 74357510 | A | G | 439 | 0.003416856 | 3 | 21.3012101 | 4.41516106 | 1.40E-06 | NA |
| rs190251199 | 14 | 105590577 | T | C | 439 | 0.003416856 | 3 | 22.6385061 | 4.76043817 | 1.98E-06 | NA |
| rs187281112 | 15 | 33113387 | C | T | 439 | 0.005694761 | 5 | 14.7343356 | 3.16793649 | 3.30E-06 | NA |
| rs56324718 | 15 | 40280073 | G | A | 439 | 0.010250569 | 9 | 12.5117374 | 2.5288888 | 7.52E-07 | NA |
| rs80212581 | 16 | 6412967 | C | T | 439 | 0.003416856 | 3 | 22.2762003 | 4.47237333 | 6.33E-07 | NA |
| rs140276610 | 16 | 6433735 | C | T | 439 | 0.003416856 | 4 | 22.1806577 | 4.64062401 | 1.76E-06 | NA |
| rs138164904 | 16 | 6858239 | C | T | 439 | 0.003416856 | 3 | 23.892641 | 4.3369669 | 3.61E-08 | NA |
| rs116897913 | 17 | 16839162 | C | T | 439 | 0.003416856 | 3 | 20.1396765 | 4.40294288 | 4.78E-06 | NA |
| rs188353596 | 17 | 46078602 | C | T | 439 | 0.003416856 | 3 | 21.8057947 | 4.39141941 | 6.85E-07 | NA |
| rs112148840 | 17 | 66233774 | C | T | 439 | 0.002277904 | 3 | 24.0241075 | 5.1994476 | 3.83E-06 | NA |
| rs147669485 | 18 | 26603650 | G | T | 439 | 0.003416856 | 4 | 22.1107639 | 4.14923198 | 9.88E-08 | NA |
| rs559152067 | 18 | 66029612 | G | A | 439 | 0.002277904 | 3 | 24.7443692 | 5.18343862 | 1.81E-06 | NA |
| rs367732718 | 19 | 35918004 | G | A | 439 | 0.002277904 | 3 | 23.8784923 | 5.13995752 | 3.39E-06 | NA |
| rs562831582 | 19 | 51234310 | A | C | 439 | 0.003416856 | 4 | 21.0856548 | 4.21624838 | 5.70E-07 | NA |
| rs547186621 | 20 | 6106741 | A | G | 439 | 0.003416856 | 4 | 21.5510829 | 4.5690881 | 2.40E-06 | NA |
| rs138733283 | 20 | 30675687 | C | T | 439 | 0.004555809 | 5 | 18.2744744 | 3.65830138 | 5.87E-07 | NA |
| rs149859280 | 20 | 30678170 | C | T | 439 | 0.004555809 | 5 | 18.3057707 | 3.66075889 | 5.72E-07 | NA |
| rs146249289 | 20 | 30682231 | C | T | 439 | 0.004555809 | 4 | 19.6109527 | 3.77306035 | 2.02E-07 | NA |
| rs145791959 | 20 | 30722429 | G | A | 439 | 0.005694761 | 5 | 17.9091953 | 3.64034112 | 8.67E-07 | NA |
| rs193041547 | 20 | 30771880 | T | C | 439 | 0.004555809 | 4 | 19.6791358 | 3.77081146 | 1.80E-07 | NA |
| rs138055631 | 20 | 30780644 | G | A | 439 | 0.005694761 | 5 | 17.6647762 | 3.62004915 | 1.06E-06 | NA |
| rs145421321 | 20 | 30862126 | C | T | 439 | 0.005694761 | 6 | 17.0212611 | 3.55829648 | 1.72E-06 | NA |
| rs143432612 | 20 | 30892775 | C | T | 439 | 0.005694761 | 6 | 16.9387232 | 3.54318154 | 1.75E-06 | NA |
| rs200198574 | 20 | 30946654 | G | A | 439 | 0.005694761 | 6 | 16.5282735 | 3.50702674 | 2.44E-06 | NA |
| rs148157126 | 20 | 30948839 | C | T | 439 | 0.004555809 | 5 | 19.7865001 | 3.76042103 | 1.43E-07 | NA |
| rs192855100 | 20 | 31108741 | G | A | 439 | 0.004555809 | 4 | 20.1986097 | 3.99111726 | 4.17E-07 | NA |
| rs386818616 | 21 | 41308948 | A | G | 439 | 0.887243736 | 112 | -4.4521096 | 0.85237138 | 1.76E-07 | NA |

**Supplementary Table S7. Independent Replication**

Chennai-1 cohort, case-control data, comparing to discovery 3M quantitative trait (QT) data

| | |
|---|---|
| Threshold for replicative significance: | 5.00E-02 |
| Total SNPs found: | 122 |
| Total risk loci found: | 38 |

Gray shading: SNPs and target genes of genome-wide significance in the discovery cohort
NA: not applicable

| rsid | chr | pos_37 | REF | ALT | n.obs | caf | MAC | Est | Est.SE | Score.pval | pval.chennai | adj pval SNPs | adj pval loci | Closest gene | Prioritized gene |
|---|---|---|---|---|---|---|---|---|---|---|---|---|---|---|---|
| rs17017794 | 4 | 91825885 | T | C | 421 | 0.0391924 | 33 | 5.83240545 | 1.14450838 | 3.47E-07 | 6.39E-02 | 1.00E+00 | 1.00E+00 | CCSER1 | CCSER1 |
| rs75334617 | 10 | 102956152 | G | A | 421 | 0.03800475 | 32 | 5.84603405 | 1.18027345 | 7.30E-07 | 6.39E-02 | 1.00E+00 | 1.00E+00 | LINC01514 | KAZALD1 |
| rs76098744 | 1 | 112349372 | C | T | 421 | 0.01662708 | 14 | 8.38612452 | 1.72704116 | 1.20E-06 | 6.78E-02 | 1.00E+00 | 1.00E+00 | KCND3 | KCND3 |
| rs74683551 | 1 | 112354418 | G | A | 421 | 0.01662708 | 14 | 8.38252043 | 1.72705812 | 1.21E-06 | 6.78E-02 | 1.00E+00 | 1.00E+00 | KCND3 | KCND3 |
| rs17510814 | 12 | 28468969 | A | C | 421 | 0.00712589 | 6 | 14.0387924 | 2.71409689 | 2.31E-07 | 6.78E-02 | 1.00E+00 | 1.00E+00 | CCDC91 | CCDC91 |
| rs141756120 | 12 | 28511096 | A | C | 421 | 0.00831354 | 6 | 14.278403 | 2.76458538 | 2.41E-07 | 6.78E-02 | 1.00E+00 | 1.00E+00 | CCDC91 | CCDC91 |
| rs143371352 | 13 | 47383834 | C | T | 421 | 0.00356295 | 3 | 20.5341625 | 3.87928964 | 1.20E-07 | 6.78E-02 | 1.00E+00 | 1.00E+00 | HTR2A | HTR2A |
| rs9643828 | 8 | 55529073 | C | T | 421 | 0.67695962 | 279 | -2.4236294 | 0.46846911 | 2.30E-07 | 8.50E-02 | 1.00E+00 | 1.00E+00 | RP1 | SOX17 |
| rs147601511 | 8 | 22322319 | A | G | 421 | 0.00475059 | 4 | 16.4471181 | 3.31777038 | 7.15E-07 | 9.08E-02 | 1.00E+00 | 1.00E+00 | PPP3CC | PPP3CC |
| rs112007361 | 11 | 99188380 | A | C | 421 | 0.03087886 | 26 | 6.35658468 | 1.22825758 | 2.28E-07 | 9.08E-02 | 1.00E+00 | 1.00E+00 | CNTN5 | CNTN5 |
| rs76327548 | 12 | 101182966 | G | A | 421 | 0.01306413 | 11 | 9.17842714 | 1.93235231 | 2.04E-06 | 1.13E-01 | 1.00E+00 | 1.00E+00 | ANO4 | ANO4 |
| rs10105693 | 8 | 55640472 | C | T | 421 | 0.32897862 | 276 | 2.2928391 | 0.44965282 | 3.41E-07 | 1.28E-01 | 1.00E+00 | 1.00E+00 | RP1 | SOX17 |
| rs11987234 | 8 | 55669829 | A | G | 421 | 0.32897862 | 276 | 2.29599259 | 0.44884532 | 3.13E-07 | 1.28E-01 | 1.00E+00 | 1.00E+00 | RP1 | SOX17 |
| rs7822082 | 8 | 55690220 | T | C | 421 | 0.33016627 | 277 | 2.3570878 | 0.44879289 | 1.50E-07 | 1.28E-01 | 1.00E+00 | 1.00E+00 | RP1 | SOX17 |
| rs149421869 | 2 | 53483429 | G | T | 421 | 0.00831354 | 8 | 11.7382186 | 2.47204376 | 2.05E-06 | 1.38E-01 | 1.00E+00 | 1.00E+00 | ASB3 | ASB3 |
| rs17776100 | 7 | 6426479 | G | A | 421 | 0.02969121 | 25 | 6.63697397 | 1.32177185 | 5.13E-07 | 1.38E-01 | 1.00E+00 | 1.00E+00 | RAC1 | RAC1 |
| rs2274997 | 1 | 229804646 | A | G | 421 | 0.0415677 | 35 | 5.58731228 | 1.1238603 | 6.64E-07 | 1.44E-01 | 1.00E+00 | 1.00E+00 | URB2 | URB2 |
| rs74521112 | 11 | 99089147 | G | T | 421 | 0.03206651 | 27 | 6.25632387 | 1.22787586 | 3.48E-07 | 1.69E-01 | 1.00E+00 | 1.00E+00 | CNTN5 | CNTN5 |
| rs79213709 | 11 | 99093455 | G | A | 421 | 0.03206651 | 26 | 6.17081884 | 1.23453654 | 5.78E-07 | 1.69E-01 | 1.00E+00 | 1.00E+00 | CNTN5 | CNTN5 |
| rs1498183 | 8 | 55716905 | C | T | 421 | 0.39429929 | 332 | 2.0245746 | 0.44255141 | 4.77E-06 | 1.83E-01 | 1.00E+00 | 1.00E+00 | RP1 | SOX17 |
| rs858397 | 8 | 55614690 | A | G | 421 | 0.33135392 | 278 | 2.2301348 | 0.44990341 | 7.16E-07 | 1.85E-01 | 1.00E+00 | 1.00E+00 | RP1 | SOX17 |
| rs13278605 | 8 | 55688171 | C | T | 421 | 0.32897862 | 277 | 2.27377348 | 0.44593165 | 3.42E-07 | 1.85E-01 | 1.00E+00 | 1.00E+00 | RP1 | SOX17 |
| rs13276543 | 8 | 55688174 | G | T | 421 | 0.32897862 | 276 | 2.29263157 | 0.44823093 | 3.14E-07 | 1.85E-01 | 1.00E+00 | 1.00E+00 | RP1 | SOX17 |
| rs2375219 | 8 | 55698295 | C | T | 421 | 0.39311164 | 330 | 2.05700167 | 0.44073069 | 3.05E-06 | 1.85E-01 | 1.00E+00 | 1.00E+00 | RP1 | SOX17 |
| rs12502861 | 4 | 2426305 | T | C | 421 | 0.01068884 | 9 | 11.1588472 | 2.36961887 | 2.49E-06 | 1.98E-01 | 1.00E+00 | 1.00E+00 | CFAP99 | CFAP99 |
| rs2274996 | 1 | 229804538 | C | T | 421 | 0.0415677 | 35 | 5.58527433 | 1.12406363 | 6.74E-07 | 2.43E-01 | 1.00E+00 | 1.00E+00 | URB2 | URB2 |
| rs2891865 | 1 | 229806368 | A | G | 421 | 0.0415677 | 35 | 5.58181376 | 1.12378916 | 6.80E-07 | 2.43E-01 | 1.00E+00 | 1.00E+00 | URB2 | URB2 |
| rs2385790 | 1 | 229807492 | C | T | 421 | 0.0415677 | 35 | 5.58224822 | 1.12394723 | 6.81E-07 | 2.43E-01 | 1.00E+00 | 1.00E+00 | URB2 | URB2 |
| rs12024557 | 1 | 229812357 | A | C | 421 | 0.04275534 | 35 | 5.56930969 | 1.12195729 | 6.91E-07 | 2.43E-01 | 1.00E+00 | 1.00E+00 | URB2 | URB2 |
| rs4562666 | 1 | 229824770 | T | C | 421 | 0.04275534 | 36 | 5.51922599 | 1.11457457 | 7.35E-07 | 2.43E-01 | 1.00E+00 | 1.00E+00 | URB2 | URB2 |
| rs12036586 | 1 | 229826378 | G | A | 421 | 0.04750594 | 40 | 5.22212176 | 1.07114093 | 1.09E-06 | 2.43E-01 | 1.00E+00 | 1.00E+00 | URB2 | URB2 |
| rs16850124 | 1 | 229831331 | T | C | 421 | 0.04394299 | 37 | 5.18095662 | 1.09435188 | 2.20E-06 | 2.43E-01 | 1.00E+00 | 1.00E+00 | URB2 | URB2 |
| rs12045643 | 1 | 229834050 | C | T | 421 | 0.04275534 | 36 | 5.48149152 | 1.10708948 | 7.37E-07 | 2.43E-01 | 1.00E+00 | 1.00E+00 | URB2 | URB2 |
| rs2375536 | 8 | 55640722 | T | C | 421 | 0.347981 | 292 | 2.12635806 | 0.44857091 | 2.13E-06 | 2.58E-01 | 1.00E+00 | 1.00E+00 | RP1 | SOX17 |
| rs4737674 | 8 | 55661654 | C | A | 421 | 0.33016627 | 277 | 2.30559554 | 0.44921988 | 2.86E-07 | 2.58E-01 | 1.00E+00 | 1.00E+00 | RP1 | SOX17 |
| rs13277510 | 8 | 55674149 | G | A | 421 | 0.33016627 | 277 | 2.30494752 | 0.44909942 | 2.86E-07 | 2.58E-01 | 1.00E+00 | 1.00E+00 | RP1 | SOX17 |
| rs12548593 | 8 | 55674617 | G | T | 421 | 0.33254157 | 279 | 2.28269961 | 0.44752402 | 3.38E-07 | 2.58E-01 | 1.00E+00 | 1.00E+00 | RP1 | SOX17 |
| rs1812506 | 8 | 55676101 | A | G | 421 | 0.3456057 | 291 | 2.12258868 | 0.4466178 | 2.01E-06 | 2.58E-01 | 1.00E+00 | 1.00E+00 | RP1 | SOX17 |
| rs16920698 | 8 | 55678434 | G | A | 421 | 0.33016627 | 277 | 2.30492695 | 0.44908177 | 2.86E-07 | 2.58E-01 | 1.00E+00 | 1.00E+00 | RP1 | SOX17 |
| rs1561297 | 8 | 55678538 | A | C | 421 | 0.33254157 | 279 | 2.28315237 | 0.44743146 | 3.35E-07 | 2.58E-01 | 1.00E+00 | 1.00E+00 | RP1 | SOX17 |
| rs4737676 | 8 | 55679546 | G | A | 421 | 0.33016627 | 277 | 2.30492057 | 0.44908192 | 2.86E-07 | 2.58E-01 | 1.00E+00 | 1.00E+00 | RP1 | SOX17 |
| rs2083123 | 8 | 55680318 | C | T | 421 | 0.33254157 | 279 | 2.27910794 | 0.44707491 | 3.44E-07 | 2.58E-01 | 1.00E+00 | 1.00E+00 | RP1 | SOX17 |
| rs983248 | 8 | 55680792 | C | T | 421 | 0.33016627 | 277 | 2.30260428 | 0.44871461 | 2.87E-07 | 2.58E-01 | 1.00E+00 | 1.00E+00 | RP1 | SOX17 |

| | | | | | | | | | | | | | | | |
|---|---|---|---|---|---|---|---|---|---|---|---|---|---|---|---|
| rs1391463 | 8 | 55681876 | T | G | 421 | 0.33016627 | 277 | 2.30258865 | 0.44871307 | 2.87E-07 | 2.58E-01 | 1.00E+00 | 1.00E+00 | RP1 | SOX17 |
| rs4737201 | 8 | 55691458 | C | T | 421 | 0.33016627 | 277 | 2.30197734 | 0.44869179 | 2.89E-07 | 2.58E-01 | 1.00E+00 | 1.00E+00 | RP1 | SOX17 |
| rs7843693 | 8 | 55692112 | G | A | 421 | 0.39786223 | 334 | 1.99856756 | 0.43695394 | 4.79E-06 | 2.58E-01 | 1.00E+00 | 1.00E+00 | RP1 | SOX17 |
| rs1396896 | 8 | 55695310 | A | G | 421 | 0.39786223 | 334 | 1.99853327 | 0.43690452 | 4.78E-06 | 2.58E-01 | 1.00E+00 | 1.00E+00 | RP1 | SOX17 |
| rs1391462 | 8 | 55699781 | C | A | 421 | 0.39786223 | 334 | 1.9984766 | 0.43696027 | 4.79E-06 | 2.58E-01 | 1.00E+00 | 1.00E+00 | RP1 | SOX17 |
| rs12678939 | 8 | 55705021 | A | G | 421 | 0.39429929 | 331 | 2.01975618 | 0.44220051 | 4.94E-06 | 2.58E-01 | 1.00E+00 | 1.00E+00 | RP1 | SOX17 |
| rs12266995 | 10 | 24852783 | T | C | 421 | 0.03087886 | 26 | 5.96782535 | 1.29311959 | 3.93E-06 | 2.85E-01 | 1.00E+00 | 1.00E+00 | ARHGAP21 | ARHGAP21 |
| rs149421869 | 2 | 52563371 | C | T | 421 | 0.00356295 | 3 | 19.7446169 | 3.72634921 | 1.17E-07 | 2.96E-01 | 1.00E+00 | 1.00E+00 | ASB3 | ASB3 |
| rs187520610 | 2 | 53360041 | G | A | 421 | 0.00475059 | 4 | 18.8010396 | 3.19364595 | 3.93E-09 | 2.96E-01 | 1.00E+00 | 1.00E+00 | ASB3 | ASB3 |
| rs116651654 | 4 | 163238743 | C | T | 421 | 0.00712589 | 6 | 15.6223694 | 3.06624451 | 3.49E-07 | 2.96E-01 | 1.00E+00 | 1.00E+00 | FSTL5 | FSTL5 |
| rs290120 | 5 | 163268244 | T | G | 421 | 0.00712589 | 6 | 13.3587764 | 2.83260801 | 2.40E-06 | 2.96E-01 | 1.00E+00 | 1.00E+00 | MAT2B | MAT2B |
| rs73586304 | 6 | 142839425 | C | T | 421 | 0.00356295 | 3 | 18.2405116 | 3.99428202 | 4.96E-06 | 2.96E-01 | 1.00E+00 | 1.00E+00 | VTA1 | VTA1 |
| rs181259864 | 7 | 97488823 | C | A | 421 | 0.00356295 | 3 | 17.1360821 | 3.68242789 | 3.26E-06 | 2.96E-01 | 1.00E+00 | 1.00E+00 | CZ1P-ASNS | ASNS |
| rs192750513 | 7 | 97577830 | A | G | 421 | 0.00356295 | 3 | 17.1892469 | 3.72514218 | 3.94E-06 | 2.96E-01 | 1.00E+00 | 1.00E+00 | CZ1P-ASNS | ASNS |
| rs151115079 | 11 | 18655741 | T | C | 421 | 0.00475059 | 5 | 17.8273587 | 3.2680205 | 4.89E-08 | 2.96E-01 | 1.00E+00 | 1.00E+00 | SPTY2D1 | SPTY2D1 |
| rs138414342 | 11 | 18679398 | G | A | 421 | 0.00475059 | 5 | 18.1582505 | 3.28214943 | 3.16E-08 | 2.96E-01 | 1.00E+00 | 1.00E+00 | SPTY2D1 | SPTY2D1 |
| rs151323346 | 12 | 21012024 | T | C | 421 | 0.00593824 | 4 | 15.8903888 | 3.38034868 | 2.59E-06 | 2.96E-01 | 1.00E+00 | 1.00E+00 | PDE3A | PDE3A |
| rs75186966 | 13 | 47395758 | A | C | 421 | 0.00356295 | 3 | 20.2281345 | 3.83225523 | 1.30E-07 | 2.96E-01 | 1.00E+00 | 1.00E+00 | HTR2A | HTR2A |
| rs146442492 | 15 | 58982115 | C | T | 421 | 0.02731591 | 24 | 7.05925858 | 1.51661316 | 3.25E-06 | 2.96E-01 | 1.00E+00 | 1.00E+00 | ADAM10 | ADAM10 |
| rs2327968 | 20 | 15813491 | C | T | 421 | 0.02494062 | 21 | 6.15899856 | 1.34390809 | 4.59E-06 | 2.96E-01 | 1.00E+00 | 1.00E+00 | MACROD2 | MACROD2 |
| rs2876414 | 20 | 15813704 | G | T | 421 | 0.02256532 | 19 | 6.81910907 | 1.48530907 | 4.41E-06 | 2.96E-01 | 1.00E+00 | 1.00E+00 | MACROD2 | MACROD2 |
| rs140788628 | 20 | 15858501 | C | A | 421 | 0.01068884 | 8 | 11.8039284 | 2.2826396 | 2.33E-07 | 2.96E-01 | 1.00E+00 | 1.00E+00 | MACROD2 | MACROD2 |
| rs2365739 | 1 | 62484462 | G | A | 421 | 0.02137767 | 18 | 6.86815917 | 1.43887996 | 1.81E-06 | 3.34E-01 | 1.00E+00 | 1.00E+00 | PATJ | PATJ |
| rs1856085 | 1 | 104114545 | G | A | 421 | 0.0023753 | 3 | 21.51144 | 4.26747898 | 4.64E-07 | 3.34E-01 | 1.00E+00 | 1.00E+00 | AMY2B | AMY2B |
| rs143597860 | 1 | 104157143 | A | G | 421 | 0.0023753 | 3 | 21.5748154 | 4.2760379 | 4.52E-07 | 3.34E-01 | 1.00E+00 | 1.00E+00 | AMY2B | AMY2B |
| rs144541665 | 1 | 104310729 | G | A | 421 | 0.0023753 | 3 | 21.8734611 | 4.30339839 | 3.72E-07 | 3.34E-01 | 1.00E+00 | 1.00E+00 | AMY2B | AMY2B |
| rs113651406 | 4 | 990967 | C | T | 421 | 0.00356295 | 3 | 19.4054026 | 3.7635826 | 2.52E-07 | 3.34E-01 | 1.00E+00 | 1.00E+00 | IDUA | IDUA |
| rs140797780 | 8 | 22087792 | C | T | 421 | 0.00356295 | 3 | 19.2908396 | 3.69147785 | 1.73E-07 | 3.34E-01 | 1.00E+00 | 1.00E+00 | PHYHIP | PHYHIP |
| rs76904423 | 12 | 101188744 | G | A | 421 | 0.01068884 | 10 | 9.80576231 | 2.11568908 | 3.57E-06 | 3.34E-01 | 1.00E+00 | 1.00E+00 | ANO4 | ANO4 |
| rs150586237 | 6 | 24491348 | C | T | 421 | 0.00356295 | 3 | 22.0461443 | 3.78094793 | 5.51E-09 | 3.46E-01 | 1.00E+00 | 1.00E+00 | GPLD1 | GPLD1 |
| rs423841 | 8 | 55556069 | G | A | 421 | 0.66270784 | 291 | -2.1495726 | 0.45846846 | 2.75E-06 | 3.49E-01 | 1.00E+00 | 1.00E+00 | RP1 | SOX17 |
| rs433324 | 8 | 55564609 | A | G | 421 | 0.66627078 | 287 | -2.2483462 | 0.45814252 | 9.22E-07 | 3.49E-01 | 1.00E+00 | 1.00E+00 | RP1 | SOX17 |
| rs369623 | 8 | 55571940 | A | C | 421 | 0.66627078 | 287 | -2.2359815 | 0.45415557 | 8.51E-07 | 3.49E-01 | 1.00E+00 | 1.00E+00 | RP1 | SOX17 |
| rs446222 | 8 | 55574960 | G | A | 421 | 0.66627078 | 288 | -2.247819 | 0.45397995 | 7.37E-07 | 3.49E-01 | 1.00E+00 | 1.00E+00 | RP1 | SOX17 |
| rs432393 | 8 | 55580298 | C | T | 421 | 0.66270784 | 291 | -2.1875382 | 0.45096346 | 1.23E-06 | 3.49E-01 | 1.00E+00 | 1.00E+00 | RP1 | SOX17 |
| rs3098298 | 8 | 55582838 | C | T | 421 | 0.66270784 | 291 | -2.1875635 | 0.45094791 | 1.23E-06 | 3.49E-01 | 1.00E+00 | 1.00E+00 | RP1 | SOX17 |
| rs367179 | 8 | 55587616 | T | C | 421 | 0.66270784 | 291 | -2.1875635 | 0.45094791 | 1.23E-06 | 3.49E-01 | 1.00E+00 | 1.00E+00 | RP1 | SOX17 |
| rs382476 | 8 | 55590975 | G | A | 421 | 0.66627078 | 288 | -2.2481938 | 0.4540554 | 7.37E-07 | 3.49E-01 | 1.00E+00 | 1.00E+00 | RP1 | SOX17 |
| rs384543 | 8 | 55591609 | G | A | 421 | 0.66627078 | 288 | -2.2481938 | 0.4540554 | 7.37E-07 | 3.49E-01 | 1.00E+00 | 1.00E+00 | RP1 | SOX17 |
| rs405226 | 8 | 55592336 | A | G | 421 | 0.66270784 | 291 | -2.1881305 | 0.45101874 | 1.23E-06 | 3.49E-01 | 1.00E+00 | 1.00E+00 | RP1 | SOX17 |
| rs384127 | 8 | 55597489 | G | A | 421 | 0.66627078 | 288 | -2.2481559 | 0.45404811 | 7.37E-07 | 3.49E-01 | 1.00E+00 | 1.00E+00 | RP1 | SOX17 |
| rs2375537 | 8 | 55619508 | C | T | 421 | 0.33254157 | 280 | 2.23876848 | 0.44938849 | 6.30E-07 | 3.49E-01 | 1.00E+00 | 1.00E+00 | RP1 | SOX17 |
| rs720372 | 8 | 55628637 | G | A | 421 | 0.34679335 | 292 | 2.09872734 | 0.44886259 | 2.93E-06 | 3.49E-01 | 1.00E+00 | 1.00E+00 | RP1 | SOX17 |
| rs1437781 | 8 | 55629852 | T | C | 421 | 0.33254157 | 280 | 2.23854673 | 0.44940766 | 6.32E-07 | 3.49E-01 | 1.00E+00 | 1.00E+00 | RP1 | SOX17 |
| rs1595406 | 8 | 55630615 | A | G | 421 | 0.3456057 | 291 | 2.10018889 | 0.44830064 | 2.80E-06 | 3.49E-01 | 1.00E+00 | 1.00E+00 | RP1 | SOX17 |
| rs1437782 | 8 | 55632762 | C | T | 421 | 0.33016627 | 278 | 2.2579585 | 0.45098781 | 5.54E-07 | 3.49E-01 | 1.00E+00 | 1.00E+00 | RP1 | SOX17 |
| rs10958428 | 8 | 55685641 | A | G | 421 | 0.33372922 | 280 | 2.23996885 | 0.44568673 | 5.01E-07 | 3.49E-01 | 1.00E+00 | 1.00E+00 | RP1 | SOX17 |
| rs75689761 | 7 | 18406573 | C | T | 421 | 0.00831354 | 7 | 11.8482917 | 2.41320258 | 9.12E-07 | 4.27E-01 | 1.00E+00 | 1.00E+00 | HDAC9 | HDAC9 |
| rs77346868 | 7 | 18406599 | A | G | 421 | 0.00950119 | 8 | 11.0210129 | 2.26481172 | 1.14E-06 | 4.27E-01 | 1.00E+00 | 1.00E+00 | HDAC9 | HDAC9 |
| rs78225611 | 7 | 18407464 | A | C | 421 | 0.00950119 | 8 | 11.0294387 | 2.26622466 | 1.13E-06 | 4.27E-01 | 1.00E+00 | 1.00E+00 | HDAC9 | HDAC9 |
| rs77300464 | 7 | 18408761 | A | G | 421 | 0.00712589 | 6 | 14.6659794 | 2.6169516 | 2.09E-08 | 4.27E-01 | 1.00E+00 | 1.00E+00 | HDAC9 | HDAC9 |
| rs79602997 | 7 | 18410250 | G | A | 421 | 0.00831354 | 7 | 11.8608211 | 2.41901626 | 9.43E-07 | 4.27E-01 | 1.00E+00 | 1.00E+00 | HDAC9 | HDAC9 |

| | | | | | | | | | | | | | | | |
|---|---|---|---|---|---|---|---|---|---|---|---|---|---|---|---|
| rs75773869 | 7 | 18410845 | G | T | 421 | 0.00831354 | 7 | 11.8642716 | 2.41902322 | 9.36E-07 | 4.27E-01 | 1.00E+00 | 1.00E+00 | HDAC9 | HDAC9 |
| rs75606013 | 7 | 18414613 | G | A | 421 | 0.00712589 | 6 | 14.6617119 | 2.61692421 | 2.11E-08 | 4.27E-01 | 1.00E+00 | 1.00E+00 | HDAC9 | HDAC9 |
| rs61434999 | 7 | 18418351 | A | G | 421 | 0.00831354 | 7 | 11.8411511 | 2.41284027 | 9.22E-07 | 4.27E-01 | 1.00E+00 | 1.00E+00 | HDAC9 | HDAC9 |
| rs78907958 | 7 | 18425017 | T | G | 421 | 0.00831354 | 7 | 12.0901322 | 2.40514961 | 4.99E-07 | 4.27E-01 | 1.00E+00 | 1.00E+00 | HDAC9 | HDAC9 |
| rs76526501 | 7 | 18431110 | G | A | 421 | 0.00712589 | 6 | 14.8397312 | 2.5778114 | 8.58E-09 | 4.27E-01 | 1.00E+00 | 1.00E+00 | HDAC9 | HDAC9 |
| rs74455595 | 7 | 18431784 | A | G | 421 | 0.00712589 | 6 | 14.8397312 | 2.5778114 | 8.58E-09 | 4.27E-01 | 1.00E+00 | 1.00E+00 | HDAC9 | HDAC9 |
| rs79182806 | 7 | 18433827 | T | C | 421 | 0.00831354 | 7 | 12.0972603 | 2.38956889 | 4.14E-07 | 4.27E-01 | 1.00E+00 | 1.00E+00 | HDAC9 | HDAC9 |
| rs10279777 | 7 | 18441589 | G | A | 421 | 0.00950119 | 8 | 12.8681335 | 2.27961694 | 1.65E-08 | 4.27E-01 | 1.00E+00 | 1.00E+00 | HDAC9 | HDAC9 |
| rs77867199 | 7 | 18442275 | G | T | 421 | 0.01068884 | 9 | 10.8221282 | 2.13363365 | 3.93E-07 | 4.27E-01 | 1.00E+00 | 1.00E+00 | HDAC9 | HDAC9 |
| rs80156375 | 7 | 18443215 | A | C | 421 | 0.00950119 | 8 | 11.5406255 | 2.25971308 | 3.27E-07 | 4.27E-01 | 1.00E+00 | 1.00E+00 | HDAC9 | HDAC9 |
| rs17169602 | 7 | 18446741 | G | A | 421 | 0.00950119 | 8 | 12.7273717 | 2.25993984 | 1.78E-08 | 4.27E-01 | 1.00E+00 | 1.00E+00 | HDAC9 | HDAC9 |
| rs10486295 | 7 | 18446807 | G | A | 421 | 0.00950119 | 8 | 12.76229 | 2.26480456 | 1.75E-08 | 4.27E-01 | 1.00E+00 | 1.00E+00 | HDAC9 | HDAC9 |
| rs75090694 | 7 | 18447436 | A | G | 421 | 0.00831354 | 7 | 13.8314005 | 2.41427081 | 1.01E-08 | 4.27E-01 | 1.00E+00 | 1.00E+00 | HDAC9 | HDAC9 |
| rs12315614 | 12 | 64920957 | C | A | 421 | 0.0760095 | 64 | 3.77710662 | 0.8201959 | 4.12E-06 | 5.64E-01 | 1.00E+00 | 1.00E+00 | TBK1 | TBK1 |
| rs147630370 | 4 | 87450675 | T | C | 421 | 0.00475059 | 4 | 16.0013633 | 3.23339786 | 7.47E-07 | 6.04E-01 | 1.00E+00 | 1.00E+00 | MAPK10 | MAPK10 |
| rs142993106 | 4 | 90957372 | G | A | 421 | 0.01781473 | 15 | 8.08463611 | 1.73306162 | 3.09E-06 | 6.04E-01 | 1.00E+00 | 1.00E+00 | CCSER1 | CCSER1 |
| rs146526206 | 4 | 90993018 | T | C | 421 | 0.01781473 | 15 | 8.09176656 | 1.69535286 | 1.82E-06 | 6.04E-01 | 1.00E+00 | 1.00E+00 | CCSER1 | CCSER1 |
| rs139493286 | 18 | 28816019 | G | A | 421 | 0.00356295 | 3 | 17.4603093 | 3.75076428 | 3.24E-06 | 7.11E-01 | 1.00E+00 | 1.00E+00 | DSG1 | DSG1 |
| rs1686289 | 14 | 46260982 | G | A | 421 | 0.67814727 | 277 | -2.2551539 | 0.48625976 | 3.52E-06 | 8.30E-01 | 1.00E+00 | 1.00E+00 | LINC02303 | MIS18BP1 |
| rs176786 | 14 | 46282970 | T | C | 421 | 0.32304038 | 268 | 2.28475641 | 0.49171936 | 3.38E-06 | 8.30E-01 | 1.00E+00 | 1.00E+00 | LINC02303 | MIS18BP1 |
| rs79539453 | 11 | 125266353 | C | T | 421 | 0.0023753 | 3 | 20.8220616 | 4.34542406 | 1.65E-06 | 9.23E-01 | 1.00E+00 | 1.00E+00 | PKNOX2 | PKNOX2 |
| rs78547898 | 22 | 32824278 | G | A | 421 | 0.00356295 | 3 | 20.9098395 | 4.15538274 | 4.85E-07 | 9.27E-01 | 1.00E+00 | 1.00E+00 | BPIFC | FOX07 |
| rs7104959 | 11 | 129846126 | C | T | 421 | 0.00356295 | 3 | 17.9204611 | 3.84366851 | 3.13E-06 | 9.37E-01 | 1.00E+00 | 1.00E+00 | PRDM10 | PRDM10 |
| rs10494861 | 1 | 205331874 | G | A | 421 | 0.00356295 | 3 | 19.1820498 | 4.0729134 | 2.48E-06 | 9.56E-01 | 1.00E+00 | 1.00E+00 | KLHDC8A | KLHDC8A |
| rs1877768 | 6 | 16534923 | C | T | 421 | 0.01781473 | 15 | 8.28754685 | 1.76830748 | 2.78E-06 | 9.56E-01 | 1.00E+00 | 1.00E+00 | ATXN1 | ATXN1 |
| rs111391231 | 7 | 89497045 | T | C | 421 | 0.01425178 | 12 | 8.31717755 | 1.80720714 | 4.18E-06 | 9.56E-01 | 1.00E+00 | 1.00E+00 | STEAP2 | STEAP2 |
| rs111900874 | 7 | 89516669 | G | A | 421 | 0.01425178 | 12 | 8.42424013 | 1.82258334 | 3.80E-06 | 9.56E-01 | 1.00E+00 | 1.00E+00 | STEAP2 | STEAP2 |
| rs115348382 | 1 | 9655903 | G | A | 421 | 0.00356295 | 3 | 18.2107495 | 3.84721698 | 2.21E-06 | NA | | | | |
| rs186532456 | 1 | 18948328 | C | T | 421 | 0.00356295 | 3 | 20.1730343 | 4.11737745 | 9.61E-07 | NA | | | | |
| rs562032622 | 1 | 18959333 | A | C | 421 | 0.00356295 | 3 | 21.0267115 | 4.3197503 | 1.13E-06 | NA | | | | |
| rs149493615 | 1 | 79880089 | G | A | 421 | 0.00831354 | 8 | 11.750632 | 2.43451244 | 1.39E-06 | NA | | | | |
| rs143811231 | 1 | 79968131 | T | C | 421 | 0.00831354 | 8 | 11.5747409 | 2.42066027 | 1.74E-06 | NA | | | | |
| rs34270375 | 1 | 89370702 | G | A | 421 | 0.02612827 | 21 | 7.71147017 | 1.57216792 | 9.34E-07 | NA | | | | |
| rs187518659 | 1 | 99455745 | T | G | 421 | 0.00950119 | 8 | 11.6819623 | 2.46564537 | 2.16E-06 | NA | | | | |
| rs140420703 | 1 | 102803484 | T | G | 421 | 0.00475059 | 5 | 16.487654 | 3.46213891 | 1.91E-06 | NA | | | | |
| rs563167766 | 1 | 102866365 | G | A | 421 | 0.0023753 | 3 | 19.415739 | 4.24240395 | 4.73E-06 | NA | | | | |
| rs77180278 | 1 | 102961882 | T | C | 421 | 0.02850356 | 24 | 7.33619996 | 1.37404112 | 9.34E-08 | NA | | | | |
| rs112351653 | 1 | 103220360 | T | C | 421 | 0.02850356 | 24 | 7.42386947 | 1.33286772 | 2.55E-08 | NA | | | | |
| rs180926150 | 1 | 103226326 | C | T | 421 | 0.0023753 | 3 | 19.641918 | 4.24793649 | 3.77E-06 | NA | | | | |
| rs114413507 | 1 | 103419168 | T | C | 421 | 0.02850356 | 24 | 7.42216029 | 1.33294954 | 2.57E-08 | NA | | | | |
| rs116672066 | 1 | 103472916 | G | A | 421 | 0.02731591 | 23 | 8.04769476 | 1.37187844 | 4.46E-09 | NA | | | | |
| rs111928960 | 1 | 103633635 | G | A | 421 | 0.02850356 | 22 | 8.60257208 | 1.45827516 | 3.65E-09 | NA | | | | |
| rs113221952 | 1 | 103753974 | A | G | 421 | 0.02137767 | 19 | 7.92350704 | 1.61421682 | 9.17E-07 | NA | | | | |
| rs76617932 | 1 | 180930424 | T | C | 421 | 0.01187648 | 9 | 10.5449608 | 2.25040726 | 2.79E-06 | NA | | | | |
| rs183180157 | 1 | 181247121 | A | C | 421 | 0.00950119 | 7 | 12.61386 | 2.68921121 | 2.72E-06 | NA | | | | |
| rs375790303 | 1 | 184530482 | G | A | 421 | 0.00831354 | 7 | 11.4240393 | 2.49719192 | 4.77E-06 | NA | | | | |
| rs138480898 | 1 | 184955657 | C | T | 421 | 0.00356295 | 3 | 16.9873974 | 3.66972373 | 3.67E-06 | NA | | | | |
| rs145766563 | 1 | 185129502 | G | A | 421 | 0.00356295 | 4 | 16.8682102 | 3.54127008 | 1.90E-06 | NA | | | | |
| rs147032554 | 1 | 186148864 | T | G | 421 | 0.00356295 | 3 | 18.162609 | 3.65402821 | 6.68E-07 | NA | | | | |
| rs180989936 | 1 | 193044178 | A | G | 421 | 0.00475059 | 4 | 17.1793579 | 3.70215494 | 3.48E-06 | NA | | | | |
| rs559559983 | 1 | 246977132 | C | A | 421 | 0.00712589 | 5 | 13.811132 | 2.89725588 | 1.87E-06 | NA | | | | |
| rs550763536 | 2 | 7452347 | T | G | 421 | 0.00593824 | 5 | 14.4871647 | 3.10349768 | 3.04E-06 | NA | | | | |

| | | | | | | | | | | | |
|---|---|---|---|---|---|---|---|---|---|---|---|
| rs558553658 | 2 | 15682724 | C | T | 421 | 0.00475059 | 3 | 17.1099107 | 3.29108061 | 2.01E-07 | NA |
| rs536781978 | 2 | 29733681 | A | G | 421 | 0.00356295 | 3 | 17.5870824 | 3.70259451 | 2.03E-06 | NA |
| rs568321148 | 2 | 29870348 | T | G | 421 | 0.00356295 | 3 | 17.7125046 | 3.7201251 | 1.92E-06 | NA |
| rs76777840 | 2 | 48312950 | G | A | 421 | 0.00593824 | 5 | 14.3500784 | 2.8927894 | 7.03E-07 | NA |
| rs145080832 | 2 | 48473143 | G | A | 421 | 0.00475059 | 4 | 17.1618477 | 3.25709537 | 1.37E-07 | NA |
| rs184220112 | 2 | 48631743 | C | A | 421 | 0.00475059 | 4 | 16.4879043 | 3.22267606 | 3.12E-07 | NA |
| rs189890455 | 2 | 48655397 | C | T | 421 | 0.00593824 | 5 | 15.1467391 | 2.99355083 | 4.20E-07 | NA |
| rs181193202 | 2 | 52527354 | T | C | 421 | 0.00356295 | 3 | 19.6312088 | 3.74776015 | 1.62E-07 | NA |
| rs190193113 | 2 | 53085778 | G | A | 421 | 0.00356295 | 3 | 19.9533602 | 3.7619559 | 1.13E-07 | NA |
| rs146479102 | 2 | 65825759 | G | A | 421 | 0.00593824 | 5 | 15.4846768 | 3.26980199 | 2.18E-06 | NA |
| rs528288879 | 2 | 65875930 | C | T | 421 | 0.00593824 | 6 | 14.7878085 | 2.95121929 | 5.42E-07 | NA |
| rs11690187 | 2 | 67565909 | A | C | 421 | 0.00593824 | 5 | 13.6310532 | 2.9751322 | 4.61E-06 | NA |
| rs151272830 | 2 | 67682724 | G | T | 421 | 0.00712589 | 6 | 13.3323479 | 2.84806076 | 2.85E-06 | NA |
| rs186142189 | 2 | 67702707 | G | A | 421 | 0.00712589 | 6 | 12.7361927 | 2.71405368 | 2.70E-06 | NA |
| rs184200893 | 2 | 69260913 | C | T | 421 | 0.00356295 | 3 | 19.9580356 | 4.21905487 | 2.24E-06 | NA |
| rs111927235 | 2 | 74483954 | A | G | 421 | 0.00475059 | 4 | 17.0468955 | 3.46111889 | 8.43E-07 | NA |
| rs111838310 | 2 | 74673491 | C | A | 421 | 0.00475059 | 5 | 17.7963938 | 3.48429066 | 3.26E-07 | NA |
| rs112983626 | 2 | 74697150 | G | A | 421 | 0.00475059 | 4 | 17.7848079 | 3.48532494 | 3.35E-07 | NA |
| rs113006316 | 2 | 74802360 | A | G | 421 | 0.0023753 | 3 | 22.8886792 | 4.92411335 | 3.35E-06 | NA |
| rs17746486 | 2 | 95722609 | C | T | 421 | 0.03206651 | 29 | 6.90851358 | 1.43477175 | 1.47E-06 | NA |
| rs76554191 | 2 | 95967628 | G | A | 421 | 0.04038005 | 34 | 5.72148039 | 1.2389174 | 3.87E-06 | NA |
| rs140352232 | 2 | 108038112 | G | A | 421 | 0.0023753 | 3 | 21.0576511 | 4.56013129 | 3.88E-06 | NA |
| rs116189766 | 2 | 126393864 | T | C | 421 | 0.0023753 | 3 | 21.190764 | 4.36037289 | 1.17E-06 | NA |
| rs139877408 | 2 | 129606273 | A | G | 421 | 0.0023753 | 3 | 20.3067129 | 4.43052497 | 4.58E-06 | NA |
| rs142894171 | 2 | 151438271 | G | T | 421 | 0.00356295 | 3 | 18.0710394 | 3.74586836 | 1.41E-06 | NA |
| rs541508507 | 2 | 170023265 | G | A | 421 | 0.00475059 | 4 | 15.4630033 | 3.27389618 | 2.32E-06 | NA |
| rs142549310 | 2 | 170030506 | C | T | 421 | 0.00475059 | 4 | 15.2501713 | 3.24983634 | 2.70E-06 | NA |
| rs556293455 | 2 | 176950420 | G | A | 421 | 0.00356295 | 3 | 14.3860463 | 2.96998402 | 1.27E-06 | NA |
| rs184098071 | 2 | 177116420 | G | A | 421 | 0.00356295 | 3 | 13.947724 | 2.86782452 | 1.15E-06 | NA |
| rs532416695 | 2 | 177486790 | G | A | 421 | 0.00356295 | 3 | 19.8726656 | 3.76737683 | 1.33E-07 | NA |
| rs112557251 | 2 | 188375400 | T | C | 421 | 0.0023753 | 3 | 20.6510064 | 4.44850106 | 3.45E-06 | NA |
| rs185158855 | 2 | 223650026 | C | A | 421 | 0.00475059 | 4 | 16.6678864 | 3.60266524 | 3.72E-06 | NA |
| rs185510569 | 2 | 223814861 | G | A | 421 | 0.00356295 | 3 | 19.5886906 | 3.60490977 | 5.51E-08 | NA |
| rs147559909 | 2 | 237051523 | T | C | 421 | 0.00593824 | 5 | 17.0840975 | 3.03209195 | 1.76E-08 | NA |
| rs181217257 | 2 | 239989119 | C | T | 421 | 0.00475059 | 4 | 21.5677368 | 3.47296989 | 5.29E-10 | NA |
| rs188076929 | 2 | 239993719 | T | C | 421 | 0.00475059 | 4 | 19.9775614 | 3.27246746 | 1.03E-09 | NA |
| rs112475378 | 3 | 1626661 | T | C | 421 | 0.01662708 | 14 | 9.55085929 | 1.97628138 | 1.35E-06 | NA |
| rs145676540 | 3 | 2044635 | C | T | 421 | 0.00593824 | 5 | 13.980658 | 2.88447377 | 1.25E-06 | NA |
| rs146007933 | 3 | 28227423 | T | C | 421 | 0.02137767 | 18 | 7.51149453 | 1.55442805 | 1.35E-06 | NA |
| rs73057656 | 3 | 33940571 | A | G | 421 | 0.0368171 | 33 | 5.9238329 | 1.27589711 | 3.44E-06 | NA |
| rs73085348 | 3 | 42711221 | A | G | 421 | 0.01187648 | 11 | 10.139317 | 2.03504571 | 6.28E-07 | NA |
| rs142684595 | 3 | 55319301 | T | C | 421 | 0.00593824 | 5 | 16.7729431 | 3.11364755 | 7.17E-08 | NA |
| rs80203220 | 3 | 122722331 | C | T | 421 | 0.00593824 | 6 | 13.426042 | 2.84218656 | 2.31E-06 | NA |
| rs192443987 | 3 | 135532345 | G | A | 421 | 0.00475059 | 4 | 17.7540285 | 3.67564302 | 1.36E-06 | NA |
| rs148248743 | 3 | 136134595 | C | T | 421 | 0.0023753 | 3 | 22.839379 | 4.62044689 | 7.69E-07 | NA |
| rs576124203 | 3 | 141841196 | T | G | 421 | 0.00356295 | 3 | 19.4039925 | 3.84392582 | 4.47E-07 | NA |
| rs545552231 | 3 | 141864350 | C | T | 421 | 0.00356295 | 3 | 19.8641332 | 3.94582563 | 4.80E-07 | NA |
| rs193153124 | 3 | 148330710 | A | G | 421 | 0.00593824 | 5 | 14.3657313 | 3.06514594 | 2.78E-06 | NA |
| rs188720948 | 3 | 150069880 | T | C | 421 | 0.00356295 | 3 | 17.7032653 | 3.59722874 | 8.59E-07 | NA |
| rs16823323 | 3 | 153657202 | G | A | 421 | 0.01662708 | 14 | 9.48464065 | 1.71769795 | 3.36E-08 | NA |
| rs139943877 | 3 | 155451289 | G | A | 421 | 0.00712589 | 7 | 13.0535902 | 2.76041594 | 2.26E-06 | NA |
| rs187047882 | 3 | 164285621 | G | A | 421 | 0.00356295 | 3 | 18.5641361 | 3.99725417 | 3.41E-06 | NA |

| | | | | | | | | | | | |
|---|---|---|---|---|---|---|---|---|---|---|---|
| rs141169929 | 3 | 164808462 | A | G | 421 | 0.00475059 | 5 | 14.8454178 | 3.1738839 | 2.91E-06 | NA |
| rs186649043 | 3 | 174932293 | C | T | 421 | 0.00356295 | 3 | 19.2134209 | 4.06588786 | 2.30E-06 | NA |
| rs189709453 | 3 | 177574308 | G | A | 421 | 0.00356295 | 3 | 21.6581488 | 4.05916142 | 9.52E-08 | NA |
| rs186767531 | 3 | 177639777 | T | C | 421 | 0.00356295 | 3 | 21.2013243 | 4.04780758 | 1.63E-07 | NA |
| rs182868205 | 3 | 177712448 | C | T | 421 | 0.00356295 | 3 | 20.9548768 | 3.97841839 | 1.39E-07 | NA |
| rs191792521 | 3 | 195646605 | G | A | 421 | 0.00831354 | 6 | 13.9515484 | 2.96443599 | 2.52E-06 | NA |
| rs189765693 | 4 | 4324793 | T | C | 421 | 0.00475059 | 4 | 15.5191341 | 3.29924607 | 2.55E-06 | NA |
| rs183962155 | 4 | 21259643 | A | C | 421 | 0.00593824 | 6 | 13.7646008 | 2.97247759 | 3.64E-06 | NA |
| rs113751774 | 4 | 23428854 | C | T | 421 | 0.00712589 | 7 | 14.9103274 | 2.96657459 | 5.01E-07 | NA |
| rs113063005 | 4 | 23509067 | T | C | 421 | 0.00593824 | 6 | 21.2073894 | 3.15520096 | 1.80E-11 | NA |
| rs145875128 | 4 | 32073136 | G | A | 421 | 0.00356295 | 3 | 24.479986 | 4.60639213 | 1.07E-07 | NA |
| rs143287889 | 4 | 35572280 | C | T | 421 | 0.0023753 | 3 | 19.96741 | 4.34406 | 4.30E-06 | NA |
| rs77141817 | 4 | 37053759 | T | C | 421 | 0.00356295 | 3 | 20.6110292 | 3.64214866 | 1.52E-08 | NA |
| rs190822761 | 4 | 37099356 | G | T | 421 | 0.00356295 | 3 | 20.5846354 | 3.64346559 | 1.61E-08 | NA |
| rs999769259 | 4 | 62511965 | G | A | 421 | 0.00356295 | 3 | 22.1246725 | 4.77597881 | 3.61E-06 | NA |
| rs147171192 | 4 | 89135588 | A | G | 421 | 0.00712589 | 5 | 13.3044047 | 2.89758492 | 4.40E-06 | NA |
| rs145116559 | 4 | 112677596 | T | C | 421 | 0.00356295 | 4 | 18.2441224 | 3.9932536 | 4.91E-06 | NA |
| rs181415102 | 4 | 112689266 | T | C | 421 | 0.00356295 | 4 | 18.2631019 | 3.99387944 | 4.81E-06 | NA |
| rs191423619 | 4 | 126516627 | G | T | 421 | 0.0023753 | 3 | 21.694611 | 4.20667337 | 2.51E-07 | NA |
| rs149298750 | 4 | 127158701 | A | C | 421 | 0.00356295 | 3 | 22.184052 | 4.26573246 | 1.99E-07 | NA |
| rs112679237 | 4 | 139140860 | T | C | 421 | 0.01781473 | 18 | 8.71202752 | 1.67680671 | 2.04E-07 | NA |
| rs531769270 | 4 | 154411043 | T | C | 421 | 0.00356295 | 3 | 16.5945338 | 3.51936092 | 2.41E-06 | NA |
| rs567982164 | 5 | 25631297 | G | A | 421 | 0.0023753 | 3 | 21.8356286 | 4.36706703 | 5.73E-07 | NA |
| rs191986449 | 5 | 25745687 | C | T | 421 | 0.00475059 | 3 | 18.4978793 | 4.03679831 | 4.60E-06 | NA |
| rs185771987 | 5 | 73285489 | T | C | 421 | 0.00356295 | 3 | 18.2344824 | 3.97675014 | 4.53E-06 | NA |
| rs139360368 | 5 | 73372109 | A | C | 421 | 0.00356295 | 4 | 18.3590487 | 3.78701272 | 1.25E-06 | NA |
| rs181933850 | 5 | 91465647 | A | G | 421 | 0.00831354 | 7 | 11.0436586 | 2.37544216 | 3.33E-06 | NA |
| rs190190051 | 5 | 91475485 | G | A | 421 | 0.00831354 | 7 | 11.0589887 | 2.38317975 | 3.48E-06 | NA |
| rs182531466 | 5 | 91530073 | C | A | 421 | 0.00712589 | 6 | 12.6804978 | 2.75685261 | 4.23E-06 | NA |
| rs187236873 | 5 | 91530447 | G | A | 421 | 0.00712589 | 6 | 12.44907 | 2.61780937 | 1.98E-06 | NA |
| rs183816745 | 5 | 91631888 | A | G | 421 | 0.00593824 | 5 | 17.3487909 | 3.20195556 | 6.02E-08 | NA |
| rs111407636 | 5 | 95080029 | C | T | 421 | 0.00356295 | 3 | 18.642774 | 3.74168052 | 6.28E-07 | NA |
| rs111676272 | 5 | 95094298 | C | A | 421 | 0.00356295 | 3 | 18.6068625 | 3.74429748 | 6.72E-07 | NA |
| rs75848314 | 5 | 95098340 | T | C | 421 | 0.00356295 | 3 | 20.0998481 | 3.78700451 | 1.11E-07 | NA |
| rs111846247 | 5 | 95111643 | T | C | 421 | 0.00356295 | 3 | 20.4865772 | 3.85467205 | 1.07E-07 | NA |
| rs137873790 | 5 | 97087041 | A | G | 421 | 0.01306413 | 11 | 9.22240881 | 2.01095379 | 4.52E-06 | NA |
| rs189912648 | 5 | 134765439 | C | T | 421 | 0.00356295 | 4 | 16.9732599 | 3.60846227 | 2.55E-06 | NA |
| rs191006910 | 5 | 154216009 | G | A | 421 | 0.00356295 | 3 | 17.6173803 | 3.79049689 | 3.36E-06 | NA |
| rs74343174 | 5 | 161493182 | C | A | 421 | 0.00593824 | 5 | 14.7671824 | 3.13913408 | 2.55E-06 | NA |
| rs775626702 | 5 | 162630067 | A | C | 421 | 0.00356295 | 3 | 18.9484137 | 4.07023372 | 3.23E-06 | NA |
| rs371245624 | 5 | 162880956 | T | C | 421 | 0.00356295 | 3 | 19.4795931 | 4.06572011 | 1.66E-06 | NA |
| rs545428520 | 5 | 167821266 | T | C | 421 | 0.00356295 | 4 | 21.566035 | 3.66407988 | 3.96E-09 | NA |
| rs528404963 | 5 | 167852025 | T | C | 421 | 0.00356295 | 4 | 21.1393977 | 3.65303517 | 7.17E-09 | NA |
| rs72832764 | 5 | 170004673 | G | A | 421 | 0.00356295 | 3 | 18.2159757 | 3.88196743 | 2.70E-06 | NA |
| rs72837643 | 5 | 170188955 | T | C | 421 | 0.00356295 | 4 | 16.9302874 | 3.67743479 | 4.15E-06 | NA |
| rs142311947 | 5 | 177384469 | G | A | 421 | 0.00712589 | 8 | 12.0339013 | 2.57618116 | 2.99E-06 | NA |
| rs151015676 | 5 | 177390937 | T | G | 421 | 0.0023753 | 3 | 20.1421367 | 4.37074482 | 4.06E-06 | NA |
| rs571986619 | 6 | 85678832 | A | G | 421 | 0.0023753 | 3 | 20.7858067 | 4.34301983 | 1.70E-06 | NA |
| rs56224400 | 6 | 98092675 | T | C | 421 | 0.01662708 | 14 | 8.4433064 | 1.72251471 | 9.50E-07 | NA |
| rs147627638 | 6 | 99173116 | A | G | 421 | 0.00712589 | 6 | 13.0785306 | 2.79364254 | 2.85E-06 | NA |
| rs141326851 | 6 | 134833127 | A | C | 421 | 0.01662708 | 14 | 8.57849688 | 1.81053268 | 2.16E-06 | NA |
| rs146048121 | 6 | 142198955 | G | A | 421 | 0.0023753 | 3 | 20.0214379 | 4.30086025 | 3.24E-06 | NA |

| | | | | | | | | | | | |
|---|---|---|---|---|---|---|---|---|---|---|---|
| rs142106992 | 6 | 142269885 | C | A | 421 | 0.00356295 | 4 | 22.8345886 | 3.6758699 | 5.23E-10 | NA |
| rs72983831 | 6 | 142307100 | T | G | 421 | 0.00593824 | 6 | 15.6733239 | 3.17294911 | 7.83E-07 | NA |
| rs72986533 | 6 | 142611258 | T | C | 421 | 0.00831354 | 8 | 13.2812726 | 2.53063629 | 1.54E-07 | NA |
| rs148532212 | 6 | 165499857 | T | C | 421 | 0.00356295 | 3 | 19.3535457 | 3.70945693 | 1.81E-07 | NA |
| rs117498042 | 6 | 165514281 | C | T | 421 | 0.00356295 | 3 | 19.335471 | 3.70913521 | 1.86E-07 | NA |
| rs148153037 | 6 | 167501386 | G | A | 421 | 0.00831354 | 8 | 14.3359246 | 2.42128102 | 3.20E-09 | NA |
| rs184487573 | 6 | 167513471 | A | G | 421 | 0.00712589 | 7 | 13.5009635 | 2.59268014 | 1.92E-07 | NA |
| rs187978759 | 7 | 11711845 | G | A | 421 | 0.00356295 | 3 | 20.4555522 | 4.3637719 | 2.76E-06 | NA |
| rs117166500 | 7 | 17052778 | G | T | 421 | 0.00712589 | 7 | 12.5146865 | 2.62694198 | 1.90E-06 | NA |
| rs55844051 | 7 | 23360363 | T | C | 421 | 0.00356295 | 3 | 18.3875896 | 3.86268704 | 1.93E-06 | NA |
| rs1301444047 | 7 | 36574504 | G | A | 421 | 0.04275534 | 43 | 5.12842209 | 1.08835549 | 2.45E-06 | NA |
| rs62447184 | 7 | 36574504 | G | A | 421 | 0.04275534 | 43 | 5.12842209 | 1.08835549 | 2.45E-06 | NA |
| rs574076561 | 7 | 49544747 | A | G | 421 | 0.00356295 | 3 | 19.6434766 | 4.25225723 | 3.85E-06 | NA |
| rs539713344 | 7 | 100474786 | G | A | 421 | 0.0023753 | 3 | 20.044756 | 4.21957545 | 2.03E-06 | NA |
| rs188028357 | 7 | 100668425 | C | T | 421 | 0.0023753 | 3 | 21.8425578 | 4.35791809 | 5.38E-07 | NA |
| rs536023430 | 7 | 146865072 | T | C | 421 | 0.00356295 | 3 | 19.4038683 | 4.13084856 | 2.64E-06 | NA |
| rs187384541 | 8 | 1632241 | A | G | 421 | 0.03562945 | 26 | 6.88745117 | 1.47776668 | 3.15E-06 | NA |
| rs575473987 | 8 | 5577494 | C | T | 421 | 0.00356295 | 3 | 22.2248653 | 3.88625754 | 1.07E-08 | NA |
| rs139062456 | 8 | 13251991 | C | T | 421 | 0.01781473 | 15 | 7.83515809 | 1.69687423 | 3.89E-06 | NA |
| rs185874707 | 8 | 18030674 | C | T | 421 | 0.01187648 | 10 | 9.13934765 | 1.94918644 | 2.75E-06 | NA |
| rs188415494 | 8 | 25613298 | C | T | 421 | 0.00356295 | 3 | 18.5169478 | 3.9914865 | 3.50E-06 | NA |
| rs1221830047 | 8 | 55690220 | T | C | 421 | 0.33016627 | 277 | 2.3570878 | 0.44879289 | 1.50E-07 | NA |
| rs117816016 | 8 | 103751262 | C | T | 421 | 0.00356295 | 3 | 19.8807023 | 4.26960212 | 3.22E-06 | NA |
| rs567383525 | 8 | 115029494 | C | T | 421 | 0.00356295 | 3 | 17.7455227 | 3.71970627 | 1.84E-06 | NA |
| rs545550279 | 8 | 115552429 | G | T | 421 | 0.00356295 | 3 | 19.1812421 | 4.19212988 | 4.75E-06 | NA |
| rs536803366 | 8 | 123001411 | T | C | 421 | 0.0023753 | 3 | 19.2098639 | 4.00952057 | 1.66E-06 | NA |
| rs555249476 | 8 | 123001411 | T | C | 421 | 0.0023753 | 3 | 19.2098639 | 4.00952057 | 1.66E-06 | NA |
| rs532513136 | 8 | 135813748 | C | A | 421 | 0.00356295 | 3 | 19.9490594 | 4.03118599 | 7.47E-07 | NA |
| rs532730683 | 9 | 1784492 | G | T | 421 | 0.00356295 | 3 | 18.726838 | 3.8314675 | 1.02E-06 | NA |
| rs540065886 | 9 | 2770228 | T | C | 421 | 0.00356295 | 3 | 19.9489151 | 4.18802805 | 1.90E-06 | NA |
| rs543844012 | 9 | 30107023 | C | T | 421 | 0.00356295 | 3 | 18.1160485 | 3.9446103 | 4.38E-06 | NA |
| rs148556485 | 9 | 84023826 | A | C | 421 | 0.0023753 | 3 | 21.3800152 | 4.26618316 | 5.40E-07 | NA |
| rs140782222 | 9 | 84028894 | T | C | 421 | 0.0023753 | 3 | 21.791526 | 4.31928461 | 4.53E-07 | NA |
| rs188034471 | 9 | 85937714 | G | A | 421 | 0.00475059 | 4 | 14.7559999 | 3.14925113 | 2.79E-06 | NA |
| rs190294315 | 9 | 85945465 | C | T | 421 | 0.00593824 | 5 | 14.5664276 | 2.88126649 | 4.29E-07 | NA |
| rs545690161 | 9 | 93030699 | G | A | 421 | 0.00593824 | 6 | 17.1436874 | 3.00318779 | 1.14E-08 | NA |
| rs565682685 | 9 | 93222328 | T | C | 421 | 0.00475059 | 4 | 18.938397 | 3.7206174 | 3.58E-07 | NA |
| rs183737367 | 9 | 93330047 | T | C | 421 | 0.00356295 | 3 | 23.3452306 | 4.24471431 | 3.80E-08 | NA |
| rs187213609 | 9 | 93415465 | C | T | 421 | 0.00356295 | 3 | 23.1975304 | 4.27412656 | 5.72E-08 | NA |
| rs150027952 | 9 | 103385416 | A | G | 421 | 0.0023753 | 3 | 21.8616648 | 4.51464643 | 1.28E-06 | NA |
| rs146207930 | 9 | 129042336 | A | G | 421 | 0.00712589 | 6 | 11.8624321 | 2.51502465 | 2.40E-06 | NA |
| rs78296164 | 9 | 135266715 | C | T | 421 | 0.00831354 | 7 | 11.6095338 | 2.33917557 | 6.94E-07 | NA |
| rs77871739 | 9 | 138552309 | G | A | 421 | 0.00475059 | 4 | 15.0724603 | 3.23606181 | 3.20E-06 | NA |
| rs118040657 | 10 | 3472846 | C | T | 421 | 0.00831354 | 8 | 12.1147712 | 2.64434694 | 4.62E-06 | NA |
| rs184425183 | 10 | 13457520 | A | G | 421 | 0.00356295 | 3 | 22.6683174 | 3.77549648 | 1.92E-09 | NA |
| rs184458518 | 10 | 13471195 | T | G | 421 | 0.00356295 | 3 | 22.8015789 | 3.77123942 | 1.48E-09 | NA |
| rs117998251 | 10 | 13497976 | C | T | 421 | 0.00356295 | 3 | 22.9543311 | 3.77480488 | 1.19E-09 | NA |
| rs117025967 | 10 | 20207052 | C | A | 421 | 0.01306413 | 11 | 9.66973563 | 1.927439 | 5.25E-07 | NA |
| rs529011661 | 10 | 20372316 | G | A | 421 | 0.00475059 | 4 | 15.6393469 | 3.3125339 | 2.34E-06 | NA |
| rs138249376 | 10 | 63515829 | T | G | 421 | 0.00475059 | 4 | 16.4300711 | 3.59466791 | 4.86E-06 | NA |
| rs140277951 | 10 | 82359100 | G | A | 421 | 0.00831354 | 6 | 13.7463158 | 2.64333932 | 1.99E-07 | NA |
| rs566018180 | 10 | 86755052 | C | T | 421 | 0.00356295 | 3 | 18.563552 | 4.04215862 | 4.38E-06 | NA |

| | | | | | | | | | | | |
|---|---|---|---|---|---|---|---|---|---|---|---|
| rs140706881 | 10 | 96217535 | G | A | 421 | 0.00475059 | 4 | 18.974805 | 3.93288931 | 1.40E-06 | NA |
| rs117913371 | 10 | 102918486 | G | A | 421 | 0.02494062 | 21 | 7.86609555 | 1.40785512 | 2.31E-08 | NA |
| rs752259256 | 10 | 104925319 | T | C | 421 | 0.00356295 | 3 | 20.4258639 | 4.45411591 | 4.52E-06 | NA |
| rs180828621 | 10 | 124533409 | G | A | 421 | 0.00712589 | 6 | 15.1380574 | 2.79957627 | 6.40E-08 | NA |
| rs147393020 | 10 | 124779274 | A | G | 421 | 0.00593824 | 5 | 14.1853505 | 2.95980896 | 1.65E-06 | NA |
| rs193093906 | 10 | 126705489 | G | A | 421 | 0.00950119 | 9 | 10.5219537 | 2.25840517 | 3.18E-06 | NA |
| rs541653703 | 11 | 18701786 | G | A | 421 | 0.00475059 | 4 | 18.3169499 | 3.27243418 | 2.18E-08 | NA |
| rs118093638 | 11 | 18718324 | C | T | 421 | 0.00593824 | 5 | 14.2657259 | 2.91296506 | 9.72E-07 | NA |
| rs181812512 | 11 | 66665729 | C | T | 421 | 0.00356295 | 3 | 19.5093168 | 4.24219816 | 4.25E-06 | NA |
| rs529345909 | 11 | 67110852 | A | G | 421 | 0.00356295 | 3 | 18.70153 | 4.09206035 | 4.87E-06 | NA |
| rs544042801 | 11 | 68460243 | G | A | 421 | 0.00475059 | 4 | 15.8869866 | 3.40519019 | 3.08E-06 | NA |
| rs149949098 | 11 | 95099866 | G | A | 421 | 0.01662708 | 13 | 8.55927625 | 1.81883308 | 2.53E-06 | NA |
| rs148781275 | 11 | 103640603 | A | G | 421 | 0.00475059 | 5 | 15.7111853 | 3.32183832 | 2.25E-06 | NA |
| rs141281289 | 11 | 123693024 | A | G | 421 | 0.00593824 | 6 | 15.811518 | 3.0889522 | 3.08E-07 | NA |
| rs528140343 | 11 | 125719227 | A | C | 421 | 0.00356295 | 3 | 18.2835521 | 3.72738029 | 9.33E-07 | NA |
| rs546409459 | 11 | 125754989 | A | G | 421 | 0.00356295 | 3 | 18.9195474 | 4.00286836 | 2.28E-06 | NA |
| rs528609331 | 11 | 125842195 | C | T | 421 | 0.00356295 | 3 | 19.5812418 | 3.94832705 | 7.07E-07 | NA |
| rs189360484 | 12 | 1870510 | A | G | 421 | 0.00475059 | 4 | 16.8078794 | 3.31424855 | 3.95E-07 | NA |
| rs141754456 | 12 | 20151132 | T | C | 421 | 0.00712589 | 7 | 12.1460333 | 2.32842921 | 1.82E-07 | NA |
| rs118184666 | 12 | 20424749 | G | A | 421 | 0.00712589 | 6 | 10.9509803 | 2.32477242 | 2.47E-06 | NA |
| rs549931083 | 12 | 20516286 | A | C | 421 | 0.00356295 | 3 | 14.3282694 | 2.79891395 | 3.07E-07 | NA |
| rs371879555 | 12 | 23015962 | T | C | 421 | 0.00356295 | 3 | 20.1033765 | 4.20002323 | 1.70E-06 | NA |
| rs183466664 | 12 | 26821687 | A | G | 421 | 0.00475059 | 4 | 16.0181882 | 3.45609016 | 3.57E-06 | NA |
| rs77353774 | 12 | 28248852 | G | A | 421 | 0.00712589 | 6 | 13.1054362 | 2.67954103 | 1.00E-06 | NA |
| rs113167689 | 12 | 28435962 | C | T | 421 | 0.00712589 | 6 | 14.1364135 | 2.73335629 | 2.32E-07 | NA |
| rs117991215 | 12 | 28511473 | T | C | 421 | 0.00831354 | 6 | 14.333666 | 2.77918736 | 2.50E-07 | NA |
| rs191930622 | 12 | 48284655 | G | A | 421 | 0.0023753 | 3 | 19.6783579 | 4.30610368 | 4.88E-06 | NA |
| rs56302696 | 12 | 48292830 | G | A | 421 | 0.0023753 | 3 | 19.8959041 | 4.31584707 | 4.03E-06 | NA |
| rs185620578 | 12 | 48569399 | C | T | 421 | 0.0023753 | 3 | 20.7232267 | 4.39505442 | 2.42E-06 | NA |
| rs190806532 | 12 | 48862147 | G | T | 421 | 0.0023753 | 3 | 21.326955 | 4.43856043 | 1.55E-06 | NA |
| rs568658857 | 12 | 49853998 | G | A | 421 | 0.00593824 | 5 | 14.2361193 | 3.09423422 | 4.21E-06 | NA |
| rs137880949 | 12 | 63306297 | T | C | 421 | 0.00475059 | 3 | 21.6084034 | 4.13589168 | 1.75E-07 | NA |
| rs191053292 | 12 | 63445280 | T | C | 421 | 0.00356295 | 3 | 22.9266488 | 4.11439732 | 2.51E-08 | NA |
| rs182437250 | 12 | 63608466 | T | C | 421 | 0.00475059 | 4 | 18.8083451 | 3.79343554 | 7.12E-07 | NA |
| rs180764936 | 12 | 101498621 | T | C | 421 | 0.00356295 | 3 | 19.3473053 | 4.0200238 | 1.49E-06 | NA |
| rs185855183 | 12 | 101505126 | C | T | 421 | 0.00356295 | 3 | 18.7397727 | 3.86137343 | 1.22E-06 | NA |
| rs139598422 | 13 | 23887014 | A | G | 421 | 0.00356295 | 4 | 20.4886522 | 3.64167194 | 1.84E-08 | NA |
| rs150077525 | 13 | 57979549 | A | G | 421 | 0.00712589 | 7 | 12.9984875 | 2.71312414 | 1.66E-06 | NA |
| rs534845494 | 13 | 58213864 | A | G | 421 | 0.00475059 | 5 | 15.9431765 | 3.18484007 | 5.56E-07 | NA |
| rs140062526 | 13 | 59286033 | G | A | 421 | 0.00593824 | 5 | 14.0926088 | 3.02052903 | 3.08E-06 | NA |
| rs546286713 | 13 | 91411079 | G | A | 421 | 0.00475059 | 4 | 15.7027841 | 3.3611216 | 2.98E-06 | NA |
| rs567080482 | 13 | 95075405 | T | C | 421 | 0.00475059 | 4 | 15.4999561 | 3.31620837 | 2.95E-06 | NA |
| rs142928734 | 13 | 101601082 | G | A | 421 | 0.00356295 | 3 | 17.009593 | 3.70044989 | 4.29E-06 | NA |
| rs556680896 | 13 | 101602415 | C | T | 421 | 0.00356295 | 3 | 17.0254659 | 3.70286799 | 4.27E-06 | NA |
| rs184265355 | 13 | 108011352 | A | C | 421 | 0.00593824 | 6 | 14.4271719 | 2.91829792 | 7.67E-07 | NA |
| rs572961122 | 13 | 108012985 | C | T | 421 | 0.00593824 | 6 | 13.3354896 | 2.85827607 | 3.08E-06 | NA |
| rs528809914 | 13 | 113041256 | G | A | 421 | 0.00356295 | 3 | 19.5048552 | 3.76867111 | 2.27E-07 | NA |
| rs180765647 | 13 | 114427311 | G | T | 421 | 0.0023753 | 3 | 20.1246521 | 4.37397806 | 4.20E-06 | NA |
| rs138215817 | 14 | 22641516 | A | G | 421 | 0.00475059 | 4 | 16.311019 | 3.27367022 | 6.28E-07 | NA |
| rs74704551 | 14 | 30161887 | C | T | 421 | 0.00356295 | 3 | 24.1990705 | 4.31064693 | 1.98E-08 | NA |
| rs176783 | 14 | 46280913 | A | G | 421 | 0.32185273 | 267 | 2.28010147 | 0.49376374 | 3.88E-06 | NA |
| rs143048774 | 14 | 46294660 | A | C | 421 | 0.31710214 | 265 | 2.28482299 | 0.49630858 | 4.15E-06 | NA |

| | | | | | | | | | | | |
|---|---|---|---|---|---|---|---|---|---|---|---|
| rs428110 | 14 | 46294660 | A | C | 421 | 0.31710214 | 265 | 2.28482299 | 0.49630858 | 4.15E-06 | NA |
| rs116862847 | 14 | 64141677 | C | T | 421 | 0.00712589 | 7 | 15.3160369 | 2.95194262 | 2.12E-07 | NA |
| rs569916471 | 14 | 75891342 | G | A | 421 | 0.00475059 | 5 | 15.6404088 | 3.3537133 | 3.11E-06 | NA |
| rs113767990 | 14 | 81717563 | G | A | 421 | 0.00475059 | 4 | 14.5811155 | 3.19363095 | 4.98E-06 | NA |
| rs190251199 | 14 | 105590577 | T | C | 421 | 0.00356295 | 3 | 20.0125292 | 4.01627173 | 6.27E-07 | NA |
| rs185155853 | 15 | 41244100 | C | T | 421 | 0.00475059 | 5 | 15.9164657 | 3.40838631 | 3.02E-06 | NA |
| rs144026361 | 15 | 41248669 | C | T | 421 | 0.00475059 | 5 | 16.1372994 | 3.43734659 | 2.67E-06 | NA |
| rs558614420 | 15 | 41810870 | C | T | 421 | 0.00593824 | 6 | 13.9750831 | 3.02772732 | 3.92E-06 | NA |
| rs138109686 | 15 | 42051442 | A | G | 421 | 0.00593824 | 6 | 13.5956705 | 2.96761718 | 4.62E-06 | NA |
| rs145896760 | 15 | 42119222 | G | A | 421 | 0.00593824 | 6 | 13.3688825 | 2.92346809 | 4.81E-06 | NA |
| rs140642138 | 15 | 42125165 | G | A | 421 | 0.00593824 | 6 | 13.5832221 | 2.96274049 | 4.55E-06 | NA |
| rs6080 | 15 | 58837933 | C | A | 421 | 0.04394299 | 36 | 5.66476745 | 1.2393253 | 4.86E-06 | NA |
| rs145439370 | 15 | 58879765 | T | C | 421 | 0.03087886 | 26 | 6.89328891 | 1.40339117 | 9.02E-07 | NA |
| rs149425014 | 15 | 58951660 | T | C | 421 | 0.02612827 | 22 | 7.55224792 | 1.58320604 | 1.84E-06 | NA |
| rs193253461 | 15 | 59229353 | A | G | 421 | 0.01306413 | 12 | 10.5374468 | 2.07404763 | 3.76E-07 | NA |
| rs184117160 | 15 | 59404306 | C | T | 421 | 0.01425178 | 11 | 10.3417236 | 2.13366338 | 1.25E-06 | NA |
| rs80292573 | 15 | 59435086 | T | G | 421 | 0.03444181 | 30 | 6.55323491 | 1.32696583 | 7.87E-07 | NA |
| rs182303755 | 15 | 59634792 | A | C | 421 | 0.01306413 | 12 | 10.5479917 | 2.09359291 | 4.70E-07 | NA |
| rs138217865 | 15 | 94392882 | C | T | 421 | 0.00475059 | 4 | 16.709641 | 3.45193896 | 1.29E-06 | NA |
| rs553840536 | 16 | 25697895 | A | G | 421 | 0.00356295 | 3 | 19.6203442 | 4.23547954 | 3.61E-06 | NA |
| rs183817723 | 16 | 59302775 | C | T | 421 | 0.00356295 | 4 | 17.5860507 | 3.7341919 | 2.48E-06 | NA |
| rs144954214 | 16 | 76179362 | A | G | 421 | 0.0023753 | 3 | 21.8113263 | 4.43942529 | 8.96E-07 | NA |
| rs529523094 | 16 | 77715551 | A | G | 421 | 0.00356295 | 3 | 18.1584346 | 3.72617419 | 1.10E-06 | NA |
| rs146728064 | 17 | 19265440 | G | A | 421 | 0.00712589 | 6 | 12.6400411 | 2.73467115 | 3.80E-06 | NA |
| rs184613584 | 17 | 48508221 | A | C | 421 | 0.00712589 | 6 | 14.1377974 | 2.8212234 | 5.41E-07 | NA |
| rs191271637 | 17 | 52123260 | A | G | 421 | 0.00356295 | 3 | 18.379836 | 3.92083355 | 2.76E-06 | NA |
| rs185819304 | 18 | 27001580 | G | A | 421 | 0.00356295 | 3 | 19.9401113 | 3.94501953 | 4.32E-07 | NA |
| rs187942235 | 18 | 27030430 | C | T | 421 | 0.00356295 | 3 | 20.3968658 | 3.96963025 | 2.77E-07 | NA |
| rs143538552 | 18 | 29050262 | A | G | 421 | 0.00356295 | 3 | 19.1457218 | 3.63679703 | 1.41E-07 | NA |
| rs373746073 | 18 | 29058384 | C | A | 421 | 0.00356295 | 3 | 19.0847197 | 3.63798064 | 1.55E-07 | NA |
| rs146333745 | 18 | 55497457 | C | T | 421 | 0.00356295 | 4 | 17.4215784 | 3.56782335 | 1.04E-06 | NA |
| rs185464792 | 19 | 18797371 | C | T | 421 | 0.0023753 | 3 | 22.5896015 | 4.50191473 | 5.23E-07 | NA |
| rs186768950 | 19 | 18806124 | C | A | 421 | 0.0023753 | 3 | 22.7152061 | 4.50776479 | 4.68E-07 | NA |
| rs541288561 | 19 | 18869445 | T | G | 421 | 0.00475059 | 5 | 15.6119012 | 3.13183483 | 6.20E-07 | NA |
| rs559008174 | 19 | 18876059 | C | T | 421 | 0.00475059 | 5 | 15.6025661 | 3.12812006 | 6.11E-07 | NA |
| rs570407448 | 19 | 18880030 | G | A | 421 | 0.00475059 | 5 | 15.4977984 | 3.12412555 | 7.02E-07 | NA |
| rs546144116 | 19 | 19563339 | C | T | 421 | 0.0023753 | 3 | 23.1354815 | 4.47162758 | 2.29E-07 | NA |
| rs560206697 | 19 | 20729098 | C | T | 421 | 0.0023753 | 3 | 24.9989943 | 4.55350673 | 4.02E-08 | NA |
| rs111285015 | 19 | 23123198 | G | A | 421 | 0.00356295 | 3 | 27.3347642 | 4.62083983 | 3.31E-09 | NA |
| rs1008091735 | 19 | 31090099 | T | C | 421 | 0.00356295 | 3 | 18.4689243 | 3.69079909 | 5.61E-07 | NA |
| rs148433854 | 19 | 31096478 | G | A | 421 | 0.00356295 | 3 | 18.6267585 | 3.68806754 | 4.41E-07 | NA |
| rs559228693 | 20 | 15963329 | G | A | 421 | 0.00593824 | 5 | 14.3643551 | 3.08196497 | 3.15E-06 | NA |
| rs557092705 | 20 | 34189911 | C | T | 421 | 0.00356295 | 3 | 19.4769546 | 4.2480073 | 4.54E-06 | NA |
| rs184785969 | 21 | 17171431 | C | A | 421 | 0.00356295 | 3 | 19.8736486 | 3.70683937 | 8.26E-08 | NA |
| rs117280553 | 21 | 17207163 | T | C | 421 | 0.00356295 | 3 | 20.087417 | 3.70274826 | 5.80E-08 | NA |
| rs79486609 | 21 | 17245006 | G | A | 421 | 0.00356295 | 3 | 20.6624886 | 3.77824957 | 4.53E-08 | NA |
| rs73227413 | 21 | 23136973 | G | A | 421 | 0.03444181 | 28 | 5.96205514 | 1.27150176 | 2.75E-06 | NA |
| rs75024143 | 21 | 23156546 | G | T | 421 | 0.01425178 | 12 | 9.81254241 | 2.0684542 | 2.10E-06 | NA |
| rs192134381 | 21 | 23450714 | T | C | 421 | 0.00356295 | 3 | 21.788812 | 3.73125357 | 5.23E-09 | NA |
| rs397836601 | 21 | 23450714 | T | C | 421 | 0.00356295 | 3 | 21.788812 | 3.73125357 | 5.23E-09 | NA |
| rs118183140 | 21 | 35477486 | C | T | 421 | 0.02019002 | 17 | 7.5893248 | 1.56122232 | 1.17E-06 | NA |
| rs183586634 | 21 | 38763032 | G | A | 421 | 0.00593824 | 5 | 14.3132202 | 2.99210048 | 1.72E-06 | NA |

| | | | | | | | | | | | |
|---|---|---|---|---|---|---|---|---|---|---|---|
| rs117185941 | 21 | 38766484 | G | A | 421 | 0.00593824 | 5 | 15.3808786 | 3.17143013 | 1.24E-06 | NA |
| rs1329159859 | 21 | 38766484 | G | A | 421 | 0.00593824 | 5 | 15.3808786 | 3.17143013 | 1.24E-06 | NA |
| rs118084887 | 21 | 38863820 | T | C | 421 | 0.00593824 | 5 | 14.9382616 | 3.13674741 | 1.91E-06 | NA |
| rs150539922 | 21 | 43276916 | T | C | 421 | 0.00356295 | 3 | 16.6696937 | 3.64477381 | 4.79E-06 | NA |
| rs113625788 | 22 | 19969182 | C | T | 421 | 0.00831354 | 7 | 11.8084829 | 2.42941561 | 1.17E-06 | NA |
| rs796777817 | 22 | 32824278 | G | A | 421 | 0.00356295 | 3 | 20.9098395 | 4.15538274 | 4.85E-07 | NA |
| rs541680196 | 22 | 40528090 | G | A | 421 | 0.00593824 | 5 | 14.3867808 | 2.94903659 | 1.07E-06 | NA |
| rs185139807 | 22 | 40594781 | G | A | 421 | 0.00593824 | 5 | 14.4330526 | 2.95312686 | 1.02E-06 | NA |
| rs141127122 | 22 | 40604439 | G | A | 421 | 0.00475059 | 4 | 15.3824875 | 3.30083549 | 3.16E-06 | NA |
| rs148998974 | 22 | 40620530 | A | G | 421 | 0.00593824 | 5 | 14.5129532 | 2.94976024 | 8.65E-07 | NA |
| rs555040883 | 22 | 40631476 | G | A | 421 | 0.00475059 | 4 | 15.4273069 | 3.30542717 | 3.05E-06 | NA |
| rs182959028 | 22 | 45823032 | T | C | 421 | 0.00831354 | 7 | 11.944461 | 2.53146 | 2.38E-06 | NA |
| rs150946694 | 22 | 46853180 | T | C | 421 | 0.00475059 | 4 | 15.3082479 | 3.29125059 | 3.30E-06 | NA |

**Supplementary Table S7. Independent Replication**

Chennai-2 cohort, case-control data, comparing to discovery 12M quantitative trait (QT) data

| | |
|---|---|
| Threshold for replicative significance = | 5.00E-02 |
| Total SNPs found: | 41 |
| Total risk loci found: | 22 |

Thick boxed: SNPs that attained adjusted p-values of replication significance
NA: not applicable

| rsid | chr | pos_37 | REF | ALT | n.obs | caf | MAC | Est | Est.SE | Score.pval | pval.chennai | adj pval SNPs | adj pval loci | Closest gene | Prioritized gene |
|---|---|---|---|---|---|---|---|---|---|---|---|---|---|---|---|
| rs2427460 | 20 | 61590782 | T | C | 439 | 0.48633257 | 420 | -2.574044 | 0.51325892 | 5.30E-07 | 2.80E-04 | 1.15E-02 | 6.17E-03 | SLC17A9 | SLC17A9 |
| rs9636964 | 21 | 41304765 | A | G | 439 | 0.88496583 | 115 | -4.2543192 | 0.82744371 | 2.73E-07 | 3.07E-02 | 1.00E+00 | 6.76E-01 | PCP4 | PCP4 |
| rs147944608 | 10 | 110423709 | C | T | 439 | 0.01252847 | 11 | 11.2205243 | 2.29229161 | 9.84E-07 | 5.91E-02 | 1.00E+00 | 1.00E+00 | XPNPEP1 | XPNPEP1 |
| rs144138711 | 9 | 99922419 | T | C | 439 | 0.04555809 | 40 | 6.08547131 | 1.26091918 | 1.39E-06 | 5.95E-02 | 1.00E+00 | 1.00E+00 | ANKRD18CP | CCDC180 |
| rs117591241 | 3 | 190584679 | A | G | 439 | 0.00341686 | 4 | 19.9014028 | 4.33331389 | 4.38E-06 | 8.12E-02 | 1.00E+00 | 1.00E+00 | GMNC | GMNC |
| rs1472243465 | 3 | 190584679 | A | G | 439 | 0.00341686 | 4 | 19.9014028 | 4.33331389 | 4.38E-06 | 8.12E-02 | 1.00E+00 | 1.00E+00 | GMNC | GMNC |
| rs74572772 | 22 | 34492649 | A | G | 439 | 0.01822323 | 16 | 10.2225183 | 1.94126774 | 1.40E-07 | 1.09E-01 | 1.00E+00 | 1.00E+00 | LARGE1 | LARGE1 |
| rs61918041 | 12 | 6681868 | C | T | 439 | 0.023918 | 21 | 7.97264643 | 1.73636056 | 4.40E-06 | 1.93E-01 | 1.00E+00 | 1.00E+00 | CHD4 | CHD4 |
| rs56821264 | 6 | 148703901 | C | T | 439 | 0.00683371 | 6 | 14.7509213 | 3.11419566 | 2.17E-06 | 2.05E-01 | 1.00E+00 | 1.00E+00 | SASH1 | SASH1 |
| rs917335559 | 6 | 148703901 | C | T | 439 | 0.00683371 | 6 | 14.7509213 | 3.11419566 | 2.17E-06 | 2.05E-01 | 1.00E+00 | 1.00E+00 | SASH1 | SASH1 |
| rs1162141034 | 2 | 67825685 | C | T | 439 | 0.07972665 | 71 | 4.55991411 | 0.98044721 | 3.31E-06 | 2.13E-01 | 1.00E+00 | 1.00E+00 | ETAA1 | ETAA1 |
| rs146465493 | 2 | 67825685 | C | T | 439 | 0.07972665 | 71 | 4.55991411 | 0.98044721 | 3.31E-06 | 2.13E-01 | 1.00E+00 | 1.00E+00 | ETAA1 | ETAA1 |
| rs2902021 | 2 | 67825685 | C | T | 439 | 0.07972665 | 71 | 4.55991411 | 0.98044721 | 3.31E-06 | 2.13E-01 | 1.00E+00 | 1.00E+00 | ETAA1 | ETAA1 |
| rs75082290 | 2 | 67831153 | T | G | 439 | 0.0785877 | 69 | 4.69395387 | 0.98072733 | 1.70E-06 | 2.58E-01 | 1.00E+00 | 1.00E+00 | ETAA1 | ETAA1 |
| rs9974985 | 21 | 41307573 | G | A | 439 | 0.88610478 | 114 | -4.3459395 | 0.82924282 | 1.60E-07 | 3.20E-01 | 1.00E+00 | 1.00E+00 | PCP4 | PCP4 |
| rs7534177 | 1 | 65965720 | A | G | 439 | 0.05694761 | 49 | 4.96123378 | 1.0598616 | 2.85E-06 | 3.28E-01 | 1.00E+00 | 1.00E+00 | LEPR | LEPR |
| rs4896997 | 6 | 148683924 | C | T | 439 | 0.00683371 | 6 | 14.4320923 | 3.09188376 | 3.05E-06 | 3.29E-01 | 1.00E+00 | 1.00E+00 | SASH1 | SASH1 |
| rs62515405 | 8 | 57055978 | G | A | 439 | 0.023918 | 21 | 8.07691204 | 1.76428755 | 4.69E-06 | 3.29E-01 | 1.00E+00 | 1.00E+00 | PLAG1 | PLAG1 |
| rs17078283 | 6 | 148696920 | C | T | 439 | 0.00683371 | 6 | 14.4302821 | 3.0918048 | 3.05E-06 | 3.42E-01 | 1.00E+00 | 1.00E+00 | SASH1 | SASH1 |
| rs17543620 | 4 | 169934725 | T | C | 439 | 0.0501139 | 44 | 5.42796755 | 1.17982097 | 4.21E-06 | 3.47E-01 | 1.00E+00 | 1.00E+00 | CBR4 | CBR4 |
| rs1005412 | 21 | 41308948 | A | G | 439 | 0.88724374 | 112 | -4.4521096 | 0.85237138 | 1.76E-07 | 3.96E-01 | 1.00E+00 | 1.00E+00 | PCP4 | PCP4 |
| rs386818616 | 21 | 41308948 | A | G | 439 | 0.88724374 | 112 | -4.4521096 | 0.85237138 | 1.76E-07 | 3.96E-01 | 1.00E+00 | 1.00E+00 | PCP4 | PCP4 |
| rs117699122 | 12 | 560262 | T | C | 439 | 0.00341686 | 4 | 21.4238256 | 4.40952328 | 1.18E-06 | 4.03E-01 | 1.00E+00 | 1.00E+00 | CCDC77 | CCDC77 |
| rs9305683 | 21 | 41305720 | G | A | 439 | 0.88952164 | 112 | -4.1163679 | 0.83634363 | 8.57E-07 | 4.08E-01 | 1.00E+00 | 1.00E+00 | PCP4 | PCP4 |
| rs12114488 | 8 | 62672723 | G | A | 439 | 0.30523918 | 266 | 2.62307914 | 0.54257702 | 1.33E-06 | 4.11E-01 | 1.00E+00 | 1.00E+00 | MIR4470 | ADPH |
| rs139816293 | 20 | 30921343 | C | T | 439 | 0.00569476 | 6 | 17.1384248 | 3.52537223 | 1.17E-06 | 4.23E-01 | 1.00E+00 | 1.00E+00 | KIF3B | KIF3B |
| rs74683551 | 1 | 112354418 | G | A | 439 | 0.01594533 | 14 | 9.54722832 | 2.04640679 | 3.08E-06 | 4.23E-01 | 1.00E+00 | 1.00E+00 | KCND3 | KCND3 |
| rs76356799 | 3 | 179593768 | G | A | 439 | 0.00455581 | 4 | 19.2867007 | 3.76039319 | 2.91E-07 | 5.66E-01 | 1.00E+00 | 1.00E+00 | PEX5L | PEX5L |
| rs143686474 | 7 | 16291999 | A | C | 439 | 0.01252847 | 11 | 10.8531498 | 2.37513184 | 4.89E-06 | 5.72E-01 | 1.00E+00 | 1.00E+00 | CRPPA | SOSTDC1 |
| rs62192733 | 20 | 1928157 | A | G | 439 | 0.00683371 | 7 | 15.744618 | 3.35866563 | 2.76E-06 | 5.76E-01 | 1.00E+00 | 1.00E+00 | PDYN | SIRPA |
| rs147669485 | 18 | 26603650 | G | T | 439 | 0.00341686 | 4 | 22.1107639 | 4.14923198 | 9.88E-08 | 5.80E-01 | 1.00E+00 | 1.00E+00 | CDH2 | CDH2 |
| rs7275595 | 21 | 41307923 | G | A | 439 | 0.88838269 | 113 | -4.2609754 | 0.83407868 | 3.25E-07 | 6.02E-01 | 1.00E+00 | 1.00E+00 | PCP4 | PCP4 |
| rs2606194 | 17 | 77210823 | A | G | 439 | 0.94874715 | 44 | -5.6956979 | 1.16435773 | 1.00E-06 | 6.65E-01 | 1.00E+00 | 1.00E+00 | RBFOX3 | RBFOX3 |
| rs17127656 | 1 | 65943471 | C | T | 439 | 0.05580866 | 49 | 5.07046538 | 1.05780641 | 1.64E-06 | 6.74E-01 | 1.00E+00 | 1.00E+00 | LEPR | LEPR |
| rs77297738 | 22 | 34473543 | C | T | 439 | 0.01822323 | 16 | 10.2480572 | 1.97167592 | 2.02E-07 | 6.78E-01 | 1.00E+00 | 1.00E+00 | LARGE1 | LARGE1 |
| rs17615362 | 4 | 169934087 | G | A | 439 | 0.0501139 | 44 | 5.41974472 | 1.17918425 | 4.30E-06 | 7.04E-01 | 1.00E+00 | 1.00E+00 | CBR4 | CBR4 |
| rs7518849 | 1 | 65948791 | T | C | 439 | 0.05580866 | 49 | 5.07456956 | 1.05964469 | 1.68E-06 | 7.24E-01 | 1.00E+00 | 1.00E+00 | LEPR | LEPR |
| rs9981433 | 21 | 41309565 | G | T | 439 | 0.88154897 | 116 | -4.1220124 | 0.84715422 | 1.14E-06 | 8.07E-01 | 1.00E+00 | 1.00E+00 | LEPR | LEPR |
| rs80019988 | 22 | 34493647 | G | A | 439 | 0.01822323 | 16 | 10.0623306 | 1.91665407 | 1.52E-07 | 8.18E-01 | 1.00E+00 | 1.00E+00 | LARGE1 | LARGE1 |
| rs4131286 | 6 | 148688963 | G | T | 439 | 0.00683371 | 6 | 14.4284672 | 3.09172532 | 3.06E-06 | 9.26E-01 | 1.00E+00 | 1.00E+00 | SASH1 | SASH1 |
| rs11579567 | 1 | 65957141 | C | A | 439 | 0.05580866 | 49 | 5.02973793 | 1.06097379 | 2.13E-06 | 9.78E-01 | 1.00E+00 | 1.00E+00 | LEPR | LEPR |
| rs142934021 | 5 | 52662672 | G | A | 439 | 0.00797267 | 7 | 18.824715 | 3.31820294 | 1.40E-08 | NA | | | | |
| rs138164904 | 16 | 6858239 | C | T | 439 | 0.00341686 | 3 | 23.892641 | 4.3369669 | 3.61E-08 | NA | | | | |

| | | | | | | | | | | | |
|---|---|---|---|---|---|---|---|---|---|---|---|
| rs138138661 | 3 | 147657814 | C | T | 439 | 0.00455581 | 4 | 23.1347457 | 4.25695836 | 5.49E-08 | NA |
| rs148997617 | 3 | 147755576 | C | T | 439 | 0.00455581 | 4 | 20.8774277 | 3.92648902 | 1.05E-07 | NA |
| rs148157126 | 20 | 30948839 | C | T | 439 | 0.00455581 | 5 | 19.7865001 | 3.76042103 | 1.43E-07 | NA |
| rs186792608 | 2 | 43096699 | A | G | 439 | 0.00341686 | 3 | 22.4512499 | 4.28172841 | 1.58E-07 | NA |
| rs113154814 | 2 | 68289497 | T | C | 439 | 0.00569476 | 5 | 19.8310188 | 3.7892894 | 1.66E-07 | NA |
| rs193041547 | 20 | 30771880 | T | C | 439 | 0.00455581 | 4 | 19.6791358 | 3.77081146 | 1.80E-07 | NA |
| rs146249289 | 20 | 30682231 | C | T | 439 | 0.00455581 | 4 | 19.6109527 | 3.77306035 | 2.02E-07 | NA |
| rs192855100 | 20 | 31108741 | G | A | 439 | 0.00455581 | 4 | 20.1986097 | 3.99111726 | 4.17E-07 | NA |
| rs562831582 | 19 | 51234310 | A | C | 439 | 0.00341686 | 4 | 21.0856548 | 4.21624838 | 5.70E-07 | NA |
| rs149859280 | 20 | 30678170 | C | T | 439 | 0.00455581 | 5 | 18.3057707 | 3.66075889 | 5.72E-07 | NA |
| rs138733283 | 20 | 30675687 | C | T | 439 | 0.00455581 | 5 | 18.2744744 | 3.65830138 | 5.87E-07 | NA |
| rs80212581 | 16 | 6412967 | C | T | 439 | 0.00341686 | 3 | 22.2762003 | 4.47237333 | 6.33E-07 | NA |
| rs148248743 | 3 | 136134595 | C | T | 439 | 0.0022779 | 3 | 27.1548587 | 5.46352726 | 6.69E-07 | NA |
| rs188353596 | 17 | 46078602 | C | T | 439 | 0.00341686 | 3 | 21.8057947 | 4.39141941 | 6.85E-07 | NA |
| rs113791989 | 13 | 61918888 | G | A | 439 | 0.0022779 | 3 | 25.2049582 | 5.08402789 | 7.13E-07 | NA |
| rs56324718 | 15 | 40280073 | G | A | 439 | 0.01025057 | 9 | 12.5117374 | 2.5288888 | 7.52E-07 | NA |
| rs191298981 | 2 | 68110046 | C | T | 439 | 0.00455581 | 3 | 24.0078714 | 4.85703224 | 7.70E-07 | NA |
| rs146919974 | 2 | 115252863 | T | C | 439 | 0.00455581 | 5 | 19.7280466 | 3.99554232 | 7.91E-07 | NA |
| rs145791959 | 20 | 30722429 | G | A | 439 | 0.00569476 | 5 | 17.9091953 | 3.64034112 | 8.67E-07 | NA |
| rs142956968 | 10 | 23490165 | C | T | 439 | 0.01594533 | 14 | 10.4167992 | 2.11909083 | 8.85E-07 | NA |
| rs532269430 | 13 | 74054452 | T | C | 439 | 0.00455581 | 5 | 18.6173752 | 3.7963004 | 9.39E-07 | NA |
| rs113063005 | 4 | 23509067 | T | C | 439 | 0.00683371 | 7 | 16.61763 | 3.39113209 | 9.57E-07 | NA |
| rs113537164 | 2 | 68234930 | A | C | 439 | 0.00455581 | 4 | 20.3890413 | 4.17097432 | 1.02E-06 | NA |
| rs138055631 | 20 | 30780644 | G | A | 439 | 0.00569476 | 5 | 17.6647762 | 3.62004915 | 1.06E-06 | NA |
| rs566724618 | 13 | 23463839 | C | T | 439 | 0.00341686 | 3 | 22.3433705 | 4.58061605 | 1.07E-06 | NA |
| rs145764464 | 10 | 23601847 | G | A | 439 | 0.01708428 | 15 | 9.5967306 | 1.96976348 | 1.10E-06 | NA |
| rs529110230 | 7 | 117175907 | T | C | 439 | 0.00341686 | 3 | 20.8583633 | 4.30235088 | 1.25E-06 | NA |
| rs182868205 | 3 | 177712448 | C | T | 439 | 0.00341686 | 3 | 22.8544303 | 4.73215662 | 1.37E-06 | NA |
| rs372194899 | 13 | 74357510 | A | G | 439 | 0.00341686 | 3 | 21.3012101 | 4.41516106 | 1.40E-06 | NA |
| rs368187808 | 13 | 74357508 | A | C | 439 | 0.00341686 | 3 | 21.2991251 | 4.41501637 | 1.41E-06 | NA |
| rs145804766 | 1 | 202122499 | C | T | 439 | 0.01025057 | 9 | 12.5864465 | 2.61118962 | 1.43E-06 | NA |
| rs182211730 | 7 | 116003615 | G | A | 439 | 0.00455581 | 4 | 18.7746033 | 3.8971981 | 1.45E-06 | NA |
| rs558784715 | 7 | 117043294 | A | G | 439 | 0.00341686 | 3 | 20.7591787 | 4.31689475 | 1.52E-06 | NA |
| rs189709453 | 3 | 177574308 | G | A | 439 | 0.00341686 | 3 | 23.2009936 | 4.82954374 | 1.56E-06 | NA |
| rs142721557 | 7 | 117218328 | A | C | 439 | 0.00341686 | 3 | 20.7329975 | 4.32245894 | 1.61E-06 | NA |
| rs576047962 | 7 | 117109021 | A | G | 439 | 0.00341686 | 3 | 20.7261549 | 4.32140182 | 1.62E-06 | NA |
| rs142215699 | 7 | 117199874 | T | C | 439 | 0.00341686 | 3 | 20.7292287 | 4.32245361 | 1.62E-06 | NA |
| rs143281973 | 7 | 116007996 | C | T | 439 | 0.00455581 | 4 | 18.2750778 | 3.81098198 | 1.62E-06 | NA |
| rs201355675 | 7 | 117225781 | G | A | 439 | 0.00341686 | 3 | 20.718124 | 4.32246722 | 1.64E-06 | NA |
| rs186767531 | 3 | 177639777 | T | C | 439 | 0.00341686 | 3 | 23.0749977 | 4.81664642 | 1.66E-06 | NA |
| rs181132315 | 4 | 129647369 | C | T | 439 | 0.00341686 | 3 | 22.1324679 | 4.62266164 | 1.69E-06 | NA |
| rs145421321 | 20 | 30862126 | C | T | 439 | 0.00569476 | 6 | 17.0212611 | 3.55829648 | 1.72E-06 | NA |
| rs143432612 | 20 | 30892775 | C | T | 439 | 0.00569476 | 6 | 16.9387232 | 3.54318154 | 1.75E-06 | NA |
| rs140276610 | 16 | 6433735 | C | T | 439 | 0.00341686 | 4 | 22.1806577 | 4.64062401 | 1.76E-06 | NA |
| rs559152067 | 18 | 66029612 | G | A | 439 | 0.0022779 | 3 | 24.7443692 | 5.18343862 | 1.81E-06 | NA |
| rs147221953 | 13 | 23485722 | G | A | 439 | 0.00341686 | 3 | 22.5809676 | 4.7360336 | 1.86E-06 | NA |
| rs556274646 | 11 | 439012 | C | T | 439 | 0.01025057 | 9 | 12.8914592 | 2.70847728 | 1.94E-06 | NA |
| rs190251199 | 14 | 105590577 | T | C | 439 | 0.00341686 | 3 | 22.6385061 | 4.76043817 | 1.98E-06 | NA |
| rs188993522 | 7 | 117274731 | T | C | 439 | 0.00341686 | 3 | 21.2984827 | 4.47867266 | 1.98E-06 | NA |
| rs568321148 | 2 | 29870348 | T | G | 439 | 0.00341686 | 3 | 20.7400203 | 4.36652717 | 2.04E-06 | NA |
| rs148815783 | 11 | 1013992 | C | T | 439 | 0.00797267 | 8 | 13.5980313 | 2.8647171 | 2.07E-06 | NA |
| rs76220567 | 2 | 13059945 | T | C | 439 | 0.00341686 | 4 | 20.6828868 | 4.35776216 | 2.07E-06 | NA |

| | | | | | | | | | | | |
|---|---|---|---|---|---|---|---|---|---|---|---|
| rs35145334 | 1 | 23465122 | A | G | 439 | 0.01708428 | 14 | 10.6552656 | 2.24620792 | 2.10E-06 | NA |
| rs114067899 | 1 | 85191044 | G | A | 439 | 0.00797267 | 7 | 13.7760673 | 2.90836365 | 2.17E-06 | NA |
| rs536781978 | 2 | 29733681 | A | G | 439 | 0.00341686 | 3 | 20.5561882 | 4.34258099 | 2.21E-06 | NA |
| rs147267707 | 4 | 43026558 | G | A | 439 | 0.00455581 | 4 | 18.0094385 | 3.80818408 | 2.25E-06 | NA |
| rs77618729 | 4 | 43050968 | T | C | 439 | 0.00455581 | 4 | 18.0130153 | 3.80914173 | 2.26E-06 | NA |
| rs547186621 | 20 | 6106741 | A | G | 439 | 0.00341686 | 4 | 21.5510829 | 4.5690881 | 2.40E-06 | NA |
| rs200198574 | 20 | 30946654 | G | A | 439 | 0.00569476 | 6 | 16.5282735 | 3.50702674 | 2.44E-06 | NA |
| rs532464521 | 4 | 43571039 | T | C | 439 | 0.00455581 | 4 | 17.9895528 | 3.81820438 | 2.46E-06 | NA |
| rs79252854 | 3 | 157523157 | C | T | 439 | 0.00455581 | 5 | 17.5464336 | 3.72541417 | 2.48E-06 | NA |
| rs79935606 | 2 | 177338635 | T | C | 439 | 0.00569476 | 5 | 16.3396833 | 3.47376974 | 2.55E-06 | NA |
| rs556435089 | 4 | 43551033 | G | A | 439 | 0.00455581 | 4 | 17.939079 | 3.81772146 | 2.62E-06 | NA |
| rs139390630 | 1 | 84866196 | G | A | 439 | 0.01138952 | 13 | 11.6975971 | 2.4994863 | 2.87E-06 | NA |
| rs149706477 | 3 | 66148228 | A | G | 439 | 0.00341686 | 3 | 20.9828687 | 4.4842531 | 2.88E-06 | NA |
| rs189094663 | 4 | 11623450 | G | A | 439 | 0.0022779 | 3 | 23.895416 | 5.10822708 | 2.90E-06 | NA |
| rs181182636 | 7 | 115654558 | A | G | 439 | 0.00341686 | 3 | 21.9472639 | 4.69244743 | 2.91E-06 | NA |
| rs76098744 | 1 | 112349372 | C | T | 439 | 0.01594533 | 14 | 9.5685657 | 2.04635193 | 2.93E-06 | NA |
| rs73202425 | 7 | 109155716 | T | C | 439 | 0.01252847 | 12 | 10.761844 | 2.30582166 | 3.05E-06 | NA |
| rs139055031 | 3 | 158586849 | T | C | 439 | 0.00569476 | 6 | 16.2853761 | 3.4955337 | 3.18E-06 | NA |
| rs138873576 | 7 | 116025359 | G | T | 439 | 0.00455581 | 4 | 17.6151711 | 3.78331079 | 3.22E-06 | NA |
| rs148483098 | 3 | 157382051 | C | A | 439 | 0.00569476 | 5 | 15.7357844 | 3.38211024 | 3.28E-06 | NA |
| rs187281112 | 15 | 33113387 | C | T | 439 | 0.00569476 | 5 | 14.7343356 | 3.16793649 | 3.30E-06 | NA |
| rs189912648 | 5 | 134765439 | C | T | 439 | 0.00341686 | 4 | 19.8604449 | 4.2716652 | 3.33E-06 | NA |
| rs557276277 | 4 | 19013698 | T | C | 439 | 0.00569476 | 5 | 19.0146344 | 4.09137627 | 3.36E-06 | NA |
| rs147869916 | 1 | 227307652 | A | G | 439 | 0.00569476 | 5 | 17.5964551 | 3.78625881 | 3.36E-06 | NA |
| rs367732718 | 19 | 35918004 | G | A | 439 | 0.0022779 | 3 | 23.8784923 | 5.13995752 | 3.39E-06 | NA |
| rs62515436 | 8 | 57141203 | G | T | 439 | 0.01708428 | 15 | 9.27545902 | 1.99804812 | 3.45E-06 | NA |
| rs189234695 | 3 | 147658158 | T | C | 439 | 0.00341686 | 3 | 23.9697134 | 5.16542129 | 3.48E-06 | NA |
| rs186893139 | 7 | 116096970 | C | T | 439 | 0.00455581 | 4 | 18.602598 | 4.01258383 | 3.55E-06 | NA |
| rs73384619 | 6 | 20458179 | C | T | 439 | 0.02277904 | 21 | 8.3916702 | 1.81241389 | 3.65E-06 | NA |
| rs114280794 | 3 | 132842203 | G | A | 439 | 0.01594533 | 14 | 8.76296877 | 1.89632016 | 3.82E-06 | NA |
| rs112148840 | 17 | 66233774 | C | T | 439 | 0.0022779 | 3 | 24.0241075 | 5.1994476 | 3.83E-06 | NA |
| rs147455971 | 4 | 140931712 | T | C | 439 | 0.00797267 | 5 | 17.2160142 | 3.72793057 | 3.87E-06 | NA |
| rs144788248 | 3 | 19586745 | T | C | 439 | 0.01138952 | 10 | 11.9086237 | 2.57879124 | 3.88E-06 | NA |
| rs111512950 | 5 | 153680427 | C | T | 439 | 0.00911162 | 8 | 13.567443 | 2.94598493 | 4.12E-06 | NA |
| rs149616342 | 12 | 99178813 | C | T | 439 | 0.01138952 | 10 | 11.2978214 | 2.454657 | 4.17E-06 | NA |
| rs190354334 | 7 | 116075846 | G | A | 439 | 0.00455581 | 5 | 17.3392581 | 3.7718663 | 4.29E-06 | NA |
| rs113244573 | 12 | 6684385 | G | A | 439 | 0.023918 | 21 | 7.97095746 | 1.73766156 | 4.49E-06 | NA |
| rs116897913 | 17 | 16839162 | C | T | 439 | 0.00341686 | 3 | 20.1396765 | 4.40294288 | 4.78E-06 | NA |
| rs143669489 | 3 | 159392885 | G | A | 439 | 0.00341686 | 3 | 24.2253199 | 5.30400674 | 4.94E-06 | NA |

**Supplementary Table S7. Independent Replication**

Chennai-2 cohort, case-control data, comparing to discovery 3M quantitative trait (QT) data

| | |
|---|---|
| Threshold for replicative significance: | 5.00E-02 |
| Total SNPs found: | 123 |
| Total risk loci found: | 41 |

Gray shading: SNPs and target genes of genome-wide significance in the discovery cohort
Thick boxed: SNPs that attained adjusted p-values of replication significance
NA: not applicable

| rsid | chr | pos_37 | REF | ALT | n.obs | caf | MAC | Est | Est.SE | Score.pval | pval.chennai | adj pval SNPs | adj pval loci | Closest gene | Prioritized gene |
|---|---|---|---|---|---|---|---|---|---|---|---|---|---|---|---|
| rs7822082 | 8 | 55690220 | T | C | 421 | 0.33016627 | 277 | 2.3570878 | 0.44879289 | 1.50E-07 | 8.66E-06 | 1.07E-03 | 3.55E-04 | RP1 | SOX17 |
| rs1561297 | 8 | 55678538 | A | C | 421 | 0.33254157 | 279 | 2.28315237 | 0.44743146 | 3.35E-07 | 7.41E-05 | 9.11E-03 | 3.04E-03 | RP1 | SOX17 |
| rs193153124 | 3 | 148330710 | A | G | 421 | 0.00593824 | 5 | 14.3657313 | 3.06514594 | 2.78E-06 | 3.56E-04 | 4.38E-02 | 1.46E-02 | AGTR1 | AGTR1 |
| rs12678939 | 8 | 55705021 | A | G | 421 | 0.39429929 | 331 | 2.01975618 | 0.44220051 | 4.94E-06 | 1.66E-03 | 2.04E-01 | 6.81E-02 | RP1 | SOX17 |
| rs446222 | 8 | 55574960 | G | A | 421 | 0.66627078 | 288 | -2.247819 | 0.45397995 | 7.37E-07 | 1.95E-03 | 2.40E-01 | 8.00E-02 | RP1 | SOX17 |
| rs2375537 | 8 | 55619508 | C | T | 421 | 0.33254157 | 280 | 2.23876848 | 0.44938849 | 6.30E-07 | 3.33E-03 | 4.10E-01 | 1.37E-01 | RP1 | SOX17 |
| rs2375536 | 8 | 55640722 | T | C | 421 | 0.347981 | 292 | 2.12635806 | 0.44857091 | 2.13E-06 | 3.38E-03 | 4.16E-01 | 1.39E-01 | RP1 | SOX17 |
| rs1812506 | 8 | 55676101 | A | G | 421 | 0.3456057 | 291 | 2.12258868 | 0.4466178 | 2.01E-06 | 5.06E-03 | 6.22E-01 | 2.07E-01 | RP1 | SOX17 |
| rs858397 | 8 | 55614690 | A | G | 421 | 0.33135392 | 278 | 2.2301348 | 0.44990341 | 7.16E-07 | 1.03E-02 | 1.00E+00 | 4.22E-01 | RP1 | SOX17 |
| rs382476 | 8 | 55590975 | G | A | 421 | 0.66627078 | 288 | -2.2481938 | 0.4540554 | 7.37E-07 | 1.07E-02 | 1.00E+00 | 4.39E-01 | RP1 | SOX17 |
| rs1595406 | 8 | 55630615 | A | G | 421 | 0.3456057 | 291 | 2.10018889 | 0.44830064 | 2.80E-06 | 1.27E-02 | 1.00E+00 | 5.21E-01 | RP1 | SOX17 |
| rs13278605 | 8 | 55688171 | C | T | 421 | 0.32897862 | 277 | 2.27377348 | 0.44593165 | 3.42E-07 | 1.37E-02 | 1.00E+00 | 5.62E-01 | RP1 | SOX17 |
| rs13276543 | 8 | 55688174 | G | T | 421 | 0.32897862 | 276 | 2.29263157 | 0.44823093 | 3.14E-07 | 1.37E-02 | 1.00E+00 | 5.62E-01 | RP1 | SOX17 |
| rs405226 | 8 | 55592336 | A | G | 421 | 0.66270784 | 291 | -2.1881305 | 0.45101874 | 1.23E-06 | 1.41E-02 | 1.00E+00 | 5.78E-01 | RP1 | SOX17 |
| rs1391462 | 8 | 55699781 | C | A | 421 | 0.39786223 | 334 | 1.9984766 | 0.43696027 | 4.79E-06 | 1.84E-02 | 1.00E+00 | 7.54E-01 | RP1 | SOX17 |
| rs1437782 | 8 | 55632762 | C | T | 421 | 0.33016627 | 278 | 2.2579585 | 0.45098781 | 5.54E-07 | 1.98E-02 | 1.00E+00 | 8.12E-01 | RP1 | SOX17 |
| rs2375219 | 8 | 55698295 | C | T | 421 | 0.39311164 | 330 | 2.05700167 | 0.44073069 | 3.05E-06 | 2.18E-02 | 1.00E+00 | 8.94E-01 | RP1 | SOX17 |
| rs16920698 | 8 | 55678434 | G | A | 421 | 0.33016627 | 277 | 2.30492695 | 0.44908177 | 2.86E-07 | 2.28E-02 | 1.00E+00 | 9.35E-01 | RP1 | SOX17 |
| rs4737676 | 8 | 55679546 | G | A | 421 | 0.33016627 | 277 | 2.30492057 | 0.44908192 | 2.86E-07 | 2.53E-02 | 1.00E+00 | 1.00E+00 | RP1 | SOX17 |
| rs1396896 | 8 | 55695310 | A | G | 421 | 0.39786223 | 334 | 1.99853327 | 0.43690452 | 4.78E-06 | 2.66E-02 | 1.00E+00 | 1.00E+00 | RP1 | SOX17 |
| rs9643828 | 8 | 55529073 | C | T | 421 | 0.67695962 | 279 | -2.4236294 | 0.46846911 | 2.30E-07 | 2.99E-02 | 1.00E+00 | 1.00E+00 | RP1 | SOX17 |
| rs1437781 | 8 | 55629852 | T | C | 421 | 0.33254157 | 280 | 2.23854673 | 0.44940766 | 6.32E-07 | 3.11E-02 | 1.00E+00 | 1.00E+00 | RP1 | SOX17 |
| rs12548593 | 8 | 55674617 | G | T | 421 | 0.33254157 | 279 | 2.28269961 | 0.44752402 | 3.38E-07 | 3.14E-02 | 1.00E+00 | 1.00E+00 | RP1 | SOX17 |
| rs2083123 | 8 | 55680318 | C | T | 421 | 0.33254157 | 279 | 2.27910794 | 0.44707491 | 3.44E-07 | 3.14E-02 | 1.00E+00 | 1.00E+00 | RP1 | SOX17 |
| rs720372 | 8 | 55628637 | G | A | 421 | 0.34679335 | 292 | 2.09872734 | 0.44886259 | 2.93E-06 | 3.58E-02 | 1.00E+00 | 1.00E+00 | RP1 | SOX17 |
| rs7843693 | 8 | 55692112 | G | A | 421 | 0.39786223 | 334 | 1.99856756 | 0.43695394 | 4.79E-06 | 3.58E-02 | 1.00E+00 | 1.00E+00 | RP1 | SOX17 |
| rs13277510 | 8 | 55674149 | G | A | 421 | 0.33016627 | 277 | 2.30494752 | 0.44909942 | 2.86E-07 | 3.62E-02 | 1.00E+00 | 1.00E+00 | RP1 | SOX17 |
| rs983248 | 8 | 55680792 | C | T | 421 | 0.33016627 | 277 | 2.30260428 | 0.44871461 | 2.87E-07 | 3.75E-02 | 1.00E+00 | 1.00E+00 | RP1 | SOX17 |
| rs432393 | 8 | 55580298 | C | T | 421 | 0.66270784 | 291 | -2.1875382 | 0.45096346 | 1.23E-06 | 4.10E-02 | 1.00E+00 | 1.00E+00 | RP1 | SOX17 |
| rs12024557 | 1 | 229812357 | A | C | 421 | 0.04275534 | 35 | 5.56930969 | 1.12195729 | 6.91E-07 | 4.44E-02 | 1.00E+00 | 1.00E+00 | URB2 | URB2 |
| rs16823323 | 3 | 153657202 | G | A | 421 | 0.01662708 | 14 | 9.48464065 | 1.71769795 | 3.36E-08 | 5.30E-02 | 1.00E+00 | 1.00E+00 | ARHGEF26 | ARHGEF26 |
| rs79182806 | 7 | 18433827 | T | C | 421 | 0.00831354 | 7 | 12.0972603 | 2.38956889 | 4.14E-07 | 5.30E-02 | 1.00E+00 | 1.00E+00 | HDAC9 | HDAC9 |
| rs369623 | 8 | 55571940 | A | C | 421 | 0.66627078 | 287 | -2.2359815 | 0.45415557 | 8.51E-07 | 6.33E-02 | 1.00E+00 | 1.00E+00 | RP1 | SOX17 |
| rs384127 | 8 | 55597489 | G | A | 421 | 0.66627078 | 288 | -2.2481559 | 0.45404811 | 7.37E-07 | 6.35E-02 | 1.00E+00 | 1.00E+00 | RP1 | SOX17 |
| rs184613584 | 17 | 48508221 | A | C | 421 | 0.00712589 | 6 | 14.1377974 | 2.8212234 | 5.41E-07 | 6.48E-02 | 1.00E+00 | 1.00E+00 | ACSF2 | ACSF2 |
| rs182959028 | 22 | 45823032 | T | C | 421 | 0.00831354 | 7 | 11.944461 | 2.53146 | 2.38E-06 | 6.48E-02 | 1.00E+00 | 1.00E+00 | RIBC2 | RIBC2 |
| rs433324 | 8 | 55564609 | A | G | 421 | 0.66627078 | 287 | -2.2483462 | 0.45814252 | 9.22E-07 | 6.83E-02 | 1.00E+00 | 1.00E+00 | RP1 | SOX17 |
| rs1301444047 | 7 | 36574504 | G | A | 421 | 0.04275534 | 43 | 5.12842209 | 1.08835549 | 2.45E-06 | 7.23E-02 | 1.00E+00 | 1.00E+00 | AOAH | AOAH |
| rs62447184 | 7 | 36574504 | G | A | 421 | 0.04275534 | 43 | 5.12842209 | 1.08835549 | 2.45E-06 | 7.23E-02 | 1.00E+00 | 1.00E+00 | AOAH | AOAH |
| rs541288561 | 19 | 18869445 | T | G | 421 | 0.00475059 | 5 | 15.6119012 | 3.13183483 | 6.20E-07 | 7.23E-02 | 1.00E+00 | 1.00E+00 | CRTC1 | CRTC1 |
| rs570407448 | 19 | 18880030 | G | A | 421 | 0.00475059 | 5 | 15.4977984 | 3.12412555 | 7.02E-07 | 7.23E-02 | 1.00E+00 | 1.00E+00 | CRTC1 | CRTC1 |
| rs423841 | 8 | 55556069 | G | A | 421 | 0.66270784 | 291 | -2.1495726 | 0.45846846 | 2.75E-06 | 7.39E-02 | 1.00E+00 | 1.00E+00 | RP1 | SOX17 |
| rs117913371 | 10 | 102918486 | G | A | 421 | 0.02494062 | 21 | 7.86609555 | 1.40785512 | 2.31E-08 | 7.46E-02 | 1.00E+00 | 1.00E+00 | LINC01514 | KAZALD1 |

| | | | | | | | | | | | | | | | |
|---|---|---|---|---|---|---|---|---|---|---|---|---|---|---|---|
| rs147601511 | 8 | 22322319 | A | G | 421 | 0.00475059 | 4 | 16.4471181 | 3.31777038 | 7.15E-07 | 7.96E-02 | 1.00E+00 | 1.00E+00 | PPP3CC | PPP3CC |
| rs1391463 | 8 | 55681876 | T | G | 421 | 0.33016627 | 277 | 2.30258865 | 0.44871307 | 2.87E-07 | 9.01E-02 | 1.00E+00 | 1.00E+00 | RP1 | SOX17 |
| rs78225611 | 7 | 18407464 | A | C | 421 | 0.00950119 | 8 | 11.0294387 | 2.26622466 | 1.13E-06 | 1.05E-01 | 1.00E+00 | 1.00E+00 | HDAC9 | HDAC9 |
| rs77300464 | 7 | 18408761 | A | G | 421 | 0.00712589 | 6 | 14.6659794 | 2.6169516 | 2.09E-08 | 1.10E-01 | 1.00E+00 | 1.00E+00 | HDAC9 | HDAC9 |
| rs4737674 | 8 | 55661654 | C | A | 421 | 0.33016627 | 277 | 2.30559554 | 0.44921988 | 2.86E-07 | 1.16E-01 | 1.00E+00 | 1.00E+00 | RP1 | SOX17 |
| rs11987234 | 8 | 55669829 | A | G | 421 | 0.32897862 | 276 | 2.29599259 | 0.44884532 | 3.13E-07 | 1.20E-01 | 1.00E+00 | 1.00E+00 | RP1 | SOX17 |
| rs17169602 | 7 | 18446741 | G | A | 421 | 0.00950119 | 8 | 12.7273717 | 2.25993984 | 1.78E-08 | 1.25E-01 | 1.00E+00 | 1.00E+00 | HDAC9 | HDAC9 |
| rs3098298 | 8 | 55582838 | C | T | 421 | 0.66270784 | 291 | -2.1875635 | 0.45094791 | 1.23E-06 | 1.25E-01 | 1.00E+00 | 1.00E+00 | RP1 | SOX17 |
| rs61434999 | 7 | 18418351 | A | G | 421 | 0.00831354 | 7 | 11.8411511 | 2.41284027 | 9.22E-07 | 1.28E-01 | 1.00E+00 | 1.00E+00 | HDAC9 | HDAC9 |
| rs77346868 | 7 | 18406599 | A | G | 421 | 0.00950119 | 8 | 11.0210129 | 2.26481172 | 1.14E-06 | 1.31E-01 | 1.00E+00 | 1.00E+00 | HDAC9 | HDAC9 |
| rs139493286 | 18 | 28816019 | G | A | 421 | 0.00356295 | 3 | 17.4603093 | 3.75076428 | 3.24E-06 | 1.33E-01 | 1.00E+00 | 1.00E+00 | DSG1 | DSG1 |
| rs1877768 | 6 | 16534923 | C | T | 421 | 0.01781473 | 15 | 8.28754685 | 1.76830748 | 2.78E-06 | 1.60E-01 | 1.00E+00 | 1.00E+00 | ATXN1 | ATXN1 |
| rs1686289 | 14 | 46260982 | G | A | 421 | 0.67814727 | 277 | -2.2551539 | 0.48625976 | 3.52E-06 | 1.63E-01 | 1.00E+00 | 1.00E+00 | LINC02303 | MIS18BP1 |
| rs150586237 | 6 | 24491348 | C | T | 421 | 0.00356295 | 3 | 22.0461443 | 3.78094793 | 5.51E-09 | 1.64E-01 | 1.00E+00 | 1.00E+00 | GPLD1 | GPLD1 |
| rs10105693 | 8 | 55640472 | C | T | 421 | 0.32897862 | 276 | 2.2928391 | 0.44965282 | 3.41E-07 | 1.68E-01 | 1.00E+00 | 1.00E+00 | RP1 | SOX17 |
| rs79539453 | 11 | 125266353 | C | T | 421 | 0.0023753 | 3 | 20.8220616 | 4.34542406 | 1.65E-06 | 1.73E-01 | 1.00E+00 | 1.00E+00 | PKNOX2 | PKNOX2 |
| rs10279777 | 7 | 18441589 | G | A | 421 | 0.00950119 | 8 | 12.8681335 | 2.27961694 | 1.65E-08 | 1.97E-01 | 1.00E+00 | 1.00E+00 | HDAC9 | HDAC9 |
| rs367179 | 8 | 55587616 | T | C | 421 | 0.66270784 | 291 | -2.1875635 | 0.45094791 | 1.23E-06 | 2.03E-01 | 1.00E+00 | 1.00E+00 | RP1 | SOX17 |
| rs384543 | 8 | 55591609 | G | A | 421 | 0.66627078 | 288 | -2.2481938 | 0.4540554 | 7.37E-07 | 2.18E-01 | 1.00E+00 | 1.00E+00 | RP1 | SOX17 |
| rs2385790 | 1 | 229807492 | C | T | 421 | 0.0415677 | 35 | 5.58224822 | 1.12394723 | 6.81E-07 | 2.21E-01 | 1.00E+00 | 1.00E+00 | URB2 | URB2 |
| rs78907958 | 7 | 18425017 | T | G | 421 | 0.00831354 | 7 | 12.0901322 | 2.40514961 | 4.99E-07 | 2.36E-01 | 1.00E+00 | 1.00E+00 | HDAC9 | HDAC9 |
| rs2274997 | 1 | 229804646 | A | G | 421 | 0.0415677 | 35 | 5.58731228 | 1.1238603 | 6.64E-07 | 2.43E-01 | 1.00E+00 | 1.00E+00 | URB2 | URB2 |
| rs1498183 | 8 | 55716905 | C | T | 421 | 0.39429929 | 332 | 2.0245746 | 0.44255141 | 4.77E-06 | 2.53E-01 | 1.00E+00 | 1.00E+00 | RP1 | SOX17 |
| rs10958428 | 8 | 55685641 | A | G | 421 | 0.33372922 | 280 | 2.23996885 | 0.44568673 | 5.01E-07 | 2.78E-01 | 1.00E+00 | 1.00E+00 | RP1 | SOX17 |
| rs80156375 | 7 | 18443215 | A | C | 421 | 0.00950119 | 8 | 11.5406255 | 2.25971308 | 3.27E-07 | 2.83E-01 | 1.00E+00 | 1.00E+00 | HDAC9 | HDAC9 |
| rs79213709 | 11 | 99093455 | G | A | 421 | 0.03206651 | 26 | 6.17081884 | 1.23453654 | 5.78E-07 | 2.83E-01 | 1.00E+00 | 1.00E+00 | CNTN5 | CNTN5 |
| rs12502861 | 4 | 2426305 | T | C | 421 | 0.01068884 | 9 | 11.1588472 | 2.36961887 | 2.49E-06 | 3.29E-01 | 1.00E+00 | 1.00E+00 | CFAP99 | CFAP99 |
| rs557092705 | 20 | 34189911 | C | T | 421 | 0.00356295 | 3 | 19.4769546 | 4.2480073 | 4.54E-06 | 3.29E-01 | 1.00E+00 | 1.00E+00 | FER1L4 | ERGIC3 |
| rs75773869 | 7 | 18410845 | G | T | 421 | 0.00831354 | 7 | 11.8642716 | 2.41902322 | 9.36E-07 | 3.31E-01 | 1.00E+00 | 1.00E+00 | HDAC9 | HDAC9 |
| rs74455595 | 7 | 18431784 | A | G | 421 | 0.00712589 | 6 | 14.8397312 | 2.5778114 | 8.58E-09 | 3.40E-01 | 1.00E+00 | 1.00E+00 | HDAC9 | HDAC9 |
| rs79602997 | 7 | 18410250 | G | A | 421 | 0.00831354 | 7 | 11.8608211 | 2.41901626 | 9.43E-07 | 3.50E-01 | 1.00E+00 | 1.00E+00 | HDAC9 | HDAC9 |
| rs75090694 | 7 | 18447436 | A | G | 421 | 0.00831354 | 7 | 13.8314005 | 2.41427081 | 1.01E-08 | 3.59E-01 | 1.00E+00 | 1.00E+00 | HDAC9 | HDAC9 |
| rs4737201 | 8 | 55691458 | C | T | 421 | 0.33016627 | 277 | 2.30197734 | 0.44869179 | 2.89E-07 | 3.65E-01 | 1.00E+00 | 1.00E+00 | RP1 | SOX17 |
| rs17746486 | 2 | 95722609 | C | T | 421 | 0.03206651 | 29 | 6.90851358 | 1.43477175 | 1.47E-06 | 3.80E-01 | 1.00E+00 | 1.00E+00 | MAL | MAL |
| rs118040657 | 10 | 3472846 | C | T | 421 | 0.00831354 | 8 | 12.1147712 | 2.64434694 | 4.62E-06 | 3.80E-01 | 1.00E+00 | 1.00E+00 | LOC10537636( | KLF6 |
| rs113651406 | 4 | 990967 | C | T | 421 | 0.00356295 | 3 | 19.4054026 | 3.7635826 | 2.52E-07 | 3.88E-01 | 1.00E+00 | 1.00E+00 | IDUA | IDUA |
| rs75689761 | 7 | 18406573 | C | T | 421 | 0.00831354 | 7 | 11.8482917 | 2.41320258 | 9.12E-07 | 3.91E-01 | 1.00E+00 | 1.00E+00 | HDAC9 | HDAC9 |
| rs147171192 | 4 | 89135588 | A | G | 421 | 0.00712589 | 5 | 13.3044047 | 2.89758492 | 4.40E-06 | 4.15E-01 | 1.00E+00 | 1.00E+00 | ABCG2 | ABCG2 |
| rs10486295 | 7 | 18446807 | G | A | 421 | 0.00950119 | 8 | 12.76229 | 2.26480456 | 1.75E-08 | 4.22E-01 | 1.00E+00 | 1.00E+00 | HDAC9 | HDAC9 |
| rs74683551 | 1 | 112354418 | G | A | 421 | 0.01662708 | 14 | 8.38252043 | 1.72705812 | 1.21E-06 | 4.23E-01 | 1.00E+00 | 1.00E+00 | KCND3 | KCND3 |
| rs147630370 | 4 | 87450675 | T | C | 421 | 0.00475059 | 4 | 16.0013633 | 3.23339786 | 7.47E-07 | 4.23E-01 | 1.00E+00 | 1.00E+00 | MAPK10 | MAPK10 |
| rs111391231 | 7 | 89497045 | T | C | 421 | 0.01425178 | 12 | 8.31717755 | 1.80720714 | 4.18E-06 | 4.23E-01 | 1.00E+00 | 1.00E+00 | STEAP2 | STEAP2 |
| rs112007361 | 11 | 99188380 | A | C | 421 | 0.03087886 | 26 | 6.35658468 | 1.22825758 | 2.28E-07 | 4.23E-01 | 1.00E+00 | 1.00E+00 | CNTN5 | CNTN5 |
| rs10494861 | 1 | 205331874 | G | A | 421 | 0.00356295 | 3 | 19.1820498 | 4.0729134 | 2.48E-06 | 4.27E-01 | 1.00E+00 | 1.00E+00 | KLHDC8A | KLHDC8A |
| rs142894171 | 2 | 151438271 | G | T | 421 | 0.00356295 | 3 | 18.0710394 | 3.74586836 | 1.41E-06 | 4.27E-01 | 1.00E+00 | 1.00E+00 | LINC02612 | RND3 |
| rs176786 | 14 | 46282970 | T | C | 421 | 0.32304038 | 268 | 2.28475641 | 0.49171936 | 3.38E-06 | 4.32E-01 | 1.00E+00 | 1.00E+00 | LINC02303 | MIS18BP1 |
| rs76327548 | 12 | 101182966 | G | A | 421 | 0.01306413 | 11 | 9.17842714 | 1.93235231 | 2.04E-06 | 4.39E-01 | 1.00E+00 | 1.00E+00 | ANO4 | ANO4 |
| rs12045643 | 1 | 229834050 | C | T | 421 | 0.04275534 | 36 | 5.48149152 | 1.10708948 | 7.37E-07 | 4.45E-01 | 1.00E+00 | 1.00E+00 | URB2 | URB2 |
| rs140782222 | 9 | 84028894 | T | C | 421 | 0.0023753 | 3 | 21.791526 | 4.31928461 | 4.53E-07 | 4.53E-01 | 1.00E+00 | 1.00E+00 | TLE1 | TLE1 |
| rs12266995 | 10 | 24852783 | T | C | 421 | 0.03087886 | 26 | 5.96782535 | 1.29311959 | 3.93E-06 | 4.91E-01 | 1.00E+00 | 1.00E+00 | ARHGAP21 | ARHGAP21 |
| rs176783 | 14 | 46280913 | A | G | 421 | 0.32185273 | 267 | 2.28010147 | 0.49376374 | 3.88E-06 | 5.10E-01 | 1.00E+00 | 1.00E+00 | LINC02303 | MIS18BP1 |
| rs77867199 | 7 | 18442275 | G | T | 421 | 0.01068884 | 9 | 10.8221282 | 2.13363365 | 3.93E-07 | 5.14E-01 | 1.00E+00 | 1.00E+00 | HDAC9 | HDAC9 |

| | | | | | | | | | | | | | | | |
|---|---|---|---|---|---|---|---|---|---|---|---|---|---|---|---|
| rs2891865 | 1 | 229806368 | A | G | 421 | 0.0415677 | 35 | 5.58181376 | 1.12378916 | 6.80E-07 | 5.18E-01 | 1.00E+00 | 1.00E+00 | URB2 | URB2 |
| rs16850124 | 1 | 229831331 | T | C | 421 | 0.04394299 | 37 | 5.18095662 | 1.09435188 | 2.20E-06 | 5.34E-01 | 1.00E+00 | 1.00E+00 | URB2 | URB2 |
| rs75606013 | 7 | 18414613 | G | A | 421 | 0.00712589 | 6 | 14.6617119 | 2.61692421 | 2.11E-08 | 5.47E-01 | 1.00E+00 | 1.00E+00 | HDAC9 | HDAC9 |
| rs290120 | 5 | 163268244 | T | G | 421 | 0.00712589 | 6 | 13.3587764 | 2.83260801 | 2.40E-06 | 5.69E-01 | 1.00E+00 | 1.00E+00 | MAT2B | MAT2B |
| rs73085348 | 3 | 42711221 | A | G | 421 | 0.01187648 | 11 | 10.139317 | 2.03504571 | 6.28E-07 | 5.76E-01 | 1.00E+00 | 1.00E+00 | STEAP2 | STEAP2 |
| rs111900874 | 7 | 89516669 | G | A | 421 | 0.01425178 | 12 | 8.42424013 | 1.82258334 | 3.80E-06 | 5.76E-01 | 1.00E+00 | 1.00E+00 | STEAP2 | STEAP2 |
| rs148556485 | 9 | 84023826 | A | C | 421 | 0.0023753 | 3 | 21.3800152 | 4.26618316 | 5.40E-07 | 5.76E-01 | 1.00E+00 | 1.00E+00 | TLE1 | TLE1 |
| rs180765647 | 13 | 114427311 | G | T | 421 | 0.0023753 | 3 | 20.1246521 | 4.37397806 | 4.20E-06 | 5.76E-01 | 1.00E+00 | 1.00E+00 | GRK1 | ATP4B |
| rs2327968 | 20 | 15813491 | C | T | 421 | 0.02494062 | 21 | 6.15899856 | 1.34390809 | 4.59E-06 | 5.83E-01 | 1.00E+00 | 1.00E+00 | MACROD2 | MACROD2 |
| rs2365739 | 1 | 62484462 | G | A | 421 | 0.02137767 | 18 | 6.86815917 | 1.43887996 | 1.81E-06 | 5.88E-01 | 1.00E+00 | 1.00E+00 | PATJ | PATJ |
| rs142993106 | 4 | 90957372 | G | A | 421 | 0.01781473 | 15 | 8.08463611 | 1.73306162 | 3.09E-06 | 5.88E-01 | 1.00E+00 | 1.00E+00 | CCSER1 | CCSER1 |
| rs180828621 | 10 | 124533409 | G | A | 421 | 0.00712589 | 6 | 15.1380574 | 2.79957627 | 6.40E-08 | 5.90E-01 | 1.00E+00 | 1.00E+00 | DMBT1L1 | CUZD1 |
| rs12315614 | 12 | 64920957 | C | A | 421 | 0.0760095 | 64 | 3.77710662 | 0.8201959 | 4.12E-06 | 6.20E-01 | 1.00E+00 | 1.00E+00 | TBK1 | TBK1 |
| rs4562666 | 1 | 229824770 | T | C | 421 | 0.04275534 | 36 | 5.51922599 | 1.11457457 | 7.35E-07 | 6.58E-01 | 1.00E+00 | 1.00E+00 | URB2 | URB2 |
| rs2876414 | 20 | 15813704 | G | T | 421 | 0.02256532 | 19 | 6.81910907 | 1.48530907 | 4.41E-06 | 6.60E-01 | 1.00E+00 | 1.00E+00 | MACROD2 | MACROD2 |
| rs12036586 | 1 | 229826378 | G | A | 421 | 0.04750594 | 40 | 5.22212176 | 1.07114093 | 1.09E-06 | 6.73E-01 | 1.00E+00 | 1.00E+00 | URB2 | URB2 |
| rs75334617 | 10 | 102956152 | G | A | 421 | 0.03800475 | 32 | 5.84603405 | 1.18027345 | 7.30E-07 | 7.59E-01 | 1.00E+00 | 1.00E+00 | LINC01514 | KAZALD1 |
| rs78547898 | 22 | 32824278 | G | A | 421 | 0.00356295 | 3 | 20.9098395 | 4.15538274 | 4.85E-07 | 7.68E-01 | 1.00E+00 | 1.00E+00 | BPIFC | FOX07 |
| rs796777817 | 22 | 32824278 | G | A | 421 | 0.00356295 | 3 | 20.9098395 | 4.15538274 | 4.85E-07 | 7.68E-01 | 1.00E+00 | 1.00E+00 | BPIFC | FOX07 |
| rs2274996 | 1 | 229804538 | C | T | 421 | 0.0415677 | 35 | 5.58527433 | 1.12406363 | 6.74E-07 | 7.87E-01 | 1.00E+00 | 1.00E+00 | URB2 | URB2 |
| rs143048774 | 14 | 46294660 | A | C | 421 | 0.31710214 | 265 | 2.28482299 | 0.49630858 | 4.15E-06 | 8.12E-01 | 1.00E+00 | 1.00E+00 | LINC02303 | MIS18BP1 |
| rs428110 | 14 | 46294660 | A | C | 421 | 0.31710214 | 265 | 2.28482299 | 0.49630858 | 4.15E-06 | 8.12E-01 | 1.00E+00 | 1.00E+00 | LINC02303 | MIS18BP1 |
| rs76526501 | 7 | 18431110 | G | A | 421 | 0.00712589 | 6 | 14.8397312 | 2.5778114 | 8.58E-09 | 8.23E-01 | 1.00E+00 | 1.00E+00 | HDAC9 | HDAC9 |
| rs146526206 | 4 | 90993018 | T | C | 421 | 0.01781473 | 15 | 8.09176656 | 1.69535286 | 1.82E-06 | 8.30E-01 | 1.00E+00 | 1.00E+00 | CCSER1 | CCSER1 |
| rs17017794 | 4 | 91825885 | T | C | 421 | 0.0391924 | 33 | 5.83240545 | 1.14450838 | 3.47E-07 | 9.15E-01 | 1.00E+00 | 1.00E+00 | CCSER1 | CCSER1 |
| rs147559909 | 2 | 237051523 | T | C | 421 | 0.00593824 | 5 | 17.0840975 | 3.03209195 | 1.76E-08 | 9.53E-01 | 1.00E+00 | 1.00E+00 | AGAP1 | AGAP1 |
| rs74521112 | 11 | 99089147 | G | T | 421 | 0.03206651 | 27 | 6.25632387 | 1.22787586 | 3.48E-07 | 9.53E-01 | 1.00E+00 | 1.00E+00 | CNTN5 | CNTN5 |
| rs115348382 | 1 | 9655903 | G | A | 421 | 0.00356295 | 3 | 18.2107495 | 3.84721698 | 2.21E-06 | NA | | | | |
| rs186532456 | 1 | 18948328 | C | T | 421 | 0.00356295 | 3 | 20.1730343 | 4.11737745 | 9.61E-07 | NA | | | | |
| rs562032622 | 1 | 18959333 | A | C | 421 | 0.00356295 | 3 | 21.0267115 | 4.3197503 | 1.13E-06 | NA | | | | |
| rs149493615 | 1 | 79880089 | G | A | 421 | 0.00831354 | 8 | 11.750632 | 2.43451244 | 1.39E-06 | NA | | | | |
| rs143811231 | 1 | 79968131 | T | C | 421 | 0.00831354 | 8 | 11.5747409 | 2.42066027 | 1.74E-06 | NA | | | | |
| rs34270375 | 1 | 89370702 | G | A | 421 | 0.02612827 | 21 | 7.71147017 | 1.57216792 | 9.34E-07 | NA | | | | |
| rs187518659 | 1 | 99455745 | T | G | 421 | 0.00950119 | 8 | 11.6819623 | 2.46564537 | 2.16E-06 | NA | | | | |
| rs140420703 | 1 | 102803484 | T | G | 421 | 0.00475059 | 5 | 16.487654 | 3.46213891 | 1.91E-06 | NA | | | | |
| rs563167766 | 1 | 102866365 | G | A | 421 | 0.0023753 | 3 | 19.415739 | 4.24240395 | 4.73E-06 | NA | | | | |
| rs77180278 | 1 | 102961882 | T | C | 421 | 0.02850356 | 24 | 7.33619996 | 1.37404112 | 9.34E-08 | NA | | | | |
| rs112351653 | 1 | 103220360 | T | C | 421 | 0.02850356 | 24 | 7.42386947 | 1.33286772 | 2.55E-08 | NA | | | | |
| rs180926150 | 1 | 103226326 | C | T | 421 | 0.0023753 | 3 | 19.641918 | 4.24793649 | 3.77E-06 | NA | | | | |
| rs114413507 | 1 | 103419168 | T | C | 421 | 0.02850356 | 24 | 7.42216029 | 1.33294954 | 2.57E-08 | NA | | | | |
| rs116672066 | 1 | 103472916 | G | A | 421 | 0.02731591 | 23 | 8.04769476 | 1.37187844 | 4.46E-09 | NA | | | | |
| rs111928960 | 1 | 103633635 | G | A | 421 | 0.02850356 | 22 | 8.60257208 | 1.45827516 | 3.65E-09 | NA | | | | |
| rs113221952 | 1 | 103753974 | A | G | 421 | 0.02137767 | 19 | 7.92350704 | 1.61421682 | 9.17E-07 | NA | | | | |
| rs1856085 | 1 | 104114545 | G | A | 421 | 0.0023753 | 3 | 21.51144 | 4.26747898 | 4.64E-07 | NA | | | | |
| rs143597860 | 1 | 104157143 | A | G | 421 | 0.0023753 | 3 | 21.5748154 | 4.2760379 | 4.52E-07 | NA | | | | |
| rs144541665 | 1 | 104310729 | G | A | 421 | 0.0023753 | 3 | 21.8734611 | 4.30339839 | 3.72E-07 | NA | | | | |
| rs76098744 | 1 | 112349372 | C | T | 421 | 0.01662708 | 14 | 8.38612452 | 1.72704116 | 1.20E-06 | NA | | | | |
| rs76617932 | 1 | 180930424 | T | C | 421 | 0.01187648 | 9 | 10.5449608 | 2.25040726 | 2.79E-06 | NA | | | | |
| rs183180157 | 1 | 181247121 | A | C | 421 | 0.00950119 | 7 | 12.61386 | 2.68921121 | 2.72E-06 | NA | | | | |
| rs375790303 | 1 | 184530482 | G | A | 421 | 0.00831354 | 7 | 11.4240393 | 2.49719192 | 4.77E-06 | NA | | | | |
| rs138480898 | 1 | 184955657 | C | T | 421 | 0.00356295 | 3 | 16.9873974 | 3.66972373 | 3.67E-06 | NA | | | | |
| rs145766563 | 1 | 185129502 | G | A | 421 | 0.00356295 | 4 | 16.8682102 | 3.54127008 | 1.90E-06 | NA | | | | |

| | | | | | | | | | | | |
|---|---|---|---|---|---|---|---|---|---|---|---|
| rs147032554 | 1 | 186148864 | T | G | 421 | 0.00356295 | 3 | 18.162609 | 3.65402821 | 6.68E-07 | NA |
| rs180989936 | 1 | 193044178 | A | G | 421 | 0.00475059 | 4 | 17.1793579 | 3.70215494 | 3.48E-06 | NA |
| rs559559983 | 1 | 246977132 | C | A | 421 | 0.00712589 | 5 | 13.811132 | 2.89725588 | 1.87E-06 | NA |
| rs550763536 | 2 | 7452347 | T | G | 421 | 0.00593824 | 5 | 14.4871647 | 3.10349768 | 3.04E-06 | NA |
| rs558553658 | 2 | 15682724 | C | T | 421 | 0.00475059 | 3 | 17.1099107 | 3.29108061 | 2.01E-07 | NA |
| rs536781978 | 2 | 29733681 | A | G | 421 | 0.00356295 | 3 | 17.5870824 | 3.70259451 | 2.03E-06 | NA |
| rs568321148 | 2 | 29870348 | T | G | 421 | 0.00356295 | 3 | 17.7125046 | 3.7201251 | 1.92E-06 | NA |
| rs76777840 | 2 | 48312950 | G | A | 421 | 0.00593824 | 5 | 14.3500784 | 2.8927894 | 7.03E-07 | NA |
| rs145080832 | 2 | 48473143 | G | A | 421 | 0.00475059 | 4 | 17.1618477 | 3.25709537 | 1.37E-07 | NA |
| rs184220112 | 2 | 48631743 | C | A | 421 | 0.00475059 | 4 | 16.4879043 | 3.22267606 | 3.12E-07 | NA |
| rs189890455 | 2 | 48655397 | C | T | 421 | 0.00593824 | 5 | 15.1467391 | 2.99355083 | 4.20E-07 | NA |
| rs181193202 | 2 | 52527354 | T | C | 421 | 0.00356295 | 3 | 19.6312088 | 3.74776015 | 1.62E-07 | NA |
| rs183378658 | 2 | 52563371 | C | T | 421 | 0.00356295 | 3 | 19.7446169 | 3.72634921 | 1.17E-07 | NA |
| rs190193113 | 2 | 53085778 | G | A | 421 | 0.00356295 | 3 | 19.9533602 | 3.7619559 | 1.13E-07 | NA |
| rs187520610 | 2 | 53360041 | G | A | 421 | 0.00475059 | 4 | 18.8010396 | 3.19364595 | 3.93E-09 | NA |
| rs149421869 | 2 | 53483429 | G | T | 421 | 0.00831354 | 8 | 11.7382186 | 2.47204376 | 2.05E-06 | NA |
| rs146479102 | 2 | 65825759 | G | A | 421 | 0.00593824 | 5 | 15.4846768 | 3.26980199 | 2.18E-06 | NA |
| rs528288879 | 2 | 65875930 | C | T | 421 | 0.00593824 | 6 | 14.7878085 | 2.95121929 | 5.42E-07 | NA |
| rs11690187 | 2 | 67565909 | A | C | 421 | 0.00593824 | 5 | 13.6310532 | 2.9751322 | 4.61E-06 | NA |
| rs151272830 | 2 | 67682724 | G | T | 421 | 0.00712589 | 6 | 13.3323479 | 2.84806076 | 2.85E-06 | NA |
| rs186142189 | 2 | 67702707 | G | A | 421 | 0.00712589 | 6 | 12.7361927 | 2.71405368 | 2.70E-06 | NA |
| rs184200893 | 2 | 69260913 | C | T | 421 | 0.00356295 | 3 | 19.9580356 | 4.21905487 | 2.24E-06 | NA |
| rs111927235 | 2 | 74483954 | A | G | 421 | 0.00475059 | 4 | 17.0468955 | 3.46111889 | 8.43E-07 | NA |
| rs111838310 | 2 | 74673491 | C | A | 421 | 0.00475059 | 5 | 17.7963938 | 3.48429066 | 3.26E-07 | NA |
| rs112983626 | 2 | 74697150 | G | A | 421 | 0.00475059 | 4 | 17.7848079 | 3.48532494 | 3.35E-07 | NA |
| rs113006316 | 2 | 74802360 | A | G | 421 | 0.0023753 | 3 | 22.8886792 | 4.92411335 | 3.35E-06 | NA |
| rs76554191 | 2 | 95967628 | G | A | 421 | 0.04038005 | 34 | 5.72148039 | 1.2389174 | 3.87E-06 | NA |
| rs140352232 | 2 | 108038112 | G | A | 421 | 0.0023753 | 3 | 21.0576511 | 4.56013129 | 3.88E-06 | NA |
| rs116189766 | 2 | 126393864 | T | C | 421 | 0.0023753 | 3 | 21.190764 | 4.36037289 | 1.17E-06 | NA |
| rs139877408 | 2 | 129606273 | A | G | 421 | 0.0023753 | 3 | 20.3067129 | 4.43052497 | 4.58E-06 | NA |
| rs541508507 | 2 | 170023265 | G | A | 421 | 0.00475059 | 4 | 15.4630033 | 3.27389618 | 2.32E-06 | NA |
| rs142549310 | 2 | 170030506 | C | T | 421 | 0.00475059 | 4 | 15.2501713 | 3.24983634 | 2.70E-06 | NA |
| rs556293455 | 2 | 176950420 | G | A | 421 | 0.00356295 | 3 | 14.3860463 | 2.96998402 | 1.27E-06 | NA |
| rs184098071 | 2 | 177116420 | G | A | 421 | 0.00356295 | 3 | 13.947724 | 2.86782452 | 1.15E-06 | NA |
| rs532416695 | 2 | 177486790 | G | A | 421 | 0.00356295 | 3 | 19.8726656 | 3.76737683 | 1.33E-07 | NA |
| rs112557251 | 2 | 188375400 | T | C | 421 | 0.0023753 | 3 | 20.6510064 | 4.44850106 | 3.45E-06 | NA |
| rs185158855 | 2 | 223650026 | C | A | 421 | 0.00475059 | 4 | 16.6678864 | 3.60266524 | 3.72E-06 | NA |
| rs185510569 | 2 | 223814861 | G | A | 421 | 0.00356295 | 3 | 19.5886906 | 3.60490977 | 5.51E-08 | NA |
| rs181217257 | 2 | 239989119 | C | T | 421 | 0.00475059 | 4 | 21.5677368 | 3.47296989 | 5.29E-10 | NA |
| rs188076929 | 2 | 239993719 | T | C | 421 | 0.00475059 | 4 | 19.9775614 | 3.27246746 | 1.03E-09 | NA |
| rs112475378 | 3 | 1626661 | T | C | 421 | 0.01662708 | 14 | 9.55085929 | 1.97628138 | 1.35E-06 | NA |
| rs145676540 | 3 | 2044635 | C | T | 421 | 0.00593824 | 5 | 13.980658 | 2.88447377 | 1.25E-06 | NA |
| rs146007933 | 3 | 28227423 | T | C | 421 | 0.02137767 | 18 | 7.51149453 | 1.55442805 | 1.35E-06 | NA |
| rs73057656 | 3 | 33940571 | A | G | 421 | 0.0368171 | 33 | 5.9238329 | 1.27589711 | 3.44E-06 | NA |
| rs142684595 | 3 | 55319301 | T | C | 421 | 0.00593824 | 5 | 16.7729431 | 3.11364755 | 7.17E-08 | NA |
| rs80203220 | 3 | 122722331 | C | T | 421 | 0.00593824 | 6 | 13.426042 | 2.84218656 | 2.31E-06 | NA |
| rs192443987 | 3 | 135532345 | G | A | 421 | 0.00475059 | 4 | 17.7540285 | 3.67564302 | 1.36E-06 | NA |
| rs148248743 | 3 | 136134595 | C | T | 421 | 0.0023753 | 3 | 22.839379 | 4.62044689 | 7.69E-07 | NA |
| rs576124203 | 3 | 141841196 | T | G | 421 | 0.00356295 | 3 | 19.4039925 | 3.84392582 | 4.47E-07 | NA |
| rs545552231 | 3 | 141864350 | C | T | 421 | 0.00356295 | 3 | 19.8641332 | 3.94582563 | 4.80E-07 | NA |
| rs188720948 | 3 | 150069880 | T | C | 421 | 0.00356295 | 3 | 17.7032653 | 3.59722874 | 8.59E-07 | NA |
| rs139943877 | 3 | 155451289 | G | A | 421 | 0.00712589 | 7 | 13.0535902 | 2.76041594 | 2.26E-06 | NA |

| | | | | | | | | | | | |
|---|---|---|---|---|---|---|---|---|---|---|---|
| rs187047882 | 3 | 164285621 | G | A | 421 | 0.00356295 | 3 | 18.5641361 | 3.99725417 | 3.41E-06 | NA |
| rs141169929 | 3 | 164808462 | A | G | 421 | 0.00475059 | 5 | 14.8454178 | 3.1738839 | 2.91E-06 | NA |
| rs186649043 | 3 | 174932293 | C | T | 421 | 0.00356295 | 3 | 19.2134209 | 4.06588786 | 2.30E-06 | NA |
| rs189709453 | 3 | 177574308 | G | A | 421 | 0.00356295 | 3 | 21.6581488 | 4.05916142 | 9.52E-08 | NA |
| rs186767531 | 3 | 177639777 | T | C | 421 | 0.00356295 | 3 | 21.2013243 | 4.04780758 | 1.63E-07 | NA |
| rs182868205 | 3 | 177712448 | C | T | 421 | 0.00356295 | 3 | 20.9548768 | 3.97841839 | 1.39E-07 | NA |
| rs191792521 | 3 | 195646605 | G | A | 421 | 0.00831354 | 6 | 13.9515484 | 2.96443599 | 2.52E-06 | NA |
| rs189765693 | 4 | 4324793 | T | C | 421 | 0.00475059 | 4 | 15.5191341 | 3.29924607 | 2.55E-06 | NA |
| rs183962155 | 4 | 21259643 | A | C | 421 | 0.00593824 | 6 | 13.7646008 | 2.97247759 | 3.64E-06 | NA |
| rs113751774 | 4 | 23428854 | C | T | 421 | 0.00712589 | 7 | 14.9103274 | 2.96657459 | 5.01E-07 | NA |
| rs113063005 | 4 | 23509067 | T | C | 421 | 0.00593824 | 6 | 21.2073894 | 3.15520096 | 1.80E-11 | NA |
| rs145875128 | 4 | 32073136 | G | A | 421 | 0.00356295 | 3 | 24.479986 | 4.60639213 | 1.07E-07 | NA |
| rs143287889 | 4 | 35572280 | C | T | 421 | 0.0023753 | 3 | 19.96741 | 4.34406 | 4.30E-06 | NA |
| rs77141817 | 4 | 37053759 | T | C | 421 | 0.00356295 | 3 | 20.6110292 | 3.64214866 | 1.52E-08 | NA |
| rs190822761 | 4 | 37099356 | G | T | 421 | 0.00356295 | 3 | 20.5846354 | 3.64346559 | 1.61E-08 | NA |
| rs999769259 | 4 | 62511965 | G | A | 421 | 0.00356295 | 3 | 22.1246725 | 4.77597881 | 3.61E-06 | NA |
| rs145116559 | 4 | 112677596 | T | C | 421 | 0.00356295 | 4 | 18.2441224 | 3.9932536 | 4.91E-06 | NA |
| rs181415102 | 4 | 112689266 | T | C | 421 | 0.00356295 | 4 | 18.2631019 | 3.99387944 | 4.81E-06 | NA |
| rs191423619 | 4 | 126516627 | G | T | 421 | 0.0023753 | 3 | 21.694611 | 4.20667337 | 2.51E-07 | NA |
| rs149298750 | 4 | 127158701 | A | C | 421 | 0.00356295 | 3 | 22.184052 | 4.26573246 | 1.99E-07 | NA |
| rs112679237 | 4 | 139140860 | T | C | 421 | 0.01781473 | 18 | 8.71202752 | 1.67680671 | 2.04E-07 | NA |
| rs531769270 | 4 | 154411043 | T | C | 421 | 0.00356295 | 3 | 16.5945338 | 3.51936092 | 2.41E-06 | NA |
| rs116651654 | 4 | 163238743 | C | T | 421 | 0.00712589 | 6 | 15.6223694 | 3.06624451 | 3.49E-07 | NA |
| rs567982164 | 5 | 25631297 | G | A | 421 | 0.0023753 | 3 | 21.8356286 | 4.36706703 | 5.73E-07 | NA |
| rs191986449 | 5 | 25745687 | C | T | 421 | 0.00475059 | 3 | 18.4978793 | 4.03679831 | 4.60E-06 | NA |
| rs185771987 | 5 | 73285489 | T | C | 421 | 0.00356295 | 3 | 18.2344824 | 3.97675014 | 4.53E-06 | NA |
| rs139360368 | 5 | 73372109 | A | C | 421 | 0.00356295 | 4 | 18.3590487 | 3.78701272 | 1.25E-06 | NA |
| rs181933850 | 5 | 91465647 | A | G | 421 | 0.00831354 | 7 | 11.0436586 | 2.37544216 | 3.33E-06 | NA |
| rs190190051 | 5 | 91475485 | G | A | 421 | 0.00831354 | 7 | 11.0589887 | 2.38317975 | 3.48E-06 | NA |
| rs182531466 | 5 | 91530073 | C | A | 421 | 0.00712589 | 6 | 12.6804978 | 2.75685261 | 4.23E-06 | NA |
| rs187236873 | 5 | 91530447 | G | A | 421 | 0.00712589 | 6 | 12.44907 | 2.61780937 | 1.98E-06 | NA |
| rs183816745 | 5 | 91631888 | A | G | 421 | 0.00593824 | 5 | 17.3487909 | 3.20195556 | 6.02E-08 | NA |
| rs111407636 | 5 | 95080029 | C | T | 421 | 0.00356295 | 3 | 18.642774 | 3.74168052 | 6.28E-07 | NA |
| rs111676272 | 5 | 95094298 | C | A | 421 | 0.00356295 | 3 | 18.6068625 | 3.74429748 | 6.72E-07 | NA |
| rs75848314 | 5 | 95098340 | T | C | 421 | 0.00356295 | 3 | 20.0998481 | 3.78700451 | 1.11E-07 | NA |
| rs111846247 | 5 | 95111643 | T | C | 421 | 0.00356295 | 3 | 20.4865772 | 3.85467205 | 1.07E-07 | NA |
| rs137873790 | 5 | 97087041 | A | G | 421 | 0.01306413 | 11 | 9.22240881 | 2.01095379 | 4.52E-06 | NA |
| rs189912648 | 5 | 134765439 | C | T | 421 | 0.00356295 | 4 | 16.9732599 | 3.60846227 | 2.55E-06 | NA |
| rs191006910 | 5 | 154216009 | G | A | 421 | 0.00356295 | 3 | 17.6173803 | 3.79049689 | 3.36E-06 | NA |
| rs74343174 | 5 | 161493182 | C | A | 421 | 0.00593824 | 5 | 14.7671824 | 3.13913408 | 2.55E-06 | NA |
| rs775626702 | 5 | 162630067 | A | C | 421 | 0.00356295 | 3 | 18.9484137 | 4.07023372 | 3.23E-06 | NA |
| rs371245624 | 5 | 162880956 | T | C | 421 | 0.00356295 | 3 | 19.4795931 | 4.06572011 | 1.66E-06 | NA |
| rs545428520 | 5 | 167821266 | T | C | 421 | 0.00356295 | 4 | 21.566035 | 3.66407988 | 3.96E-09 | NA |
| rs528404963 | 5 | 167852025 | T | C | 421 | 0.00356295 | 4 | 21.1393977 | 3.65303517 | 7.17E-09 | NA |
| rs72832764 | 5 | 170004673 | G | A | 421 | 0.00356295 | 3 | 18.2159757 | 3.88196743 | 2.70E-06 | NA |
| rs72837643 | 5 | 170188955 | T | C | 421 | 0.00356295 | 4 | 16.9302874 | 3.67743479 | 4.15E-06 | NA |
| rs142311947 | 5 | 177384469 | G | A | 421 | 0.00712589 | 8 | 12.0339013 | 2.57618116 | 2.99E-06 | NA |
| rs151015676 | 5 | 177390937 | T | G | 421 | 0.0023753 | 3 | 20.1421367 | 4.37074482 | 4.06E-06 | NA |
| rs571986619 | 6 | 85678832 | A | G | 421 | 0.0023753 | 3 | 20.7858067 | 4.34301983 | 1.70E-06 | NA |
| rs56224400 | 6 | 98092675 | T | C | 421 | 0.01662708 | 14 | 8.4433064 | 1.72251471 | 9.50E-07 | NA |
| rs147627638 | 6 | 99173116 | A | G | 421 | 0.00712589 | 6 | 13.0785306 | 2.79364254 | 2.85E-06 | NA |
| rs141326851 | 6 | 134833127 | A | C | 421 | 0.01662708 | 14 | 8.57849688 | 1.81053268 | 2.16E-06 | NA |

| | | | | | | | | | | | |
|---|---|---|---|---|---|---|---|---|---|---|---|
| rs146048121 | 6 | 142198955 | G | A | 421 | 0.0023753 | 3 | 20.0214379 | 4.30086025 | 3.24E-06 | NA |
| rs142106992 | 6 | 142269885 | C | A | 421 | 0.00356295 | 4 | 22.8345886 | 3.6758699 | 5.23E-10 | NA |
| rs72983831 | 6 | 142307100 | T | G | 421 | 0.00593824 | 6 | 15.6733239 | 3.17294911 | 7.83E-07 | NA |
| rs72986533 | 6 | 142611258 | T | C | 421 | 0.00831354 | 8 | 13.2812726 | 2.53063629 | 1.54E-07 | NA |
| rs73586304 | 6 | 142839425 | C | T | 421 | 0.00356295 | 3 | 18.2405116 | 3.99428202 | 4.96E-06 | NA |
| rs148532212 | 6 | 165499857 | T | C | 421 | 0.00356295 | 3 | 19.3535457 | 3.70945693 | 1.81E-07 | NA |
| rs117498042 | 6 | 165514281 | C | T | 421 | 0.00356295 | 3 | 19.335471 | 3.70913521 | 1.86E-07 | NA |
| rs148153037 | 6 | 167501386 | G | A | 421 | 0.00831354 | 8 | 14.3359246 | 2.42128102 | 3.20E-09 | NA |
| rs184487573 | 6 | 167513471 | A | G | 421 | 0.00712589 | 7 | 13.5009635 | 2.59268014 | 1.92E-07 | NA |
| rs17776100 | 7 | 6426479 | G | A | 421 | 0.02969121 | 25 | 6.63697397 | 1.32177185 | 5.13E-07 | NA |
| rs187978759 | 7 | 11711845 | G | A | 421 | 0.00356295 | 3 | 20.4555522 | 4.3637719 | 2.76E-06 | NA |
| rs117166500 | 7 | 17052778 | G | T | 421 | 0.00712589 | 7 | 12.5146865 | 2.62694198 | 1.90E-06 | NA |
| rs55844051 | 7 | 23360363 | T | C | 421 | 0.00356295 | 3 | 18.3875896 | 3.86268704 | 1.93E-06 | NA |
| rs574076561 | 7 | 49544747 | A | G | 421 | 0.00356295 | 3 | 19.6434766 | 4.25225723 | 3.85E-06 | NA |
| rs181259864 | 7 | 97488823 | C | A | 421 | 0.00356295 | 3 | 17.1360821 | 3.68242789 | 3.26E-06 | NA |
| rs192750513 | 7 | 97577830 | A | G | 421 | 0.00356295 | 3 | 17.1892469 | 3.72514218 | 3.94E-06 | NA |
| rs539713344 | 7 | 100474786 | G | A | 421 | 0.0023753 | 3 | 20.044756 | 4.21957545 | 2.03E-06 | NA |
| rs188028357 | 7 | 100668425 | C | T | 421 | 0.0023753 | 3 | 21.8425578 | 4.35791809 | 5.38E-07 | NA |
| rs536023430 | 7 | 146865072 | T | C | 421 | 0.00356295 | 3 | 19.4038683 | 4.13084856 | 2.64E-06 | NA |
| rs187384541 | 8 | 1632241 | A | G | 421 | 0.03562945 | 26 | 6.88745117 | 1.47776668 | 3.15E-06 | NA |
| rs575473987 | 8 | 5577494 | C | T | 421 | 0.00356295 | 3 | 22.2248653 | 3.88625754 | 1.07E-08 | NA |
| rs139062456 | 8 | 13251991 | C | T | 421 | 0.01781473 | 15 | 7.83515809 | 1.69687423 | 3.89E-06 | NA |
| rs185874707 | 8 | 18030674 | C | T | 421 | 0.01187648 | 10 | 9.13934765 | 1.94918644 | 2.75E-06 | NA |
| rs140797780 | 8 | 22087792 | C | T | 421 | 0.00356295 | 3 | 19.2908396 | 3.69147785 | 1.73E-07 | NA |
| rs188415494 | 8 | 25613298 | C | T | 421 | 0.00356295 | 3 | 18.5169478 | 3.9914865 | 3.50E-06 | NA |
| rs117816016 | 8 | 103751262 | C | T | 421 | 0.00356295 | 3 | 19.8807023 | 4.26960212 | 3.22E-06 | NA |
| rs567383525 | 8 | 115029494 | C | T | 421 | 0.00356295 | 3 | 17.7455227 | 3.71970627 | 1.84E-06 | NA |
| rs545550279 | 8 | 115552429 | G | T | 421 | 0.00356295 | 3 | 19.1812421 | 4.19212988 | 4.75E-06 | NA |
| rs536803366 | 8 | 123001411 | T | C | 421 | 0.0023753 | 3 | 19.2098639 | 4.00952057 | 1.66E-06 | NA |
| rs555249476 | 8 | 123001411 | T | C | 421 | 0.0023753 | 3 | 19.2098639 | 4.00952057 | 1.66E-06 | NA |
| rs532513136 | 8 | 135813748 | C | A | 421 | 0.00356295 | 3 | 19.9490594 | 4.03118599 | 7.47E-07 | NA |
| rs532730683 | 9 | 1784492 | G | T | 421 | 0.00356295 | 3 | 18.726838 | 3.8314675 | 1.02E-06 | NA |
| rs540065886 | 9 | 2770228 | T | C | 421 | 0.00356295 | 3 | 19.9489151 | 4.18802805 | 1.90E-06 | NA |
| rs543844012 | 9 | 30107023 | C | T | 421 | 0.00356295 | 3 | 18.1160485 | 3.9446103 | 4.38E-06 | NA |
| rs188034471 | 9 | 85937714 | G | A | 421 | 0.00475059 | 4 | 14.7559999 | 3.14925113 | 2.79E-06 | NA |
| rs190294315 | 9 | 85945465 | C | T | 421 | 0.00593824 | 5 | 14.5664276 | 2.88126649 | 4.29E-07 | NA |
| rs545690161 | 9 | 93030699 | G | A | 421 | 0.00593824 | 6 | 17.1436874 | 3.00318779 | 1.14E-08 | NA |
| rs565682685 | 9 | 93222328 | T | C | 421 | 0.00475059 | 4 | 18.938397 | 3.7206174 | 3.58E-07 | NA |
| rs183737367 | 9 | 93330047 | T | C | 421 | 0.00356295 | 3 | 23.3452306 | 4.24471431 | 3.80E-08 | NA |
| rs187213609 | 9 | 93415465 | C | T | 421 | 0.00356295 | 3 | 23.1975304 | 4.27412656 | 5.72E-08 | NA |
| rs150027952 | 9 | 103385416 | A | G | 421 | 0.0023753 | 3 | 21.8616648 | 4.51464643 | 1.28E-06 | NA |
| rs146207930 | 9 | 129042336 | A | G | 421 | 0.00712589 | 6 | 11.8624321 | 2.51502465 | 2.40E-06 | NA |
| rs78296164 | 9 | 135266715 | C | T | 421 | 0.00831354 | 7 | 11.6095338 | 2.33917557 | 6.94E-07 | NA |
| rs77871739 | 9 | 138552309 | G | A | 421 | 0.00475059 | 4 | 15.0724603 | 3.23606181 | 3.20E-06 | NA |
| rs184425183 | 10 | 13457520 | A | G | 421 | 0.00356295 | 3 | 22.6683174 | 3.77549648 | 1.92E-09 | NA |
| rs184458518 | 10 | 13471195 | T | G | 421 | 0.00356295 | 3 | 22.8015789 | 3.77123942 | 1.48E-09 | NA |
| rs117998251 | 10 | 13497976 | C | T | 421 | 0.00356295 | 3 | 22.9543311 | 3.77480488 | 1.19E-09 | NA |
| rs117025967 | 10 | 20207052 | C | A | 421 | 0.01306413 | 11 | 9.66973563 | 1.927439 | 5.25E-07 | NA |
| rs529011661 | 10 | 20372316 | G | A | 421 | 0.00475059 | 4 | 15.6393469 | 3.3125339 | 2.34E-06 | NA |
| rs138249376 | 10 | 63515829 | T | G | 421 | 0.00475059 | 4 | 16.4300711 | 3.59466791 | 4.86E-06 | NA |
| rs140277951 | 10 | 82359100 | G | A | 421 | 0.00831354 | 6 | 13.7463158 | 2.64333932 | 1.99E-07 | NA |
| rs566018180 | 10 | 86755052 | C | T | 421 | 0.00356295 | 3 | 18.563552 | 4.04215862 | 4.38E-06 | NA |

| | | | | | | | | | | | |
|---|---|---|---|---|---|---|---|---|---|---|---|
| rs140706881 | 10 | 96217535 | G | A | 421 | 0.00475059 | 4 | 18.974805 | 3.93288931 | 1.40E-06 | NA |
| rs752259256 | 10 | 104925319 | T | C | 421 | 0.00356295 | 3 | 20.4258639 | 4.45411591 | 4.52E-06 | NA |
| rs147393020 | 10 | 124779274 | A | G | 421 | 0.00593824 | 5 | 14.1853505 | 2.95980896 | 1.65E-06 | NA |
| rs193093906 | 10 | 126705489 | G | A | 421 | 0.00950119 | 9 | 10.5219537 | 2.25840517 | 3.18E-06 | NA |
| rs151115079 | 11 | 18655741 | T | C | 421 | 0.00475059 | 5 | 17.8273587 | 3.2680205 | 4.89E-08 | NA |
| rs138414342 | 11 | 18679398 | G | A | 421 | 0.00475059 | 5 | 18.1582505 | 3.28214943 | 3.16E-08 | NA |
| rs541653703 | 11 | 18701786 | G | A | 421 | 0.00475059 | 4 | 18.3169499 | 3.27243418 | 2.18E-08 | NA |
| rs118093638 | 11 | 18718324 | C | T | 421 | 0.00593824 | 5 | 14.2657259 | 2.91296506 | 9.72E-07 | NA |
| rs181812512 | 11 | 66665729 | C | T | 421 | 0.00356295 | 3 | 19.5093168 | 4.24219816 | 4.25E-06 | NA |
| rs529345909 | 11 | 67110852 | A | G | 421 | 0.00356295 | 3 | 18.70153 | 4.09206035 | 4.87E-06 | NA |
| rs544042801 | 11 | 68460243 | G | A | 421 | 0.00475059 | 4 | 15.8869866 | 3.40519019 | 3.08E-06 | NA |
| rs149949098 | 11 | 95099866 | G | A | 421 | 0.01662708 | 13 | 8.55927625 | 1.81883308 | 2.53E-06 | NA |
| rs148781275 | 11 | 103640603 | A | G | 421 | 0.00475059 | 5 | 15.7111853 | 3.32183832 | 2.25E-06 | NA |
| rs141281289 | 11 | 123693024 | A | G | 421 | 0.00593824 | 6 | 15.811518 | 3.0889522 | 3.08E-07 | NA |
| rs528140343 | 11 | 125719227 | A | C | 421 | 0.00356295 | 3 | 18.2835521 | 3.72738029 | 9.33E-07 | NA |
| rs546409459 | 11 | 125754989 | A | G | 421 | 0.00356295 | 3 | 18.9195474 | 4.00286836 | 2.28E-06 | NA |
| rs528609331 | 11 | 125842195 | C | T | 421 | 0.00356295 | 3 | 19.5812418 | 3.94832705 | 7.07E-07 | NA |
| rs7104959 | 11 | 129846126 | C | T | 421 | 0.00356295 | 3 | 17.9204611 | 3.84366851 | 3.13E-06 | NA |
| rs189360484 | 12 | 1870510 | A | G | 421 | 0.00475059 | 4 | 16.8078794 | 3.31424855 | 3.95E-07 | NA |
| rs141754456 | 12 | 20151132 | T | C | 421 | 0.00712589 | 7 | 12.1460333 | 2.32842921 | 1.82E-07 | NA |
| rs118184666 | 12 | 20424749 | G | A | 421 | 0.00712589 | 6 | 10.9509803 | 2.32477242 | 2.47E-06 | NA |
| rs549931083 | 12 | 20516286 | A | C | 421 | 0.00356295 | 3 | 14.3282694 | 2.79891395 | 3.07E-07 | NA |
| rs151323346 | 12 | 21012024 | T | C | 421 | 0.00593824 | 4 | 15.8903888 | 3.38034868 | 2.59E-06 | NA |
| rs371879555 | 12 | 23015962 | T | C | 421 | 0.00356295 | 3 | 20.1033765 | 4.20002323 | 1.70E-06 | NA |
| rs183466664 | 12 | 26821687 | A | G | 421 | 0.00475059 | 4 | 16.0181882 | 3.45609016 | 3.57E-06 | NA |
| rs77353774 | 12 | 28248852 | G | A | 421 | 0.00712589 | 6 | 13.1054362 | 2.67954103 | 1.00E-06 | NA |
| rs113167689 | 12 | 28435962 | C | T | 421 | 0.00712589 | 6 | 14.1364135 | 2.73335629 | 2.32E-07 | NA |
| rs17510814 | 12 | 28468969 | A | C | 421 | 0.00712589 | 6 | 14.0387924 | 2.71409689 | 2.31E-07 | NA |
| rs141756120 | 12 | 28511096 | A | C | 421 | 0.00831354 | 6 | 14.278403 | 2.76458538 | 2.41E-07 | NA |
| rs117991215 | 12 | 28511473 | T | C | 421 | 0.00831354 | 6 | 14.333666 | 2.77918736 | 2.50E-07 | NA |
| rs191930622 | 12 | 48284655 | G | A | 421 | 0.0023753 | 3 | 19.6783579 | 4.30610368 | 4.88E-06 | NA |
| rs56302696 | 12 | 48292830 | G | A | 421 | 0.0023753 | 3 | 19.8959041 | 4.31584707 | 4.03E-06 | NA |
| rs185620578 | 12 | 48569399 | C | T | 421 | 0.0023753 | 3 | 20.7232267 | 4.39505442 | 2.42E-06 | NA |
| rs190806532 | 12 | 48862147 | G | T | 421 | 0.0023753 | 3 | 21.326955 | 4.43856043 | 1.55E-06 | NA |
| rs568658857 | 12 | 49853998 | G | A | 421 | 0.00593824 | 5 | 14.2361193 | 3.09423422 | 4.21E-06 | NA |
| rs137880949 | 12 | 63306297 | T | C | 421 | 0.00475059 | 3 | 21.6084034 | 4.13589168 | 1.75E-07 | NA |
| rs191053292 | 12 | 63445280 | T | C | 421 | 0.00356295 | 3 | 22.9266488 | 4.11439732 | 2.51E-08 | NA |
| rs182437250 | 12 | 63608466 | T | C | 421 | 0.00475059 | 4 | 18.8083451 | 3.79343554 | 7.12E-07 | NA |
| rs76904423 | 12 | 101188744 | G | A | 421 | 0.01068884 | 10 | 9.80576231 | 2.11568908 | 3.57E-06 | NA |
| rs180764936 | 12 | 101498621 | T | C | 421 | 0.00356295 | 3 | 19.3473053 | 4.0200238 | 1.49E-06 | NA |
| rs185855183 | 12 | 101505126 | C | T | 421 | 0.00356295 | 3 | 18.7397727 | 3.86137343 | 1.22E-06 | NA |
| rs139598422 | 13 | 23887014 | A | G | 421 | 0.00356295 | 4 | 20.4886522 | 3.64167194 | 1.84E-08 | NA |
| rs143371352 | 13 | 47383834 | C | T | 421 | 0.00356295 | 3 | 20.5341625 | 3.87928964 | 1.20E-07 | NA |
| rs75186966 | 13 | 47395758 | A | C | 421 | 0.00356295 | 3 | 20.2281345 | 3.83225523 | 1.30E-07 | NA |
| rs150077525 | 13 | 57979549 | A | G | 421 | 0.00712589 | 7 | 12.9984875 | 2.71312414 | 1.66E-06 | NA |
| rs534845494 | 13 | 58213864 | A | G | 421 | 0.00475059 | 5 | 15.9431765 | 3.18484007 | 5.56E-07 | NA |
| rs140062526 | 13 | 59286033 | G | A | 421 | 0.00593824 | 5 | 14.0926088 | 3.02052903 | 3.08E-06 | NA |
| rs546286713 | 13 | 91411079 | G | A | 421 | 0.00475059 | 4 | 15.7027841 | 3.3611216 | 2.98E-06 | NA |
| rs567080482 | 13 | 95075405 | T | C | 421 | 0.00475059 | 4 | 15.4999561 | 3.31620837 | 2.95E-06 | NA |
| rs142928734 | 13 | 101601082 | G | A | 421 | 0.00356295 | 3 | 17.009593 | 3.70044989 | 4.29E-06 | NA |
| rs556680896 | 13 | 101602415 | C | T | 421 | 0.00356295 | 3 | 17.0254659 | 3.70286799 | 4.27E-06 | NA |
| rs184265355 | 13 | 108011352 | A | C | 421 | 0.00593824 | 6 | 14.4271719 | 2.91829792 | 7.67E-07 | NA |

| | | | | | | | | | | | |
|---|---|---|---|---|---|---|---|---|---|---|---|
| rs572961122 | 13 | 108012985 | C | T | 421 | 0.00593824 | 6 | 13.3354896 | 2.85827607 | 3.08E-06 | NA |
| rs528809914 | 13 | 113041256 | G | A | 421 | 0.00356295 | 3 | 19.5048552 | 3.76867111 | 2.27E-07 | NA |
| rs138215817 | 14 | 22641516 | A | G | 421 | 0.00475059 | 4 | 16.311019 | 3.27367022 | 6.28E-07 | NA |
| rs74704551 | 14 | 30161887 | C | T | 421 | 0.00356295 | 3 | 24.1990705 | 4.31064693 | 1.98E-08 | NA |
| rs116862847 | 14 | 64141677 | C | T | 421 | 0.00712589 | 7 | 15.3160369 | 2.95194262 | 2.12E-07 | NA |
| rs569916471 | 14 | 75891342 | G | A | 421 | 0.00475059 | 5 | 15.6404088 | 3.3537133 | 3.11E-06 | NA |
| rs113767990 | 14 | 81717563 | G | A | 421 | 0.00475059 | 4 | 14.5811155 | 3.19363095 | 4.98E-06 | NA |
| rs190251199 | 14 | 105590577 | T | C | 421 | 0.00356295 | 3 | 20.0125292 | 4.01627173 | 6.27E-07 | NA |
| rs185155853 | 15 | 41244100 | C | T | 421 | 0.00475059 | 5 | 15.9164657 | 3.40838631 | 3.02E-06 | NA |
| rs144026361 | 15 | 41248669 | C | T | 421 | 0.00475059 | 5 | 16.1372994 | 3.43734659 | 2.67E-06 | NA |
| rs558614420 | 15 | 41810870 | C | T | 421 | 0.00593824 | 6 | 13.9750831 | 3.02772732 | 3.92E-06 | NA |
| rs138109686 | 15 | 42051442 | A | G | 421 | 0.00593824 | 6 | 13.5956705 | 2.96761718 | 4.62E-06 | NA |
| rs145896760 | 15 | 42119222 | G | A | 421 | 0.00593824 | 6 | 13.3688825 | 2.92346809 | 4.81E-06 | NA |
| rs140642138 | 15 | 42125165 | G | A | 421 | 0.00593824 | 6 | 13.5832221 | 2.96274049 | 4.55E-06 | NA |
| rs6080 | 15 | 58837933 | C | A | 421 | 0.04394299 | 36 | 5.66476745 | 1.2393253 | 4.86E-06 | NA |
| rs145439370 | 15 | 58879765 | T | C | 421 | 0.03087886 | 26 | 6.89328891 | 1.40339117 | 9.02E-07 | NA |
| rs149425014 | 15 | 58951660 | T | C | 421 | 0.02612827 | 22 | 7.55224792 | 1.58320604 | 1.84E-06 | NA |
| rs146442492 | 15 | 58982115 | C | T | 421 | 0.02731591 | 24 | 7.05925858 | 1.51661316 | 3.25E-06 | NA |
| rs193253461 | 15 | 59229353 | A | G | 421 | 0.01306413 | 12 | 10.5374468 | 2.07404763 | 3.76E-07 | NA |
| rs184117160 | 15 | 59404306 | C | T | 421 | 0.01425178 | 11 | 10.3417236 | 2.13366338 | 1.25E-06 | NA |
| rs80292573 | 15 | 59435086 | T | G | 421 | 0.03444181 | 30 | 6.55323491 | 1.32696583 | 7.87E-07 | NA |
| rs182303755 | 15 | 59634792 | A | C | 421 | 0.01306413 | 12 | 10.5479917 | 2.09359291 | 4.70E-07 | NA |
| rs138217865 | 15 | 94392882 | C | T | 421 | 0.00475059 | 4 | 16.709641 | 3.45193896 | 1.29E-06 | NA |
| rs553840536 | 16 | 25697895 | A | G | 421 | 0.00356295 | 3 | 19.6203442 | 4.23547954 | 3.61E-06 | NA |
| rs183817723 | 16 | 59302775 | C | T | 421 | 0.00356295 | 4 | 17.5860507 | 3.7341919 | 2.48E-06 | NA |
| rs144954214 | 16 | 76179362 | A | G | 421 | 0.0023753 | 3 | 21.8113263 | 4.43942529 | 8.96E-07 | NA |
| rs529523094 | 16 | 77715551 | A | G | 421 | 0.00356295 | 3 | 18.1584346 | 3.72617419 | 1.10E-06 | NA |
| rs146728064 | 17 | 19265440 | G | A | 421 | 0.00712589 | 6 | 12.6400411 | 2.73467115 | 3.80E-06 | NA |
| rs191271637 | 17 | 52123260 | A | G | 421 | 0.00356295 | 3 | 18.379836 | 3.92083355 | 2.76E-06 | NA |
| rs185819304 | 18 | 27001580 | G | A | 421 | 0.00356295 | 3 | 19.9401113 | 3.94501953 | 4.32E-07 | NA |
| rs187942235 | 18 | 27030430 | C | T | 421 | 0.00356295 | 3 | 20.3968658 | 3.96963025 | 2.77E-07 | NA |
| rs143538552 | 18 | 29050262 | A | G | 421 | 0.00356295 | 3 | 19.1457218 | 3.63679703 | 1.41E-07 | NA |
| rs373746073 | 18 | 29058384 | C | A | 421 | 0.00356295 | 3 | 19.0847197 | 3.63798064 | 1.55E-07 | NA |
| rs146333745 | 18 | 55497457 | C | T | 421 | 0.00356295 | 4 | 17.4215784 | 3.56782335 | 1.04E-06 | NA |
| rs185464792 | 19 | 18797371 | C | T | 421 | 0.0023753 | 3 | 22.5896015 | 4.50191473 | 5.23E-07 | NA |
| rs186768950 | 19 | 18806124 | C | A | 421 | 0.0023753 | 3 | 22.7152061 | 4.50776479 | 4.68E-07 | NA |
| rs559008174 | 19 | 18876059 | C | T | 421 | 0.00475059 | 5 | 15.6025661 | 3.12812006 | 6.11E-07 | NA |
| rs546144116 | 19 | 19563339 | C | T | 421 | 0.0023753 | 3 | 23.1354815 | 4.47162758 | 2.29E-07 | NA |
| rs560206697 | 19 | 20729098 | C | T | 421 | 0.0023753 | 3 | 24.9989943 | 4.55350673 | 4.02E-08 | NA |
| rs111285015 | 19 | 23123198 | G | A | 421 | 0.00356295 | 3 | 27.3347642 | 4.62083983 | 3.31E-09 | NA |
| rs1008091735 | 19 | 31090099 | T | C | 421 | 0.00356295 | 3 | 18.4689243 | 3.69079909 | 5.61E-07 | NA |
| rs148433854 | 19 | 31096478 | G | A | 421 | 0.00356295 | 3 | 18.6267585 | 3.68806754 | 4.41E-07 | NA |
| rs140788628 | 20 | 15858501 | C | A | 421 | 0.01068884 | 8 | 11.8039284 | 2.2826396 | 2.33E-07 | NA |
| rs559228693 | 20 | 15963329 | G | A | 421 | 0.00593824 | 5 | 14.3643551 | 3.08196497 | 3.15E-06 | NA |
| rs184785969 | 21 | 17171431 | C | A | 421 | 0.00356295 | 3 | 19.8736486 | 3.70683937 | 8.26E-08 | NA |
| rs117280553 | 21 | 17207163 | T | C | 421 | 0.00356295 | 3 | 20.087417 | 3.70274826 | 5.80E-08 | NA |
| rs79486609 | 21 | 17245006 | G | A | 421 | 0.00356295 | 3 | 20.6624886 | 3.77824957 | 4.53E-08 | NA |
| rs73227413 | 21 | 23136973 | G | A | 421 | 0.03444181 | 28 | 5.96205514 | 1.27150176 | 2.75E-06 | NA |
| rs75024143 | 21 | 23156546 | G | T | 421 | 0.01425178 | 12 | 9.81254241 | 2.0684542 | 2.10E-06 | NA |
| rs192134381 | 21 | 23450714 | T | C | 421 | 0.00356295 | 3 | 21.788812 | 3.73125357 | 5.23E-09 | NA |
| rs397836601 | 21 | 23450714 | T | C | 421 | 0.00356295 | 3 | 21.788812 | 3.73125357 | 5.23E-09 | NA |
| rs118183140 | 21 | 35477486 | C | T | 421 | 0.02019002 | 17 | 7.5893248 | 1.56122232 | 1.17E-06 | NA |

| | | | | | | | | | | | |
|---|---|---|---|---|---|---|---|---|---|---|---|
| rs183586634 | 21 | 38763032 | G | A | 421 | 0.00593824 | 5 | 14.3132202 | 2.99210048 | 1.72E-06 | NA |
| rs117185941 | 21 | 38766484 | G | A | 421 | 0.00593824 | 5 | 15.3808786 | 3.17143013 | 1.24E-06 | NA |
| rs1329159859 | 21 | 38766484 | G | A | 421 | 0.00593824 | 5 | 15.3808786 | 3.17143013 | 1.24E-06 | NA |
| rs118084887 | 21 | 38863820 | T | C | 421 | 0.00593824 | 5 | 14.9382616 | 3.13674741 | 1.91E-06 | NA |
| rs150539922 | 21 | 43276916 | T | C | 421 | 0.00356295 | 3 | 16.6696937 | 3.64477381 | 4.79E-06 | NA |
| rs113625788 | 22 | 19969182 | C | T | 421 | 0.00831354 | 7 | 11.8084829 | 2.42941561 | 1.17E-06 | NA |
| rs541680196 | 22 | 40528090 | G | A | 421 | 0.00593824 | 5 | 14.3867808 | 2.94903659 | 1.07E-06 | NA |
| rs185139807 | 22 | 40594781 | G | A | 421 | 0.00593824 | 5 | 14.4330526 | 2.95312686 | 1.02E-06 | NA |
| rs141127122 | 22 | 40604439 | G | A | 421 | 0.00475059 | 4 | 15.3824875 | 3.30083549 | 3.16E-06 | NA |
| rs148998974 | 22 | 40620530 | A | G | 421 | 0.00593824 | 5 | 14.5129532 | 2.94976024 | 8.65E-07 | NA |
| rs555040883 | 22 | 40631476 | G | A | 421 | 0.00475059 | 4 | 15.4273069 | 3.30542717 | 3.05E-06 | NA |
| rs150946694 | 22 | 46853180 | T | C | 421 | 0.00475059 | 4 | 15.3082479 | 3.29125059 | 3.30E-06 | NA |
