## Supplementary material for "Pharmacogenomics of steroid-induced ocular hypertension: relationship to high-tension glaucomas and new pathophysiologic insight": Suppl Table S8

**Supplementary Table S8. GWAS Results Indianapolis-2 Replication Cohort**
**12 month quantitative trait (QT), P-value ordered**

| ID | chr | POS_37 | freq | MAC | Score | Score.SE | Score.Stat | Score.pval | Func.refGene | Gene.refGene | GeneDetail.refGene | rsID | gnomAD_ genome_ALL | gnomAD_ genome_ NFE | Rsq | hwe | chr_38 | POS_38 |
|---|---|---|---|---|---|---|---|---|---|---|---|---|---|---|---|---|---|---|
| 18:6045905:T:A | 18 | 6045905 | 0.049893204 | 10 | 2.439599988 | 0.449001371 | 5.43339095 | 5.52931E-08 | intronic | L3MBTL4 | . | rs74649788 | 0.0811 | 0.0475 | 0.923243 | 1 | chr18 | 6045906 |
| 13:30768424:G:A | 13 | 30768424 | 0.029165049 | 6 | 1.909228275 | 0.360021719 | 5.303091934 | 1.13858E-07 | intergenic | LINC00365;KATNAL1 | dist=85412;dist=8343 | rs74833295 | 0.0153 | 0.0232 | 0.9983 | 1 | chr13 | 30194287 |
| 3:146048618:G:A | 3 | 146048618 | 0.023815534 | 5 | 1.481987222 | 0.287749854 | 5.150262284 | 2.60122E-07 | intergenic | PLSCR4;PLSCR2 | dist=79652;dist=102457 | rs73150883 | 0.0163 | 0.0252 | 0.848249 | 1 | chr3 | 146330831 |
| 8:23542722:T:A | 8 | 23542722 | 0.016383495 | 3 | 1.2982161 | 0.255687499 | 5.077354608 | 3.82726E-07 | intergenic | NKX3-1;NKX2-6 | dist=2272;dist=17242 | rs189490695 | 0.0070 | 0.0092 | 0.874937 | 1 | chr8 | 23685209 |
| 8:23462708:A:G | 8 | 23462708 | 0.016941748 | 3 | 1.341800652 | 0.264601135 | 5.071031363 | 3.95666E-07 | intergenic | SLC25A37;NKX3-1 | dist=32645;dist=73498 | rs539324287 | 0.0033 | 0.0057 | 0.907302 | 1 | chr8 | 23605195 |
| 17:75157900:C:T | 17 | 75157900 | 0.033300971 | 7 | 1.815437814 | 0.358406655 | 5.065301631 | 4.07754E-07 | intronic | SEC14L1 | . | rs113199364 | 0.0230 | 0.0364 | 0.893794 | 1 | chr17 | 77161818 |
| 3:75482532:G:C | 3 | 75482532 | 0.011626214 | 2 | 0.80016771 | 0.159019081 | 5.031897471 | 4.85649E-07 | ncRNA_intronic | FAM86DP | . | rs185293133 | 0.0054 | 0.0078 | 0.492536 | 1 | chr3 | 75433381 |
| 18:6049551:C:A | 18 | 6049551 | 0.044349515 | 9 | 2.087223432 | 0.417798884 | 4.995761149 | 5.86042E-07 | intronic | L3MBTL4 | . | rs58503971 | 0.0786 | 0.0441 | 0.959099 | 1 | chr18 | 6049552 |
| 18:6049026:T:A | 18 | 6049026 | 0.044359223 | 9 | 2.087445553 | 0.417849208 | 4.995691057 | 5.86254E-07 | intronic | L3MBTL4 | . | rs79381542 | 0.0807 | 0.0470 | 0.959092 | 1 | chr18 | 6049027 |
| 5:115979852:T:C | 5 | 115979852 | 0.029092233 | 6 | 1.782650366 | 0.361233296 | 4.934900476 | 8.01916E-07 | intergenic | SEMA6A;LOC102467223 | dist=69230;dist=99146 | rs114241096 | 0.0185 | 0.0229 | 0.998458 | 1 | chr5 | 116644156 |
| 5:115980038:G:C | 5 | 115980038 | 0.029092233 | 6 | 1.782650366 | 0.361233296 | 4.934900476 | 8.01916E-07 | intergenic | SEMA6A;LOC102467223 | dist=69416;dist=98960 | rs114587121 | 0.0185 | 0.0229 | 0.998458 | 1 | chr5 | 116644342 |
| 5:115980724:A:G | 5 | 115980724 | 0.029097087 | 6 | 1.782403345 | 0.361231089 | 4.934246796 | 8.04606E-07 | intergenic | SEMA6A;LOC102467223 | dist=70102;dist=98274 | rs79516960 | 0.0171 | 0.0229 | 0.998286 | 1 | chr5 | 116645028 |
| 5:115980901:A:C | 5 | 115980901 | 0.029097087 | 6 | 1.782403345 | 0.361231089 | 4.934246796 | 8.04606E-07 | intergenic | SEMA6A;LOC102467223 | dist=70279;dist=98097 | rs78085382 | 0.0186 | 0.0228 | 0.998286 | 1 | chr5 | 116645205 |
| 5:115981236:C:G | 5 | 115981236 | 0.029097087 | 6 | 1.782403345 | 0.361231089 | 4.934246796 | 8.04606E-07 | intergenic | SEMA6A;LOC102467223 | dist=70614;dist=97762 | rs79794218 | 0.0170 | 0.0229 | 0.998286 | 1 | chr5 | 116645540 |
| 5:115981990:C:G | 5 | 115981990 | 0.029097087 | 6 | 1.782403345 | 0.361231089 | 4.934246796 | 8.04606E-07 | intergenic | SEMA6A;LOC102467223 | dist=71368;dist=97008 | rs114108437 | 0.0170 | 0.0229 | 0.998286 | 1 | chr5 | 116646294 |
| 5:115991913:A:T | 5 | 115991913 | 0.029087379 | 6 | 1.782276135 | 0.361233864 | 4.933856736 | 8.06216E-07 | intergenic | SEMA6A;LOC102467223 | dist=81291;dist=87085 | rs76006987 | 0.0185 | 0.0225 | 0.998629 | 1 | chr5 | 116656217 |
| 5:115988180:G:A | 5 | 115988180 | 0.029126214 | 6 | 1.784300998 | 0.361724673 | 4.932759998 | 8.10758E-07 | intergenic | SEMA6A;LOC102467223 | dist=77558;dist=90818 | rs77857559 | 0.0165 | 0.0221 | 1 | 1 | chr5 | 116652484 |
| 5:115988843:C:T | 5 | 115988843 | 0.029126214 | 6 | 1.784300998 | 0.361724673 | 4.932759998 | 8.10758E-07 | intergenic | SEMA6A;LOC102467223 | dist=78221;dist=90155 | rs115917626 | 0.0167 | 0.0223 | 1 | 1 | chr5 | 116653147 |
| 13:52703883:C:G | 13 | 52703883 | 0.036063107 | 7 | 1.896523445 | 0.386307062 | 4.90936778 | 9.13705E-07 | ncRNA_exonic | LOC101929657 | . | rs148654242 | 0.0202 | 0.0302 | 0.890541 | 1 | chr13 | 52129747 |
| 5:115976909:A:C | 5 | 115976909 | 0.029834951 | 6 | 1.764770451 | 0.360470372 | 4.895743415 | 9.79348E-07 | intergenic | SEMA6A;LOC102467223 | dist=66287;dist=102089 | rs76433992 | 0.0168 | 0.0226 | 0.974211 | 1 | chr5 | 116641213 |
| 2:50062774:T:G | 2 | 50062774 | 0.030864078 | 6 | 1.721021507 | 0.356977463 | 4.821092882 | 1.42774E-06 | intergenic | FSHR;NRXN1 | dist=681108;dist=82869 | rs138665118 | 0.0114 | 0.0171 | 0.892953 | 1 | chr2 | 49835636 |
| 5:162653096:T:C | 5 | 162653096 | 0.02438835 | 5 | 1.593867941 | 0.333478221 | 4.779526333 | 1.75709E-06 | intergenic | GABRG2;CCNG1 | dist=1070551;dist=211481 | rs116626718 | 0.0249 | 0.0325 | 0.995221 | 1 | chr5 | 163226090 |
| 2:126913427:C:G | 2 | 126913427 | 0.033174757 | 7 | 1.808450116 | 0.378512536 | 4.777781303 | 1.7724E-06 | intergenic | CNTNAP5;GYPC | dist=1240473;dist=500084 | rs28798228 | 0.0111 | 0.0144 | 0.961063 | 1 | chr2 | 126155850 |
| 2:74482548:G:T | 2 | 74482548 | 0.032208738 | 7 | 1.780987998 | 0.373137772 | 4.77300378 | 1.81498E-06 | intronic | SLC4A5 | . | rs12621729 | 0.1479 | 0.0335 | 0.942493 | 1 | chr2 | 74255421 |
| 5:162646718:G:C | 5 | 162646718 | 0.024461165 | 5 | 1.583713858 | 0.331858007 | 4.772263512 | 1.82167E-06 | intergenic | GABRG2;CCNG1 | dist=1064173;dist=217859 | rs115331238 | 0.0236 | 0.0306 | 0.983743 | 1 | chr5 | 163219712 |
| 15:27636953:T:G | 15 | 27636953 | 0.067961165 | 14 | 2.448622014 | 0.514411414 | 4.760046044 | 1.93549E-06 | intronic | GABRG3 | . | rs72705714 | 0.0522 | 0.0496 | 1 | 1 | chr15 | 27391807 |
| 15:27636387:G:A | 15 | 27636387 | 0.067961166 | 14 | 2.444243265 | 0.513892292 | 4.756333777 | 1.9714E-06 | intronic | GABRG3 | . | rs72705712 | 0.0463 | 0.0496 | 0.998164 | 1 | chr15 | 27391241 |
| 15:27637194:T:C | 15 | 27637194 | 0.067961166 | 14 | 2.444243265 | 0.513892292 | 4.756333777 | 1.9714E-06 | intronic | GABRG3 | . | rs72705715 | 0.0428 | 0.0496 | 0.998164 | 1 | chr15 | 27392048 |
| 15:27635356:C:G | 15 | 27635356 | 0.067970874 | 14 | 2.443670474 | 0.513826908 | 4.75582426 | 1.97638E-06 | intronic | GABRG3 | . | rs72705711 | 0.0601 | 0.0498 | 0.997858 | 1 | chr15 | 27390210 |
| 2:126955214:G:A | 2 | 126955214 | 0.032441747 | 7 | 1.7618909 | 0.370731447 | 4.752472211 | 2.00944E-06 | intergenic | CNTNAP5;GYPC | dist=1282260;dist=458297 | rs28796094 | 0.0111 | 0.0144 | 0.939443 | 1 | chr2 | 126197637 |
| 7:9489611:T:G | 7 | 9489611 | 0.019781553 | 4 | 1.317750936 | 0.277733732 | 4.744655705 | 2.08861E-06 | intergenic | NXPH1;PER4 | dist=697018;dist=184289 | rs17206867 | 0.0067 | 0.0108 | 0.886061 | 1 | chr7 | 9449981 |
| 15:31383118:A:C | 15 | 31383118 | 0.019359223 | 4 | 1.431828899 | 0.302121941 | 4.739241691 | 2.1452E-06 | intronic | TRPM1 | . | rs75968509 | 0.0124 | 0.0149 | 0.979554 | 1 | chr15 | 31090915 |
| 5:162661735:C:T | 5 | 162661735 | 0.025213592 | 5 | 1.5722456 | 0.331875986 | 4.73744913 | 2.16425E-06 | intergenic | GABRG2;CCNG1 | dist=1079190;dist=202842 | rs114008181 | 0.0256 | 0.0337 | 0.958521 | 1 | chr5 | 163234729 |
| 15:27633984:A:C | 15 | 27633984 | 0.068470875 | 14 | 2.432058671 | 0.513440775 | 4.736785214 | 2.17135E-06 | intronic | GABRG3 | . | rs72705708 | 0.0450 | 0.0491 | 0.990527 | 1 | chr15 | 27388838 |
| 5:162629778:G:A | 5 | 162629778 | 0.024538835 | 5 | 1.5321512 | 0.32364261 | 4.734083693 | 2.20047E-06 | intergenic | GABRG2;CCNG1 | dist=1047233;dist=234799 | rs116189826 | 0.0218 | 0.0286 | 0.937899 | 1 | chr5 | 163202772 |
| 15:27634215:C:T | 15 | 27634215 | 0.068349515 | 14 | 2.429618929 | 0.513388602 | 4.732514355 | 2.21756E-06 | intronic | GABRG3 | . | rs72705709 | 0.0448 | 0.0489 | 0.992399 | 1 | chr15 | 27389069 |
| 15:27634405:G:C | 15 | 27634405 | 0.068320389 | 14 | 2.428373305 | 0.513251522 | 4.731351398 | 2.2303E-06 | intronic | GABRG3 | . | rs72705710 | 0.0448 | 0.0489 | 0.992701 | 1 | chr15 | 27389259 |
| 8:133999440:A:C | 8 | 133999440 | 0.014558252 | 3 | 1.200642195 | 0.254591569 | 4.715954269 | 2.4058E-06 | intronic | TG | . | rs74591804 | 0.0132 | 0.0217 | 0.999662 | 1 | chr8 | 132987195 |
| 12:11046142:C:A | 12 | 11046142 | 0.03042233 | 6 | 1.581110295 | 0.336660743 | 4.696449854 | 2.64722E-06 | ncRNA_intronic | PRH1-PRR4 | . | rs111621289 | 0.0242 | 0.0300 | 0.810976 | 1 | chr12 | 10893543 |
| 6:126490324:C:T | 6 | 126490324 | 0.016917476 | 3 | 1.203746268 | 0.259271271 | 4.642806213 | 3.43709E-06 | intergenic | MIR5695;CENPW | dist=46562;dist=170611 | rs148294287 | 0.0107 | 0.0165 | 0.861892 | 1 | chr6 | 126169178 |
| 6:31248568:G:A | 6 | 31248568 | 0.024305825 | 5 | 1.53280058 | 0.332764823 | 4.606257864 | 4.0998E-06 | intergenic | HLA-C;HLA-B | dist=8655;dist=73081 | rs9380234 | 0.0429 | 0.0402 | 0.995714 | 1 | chr6 | 31280791 |
| 6:31238801:C:G | 6 | 31238801 | 0.024349515 | 5 | 1.533218592 | 0.332869958 | 4.606058777 | 4.10372E-06 | intronic | HLA-C | . | rs41544614 | 0.0428 | 0.0404 | 0.994693 | 1 | chr6 | 31271024 |
| 6:158989974:G:A | 6 | 158989974 | 0.015582524 | 3 | 1.102833648 | 0.239463028 | 4.60544434 | 4.11586E-06 | intronic | TMEM181 | . | rs142006494 | 0.0339 | 0.0408 | 0.909469 | 1 | chr6 | 158568942 |
| 6:31240096:G:A | 6 | 31240096 | 0.024271845 | 5 | 1.534745688 | 0.333295989 | 4.604752947 | 4.12956E-06 | upstream | HLA-C | dist=183 | rs9366775 | 0.0429 | 0.0404 | 1 | 1 | chr6 | 31272319 |
| 6:31240479:T:G | 6 | 31240479 | 0.024271845 | 5 | 1.534745688 | 0.333295989 | 4.604752947 | 4.12956E-06 | upstream | HLA-C | dist=566 | rs9357121 | 0.0431 | 0.0406 | 1 | 1 | chr6 | 31272702 |
| 6:31245080:G:A | 6 | 31245080 | 0.024271845 | 5 | 1.534745688 | 0.333295989 | 4.604752947 | 4.12956E-06 | intergenic | HLA-C;HLA-B | dist=5167;dist=76569 | rs9391714 | 0.0430 | 0.0403 | 1 | 1 | chr6 | 31277303 |
| 6:31247267:C:T | 6 | 31247267 | 0.024237864 | 5 | 1.532381559 | 0.332833918 | 4.60404267 | 4.14368E-06 | intergenic | HLA-C;HLA-B | dist=7354;dist=74382 | rs56356836 | 0.0429 | 0.0402 | 0.998566 | 1 | chr6 | 31279490 |
| 6:31247998:T:C | 6 | 31247998 | 0.024237864 | 5 | 1.532381559 | 0.332833918 | 4.60404267 | 4.14368E-06 | intergenic | HLA-C;HLA-B | dist=8085;dist=73651 | rs9405016 | 0.0430 | 0.0403 | 0.998566 | 1 | chr6 | 31280221 |
| 6:31248262:G:A | 6 | 31248262 | 0.024237864 | 5 | 1.532381559 | 0.332833918 | 4.60404267 | 4.14368E-06 | intergenic | HLA-C;HLA-B | dist=8349;dist=73387 | rs12529015 | 0.0429 | 0.0401 | 0.998566 | 1 | chr6 | 31280485 |
| 6:31248493:T:C | 6 | 31248493 | 0.024237864 | 5 | 1.532381559 | 0.332833918 | 4.60404267 | 4.14368E-06 | intergenic | HLA-C;HLA-B | dist=8580;dist=73156 | rs9368669 | 0.0429 | 0.0402 | 0.998566 | 1 | chr6 | 31280716 |
| 6:31256026:C:G | 6 | 31256026 | 0.024237864 | 5 | 1.532381559 | 0.332833918 | 4.60404267 | 4.14368E-06 | intergenic | HLA-C;HLA-B | dist=16113;dist=65623 | rs17198734 | 0.0253 | 0.0303 | 0.998566 | 1 | chr6 | 31288249 |
| 6:31256058:G:T | 6 | 31256058 | 0.024237864 | 5 | 1.532381559 | 0.332833918 | 4.60404267 | 4.14368E-06 | intergenic | HLA-C;HLA-B | dist=16145;dist=65591 | rs17192386 | 0.0358 | 0.0305 | 0.998566 | 1 | chr6 | 31288281 |
| 6:31238068:C:T | 6 | 31238068 | 0.024432039 | 5 | 1.532787887 | 0.332980302 | 4.603238922 | 4.15971E-06 | exonic | HLA-C | . | rs1050276 | 0.0426 | 0.0400 | 0.991508 | 1 | chr6 | 31270291 |
| 6:31239727:C:A | 6 | 31239727 | 0.024334952 | 5 | 1.529466801 | 0.332272783 | 4.603045682 | 4.16357E-06 | intronic | HLA-C | . | rs29029490 | 0.0429 | 0.0404 | 0.991645 | 1 | chr6 | 31271950 |
| 6:31240818:G:A | 6 | 31240818 | 0.024203884 | 5 | 1.529478709 | 0.332384994 | 4.601527555 | 4.19404E-06 | upstream | HLA-C | dist=905 | rs4298345 | 0.0429 | 0.0404 | 0.997144 | 1 | chr6 | 31273041 |

| | | | | | | | | | | | | | | | | | | |
|---|---|---|---|---|---|---|---|---|---|---|---|---|---|---|---|---|---|---|
| 3:196625842:T:C | 3 | 196625842 | 0.97179126 | 6 | -1.52139796 | 0.331189308 | -4.593741177 | 4.35369E-06 | intronic | SENP5 | . | rs6583185 | 0.9845 | 0.9781 | 0.853106 | 1 | chr3 | 196898971 |
| 5:162496418:T:C | 5 | 162496418 | 0.028330097 | 6 | 1.557183414 | 0.34021336 | 4.577078964 | 4.71514E-06 | intergenic | GABRG2;CCNG1 | dist=913873;dist=368159 | rs114500719 | 0.0243 | 0.0327 | 0.909911 | 1 | chr5 | 163069412 |
| 2:134644599:A:G | 2 | 134644599 | 0.014786408 | 3 | 1.137571373 | 0.248782585 | 4.572552272 | 4.81819E-06 | intergenic | NCKAP5;MIR3679 | dist=318568;dist=240097 | rs189440995 | 0.0060 | 0.0093 | 0.899627 | 1 | chr2 | 133887028 |
| 10:72790290:G:A | 10 | 72790290 | 0.038834951 | 8 | 1.881493073 | 0.411573362 | 4.571464641 | 4.84327E-06 | intergenic | PCBD1;UNC5B | dist=141747;dist=182002 | rs77364739 | 0.0865 | 0.0588 | 1 | 1 | chr10 | 71030533 |
| 4:125382184:C:G | 4 | 125382184 | 0.019456311 | 4 | 1.343334873 | 0.29403913 | 4.568558189 | 4.91091E-06 | intergenic | LINC01091;LOC101927087 | dist=530666;dist=38913 | rs72678487 | 0.0073 | 0.0115 | 0.987952 | 1 | chr4 | 124461029 |

**Supplementary Table S8. GWAS Results Indianapolis-2 Replication Cohort**
**12 month quantitative trait (QT), P-value ordered**

| ID | chr | POS_37 | freq | MAC | Score | Score.SE | Score.Stat | Score.pval | Func.refGene | Gene.refGene | GeneDetail.refGene | rsID | gnomAD_ genome_ALL | gnomAD_ genome_ NFE | Rsq | hwe | chr_38 | POS_38 |
|---|---|---|---|---|---|---|---|---|---|---|---|---|---|---|---|---|---|---|
| 13:71228466:C:A | 13 | 71228466 | 0.066820388 | 14 | 3.693410575 | 0.700908722 | 5.269460143 | 1.36826E-07 | intergenic | ATXN8OS;LINC00348 | dist=514581;dist=360807 | rs9592740 | 0.0651 | 0.0859 | 0.979474 | 1 | chr13 | 70654334 |
| 10:115449191:G:A | 10 | 115449191 | 0.01631068 | 3 | 1.860675373 | 0.358119043 | 5.195689556 | 2.03962E-07 | intronic | CASP7 | . | rs150977401 | 0.0073 | 0.0111 | 0.870763 | 1 | chr10 | 113689432 |
| 13:71242428:T:G | 13 | 71242428 | 0.065 | 13 | 3.560256036 | 0.685315257 | 5.195063146 | 2.0465E-07 | intergenic | ATXN8OS;LINC00348 | dist=528543;dist=346845 | rs9599709 | 0.0654 | 0.0860 | 0.958512 | 1 | chr13 | 70668296 |
| 2:130563530:G:A | 2 | 130563530 | 0.248378641 | 51 | 6.844310493 | 1.325444312 | 5.163785781 | 2.42005E-07 | intergenic | LOC151121;LOC389033 | dist=532066;dist=116905 | rs4337430 | 0.2131 | 0.2653 | 0.973034 | 0.279636 | chr2 | 129805957 |
| 2:151347581:A:G | 2 | 151347581 | 0.013723301 | 3 | 1.731904782 | 0.335798226 | 5.157575737 | 2.50168E-07 | intergenic | RND3;LOC101929260 | dist=3372;dist=61465 | rs145531574 | 0.0088 | 0.0141 | 0.8698 | 1 | chr2 | 150491067 |
| 15:61531695:C:G | 15 | 61531695 | 0.014752427 | 3 | 1.816785354 | 0.35534236 | 5.112774488 | 3.17461E-07 | intergenic | RORA;VPS13C | dist=10193;dist=612895 | rs143862437 | 0.0083 | 0.0139 | 0.9521 | 1 | chr15 | 61239496 |
| 15:61534825:G:A | 15 | 61534825 | 0.014762136 | 3 | 1.816594932 | 0.35534099 | 5.112258325 | 3.1833E-07 | intergenic | RORA;VPS13C | dist=13323;dist=609765 | rs182462684 | 0.0084 | 0.0138 | 0.951481 | 1 | chr15 | 61242626 |
| 12:78530991:C:T | 12 | 78530991 | 0.028985437 | 6 | 2.517383543 | 0.494187455 | 5.093985122 | 3.50614E-07 | exonic | NAV3 | . | rs61754236 | 0.0085 | 0.0125 | 0.995116 | 1 | chr12 | 78137211 |
| 12:898065:G:T | 12 | 898065 | 0.01511165 | 3 | 1.72959212 | 0.339602326 | 5.092992566 | 3.52456E-07 | intronic | WNK1 | . | rs2014160 | 0.0005 | 0.0007 | 0.853667 | 1 | chr12 | 788899 |
| 12:78545400:G:A | 12 | 78545400 | 0.02904369 | 6 | 2.519911214 | 0.495194249 | 5.088732794 | 3.60464E-07 | intronic | NAV3 | . | rs140686116 | 0.0063 | 0.0103 | 0.997108 | 1 | chr12 | 78151620 |
| 8:39944917:T:A | 8 | 39944917 | 0.030781554 | 6 | 2.577314922 | 0.506628696 | 5.087187008 | 3.63413E-07 | intergenic | IDO2;C8orf4 | dist=71007;dist=66070 | rs138697513 | 0.0130 | 0.0201 | 0.884291 | 1 | chr8 | 40087398 |
| 8:39944783:C:T | 8 | 39944783 | 0.030776699 | 6 | 2.576605742 | 0.50654249 | 5.086652731 | 3.64438E-07 | intergenic | IDO2;C8orf4 | dist=70873;dist=66204 | rs145892996 | 0.0131 | 0.0201 | 0.884119 | 1 | chr8 | 40087264 |
| 2:130565338:C:A | 2 | 130565338 | 0.253538835 | 52 | 6.70870463 | 1.334874447 | 5.025719568 | 5.01548E-07 | intergenic | LOC151121;LOC389033 | dist=533874;dist=115097 | rs34017523 | 0.2123 | 0.2644 | 0.978903 | 0.420041 | chr2 | 129807765 |
| 2:130567425:T:A | 2 | 130567425 | 0.253538835 | 52 | 6.70870463 | 1.334874447 | 5.025719568 | 5.01548E-07 | intergenic | LOC151121;LOC389033 | dist=535961;dist=113010 | rs6720381 | 0.2065 | 0.2652 | 0.978903 | 0.420041 | chr2 | 129809852 |
| 2:130573252:C:A | 2 | 130573252 | 0.254174757 | 52 | 6.714995733 | 1.337157087 | 5.021845077 | 5.11775E-07 | intergenic | LOC151121;LOC389033 | dist=541788;dist=107183 | rs34895266 | 0.2140 | 0.2654 | 0.980963 | 0.420041 | chr2 | 129815679 |
| 19:46480407:T:C | 19 | 46480407 | 0.019417476 | 4 | 2.066073669 | 0.41173362 | 5.017986311 | 5.22159E-07 | intergenic | NOVA2;CCDC61 | dist=3750;dist=18312 | rs4239537 | 0.0835 | 0.0109 | 1 | 1 | chr19 | 45977149 |
| 22:19080386:G:A | 22 | 19080386 | 0.024252427 | 5 | 2.149293074 | 0.429334633 | 5.006102254 | 5.55433E-07 | intronic | DGCR2 | . | rs150428353 | 0.0172 | 0.0230 | 0.783413 | 1 | chr22 | 19092873 |
| 15:61567547:C:T | 15 | 61567547 | 0.01557767 | 3 | 1.784212495 | 0.357897115 | 4.985266493 | 6.18765E-07 | intergenic | RORA;VPS13C | dist=46045;dist=577043 | rs144327698 | 0.0097 | 0.0159 | 0.916644 | 1 | chr15 | 61275348 |
| 15:61555055:G:C | 15 | 61555055 | 0.015490291 | 3 | 1.774737275 | 0.356244746 | 4.98179214 | 6.29981E-07 | intergenic | RORA;VPS13C | dist=33553;dist=589535 | rs143096638 | 0.0093 | 0.0154 | 0.913405 | 1 | chr15 | 61262856 |
| 3:64559845:T:C | 3 | 64559845 | 0.014699029 | 3 | 1.72126431 | 0.347407762 | 4.954593705 | 7.24816E-07 | ncRNA_intronic | ADAMTS9-AS1 | . | rs146268777 | 0.0046 | 0.0068 | 0.94425 | 1 | chr3 | 64574169 |
| 2:130584142:C:T | 2 | 130584142 | 0.222898058 | 46 | 6.124040336 | 1.238717369 | 4.943856031 | 7.65923E-07 | intergenic | LOC151121;LOC389033 | dist=552678;dist=96293 | rs35748618 | 0.1481 | 0.2177 | 0.96226 | 1 | chr2 | 129826569 |
| 5:73263379:A:G | 5 | 73263379 | 0.024257281 | 5 | 2.247608363 | 0.454880737 | 4.941093741 | 7.76855E-07 | intergenic | ARHGEF28;LINC01335 | dist=25561;dist=338856 | rs190652742 | 0.0104 | 0.0094 | 0.999387 | 1 | chr5 | 73967554 |
| 2:130592585:C:T | 2 | 130592585 | 0.258529127 | 53 | 6.558142102 | 1.329599407 | 4.932419546 | 8.12172E-07 | intergenic | LOC151121;LOC389033 | dist=561121;dist=87850 | rs6742395 | 0.2225 | 0.2681 | 0.976222 | 0.430075 | chr2 | 129835012 |
| X:86181688:A:G | X | 86181688 | 0.013970874 | 3 | 1.634757242 | 0.331433352 | 4.932386049 | 8.12312E-07 | intergenic | DACH2;KLHL4 | dist=94083;dist=591027 | rs186620466 | 0.0162 | 0.0191 | 0.873035 | 1 | chrX | 86926685 |
| 10:21883439:G:A | 10 | 21883439 | 0.015543689 | 3 | 1.645849191 | 0.333793955 | 4.93073396 | 8.19212E-07 | intronic | MLLT10 | . | rs191906004 | 0.0058 | 0.0077 | 0.843604 | 1 | chr10 | 21594510 |
| 10:21970382:G:A | 10 | 21970382 | 0.014849515 | 3 | 1.594035309 | 0.323294895 | 4.930592272 | 8.19807E-07 | intronic | MLLT10 | . | rs188790792 | 0.0052 | 0.0080 | 0.823859 | 1 | chr10 | 21681453 |
| 2:130586642:C:T | 2 | 130586642 | 0.257281553 | 53 | 6.683699411 | 1.358232147 | 4.920881476 | 8.61553E-07 | intergenic | LOC151121;LOC389033 | dist=555178;dist=93793 | rs4277471 | 0.1953 | 0.2581 | 1 | 0.300358 | chr2 | 129829069 |
| 1:53720723:T:C | 1 | 53720723 | 0.040257281 | 8 | 2.387313586 | 0.485212014 | 4.920145248 | 8.648E-07 | intronic | LRP8 | . | rs12116501 | 0.0311 | 0.0437 | 0.807097 | 1 | chr1 | 53255051 |
| 1:240998165:G:A | 1 | 240998165 | 0.019203883 | 4 | 1.815512107 | 0.369826911 | 4.909085989 | 9.15019E-07 | intronic | RGS7 | . | rs72754890 | 0.0051 | 0.0073 | 0.806371 | 1 | chr1 | 240834865 |
| 7:89853957:C:T | 7 | 89853957 | 0.018815534 | 4 | 1.879423001 | 0.384569904 | 4.887077696 | 1.02344E-06 | intronic | STEAP2 | . | rs149916697 | 0.0065 | 0.0095 | 0.928405 | 1 | chr7 | 90224643 |
| 7:89988476:T:C | 7 | 89988476 | 0.018800971 | 4 | 1.89404647 | 0.387759417 | 4.8845918 | 1.03643E-06 | intronic | GTPBP10 | . | rs79847565 | 0.0069 | 0.0100 | 0.945469 | 1 | chr7 | 90359162 |
| 7:90013529:G:A | 7 | 90013529 | 0.018776699 | 4 | 1.910374378 | 0.392033885 | 4.872982793 | 1.09926E-06 | intronic | GTPBP10 | . | rs187685448 | 0.0067 | 0.0098 | 0.967919 | 1 | chr7 | 90384215 |
| 2:130589408:A:T | 2 | 130589408 | 0.263592233 | 54 | 6.506401141 | 1.335621499 | 4.871440856 | 1.10787E-06 | intergenic | LOC151121;LOC389033 | dist=557944;dist=91027 | rs1882610 | 0.2367 | 0.2795 | 0.977378 | 0.601073 | chr2 | 129831835 |
| 7:90047115:A:G | 7 | 90047115 | 0.018825242 | 4 | 1.913164032 | 0.392804365 | 4.870526408 | 1.11301E-06 | intergenic | CLDN12;CDK14 | dist=1847;dist=178561 | rs143425398 | 0.0066 | 0.0097 | 0.970376 | 1 | chr7 | 90417801 |
| 2:130582408:G:C | 2 | 130582408 | 0.264009709 | 54 | 6.522687428 | 1.33928955 | 4.87025933 | 1.11452E-06 | intergenic | LOC151121;LOC389033 | dist=550944;dist=98027 | rs12621133 | 0.2291 | 0.2789 | 0.979973 | 0.601073 | chr2 | 129824835 |
| 7:90253061:G:A | 7 | 90253061 | 0.01892233 | 4 | 1.921233543 | 0.394605718 | 4.868742276 | 1.12311E-06 | intronic | CDK14 | . | rs150148213 | 0.0066 | 0.0097 | 0.973808 | 1 | chr7 | 90623747 |
| 1:12215283:T:C | 1 | 12215283 | 0.013373786 | 3 | 1.580945998 | 0.324859599 | 4.866551587 | 1.13562E-06 | intergenic | TNFRSF8;MIR7846 | dist=11019;dist=11717 | rs147985761 | 0.0081 | 0.0132 | 0.894028 | 1 | chr1 | 12155226 |
| 9:36357606:C:A | 9 | 36357606 | 0.023582524 | 5 | 2.18417045 | 0.448849347 | 4.866154904 | 1.1379E-06 | intronic | RNF38 | . | rs4879986 | 0.0112 | 0.0117 | 0.970686 | 1 | chr9 | 36357609 |
| 9:36381902:C:T | 9 | 36381902 | 0.97573301 | 5 | -2.242692534 | 0.461738278 | -4.857064356 | 1.19139E-06 | intronic | RNF38 | . | rs2248297 | 0.9692 | 0.9879 | 0.999795 | 1 | chr9 | 36381905 |
| 4:163976364:T:C | 4 | 163976364 | 0.028898058 | 6 | 2.023347127 | 0.417548067 | 4.845782525 | 1.26114E-06 | intergenic | FSTL5;MIR4454 | dist=891178;dist=38362 | rs146154846 | 0.0182 | 0.0245 | 0.711952 | 1 | chr4 | 163055212 |
| 2:117348669:G:A | 2 | 117348669 | 0.020179612 | 4 | 2.005609812 | 0.414444146 | 4.839276486 | 1.30313E-06 | intergenic | DPP10;DDX18 | dist=746343;dist=1223586 | rs75685045 | 0.0065 | 0.0111 | 0.944983 | 1 | chr2 | 116591093 |
| 9:36360845:T:C | 9 | 36360845 | 0.024014563 | 5 | 2.210438405 | 0.456810098 | 4.838856266 | 1.30588E-06 | intronic | RNF38 | . | rs79319828 | 0.0116 | 0.0120 | 0.9887 | 1 | chr9 | 36360848 |
| 9:36386865:T:G | 9 | 36386865 | 0.024029126 | 5 | 2.210045419 | 0.456790677 | 4.838201677 | 1.31019E-06 | intronic | RNF38 | . | rs76774745 | 0.0117 | 0.0120 | 0.988086 | 1 | chr9 | 36386868 |
| 17:43780479:T:C | 17 | 43780479 | 0.0145 | 3 | 1.703938501 | 0.352883611 | 4.828613304 | 1.37487E-06 | intronic | MGC57346-CRHR1 | . | rs79985283 | 0.0211 | 0.0077 | 0.994938 | 1 | chr17 | 45703113 |
| 10:72790290:G:A | 10 | 72790290 | 0.038834951 | 8 | 2.690625403 | 0.557664123 | 4.824813526 | 1.40134E-06 | intergenic | PCBD1;UNC5B | dist=141747;dist=182002 | rs77364739 | 0.0865 | 0.0588 | 1 | 1 | chr10 | 71030533 |
| 1:70886329:A:G | 1 | 70886329 | 0.038961165 | 8 | 2.731688539 | 0.567885471 | 4.81028073 | 1.50718E-06 | intronic | CTH | . | rs79934683 | 0.0306 | 0.0416 | 0.993133 | 1 | chr1 | 70420646 |
| 20:6955818:G:T | 20 | 6955818 | 0.017223301 | 4 | 1.82640918 | 0.380591181 | 4.798874146 | 1.5956E-06 | intergenic | BMP2;LINC01428 | dist=194893;dist=171296 | rs146812516 | 0.0161 | 0.0210 | 0.862977 | 1 | chr20 | 6975171 |
| 14:24001662:C:T | 14 | 24001662 | 0.024456311 | 5 | 1.768121176 | 0.369052815 | 4.790970568 | 1.65976E-06 | intronic | ZFHX2 | . | rs41307102 | 0.0265 | 0.0375 | 0.636713 | 1 | chr14 | 23532453 |
| 8:40049692:T:G | 8 | 40049692 | 0.044912622 | 9 | 2.785140072 | 0.582245322 | 4.783447747 | 1.72314E-06 | intergenic | C8orf4;ZMAT4 | dist=36865;dist=338419 | rs150470417 | 0.0259 | 0.0383 | 0.960377 | 1 | chr8 | 40192173 |
| 8:40027101:T:C | 8 | 40027101 | 0.04368932 | 9 | 2.810807743 | 0.588520937 | 4.776053945 | 1.78768E-06 | intergenic | C8orf4;ZMAT4 | dist=14274;dist=361010 | rs116259399 | 0.0402 | 0.0401 | 1 | 1 | chr8 | 40169582 |
| 5:5233724:G:C | 5 | 5233724 | 0.014830097 | 3 | 1.706394641 | 0.358423256 | 4.760836832 | 1.92792E-06 | intronic | ADAMTS16 | . | rs115149137 | 0.0104 | 0.0175 | 0.967403 | 1 | chr5 | 5233611 |
| 7:89801861:C:T | 7 | 89801861 | 0.017533981 | 4 | 1.680077446 | 0.353443095 | 4.753459521 | 1.99965E-06 | ncRNA_intronic | STEAP2-AS1 | . | rs7800716 | 0.0069 | 0.0101 | 0.830555 | 1 | chr7 | 90172547 |
| 4:40948164:G:A | 4 | 40948164 | 0.037621359 | 8 | 2.919015192 | 0.615000485 | 4.746362423 | 2.07107E-06 | intronic | APBB2 | . | rs113189296 | 0.0591 | 0.0852 | 0.970482 | 1 | chr4 | 40946147 |
| 1:240988921:C:T | 1 | 240988921 | 0.020087379 | 4 | 1.824932693 | 0.38452179 | 4.74597992 | 2.07499E-06 | intronic | RGS7 | . | rs72754888 | 0.0047 | 0.0067 | 0.836433 | 1 | chr1 | 240825621 |
| 5:5241203:C:A | 5 | 5241203 | 0.014868932 | 3 | 1.68070787 | 0.35442199 | 4.742109459 | 2.11504E-06 | intronic | ADAMTS16 | . | rs145155685 | 0.0080 | 0.0129 | 0.944326 | 1 | chr5 | 5241090 |
| 7:48396066:G:A | 7 | 48396066 | 0.014519418 | 3 | 1.695949216 | 0.358675926 | 4.728360877 | 2.2634E-06 | intronic | ABCA13 | . | rs189164061 | 0.0077 | 0.0119 | 0.996283 | 1 | chr7 | 48356469 |
| 11:44085975:C:A | 11 | 44085975 | 0.014893204 | 3 | 1.699510358 | 0.360057984 | 4.720101852 | 2.35727E-06 | intergenic | ACCSL;ACCS | dist=4448;dist=1754 | rs76143420 | 0.0134 | 0.0215 | 0.971325 | 1 | chr11 | 44064425 |

| | | | | | | | | | | | | | | | | | | |
|---|---|---|---|---|---|---|---|---|---|---|---|---|---|---|---|---|---|---|
| 13:102628784:G:A | 13 | 102628784 | 0.00926699 | 2 | 1.077369422 | 0.228342774 | 4.718211147 | 2.37927E-06 | intronic | FGF14 | . | rs75197268 | 0.0122 | 0.0178 | 0.53936 | 1 | chr13 | 101976434 |
| 18:39394472:G:C | 18 | 39394472 | 0.023024272 | 5 | 1.975849294 | 0.419524828 | 4.709731495 | 2.48043E-06 | intergenic | KC6;PIK3C3 | dist=293911;dist=140691 | rs113484345 | 0.0229 | 0.0286 | 0.890728 | 1 | chr18 | 41814507 |
| 4:173193727:G:A | 4 | 173193727 | 0.014470874 | 3 | 1.644356357 | 0.349394212 | 4.706306808 | 2.52245E-06 | intronic | GALNTL6 | . | rs187567621 | 0.0019 | 0.0033 | 0.94795 | 1 | chr4 | 172272576 |
| 10:98558540:C:A | 10 | 98558540 | 0.013485437 | 3 | 1.29789842 | 0.276292266 | 4.697556103 | 2.63293E-06 | intergenic | PIK3AP1;MIR607 | dist=78261;dist=29886 | rs61856855 | 0.0113 | 0.0141 | 0.63836 | 1 | chr10 | 96798783 |
| 13:61672508:T:C | 13 | 61672508 | 0.027004854 | 6 | 2.031117161 | 0.432525467 | 4.695948131 | 2.65373E-06 | intergenic | LINC00378;MIR3169 | dist=402574;dist=101424 | rs147220861 | 0.0080 | 0.0082 | 0.826016 | 1 | chr13 | 61098374 |
| 13:109580621:C:T | 13 | 109580621 | 0.022786408 | 5 | 2.002149238 | 0.426532153 | 4.694017139 | 2.67892E-06 | intronic | MYO16 | . | rs145442123 | 0.0106 | 0.0163 | 0.930524 | 1 | chr13 | 108928273 |
| 12:62144998:G:A | 12 | 62144998 | 0.01723301 | 4 | 1.744080472 | 0.37159489 | 4.693499608 | 2.68571E-06 | intronic | FAM19A2 | . | rs76854078 | 0.0089 | 0.0135 | 0.904535 | 1 | chr12 | 61751217 |
| 11:44073753:C:T | 11 | 44073753 | 0.01565534 | 3 | 1.691379055 | 0.360755196 | 4.688439897 | 2.75296E-06 | intronic | ACCSL | . | rs149030097 | 0.0134 | 0.0213 | 0.931546 | 1 | chr11 | 44052203 |
| 4:164000229:C:T | 4 | 164000229 | 0.02361165 | 5 | 1.814612257 | 0.387671579 | 4.680797751 | 2.85761E-06 | intergenic | FSTL5;MIR4454 | dist=915043;dist=14497 | rs145034465 | 0.0184 | 0.0231 | 0.732815 | 1 | chr4 | 163079077 |
| 10:72806820:G:A | 10 | 72806820 | 0.036461165 | 8 | 2.445966686 | 0.523449162 | 4.672787474 | 2.97139E-06 | intergenic | PCBD1;UNC5B | dist=158277;dist=165472 | rs75505956 | 0.0366 | 0.0553 | 0.953209 | 1 | chr10 | 71047063 |
| 17:52366146:G:A | 17 | 52366146 | 0.023776699 | 5 | 1.836346208 | 0.394018804 | 4.660554743 | 3.15358E-06 | intergenic | KIF2B;TOM1L1 | dist=463573;dist=611906 | rs62072692 | 0.0232 | 0.0371 | 0.819939 | 1 | chr17 | 54288785 |
| 2:130568556:T:C | 2 | 130568556 | 0.297684466 | 61 | 6.436745057 | 1.381972449 | 4.657650781 | 3.19838E-06 | intergenic | LOC151121;LOC389033 | dist=537092;dist=111879 | rs4355062 | 0.2389 | 0.2895 | 0.981992 | 0.478458 | chr2 | 129810983 |
| 2:72629234:T:G | 2 | 72629234 | 0.018174757 | 4 | 1.80424283 | 0.387623961 | 4.654621515 | 3.24576E-06 | intronic | EXOC6B | . | rs185879164 | 0.0118 | 0.0192 | 0.929494 | 1 | chr2 | 72402105 |
| 10:21749029:G:A | 10 | 21749029 | 0.011912621 | 2 | 1.22791161 | 0.263818829 | 4.654374424 | 3.24966E-06 | intergenic | NEBL-AS1;CASC10 | dist=285177;dist=34392 | rs183818481 | 0.0059 | 0.0081 | 0.674237 | 1 | chr10 | 21460100 |
| 15:45198007:A:C | 15 | 45198007 | 0.048796117 | 10 | 2.632234639 | 0.565845842 | 4.651858229 | 3.28957E-06 | intergenic | TRIM69;C15orf43 | dist=137980;dist=50893 | rs28631454 | 0.3216 | 0.1096 | 0.89064 | 1 | chr15 | 44905809 |
| 20:45720309:A:G | 20 | 45720309 | 0.032533981 | 7 | 2.316388446 | 0.498848234 | 4.643473279 | 3.426E-06 | intronic | EYA2 | . | rs146684353 | 0.0232 | 0.0301 | 0.918602 | 1 | chr20 | 47091670 |
| 6:77472188:A:G | 6 | 77472188 | 0.019490291 | 4 | 1.93070443 | 0.415892439 | 4.642316741 | 3.44524E-06 | intergenic | IMPG1;HTR1B | dist=689793;dist=698377 | rs55850402 | 0.0070 | 0.0099 | 0.988316 | 1 | chr6 | 76762471 |
| 6:77460389:C:T | 6 | 77460389 | 0.019490291 | 4 | 1.930446837 | 0.415888081 | 4.641745998 | 3.45477E-06 | intergenic | IMPG1;HTR1B | dist=677994;dist=710176 | rs191563430 | 0.0071 | 0.0101 | 0.988316 | 1 | chr6 | 76750672 |
| 2:72852782:C:A | 2 | 72852782 | 0.018898058 | 4 | 1.818657772 | 0.391871559 | 4.640953721 | 3.46805E-06 | intronic | EXOC6B | . | rs115540332 | 0.0116 | 0.0188 | 0.914669 | 1 | chr2 | 72625653 |
| 5:53390040:G:C | 5 | 53390040 | 0.306907767 | 63 | 6.436279804 | 1.389496475 | 4.632095094 | 3.61984E-06 | intronic | ARL15 | . | rs42872 | 0.2490 | 0.2995 | 0.980711 | 0.812933 | chr5 | 54094210 |
| X:20935027:G:A | X | 20935027 | 0.015194175 | 3 | 1.517881729 | 0.327834067 | 4.630030494 | 3.65612E-06 | intergenic | RPS6KA3;CNKSR2 | dist=650277;dist=457509 | rs144178739 | 0.0231 | 0.0199 | 0.814532 | 1 | chrX | 20916909 |
| 5:33636035:T:C | 5 | 33636035 | 0.155339806 | 32 | 5.112931608 | 1.104845474 | 4.627734581 | 3.69687E-06 | intronic | ADAMTS12 | . | rs13164825 | 0.1577 | 0.1968 | 1 | 0.705556 | chr5 | 33635930 |
| 4:31782754:A:G | 4 | 31782754 | 0.053990291 | 11 | 3.25806796 | 0.70424346 | 4.626337545 | 3.72189E-06 | intergenic | LOC102723778;LOC102723828 | dist=569458;dist=216247 | rs79231845 | 0.0271 | 0.0314 | 0.964283 | 1 | chr4 | 31781132 |
| 6:77429812:T:G | 6 | 77429812 | 0.019640777 | 4 | 1.923001054 | 0.415738055 | 4.625511258 | 3.73676E-06 | intergenic | IMPG1;HTR1B | dist=647417;dist=740753 | rs77029592 | 0.0083 | 0.0100 | 0.980847 | 1 | chr6 | 76720095 |
| 10:22083617:A:G | 10 | 22083617 | 0.011519417 | 2 | 1.283845844 | 0.277566907 | 4.625356308 | 3.73955E-06 | intronic | DNAJC1 | . | rs181139900 | 0.0064 | 0.0085 | 0.775781 | 1 | chr10 | 21794688 |
| 5:33646456:G:C | 5 | 33646456 | 0.164631068 | 34 | 4.973632977 | 1.076119596 | 4.621821771 | 3.80385E-06 | intronic | ADAMTS12 | . | rs10057508 | 0.1583 | 0.2157 | 0.991904 | 1 | chr5 | 33646351 |
| 6:77420028:C:A | 6 | 77420028 | 0.019699029 | 4 | 1.920385778 | 0.415757928 | 4.618999777 | 3.85594E-06 | intergenic | IMPG1;HTR1B | dist=637633;dist=750537 | rs78549750 | 0.0160 | 0.0101 | 0.978334 | 1 | chr6 | 76710311 |
| 6:77414670:G:C | 6 | 77414670 | 0.019820388 | 4 | 1.925532437 | 0.417298507 | 4.614280674 | 3.94459E-06 | intergenic | IMPG1;HTR1B | dist=632275;dist=755895 | rs56232065 | 0.0103 | 0.0099 | 0.980026 | 1 | chr6 | 76704953 |
| 5:33647634:G:A | 5 | 33647634 | 0.159796116 | 33 | 4.948363002 | 1.073508457 | 4.609524006 | 4.03592E-06 | intronic | ADAMTS12 | . | rs13176485 | 0.1519 | 0.2149 | 0.991564 | 1 | chr5 | 33647529 |
| 5:33644123:G:T | 5 | 33644123 | 0.15973301 | 33 | 4.946113148 | 1.073435357 | 4.60774197 | 4.07065E-06 | intronic | ADAMTS12 | . | rs35388476 | 0.1520 | 0.2154 | 0.992101 | 1 | chr5 | 33644018 |
| 21:35375126:G:A | 21 | 35375126 | 0.007194175 | 1 | 0.846200633 | 0.183711704 | 4.606133495 | 4.10225E-06 | intergenic | LINC00649;MRPS6 | dist=38864;dist=70697 | rs144499025 | 0.0048 | 0.0076 | 0.538813 | 1 | chr21 | 34002825 |
| 11:21775222:T:A | 11 | 21775222 | 0.012529126 | 3 | 1.341788897 | 0.291618921 | 4.601172279 | 4.2012E-06 | intergenic | NELL1;ANO5 | dist=177993;dist=439500 | rs147118669 | 0.0050 | 0.0085 | 0.79684 | 1 | chr11 | 21753676 |
| 10:22068468:C:A | 10 | 22068468 | 0.011684466 | 2 | 1.295186101 | 0.281543412 | 4.600306895 | 4.21869E-06 | intronic | DNAJC1 | . | rs184920540 | 0.0059 | 0.0079 | 0.788199 | 1 | chr10 | 21779539 |
| 5:33644614:T:C | 5 | 33644614 | 0.160194175 | 33 | 4.958247799 | 1.078142852 | 4.598878331 | 4.24772E-06 | intronic | ADAMTS12 | . | rs67267661 | 0.1519 | 0.2153 | 1 | 1 | chr5 | 33644509 |
| 4:31766001:G:A | 4 | 31766001 | 0.053223301 | 11 | 3.28160902 | 0.713701131 | 4.598015718 | 4.26534E-06 | intergenic | LOC102723778;LOC102723828 | dist=552705;dist=233000 | rs16885796 | 0.0382 | 0.0334 | 0.994282 | 1 | chr4 | 31764379 |
| 2:66301595:G:A | 2 | 66301595 | 0.024257282 | 5 | 2.107857413 | 0.458486846 | 4.597421782 | 4.27751E-06 | intergenic | SPRED2;MIR4778 | dist=641939;dist=283786 | rs11676376 | 0.0435 | 0.0423 | 0.998566 | 1 | chr2 | 66074461 |
| 10:22216057:G:C | 10 | 22216057 | 0.013985437 | 3 | 1.470044503 | 0.319890854 | 4.595456497 | 4.31803E-06 | intronic | DNAJC1 | . | rs45496395 | 0.0063 | 0.0084 | 0.8731 | 1 | chr10 | 21927128 |
| 4:31779863:T:C | 4 | 31779863 | 0.053402913 | 11 | 3.29083276 | 0.717091233 | 4.589140975 | 4.45074E-06 | intergenic | LOC102723778;LOC102723828 | dist=566567;dist=219138 | rs115916744 | 0.0283 | 0.0312 | 0.999904 | 0.247516 | chr4 | 31778241 |
| 4:31772884:C:T | 4 | 31772884 | 0.053398058 | 11 | 3.290738644 | 0.717096412 | 4.588976583 | 4.45424E-06 | intergenic | LOC102723778;LOC102723828 | dist=559588;dist=226117 | rs116957173 | 0.0270 | 0.0312 | 1 | 0.245321 | chr4 | 31771262 |
| 5:3562095:T:A | 5 | 3562095 | 0.00734466 | 2 | 0.983175268 | 0.214354222 | 4.58668487 | 4.5034E-06 | intergenic | LINC01019;IRX1 | dist=25887;dist=34073 | rs143503046 | 0.0027 | 0.0041 | 0.686588 | 1 | chr5 | 3561981 |
| 4:31777594:G:A | 4 | 31777594 | 0.053165049 | 11 | 3.264991759 | 0.71189775 | 4.586321223 | 4.51124E-06 | intergenic | LOC102723778;LOC102723828 | dist=564298;dist=221407 | rs116168704 | 0.0285 | 0.0314 | 0.991203 | 1 | chr4 | 31775972 |
| 1:18614493:G:T | 1 | 18614493 | 0.014364078 | 3 | 1.46894969 | 0.320848564 | 4.578327141 | 4.68709E-06 | intronic | IGSF21 | . | rs115263712 | 0.0101 | 0.0137 | 0.8337 | 1 | chr1 | 18287999 |
| 4:189389929:T:C | 4 | 189389929 | 0.010296117 | 2 | 1.359541054 | 0.297050321 | 4.576803849 | 4.72134E-06 | ncRNA_intronic | LINC01060 | . | rs560030667 | 0.0037 | 0.0047 | 0.944465 | 1 | chr4 | 188468775 |
| 4:189389927:T:C | 4 | 189389927 | 0.010276699 | 2 | 1.359541791 | 0.297112351 | 4.575850811 | 4.74288E-06 | ncRNA_intronic | LINC01060 | . | rs544973300 | 0.0035 | 0.0050 | 0.946406 | 1 | chr4 | 188468773 |
| 5:112013914:G:A | 5 | 112013914 | 0.980427185 | 4 | -1.381862916 | 0.302073383 | -4.574593434 | 4.77146E-06 | ncRNA_intronic | LOC102467216 | . | rs12153142 | 0.9143 | 0.9991 | 0.499216 | 1 | chr5 | 112678217 |

**Supplementary Table S8. GWAS Results Indianapolis-2 Replication Cohort**
**12 month quantitative trait (QT), risk locus ordered**

Shaded cells in Gene.refGene column indicate risk loci that co-localize with risk loci identified in the Indianapolis-1 GWAS

| ID_37 | chr_37 | POS_37 | rsID | chr_38 | POS_38 | freq | MAC | Score | Score.SE | Score.Stat | Score.pval | Func.refGene | Gene.refGene | GeneDetail.refGene | AOP-expressed refGene (protein coding only) | gnomAD_ genome_ALL | gnomAD_ genome_ NFE | Rsq | hwe | AOP-expressed gene name |
|---|---|---|---|---|---|---|---|---|---|---|---|---|---|---|---|---|---|---|---|---|
| 2:50062774:T:G | 2 | 50062774 | rs138665118 | chr2 | 49835636 | 0.03086408 | 6 | 1.721021507 | 0.35697746 | 4.821092882 | 1.42774E-06 | intergenic | FSHR;NRXN1 | dist=681108;dist=82869 | NRXN1 | 0.0114 | 0.0171 | 0.892953 | 1 | Neurexin 1 |
| 2:74482548:G:T | 2 | 74482548 | rs12621729 | chr2 | 74255421 | 0.03220874 | 7 | 1.780987998 | 0.37313777 | 4.77300378 | 1.81498E-06 | intronic | SLC4A5 | . | SLC4A5 | 0.1479 | 0.0335 | 0.942493 | 1 | Solute Carrier Family 4 Member 5 (sodium bicarbonate transporter) |
| 2:126913427:C:G | 2 | 126913427 | rs28798228 | chr2 | 126155850 | 0.03317476 | 7 | 1.808450116 | 0.37851254 | 4.777781303 | 1.7724E-06 | intergenic | CNTNAP5;GYPC | dist=1240473;dist=500084 | GYPC | 0.0111 | 0.0144 | 0.961063 | 1 | Glycophorin C (Gerbich Blood Group) |
| 2:126955214:G:A | 2 | 126955214 | rs28796094 | chr2 | 126197637 | 0.03244175 | 7 | 1.7618909 | 0.37073145 | 4.752472211 | 2.00944E-06 | intergenic | CNTNAP5;GYPC | dist=1282260;dist=458297 | | 0.0111 | 0.0144 | 0.939443 | 1 | |
| 2:134644599:A:G | 2 | 134644599 | rs189440995 | chr2 | 133887028 | 0.01478641 | 3 | 1.137571373 | 0.24878258 | 4.572552272 | 4.81819E-06 | intergenic | NCKAP5;MIR3679 | dist=318568;dist=240097 | NCKAP5 | 0.0060 | 0.0093 | 0.899627 | 1 | NCK Associated Protein 5 |
| 3:75482532:G:C | 3 | 75482532 | rs185293133 | chr3 | 75433381 | 0.01162621 | 2 | 0.80016771 | 0.15901908 | 5.031897471 | 4.85649E-07 | ncRNA_intronic | FAM86DP | . | | 0.0054 | 0.0078 | 0.492536 | 1 | |
| 3:146048618:G:A | 3 | 146048618 | rs73150883 | chr3 | 146330831 | 0.02381553 | 5 | 1.481987222 | 0.28774985 | 5.150262284 | 2.60122E-07 | intergenic | PLSCR4;PLSCR2 | dist=79652;dist=102457 | PLSCR4;PLSCR2 | 0.0163 | 0.0252 | 0.848249 | 1 | Phospholipid Scramblase 4;Phospholipid Scramblase 2 |
| 3:196625842:T:C | 3 | 196625842 | rs6583185 | chr3 | 196898971 | 0.97179126 | 6 | -1.52139796 | 0.33118931 | -4.593741177 | 4.35369E-06 | intronic | SENP5 | . | SENP5 | 0.9845 | 0.9781 | 0.853106 | 1 | SUMO Specific Peptidase 5 |
| 4:125382184:C:G | 4 | 125382184 | rs72678487 | chr4 | 124461029 | 0.01945631 | 4 | 1.343334873 | 0.29403913 | 4.568558189 | 4.91091E-06 | intergenic | LINC01091;LOC101927087 | dist=530666;dist=38913 | | 0.0073 | 0.0115 | 0.987952 | 1 | |
| 5:115976909:A:C | 5 | 115976909 | rs76433992 | chr5 | 116641213 | 0.02983495 | 6 | 1.764770451 | 0.36047037 | 4.895743415 | 9.79348E-07 | intergenic | SEMA6A;LOC102467223 | dist=66287;dist=102089 | SEMA6A | 0.0168 | 0.0226 | 0.974211 | 1 | Semaphorin 6A |
| 5:115979852:T:C | 5 | 115979852 | rs114241096 | chr5 | 116644156 | 0.02909223 | 6 | 1.782650366 | 0.3612333 | 4.934900476 | 8.01916E-07 | intergenic | SEMA6A;LOC102467223 | dist=69230;dist=99146 | | 0.0185 | 0.0229 | 0.998458 | 1 | |
| 5:115980038:G:C | 5 | 115980038 | rs114587121 | chr5 | 116644342 | 0.02909223 | 6 | 1.782650366 | 0.3612333 | 4.934900476 | 8.01916E-07 | intergenic | SEMA6A;LOC102467223 | dist=69416;dist=98960 | | 0.0185 | 0.0229 | 0.998458 | 1 | |
| 5:115980724:A:G | 5 | 115980724 | rs79516960 | chr5 | 116645028 | 0.02909709 | 6 | 1.782403345 | 0.36123109 | 4.934246796 | 8.04606E-07 | intergenic | SEMA6A;LOC102467223 | dist=70102;dist=98274 | | 0.0171 | 0.0229 | 0.998286 | 1 | |
| 5:115980901:A:C | 5 | 115980901 | rs78085382 | chr5 | 116645205 | 0.02909709 | 6 | 1.782403345 | 0.36123109 | 4.934246796 | 8.04606E-07 | intergenic | SEMA6A;LOC102467223 | dist=70279;dist=98097 | | 0.0186 | 0.0228 | 0.998286 | 1 | |
| 5:115981236:C:G | 5 | 115981236 | rs79794218 | chr5 | 116645540 | 0.02909709 | 6 | 1.782403345 | 0.36123109 | 4.934246796 | 8.04606E-07 | intergenic | SEMA6A;LOC102467223 | dist=70614;dist=97762 | | 0.0170 | 0.0229 | 0.998286 | 1 | |
| 5:115981990:C:G | 5 | 115981990 | rs114108437 | chr5 | 116646294 | 0.02909709 | 6 | 1.782403345 | 0.36123109 | 4.934246796 | 8.04606E-07 | intergenic | SEMA6A;LOC102467223 | dist=71368;dist=97008 | | 0.0170 | 0.0229 | 0.998286 | 1 | |
| 5:115988180:G:A | 5 | 115988180 | rs77857559 | chr5 | 116652484 | 0.02912621 | 6 | 1.784300998 | 0.36172467 | 4.932759998 | 8.10758E-07 | intergenic | SEMA6A;LOC102467223 | dist=77558;dist=90818 | | 0.0165 | 0.0221 | 1 | 1 | |
| 5:115988843:C:T | 5 | 115988843 | rs115917626 | chr5 | 116653147 | 0.02912621 | 6 | 1.784300998 | 0.36172467 | 4.932759998 | 8.10758E-07 | intergenic | SEMA6A;LOC102467223 | dist=78221;dist=90155 | | 0.0167 | 0.0223 | 1 | 1 | |
| 5:115991913:A:T | 5 | 115991913 | rs76006987 | chr5 | 116656217 | 0.02908738 | 6 | 1.782276135 | 0.36123386 | 4.933856736 | 8.06216E-07 | intergenic | SEMA6A;LOC102467223 | dist=81291;dist=87085 | | 0.0185 | 0.0225 | 0.998629 | 1 | |
| 5:162496418:T:C | 5 | 162496418 | rs114500719 | chr5 | 163069412 | 0.0283301 | 6 | 1.557183414 | 0.34021336 | 4.577078964 | 4.71514E-06 | intergenic | GABRG2;CCNG1 | dist=913873;dist=368159 | GABRG2 | 0.0243 | 0.0327 | 0.909911 | 1 | Gamma-Aminobutyric Acid Type A Receptor Subunit Gamma2 |
| 5:162629778:G:A | 5 | 162629778 | rs116189826 | chr5 | 163202772 | 0.02453883 | 5 | 1.5321512 | 0.32364261 | 4.734083693 | 2.20047E-06 | intergenic | GABRG2;CCNG1 | dist=1047233;dist=234799 | CCNG1 | 0.0218 | 0.0286 | 0.937899 | 1 | Cyclin G1 |
| 5:162646718:G:C | 5 | 162646718 | rs115331238 | chr5 | 163219712 | 0.02446116 | 5 | 1.583713858 | 0.33185801 | 4.772263512 | 1.82167E-06 | intergenic | GABRG2;CCNG1 | dist=1064173;dist=217859 | | 0.0236 | 0.0306 | 0.983743 | 1 | |
| 5:162653096:T:C | 5 | 162653096 | rs116626718 | chr5 | 163226090 | 0.02438835 | 5 | 1.593867941 | 0.33347822 | 4.779526333 | 1.75709E-06 | intergenic | GABRG2;CCNG1 | dist=1070551;dist=211481 | | 0.0249 | 0.0325 | 0.995221 | 1 | |
| 5:162661735:C:T | 5 | 162661735 | rs114008181 | chr5 | 163234729 | 0.02521359 | 5 | 1.5722456 | 0.33187599 | 4.73744913 | 2.16425E-06 | intergenic | GABRG2;CCNG1 | dist=1079190;dist=202842 | | 0.0256 | 0.0337 | 0.958521 | 1 | |
| 6:31238068:C:T | 6 | 31238068 | rs1050276 | chr6 | 31270291 | 0.02443204 | 5 | 1.532787887 | 0.3329803 | 4.603238922 | 4.15971E-06 | exonic | HLA-C | . | | 0.0426 | 0.0400 | 0.991508 | 1 | |
| 6:31238801:C:G | 6 | 31238801 | rs41544614 | chr6 | 31271024 | 0.02434951 | 5 | 1.533218592 | 0.33286996 | 4.606058777 | 4.10372E-06 | intronic | HLA-C | . | | 0.0428 | 0.0404 | 0.994693 | 1 | |
| 6:31239727:C:A | 6 | 31239727 | rs29029490 | chr6 | 31271950 | 0.02433495 | 5 | 1.529466801 | 0.33227278 | 4.603045682 | 4.16357E-06 | intronic | HLA-C | . | | 0.0429 | 0.0404 | 0.991645 | 1 | |
| 6:31240096:G:A | 6 | 31240096 | rs9366775 | chr6 | 31272319 | 0.02427184 | 5 | 1.534745688 | 0.33329599 | 4.604752947 | 4.12956E-06 | upstream | HLA-C | dist=183 | | 0.0429 | 0.0404 | 1 | 1 | |
| 6:31240479:T:G | 6 | 31240479 | rs9357121 | chr6 | 31272702 | 0.02427184 | 5 | 1.534745688 | 0.33329599 | 4.604752947 | 4.12956E-06 | upstream | HLA-C | dist=566 | | 0.0431 | 0.0406 | 1 | 1 | |
| 6:31240818:G:A | 6 | 31240818 | rs4298345 | chr6 | 31273041 | 0.02420388 | 5 | 1.529478709 | 0.33238499 | 4.601527555 | 4.19404E-06 | upstream | HLA-C | dist=905 | | 0.0429 | 0.0404 | 0.997144 | 1 | |
| 6:31245080:G:A | 6 | 31245080 | rs9391714 | chr6 | 31277303 | 0.02427184 | 5 | 1.534745688 | 0.33329599 | 4.604752947 | 4.12956E-06 | intergenic | HLA-C;HLA-B | dist=5167;dist=76569 | HLA-C | 0.0430 | 0.0403 | 1 | 1 | Major Histocompatibility Complex, Class I, C |
| 6:31247267:C:T | 6 | 31247267 | rs56356836 | chr6 | 31279490 | 0.02423786 | 5 | 1.532381559 | 0.33283392 | 4.60404267 | 4.14368E-06 | intergenic | HLA-C;HLA-B | dist=7354;dist=74382 | HLA-B | 0.0429 | 0.0402 | 0.998566 | 1 | Major Histocompatibility Complex, Class I, B |
| 6:31247998:T:C | 6 | 31247998 | rs9405016 | chr6 | 31280221 | 0.02423786 | 5 | 1.532381559 | 0.33283392 | 4.60404267 | 4.14368E-06 | intergenic | HLA-C;HLA-B | dist=8085;dist=73651 | | 0.0430 | 0.0403 | 0.998566 | 1 | |
| 6:31248262:G:A | 6 | 31248262 | rs12529015 | chr6 | 31280485 | 0.02423786 | 5 | 1.532381559 | 0.33283392 | 4.60404267 | 4.14368E-06 | intergenic | HLA-C;HLA-B | dist=8349;dist=73387 | | 0.0429 | 0.0401 | 0.998566 | 1 | |
| 6:31248493:T:C | 6 | 31248493 | rs9368669 | chr6 | 31280716 | 0.02423786 | 5 | 1.532381559 | 0.33283392 | 4.60404267 | 4.14368E-06 | intergenic | HLA-C;HLA-B | dist=8580;dist=73156 | | 0.0429 | 0.0402 | 0.998566 | 1 | |
| 6:31248568:G:A | 6 | 31248568 | rs9380234 | chr6 | 31280791 | 0.02430583 | 5 | 1.53280058 | 0.33276482 | 4.606257864 | 4.0998E-06 | intergenic | HLA-C;HLA-B | dist=8655;dist=73081 | | 0.0429 | 0.0402 | 0.995714 | 1 | |
| 6:31256026:C:G | 6 | 31256026 | rs17198734 | chr6 | 31288249 | 0.02423786 | 5 | 1.532381559 | 0.33283392 | 4.60404267 | 4.14368E-06 | intergenic | HLA-C;HLA-B | dist=16113;dist=65623 | | 0.0253 | 0.0303 | 0.998566 | 1 | |
| 6:31256058:G:T | 6 | 31256058 | rs17192386 | chr6 | 31288281 | 0.02423786 | 5 | 1.532381559 | 0.33283392 | 4.60404267 | 4.14368E-06 | intergenic | HLA-C;HLA-B | dist=16145;dist=65591 | | 0.0358 | 0.0305 | 0.998566 | 1 | |
| 6:126490324:C:T | 6 | 126490324 | rs148294287 | chr6 | 126169178 | 0.01691748 | 3 | 1.203746268 | 0.25927127 | 4.642806213 | 3.43709E-06 | intergenic | MIR5695;CENPW | dist=46562;dist=170611 | CENPW | 0.0107 | 0.0165 | 0.861892 | 1 | Centromere Protein W |
| 6:158989974:G:A | 6 | 158989974 | rs142006494 | chr6 | 158568942 | 0.01558252 | 3 | 1.102833648 | 0.23946303 | 4.60544434 | 4.11586E-06 | intronic | TMEM181 | . | TMEM181 | 0.0339 | 0.0408 | 0.909469 | 1 | Transmembrane Protein 181 |
| 7:9489611:T:G | 7 | 9489611 | rs17206867 | chr7 | 9449981 | 0.01978155 | 4 | 1.317750936 | 0.27773373 | 4.744655705 | 2.08861E-06 | intergenic | NXPH1;PER4 | dist=697018;dist=184289 | | 0.0067 | 0.0108 | 0.886061 | 1 | |
| 8:23462708:A:G | 8 | 23462708 | rs539324287 | chr8 | 23605195 | 0.01694175 | 3 | 1.341800652 | 0.26460113 | 5.071031363 | 3.95666E-07 | intergenic | SLC25A37;NKX3-1 | dist=32645;dist=73498 | SLC25A37 | 0.0033 | 0.0057 | 0.907302 | 1 | Solute Carrier Family 25 Member 37 (mitochondrial) |
| 8:23542722:T:A | 8 | 23542722 | rs189490695 | chr8 | 23685209 | 0.0163835 | 3 | 1.2982161 | 0.2556875 | 5.077354608 | 3.82726E-07 | intergenic | NKX3-1;NKX2-6 | dist=2272;dist=17242 | NKX3-1 | 0.0070 | 0.0092 | 0.874937 | 1 | NK3 Homeobox 1 |
| 8:133999440:A:C | 8 | 133999440 | rs74591804 | chr8 | 132987195 | 0.01455825 | 3 | 1.200642195 | 0.25459157 | 4.715954269 | 2.4058E-06 | intronic | TG | . | TG | 0.0132 | 0.0217 | 0.999662 | 1 | Thyroglobulin |
| 10:72790290:G:A | 10 | 72790290 | rs77364739 | chr10 | 71030533 | 0.03883495 | 8 | 1.881493073 | 0.41157336 | 4.571464641 | 4.84327E-06 | intergenic | PCBD1;UNC5B | dist=141747;dist=182002 | PCBD1;UNC5B | 0.0865 | 0.0588 | 1 | 1 | Pterin-4 Alpha-Carbinolamine Dehydratase 1;Unc-5 Netrin Receptor |
| 12:11046142:C:A | 12 | 11046142 | rs111621289 | chr12 | 10893543 | 0.03042233 | 6 | 1.581110295 | 0.33666074 | 4.696449854 | 2.64722E-06 | ncRNA_intronic | PRH1-PRR4 | . | | 0.0242 | 0.0300 | 0.810976 | 1 | |
| 13:30768424:G:A | 13 | 30768424 | rs74833295 | chr13 | 30194287 | 0.02916505 | 6 | 1.909228275 | 0.36002172 | 5.303091934 | 1.13858E-07 | intergenic | LINC00365;KATNAL1 | dist=85412;dist=8343 | KATNAL1 | 0.0153 | 0.0232 | 0.9983 | 1 | Katanin Catalytic Subunit A1 Like 1 |
| 13:52703883:C:G | 13 | 52703883 | rs148654242 | chr13 | 52129747 | 0.03606311 | 7 | 1.896523445 | 0.38630706 | 4.90936778 | 9.13705E-07 | ncRNA_exonic | LOC101929657 | . | | 0.0202 | 0.0302 | 0.890541 | 1 | |
| 15:27633984:A:C | 15 | 27633984 | rs72705708 | chr15 | 27388838 | 0.06847087 | 14 | 2.432058671 | 0.51344077 | 4.736785214 | 2.17135E-06 | intronic | GABRG3 | . | GABRG3 | 0.0450 | 0.0491 | 0.990527 | 1 | Gamma-Aminobutyric Acid Type A Receptor Subunit Gamma3 |
| 15:27634215:C:T | 15 | 27634215 | rs72705709 | chr15 | 27389069 | 0.06834952 | 14 | 2.429618929 | 0.5133886 | 4.732514355 | 2.21756E-06 | intronic | GABRG3 | . | | 0.0448 | 0.0489 | 0.992399 | 1 | |
| 15:27634405:G:C | 15 | 27634405 | rs72705710 | chr15 | 27389259 | 0.06832039 | 14 | 2.428373305 | 0.51325152 | 4.731351398 | 2.2303E-06 | intronic | GABRG3 | . | | 0.0448 | 0.0489 | 0.992701 | 1 | |
| 15:27635356:C:G | 15 | 27635356 | rs72705711 | chr15 | 27390210 | 0.06797087 | 14 | 2.443670474 | 0.51382691 | 4.75582426 | 1.97638E-06 | intronic | GABRG3 | . | | 0.0601 | 0.0498 | 0.997858 | 1 | |
| 15:27636387:G:A | 15 | 27636387 | rs72705712 | chr15 | 27391241 | 0.06796117 | 14 | 2.444243265 | 0.51389229 | 4.756333777 | 1.9714E-06 | intronic | GABRG3 | . | | 0.0463 | 0.0496 | 0.998164 | 1 | |
| 15:27636953:T:G | 15 | 27636953 | rs72705714 | chr15 | 27391807 | 0.06796117 | 14 | 2.448622014 | 0.51441141 | 4.760046044 | 1.93549E-06 | intronic | GABRG3 | . | | 0.0522 | 0.0496 | 1 | 1 | |
| 15:27637194:T:C | 15 | 27637194 | rs72705715 | chr15 | 27392048 | 0.06796117 | 14 | 2.444243265 | 0.51389229 | 4.756333777 | 1.9714E-06 | intronic | GABRG3 | . | | 0.0428 | 0.0496 | 0.998164 | 1 | |
| 15:31383118:A:C | 15 | 31383118 | rs75968509 | chr15 | 31090915 | 0.01935922 | 4 | 1.431828899 | 0.30212194 | 4.739241691 | 2.1452E-06 | intronic | TRPM1 | . | TRPM1 | 0.0124 | 0.0149 | 0.979554 | 1 | Transient Receptor Potential Cation Channel Subfamily M Member |
| 17:75157900:C:T | 17 | 75157900 | rs113199364 | chr17 | 77161818 | 0.03330097 | 7 | 1.815437814 | 0.35840665 | 5.065301631 | 4.07754E-07 | intronic | SEC14L1 | . | SEC14L1 | 0.0230 | 0.0364 | 0.893794 | 1 | SEC14 Like Lipid Binding 1 |
| 18:6045905:T:A | 18 | 6045905 | rs74649788 | chr18 | 6045906 | 0.0498932 | 10 | 2.439599988 | 0.44900137 | 5.43339095 | 5.52931E-08 | intronic | L3MBTL4 | . | L3MBTL4 | 0.0811 | 0.0475 | 0.923243 | 1 | L3MBTL Histone Methyl-Lysine Binding Protein 4 |
| 18:6049026:T:A | 18 | 6049026 | rs79381542 | chr18 | 6049027 | 0.04435922 | 9 | 2.087445553 | 0.41784921 | 4.995691057 | 5.86254E-07 | intronic | L3MBTL4 | . | | 0.0807 | 0.0470 | 0.959092 | 1 | |
| 18:6049551:C:A | 18 | 6049551 | rs58503971 | chr18 | 6049552 | 0.04434952 | 9 | 2.087223432 | 0.41779888 | 4.995761149 | 5.86042E-07 | intronic | L3MBTL4 | . | | 0.0786 | 0.0441 | 0.959099 | 1 | |
| | | | | | | | | | | | | | 25 risk loci | | | | | | | |
| | | | | | | | | | | | | | one overlaps with 3 month: PCBD1;UNC5B | | | | | | | |
| | | | | | | | | | | | | | Thus 57+25-1 = 81 total risk loci | | | | | | | |
| | | | | | | | | | | | | | Gray cells = risk loci overlapping with Indianapolis-1 GWAS; 4/: | 16.00% | | | | | | |

**Supplementary Table S8. GWAS Results Indianapolis-2 Replication Cohort**
**3 month quantitative trait (QT), risk locus ordered**

Shaded cells in Gene.refGene column indicate risk loci that co-localize with risk loci identified in the Indianapolis-1 GWAS

| ID_37 | chr_37 | POS_37 | rsID | chr_38 | POS_38 | freq | MAC | Score | Score.SE | Score.Stat | Score.pval | Func.refGene | Gene.refGene | GeneDetail.refGene | AOP-expressed refGene (protein coding only) | gnomAD_genome_ALL | gnomAD_genome_NFE | Rsq | hwe | AOP-expressed gene name |
|---|---|---|---|---|---|---|---|---|---|---|---|---|---|---|---|---|---|---|---|---|
| 1:12215283:T:C | 1 | 12215283 | rs147985761 | chr1 | 12155226 | 0.013373786 | 3 | 1.580945998 | 0.324859599 | 4.866551587 | 1.13562E-06 | intergenic | TNFRSF8;MIR7846 | dist=11019;dist=11717 | TNFRSF8 | 0.0081 | 0.0132 | 0.894028 | 1 | TNF Receptor Superfamily Member 8 |
| 1:18614493:G:T | 1 | 18614493 | rs115263712 | chr1 | 18287999 | 0.014364078 | 3 | 1.46894969 | 0.320848564 | 4.578327141 | 4.68709E-06 | intronic | IGSF21 | . | IGSF21 | 0.0101 | 0.0137 | 0.8337 | 1 | Immunoglobin Superfamily Member 21 |
| 1:53720723:T:C | 1 | 53720723 | rs12116501 | chr1 | 53255051 | 0.040257281 | 8 | 2.387313586 | 0.485212014 | 4.920145248 | 8.648E-07 | intronic | LRP8 | . | LRP8 | 0.0311 | 0.0437 | 0.807097 | 1 | LDL Receptor Related Protein 8 |
| 1:70886329:A:G | 1 | 70886329 | rs79934683 | chr1 | 70420646 | 0.038961165 | 8 | 2.731688539 | 0.567885471 | 4.81028073 | 1.50718E-06 | intronic | CTH | . | CTH | 0.0306 | 0.0416 | 0.993133 | 1 | Cystathionine Gamma-Lyase |
| 1:240988921:C:T | 1 | 240988921 | rs72754888 | chr1 | 240825621 | 0.020087379 | 4 | 1.824932693 | 0.38452179 | 4.74597992 | 2.07499E-06 | intronic | RGS7 | . | RGS7 | 0.0047 | 0.0067 | 0.836433 | 1 | Regulator Of G Protein Signaling 7 |
| 1:240998165:G:A | 1 | 240998165 | rs72754890 | chr1 | 240834865 | 0.019203883 | 4 | 1.815512107 | 0.369826911 | 4.909085989 | 9.15019E-07 | intronic | RGS7 | . | | 0.0051 | 0.0073 | 0.806371 | 1 | |
| 2:66301595:G:A | 2 | 66301595 | rs11676376 | chr2 | 66074461 | 0.024257282 | 5 | 2.107857413 | 0.458486846 | 4.597421782 | 4.27751E-06 | intergenic | SPRED2;MIR4778 | dist=641939;dist=283786 | SPRED2 | 0.0435 | 0.0423 | 0.998566 | 1 | Sprouty Related EVH1 Domain Containing 2 |
| 2:72629234:T:G | 2 | 72629234 | rs185879164 | chr2 | 72402105 | 0.018174757 | 4 | 1.80424283 | 0.387623961 | 4.654621515 | 3.24576E-06 | intronic | EXOC6B | . | EXOC6B | 0.0118 | 0.0192 | 0.929494 | 1 | Exocyst Complex Component 6B (exocytosis) |
| 2:72852782:C:A | 2 | 72852782 | rs115540332 | chr2 | 72625653 | 0.018898058 | 4 | 1.818657772 | 0.391871559 | 4.640953721 | 3.46805E-06 | intronic | EXOC6B | . | | 0.0116 | 0.0188 | 0.914669 | 1 | |
| 2:117348669:G:A | 2 | 117348669 | rs75685045 | chr2 | 116591093 | 0.020179612 | 4 | 2.005609812 | 0.414444146 | 4.839276486 | 1.30313E-06 | intergenic | DPP10;DDX18 | dist=746343;dist=1223586 | DPP10;DDX18 | 0.0065 | 0.0111 | 0.944983 | 1 | Dipeptidyl Peptidase Like 10 (modifies KCND3); DEAD-Box Helicase 18 |
| 2:130563530:G:A | 2 | 130563530 | rs4337430 | chr2 | 129805957 | 0.248378641 | 51 | 6.844310493 | 1.325444312 | 5.163785781 | 2.42005E-07 | intergenic | LOC151121;LOC389033 | dist=532066;dist=116905 | | 0.2131 | 0.2653 | 0.973034 | 0.279636 | |
| 2:130565338:C:A | 2 | 130565338 | rs34017523 | chr2 | 129807765 | 0.253538835 | 52 | 6.70870463 | 1.334874447 | 5.025719568 | 5.01548E-07 | intergenic | LOC151121;LOC389033 | dist=533874;dist=115097 | | 0.2123 | 0.2644 | 0.978903 | 0.420041 | |
| 2:130567425:T:A | 2 | 130567425 | rs6720381 | chr2 | 129809852 | 0.253538835 | 52 | 6.70870463 | 1.334874447 | 5.025719568 | 5.01548E-07 | intergenic | LOC151121;LOC389033 | dist=535961;dist=113010 | | 0.2065 | 0.2652 | 0.978903 | 0.420041 | |
| 2:130568556:T:C | 2 | 130568556 | rs4355062 | chr2 | 129810983 | 0.297684466 | 61 | 6.436745057 | 1.381972449 | 4.657650781 | 3.19838E-06 | intergenic | LOC151121;LOC389033 | dist=537092;dist=111879 | | 0.2389 | 0.2895 | 0.981992 | 0.478458 | |
| 2:130573252:C:A | 2 | 130573252 | rs34895266 | chr2 | 129815679 | 0.254174757 | 52 | 6.714995733 | 1.337157087 | 5.021845077 | 5.11775E-07 | intergenic | LOC151121;LOC389033 | dist=541788;dist=107183 | | 0.2140 | 0.2654 | 0.980963 | 0.420041 | |
| 2:130582408:G:C | 2 | 130582408 | rs12621133 | chr2 | 129824835 | 0.264009709 | 54 | 6.522687428 | 1.33928955 | 4.87025933 | 1.11452E-06 | intergenic | LOC151121;LOC389033 | dist=550944;dist=98027 | | 0.2291 | 0.2789 | 0.979973 | 0.601073 | |
| 2:130584142:C:T | 2 | 130584142 | rs35748618 | chr2 | 129826569 | 0.222898058 | 46 | 6.124040336 | 1.238717369 | 4.943856031 | 7.65923E-07 | intergenic | LOC151121;LOC389033 | dist=552678;dist=96293 | | 0.1481 | 0.2177 | 0.96226 | 1 | |
| 2:130586642:C:T | 2 | 130586642 | rs4277471 | chr2 | 129829069 | 0.257281553 | 53 | 6.683699411 | 1.358232147 | 4.920881476 | 8.61553E-07 | intergenic | LOC151121;LOC389033 | dist=555178;dist=93793 | | 0.1953 | 0.2581 | 1 | 0.300358 | |
| 2:130589408:A:T | 2 | 130589408 | rs1882610 | chr2 | 129831835 | 0.263592233 | 54 | 6.506401141 | 1.335621499 | 4.871440856 | 1.10787E-06 | intergenic | LOC151121;LOC389033 | dist=557944;dist=91027 | | 0.2367 | 0.2795 | 0.977378 | 0.601073 | |
| 2:130592585:C:T | 2 | 130592585 | rs6742395 | chr2 | 129835012 | 0.258529127 | 53 | 6.558142102 | 1.329599407 | 4.932419546 | 8.12172E-07 | intergenic | LOC151121;LOC389033 | dist=561121;dist=87850 | | 0.2225 | 0.2681 | 0.976222 | 0.430075 | |
| 2:151347581:A:G | 2 | 151347581 | rs145531574 | chr2 | 150491067 | 0.013723301 | 3 | 1.731904782 | 0.335798226 | 5.157575737 | 2.50168E-07 | intergenic | RND3;LOC101929260 | dist=3372;dist=61465 | RND3 | 0.0088 | 0.0141 | 0.8698 | 1 | Rho Family GTPase 3 |
| 3:64559845:T:C | 3 | 64559845 | rs146268777 | chr3 | 64574169 | 0.014699029 | 3 | 1.72126431 | 0.347407762 | 4.954593705 | 7.24816E-07 | ncRNA_intronic | ADAMTS9-AS1 | . | ADAMTS9 | 0.0046 | 0.0068 | 0.94425 | 1 | ADAMTS9 |
| 4:31766001:G:A | 4 | 31766001 | rs16885796 | chr4 | 31764379 | 0.053223301 | 11 | 3.28160902 | 0.713701131 | 4.598015718 | 4.26534E-06 | intergenic | LOC102723778;LOC102723828 | dist=552705;dist=233000 | | 0.0382 | 0.0334 | 0.994282 | 1 | |
| 4:31772884:C:T | 4 | 31772884 | rs116957173 | chr4 | 31771262 | 0.053398058 | 11 | 3.290738644 | 0.717096412 | 4.588976583 | 4.45424E-06 | intergenic | LOC102723778;LOC102723828 | dist=559588;dist=226117 | | 0.0270 | 0.0312 | 1 | 0.245321 | |
| 4:31777594:G:A | 4 | 31777594 | rs116168704 | chr4 | 31775972 | 0.053165049 | 11 | 3.264991759 | 0.71189775 | 4.586321223 | 4.51124E-06 | intergenic | LOC102723778;LOC102723828 | dist=564298;dist=221407 | | 0.0285 | 0.0314 | 0.991203 | 1 | |
| 4:31779863:T:C | 4 | 31779863 | rs115916744 | chr4 | 31778241 | 0.053402913 | 11 | 3.29083276 | 0.717091233 | 4.589140975 | 4.45074E-06 | intergenic | LOC102723778;LOC102723828 | dist=566567;dist=219138 | | 0.0283 | 0.0312 | 0.999904 | 0.247516 | |
| 4:31782754:A:G | 4 | 31782754 | rs79231845 | chr4 | 31781132 | 0.053990291 | 11 | 3.25806796 | 0.70424346 | 4.626337545 | 3.72189E-06 | intergenic | LOC102723778;LOC102723828 | dist=569458;dist=216247 | | 0.0271 | 0.0314 | 0.964283 | 1 | |
| 4:40948164:G:A | 4 | 40948164 | rs113189296 | chr4 | 40946147 | 0.037621359 | 8 | 2.919015192 | 0.615000485 | 4.746362423 | 2.07107E-06 | intronic | APBB2 | . | APBB2 | 0.0591 | 0.0852 | 0.970482 | 1 | Amyloid Beta Precursor Protein Binding Family B Member 2 |
| 4:163976364:T:C | 4 | 163976364 | rs146154846 | chr4 | 163055212 | 0.028898058 | 6 | 2.023347127 | 0.417548067 | 4.845782525 | 1.26114E-06 | intergenic | FSTL5;MIR4454 | dist=891178;dist=38362 | FSTL5 | 0.0182 | 0.0245 | 0.711952 | 1 | Follistatin-like 5 |
| 4:164000229:C:T | 4 | 164000229 | rs145034465 | chr4 | 163079077 | 0.02361165 | 5 | 1.814612257 | 0.387671579 | 4.680797751 | 2.85761E-06 | intergenic | FSTL5;MIR4454 | dist=915043;dist=14497 | | 0.0184 | 0.0231 | 0.732815 | 1 | |
| 4:173193727:G:A | 4 | 173193727 | rs187567621 | chr4 | 172272576 | 0.014470874 | 3 | 1.644356357 | 0.349394212 | 4.706306808 | 2.52245E-06 | intronic | GALNTL6 | . | GALNTL6 | 0.0019 | 0.0033 | 0.94795 | 1 | Polypeptide N-Acetylgalactosaminyltransferase Like 6 |
| 4:189389927:T:C | 4 | 189389927 | rs544973300 | chr4 | 188468773 | 0.010276699 | 2 | 1.359541791 | 0.297112351 | 4.575850811 | 4.74288E-06 | ncRNA_intronic | LINC01060 | . | | 0.0035 | 0.0050 | 0.946406 | 1 | |
| 4:189389929:T:C | 4 | 189389929 | rs560030667 | chr4 | 188468775 | 0.010296117 | 2 | 1.359541054 | 0.297050321 | 4.576803849 | 4.72134E-06 | ncRNA_intronic | LINC01060 | . | | 0.0037 | 0.0047 | 0.944465 | 1 | |
| 5:3562095:T:A | 5 | 3562095 | rs143503046 | chr5 | 3561981 | 0.00734466 | 2 | 0.983175268 | 0.214354222 | 4.58668487 | 4.5034E-06 | intergenic | LINC01019;IRX1 | dist=25887;dist=34073 | IRX1 | 0.0027 | 0.0041 | 0.686588 | 1 | Iroquois Homeobox 1 |
| 5:5233724:G:C | 5 | 5233724 | rs115149137 | chr5 | 5233611 | 0.014830097 | 3 | 1.706394641 | 0.358423256 | 4.760836832 | 1.92792E-06 | intronic | ADAMTS16 | . | | 0.0104 | 0.0175 | 0.967403 | 1 | |
| 5:5241203:C:A | 5 | 5241203 | rs145155685 | chr5 | 5241090 | 0.014868932 | 3 | 1.68070787 | 0.35442199 | 4.742109459 | 2.11504E-06 | intronic | ADAMTS16 | . | | 0.0080 | 0.0129 | 0.944326 | 1 | |
| 5:33636035:T:C | 5 | 33636035 | rs13164825 | chr5 | 33635930 | 0.155339806 | 32 | 5.112931608 | 1.104845474 | 4.627734581 | 3.69687E-06 | intronic | ADAMTS12 | . | ADAMTS12 | 0.1577 | 0.1968 | 1 | 0.705556 | ADAMTS12 |
| 5:33644123:G:T | 5 | 33644123 | rs35388476 | chr5 | 33644018 | 0.15973301 | 33 | 4.946113148 | 1.073435357 | 4.60774197 | 4.07065E-06 | intronic | ADAMTS12 | . | | 0.1520 | 0.2154 | 0.992101 | 1 | |
| 5:33644614:T:C | 5 | 33644614 | rs67267661 | chr5 | 33644509 | 0.160194175 | 33 | 4.958247799 | 1.078142852 | 4.598878331 | 4.24772E-06 | intronic | ADAMTS12 | . | | 0.1519 | 0.2153 | 1 | 1 | |
| 5:33646456:G:C | 5 | 33646456 | rs10057508 | chr5 | 33646351 | 0.164631068 | 34 | 4.973632977 | 1.076119596 | 4.621821771 | 3.80385E-06 | intronic | ADAMTS12 | . | | 0.1583 | 0.2157 | 0.991904 | 1 | |
| 5:33647634:G:A | 5 | 33647634 | rs13176485 | chr5 | 33647529 | 0.159796116 | 33 | 4.948363002 | 1.073508457 | 4.609524006 | 4.03592E-06 | intronic | ADAMTS12 | . | | 0.1519 | 0.2149 | 0.991564 | 1 | |
| 5:53390040:G:C | 5 | 53390040 | rs42872 | chr5 | 54094210 | 0.306907767 | 63 | 6.436279804 | 1.389496475 | 4.632095094 | 3.61984E-06 | intronic | ARL15 | . | ARL15 | 0.2490 | 0.2995 | 0.980711 | 0.812933 | ARF Like GTPase 15 |
| 5:73263379:A:G | 5 | 73263379 | rs190652742 | chr5 | 73967554 | 0.024257281 | 5 | 2.247608363 | 0.454880737 | 4.941093741 | 7.76855E-07 | intergenic | ARHGEF28;LINC01335 | dist=25561;dist=338856 | ARHGEF28 | 0.0104 | 0.0094 | 0.999387 | 1 | Rho Guanine Nucleotide Exchange Factor 2 |
| 5:112013914:G:A | 5 | 112013914 | rs12153142 | chr5 | 112678217 | 0.980427185 | 4 | -1.381862916 | 0.302073383 | -4.574593434 | 4.77146E-06 | ncRNA_intronic | LOC102467216 | . | | 0.9143 | 0.9991 | 0.499216 | 1 | |
| 6:77414670:G:C | 6 | 77414670 | rs56232065 | chr6 | 76704953 | 0.019820388 | 4 | 1.925532437 | 0.417298507 | 4.614280674 | 3.94459E-06 | intergenic | IMPG1;HTR1B | dist=632275;dist=755895 | | 0.0103 | 0.0099 | 0.980026 | 1 | Interphotoreceptor Matrix Proteoglycan 1; 5-Hydroxytryptamine Receptor 1B (serat |
| 6:77420028:C:A | 6 | 77420028 | rs78549750 | chr6 | 76710311 | 0.019699029 | 4 | 1.920385778 | 0.415757928 | 4.618999777 | 3.85594E-06 | intergenic | IMPG1;HTR1B | dist=637633;dist=750537 | | 0.0160 | 0.0101 | 0.978334 | 1 | |
| 6:77429812:T:G | 6 | 77429812 | rs77029592 | chr6 | 76720095 | 0.019640777 | 4 | 1.923001054 | 0.415738055 | 4.625511258 | 3.73676E-06 | intergenic | IMPG1;HTR1B | dist=647417;dist=740753 | | 0.0083 | 0.0100 | 0.980847 | 1 | |
| 6:77460389:C:T | 6 | 77460389 | rs191563430 | chr6 | 76750672 | 0.019490291 | 4 | 1.930446837 | 0.415888081 | 4.641745998 | 3.45477E-06 | intergenic | IMPG1;HTR1B | dist=677994;dist=710176 | | 0.0071 | 0.0101 | 0.988316 | 1 | |
| 6:77472188:A:G | 6 | 77472188 | rs55850402 | chr6 | 76762471 | 0.019490291 | 4 | 1.93070443 | 0.415892439 | 4.642316741 | 3.44524E-06 | intergenic | IMPG1;HTR1B | dist=689793;dist=698377 | | 0.0070 | 0.0099 | 0.988316 | 1 | |
| 7:48396066:G:A | 7 | 48396066 | rs189164061 | chr7 | 48356469 | 0.014519418 | 3 | 1.695949216 | 0.358675926 | 4.728360877 | 2.2634E-06 | intronic | ABCA13 | . | ABCA13 | 0.0077 | 0.0119 | 0.996283 | 1 | ATP Binding Cassette Subfamily A Member 13 |
| 7:89801861:C:T | 7 | 89801861 | rs7800716 | chr7 | 90172547 | 0.017533981 | 4 | 1.680077446 | 0.353443095 | 4.753459521 | 1.99965E-06 | ncRNA_intronic | STEAP2-AS1 | . | STEAP2 | 0.0069 | 0.0101 | 0.830555 | 1 | STEAP2 Metalloreductase |
| 7:89853957:C:T | 7 | 89853957 | rs149916697 | chr7 | 90224643 | 0.018815534 | 4 | 1.879423001 | 0.384569904 | 4.887077696 | 1.02344E-06 | intronic | STEAP2 | . | | 0.0065 | 0.0095 | 0.928405 | 1 | |
| 7:89988476:T:C | 7 | 89988476 | rs79847565 | chr7 | 90359162 | 0.018800971 | 4 | 1.89404647 | 0.387759417 | 4.8845918 | 1.03643E-06 | intronic | GTPBP10 | . | GTPBP10 | 0.0069 | 0.0100 | 0.945469 | 1 | GTP Binding Protein 10 |
| 7:90013529:G:A | 7 | 90013529 | rs187685448 | chr7 | 90384215 | 0.018776699 | 4 | 1.910374378 | 0.392033885 | 4.872982793 | 1.09926E-06 | intronic | GTPBP10 | . | | 0.0067 | 0.0098 | 0.967919 | 1 | |
| 7:90047115:A:G | 7 | 90047115 | rs143425398 | chr7 | 90417801 | 0.018825242 | 4 | 1.913164032 | 0.392804365 | 4.870526408 | 1.11301E-06 | intergenic | CLDN12;CDK14 | dist=1847;dist=178561 | CDK14 | 0.0066 | 0.0097 | 0.970376 | 1 | Cyclin Dependent Kinase 14 |
| 7:90253061:G:A | 7 | 90253061 | rs150148213 | chr7 | 90623747 | 0.01892233 | 4 | 1.921233543 | 0.394605718 | 4.868742276 | 1.12311E-06 | intronic | CDK14 | . | | 0.0066 | 0.0097 | 0.973808 | 1 | |
| 8:39944783:C:T | 8 | 39944783 | rs145892996 | chr8 | 40087264 | 0.030776699 | 6 | 2.576605742 | 0.50654249 | 5.086652731 | 3.64438E-07 | intergenic | IDO2;C8orf4 | dist=70873;dist=66204 | C8orf4 | 0.0131 | 0.0201 | 0.884119 | 1 | TCIM - Transcriptional And Immune Response Regulator |
| 8:39944917:T:A | 8 | 39944917 | rs138697513 | chr8 | 40087398 | 0.030781554 | 6 | 2.577314922 | 0.506628696 | 5.087187008 | 3.63413E-07 | intergenic | IDO2;C8orf4 | dist=71007;dist=66070 | | 0.0130 | 0.0201 | 0.884291 | 1 | |
| 8:40027101:T:C | 8 | 40027101 | rs116259399 | chr8 | 40169582 | 0.04368932 | 9 | 2.810807743 | 0.588520937 | 4.776053945 | 1.78768E-06 | intergenic | C8orf4;ZMAT4 | dist=14274;dist=361010 | ZMAT4 | 0.0402 | 0.0401 | 1 | 1 | Zinc Finger Matrin-Type 4 |
| 8:40049692:T:G | 8 | 40049692 | rs150470417 | chr8 | 40192173 | 0.044912622 | 9 | 2.785140072 | 0.582245322 | 4.783447747 | 1.72314E-06 | intergenic | C8orf4;ZMAT4 | dist=36865;dist=338419 | | 0.0259 | 0.0383 | 0.960377 | 1 | |
| 9:36357606:C:A | 9 | 36357606 | rs4879986 | chr9 | 36357609 | 0.023582524 | 5 | 2.18417045 | 0.448849347 | 4.866154904 | 1.1379E-06 | intronic | RNF38 | . | RNF38 | 0.0112 | 0.0117 | 0.970686 | 1 | Ring Finger Protein 38 |
| 9:36360845:T:C | 9 | 36360845 | rs79319828 | chr9 | 36360848 | 0.024014563 | 5 | 2.210438405 | 0.456810098 | 4.838856266 | 1.30588E-06 | intronic | RNF38 | . | | 0.0116 | 0.0120 | 0.9887 | 1 | |
| 9:36381902:C:T | 9 | 36381902 | rs2248297 | chr9 | 36381905 | 0.97573301 | 5 | -2.242692534 | 0.461738278 | -4.857064356 | 1.19139E-06 | intronic | RNF38 | . | | 0.9692 | 0.9879 | 0.999795 | 1 | |
| 9:36386865:T:G | 9 | 36386865 | rs76774745 | chr9 | 36386868 | 0.024029126 | 5 | 2.210045419 | 0.456790677 | 4.838201677 | 1.31019E-06 | intronic | RNF38 | . | | 0.0117 | 0.0120 | 0.988086 | 1 | |
| 10:21749029:G:A | 10 | 21749029 | rs183818481 | chr10 | 21460100 | 0.011912621 | 2 | 1.22791161 | 0.263818829 | 4.654374424 | 3.24966E-06 | intergenic | NEBL-AS1;CASC10 | dist=285177;dist=34392 | NEBL | 0.0059 | 0.0081 | 0.674237 | 1 | Nebulette |
| 10:21883439:G:A | 10 | 21883439 | rs191906004 | chr10 | 21594510 | 0.015543689 | 3 | 1.645849191 | 0.333793955 | 4.93073396 | 8.19212E-07 | intronic | MLLT10 | . | MLLT10 | 0.0058 | 0.0077 | 0.843604 | 1 | MLLT10 Histone Lysine Methyltransferase DOT1L Cofactor |
| 10:21970382:G:A | 10 | 21970382 | rs188790792 | chr10 | 21681453 | 0.014849515 | 3 | 1.594035309 | 0.323294895 | 4.930592272 | 8.19807E-07 | intronic | MLLT10 | . | | 0.0052 | 0.0080 | 0.823859 | 1 | |
| 10:22068468:C:A | 10 | 22068468 | rs184920540 | chr10 | 21779539 | 0.011684466 | 2 | 1.295186101 | 0.281543412 | 4.600306895 | 4.21869E-06 | intronic | DNAJC1 | . | DNAJC1 | 0.0059 | 0.0079 | 0.788199 | 1 | DnaJ Heat Shock Protein Family (Hsp40) Member C1 |
| 10:22083617:A:G | 10 | 22083617 | rs181139900 | chr10 | 21794688 | 0.011519417 | 2 | 1.283845844 | 0.277566907 | 4.625356308 | 3.73955E-06 | intronic | DNAJC1 | . | | 0.0064 | 0.0085 | 0.775781 | 1 | |
| 10:22216057:G:C | 10 | 22216057 | rs45496395 | chr10 | 21927128 | 0.013985437 | 3 | 1.470044503 | 0.319890854 | 4.595456497 | 4.31803E-06 | intronic | DNAJC1 | . | | 0.0063 | 0.0084 | 0.8731 | 1 | |
| 10:72790290:G:A | 10 | 72790290 | rs77364739 | chr10 | 71030533 | 0.038834951 | 8 | 2.690625403 | 0.557664123 | 4.824813526 | 1.40134E-06 | intergenic | PCBD1;UNC5B | dist=141747;dist=182002 | PCBD1;UNC5B | 0.0865 | 0.0588 | 1 | 1 | Pterin-4 Alpha-Carbinolamine Dehydratase 1; Unc-5 Netrin Receptor B |
| 10:72806820:G:A | 10 | 72806820 | rs75505956 | chr10 | 71047063 | 0.036461165 | 8 | 2.445966686 | 0.523449162 | 4.672787474 | 2.97139E-06 | intergenic | PCBD1;UNC5B | dist=158277;dist=165472 | | 0.0366 | 0.0553 | 0.953209 | 1 | |
| 10:98558540:C:A | 10 | 98558540 | rs61856855 | chr10 | 96798783 | 0.013485437 | 3 | 1.29789842 | 0.276292266 | 4.697556103 | 2.63293E-06 | intergenic | PIK3AP1;MIR607 | dist=78261;dist=29886 | PIK3AP1 | 0.0113 | 0.0141 | 0.63836 | 1 | Phosphoinositide-3-Kinase Adaptor Protein 1; |
| 10:115449191:G:A | 10 | 115449191 | rs150977401 | chr10 | 113689432 | 0.01631068 | 3 | 1.860675373 | 0.358119043 | 5.195689556 | 2.03962E-07 | intronic | CASP7 | . | CASP7 | 0.0073 | 0.0111 | 0.870763 | 1 | Caspase 7 |
| 11:21775222:T:A | 11 | 21775222 | rs147118669 | chr11 | 21753676 | 0.012529126 | 3 | 1.341788897 | 0.291618921 | 4.601172279 | 4.2012E-06 | intergenic | NELL1;ANO5 | dist=177993;dist=439500 | NELL1;ANO5 | 0.0050 | 0.0085 | 0.79684 | 1 | Neural EGFL Like 1; Anoctamin 5 |
| 11:44073753:C:T | 11 | 44073753 | rs149030097 | chr11 | 44052203 | 0.01565534 | 3 | 1.691379055 | 0.360755196 | 4.688439897 | 2.75296E-06 | intronic | ACCSL | . | | 0.0134 | 0.0213 | 0.931546 | 1 | |

| | | | | | | | | | | | | | | | | | | | |
|---|---|---|---|---|---|---|---|---|---|---|---|---|---|---|---|---|---|---|---|
| 11:44085975:C:A | 11 | 44085975 | rs76143420 | chr11 | 44064425 | 0.014893204 | 3 | 1.699510358 | 0.360057984 | 4.720101852 | 2.35727E-06 | intergenic | ACCSL;ACCS | dist=4448;dist=1754 | ACCS | 0.0134 | 0.0215 | 0.971325 | 1 | 1-Aminocyclopropane-1-Carboxylate Synthase Homolog (Inactive) |
| 12:898065:G:T | 12 | 898065 | rs2014160 | chr12 | 788899 | 0.01511165 | 3 | 1.72959212 | 0.339602326 | 5.092992566 | 3.52456E-07 | intronic | WNK1 | . | WNK1 | 0.0005 | 0.0007 | 0.853667 | 1 | WNK Lysine Deficient Protein Kinase 1 |
| 12:62144998:G:A | 12 | 62144998 | rs76854078 | chr12 | 61751217 | 0.01723301 | 4 | 1.744080472 | 0.37159489 | 4.693499608 | 2.68571E-06 | intronic | FAM19A2 | . | FAM19A2 | 0.0089 | 0.0135 | 0.904535 | 1 | TAFA2 - TAFA Chemokine Like Family Member 2 |
| 12:78530991:C:T | 12 | 78530991 | rs61754236 | chr12 | 78137211 | 0.028985437 | 6 | 2.517383543 | 0.494187455 | 5.093985122 | 3.50614E-07 | exonic | NAV3 | . | NAV3 | 0.0085 | 0.0125 | 0.995116 | 1 | Neuron Navigator 3 |
| 12:78545400:G:A | 12 | 78545400 | rs140686116 | chr12 | 78151620 | 0.02904369 | 6 | 2.519911214 | 0.495194249 | 5.088732794 | 3.60464E-07 | intronic | NAV3 | . | | 0.0063 | 0.0103 | 0.997108 | 1 | |
| 13:61672508:T:C | 13 | 61672508 | rs147220861 | chr13 | 61098374 | 0.027004854 | 6 | 2.031117161 | 0.432525467 | 4.695948131 | 2.65373E-06 | intergenic | LINC00378;MIR3169 | dist=402574;dist=101424 | | 0.0080 | 0.0082 | 0.826016 | 1 | |
| 13:71228466:C:A | 13 | 71228466 | rs9592740 | chr13 | 70654334 | 0.066820388 | 14 | 3.693410575 | 0.700908722 | 5.269460143 | 1.36826E-07 | intergenic | ATXN8OS;LINC00348 | dist=514581;dist=360807 | | 0.0651 | 0.0859 | 0.979474 | 1 | |
| 13:71242428:T:G | 13 | 71242428 | rs9599709 | chr13 | 70668296 | 0.065 | 13 | 3.560256036 | 0.685315257 | 5.195063146 | 2.0465E-07 | intergenic | ATXN8OS;LINC00348 | dist=528543;dist=346845 | | 0.0654 | 0.0860 | 0.958512 | 1 | |
| 13:102628784:G:A | 13 | 102628784 | rs75197268 | chr13 | 101976434 | 0.00926699 | 2 | 1.077369422 | 0.228342774 | 4.718211147 | 2.37927E-06 | intronic | FGF14 | . | FGF14 | 0.0122 | 0.0178 | 0.53936 | 1 | Fibrobast Growth Factor 14 |
| 13:109580621:C:T | 13 | 109580621 | rs145442123 | chr13 | 108928273 | 0.022786408 | 5 | 2.002149238 | 0.426532153 | 4.694017139 | 2.67892E-06 | intronic | MYO16 | . | MYO16 | 0.0106 | 0.0163 | 0.930524 | 1 | Myosin XVI |
| 14:24001662:C:T | 14 | 24001662 | rs41307102 | chr14 | 23532453 | 0.024456311 | 5 | 1.768121176 | 0.369052815 | 4.790970568 | 1.65976E-06 | intronic | ZFHX2 | . | ZFHX2 | 0.0265 | 0.0375 | 0.636713 | 1 | Zinc Finger Homeobox 2 |
| 15:45198007:A:C | 15 | 45198007 | rs28631454 | chr15 | 44905809 | 0.048796117 | 10 | 2.632234639 | 0.565845842 | 4.651858229 | 3.28957E-06 | intergenic | TRIM69;C15orf43 | dist=137980;dist=50893 | TRIM69 | 0.3216 | 0.1096 | 0.89064 | 1 | Tripartite Motif Containing 69 |
| 15:61531695:C:G | 15 | 61531695 | rs143862437 | chr15 | 61239496 | 0.014752427 | 3 | 1.816785354 | 0.35534236 | 5.112774488 | 3.17461E-07 | intergenic | RORA;VPS13C | dist=10193;dist=612895 | RORA;VPS13C | 0.0083 | 0.0139 | 0.9521 | 1 | RAR Related Orphan Receptor A; Vacuolar Protein Sorting 13 Homolog C |
| 15:61534825:G:A | 15 | 61534825 | rs182462684 | chr15 | 61242626 | 0.014762136 | 3 | 1.816594932 | 0.35534099 | 5.112258325 | 3.1833E-07 | intergenic | RORA;VPS13C | dist=13323;dist=609765 | | 0.0084 | 0.0138 | 0.951481 | 1 | |
| 15:61555055:G:C | 15 | 61555055 | rs143096638 | chr15 | 61262856 | 0.015490291 | 3 | 1.774737275 | 0.356244746 | 4.98179214 | 6.29981E-07 | intergenic | RORA;VPS13C | dist=33553;dist=589535 | | 0.0093 | 0.0154 | 0.913405 | 1 | |
| 15:61567547:C:T | 15 | 61567547 | rs144327698 | chr15 | 61275348 | 0.01557767 | 3 | 1.784212495 | 0.357897115 | 4.985266493 | 6.18765E-07 | intergenic | RORA;VPS13C | dist=46045;dist=577043 | | 0.0097 | 0.0159 | 0.916644 | 1 | |
| 17:43780479:T:C | 17 | 43780479 | rs79985283 | chr17 | 45703113 | 0.0145 | 3 | 1.703938501 | 0.352883611 | 4.828613304 | 1.37487E-06 | intronic | MGC57346-CRHR1 | . | | 0.0211 | 0.0077 | 0.994938 | 1 | |
| 17:52366146:G:A | 17 | 52366146 | rs62072692 | chr17 | 54288785 | 0.023776699 | 5 | 1.836346208 | 0.394018804 | 4.660554743 | 3.15358E-06 | intergenic | KIF2B;TOM1L1 | dist=463573;dist=611906 | TOM1L1 | 0.0232 | 0.0371 | 0.819939 | 1 | Target Of Myb1 Like 1 Membrane Trafficking Protein |
| 18:39394472:G:C | 18 | 39394472 | rs113484345 | chr18 | 41814507 | 0.023024272 | 5 | 1.975849294 | 0.419524828 | 4.709731495 | 2.48043E-06 | intergenic | KC6;PIK3C3 | dist=293911;dist=140691 | PIK3C3 | 0.0229 | 0.0286 | 0.890728 | 1 | Phosphatidylinositol 3-Kinase Catalytic Subunit Type 3 |
| 19:46480407:T:C | 19 | 46480407 | rs4239537 | chr19 | 45977149 | 0.019417476 | 4 | 2.066073669 | 0.41173362 | 5.017986311 | 5.22159E-07 | intergenic | NOVA2;CCDC61 | dist=3750;dist=18312 | NOVA2;CCDC61 | 0.0835 | 0.0109 | 1 | 1 | NOVA Alternative Splicing Regulator 2; Coiled-Coil Domain Containing 61 |
| 20:6955818:G:T | 20 | 6955818 | rs146812516 | chr20 | 6975171 | 0.017223301 | 4 | 1.82640918 | 0.380591181 | 4.798874146 | 1.5956E-06 | intergenic | BMP2;LINC01428 | dist=194893;dist=171296 | BMP2 | 0.0161 | 0.0210 | 0.862977 | 1 | Bone Morphogenetic Protein 2 |
| 20:45720309:A:G | 20 | 45720309 | rs146684353 | chr20 | 47091670 | 0.032533981 | 7 | 2.316388446 | 0.498848234 | 4.643473279 | 3.426E-06 | intronic | EYA2 | . | EYA2 | 0.0232 | 0.0301 | 0.918602 | 1 | Eyes Absent 2 |
| 21:35375126:G:A | 21 | 35375126 | rs144499025 | chr21 | 34002825 | 0.007194175 | 1 | 0.846200633 | 0.183711704 | 4.606133495 | 4.10225E-06 | intergenic | LINC00649;MRPS6 | dist=38864;dist=70697 | MRPS6 | 0.0048 | 0.0076 | 0.538813 | 1 | Mitochondrial Ribosomal Protein S6 |
| 22:19080386:G:A | 22 | 19080386 | rs150428353 | chr22 | 19092873 | 0.024252427 | 5 | 2.149293074 | 0.429334633 | 5.006102254 | 5.55433E-07 | intronic | DGCR2 | . | DGCR2 | 0.0172 | 0.0230 | 0.783413 | 1 | DiGeorge Syndrome Critical Region Gene 2 |
| X:20935027:G:A | X | 20935027 | rs144178739 | chrX | 20916909 | 0.015194175 | 3 | 1.517881729 | 0.327834067 | 4.630030494 | 3.65612E-06 | intergenic | RPS6KA3;CNKSR2 | dist=650277;dist=457509 | CNKSR2 | 0.0231 | 0.0199 | 0.814532 | 1 | Connector Enhancer Of Kinase Suppressor Of Ras 2 |
| X:86181688:A:G | X | 86181688 | rs186620466 | chrX | 86926685 | 0.013970874 | 3 | 1.634757242 | 0.331433352 | 4.932386049 | 8.12312E-07 | intergenic | DACH2;KLHL4 | dist=94083;dist=591027 | DACH2;KLHL4 | 0.0162 | 0.0191 | 0.873035 | 1 | Dachshund Family Transcription Factor 2; Kelch Like Family Member 4 |

57 risk loci

one overlaps with 12 month: PCBD1;UNC5B

Thus 57+25-1 = 81 total risk loci

Gray cells = risk loci overlapping with Indianapolis-1 GWAS; 8/57 14.04%
