## Supplementary material for "Pharmacogenomics of steroid-induced ocular hypertension: relationship to high-tension glaucomas and new pathophysiologic insight": Suppl Table S9

**Supplementary Table S9. Functional Annotation Prioritized Genes**
**Functional grouping of prioritized target genes**

| Functional category | Prioritized target genes associated with SNPs of genome-wide significance | References to GC effects, ocular hypertension, or glaucoma | Prioritized target genes associated with SNPs of suggestive significance |
|---|---|---|---|
| Chromatin remodeling | *AGAP1*<br>*HDAC4*<br>*HDAC9*<br>*SPTY2D1* | ArfGAP protein that functions as a TF, priming chromatin for GR binding, thus influencing gene expression. (BioRxiv)<br>HDAC4 regulates GC signaling (PMID: 40186004). HDAC4 plays a role in neuroprotection and may be a therapeutic target for optic nerve damage in glaucoma (US Patent Publication Number WO/2024/010709)<br>HDAC9, a histone deacetylase, plays a role in the development of endothelial cell dysfunction and changes in vessel permeability in glaucoma (PMID: 37806089) | *AEBP2*<br>*ARID5B*<br>*ATXN1*<br>*CHD4*<br>*GATAD2A*<br>*JMJD7*<br>*MACROD2*<br>*MACROH2A1*<br>*RCBTB1*<br>*SMARCA2*<br>*TBL1XR1* |
| Transcription | *BEND7*<br>*PPARGC1A*<br>*ZNF737*<br>*ZNF728* | PPARGC1A expression stimulated by GCs; PPARGC1A can potentiate GC-induced transcription (OMIM entry 604517) | *AHR*<br>*ASXL1*<br>*CRTC1*<br>*CTBP2*<br>*DCANP1*<br>*DCT*<br>*DMRT2*<br>*E2F3*<br>*EBF2*<br>*EVX2*<br>*FOXN2*<br>*GTF2B*<br>*HOXD13*<br>*ID2*<br>*JADE1*<br>*JDP2*<br>*KLF5*<br>*KLF6*<br>*KLF12*<br>*KRBOX1*<br>*MAML3*<br>*MEOX2*<br>*MGA*<br>*NR2F1*<br>*NRC32*<br>*PITX2*<br>*PKNOX2*<br>*PLAG1*<br>*PLAGL2*<br>*POU3F2*<br>*PRDM10*<br>*PRDM15*<br>*SALL2*<br>*SHOX2*<br>*SLTM*<br>*SOX17*<br>*SWT1*<br>*TBX18*<br>*TFDP2*<br>*TFEC*<br>*TLE1*<br>*TRPS1*<br>*TTF1*<br>*ZBTB47*<br>*ZBTB49*<br>*ZFAT*<br>*ZKSCAN2*<br>*ZMAT3*<br>*ZNF514*<br>*ZNF536*<br>ZNF641<br>ZNF804B |

| | | | |
|---|---|---|---|
| Alternative RNA splicing | *RBFOX1* | | *RBFOX3* |
| GTPase pathways | *AGAP1*<br>*ARHGEF26*<br>*DIRAS2*<br>*PPM1H* | ArfGAP protein that functions as a TF, priming chromatin for GR binding, thus influencing gene expression. (BioRxiv)<br>ARHGEF26 knockdown ameliorated GC-induced myofibroblast transdifferentiation in TM cells, and prevented the development of GC-induced OH in mice (PMID: 39092889).<br>DIRAS2 is upregulated in lymphocytes following GC treatment. It is a negative regulator of mTOR2 signaling, triggering autophagy (PMID: 21733849). | *ARAP2*<br>*ARHGAP21*<br>*ARHGEF28*<br>*DLC1*<br>*RAC1*<br>*RASSF3*<br>*RND3*<br>*RTKN*<br>*TBC1D12* |
| Hippo-Yap signaling | *WWC1* | WWC1 activity is affected by the Hippo pathway and GC signaling (PMID: 30627107). | *SCHIP1* |
| TGFB superfamily signaling | *FST* | FST expression increased in TM of glaucoma patients, and expression is upregulated by TGFB2 (PMID: 23010638). | *FSTL5*<br>*SOSTDC1*<br>*VWC2* |
| cAMP signaling | *PRKD1* | | |
| Energy metabolism | *PPARGC1A* | PPARGC1A expression stimulated by GCs; PPARGC1A can potentiate GC-induced transcription (OMIM entry 604517) | *ACADSB*<br>*ACSF2*<br>*ACSL3*<br>*ADIPOR2*<br>*ALDH5A1*<br>*ALDH8A1*<br>*ALK*<br>*AMY2A*<br>*AMY2B*<br>*ARID5B*<br>*ASNS*<br>*CBR4*<br>*CHCHD7*<br>*CMC1*<br>*CRTC1*<br>*CTBP2*<br>*ETF3*<br>*ETNK1*<br>*LEPR*<br>*LPROT*<br>*LIPC*<br>*MOGAT1*<br>*MSRB2*<br>*MTX2*<br>*NT5C2*<br>*PC*<br>*PHYHIP*<br>*TMEM26*<br>*TMEM86A* |
| Immunity and inflammatory response | *CCR6* | | ENDOD1<br>HCK<br>TBK1<br>TLR2<br>TNFRSF13B |
| Ubiquitination | *ASB3*<br>*C4orf19* | | *ASB8*<br>*BTRC*<br>*FBXO7*<br>*FBXO11*<br>*NEDD4L*<br>*RNF2*<br>*RNF111*<br>*RNF144A*<br>*SH3RF1*<br>*SOCS7*<br>*TBL1XR1* |

| | | | |
|---|---|---|---|
| De-ubiquitination | *USP25* | | *OTUD3*<br>*USP18* |
| GPI Anchor Cleavage | *GPLD1* | | |
| Complement Inhibition & EMT | *CSMD1* | | *CSMD3* |
| Cell-ECM Adhesion | *SGCG* | | *FERMT1* |
| ECM Assembly | *COL11A1*<br>*KAZALD1* | | |

**Supplementary Table S9. Functional Annotation Prioritized Genes**

Pathway Enrichment Analysis

**NOTES**

Events identified by Reactome; enrichment scores determined using DAVID Bioinformatics functional annotation tool

Ordered by GWAS prioritized target gene enrichment score

DEGs: genes differentially regulated by glucocorticoids (GCs)

Prioritized target genes: see Table S3

Paired eye study DEGs:see Table S4

| Reactome Events | DAVID Enrichment Scores | | |
|---|---|---|---|
| | Prioritized target genes | Paired eye study responder DEGs | Paired eye study non-responder DEGs |
| Signal transduction: signaling by NOTCH | 1.86 | Not scored | Not scored |
| Disease: diseases of signal transduction by growth factor receptors and second messengers [cancer] | 1.86 | 0.51 | Not scored |
| Muscle contraction | 1.77 | 2.52 | Not scored |
| Muscle contraction: cardiac conduction | 1.77 | 2.52 | Not scored |
| Muscle contraction: cardiac conduction: Phase 0 – rapid depolarisation | Not scored | 0.99 | Not scored |
| Muscle contraction: cardiac conduction: Phase 1 - inactivation of fast Na+ channels | 1.77 | Not scored | Not scored |
| Muscle contraction: cardiac conduction: Phase 2 - plateau phase | Not scored | Not scored | Not scored |
| Muscle contraction: cardiac conduction: Phase 3 - rapid repolorisation | Not scored | Not scored | Not scored |
| Metabolism: metabolism of carbohydrates | 1.3 | 0.2 | Not scored |
| Metabolism: metabolism of carbohydrates: glycosaminoglycan metabolism | 1.3 | 0.2 | Not scored |
| Developmental biology: adipogenesis | 1.12 | Not scored | Not scored |
| Transport of small molecules | 0.92 | Not scored | Not scored |
| Transport of small molecules: ion channel transport | 0.92 | Not scored | Not scored |
| Transport of small molecules: stimuli-sensing channels | 0.92 | Not scored | Not scored |
| Disease: infectious disease: HIV infection | 0.89 | Not scored | Not scored |
| Disease: diseases of metabolism | 0.8 | Not scored | Not scored |
| Disease: diseases of glycosylation | 0.8 | Not scored | Not scored |
| Metabolism of proteins: post-translational protein modification: O-linked glycosylation | 0.8 | Not scored | Not scored |
| Signal transduction: signaling by Rho GTPases, Miro GTPases and ROBTB3 | 0.8 | 0.14 | 0.69 |
| Developmental biology | 0.75 | Not scored | 0.74 |
| Developmental biology: nervous system development | 0.75 | Not scored | 0.74 |
| Developmental biology: axon guidance | 0.75 | Not scored | Not scored |
| Vesicle-mediated transport | 0.71 | Not scored | Not scored |
| Vesicle-mediated transport: membrane trafficking | 0.71 | Not scored | Not scored |
| Vesicle-mediated transport: clathrin-mediated endocytosis | 0.71 | Not scored | Not scored |
| Immune system: cytokine signaling in immune system: signaling by interleukins | 0.67 | Not scored | Not scored |
| Hemostasis: platelet activation, signaling and aggregation | 0.67 | 0.73 | 0.67 |
| Signal transduction: signaling by VEGF | 0.65 | Not scored | Not scored |
| Cellular response to stimuli: cellular senescence | 0.55 | Not scored | Not scored |
| Circadian clock | 0.49 | Not scored | Not scored |
| Organelle biogenesis and maintenance | 0.49 | Not scored | Not scored |
| Organelle biogenesis and maintenance: mitochondrial biogenesis | 0.49 | Not scored | Not scored |
| Disease | 0.45 | 0.19 | Not scored |
| Disease: infectious disease | 0.45 | 0.19 | Not scored |
| Disease: viral infection | 0.45 | 0.19 | Not scored |
| Signal transduction: intracellular signaling by second messengers | 0.45 | 0.15 | Not scored |
| Signal transduction: signaling by GPCR | 0.44 | 3.98 | 2.2 |
| Signal transduction: signaling by TGFB family members | 0.35 | Not scored | Not scored |
| Cell-cell communication | 0.31 | 1.7 | 0.45 |
| Immune system: cytokine signaling in immune system | 0.27 | Not scored | Not scored |
| Gene expression (transcription): epigenetic regulation of gene expression | 0.25 | 0 | Not scored |
| Chromatin organization | 0.22 | Not scored | Not scored |
| Signal transduction: signaling by nuclear receptors | 0.19 | Not scored | Not scored |
| Signal transduction: MAPK family signaling cascades | 0.18 | 0.48 | Not scored |
| Immune system: adaptive immune system | 0.16 | Not scored | Not scored |
| Immune system: adaptive immune system: antigen processing & presentation | 0.16 | Not scored | Not scored |
| Metabolism of proteins: post-translational protein modification: neddylation | 0.16 | Not scored | Not scored |

| | | | |
|---|---|---|---|
| Gene expression (transcription) | 0.15 | 0 | 0.4 |
| Gene expression (transcription): RNA polymerase II transcription | 0.15 | 0 | 0.4 |
| Cell cycle | 0.08 | Not scored | Not scored |
| Metabolism of RNA | 0.03 | Not scored | Not scored |
| Metabolism | Not scored | Not scored | Not scored |
| Signal transduction: signaling by Wnt | Not scored | 3.45 | Not scored |
| Signal transduction | Not scored | 3.45 | Not scored |
| Extracellular matrix organization | Not scored | 1.99 | Not scored |
| Immune system | Not scored | 1.26 | Not scored |
| Immune system: innate immune system | Not scored | 1.26 | Not scored |
| Immune system: innate immune system: neutrophil degranulation | Not scored | 1.26 | Not scored |
